# Turnover of SARS-CoV-2 lineages shaped the pandemic and enabled the emergence of new variants in the state of Rio de Janeiro, Brazil

**DOI:** 10.1101/2021.07.20.21260890

**Authors:** Ronaldo da Silva Francisco Junior, Alessandra P Lamarca, Luiz G P de Almeida, Liliane Cavalcante, Douglas Terra Machado, Yasmmin Martins, Otávio Brustolini, Alexandra L Gerber, Ana Paula de C Guimarães, Reinaldo Bellini Gonçalves, Cassia Alves, Diana Mariani, Thais Felix Cruz, Isabelle Vasconcellos de Souza, Erika Martins de Carvalho, Mario Sergio Ribeiro, Silvia Carvalho, Flávio Dias da Silva, Marcio Henrique de Oliveira Garcia, Leandro Magalhães de Souza, Cristiane Gomes Da Silva, Caio Luiz Pereira Ribeiro, Andréa Cony Cavalcanti, Claudia Maria Braga de Mello, Cláudio J. Struchiner, Amilcar Tanuri, Ana Tereza R Vasconcelos

**Author notes:** Both authors contributed equally (Ronaldo da Silva Francisco Junior, Alessandra P Lamarca). Both authors coordinated this work (Amilcar Tanuri, Ana Tereza R Vasconcelos).

## Abstract

In the present study, we provide a retrospective genomic epidemiology analysis of the SARS-CoV-2 pandemic in the state of Rio de Janeiro, Brazil. We gathered publicly available data from GISAD and sequenced more 1,927 new genomes sampled periodically from March 2021 to June 2021 from 91 out of the 92 cities of the state. Our results showed that the pandemic was characterized by three different phases driven by a successive replacement of lineages. All stages occurred in distinct mortality and mobility contexts, with higher evidence of social distancing measures being observed in early pandemic and relaxed in the last two phases. Interestingly, we noticed that viral supercarriers accounted for the overwhelming majority of the circulating virus (> 90%) among symptomatic individuals in the state. Moreover, SARS-CoV-2 genomic surveillance also revealed the emergence and spread of two new variants (P.5 and P.1.2) firstly reported in this study. Altogether, our findings provided important lessons learned from the different epidemiological aspects of the SARS-CoV-2 dynamic in the state of Rio de Janeiro that have a strong potential to shape future decisions aiming to improve public health management and understanding mechanisms underlying virus dispersion.

## Introduction

One and a half years have passed since the outbreak of the Severe Acute Respiratory Syndrome-related Coronavirus 2 (SARS-CoV-2) virus in late 2019 in Wuhan, China. Currently, SARS-CoV-2 has accumulated a myriad of mutations and diversified into more than a thousand recognized lineages [1]. While vaccination is advancing across the world and different treatments are being tested, the imminent emergence of resistant and more aggressive variants haunts the scientific community. In fact, lineages such as Gamma (P.1), Alpha (B.1.1.7), Beta (B.1.351), and Delta (B.1.617.2) are already known to partially evade natural- and vaccine-induced antibody response as well as have an elevated transmission rate [2–4]. These Variants of Concern (VoC), named by the World Health Organization (WHO), have become widespread in the world replacing other lineages [5–7].

The emergence of novel SARS-CoV-2 variants is particularly worrying in countries with slow vaccination rates, such as Brazil. Since March 2020, the COVID-19 pandemic has caused over 19 million cases and the loss of 530.000 lives so far. Although Brazil corresponds to approximately 2.7% of the total world population, it accounts for more than 12,5% of deaths globally. By July 2021, only 15% of the Brazilian population has completed the vaccination program recommended. This small number allows the virus to circulate freely within the population, enabling the emergence of mutations and further diversification into new lineages. As a matter of fact, previous studies analyzing the spatial progression of the pandemic in Brazil revealed that the states of São Paulo and Rio de Janeiro were pivotal to the dissemination of lineages throughout the country, including VoCs [8–11]. Rio de Janeiro was also the background to the emergence and worldwide spread of the Variants of Interest (VoI) Zeta (P.2) [12].

The high diversity of SARS-COV-2 lineages in the state of Rio de Janeiro since the beginning of the pandemic and the presence of three VoC and one VoI in the state is the perfect scenario to verify whether there is a relationship between lineage displacement and a surge in the number of cases. Here, we provide the first temporal analysis of the lineage dynamics in the state of Rio de Janeiro. By combining epidemiological and genomic data, we described the pandemic in three main phases in the state. In addition to genomes publicly available in GISAID, we have sequenced 1,927 new genomes from 91 out of the 92 cities in Rio de Janeiro, sampled periodically between March and June of 2021. Finally, we report the emergence of two new variants, P.1.2 and P.5, with the reconstruction of the within-country and worldwide dispersion.

## Materials and Methods

### Sampling, genome extraction, sequencing, and assembly

Samples from patients with SARS-CoV-2 positive nasopharyngeal RT-PCR were collected at the Noel Nutels Central Laboratory (LACEN-RJ) and Unidades de Apoio ao Diagnóstico da Covid-19 (UNADIG-RJ). The 1,927 new genomes sequenced in this study were collected between March 24, 2021 and June 06, 2021 from 91 municipalities in the state of Rio de Janeiro/Brazil. Patients were aged between 0 and 91 years old, being 47.69% male and 52.31% female. Extraction of the genetic material was performed at the Molecular Virology Laboratory (LVM-UFRJ) with QIAamp or MagMAX Viral / Pathogen Nucleic Acid Isolation kits and KingFisher automatic platform. Annealing of cDNA was conducted with 8,5 µl of viral RNA extracted from each sample. Libraries were constructed at the DFA/LNCC Genomics Unit with Illumina COVIDSeq Test (Illumina), according to the manufacturer’s protocol. Purification was then conducted using 5 µl of each library combined, and the TapeStation (Agilent) system was used for quality control. We employed the NextSeq 500/550 Mid Output Kit v2.5 (300 Cycles) to generate reads of 2×149 bp in the NextSeq (Illumina). Sequence analysis, consensus building, and variant calling were performed with DRAGEN COVID Lineage v3.5.1. The study was approved by the Ethics Committee (30161620.0.1001.5257 and 34025020.0.0000.5257). All the assembled genomes were then submitted to the GISAID database and made publicly available. Finally, the newly assembled genomes were classified according to the Pango Lineage classification using the PangoLEARN model database (v3.5.3).

### Epidemiology, human mobility, and viral load analysis

Epidemiological data were retrieved from the Centro de Informações Estratégicas e Resposta de Vigilância em Saúde (CIEVS-RJ) of the Secretaria de Saúde do Estado do Rio de Janeiro. We then estimate the time varying reproduction numbers (Rt) from the Epidemic Curve using the EpiEstim package in R [13]. To further characterize the mobility patterns during the pandemic, we downloaded mobility data for the cities in the state of Rio de Janeiro from Google Community Mobility Reports (available at: https://www.google.com/covid19/mobility/) between 1 March 2020 to 6 June 2021. Six indices for variation in the expected value of the time typically spent in residential, transit stations, parks, groceries and pharmacies, retail and recreational, and workspaces were summarized using the 7-day moving average considering the whole state.

To verify whether a small group of samples can be overrepresented the Ct values in our dataset. We selected 1,119 out of the 1,933 samples sequenced and retrieved the cycle threshold (Ct) values of the target virus genes and the endogenous gene RnaseP. We performed a relative quantification of the viral loads (RQVL) using the method of 2^-ΔCT^. The dataset table with sample ID, sex, age, and RQVL colunms was descend ordered by the RQLV values. We calculate the ratio of 2%, 10%, and 20% of the samples relative to the total sum of the RQVL. We also performed a generalized linear model using the gamma distribution with the inverse link function to verify the significance of sex and age in the RQLV distribution using stats package in R [14].

### Evolutionary analyses

In order to infer their phylogenetic tree, we have retrieved all genomes from the state of Rio de Janeiro available in GISAID by June 21^st^,2021 that were longer than 29,000 bases, had high coverage (as defined by GISAID) and had complete collection dates (*n* = 3,029). We used MAFFT 7.475 with --auto and--addfragments options [15,16] to align this dataset to the WH01 (EPI_ISL_406798) genome sampled in 2019 from Wuhan, China. The 3’ and 5’ untranslated regions were then trimmed. The subsequent alignment was used to construct the maximum likelihood tree using IQ-TREE 2 [17] with the GTR+F+I+G4 substitution model, selected with the ModelFinder algorithm [18]. Assuming a constant evolutionary rate of 8 × 10^−4^ substitutions/site/year, we rescaled the branch lengths to dates with TreeTime [19]. Tips that deviated from the clocklike model are ignored by TreeTime (Figure S1A) and were removed from the final tree.

To reconstruct the evolution of the new P.1.2 lineage described in this work, we have retrieved all sequences from this lineage available in GISAID by June 21^st^ and aligned them to our newly-sequenced samples. As an outgroup, we selected the two oldest P.1 sequences available from the state of Amazonas. We then reconstructed the maximum likelihood tree with IQ-TREE 2 and the GTR+F+I model selected with ModelFinder algorithm. We then tested the sequences’ conformity to the molecular clock model by analyzing the correlation between root-to-tip distances and tip dates and removed from the alignment all sequences that were not included within the 95% confidence interval of the regression (Figure S1B). As the two outgroup sequences selected were removed from the final dataset, we selected as new outgroups two sequences from the P.1 clade positioned closest to P.1.2 according to the maximum likelihood phylogeny of all sequences from Rio de Janeiro.

Divergence dates were estimated using a Bayesian approach with BEAST 1.10.4 [20]. Models selected for the analyses were the GTR+F+I with estimated base frequencies, the GMRF Skyride population model, and the strict clock model. The clock rate prior was defined as a uniform distribution with 6E-4 and 1E-3 as minimum and maximum values. The MCMC was run through 200,000,000 steps with sampling every 10,000th and a burnin of 10%. Lowest ESS was 204, from the “treeLength” parameter. In order to evaluate the impact of biased sampling through time on population inference, we also applied the Bayesian Nonparametric Phylodynamic Reconstruction model with and without preferential sampling correction on the consensus tree of P.1.2 using the R package phylodyn [21].

The consensus tree was used as fixed topology and branch lengths to reconstruct the locality of its ancestor nodes. We used the Symmetric trait substitution model with the Bayesian Stochastic Search Variable Selection approach for rate estimation in BEAST 1.10.4. The MCMC chain was 10,000,000 steps with sampling every 10,000th and a burnin of 10%. Lowest ESS value was 166, from the “loc.indicators.Argentina.Paraguay” parameter. Ancestor locations inferred were then converted into coordinates and mapped using the SERAPHIM package [22] in R software.

We also reconstructed the evolutionary tree of the new lineage P.5. To accomplish this we identified all the sequences available in the GISAID database by June 21^st^ that carried the mutations that characterize the lineage. We aligned these sequences to the WH01 genome using MAFFT 7.475 with --auto and --addfragments options and trimmed the 3’ and 5’ untranslated regions. The maximum likelihood tree was then inferred using IQTREE with the GTR+F+I model, selected by the ModelFinder algorithm.

### Structural analysis of spike protein of P.1 and P.1.2 lineages

The primary sequences of Spike protein from P.1 lineage and P.1.2 subclade (A262S) were submitted in the Swiss-Model web server (https://swissmodel.expasy.org/) to obtain the best templates and build their 3D structures. The stability analysis was performed using the Stability function from FoldX tool [23]. Salt bridges were measured using the EBRIS web server [24]. The docking analysis was performed between structures of P.1 and P.1.2 subclade in HDOCK [25]. We evaluated the Spike protein against ACE2 protein and four types of antibodies anti-Spike extracted from CoV-AbDab database [26]. The human antibodies were selected based on their preferences for distinct binding locations in Spike: subunits S1, S2, S1-S2, S1-RBD, and S1-notRBD. The ACE2 structure was extracted from a complex reported in PDB with RBD region. We compared the five complexes generated for each protein using the residues found in contact with ligand and belonging to the hotspot predicted area. In the first part of the docking analysis, we compared the amino acids contacts between the receptor (P.1/P.1.2) and the five ligands. The binding affinity of each complex was measured with the Prodigy tool [27]. We searched for hot spots in the receptors using the KFC (Knowledge-based Fast atomic density evaluation and Contacts) webserverer [28]. The potential profiles considering P.1.2 subclade structure and P.1 were compared by PDB2QR [29] and APBS. Visualization and structure manipulation was conducted using PyMol.

## Results

In early March 2020, Rio de Janeiro’s Secretary of Health confirmed the first case of COVID-19 in the state. Since then, the SARS-CoV-2 virus has rapidly spread causing more than 923,234 cases and 54,125 deaths in the state of Rio de Janeiro only, with multiple events of the public health system collapsing. The highest prevalence of COVID-19 cases was observed among the central and north regions in the state (Table S1). We also noticed a high number of cases in the metropolitan region, known to have high population density, workers commuting daily between neighboring cities and an elevated number of interstate corporate travels. A statistically significant difference was found across the number of cases and deaths between females and males (OR 0.73, 95% CI: 0.71-0.74; p-value<2.2×10^−16^; Figure S2A). Though more females got infected, male mortality was higher. Among confirmed cases, typically younger subjects ranging between 30-49 years old (y.o) represented the highest occurrence rate. Nevertheless, the percentages of mortality surpassed 11% in patients aged >60 y.o reaching more than 79% in the oldest group (Figure S2B).

The epidemiological findings suggest three different exponentially growing phases associated with the disease in the state (Figure 1A). Firstly, after the epidemic outbreak in March 2020, the number of cases increased, reaching a peak in May 2020 when it started to decline. In early November 2020, a period marked by the intensification of the pre-election campaigns, a new increase in the number of cases was detected and peaked around December 2020, rapidly dropping in January 2021. The third phase took place right after the Carnival holiday on February 16, with the maximum number of cases observed in March 2021. The estimation of the real-time reproduction number (Rt) in the state assumed values between 0.78 to 2.40 during the epidemic, being most of the time close to 1-1.3 (Figure 1B). These values suggest a persistent transmission of SARS-CoV-2 but not in an explosive manner. Interestingly, we observed a 2-fold increase in the daily number of cases reported in the second [mean: 2509, sd: ±1177] and third [mean: 2505, sd: ±680] phases when compared to the first one [mean: 1446, sd: ±709].

**Figure 1.**
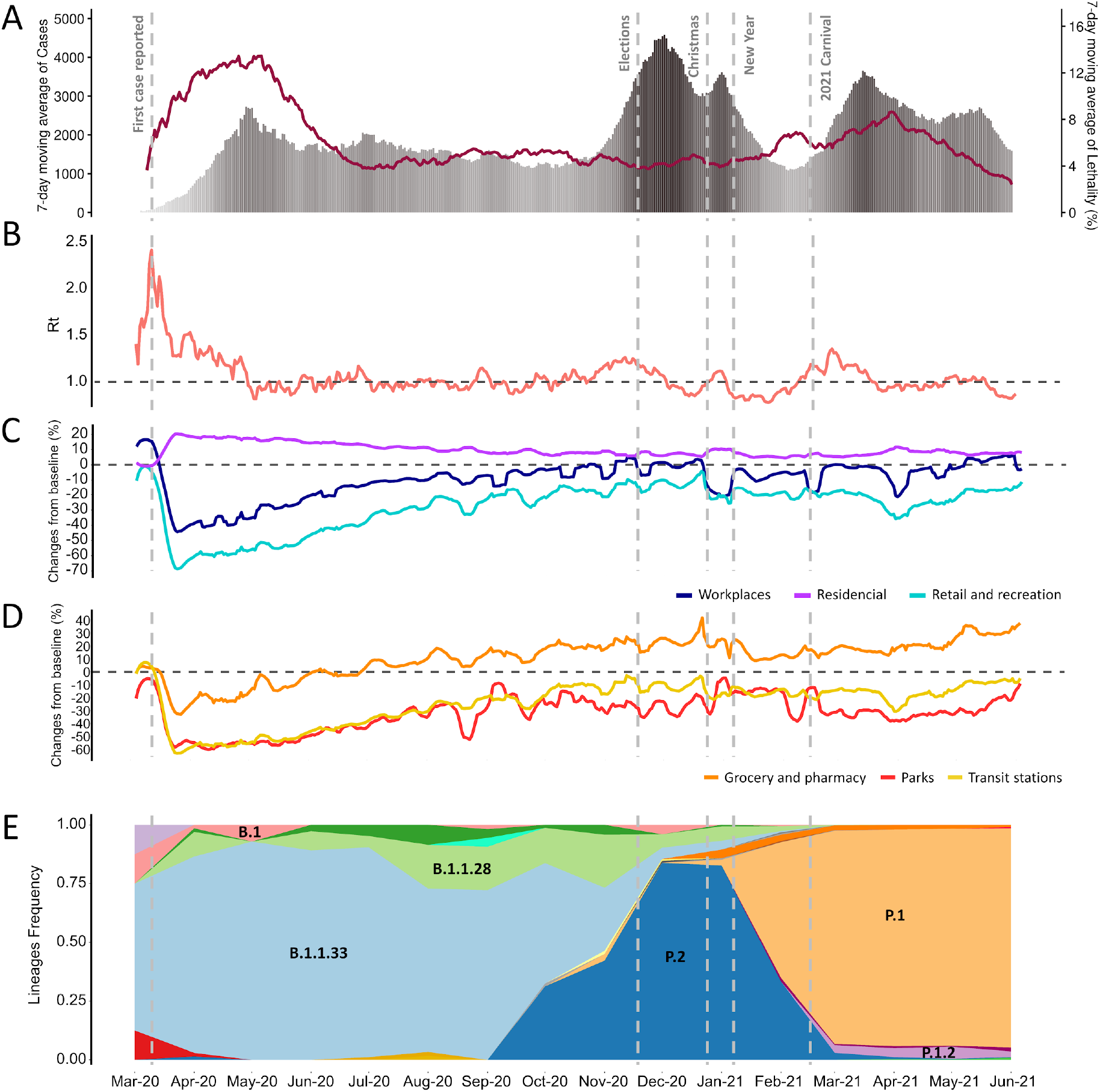
Epidemiological findings, mobility-driven changes and successive lineage replacement during SARS-CoV-2 epidemic in the state of Rio de Janeiro. A) 7-days moving average of cases and lethality reported from March 1, 2020 to June 6, 2021. Dashed lines indicate first reported case, elections, Christmas, new year and carnival 2021, respectively. The degree in gray scale shows the increases in the number of cases in each phase. B) Estimation of the real-time reproduction number (Rt). C-D) Google Community Mobility data reports with changes in time spent in workplaces, residential, retail and recreation, grocery and pharmacy, parks, and transit stations. E) Temporal distribution of the relative frequencies of SARS-CoV-2 lineages in the state of Rio de Janeiro.

The pairwise comparison between the mortality rate across the different phases revealed substantial differences among the first and second stages (p-value: 2.2×10^−4^, Wilcoxon test - Benjamini-Hochberg p-value correction), the second and third (p-value: 3.2×10^−05^) and the first and third phases (p-value: 7.9×10^−10^) (Figure S2C). Differential mortality rate was also identified among the age groups (Figure S3, Table S2), the most recent phase seemed to be more lethal among subjects ranging between 20 to 69 y.o than the others. On the other hand, the mortality in the second phase was consistently decreased among young ages and increased in subjects older than 79 y.o (Figure S3).

Remarkably, mobility-driven changes significantly accounted for the virus dispersion in the state (Figure 1C and D). The early phase was mainly characterized by a sharp decrease in activities outside home such as time spent in workplaces (−64%), retail and recreation (−74%), official national park visits (−72%), transit station (−67%), and going to grocery and pharmacy (−40%) in late March 2020. In the same period, we also observed an increase in the expected value of the typical day (baseline) in the time spent at home (+25%), evidencing the social distance measures in Rio de Janeiro. These numbers rose close to the baseline in workplace activities and decreased in households during the second and third phases as the social distancing measures were relaxed. We also noticed a continuous elevation in the percentage of going to grocery and pharmacy from -40% (1^st^ phase) to +74% (3^rd^ phase) over time, suggesting that people moved to these places more often than the baseline frequency. Therefore, both phases occurred in distinct mobility contexts with the most prominent change being reported in the first phase followed by similar patterns in the other two.

After the introductory events in early March 2020, independent efforts to sequence SARS-CoV-2 genomes from Rio de Janeiro resulted in 3,932 sequences publicly available in the GISAID database until June 2021. This number corresponded to an approximate overall average of four sequences per 1000 cases, reaching a peak of 1% of all reported cases sequenced in April and May 2021 (Table S3). We observed that the pandemic phases were characterized by successive replacement of SARS-CoV-2 lineages in the state (Figure 1E). The first phase was predominantly driven by the lineage B.1.1.33, which was the most prevalent strain circulating during eight months, from March to October 2020, being gradually replaced by P.2 during the second phase. In May 2020, B.1.1.33 achieved the higher relative frequency of approximately 92% of the genomes deposited in GISAID, coinciding with the peak reported during the first phase. Moreover, the dominance of P.2, from November 2020 to January 2021, was preceded by a continuous increase in the frequency of its ancestor, B.1.1.28, that was already circulating in a small frequency in the state since April 2020. The peak of reported cases observed in the second phase matches the highest prevalence of P.2 in December 2020. After this, P.1 became the dominant lineage reaching a frequency greater than 90% from March 2021, matching the peak of cases in the third phase. Therefore, the introduction and emergence of each lineage occurred concomitantly with the increase in the number of cases.

From the 1,927 new genomes sequenced in this study after later March 2021, 92.42% were assigned to P.1 (Gamma) lineage. We then sought to investigate the distribution of SARS-CoV-2 viral loads within a population of 1,119 samples randomly selected from the state. The cumulative distribution of the relative quantification of the viral loads (RQVL) showed that 2% (n = 22, group A) of the individuals carried 60% of the circulating virions in the period analyzed (Figure 2A). This value increases to 92.4% of the virions harbored by just 10% (n = 112, group B) of the population (Figure 2A). Furthermore, 20% (n = 223, group C) of our samples represented 98.2% of the viral loads of our dataset (Figure 2A). Next, we tested the contribution of age, sex and the interaction between age and sex to the viral loads using a gamma-generalized linear model (GLM). Both groups (A, B, and C) had approximately half of samples from each sex, nevertheless, a higher relative viral load were reported among males within-group (Figure 2 B-D). From the 22 samples (11 males and 11 females) present in group A, the viral load was 14.8% greater in males when compared to females. Among males in group B and C, our model showed that subjects aged > 40 y.o carried most of the population-circulating virions, and the older the higher the viral load observed (Table S4; Figure S4). No statistical differences were observed among females age groups.

**Figure 2.**
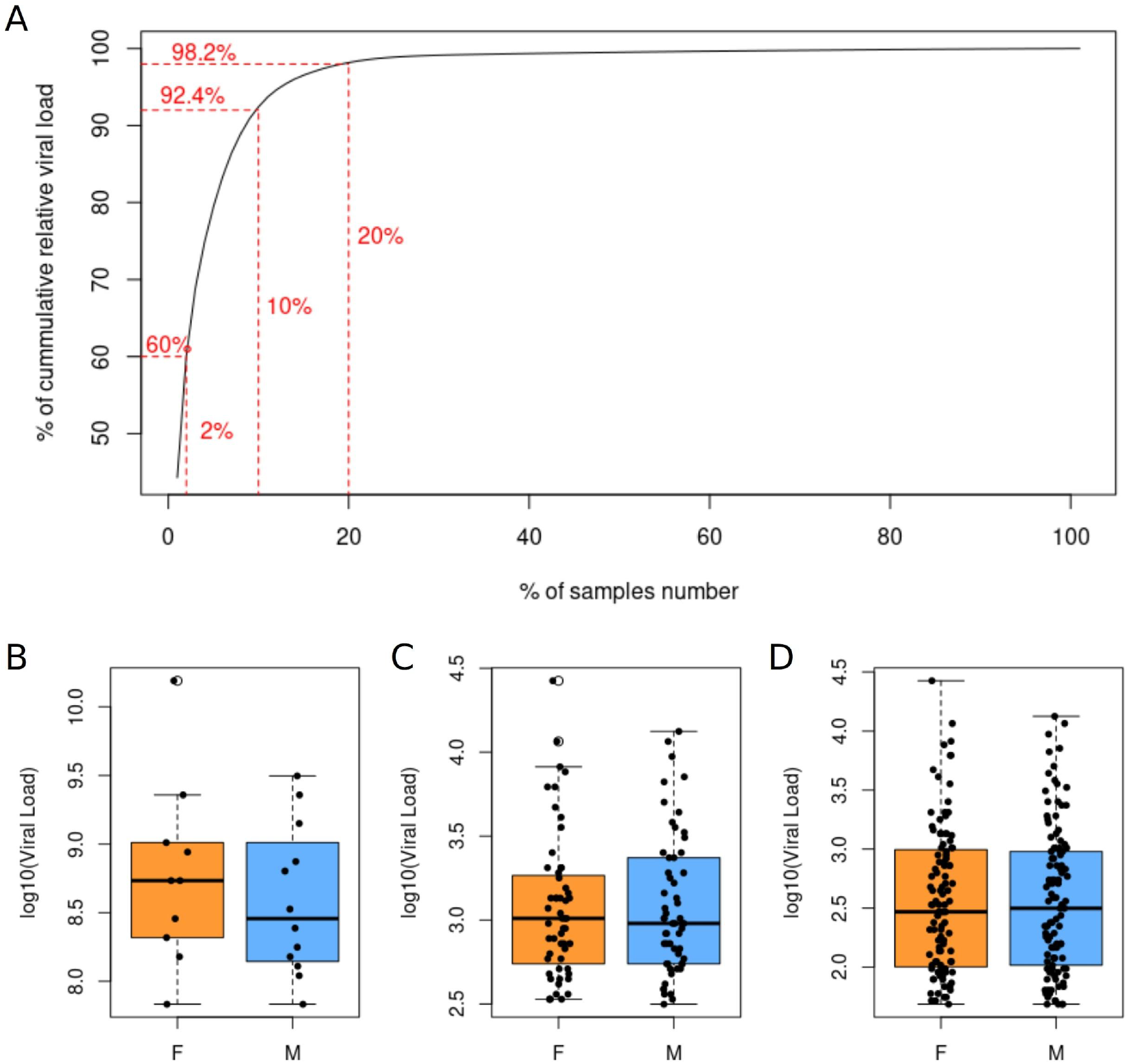
Distribution of the relative quantification of the SARS-CoV-2 viral loads (RQVL) within our population of 1,119 samples. A) Cumulative percentage of total circulating virions from nasopharynx RQVL within our population. B-D) RQVL comparison between female (orange) and male (blue) in groups A (2%), B (10%) and C (20%), respectively. Gamma-generalized linear model showed statistical significance difference among the sex of the supercarriers in each group with a higher relative viral load reported among males as demonstrated in Table S4.

SARS-CoV-2 genomic surveillance in the state of Rio de Janeiro also allowed the identification of two new variants (P.5 and P.1.2) first reported in this study (Figure 3A). The new variant assigned as P.5 by PANGO descends from B.1.1.28 with the possible origin being the Brazillian state of São Paulo. P.5 harbors 12 nonsynonymous lineage-defining mutations (ORF1ab: T1637I, A3209V, Q3729K and P4337L; S: F2L, Q14K, T95I, E484Q and N501T; N: G215V) and by June 21^st^ 2021 was restricted to Rio de Janeiro and São Paulo (Figure S5). P.1.2 derived from P.1 and was characterized by a constellation of five new mutations (ORF1ab: synC1912T, D762G and T1820I; ORF3a: D155Y; and N: synC28789T) besides the P.1 lineage-defining mutations. This variant has already spread to Argentina, Australia, Chile, England, Netherlands, Paraguay, Portugal, Spain, Uruguay and the United States. We estimated the first divergence within P.1.2 to have occurred within the Brazillian state of São Paulo, between late 2020 and early 2021 (Figure 3B). By the end of January, the lineage had already spread to all of the Brazilian states sampled, including an event of secondary dispersal (Goiás to Maranhão) and of reintroduction to São Paulo from Sergipe (Figure 3C). The lineage was also first introduced in most countries in late January; it arrived in Spain and Paraguay only in early-March (Figure 3D). This early spread is coincident to a rise in effective population size of P.1.2 lineage (Figure S6A), though this value stabilizes around late February. Temporal bias do not seem to have influenced the inference of this fluctuation (Figure S6 B-C)

**Figure 3.**
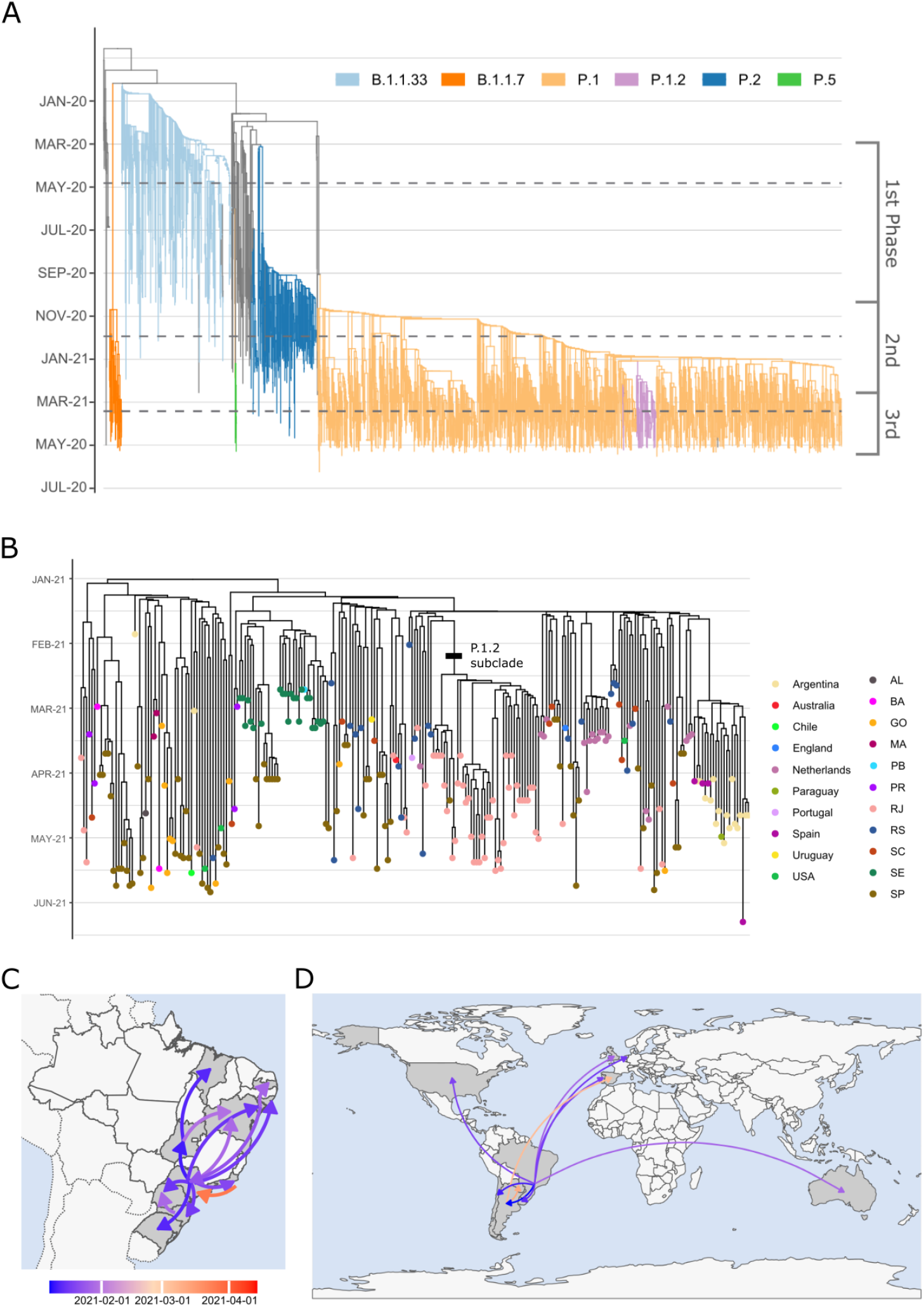
Evolutionary relationships between major lineages present in Rio de Janeiro and the evolution and dispersal of the new lineage P.1.2. A) Time tree of 3,029 genomes from the state of Rio de Janeiro, with lineages of interest highlighted by branch color. B) Divergence dates between P.1.2 sequences indicate that the lineage originated at the first days of 2021. Black bar signals the subclade with A262S mutation in Spike. Colored dots show the location of sampling. C-D) National and international transmission routes inferred for the dispersion of P.1.2. Color of arrows corresponds to the date of the first dispersal event between locations.

P.1.2 was already diversificated in a subclade that emerged in Rio de Janeiro around mid-February, being the subtype more frequently detected in the state. This subclade carries the additional mutations A262S in the Spike protein gene and the missense variant L83F in ORF3a. To further characterize the impact of A262S mutation, we performed a comparative structural analysis between the Spike protein from P.1 and the subclade of P.1.2. The 3-dimensional structure of P.1 and P.1.2 (A262S) was modeled by selecting the 7KRS model as template in PDB. This model has 3.2Å of crystal resolution with 99.09% and 99.14% of identity to P.1 and P.1.2, respectively. Quality assessment analysis showed that both predictions achieved satisfactory values by reaching 93.9% of atoms contained in P.1 and 93.84% in P.1.2 in allowed regions of the Ramachandran plot. Despite the few differences in RMSD values (P.1: 0.11 and P.1.2: 0.14) when compared to the template, stability analysis showed that P.1.2 is less stable than P.1 according to FoldX (Table S5). This difference was confirmed by estimating the number of salt bridges in both models, P.1 had more salt bridges (n = 53) than P.1.2 (n = 48), which demonstrates that P.1 folding is less flexible. We also investigated possible effects in the electrostatic surface caused by the change of an Alanine to Serine in position 262. Figure S7A and B shows that both proteins present the same potential surface although in the mutated region the cavity becomes more neutral in P.1.2, possibly altering the binding-site of the region. Interaction between both models with four antibodies and ACE2 surface protein using docking and affinity binding prediction (Table S6; Figure S7) showed that P.1.2 was more efficient in antibody interaction than P.1. P.1 also performs more stable bonds with ACE2 (−18.2 kcal/mol) than P.1.2 (−17.8 kcal/mol) and is predicted to enter easier in the cell to establish virus infection. Furthermore, we detected an active binding site in the complex S1notRBD-P.1.2 that is a candidate to alter the binding site contacts in the cavity (Figure S7).

## Discussion

In this study, we gathered epidemiological and genomic data from the outbreak of SARS-CoV-2 until June 2021 and conducted a thorough examination of the pandemic progression in the state of Rio de Janeiro, Brazil. Before this work, the state had been facing issues to keep a continuous genomic surveillance resulting in different proportions of positive samples being sequenced over time. To overcome the limitation in the number of sequences available for the state, we generated 1,927 novel SARS-CoV-2 genomes sequenced biweekly between March and June 2021, sampled from residents of 91 out of the 92 cities of Rio de Janeiro. These sequences represent ∼50% of the genomes publicly available in GISAID from the state so far, notably reaching a sequencing ratio of ∼1:100 cases reported during April and May 2021. We then analyzed lineage dynamics in Rio de Janeiro as well as the emergence and dispersion of new variants.

Three main phases defined the pandemic in the state of Rio de Janeiro, each of them represented by a different lineage. Remarkably, all phases occurred with distinct patterns of transmissibility and mortality. Whereas the daily number of cases increased during the second and third phase, more people died in the first and third ones. We hypothesized that lack of knowledge about the disease and resources to treat patients in early pandemic may explain such severity in the first phase [30–32]. After almost a year of pandemic, a significant amount of information had been already gathered about the virus and the disease, certainly improving patient care. We have observed a switch in the death profiles over the course of pandemic, while deaths were more restricted to old groups and people with comorbidities in the first phase, a substantial increase of young people was reported at the third wave. Prioritizing the vaccination of elderly people might have had a strong impact on reducing the mortality of this group [33], though it does not explain the rise in mortality in younger ages. Thus, it is plausible to assume that the evolution of new viral phenotypes are correlated to the rejuvenation in mortality observed in the third stage [34–37].

The displacement of SARS-CoV-2 lineage across each phase suggests that the new successive lineage had higher adaptive value than the last one [4,6]. Indeed, the three lineages progressively accumulated aggressive key mutations associated with higher transmissibility or evasion to the host immune system response: namely, B.1.1.33 carried D614G, P.2 harbors D614G and E484K, and P.1 carries N501Y in addition to the two previous mentioned. The observed synchronicity between lineage substitution and a rise in the number of cases may be a reflection of this increase in adaptability of the virus to human biology. This dynamic raises a red flag to the aggressiveness of the next dominant lineage. Though lineage displacement is a well-documented aspect of pathogen evolution [38–40], the mechanisms and models underlying the influence of this event on the resurgence of local outbreaks of COVID-19 is still poorly understood. Nevertheless, displacement of lineages by B.1.1.7, B.1.351, B.1.617.2 and P.1 has been reported repeatedly across the world [5,7,41–44].

We caution that direct association between each phase and the lineage distribution must be interpreted considering the temporal bias in genome sequencing, especially during the first phase (March-October 2020). Nevertheless, previous studies indicate that B.1.1.33 was widely disseminated throughout the country in the early pandemic phase [8,9], supporting our findings for Rio de Janeiro. Furthermore, the concentration of samples in the metropolitan region challenges the exact dispersion routes within-state as also highlighted by Wilkinson and colleagues [45]. Although we consider the circulation of lineages an important component that characterized each phase, it does not fully explain the number of cases and deaths, once the management of the epidemic also changed over time as demonstrated by the mobility patterns.

Our results demonstrated that returning to work-related and outside-home activities close to the pre-pandemic levels preceded the rise in transmission rates, especially in the second phase. The surge in cases, therefore, could also have been triggered by the gradual flexibilization of social distancing measures [46]. As the number of cases never shrank to pre-pandemic levels in either phase, the slight increase in transmission rates caused by this flexibilization would be enough to create the exponential rise in the virus population. Though the “lineage” and the “mobility” hypotheses could be perceived as opposing narratives, we reiterate that both endogenous and exogenous factors control the threshold of a new surge in cases. The introduction of more aggressive lineages requires the enforcement of harder non-pharmacological interventions to control the local outbreak. So far, the only way to circumvent this relationship is the reduction in viral transmission rates through vaccination. Further epidemiological modelling is still needed to better evaluate the influence of all mechanisms involved in this dynamic.

An important aspect of the disease dynamic revealed by this study was that viral supercarriers accounted for the overwhelming majority of the circulating virus in the state. Recent studies demonstrated that transmission of COVID-19 is more likely to occur in individuals with higher viral load [47–49]. Given that most of the samples in the period analyzed were assigned to P.1, such a mechanism could also shed light on the rapid spread of this VoC. Considering that the selection of RT-PCR positive samples for sequencing in our analysis is based on a pre-filtering of samples with Ct values < 27, the values of 60% and >90% of viral load of the supercarries could represent an even bigger amount of the circulating virus in the population.

Indeed, Yang et al recently described a bigger fraction of viral load (90%) being carried by just 2% of the infected individuals [50], which corroborates our findings. Though the observed viral load in our cohort could be explained by the different stages of infection of each patient, it is known that this measure is significantly variable between individuals [51,52]. Overall, supercarriers may play an important role in the dynamic of virus dissemination in the state.

It is well known that increasing viral replication rate accelerates the emergence of new mutations and, therefore, the diversification into new lineages. It is not surprising that the origin of P.1.2 is simultaneous to the countrywide propagation of P.1 in early January 2021 [10,53,54], when this rate is expected to be high. The new mutations observed in P.1.2, though, do not indicate that this lineage has a higher adaptive value than P.1. Indeed, structural protein modelling of its sub-clade suggests a higher instability and better docking with antibodies than P.1. The fast dispersal of P.1.2 within Brazil could be attributed to an opportunistic event. As P.1 was introduced in Brazilian states at the end of 2020, it induced an immune response in the host population, which temporarily prevents reinfection and limits the number of available hosts. Therefore, the P.1.2 population could only grow before P.1 had not achieved such high frequency, as observed after January [39]. Same behaviour could explain the emergence and spread across the state observed by the lineage P.5. A new opportunity of growth opens after infection-induced immune response fades in the host population, which can only be prevented through vaccination.

Finally, learning from past efforts to control the pandemic shapes the future decisions towards an improvement of the national public health, especially when embracing the findings reported by the scientific community. Currently, as this manuscript is being written, the state is at imminent risk of experiencing a fourth phase of COVID-19 epidemic due to the introduction of VoC delta (B.1.617.2) as recently reported by us [55]. Therefore, understanding the different epidemiological aspects controlling the SARS-CoV-2 dissemination, such as host mobility patterns and viral evolution, is essential to elaborate effective intervention measures that reduce the number of affected individuals. These evaluations are particularly vital in regions with high-density populations, which are prone to spread viruses worldwide.

## Data Availability

The data generated in this study is publicly available in Gisaid (www.gisaid.org) the identifiers of each sequence are reported in Table S7.

## Acknowledgement

We would like to thank all the authors and administrators of the GISAID database, which allowed this study of genomic epidemiology to be conducted properly. A full list acknowledging the authors publishing data used in this study can be found in the Supplementary File. We are very grateful to Dr. Luiz Max Fagundes de Carvalho from Fundação Getúlio Vargas for the kind aid with Bayesian models and analyses.

This work was developed in the frameworks of Corona-ômica-RJ (FAPERJ = E-26/210.179/2020). A.T.R.V. is supported by CNPq (303170/2017-4) and FAPERJ (E-26/202.903/20); A.T. by FAPERJ E-26/010.002434/2019 and E-26/210.178/2020 R.S.F.J is a recipient of a graduate fellowship from CNPq, A.P.L is granted a post-doctoral scholarship (DTI-A) from CNPq. C.J.S is supported by FAPERJ and CNPq. We acknowledge the support from the Rede Corona-ômica BR MCTI/FINEP affiliated to RedeVírus/MCTI (FINEP 01.20.0029.000462/20, CNPq 404096/2020-4).

