## Supplementary Material for "Turnover of SARS-CoV-2 lineages shaped the pandemic and enabled the emergence of new variants in the state of Rio de Janeiro, Brazil"

[#]Both authors contributed equally

[a]Both authors coordinated this work

[1] Laboratório de Bioinformática, Laboratório Nacional de Computação Científica, Petrópolis, Brazil.

[2] Departamento de Genética, Instituto de Biologia, Universidade Federal do Rio de Janeiro, Rio de Janeiro, Brazil.

[3] Unidades de Apoio ao Diagnóstico da Covid-19, Rio de Janeiro, Brazil.

[4] Secretaria Estadual de Saúde do Rio de Janeiro, Rio de Janeiro, Brazil.

[5] Secretaria Municipal de Saúde Rio de Janeiro, Rio de Janeiro, Brazil

[6] Laboratório Central de Saúde Pública Noel Nutels, Rio de Janeiro, Brazil.

[7] Fundação Getúlio Vargas, Rio de Janeiro, Brazil

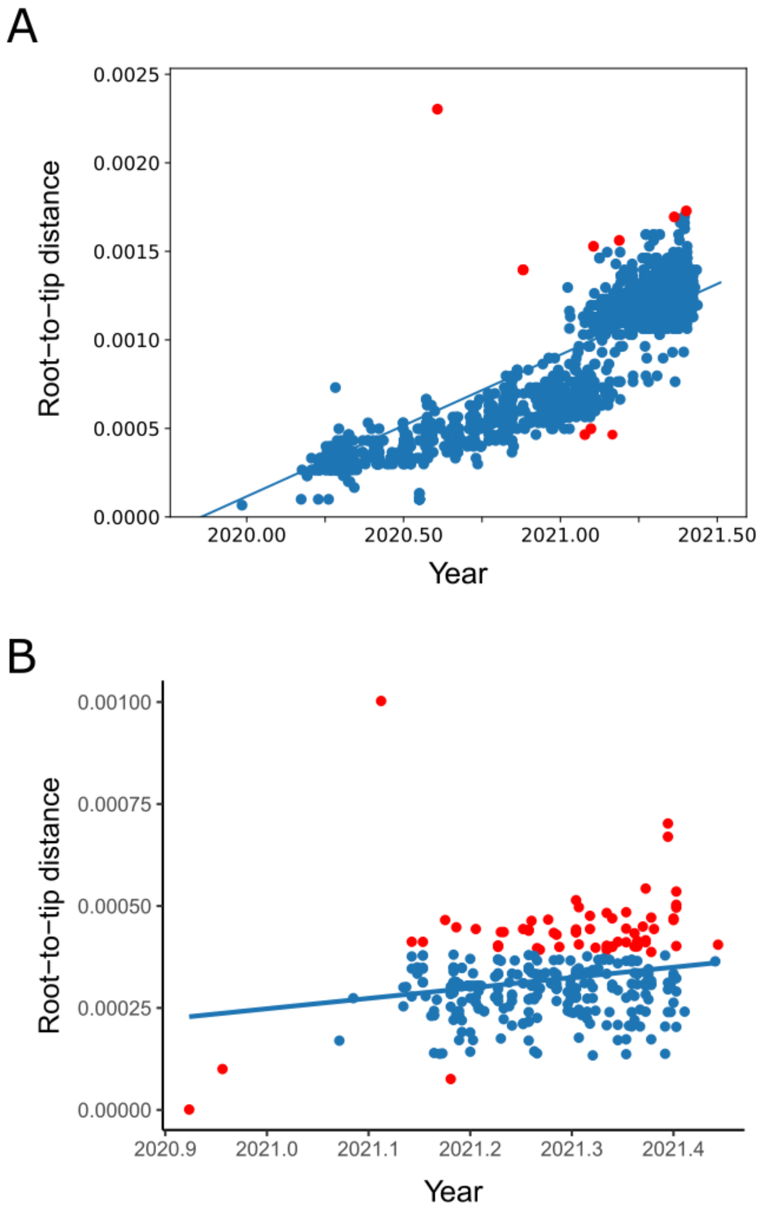


**Figure S1. Correlation between root-to-tip distance and tip sampling date.** A) Root to tip distance for the phylogenetic tree of genome sequences from Rio de Janeiro (Figure 1A). In red, the tips ignored by TreeTime to rescale the tree into dates. B) Root to tip distance for the P.1.2 tree reconstructed using maximum likelihood. In red, the tips that were not contained within the 95% confidence interval and were removed for the bayesian analysis.

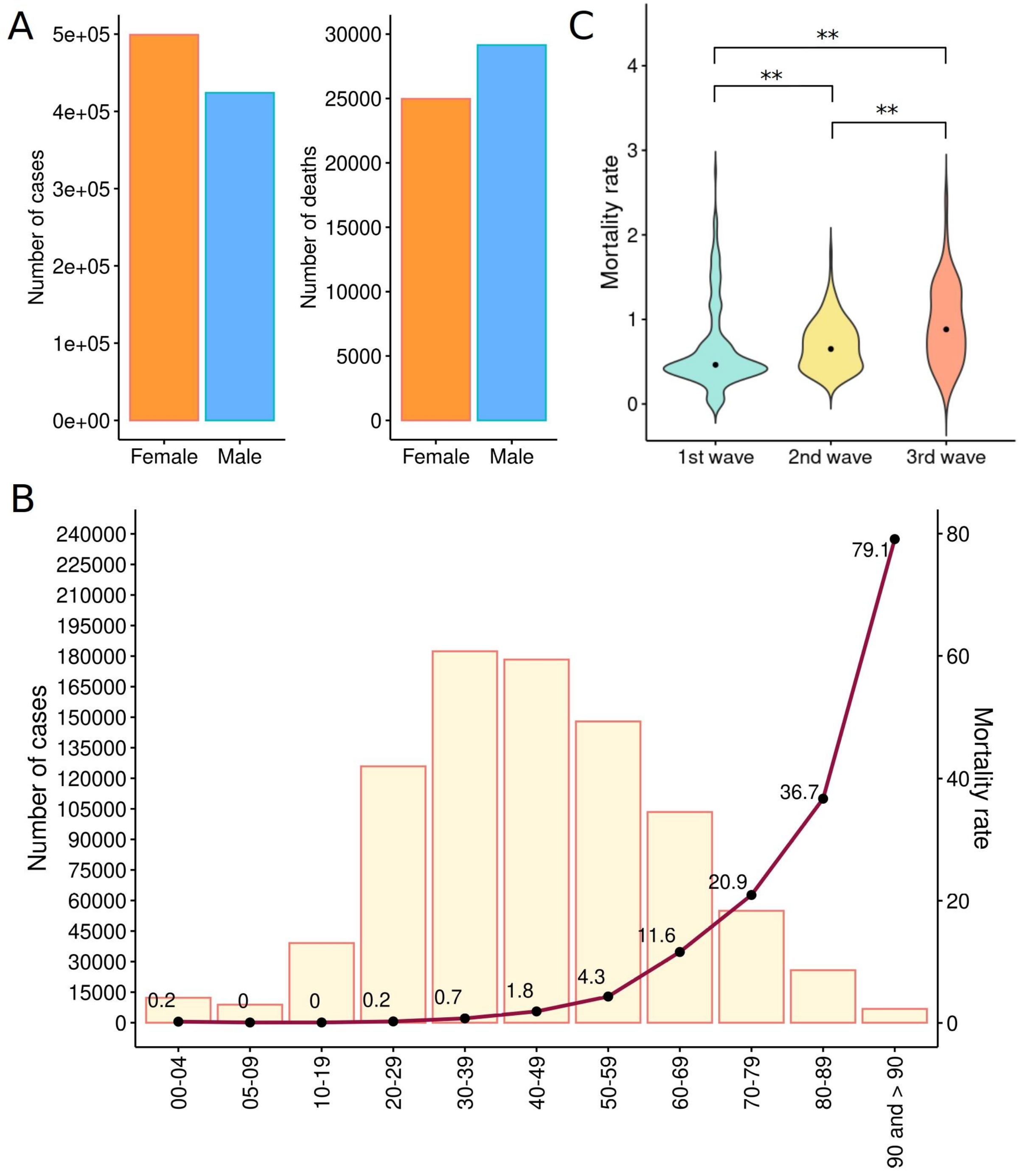


**Figure S2**. **Epidemiological description of the number of cases, deaths and the lethality rate.** A) Number of cases and deaths among females and males. B) Number of cases within age groups. Red line indicates the mortality rate within the age group. C) Differential mortality rate between the three phases using the Wilcoxon pairwise test with Benjamini-Hochberg p-value correction. Asterisk indicated statistical significant differences. The p-values are indicated as $p < 0.05$ (*) and $p < 0.01$ (**).

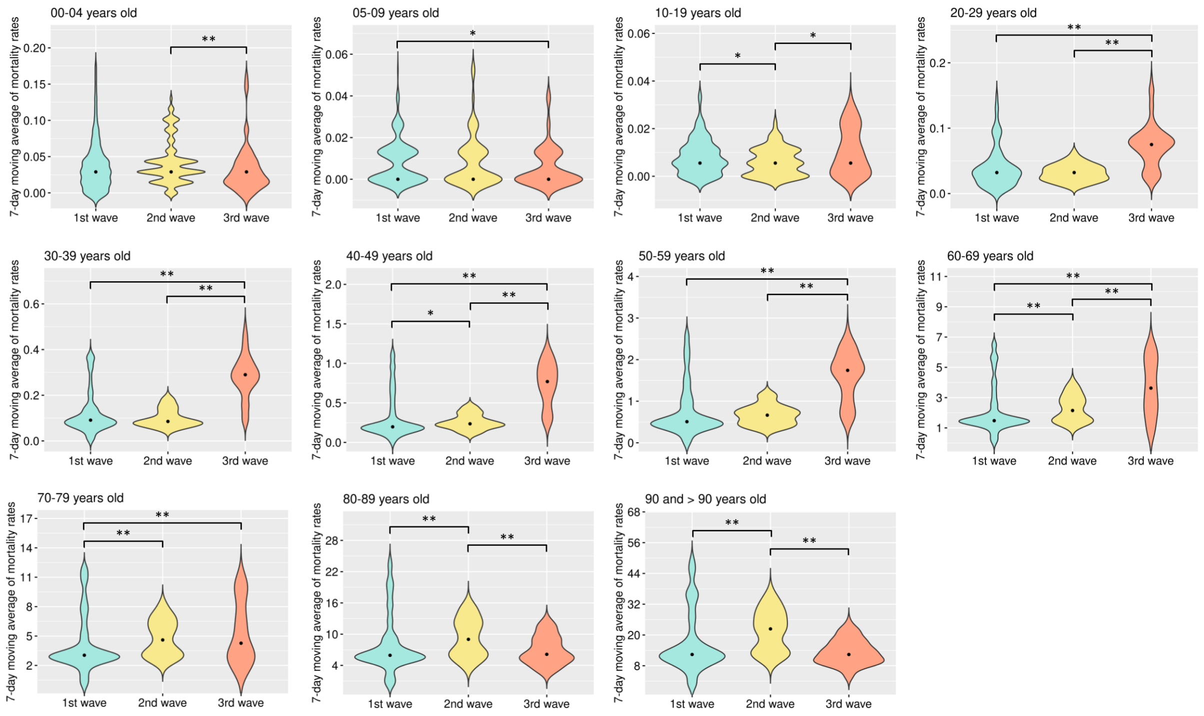


**Figure S3**. **Comparison of mortality rate within age group among the three phases.** Different ages were compared with a 7-day moving average mortality rate. The Wilcoxon pairwise test was used to compare the three phases in each age group, followed by correction by Benjamini-Hochberg. The p-values are indicated as $p < 0.05$ (*) and $p < 0.01$ (**).

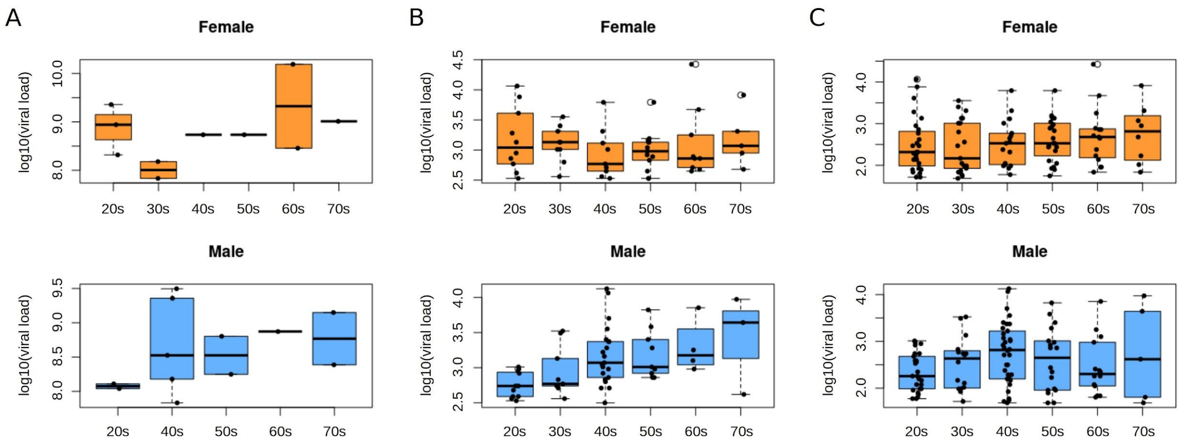


**Figure S4. Comparison between the distribution of the relative quantification of the SARS-CoV-2 viral loads (RQVL) within an age group of 1,119 samples.** Gamma-generalized linear model showed a significant difference in the RQVL among males age groups for both 2% (A), 10% (B) and 20% (C) of the population.

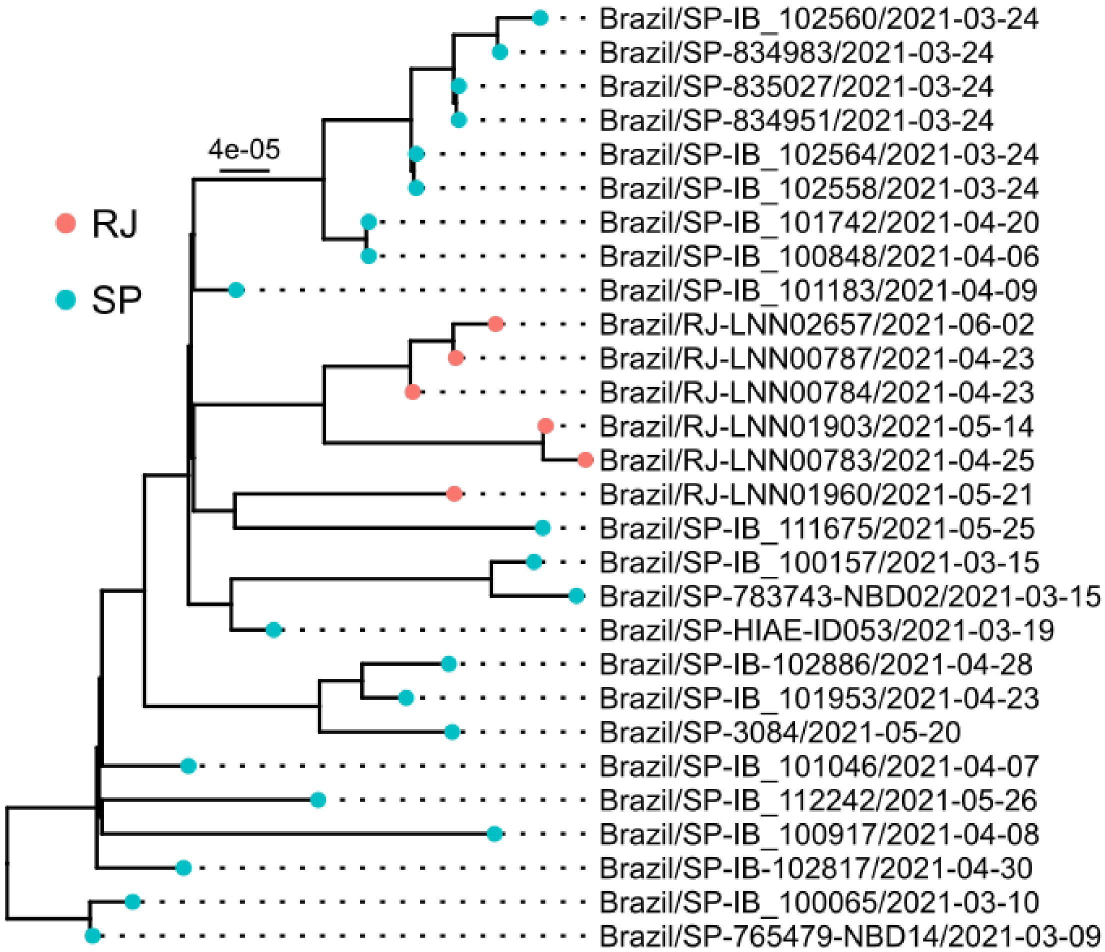


**Figure S5. Maximum likelihood tree of the 28 SARS-CoV-2 genomes classified as P.5.** Colors indicate the Brazilian state of origin of the sample.

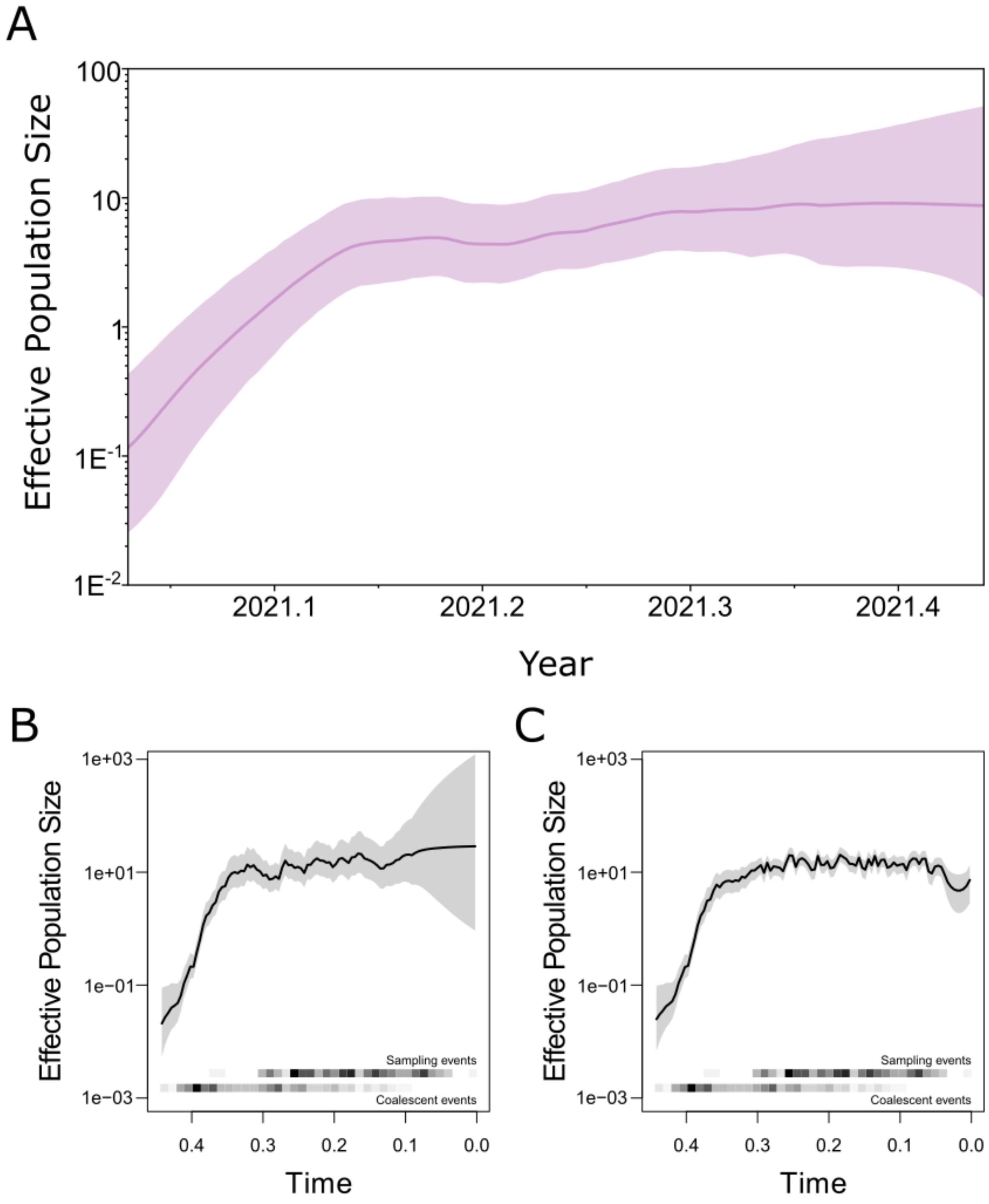


**Figure S6. Effective population size of lineage P.1.2 across time inferred with different models.** A) The GMRF Skyride population model available in BEAST was used. B-C) Bayesian Nonparametric Phylodynamic Reconstruction model was used without (B) and with (C) preferential sampling correction.

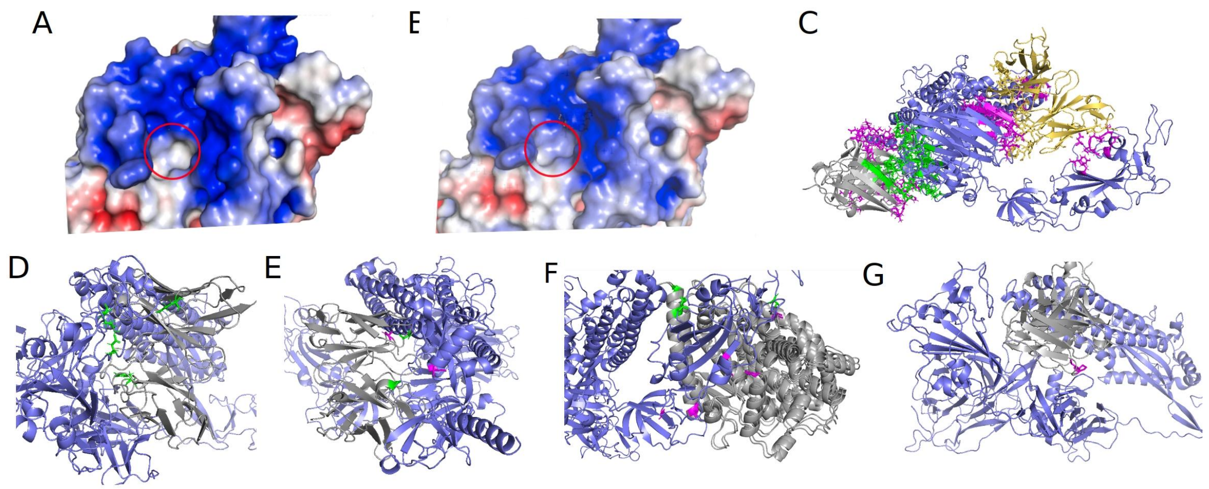


**Figure S7. Comparison of P.1 and P.1.2 protein structures using receptor-ligand docking and affinity binding prediction.** A-B) Electrostatic potential surface of P.1 and sub-clade P.1.2 structures. Red circle indicates the residuo 262 where the mutation (A262S) occurs. C-G) Receptor-ligand complexes for four interactions between Spike and neutralizing antibodies and one interaction between Spike and ACE2. In blue the receptor and the ligands in gray; P.1 exclusive residues are highlighted in pink and those exclusive from P.1.2 are colored in green. In (C), we projected the location of the antibody S1notRBD in P.1 in yellow, maintaining its location in P.1.2 as gray.

**Table S1. Prevalence of COVID-19 cases in the macroregions in the state of Rio de Janeiro**

| Region | Median | IQR |
|---|---|---|
| Central | 9.835030 | 6.050624 |
| North | 8.941276 | 4.812837 |
| Northwest | 7.548458 | 3.940123 |
| South | 7.248784 | 4.403857 |
| Lowland Coastal | 6.404055 | 4.471987 |
| Metropolitan | 5.035158 | 5.192506 |

**Table S2. Comparison of age mortality among the phases.**

| Ages | Phases | Adjusted p-value (BH) |
|---|---|---|
| 00-04 | 2nd and 3rd | 0.003 |
| 05-09 | 1st and 3rd | 0.034 |
| 10-19 | 1st and 2nd | 0.032 |
| 10-19 | 2nd and 3rd | 0.015 |
| 20-29 | 1st and 3rd | <2e-16 |
| 20-29 | 2nd and 3rd | <2e-16 |
| 30-39 | 1st and 3rd | <2e-16 |
| 30-39 | 2nd and 3rd | <2e-16 |
| 40-49 | 1st and 2nd | 0.013 |
| 40-49 | 1st and 3rd | <2e-16 |
| 40-49 | 2nd and 3rd | <2e-16 |
| 50-59 | 1st and 3rd | <2e-16 |
| 50-59 | 2nd and 3rd | <2e-16 |
| 60-69 | 1st and 2nd | 0.00014 |
| 60-69 | 1st and 3rd | 2.2e-12 |
| 60-69 | 2nd and 3rd | 1.2e-08 |
| 70-79 | 1st and 2nd | 1.5e-05 |
| 70-79 | 1st and 3rd | 0.0019 |
| 80-89 | 1st and 2nd | 1.5e-05 |
| 80-89 | 2nd and 3rd | 1.3e-06 |
| 90 and > 90 | 1st and 2nd | 9.2e-09 |
| 90 and > 90 | 2nd and 3rd | 4.6e-15 |

**Table S3. Proportion of Genomes deposited in GISAID and number of cases from the state of Rio de Janeiro.**

| Month | No. Genomes Sequenced | No. Cases Confirmed | Proportion (%) |
|---|---|---|---|
| 2020/03 | 45 | 7369 | 0.6107 |
| 2020/04 | 253 | 50535 | 0.5006 |
| 2020/05 | 54 | 60156 | 0.0898 |
| 2020/06 | 37 | 52331 | 0.0707 |
| 2020/07 | 93 | 53927 | 0.1724 |
| 2020/08 | 93 | 45997 | 0.2022 |
| 2020/09 | 54 | 41028 | 0.1316 |
| 2020/10 | 80 | 41157 | 0.1944 |
| 2020/11 | 71 | 92057 | 0.0771 |
| 2020/12 | 119 | 112478 | 0.1058 |
| 2021/01 | 162 | 70254 | 0.2306 |
| 2021/02 | 187 | 38291 | 0.4883 |
| 2021/03 | 433 | 96427 | 0.4490 |
| 2021/04 | 911 | 77517 | 1.1752 |
| 2021/05 | 913 | 74462 | 1.2261 |
| 2021/06 | 427 | 9248 | 4.6172 |

**Table S5. Energy comparison between P.1 and sub-clade P.1.2 models using FoldX tool**

| Criteria | P.1 | Sub-clade P.1.2 |
|---|---|---|
| BackHbond | -662.35 | -654.28 |
| SideHbond | -198.52 | -185.18 |
| Energy_vdwclash | 65.19 | 76.6 |
| Entropy_mainc | 1763.32 | 1754.78 |
| **Total** | 373.6 | 395.76 |

**Table S6. Number of contacts in common between P.1 and sub-clade P.1.2 in complex with the antibodies and ACE2**

| Ligand | # Common AAs | # AA P.1 | # AA Sub-clade P.1.2 | AA P1 | AA Sub-clade P.1.2 |
|---|---|---|---|---|---|
| S1-S2 | 19 | 1 | 2 | ASP-428.A | GLU-516.A;SER-459.A |
| ACE2 | 23 | 3 | 1 | PRO-561.A;THR-523.A;PRO-330.A;SER-469.A | |
| S1-Not RBD | 0 | 28 | 20 | [...] | SER-262.A;[...] |
| S1-RBD | 25 | 1 | 2 | TRP-353.A | ASP-571.A;THR-1006.A |
| S2 | 25 | 1 | 1 | TYR-707.A | GLY-1124.A |
