## Supplementary material for "Turnover of SARS-CoV-2 lineages shaped the pandemic and enabled the emergence of new variants in the state of Rio de Janeiro, Brazil": Table S7

We gratefully acknowledge the following Authors from the Originating laboratories responsible for obtaining the specimens, as well as the Submitting laboratories where the genome data were generated and shared via GISAID, on which this research is based.

All Submitters of data may be contacted directly via www.gisaid.org

Authors are sorted alphabetically.

| Accession ID | Originating Laboratory | Submitting Laboratory | Authors |
| --- | --- | --- | --- |
| EPI_ISL_1023841 | Centro de Desenvolvimento Tecnológico em Saúde - CDTS | Centro de Desenvolvimento Tecnológico em Saúde - CDTS | Souza,T.M., Fintelman-Rodrigues,N., De Paula,A.D., Saraiva,F.B., Ferreira,M.A. and Sacramento,C.Q. |
| EPI_ISL_1133266, EPI_ISL_1133268 | Laboratório Hermes Pardini | Laboratório de Biologia Integrativa, Instituto de Ciências Biológicas, Universidade Federal de Minas Gerais | Filipe Romero Rebello Moreira, Diego Menezes Bonfim, Danielle Alves Gomes Zauli, Joice do Prado Silva, Aline Brito de Lima, Frederico Scott Varella Malta, Alessandro Clayton de Souza Ferreira, Victor Cavalcanti Pardini, Daniel Costa Queiroz, Rafael Marques de Souza, Victor Emmanuel Viana Geddes, Walyson Coelho Costa, Wagner Carlos Santos Magalhaes, Rennan Garcias Moreira, Carolina Moreira Voloch, Renan Pedra de Souza, Renato Santana Aguiar. |
| EPI_ISL_1181360, EPI_ISL_1181361, EPI_ISL_1181363, EPI_ISL_1181367, EPI_ISL_1181369, EPI_ISL_1181381, EPI_ISL_1181382, EPI_ISL_1181383, EPI_ISL_1181384, EPI_ISL_1181385, EPI_ISL_1181386, EPI_ISL_1181387, EPI_ISL_1181388, EPI_ISL_1181389, EPI_ISL_1181391, EPI_ISL_1181393, EPI_ISL_1181398, EPI_ISL_1181410, EPI_ISL_1181415 | | | |
| see above | Laboratory of Respiratory Viruses and Measles, Oswaldo Cruz Institute, FIOCRUZ | Laboratory of Respiratory Viruses and Measles, Oswaldo Cruz Institute, FIOCRUZ | Paola Resende, Luciana Appolinario, Fernando Motta, Anna Carolina Paixao, Ana Carolina Mendonca, Alice Sampaio Rocha, Renata Serrano Lopes, Marilda Siqueira on behalf of the Fiocruz COVID-19 Genomic Surveillance Network |
| EPI_ISL_1181425 | Laboratorio Central de Saude Publica do Estado do Rio de Janeiro (LACEN-RJ) | Laboratory of Respiratory Viruses and Measles, Oswaldo Cruz Institute, FIOCRUZ | Paola Resende, Luciana Appolinario, Fernando Motta, Anna Carolina Paixao, Ana Carolina Mendonca, Alice Sampaio Rocha, Renata Serrano Lopes, Andrea Cony Cavalcanti, Marilda Siqueira on behalf of the Fiocruz COVID-19 Genomic Surveillance Network |
| EPI_ISL_1181430 | Laboratory of Respiratory Viruses and Measles, Oswaldo Cruz Institute, FIOCRUZ | Laboratory of Respiratory Viruses and Measles, Oswaldo Cruz Institute, FIOCRUZ | Paola Resende, Luciana Appolinario, Fernando Motta, Anna Carolina Paixao, Ana Carolina Mendonca, Alice Sampaio Rocha, Renata Serrano Lopes, Marilda Siqueira on behalf of the Fiocruz COVID-19 Genomic Surveillance Network |
| EPI_ISL_1181431 | Laboratorio Central de Saude Publica do Estado do Rio de Janeiro (LACEN-RJ) | Laboratory of Respiratory Viruses and Measles, Oswaldo Cruz Institute, FIOCRUZ | Paola Resende, Luciana Appolinario, Fernando Motta, Anna Carolina Paixao, Ana Carolina Mendonca, Alice Sampaio Rocha, Renata Serrano Lopes, Andrea Cony Cavalcanti, Marilda Siqueira on behalf of the Fiocruz COVID-19 Genomic Surveillance Network |
| EPI_ISL_1181437, EPI_ISL_1181438, EPI_ISL_1181439, EPI_ISL_1181442, EPI_ISL_1181446, EPI_ISL_1181447, EPI_ISL_1181448, EPI_ISL_1181454, EPI_ISL_1181455, EPI_ISL_1181456, EPI_ISL_1181457, EPI_ISL_1181458, EPI_ISL_1181459, EPI_ISL_1181460, EPI_ISL_1181461, EPI_ISL_1181462, EPI_ISL_1181463, EPI_ISL_1181464, EPI_ISL_1181465, EPI_ISL_1181466, EPI_ISL_1181467, EPI_ISL_1181470, EPI_ISL_1181475, EPI_ISL_1181476, EPI_ISL_1181477, EPI_ISL_1181481, EPI_ISL_1181482, EPI_ISL_1181483, EPI_ISL_1181486, EPI_ISL_1181488, EPI_ISL_1181489, EPI_ISL_1181490, EPI_ISL_1181491, EPI_ISL_1181493, EPI_ISL_1181494, EPI_ISL_1181495, EPI_ISL_1181496, EPI_ISL_1181497, EPI_ISL_1181499, EPI_ISL_1181500, EPI_ISL_1181501, EPI_ISL_1181502, EPI_ISL_1181503, EPI_ISL_1181504, EPI_ISL_1181505, EPI_ISL_1181506, EPI_ISL_1181507, EPI_ISL_1181508, EPI_ISL_1181509, EPI_ISL_1181510, EPI_ISL_1181511, EPI_ISL_1181512, EPI_ISL_1181513, EPI_ISL_1181514, EPI_ISL_1181515, EPI_ISL_1181518, EPI_ISL_1181519, EPI_ISL_1181521, EPI_ISL_1181523, EPI_ISL_1181529, EPI_ISL_1181531, EPI_ISL_1181560, EPI_ISL_1181561, EPI_ISL_1181562, EPI_ISL_1181571, EPI_ISL_1181572, EPI_ISL_1181573, EPI_ISL_1181574, EPI_ISL_1181578, EPI_ISL_1181579, EPI_ISL_1181587, EPI_ISL_1181589, EPI_ISL_1181594, EPI_ISL_1181597, EPI_ISL_1181599 | | | |
| see above | Laboratory of Respiratory Viruses and Measles, Oswaldo Cruz Institute, FIOCRUZ | Laboratory of Respiratory Viruses and Measles, Oswaldo Cruz Institute, FIOCRUZ | Paola Resende, Luciana Appolinario, Fernando Motta, Anna Carolina Paixao, Ana Carolina Mendonca, Alice Sampaio Rocha, Renata Serrano Lopes, Marilda Siqueira on behalf of the Fiocruz COVID-19 Genomic Surveillance Network |
| EPI_ISL_1181600, EPI_ISL_1181601, EPI_ISL_1181602, EPI_ISL_1181603, EPI_ISL_1181606, EPI_ISL_1181607 | Laboratorio Central de Saude Publica do Estado do Rio de Janeiro (LACEN-RJ) | Laboratory of Respiratory Viruses and Measles, Oswaldo Cruz Institute, FIOCRUZ | Paola Resende, Luciana Appolinario, Fernando Motta, Anna Carolina Paixao, Ana Carolina Mendonca, Alice Sampaio Rocha, Renata Serrano Lopes, Andrea Cony Cavalcanti, Marilda Siqueira on behalf of the Fiocruz COVID-19 Genomic Surveillance Network |
| EPI_ISL_1181608 | Laboratory of Respiratory Viruses and Measles, Oswaldo Cruz Institute, FIOCRUZ | Laboratory of Respiratory Viruses and Measles, Oswaldo Cruz Institute, FIOCRUZ | Paola Resende, Luciana Appolinario, Fernando Motta, Anna Carolina Paixao, Ana Carolina Mendonca, Alice Sampaio Rocha, Renata Serrano Lopes, Marilda Siqueira on behalf of the Fiocruz COVID-19 Genomic Surveillance Network |
| EPI_ISL_1181611, EPI_ISL_1181612, EPI_ISL_1181613 | Laboratorio Central de Saude Publica do Estado do Rio de Janeiro (LACEN-RJ) | Laboratory of Respiratory Viruses and Measles, Oswaldo Cruz Institute, FIOCRUZ | Paola Resende, Luciana Appolinario, Fernando Motta, Anna Carolina Paixao, Ana Carolina Mendonca, Alice Sampaio Rocha, Renata Serrano Lopes, Andrea Cony Cavalcanti, Marilda Siqueira on behalf of the Fiocruz COVID-19 Genomic Surveillance Network |
| EPI_ISL_1182552, EPI_ISL_1182553, EPI_ISL_1182554, EPI_ISL_1182556, EPI_ISL_1182557, EPI_ISL_1182558 | Laboratório Central do Estado do Rio de Janeiro | Coordenação Geral de Laboratórios de Saúde Pública (CGLAB/DAEVS/SVS/MS) | Vagner Fonseca, et al. |
| EPI_ISL_1213163, EPI_ISL_1213171, EPI_ISL_1213248, EPI_ISL_1213249, EPI_ISL_1213270, EPI_ISL_1213272, EPI_ISL_1213307, EPI_ISL_1213309, EPI_ISL_1213310, EPI_ISL_1213312, EPI_ISL_1213314, EPI_ISL_1213339, EPI_ISL_1213340, EPI_ISL_1213372, EPI_ISL_1213374, EPI_ISL_1213376, EPI_ISL_1213378, EPI_ISL_1213379, EPI_ISL_1213381, EPI_ISL_1213382, EPI_ISL_1213393, EPI_ISL_1213395, EPI_ISL_1213397, EPI_ISL_1213408, EPI_ISL_1213410, EPI_ISL_1213417, EPI_ISL_1213418, EPI_ISL_1213420, EPI_ISL_1213422, EPI_ISL_1213424, EPI_ISL_1213425 | | | |
| see above | Laboratório HLA/UERJ | Bioinformatics Laboratory / LNCC | Alessandra P Lamarca, Luiz G P de Almeida, Ronaldo da Silva Francisco Jr, Lucymara Fassarella Agnez Lima, Kátia Castanho Scortecci, Vinícius Pietta Perez, Otavio J. Brustolini, Eduardo Sérgio Soares Sousa, Danielle Angst Secco, Angela Maria Guimarães Santos, George Rego Albuquerque, Ana Paula Melo Mariano, Bianca Mendes Maciel, Alexandra L Gerber, Ana Paula de C Guimarães, Paulo Ricardo Nascimento, Francisco Paulo Freire Neto, Sandra Rocha Gadelha, Luís Cristóvão Porto, Eloiza Helena Campana, Selma Maria Bezerra Jeronimo, Ana Tereza R Vasconcelos |
| EPI_ISL_1239111, EPI_ISL_1239112, EPI_ISL_1239113 | Laboratório Central de Saúde Pública Noel Nutels | Coordenação Geral de Laboratórios de Saúde Pública (CGLAB) | Vagner Fonseca et al, |
| EPI_ISL_1289218 | Dutch COVID-19 response team | National Institute for Public Health and the Environment (RIVM) | Adam Meijer, Harry Vennema, Dirk Eggink, Jeroen Cremer, Sharon van den Brink, Bas van der Veer, AnneMarie van den Brandt, Florian Zwagemaker, Dennis Schmitz, Chantal Reusken, on behalf of the national COVID-19 response team |
| EPI_ISL_1324142, EPI_ISL_1324145 | UW Virology Lab | UW Virology Lab | Pavitra Roychoudhury, Hong Xie, Lasata Shrestha, Shah Mohamed Bakhash, Michelle Lin, Margaret Mills, Noah Baker, Sean Ellis, Saraswathi Sathees, Meei-Li Huang, Keith R Jerome, Alexander Greninger |
| EPI_ISL_1327150 | Lighthouse Lab in Cambridge | Wellcome Sanger Institute for the COVID-19 Genomics UK (COG-UK) Consortium | Rob Howes, The Lighthouse Lab in Cambridge and Alex Alderton, Roberto Amato, Jeffrey Barrett, Sonia Goncalves, Ewan Harrison, David K. Jackson, Ian Johnston, Dominic Kwiatkowski, Cordelia Langford, John Sillitoe on behalf of the Wellcome Sanger Institute COVID-19 Surveillance Team |
| EPI_ISL_1402430 | Laboratory of Respiratory Viruses and Measles, Oswaldo Cruz Institute, FIOCRUZ | Laboratory of Respiratory Viruses and Measles, Oswaldo Cruz Institute, FIOCRUZ | Paola Resende, Alex Pauvolid-Correa, Mia Ferreira Araujo, Ana Beatriz Machado Lima, Luciana Appolinario, Fernando Motta, Anna Carolina Paixao, Ana Carolina Mendonca, Alice Sampaio Rocha, Renata Serrano Lopes, Marilda Siqueira on behalf of the Fiocruz COVID-19 Genomic Surveillance Network |
| EPI_ISL_1416322 | PathWest Laboratory Medicine WA | PathWest Laboratory Medicine WA Microbial Surveillance Unit | PathWest Laboratory Medicine WA Microbial Surveillance Unit |
| EPI_ISL_1445142 | CS II EGIDIO BRUNHARA MORRO AGUDO | Instituto Butantan / Mendelics | Dimas Tadeu Covas, Sandra Coccuzzo Sampaio, Maria Carolina Elias, José Salvatore Leister Patané, Vincent Louis Viala, Antonio Jorge Martins, Ricardo Haddad, Claudia Renata dos Santos Barros, Elaine Cristina Marqueze, Raul Machado Neto, Debora Botequio Moretti, Bibiana Santos, João Paulo Kitajima, Erika Freitas, David Schlesinger, Simone Kashima, Evandra Strazza Rodrigues, Svetoslav Nanev Slavov, Elaine Vieira dos Santos, Rafael dos Santos Bezerra, Luiz Carlos Junior de Alcantara, Marta Giovanetti, Vagner Fonseca, Flavia Aburjaile, Rodrigo Tocantins Calado. |
| EPI_ISL_1456633, EPI_ISL_1456637, EPI_ISL_1456638, EPI_ISL_1456646, EPI_ISL_1456649, EPI_ISL_1456650, EPI_ISL_1456651, EPI_ISL_1456652, EPI_ISL_1521307, EPI_ISL_1521308, EPI_ISL_1521309, EPI_ISL_1521310, EPI_ISL_1521312, EPI_ISL_1521313, EPI_ISL_1521314, EPI_ISL_1521315 | | | |
| see above | Dutch COVID-19 response team | National Institute for Public Health and the Environment (RIVM) | Adam Meijer, Harry Vennema, Dirk Eggink, Jeroen Cremer, Sharon van den Brink, Bas van der Veer, AnneMarie van den Brandt, Lisa Wijsman, Kim Freriks, Ryanne Jaarsma, Euníce Then, Jolienke Hardeman, Lynn Aarts, Sanne Bos, Melissa van Tuil, Robert Kohl, Linda van de Nes, Sjoerd Kuiling, James Groot, Florian Zwagemaker, Dennis Schmitz, Annelies Kroneman, Karim Hajji, Chantal Reusken, on behalf of the national COVID-19 response team |
| EPI_ISL_1533977 | Laboratorio de Virologia Molecular / UFRJ | Laboratory of Respiratory Viruses and Measles, Oswaldo Cruz Institute, FIOCRUZ | Paola Resende, Carolina M Voloch, Luciana Appolinario, Fernando Motta, Anna Carolina Paixao, Ana Carolina Mendonca, Alice Sampaio Rocha, Renata Serrano Lopes, Amilcar Tanuri, Marilda Siqueira on behalf of the Fiocruz COVID-19 Genomic Surveillance Network |
| EPI_ISL_1533980, EPI_ISL_1533983 | Laboratory of Respiratory Viruses and Measles, Oswaldo Cruz Institute, FIOCRUZ | Laboratory of Respiratory Viruses and Measles, Oswaldo Cruz Institute, FIOCRUZ | Paola Resende, Luciana Appolinario, Fernando Motta, Anna Carolina Paixao, Ana Carolina Mendonca, Alice Sampaio Rocha, Renata Serrano Lopes, Marilda Siqueira on behalf of the Fiocruz COVID-19 Genomic Surveillance Network |

| | | | |
|---|---|---|---|
| EPI_ISL_1533991, EPI_ISL_1533995, EPI_ISL_1533998 | Laboratorio de Virologia Molecular / UFRJ | Laboratory of Respiratory Viruses and Measles, Oswaldo Cruz Institute, FIOCRUZ | Paola Resende, Carolina M Voloch, Luciana Appolinario, Fernando Motta, Anna Carolina Paixao, Ana Carolina Mendonca, Alice Sampaio Rocha, Renata Serrano Lopes, Amilcar Tanuri, Marilda Siqueira on behalf of the Fiocruz COVID-19 Genomic Surveillance Network |
| EPI_ISL_1534005, EPI_ISL_1534011 | Laboratory of Respiratory Viruses and Measles, Oswaldo Cruz Institute, FIOCRUZ | Laboratory of Respiratory Viruses and Measles, Oswaldo Cruz Institute, FIOCRUZ | Paola Resende, Luciana Appolinario, Fernando Motta, Anna Carolina Paixao, Ana Carolina Mendonca, Alice Sampaio Rocha, Renata Serrano Lopes, Marilda Siqueira on behalf of the Fiocruz COVID-19 Genomic Surveillance Network |
| EPI_ISL_1534012 | Laboratorio Central de Saude Publica do Estado do Rio de Janeiro (LACEN-RJ) | Laboratory of Respiratory Viruses and Measles, Oswaldo Cruz Institute, FIOCRUZ | Paola Resende, Luciana Appolinario, Fernando Motta, Anna Carolina Paixao, Ana Carolina Mendonca, Alice Sampaio Rocha, Renata Serrano Lopes, Andrea Cony Cavalcanti, Marilda Siqueira on behalf of the Fiocruz COVID-19 Genomic Surveillance Network |
| EPI_ISL_1534014, EPI_ISL_1534015, EPI_ISL_1534016 | Laboratory of Respiratory Viruses and Measles, Oswaldo Cruz Institute, FIOCRUZ | Laboratory of Respiratory Viruses and Measles, Oswaldo Cruz Institute, FIOCRUZ | Paola Resende, Luciana Appolinario, Fernando Motta, Anna Carolina Paixao, Ana Carolina Mendonca, Alice Sampaio Rocha, Renata Serrano Lopes, Marilda Siqueira on behalf of the Fiocruz COVID-19 Genomic Surveillance Network |
| EPI_ISL_1583681 | Central Public Health Laboratory - LACEN -Bahia, Salvador, Brazil | Central Public Health Laboratory - LACEN -Bahia, Salvador, Brazil | Stephane Tosta, Luciana Oliveira, Vanessa Nardy,Patrícia Cajado,Marcela Gómez, Breno Dominguez, Jaqueline Gomes, Vagner Fonseca,Marta Giovanetti,Luiz Alcantara, Felicidade Pereira, Arabela Leal |
| EPI_ISL_1592230 | Laboratório Hermes Pardini | Laboratório de Biologia Integrativa, Instituto de Ciências Biológicas, Universidade Federal de Minas Gerais | Filipe Romero Rebello Moreira, Diego Menezes Bonfim, Danielle Alves Gomes Zauli, Joice do Prado Silva, Aline Brito de Lima, Frederico Scott Varella Malta, Alessandro Clayton de Souza Ferreira, Victor Cavalcanti Pardini, Daniel Costa Queiroz, Rafael Marques de Souza, Victor Emmanuel Viana Geddes, Walyson Coelho Costa, Wagner Carlos Santos Magalhaes, Rennan Garcias Moreira, Carolina Moreira Voloch, Renan Pedra de Souza, Renato Santana Aguiar. |
| EPI_ISL_1596573 | Dutch COVID-19 response team | National Institute for Public Health and the Environment (RIVM) | Adam Meijer, Harry Vennema, Dirk Eggink, Jeroen Cremer, Sharon van den Brink, Bas van der Veer, AnneMarie van den Brandt, Lisa Wijsman, Kim Freriks, Ryanne Jaarsma, Euníce Then, Jolienke Hardeman, Lynn Aarts, Sanne Bos, Melissa van Tuil, Robert Kohl, Linda van de Nes, Sjoerd Kuiling, James Groot, Florian Zwagemaker, Dennis Schmitz, Annelies Kroneman, Karim Hajji, Chantal Reusken, on behalf of the national COVID-19 response team |
| EPI_ISL_1649154 | Aegis Sciences Corporation | Centers for Disease Control and Prevention Division of Viral Diseases, Pathogen Discovery | Dakota Howard, Dhwani Batra, Peter W. Cook, Kara Moser, Adrian Paskey, Jason Caravas, Benjamin Rambo-Martin, Shatavia Morrison, Christopher Gulvick, Scott Sammons, Yvette Unoarumhi, Darlene Wagner, Matthew Schmerer, Cyndi Clark, Patrick Campbell, Rob Case, Vikramsinha Ghorpade, Holly Houdeshell, Ola Kvalvaag, Dillon Nall, Ethan Sanders, Alec Vest, Shaun Westlund, Matthew Hardison, Clinton R. Paden, Duncan MacCannell |
| EPI_ISL_1664113, EPI_ISL_1664114, EPI_ISL_1664115, EPI_ISL_1664116, EPI_ISL_1664117, EPI_ISL_1664118, EPI_ISL_1664119, EPI_ISL_1664120, EPI_ISL_1664121, EPI_ISL_1664122, EPI_ISL_1664123, EPI_ISL_1664124, EPI_ISL_1664125, EPI_ISL_1664126, EPI_ISL_1664127, EPI_ISL_1664128, EPI_ISL_1664129, EPI_ISL_1664130, EPI_ISL_1664131, EPI_ISL_1664132, EPI_ISL_1664133, EPI_ISL_1664134, EPI_ISL_1664135, EPI_ISL_1664136, EPI_ISL_1664137, EPI_ISL_1664138, EPI_ISL_1664139, EPI_ISL_1664140, EPI_ISL_1664141, EPI_ISL_1664142, EPI_ISL_1664143, EPI_ISL_1664144, EPI_ISL_1664145, EPI_ISL_1664146, EPI_ISL_1664147, EPI_ISL_1664148, EPI_ISL_1664149, EPI_ISL_1664150, EPI_ISL_1664151, EPI_ISL_1664152, EPI_ISL_1664153, EPI_ISL_1664154, EPI_ISL_1664155, EPI_ISL_1664156, EPI_ISL_1664157, EPI_ISL_1664158, EPI_ISL_1664159, EPI_ISL_1664160, EPI_ISL_1664161, EPI_ISL_1664162, EPI_ISL_1664163, EPI_ISL_1664164, EPI_ISL_1664165, EPI_ISL_1664166, EPI_ISL_1664167, EPI_ISL_1664168, EPI_ISL_1664169, EPI_ISL_1664170, EPI_ISL_1664171, EPI_ISL_1664172, EPI_ISL_1664173, EPI_ISL_1664174, EPI_ISL_1664175, EPI_ISL_1664176, EPI_ISL_1664177, EPI_ISL_1664178, EPI_ISL_1664179, EPI_ISL_1664180, EPI_ISL_1664181, EPI_ISL_1664182, EPI_ISL_1664183, EPI_ISL_1664184, EPI_ISL_1664185, EPI_ISL_1664186, EPI_ISL_1664187, EPI_ISL_1664188, EPI_ISL_1664189, EPI_ISL_1664190, EPI_ISL_1664191, EPI_ISL_1664192, EPI_ISL_1664193, EPI_ISL_1664194, EPI_ISL_1664195, EPI_ISL_1664196, EPI_ISL_1664197, EPI_ISL_1664198, EPI_ISL_1664199, EPI_ISL_1664200, EPI_ISL_1664201, EPI_ISL_1664202 | | | |
| see above | Laboratorio Central Noel Nutels | Bioinformatics Laboratory / LNCC | Luiz G P de Almeida, Alessandra P Lamarca, Ronaldo da Silva F Jr, Liliane Cavalcante, Alexandra L Gerber, Ana Paula de C Guimarães, Douglas Terra Machado, Cassia Alves, Diana Mariani, Thais Felix Cruz, Mario Sergio Ribeiro, Silvia Carvalho, Flávio Dias da Silva, Marcio Henrique de Oliveira Garcia, Leandro Magalhães de Souza, Cristiane Gomes da Silva, Caio Luiz Pereira Ribeiro, Andréa Cony Cavalcanti, Claudia Maria Braga de Mello, Amilcar Tanuri, Ana Tereza R Vasconcelos |
| EPI_ISL_1678584, EPI_ISL_1690551, EPI_ISL_1694627, EPI_ISL_1694628, EPI_ISL_1694631, EPI_ISL_1694632, EPI_ISL_1694634, EPI_ISL_1695909 | Instituto Estadual do Cérebro Paulo Niemayer (IECPN) | Laboratório de Imunofarmacologia | Thiago Moreno Lopes Souza |
| EPI_ISL_1738831 | unknown | Instituto Nacional de Saude (INSA) | Borges et al |
| EPI_ISL_1752639 | UPA Dr Luis Atilio Losi Viana Ribeirao Preto | Instituto Adolfo Lutz, Interdiciplinary Procedures Center, Strategic Laboratory | Claudio Tavares Sacchi, Claudia Regina Gonçalves, Erica Valessa Ramos Gomes, Karoline Rodrigues Campos, Caio Vinicius Dias Lopes, Leonardo Jose Tadeu de Araujo, Katia Correa de Oliveira Santos |
| EPI_ISL_1795103 | HOSPITAL DOS FORNCEDORES | Instituto Butantan / ESALQ-Piracicaba | Instituto Butantan: Alexander Roberto Precioso, Dimas Tadeu Covas, Sandra Coccuzzo Sampaio, Maria Carolina Elias, José Salvatore Leister Patané, Vincent Louis Viala, Antonio Jorge Martins, Ricardo Haddad, Claudia Renata dos Santos Barros, Elaine Cristina Marqueze, Raul Machado Neto, Debora Botequio Moretti. Centro de Genômica Funcional da ESALQ: Luiz Lehmann Coutinho, Ricardo Augusto Brassaloti, Raquel de Lello Rocha Campos Cassano. NGS Soluções Genômicas: Pilar Drummond Sampaio Corrêa Mariani. FZEA-USP Pirassununga: Mirele Daiana Poleti, Jessika Cristina Chagas Lesbon, Elisangela Chicaroni Mattos, Heidge Fukumasu. USP-Botucatu: Rejane Maria Tommasini Grotto, Jayme A. Souza-Neto, Guilherme Targino Valente, Patricia Akemi Assato, Felipe Allan da Silva da Costa, Bianca Cechetto Carlos. Mendelics: Bibiana Santos, João Paulo Kitajima, Erika Freitas, David Schlesinger. Hemocentro Ribeirão Preto: Simone Kashima, Evandra Strazza Rodrigues, Svetoslav Nanev Slavov, Elaine Vieira dos Santos, Rafael dos Santos Bezerra, Luiz Carlos Junior de Alcantara, Marta Giovanetti, Vagner Fonseca, Flavia Aburjaile, Rodrigo Tocantins Calado. |
| EPI_ISL_1795104, EPI_ISL_1795105 | LABORATORIO MUNICIPAL DE PIRACICABA | Instituto Butantan / ESALQ-Piracicaba | Instituto Butantan: Alexander Roberto Precioso, Dimas Tadeu Covas, Sandra Coccuzzo Sampaio, Maria Carolina Elias, José Salvatore Leister Patané, Vincent Louis Viala, Antonio Jorge Martins, Ricardo Haddad, Claudia Renata dos Santos Barros, Elaine Cristina Marqueze, Raul Machado Neto, Debora Botequio Moretti. Centro de Genômica Funcional da ESALQ: Luiz Lehmann Coutinho, Ricardo Augusto Brassaloti, Raquel de Lello Rocha Campos Cassano. NGS Soluções Genômicas: Pilar Drummond Sampaio Corrêa Mariani. FZEA-USP Pirassununga: Mirele Daiana Poleti, Jessika Cristina Chagas Lesbon, Elisangela Chicaroni Mattos, Heidge Fukumasu. USP-Botucatu: Rejane Maria Tommasini Grotto, Jayme A. Souza-Neto, Guilherme Targino Valente, Patricia Akemi Assato, Felipe Allan da Silva da Costa, Bianca Cechetto Carlos. Mendelics: Bibiana Santos, João Paulo Kitajima, Erika Freitas, David Schlesinger. Hemocentro Ribeirão Preto: Simone Kashima, Evandra Strazza Rodrigues, Svetoslav Nanev Slavov, Elaine Vieira dos Santos, Rafael dos Santos Bezerra, Luiz Carlos Junior de Alcantara, Marta Giovanetti, Vagner Fonseca, Flavia Aburjaile, Rodrigo Tocantins Calado. |
| EPI_ISL_1795108 | SECRETARIA DE SAUDE DE SAO PEDRO | Instituto Butantan / ESALQ-Piracicaba | Instituto Butantan: Alexander Roberto Precioso, Dimas Tadeu Covas, Sandra Coccuzzo Sampaio, Maria Carolina Elias, José Salvatore Leister Patané, Vincent Louis Viala, Antonio Jorge Martins, Ricardo Haddad, Claudia Renata dos Santos Barros, Elaine Cristina Marqueze, Raul Machado Neto, Debora Botequio Moretti. Centro de Genômica Funcional da ESALQ: Luiz Lehmann Coutinho, Ricardo Augusto Brassaloti, Raquel de Lello Rocha Campos Cassano. NGS Soluções Genômicas: Pilar Drummond Sampaio Corrêa Mariani. FZEA-USP Pirassununga: Mirele Daiana Poleti, Jessika Cristina Chagas Lesbon, Elisangela Chicaroni Mattos, Heidge Fukumasu. USP-Botucatu: Rejane Maria Tommasini Grotto, Jayme A. Souza-Neto, Guilherme Targino Valente, Patricia Akemi Assato, Felipe Allan da Silva da Costa, Bianca Cechetto Carlos. Mendelics: Bibiana Santos, João Paulo Kitajima, Erika Freitas, David Schlesinger. Hemocentro Ribeirão Preto: Simone Kashima, Evandra Strazza Rodrigues, Svetoslav Nanev Slavov, Elaine Vieira dos Santos, Rafael dos Santos Bezerra, Luiz Carlos Junior de Alcantara, Marta Giovanetti, Vagner Fonseca, Flavia Aburjaile, Rodrigo Tocantins Calado. |
| EPI_ISL_1795223 | HOSPITAL REGIONAL DE ITAPETININGA | Instituto Butantan / ESALQ-Piracicaba | Instituto Butantan: Alexander Roberto Precioso, Dimas Tadeu Covas, Sandra Coccuzzo Sampaio, Maria Carolina Elias, José Salvatore Leister Patané, Vincent Louis Viala, Antonio Jorge Martins, Ricardo Haddad, Claudia Renata dos Santos Barros, Elaine Cristina Marqueze, Raul Machado Neto, Debora Botequio Moretti. Centro de Genômica Funcional da ESALQ: Luiz Lehmann Coutinho, Ricardo Augusto Brassaloti, Raquel de Lello Rocha Campos Cassano. NGS Soluções Genômicas: Pilar Drummond Sampaio Corrêa Mariani. FZEA-USP Pirassununga: Mirele Daiana Poleti, Jessika Cristina Chagas Lesbon, Elisangela Chicaroni Mattos, Heidge Fukumasu. USP-Botucatu: Rejane Maria Tommasini Grotto, Jayme A. Souza-Neto, Guilherme Targino Valente, Patricia Akemi Assato, Felipe Allan da Silva da Costa, Bianca Cechetto Carlos. Mendelics: Bibiana Santos, João Paulo Kitajima, Erika Freitas, David Schlesinger. Hemocentro Ribeirão Preto: Simone Kashima, Evandra Strazza Rodrigues, Svetoslav Nanev Slavov, Elaine Vieira dos Santos, Rafael dos Santos Bezerra, Luiz Carlos Junior de Alcantara, Marta Giovanetti, Vagner Fonseca, Flavia Aburjaile, Rodrigo Tocantins Calado. |
| EPI_ISL_1795329 | CENTRO DE ESPECIALIDADES DE PRIMAVERA | Instituto Butantan / ESALQ-Piracicaba | Instituto Butantan: Alexander Roberto Precioso, Dimas Tadeu Covas, Sandra Coccuzzo Sampaio, Maria Carolina Elias, José Salvatore Leister Patané, Vincent Louis Viala, Antonio Jorge Martins, Ricardo Haddad, Claudia Renata dos Santos Barros, Elaine Cristina Marqueze, Raul Machado Neto, Debora Botequio Moretti. Centro de Genômica Funcional da ESALQ: Luiz Lehmann Coutinho, Ricardo Augusto Brassaloti, Raquel de Lello Rocha Campos Cassano. NGS Soluções Genômicas: Pilar Drummond Sampaio Corrêa Mariani. FZEA-USP Pirassununga: Mirele Daiana Poleti, Jessika Cristina Chagas Lesbon, Elisangela Chicaroni Mattos, Heidge Fukumasu. USP-Botucatu: Rejane Maria Tommasini Grotto, Jayme A. Souza-Neto, Guilherme Targino Valente, Patricia Akemi Assato, Felipe Allan da Silva da Costa, Bianca Cechetto Carlos. Mendelics: Bibiana Santos, João Paulo Kitajima, Erika Freitas, David Schlesinger. Hemocentro Ribeirão Preto: Simone Kashima, Evandra Strazza Rodrigues, Svetoslav Nanev Slavov, Elaine Vieira dos Santos, Rafael dos Santos Bezerra, Luiz Carlos Junior de Alcantara, Marta Giovanetti, Vagner Fonseca, Flavia Aburjaile, Rodrigo Tocantins Calado. |

| | | | |
|---|---|---|---|
| EPI_ISL_1858267, EPI_ISL_1858269, EPI_ISL_1858271, EPI_ISL_1858273, EPI_ISL_1858275, EPI_ISL_1858277, EPI_ISL_1858279, EPI_ISL_1858281, EPI_ISL_1858282, EPI_ISL_1858284, EPI_ISL_1858286, EPI_ISL_1858288, EPI_ISL_1858290, EPI_ISL_1858292, EPI_ISL_1858294, EPI_ISL_1858295, EPI_ISL_1858297, EPI_ISL_1858299, EPI_ISL_1858301, EPI_ISL_1858303, EPI_ISL_1858304, EPI_ISL_1858306, EPI_ISL_1858308, EPI_ISL_1858310, EPI_ISL_1858312, EPI_ISL_1858314, EPI_ISL_1858315, EPI_ISL_1858317, EPI_ISL_1858319, EPI_ISL_1858321, EPI_ISL_1858323, EPI_ISL_1858325, EPI_ISL_1858327, EPI_ISL_1858329, EPI_ISL_1858331, EPI_ISL_1858333, EPI_ISL_1858334, EPI_ISL_1858336, EPI_ISL_1858338, EPI_ISL_1858340, EPI_ISL_1858342, EPI_ISL_1858344, EPI_ISL_1858346, EPI_ISL_1858348, EPI_ISL_1858350, EPI_ISL_1858352, EPI_ISL_1858354, EPI_ISL_1858356, EPI_ISL_1858357, EPI_ISL_1858359, EPI_ISL_1858361, EPI_ISL_1858363, EPI_ISL_1858365, EPI_ISL_1858367, EPI_ISL_1858369, EPI_ISL_1858371, EPI_ISL_1858373, EPI_ISL_1858375, EPI_ISL_1858377, EPI_ISL_1858379, EPI_ISL_1858380, EPI_ISL_1858383, EPI_ISL_1858384, EPI_ISL_1858387, EPI_ISL_1858389, EPI_ISL_1858391, EPI_ISL_1858392, EPI_ISL_1858395, EPI_ISL_1858397, EPI_ISL_1858399, EPI_ISL_1858401, EPI_ISL_1858403, EPI_ISL_1858409, EPI_ISL_1858411, EPI_ISL_1858414, EPI_ISL_1858416, EPI_ISL_1858418, EPI_ISL_1858420, EPI_ISL_1858422, EPI_ISL_1858423, EPI_ISL_1858426, EPI_ISL_1858428, EPI_ISL_1858430, EPI_ISL_1858432, EPI_ISL_1858434, EPI_ISL_1858436, EPI_ISL_1858438, EPI_ISL_1858440, EPI_ISL_1858442, EPI_ISL_1858444, EPI_ISL_1858446, EPI_ISL_1858449, EPI_ISL_1858451, EPI_ISL_1858453, EPI_ISL_1858454, EPI_ISL_1858456, EPI_ISL_1858458, EPI_ISL_1858460, EPI_ISL_1858462, EPI_ISL_1858464, EPI_ISL_1858466, EPI_ISL_1858468, EPI_ISL_1858470, EPI_ISL_1858473, EPI_ISL_1858475, EPI_ISL_1858477, EPI_ISL_1858479, EPI_ISL_1858481, EPI_ISL_1858483, EPI_ISL_1858486, EPI_ISL_1858488, EPI_ISL_1858490, EPI_ISL_1858492, EPI_ISL_1858494, EPI_ISL_1858496, EPI_ISL_1858498, EPI_ISL_1858500, EPI_ISL_1858502, EPI_ISL_1858504, EPI_ISL_1858506, EPI_ISL_1858508, EPI_ISL_1858512, EPI_ISL_1858514, EPI_ISL_1858516, EPI_ISL_1858518, EPI_ISL_1858520, EPI_ISL_1858522, EPI_ISL_1858526, EPI_ISL_1858528, EPI_ISL_1858530, EPI_ISL_1858532, EPI_ISL_1858536, EPI_ISL_1858538, EPI_ISL_1858540, EPI_ISL_1858542, EPI_ISL_1858544, EPI_ISL_1858546, EPI_ISL_1858548, EPI_ISL_1858549, EPI_ISL_1858551, EPI_ISL_1858554, EPI_ISL_1858555, EPI_ISL_1858557, EPI_ISL_1858559, EPI_ISL_1858561, EPI_ISL_1858563, EPI_ISL_1858565, EPI_ISL_1858567, EPI_ISL_1858571, EPI_ISL_1858572, EPI_ISL_1858575, EPI_ISL_1858577, EPI_ISL_1858578, EPI_ISL_1858580, EPI_ISL_1858582, EPI_ISL_1858584, EPI_ISL_1858586, EPI_ISL_1858588, EPI_ISL_1858590, EPI_ISL_1858592, EPI_ISL_1858594, EPI_ISL_1858596, EPI_ISL_1858598, EPI_ISL_1858599, EPI_ISL_1858603, EPI_ISL_1858606, EPI_ISL_1858607, EPI_ISL_1858610, EPI_ISL_1858611, EPI_ISL_1858613, EPI_ISL_1858615, EPI_ISL_1858617, EPI_ISL_1858619, EPI_ISL_1858621, EPI_ISL_1858623, EPI_ISL_1858625, EPI_ISL_1858627, EPI_ISL_1858629, EPI_ISL_1858631, EPI_ISL_1858633, EPI_ISL_1858636, EPI_ISL_1858637, EPI_ISL_1858639, EPI_ISL_1858641, EPI_ISL_1858643, EPI_ISL_1858645, EPI_ISL_1858648, EPI_ISL_1858650, EPI_ISL_1858651, EPI_ISL_1858655, EPI_ISL_1858656, EPI_ISL_1858658, EPI_ISL_1858660, EPI_ISL_1858662, EPI_ISL_1858664, EPI_ISL_1858666, EPI_ISL_1858668, EPI_ISL_1858670, EPI_ISL_1858672, EPI_ISL_1858674, EPI_ISL_1858675, EPI_ISL_1858677, EPI_ISL_1858680, EPI_ISL_1858682, EPI_ISL_1858684, EPI_ISL_1858686, EPI_ISL_1858688, EPI_ISL_1858691, EPI_ISL_1858693, EPI_ISL_1858694, EPI_ISL_1858697, EPI_ISL_1858701, EPI_ISL_1858703, EPI_ISL_1858705, EPI_ISL_1858707, EPI_ISL_1858709, EPI_ISL_1858711, EPI_ISL_1858713, EPI_ISL_1858715, EPI_ISL_1858717, EPI_ISL_1858719, EPI_ISL_1858721, EPI_ISL_1858722, EPI_ISL_1858724, EPI_ISL_1858726, EPI_ISL_1858728, EPI_ISL_1858730, EPI_ISL_1858732, EPI_ISL_1858734, EPI_ISL_1858736, EPI_ISL_1858738, EPI_ISL_1858740, EPI_ISL_1858741, EPI_ISL_1858743, EPI_ISL_1858745, EPI_ISL_1858747, EPI_ISL_1858751, EPI_ISL_1858752, EPI_ISL_1858754, EPI_ISL_1858756 | | | |
| see above | Laboratorio Central Noel Nutels | Bioinformatics Laboratory / LNCC | Luiz G P de Almeida, Alessandra P Lamarca, Ronaldo da Silva F Jr, Liliane Cavalcante, Alexandra L Gerber, Ana Paula de C Guimarães, Douglas Terra Machado, Cassia Alves, Diana Mariani, Thais Felix Cruz, Mario Sergio Ribeiro, Silvia Carvalho, Flávio Dias da Silva, Marcio Henrique de Oliveira Garcia, Leandro Magalhães de Souza, Cristiane Gomes da Silva, Caio Luiz Pereira Ribeiro, Andréa Cony Cavalcanti, Claudia Maria Braga de Mello, Amilcar Tanuri, Ana Tereza R Vasconcelos |
| EPI_ISL_1858758, EPI_ISL_1858761, EPI_ISL_1858763, EPI_ISL_1858765, EPI_ISL_1858767, EPI_ISL_1858771, EPI_ISL_1858775, EPI_ISL_1858777, EPI_ISL_1858779, EPI_ISL_1858781, EPI_ISL_1858783, EPI_ISL_1858786, EPI_ISL_1858788, EPI_ISL_1858790, EPI_ISL_1858792, EPI_ISL_1858794, EPI_ISL_1858796, EPI_ISL_1858798, EPI_ISL_1858800, EPI_ISL_1858802, EPI_ISL_1858803, EPI_ISL_1858805, EPI_ISL_1858807, EPI_ISL_1858809, EPI_ISL_1858811, EPI_ISL_1858814, EPI_ISL_1858816, EPI_ISL_1858818, EPI_ISL_1858819, EPI_ISL_1858822, EPI_ISL_1858824, EPI_ISL_1858826, EPI_ISL_1858828, EPI_ISL_1858830, EPI_ISL_1858832, EPI_ISL_1858834, EPI_ISL_1858836, EPI_ISL_1858838, EPI_ISL_1858840, EPI_ISL_1858842, EPI_ISL_1858843, EPI_ISL_1858846, EPI_ISL_1858847, EPI_ISL_1858850, EPI_ISL_1858852, EPI_ISL_1858853, EPI_ISL_1858856, EPI_ISL_1858858, EPI_ISL_1858860, EPI_ISL_1858862, EPI_ISL_1858864, EPI_ISL_1858866, EPI_ISL_1858868, EPI_ISL_1858870, EPI_ISL_1858872, EPI_ISL_1858878, EPI_ISL_1858880, EPI_ISL_1858882, EPI_ISL_1858884, EPI_ISL_1858886, EPI_ISL_1858888, EPI_ISL_1858890, EPI_ISL_1858892, EPI_ISL_1858894, EPI_ISL_1858896, EPI_ISL_1858898, EPI_ISL_1858900, EPI_ISL_1858902, EPI_ISL_1858904, EPI_ISL_1858906, EPI_ISL_1858908, EPI_ISL_1858910, EPI_ISL_1858912, EPI_ISL_1858914, EPI_ISL_1858916, EPI_ISL_1858918, EPI_ISL_1858920, EPI_ISL_1858922, EPI_ISL_1858924, EPI_ISL_1858926, EPI_ISL_1858928, EPI_ISL_1858930, EPI_ISL_1858933, EPI_ISL_1858934, EPI_ISL_1858936, EPI_ISL_1858938, EPI_ISL_1858940, EPI_ISL_1858942, EPI_ISL_1858944, EPI_ISL_1858946, EPI_ISL_1858948, EPI_ISL_1858950, EPI_ISL_1858952, EPI_ISL_1858954, EPI_ISL_1858957, EPI_ISL_1858960, EPI_ISL_1858963, EPI_ISL_1858964, EPI_ISL_1858966, EPI_ISL_1858968, EPI_ISL_1858969, EPI_ISL_1858970, EPI_ISL_1858971, EPI_ISL_1858973, EPI_ISL_1858975, EPI_ISL_1858977, EPI_ISL_1858979, EPI_ISL_1858981, EPI_ISL_1858984, EPI_ISL_1858986, EPI_ISL_1858988, EPI_ISL_1858990, EPI_ISL_1858992, EPI_ISL_1858994, EPI_ISL_1858996, EPI_ISL_1858998, EPI_ISL_1859000, EPI_ISL_1859002, EPI_ISL_1859005, EPI_ISL_1859007, EPI_ISL_1859008 | | | |
| see above | Unidade de apoio ao diagnóstico da COVID - UNADIG | Bioinformatics Laboratory / LNCC | Luiz G P de Almeida, Alessandra P Lamarca, Ronaldo da Silva F Jr, Liliane Cavalcante, Alexandra L Gerber, Ana Paula de C Guimarães, Douglas Terra Machado, Cassia Alves, Diana Mariani, Thais Felix Cruz, Mario Sergio Ribeiro, Silvia Carvalho, Flávio Dias da Silva, Marcio Henrique de Oliveira Garcia, Leandro Magalhães de Souza, Cristiane Gomes da Silva, Caio Luiz Pereira Ribeiro, Andréa Cony Cavalcanti, Claudia Maria Braga de Mello, Amilcar Tanuri, Ana Tereza R Vasconcelos |
| EPI_ISL_1966113 | CS II EGIDIO BRUNHARA MORRO AGUDO | Instituto Butantan / Mendelics | Instituto Butantan: Dimas Tadeu Covas, Sandra Coccuzzo Sampaio, Maria Carolina Elias, José Salvatore Leister Patané, Vincent Louis Viala, Antonio Jorge Martins, Ricardo Haddad, Claudia Renata dos Santos Barros, Elaine Cristina Marqueze, Raul Machado Neto, Debora Botequio Moretti, Jardelina de Souza Todao Bernardino, Loyze Paola Oliveira de Lima, Luiz Aurelio de Campos Crispin. Centro de Genômica Funcional da ESALQ: Luiz Lehmann Coutinho, Ricardo Augusto Brassaloti, Raquel de Lello Rocha Campos Cassano. NGS Soluções Genômicas: Pilar Drummond Sampaio Corrêa Mariani. FZEA-USP Pirassununga: Mirele Daiana Poleti, Jessika Cristina Chagas Lesbon, Elisangela Chicaroni Mattos, Heidge Fukumasu. USP-Botucatu: Rejane Maria Tommasini Grotto, Jayme A. Souza-Neto, Guilherme Targino Valente, Patricia Akemi Assato, Felipe Allan da Silva da Costa, Bianca Cechetto Carlos. Mendelics: Bibiana Santos, João Paulo Kitajima, Erika Freitas, David Schlesinger. Hemocentro Ribeirão Preto: Simone Kashima, Evandra Strazza Rodrigues, Svetoslav Nanev Slavov, Elaine Vieira dos Santos, Rafael dos Santos Bezerra, Luiz Carlos Junior de Alcantara, Marta Giovanetti, Vagner Fonseca, Flavia Aburjaile, Rodrigo Tocantins Calado. FAMERP-SJRP: Cecília Artico Banho, Lívia Sacchetto, Fábio Sossai Possebon, Leila Sabrina Ullmann, Cíntia Bittar, Guilherme Campos, Helena Lage Ferreira, Jorge A. Petroli Marchesi, Maisa C. Pereira Parra, Marília Moraes, Paula Rahal, Paulo Inacio da Costa, João Pessoa Araújo Jr., Maurício Lacerda Nogueira. Prefeitura de Sao Paulo: Melissa Palmieri. |
| EPI_ISL_1966358 | SMS SECRETARIA MUNICIPAL DE SAUDE DE BOITUVA | Instituto Butantan / Mendelics | Instituto Butantan: Dimas Tadeu Covas, Sandra Coccuzzo Sampaio, Maria Carolina Elias, José Salvatore Leister Patané, Vincent Louis Viala, Antonio Jorge Martins, Ricardo Haddad, Claudia Renata dos Santos Barros, Elaine Cristina Marqueze, Raul Machado Neto, Debora Botequio Moretti, Jardelina de Souza Todao Bernardino, Loyze Paola Oliveira de Lima, Luiz Aurelio de Campos Crispin. Centro de Genômica Funcional da ESALQ: Luiz Lehmann Coutinho, Ricardo Augusto Brassaloti, Raquel de Lello Rocha Campos Cassano. NGS Soluções Genômicas: Pilar Drummond Sampaio Corrêa Mariani. FZEA-USP Pirassununga: Mirele Daiana Poleti, Jessika Cristina Chagas Lesbon, Elisangela Chicaroni Mattos, Heidge Fukumasu. USP-Botucatu: Rejane Maria Tommasini Grotto, Jayme A. Souza-Neto, Guilherme Targino Valente, Patricia Akemi Assato, Felipe Allan da Silva da Costa, Bianca Cechetto Carlos. Mendelics: Bibiana Santos, João Paulo Kitajima, Erika Freitas, David Schlesinger. Hemocentro Ribeirão Preto: Simone Kashima, Evandra Strazza Rodrigues, Svetoslav Nanev Slavov, Elaine Vieira dos Santos, Rafael dos Santos Bezerra, Luiz Carlos Junior de Alcantara, Marta Giovanetti, Vagner Fonseca, Flavia Aburjaile, Rodrigo Tocantins Calado. FAMERP-SJRP: Cecília Artico Banho, Lívia Sacchetto, Fábio Sossai Possebon, Leila Sabrina Ullmann, Cíntia Bittar, Guilherme Campos, Helena Lage Ferreira, Jorge A. Petroli Marchesi, Maisa C. Pereira Parra, Marília Moraes, Paula Rahal, Paulo Inacio da Costa, João Pessoa Araújo Jr., Maurício Lacerda Nogueira. Prefeitura de Sao Paulo: Melissa Palmieri. |
| EPI_ISL_1966448 | NUCLEO DE SAUDE VILA FALCAO DE BAURU | Instituto Butantan / Mendelics | Instituto Butantan: Dimas Tadeu Covas, Sandra Coccuzzo Sampaio, Maria Carolina Elias, José Salvatore Leister Patané, Vincent Louis Viala, Antonio Jorge Martins, Ricardo Haddad, Claudia Renata dos Santos Barros, Elaine Cristina Marqueze, Raul Machado Neto, Debora Botequio Moretti, Jardelina de Souza Todao Bernardino, Loyze Paola Oliveira de Lima, Luiz Aurelio de Campos Crispin. Centro de Genômica Funcional da ESALQ: Luiz Lehmann Coutinho, Ricardo Augusto Brassaloti, Raquel de Lello Rocha Campos Cassano. NGS Soluções Genômicas: Pilar Drummond Sampaio Corrêa Mariani. FZEA-USP Pirassununga: Mirele Daiana Poleti, Jessika Cristina Chagas Lesbon, Elisangela Chicaroni Mattos, Heidge Fukumasu. USP-Botucatu: Rejane Maria Tommasini Grotto, Jayme A. Souza-Neto, Guilherme Targino Valente, Patricia Akemi Assato, Felipe Allan da Silva da Costa, Bianca Cechetto Carlos. Mendelics: Bibiana Santos, João Paulo Kitajima, Erika Freitas, David Schlesinger. Hemocentro Ribeirão Preto: Simone Kashima, Evandra Strazza Rodrigues, Svetoslav Nanev Slavov, Elaine Vieira dos Santos, Rafael dos Santos Bezerra, Luiz Carlos Junior de Alcantara, Marta Giovanetti, Vagner Fonseca, Flavia Aburjaile, Rodrigo Tocantins Calado. FAMERP-SJRP: Cecília Artico Banho, Lívia Sacchetto, Fábio Sossai Possebon, Leila Sabrina Ullmann, Cíntia Bittar, Guilherme Campos, Helena Lage Ferreira, Jorge A. Petroli Marchesi, Maisa C. Pereira Parra, Marília Moraes, Paula Rahal, Paulo Inacio da Costa, João Pessoa Araújo Jr., Maurício Lacerda Nogueira. Prefeitura de Sao Paulo: Melissa Palmieri. |
| EPI_ISL_1966526 | USF PAULISTA FERNANDOPOLIS ANTONIO PIVATO | Instituto Butantan / FZEA-USP (Pirassununga) | Instituto Butantan: Dimas Tadeu Covas, Sandra Coccuzzo Sampaio, Maria Carolina Elias, José Salvatore Leister Patané, Vincent Louis Viala, Antonio Jorge Martins, Ricardo Haddad, Claudia Renata dos Santos Barros, Elaine Cristina Marqueze, Raul Machado Neto, Debora Botequio Moretti, Jardelina de Souza Todao Bernardino, Loyze Paola Oliveira de Lima, Luiz Aurelio de Campos Crispin. Centro de Genômica Funcional da ESALQ: Luiz Lehmann Coutinho, Ricardo Augusto Brassaloti, Raquel de Lello Rocha Campos Cassano. NGS Soluções Genômicas: Pilar Drummond Sampaio Corrêa Mariani. FZEA-USP Pirassununga: Mirele Daiana Poleti, Jessika Cristina Chagas Lesbon, Elisangela Chicaroni Mattos, Heidge Fukumasu. USP-Botucatu: Rejane Maria Tommasini Grotto, Jayme A. Souza-Neto, Guilherme Targino Valente, Patricia Akemi Assato, Felipe Allan da Silva da Costa, Bianca Cechetto Carlos. Mendelics: Bibiana Santos, João Paulo Kitajima, Erika Freitas, David Schlesinger. Hemocentro Ribeirão Preto: Simone Kashima, Evandra Strazza Rodrigues, Svetoslav Nanev Slavov, Elaine Vieira dos Santos, Rafael dos Santos Bezerra, Luiz Carlos Junior de Alcantara, Marta Giovanetti, Vagner Fonseca, Flavia Aburjaile, Rodrigo Tocantins Calado. FAMERP-SJRP: Cecília Artico Banho, Lívia Sacchetto, Fábio Sossai Possebon, Leila Sabrina Ullmann, Cíntia Bittar, Guilherme Campos, Helena Lage Ferreira, Jorge A. Petroli Marchesi, Maisa C. Pereira Parra, Marília Moraes, Paula Rahal, Paulo Inacio da Costa, João Pessoa Araújo Jr., Maurício Lacerda Nogueira. Prefeitura de Sao Paulo: Melissa Palmieri. |
| EPI_ISL_1966567, EPI_ISL_1966578, EPI_ISL_1966580, EPI_ISL_1966581, EPI_ISL_1966582, EPI_ISL_1966585 | NUCLEO DE SAUDE VILA FALCAO DE BAURU | Instituto Butantan / Mendelics | Instituto Butantan: Dimas Tadeu Covas, Sandra Coccuzzo Sampaio, Maria Carolina Elias, José Salvatore Leister Patané, Vincent Louis Viala, Antonio Jorge Martins, Ricardo Haddad, Claudia Renata dos Santos Barros, Elaine Cristina Marqueze, Raul Machado Neto, Debora Botequio Moretti, Jardelina de Souza Todao Bernardino, Loyze Paola Oliveira de Lima, Luiz Aurelio de Campos Crispin. Centro de Genômica Funcional da ESALQ: Luiz Lehmann Coutinho, Ricardo Augusto Brassaloti, Raquel de Lello Rocha Campos Cassano. NGS Soluções Genômicas: Pilar Drummond Sampaio Corrêa Mariani. FZEA-USP Pirassununga: Mirele Daiana Poleti, Jessika Cristina Chagas Lesbon, Elisangela Chicaroni Mattos, Heidge Fukumasu. USP-Botucatu: Rejane Maria Tommasini Grotto, Jayme A. Souza-Neto, Guilherme Targino Valente, Patricia Akemi Assato, Felipe Allan da Silva da Costa, Bianca Cechetto Carlos. Mendelics: Bibiana Santos, João Paulo Kitajima, Erika Freitas, David Schlesinger. Hemocentro Ribeirão Preto: Simone Kashima, Evandra Strazza Rodrigues, Svetoslav Nanev Slavov, Elaine Vieira dos Santos, Rafael dos Santos Bezerra, Luiz Carlos Junior de Alcantara, Marta Giovanetti, Vagner |

| | | | |
|---|---|---|---|
| | | | Fonseca, Flavia Aburjaile, Rodrigo Tocantins Calado. FAMERP-SJRP: Cecília Artico Banho, Lívia Sacchetto, Fábio Sossai Possebon, Leila Sabrina Ullmann, Cíntia Bittar, Guilherme Campos, Helena Lage Ferreira, Jorge A. Petroli Marchesi, Maisa C. Pereira Parra, Marília Moraes, Paula Rahal, Paulo Inacio da Costa, João Pessoa Araújo Jr., Maurício Lacerda Nogueira. Prefeitura de Sao Paulo: Melissa Palmieri. |
| EPI_ISL_1966658 | UPA UNIDADE DE PRONTO ATENDIMENTO 24 HORAS BOM JESUS | Instituto Butantan / Mendelics | Instituto Butantan: Dimas Tadeu Covas, Sandra Coccuzzo Sampaio, Maria Carolina Elias, José Salvatore Leister Patané, Vincent Louis Viala, Antonio Jorge Martins, Ricardo Haddad, Claudia Renata dos Santos Barros, Elaine Cristina Marqueze, Raul Machado Neto, Debora Botequio Moretti, Jardelina de Souza Todao Bernardino, Loyze Paola Oliveira de Lima, Luiz Aurelio de Campos Crispin. Centro de Genômica Funcional da ESALQ: Luiz Lehmann Coutinho, Ricardo Augusto Brassaloti, Raquel de Lello Rocha Campos Cassano. NGS Soluções Genômicas: Pilar Drummond Sampaio Corrêa Mariani. FZEA-USP Pirassununga: Mirele Daiana Poleti, Jessika Cristina Chagas Lesbon, Elisangela Chicaroni Mattos, Heidge Fukumasu. USP-Botucatu: Rejane Maria Tommasini Grotto, Jayme A. Souza-Neto, Guilherme Targino Valente, Patricia Akemi Assato, Felipe Allan da Silva da Costa, Bianca Cechetto Carlos. Mendelics: Bibiana Santos, João Paulo Kitajima, Erika Freitas, David Schlesinger. Hemocentro Ribeirão Preto: Simone Kashima, Evandra Strazza Rodrigues, Svetoslav Nanev Slavov, Elaine Vieira dos Santos, Rafael dos Santos Bezerra, Luiz Carlos Junior de Alcantara, Marta Giovanetti, Vagner Fonseca, Flavia Aburjaile, Rodrigo Tocantins Calado. FAMERP-SJRP: Cecília Artico Banho, Lívia Sacchetto, Fábio Sossai Possebon, Leila Sabrina Ullmann, Cíntia Bittar, Guilherme Campos, Helena Lage Ferreira, Jorge A. Petroli Marchesi, Maisa C. Pereira Parra, Marília Moraes, Paula Rahal, Paulo Inacio da Costa, João Pessoa Araújo Jr., Maurício Lacerda Nogueira. Prefeitura de Sao Paulo: Melissa Palmieri. |
| EPI_ISL_1966926 | HOSPITAL REGIONAL DE ITAPETININGA | Instituto Butantan / ESALQ-USP (Piracicaba) | Instituto Butantan: Dimas Tadeu Covas, Sandra Coccuzzo Sampaio, Maria Carolina Elias, José Salvatore Leister Patané, Vincent Louis Viala, Antonio Jorge Martins, Ricardo Haddad, Claudia Renata dos Santos Barros, Elaine Cristina Marqueze, Raul Machado Neto, Debora Botequio Moretti, Jardelina de Souza Todao Bernardino, Loyze Paola Oliveira de Lima, Luiz Aurelio de Campos Crispin. Centro de Genômica Funcional da ESALQ: Luiz Lehmann Coutinho, Ricardo Augusto Brassaloti, Raquel de Lello Rocha Campos Cassano. NGS Soluções Genômicas: Pilar Drummond Sampaio Corrêa Mariani. FZEA-USP Pirassununga: Mirele Daiana Poleti, Jessika Cristina Chagas Lesbon, Elisangela Chicaroni Mattos, Heidge Fukumasu. USP-Botucatu: Rejane Maria Tommasini Grotto, Jayme A. Souza-Neto, Guilherme Targino Valente, Patricia Akemi Assato, Felipe Allan da Silva da Costa, Bianca Cechetto Carlos. Mendelics: Bibiana Santos, João Paulo Kitajima, Erika Freitas, David Schlesinger. Hemocentro Ribeirão Preto: Simone Kashima, Evandra Strazza Rodrigues, Svetoslav Nanev Slavov, Elaine Vieira dos Santos, Rafael dos Santos Bezerra, Luiz Carlos Junior de Alcantara, Marta Giovanetti, Vagner Fonseca, Flavia Aburjaile, Rodrigo Tocantins Calado. FAMERP-SJRP: Cecília Artico Banho, Lívia Sacchetto, Fábio Sossai Possebon, Leila Sabrina Ullmann, Cíntia Bittar, Guilherme Campos, Helena Lage Ferreira, Jorge A. Petroli Marchesi, Maisa C. Pereira Parra, Marília Moraes, Paula Rahal, Paulo Inacio da Costa, João Pessoa Araújo Jr., Maurício Lacerda Nogueira. Prefeitura de Sao Paulo: Melissa Palmieri. |
| EPI_ISL_1967284 | UBS RECREIO SAO JORGE | Instituto Butantan / Mendelics | Instituto Butantan: Dimas Tadeu Covas, Sandra Coccuzzo Sampaio, Maria Carolina Elias, José Salvatore Leister Patané, Vincent Louis Viala, Antonio Jorge Martins, Ricardo Haddad, Claudia Renata dos Santos Barros, Elaine Cristina Marqueze, Raul Machado Neto, Debora Botequio Moretti, Jardelina de Souza Todao Bernardino, Loyze Paola Oliveira de Lima, Luiz Aurelio de Campos Crispin. Centro de Genômica Funcional da ESALQ: Luiz Lehmann Coutinho, Ricardo Augusto Brassaloti, Raquel de Lello Rocha Campos Cassano. NGS Soluções Genômicas: Pilar Drummond Sampaio Corrêa Mariani. FZEA-USP Pirassununga: Mirele Daiana Poleti, Jessika Cristina Chagas Lesbon, Elisangela Chicaroni Mattos, Heidge Fukumasu. USP-Botucatu: Rejane Maria Tommasini Grotto, Jayme A. Souza-Neto, Guilherme Targino Valente, Patricia Akemi Assato, Felipe Allan da Silva da Costa, Bianca Cechetto Carlos. Mendelics: Bibiana Santos, João Paulo Kitajima, Erika Freitas, David Schlesinger. Hemocentro Ribeirão Preto: Simone Kashima, Evandra Strazza Rodrigues, Svetoslav Nanev Slavov, Elaine Vieira dos Santos, Rafael dos Santos Bezerra, Luiz Carlos Junior de Alcantara, Marta Giovanetti, Vagner Fonseca, Flavia Aburjaile, Rodrigo Tocantins Calado. FAMERP-SJRP: Cecília Artico Banho, Lívia Sacchetto, Fábio Sossai Possebon, Leila Sabrina Ullmann, Cíntia Bittar, Guilherme Campos, Helena Lage Ferreira, Jorge A. Petroli Marchesi, Maisa C. Pereira Parra, Marília Moraes, Paula Rahal, Paulo Inacio da Costa, João Pessoa Araújo Jr., Maurício Lacerda Nogueira. Prefeitura de Sao Paulo: Melissa Palmieri. |
| EPI_ISL_2003120, EPI_ISL_2003123 | Instituto Adolfo Lutz Central | Instituto Adolfo Lutz, Interdiciplinary Procedures Center, Strategic Laboratory | Claudio Tavares Sacchi, Claudia Regina Gonçalves, Erica Valessa Ramos Gomes, Karoline Rodrigues Campos, Caio Vinicius Dias Lopes, Leonardo Jose Tadeu de Araujo |
| EPI_ISL_2020090, EPI_ISL_2020097, EPI_ISL_2020105 | HOSPITAL UNIVERSITARIO SON ESPASES | HOSPITAL UNIVERSITARIO SON ESPASES | Carla López-Causapé, Pablo Fraile-Ribot, Antonio Oliver, SeqCovid |
| EPI_ISL_2031726 | Centro de Innovación en Vigilancia Epidemiológica (CiVE), Institut Pasteur Montevideo, Uruguay | Centro de Innovación en Vigilancia Epidemiológica (CiVE), Institut Pasteur Montevideo, Uruguay | Natalia Rego, Alicia Costábile, Mercedes Paz, Cecilia Salazar, Paula Perbolianachis, Tamara Fernández, Ignacio Ferrés, Rodrigo Arce, Alvaro Fajardo, Mailen Arleo, Tania Possi, Inés Bellini, Lucia Bilbao, Natalia Reyes, Ma Noel Bentancor, Andrés Lizosaín, María José Benítez, Odhille Chappos, Melissa Duquía, Belén González, Luciana Griffero, Mauricio Méndez, Ma Pía Techera, Juan Zanetti, Bernardina Rivera, Matías Maidana, Martina Alonso, Cecilia Alonso, Julio Medina, Henry Albornoz, Rodney Colina, Gregorio Iraola, Lucía Spangenberg, Gonzalo Moratorio, Pilar Moreno |
| EPI_ISL_2037442 | Instituto de Virologia "Dr. J. M. Vanella", Facultad de Ciencias Médicas, Universidad Nacional de Córdoba. | Centro de Investigaciones Agropecuarias (CIAP), Instituto Nacional de Tecnologia Agropecuaria (INTA) Cordoba, Argentina, on behalf of 'Proyecto Argentino Interinstitucional de Genomica de SARS-CoV-2 (PAIS Consortium) | Brenda Konigheim, Lorena Spinsanti, Adrian Diaz, Javier Aguilar, Sebastian Blanco, Mauricio Beranek, Maria Elisa Rivarola, Maria Belen Pisano, Viviana Re, Gonzalo Castro, Gabriela Barbas, Franco Fernandez, Humberto Debat, Nathalie Marquez, Sandra Gallego. |
| EPI_ISL_2038956 | Laboratorio Central de Saude Publica do Estado do Rio de Janeiro (LACEN-RJ) | Laboratory of Respiratory Viruses and Measles, Oswaldo Cruz Institute, FIOCRUZ | Paola Resende, Luciana Appolinario, Fernando Motta, Anna Carolina Paixao, Ana Carolina Mendonca, Alice Sampaio Rocha, Taina Venas, Elisa Cavalcante Pereira, Renata Serrano Lopes, Andrea Cony Cavalcanti, Marilda Siqueira on behalf of the Fiocruz COVID-19 Genomic Surveillance Network |
| EPI_ISL_2038961, EPI_ISL_2038962, EPI_ISL_2038963 | Laboratory of Respiratory Viruses and Measles, Oswaldo Cruz Institute, FIOCRUZ | Laboratory of Respiratory Viruses and Measles, Oswaldo Cruz Institute, FIOCRUZ | Paola Resende, Luciana Appolinario, Fernando Motta, Anna Carolina Paixao, Ana Carolina Mendonca, Alice Sampaio Rocha, Taina Venas, Elisa Cavalcante Pereira, Renata Serrano Lopes, Marilda Siqueira on behalf of the Fiocruz COVID-19 Genomic Surveillance Network |
| EPI_ISL_2086268 | Laboratory of Molecular Biology of Flavivirus, Oswaldo Cruz Institute, FIOCRUZ | Laboratory of Molecular Biology of Flavivirus, Oswaldo Cruz Institute, FIOCRUZ | Fernando Lopez Tort, Ieda Ribeiro, Lidiane Menezes, Alexandre dos Santos, Marta Pereira, Myrna Bonaldo |
| EPI_ISL_2093854, EPI_ISL_2093855, EPI_ISL_2093871 | Dutch COVID-19 response team | National Institute for Public Health and the Environment (RIVM) | Adam Meijer, Harry Vennema, Dirk Eggink, Jeroen Cremer, Sharon van den Brink, Bas van der Veer, AnneMarie van den Brandt, Lisa Wijsman, Kim Freriks, Ryanne Jaarsma, Euníce Then, Lynn Aarts, Sanne Bos, Melissa van Tuil, Linda van de Nes, Sjoerd Kuiling, James Groot, Florian Zwagemaker, Dennis Schmitz, Annelies Kroneman, Karim Hajji, Chantal Reusken, on behalf of the national COVID-19 response team |
| EPI_ISL_2101392 | Unidade de apoio ao diagnóstico da COVID - UNADIG | Bioinformatics Laboratory / LNCC | Luiz G P de Almeida, Alessandra P Lamarca, Ronaldo da Silva F Jr, Liliane Cavalcante, Alexandra L Gerber, Ana Paula de C Guimaraes, Douglas Terra Machado, Cassia Alves, Diana Mariani, Cintia Policarpo, Gleidson da Silva de Oliveira, Mario Sergio Ribeiro, Silvia Carvalho, Flavio Dias da Silva, Marcio Henrique de Oliveira Garcia, Leandro Magalhaes de Souza, Cristiane Gomes da Silva, Caio Luiz Pereira Ribeiro, Andrea Cony Cavalcanti, Claudia Maria Braga de Mello, Amilcar Tanuri, Ana Tereza R Vasconcelos |
| EPI_ISL_2101393 | Laboratorio Central Noel Nutels | Bioinformatics Laboratory / LNCC | Luiz G P de Almeida, Alessandra P Lamarca, Ronaldo da Silva F Jr, Liliane Cavalcante, Alexandra L Gerber, Ana Paula de C Guimaraes, Douglas Terra Machado, Cassia Alves, Diana Mariani, Cintia Policarpo, Gleidson da Silva de Oliveira, Mario Sergio Ribeiro, Silvia Carvalho, Flavio Dias da Silva, Marcio Henrique de Oliveira Garcia, Leandro Magalhaes de Souza, Cristiane Gomes da Silva, Caio Luiz Pereira Ribeiro, Andrea Cony Cavalcanti, Claudia Maria Braga de Mello, Amilcar Tanuri, Ana Tereza R Vasconcelos |
| EPI_ISL_2101394 | Unidade de apoio ao diagnóstico da COVID - UNADIG | Bioinformatics Laboratory / LNCC | Luiz G P de Almeida, Alessandra P Lamarca, Ronaldo da Silva F Jr, Liliane Cavalcante, Alexandra L Gerber, Ana Paula de C Guimaraes, Douglas Terra Machado, Cassia Alves, Diana Mariani, Cintia Policarpo, Gleidson da Silva de Oliveira, Mario Sergio Ribeiro, Silvia Carvalho, Flavio Dias da Silva, Marcio Henrique de Oliveira Garcia, Leandro Magalhaes de Souza, Cristiane Gomes da Silva, Caio Luiz Pereira Ribeiro, Andrea Cony Cavalcanti, Claudia Maria Braga de Mello, Amilcar Tanuri, Ana Tereza R Vasconcelos |
| EPI_ISL_2101395, EPI_ISL_2101396, EPI_ISL_2101397, EPI_ISL_2101398, EPI_ISL_2101399, EPI_ISL_2101400, EPI_ISL_2101401, EPI_ISL_2101403, EPI_ISL_2101404, EPI_ISL_2101405, EPI_ISL_2101406, EPI_ISL_2101410, EPI_ISL_2101411, EPI_ISL_2101412, EPI_ISL_2101413, EPI_ISL_2101414, EPI_ISL_2101415, EPI_ISL_2101416, EPI_ISL_2101417, EPI_ISL_2101418, EPI_ISL_2101419, EPI_ISL_2101420, EPI_ISL_2101421, EPI_ISL_2101422, EPI_ISL_2101423, EPI_ISL_2101424, EPI_ISL_2101425, EPI_ISL_2101426, EPI_ISL_2101427, EPI_ISL_2101428 | | | |
| see above | Laboratorio Central Noel Nutels | Bioinformatics Laboratory / LNCC | Luiz G P de Almeida, Alessandra P Lamarca, Ronaldo da Silva F Jr, Liliane Cavalcante, Alexandra L Gerber, Ana Paula de C Guimaraes, Douglas Terra Machado, Cassia Alves, Diana Mariani, Cintia Policarpo, Gleidson da Silva de Oliveira, Mario Sergio Ribeiro, Silvia Carvalho, Flavio Dias da Silva, Marcio Henrique de Oliveira Garcia, Leandro Magalhaes de Souza, Cristiane Gomes da Silva, Caio Luiz Pereira Ribeiro, Andrea Cony Cavalcanti, Claudia Maria Braga de Mello, Amilcar Tanuri, Ana Tereza R Vasconcelos |
| EPI_ISL_2101429, EPI_ISL_2101430, EPI_ISL_2101431, EPI_ISL_2101432, EPI_ISL_2101433 | Unidade de apoio ao diagnóstico da COVID - UNADIG | Bioinformatics Laboratory / LNCC | Luiz G P de Almeida, Alessandra P Lamarca, Ronaldo da Silva F Jr, Liliane Cavalcante, Alexandra L Gerber, Ana Paula de C Guimaraes, Douglas Terra Machado, Cassia Alves, Diana Mariani, Cintia Policarpo, Gleidson da Silva de Oliveira, Mario Sergio Ribeiro, Silvia Carvalho, Flavio Dias da Silva, Marcio Henrique de Oliveira Garcia, Leandro Magalhaes de Souza, Cristiane Gomes da Silva, Caio Luiz Pereira Ribeiro, Andrea Cony Cavalcanti, Claudia Maria Braga de Mello, Amilcar Tanuri, Ana Tereza R Vasconcelos |

| | | | |
|---|---|---|---|
| EPI_ISL_2101434, EPI_ISL_2101435, EPI_ISL_2101436, EPI_ISL_2101437, EPI_ISL_2101438, EPI_ISL_2101439, EPI_ISL_2101440, EPI_ISL_2101441, EPI_ISL_2101442 | Laboratorio Central Noel Nutels | Bioinformatics Laboratory / LNCC | Luiz G P de Almeida, Alessandra P Lamarca, Ronaldo da Silva F Jr, Liliane Cavalcante, Alexandra L Gerber, Ana Paula de C Guimaraes, Douglas Terra Machado, Cassia Alves, Diana Mariani, Cintia Policarpo, Gleidson da Silva de Oliveira, Mario Sergio Ribeiro, Silvia Carvalho, Flavio Dias da Silva, Marcio Henrique de Oliveira Garcia, Leandro Magalhaes de Souza, Cristiane Gomes da Silva, Caio Luiz Pereira Ribeiro, Andrea Cony Cavalcanti, Claudia Maria Braga de Mello, Amilcar Tanuri, Ana Tereza R Vasconcelos |
| EPI_ISL_2101443 | Unidade de apoio ao diagnóstico da COVID - UNADIG | Bioinformatics Laboratory / LNCC | Luiz G P de Almeida, Alessandra P Lamarca, Ronaldo da Silva F Jr, Liliane Cavalcante, Alexandra L Gerber, Ana Paula de C Guimaraes, Douglas Terra Machado, Cassia Alves, Diana Mariani, Cintia Policarpo, Gleidson da Silva de Oliveira, Mario Sergio Ribeiro, Silvia Carvalho, Flavio Dias da Silva, Marcio Henrique de Oliveira Garcia, Leandro Magalhaes de Souza, Cristiane Gomes da Silva, Caio Luiz Pereira Ribeiro, Andrea Cony Cavalcanti, Claudia Maria Braga de Mello, Amilcar Tanuri, Ana Tereza R Vasconcelos |
| EPI_ISL_2101444, EPI_ISL_2101445, EPI_ISL_2101446, EPI_ISL_2101447, EPI_ISL_2101448, EPI_ISL_2101449, EPI_ISL_2101450, EPI_ISL_2101451, EPI_ISL_2101452, EPI_ISL_2101453, EPI_ISL_2101454 | | | |
| see above | Laboratorio Central Noel Nutels | Bioinformatics Laboratory / LNCC | Luiz G P de Almeida, Alessandra P Lamarca, Ronaldo da Silva F Jr, Liliane Cavalcante, Alexandra L Gerber, Ana Paula de C Guimaraes, Douglas Terra Machado, Cassia Alves, Diana Mariani, Cintia Policarpo, Gleidson da Silva de Oliveira, Mario Sergio Ribeiro, Silvia Carvalho, Flavio Dias da Silva, Marcio Henrique de Oliveira Garcia, Leandro Magalhaes de Souza, Cristiane Gomes da Silva, Caio Luiz Pereira Ribeiro, Andrea Cony Cavalcanti, Claudia Maria Braga de Mello, Amilcar Tanuri, Ana Tereza R Vasconcelos |
| EPI_ISL_2101455, EPI_ISL_2101456, EPI_ISL_2101457, EPI_ISL_2101458, EPI_ISL_2101459, EPI_ISL_2101460, EPI_ISL_2101461, EPI_ISL_2101462, EPI_ISL_2101463 | Unidade de apoio ao diagnóstico da COVID - UNADIG | Bioinformatics Laboratory / LNCC | Luiz G P de Almeida, Alessandra P Lamarca, Ronaldo da Silva F Jr, Liliane Cavalcante, Alexandra L Gerber, Ana Paula de C Guimaraes, Douglas Terra Machado, Cassia Alves, Diana Mariani, Cintia Policarpo, Gleidson da Silva de Oliveira, Mario Sergio Ribeiro, Silvia Carvalho, Flavio Dias da Silva, Marcio Henrique de Oliveira Garcia, Leandro Magalhaes de Souza, Cristiane Gomes da Silva, Caio Luiz Pereira Ribeiro, Andrea Cony Cavalcanti, Claudia Maria Braga de Mello, Amilcar Tanuri, Ana Tereza R Vasconcelos |
| EPI_ISL_2101464, EPI_ISL_2101465, EPI_ISL_2101466, EPI_ISL_2101467, EPI_ISL_2101468, EPI_ISL_2101469 | Laboratorio Central Noel Nutels | Bioinformatics Laboratory / LNCC | Luiz G P de Almeida, Alessandra P Lamarca, Ronaldo da Silva F Jr, Liliane Cavalcante, Alexandra L Gerber, Ana Paula de C Guimaraes, Douglas Terra Machado, Cassia Alves, Diana Mariani, Cintia Policarpo, Gleidson da Silva de Oliveira, Mario Sergio Ribeiro, Silvia Carvalho, Flavio Dias da Silva, Marcio Henrique de Oliveira Garcia, Leandro Magalhaes de Souza, Cristiane Gomes da Silva, Caio Luiz Pereira Ribeiro, Andrea Cony Cavalcanti, Claudia Maria Braga de Mello, Amilcar Tanuri, Ana Tereza R Vasconcelos |
| EPI_ISL_2101470, EPI_ISL_2101471, EPI_ISL_2101472, EPI_ISL_2101473 | Unidade de apoio ao diagnóstico da COVID - UNADIG | Bioinformatics Laboratory / LNCC | Luiz G P de Almeida, Alessandra P Lamarca, Ronaldo da Silva F Jr, Liliane Cavalcante, Alexandra L Gerber, Ana Paula de C Guimaraes, Douglas Terra Machado, Cassia Alves, Diana Mariani, Cintia Policarpo, Gleidson da Silva de Oliveira, Mario Sergio Ribeiro, Silvia Carvalho, Flavio Dias da Silva, Marcio Henrique de Oliveira Garcia, Leandro Magalhaes de Souza, Cristiane Gomes da Silva, Caio Luiz Pereira Ribeiro, Andrea Cony Cavalcanti, Claudia Maria Braga de Mello, Amilcar Tanuri, Ana Tereza R Vasconcelos |
| EPI_ISL_2101474 | Laboratorio Central Noel Nutels | Bioinformatics Laboratory / LNCC | Luiz G P de Almeida, Alessandra P Lamarca, Ronaldo da Silva F Jr, Liliane Cavalcante, Alexandra L Gerber, Ana Paula de C Guimaraes, Douglas Terra Machado, Cassia Alves, Diana Mariani, Cintia Policarpo, Gleidson da Silva de Oliveira, Mario Sergio Ribeiro, Silvia Carvalho, Flavio Dias da Silva, Marcio Henrique de Oliveira Garcia, Leandro Magalhaes de Souza, Cristiane Gomes da Silva, Caio Luiz Pereira Ribeiro, Andrea Cony Cavalcanti, Claudia Maria Braga de Mello, Amilcar Tanuri, Ana Tereza R Vasconcelos |
| EPI_ISL_2101475 | Unidade de apoio ao diagnóstico da COVID - UNADIG | Bioinformatics Laboratory / LNCC | Luiz G P de Almeida, Alessandra P Lamarca, Ronaldo da Silva F Jr, Liliane Cavalcante, Alexandra L Gerber, Ana Paula de C Guimaraes, Douglas Terra Machado, Cassia Alves, Diana Mariani, Cintia Policarpo, Gleidson da Silva de Oliveira, Mario Sergio Ribeiro, Silvia Carvalho, Flavio Dias da Silva, Marcio Henrique de Oliveira Garcia, Leandro Magalhaes de Souza, Cristiane Gomes da Silva, Caio Luiz Pereira Ribeiro, Andrea Cony Cavalcanti, Claudia Maria Braga de Mello, Amilcar Tanuri, Ana Tereza R Vasconcelos |
| EPI_ISL_2101476, EPI_ISL_2101477, EPI_ISL_2101478, EPI_ISL_2101479, EPI_ISL_2101480, EPI_ISL_2101481, EPI_ISL_2101482 | Laboratorio Central Noel Nutels | Bioinformatics Laboratory / LNCC | Luiz G P de Almeida, Alessandra P Lamarca, Ronaldo da Silva F Jr, Liliane Cavalcante, Alexandra L Gerber, Ana Paula de C Guimaraes, Douglas Terra Machado, Cassia Alves, Diana Mariani, Cintia Policarpo, Gleidson da Silva de Oliveira, Mario Sergio Ribeiro, Silvia Carvalho, Flavio Dias da Silva, Marcio Henrique de Oliveira Garcia, Leandro Magalhaes de Souza, Cristiane Gomes da Silva, Caio Luiz Pereira Ribeiro, Andrea Cony Cavalcanti, Claudia Maria Braga de Mello, Amilcar Tanuri, Ana Tereza R Vasconcelos |
| EPI_ISL_2101483, EPI_ISL_2101484 | Unidade de apoio ao diagnóstico da COVID - UNADIG | Bioinformatics Laboratory / LNCC | Luiz G P de Almeida, Alessandra P Lamarca, Ronaldo da Silva F Jr, Liliane Cavalcante, Alexandra L Gerber, Ana Paula de C Guimaraes, Douglas Terra Machado, Cassia Alves, Diana Mariani, Cintia Policarpo, Gleidson da Silva de Oliveira, Mario Sergio Ribeiro, Silvia Carvalho, Flavio Dias da Silva, Marcio Henrique de Oliveira Garcia, Leandro Magalhaes de Souza, Cristiane Gomes da Silva, Caio Luiz Pereira Ribeiro, Andrea Cony Cavalcanti, Claudia Maria Braga de Mello, Amilcar Tanuri, Ana Tereza R Vasconcelos |
| EPI_ISL_2101485, EPI_ISL_2101486, EPI_ISL_2101487, EPI_ISL_2101488, EPI_ISL_2101489, EPI_ISL_2101490, EPI_ISL_2101491, EPI_ISL_2101492, EPI_ISL_2101493 | Laboratorio Central Noel Nutels | Bioinformatics Laboratory / LNCC | Luiz G P de Almeida, Alessandra P Lamarca, Ronaldo da Silva F Jr, Liliane Cavalcante, Alexandra L Gerber, Ana Paula de C Guimaraes, Douglas Terra Machado, Cassia Alves, Diana Mariani, Cintia Policarpo, Gleidson da Silva de Oliveira, Mario Sergio Ribeiro, Silvia Carvalho, Flavio Dias da Silva, Marcio Henrique de Oliveira Garcia, Leandro Magalhaes de Souza, Cristiane Gomes da Silva, Caio Luiz Pereira Ribeiro, Andrea Cony Cavalcanti, Claudia Maria Braga de Mello, Amilcar Tanuri, Ana Tereza R Vasconcelos |
| EPI_ISL_2101494, EPI_ISL_2101495 | Unidade de apoio ao diagnóstico da COVID - UNADIG | Bioinformatics Laboratory / LNCC | Luiz G P de Almeida, Alessandra P Lamarca, Ronaldo da Silva F Jr, Liliane Cavalcante, Alexandra L Gerber, Ana Paula de C Guimaraes, Douglas Terra Machado, Cassia Alves, Diana Mariani, Cintia Policarpo, Gleidson da Silva de Oliveira, Mario Sergio Ribeiro, Silvia Carvalho, Flavio Dias da Silva, Marcio Henrique de Oliveira Garcia, Leandro Magalhaes de Souza, Cristiane Gomes da Silva, Caio Luiz Pereira Ribeiro, Andrea Cony Cavalcanti, Claudia Maria Braga de Mello, Amilcar Tanuri, Ana Tereza R Vasconcelos |
| EPI_ISL_2101496, EPI_ISL_2101497, EPI_ISL_2101498, EPI_ISL_2101499, EPI_ISL_2101500, EPI_ISL_2101501, EPI_ISL_2101502, EPI_ISL_2101503, EPI_ISL_2101504 | Laboratorio Central Noel Nutels | Bioinformatics Laboratory / LNCC | Luiz G P de Almeida, Alessandra P Lamarca, Ronaldo da Silva F Jr, Liliane Cavalcante, Alexandra L Gerber, Ana Paula de C Guimaraes, Douglas Terra Machado, Cassia Alves, Diana Mariani, Cintia Policarpo, Gleidson da Silva de Oliveira, Mario Sergio Ribeiro, Silvia Carvalho, Flavio Dias da Silva, Marcio Henrique de Oliveira Garcia, Leandro Magalhaes de Souza, Cristiane Gomes da Silva, Caio Luiz Pereira Ribeiro, Andrea Cony Cavalcanti, Claudia Maria Braga de Mello, Amilcar Tanuri, Ana Tereza R Vasconcelos |
| EPI_ISL_2101505, EPI_ISL_2101506 | Unidade de apoio ao diagnóstico da COVID - UNADIG | Bioinformatics Laboratory / LNCC | Luiz G P de Almeida, Alessandra P Lamarca, Ronaldo da Silva F Jr, Liliane Cavalcante, Alexandra L Gerber, Ana Paula de C Guimaraes, Douglas Terra Machado, Cassia Alves, Diana Mariani, Cintia Policarpo, Gleidson da Silva de Oliveira, Mario Sergio Ribeiro, Silvia Carvalho, Flavio Dias da Silva, Marcio Henrique de Oliveira Garcia, Leandro Magalhaes de Souza, Cristiane Gomes da Silva, Caio Luiz Pereira Ribeiro, Andrea Cony Cavalcanti, Claudia Maria Braga de Mello, Amilcar Tanuri, Ana Tereza R Vasconcelos |
| EPI_ISL_2101507 | Laboratorio Central Noel Nutels | Bioinformatics Laboratory / LNCC | Luiz G P de Almeida, Alessandra P Lamarca, Ronaldo da Silva F Jr, Liliane Cavalcante, Alexandra L Gerber, Ana Paula de C Guimaraes, Douglas Terra Machado, Cassia Alves, Diana Mariani, Cintia Policarpo, Gleidson da Silva de Oliveira, Mario Sergio Ribeiro, Silvia Carvalho, Flavio Dias da Silva, Marcio Henrique de Oliveira Garcia, Leandro Magalhaes de Souza, Cristiane Gomes da Silva, Caio Luiz Pereira Ribeiro, Andrea Cony Cavalcanti, Claudia Maria Braga de Mello, Amilcar Tanuri, Ana Tereza R Vasconcelos |
| EPI_ISL_2101508, EPI_ISL_2101509, EPI_ISL_2101510, EPI_ISL_2101511, EPI_ISL_2101512, EPI_ISL_2101513 | Unidade de apoio ao diagnóstico da COVID - UNADIG | Bioinformatics Laboratory / LNCC | Luiz G P de Almeida, Alessandra P Lamarca, Ronaldo da Silva F Jr, Liliane Cavalcante, Alexandra L Gerber, Ana Paula de C Guimaraes, Douglas Terra Machado, Cassia Alves, Diana Mariani, Cintia Policarpo, Gleidson da Silva de Oliveira, Mario Sergio Ribeiro, Silvia Carvalho, Flavio Dias da Silva, Marcio Henrique de Oliveira Garcia, Leandro Magalhaes de Souza, Cristiane Gomes da Silva, Caio Luiz Pereira Ribeiro, Andrea Cony Cavalcanti, Claudia Maria Braga de Mello, Amilcar Tanuri, Ana Tereza R Vasconcelos |
| EPI_ISL_2101514, EPI_ISL_2101515, EPI_ISL_2101516, EPI_ISL_2101517, EPI_ISL_2101518, EPI_ISL_2101519 | Laboratorio Central Noel Nutels | Bioinformatics Laboratory / LNCC | Luiz G P de Almeida, Alessandra P Lamarca, Ronaldo da Silva F Jr, Liliane Cavalcante, Alexandra L Gerber, Ana Paula de C Guimaraes, Douglas Terra Machado, Cassia Alves, Diana Mariani, Cintia Policarpo, Gleidson da Silva de Oliveira, Mario Sergio Ribeiro, Silvia Carvalho, Flavio Dias da Silva, Marcio Henrique de Oliveira Garcia, Leandro Magalhaes de Souza, Cristiane Gomes da Silva, Caio Luiz Pereira Ribeiro, Andrea Cony Cavalcanti, Claudia Maria Braga de Mello, Amilcar Tanuri, Ana Tereza R Vasconcelos |
| EPI_ISL_2101520, EPI_ISL_2101521, EPI_ISL_2101522, EPI_ISL_2101523, EPI_ISL_2101524, EPI_ISL_2101525, EPI_ISL_2101526, EPI_ISL_2101527 | Unidade de apoio ao diagnóstico da COVID - UNADIG | Bioinformatics Laboratory / LNCC | Luiz G P de Almeida, Alessandra P Lamarca, Ronaldo da Silva F Jr, Liliane Cavalcante, Alexandra L Gerber, Ana Paula de C Guimaraes, Douglas Terra Machado, Cassia Alves, Diana Mariani, Cintia Policarpo, Gleidson da Silva de Oliveira, Mario Sergio Ribeiro, Silvia Carvalho, Flavio Dias da Silva, Marcio Henrique de Oliveira Garcia, Leandro Magalhaes de Souza, Cristiane Gomes da Silva, Caio Luiz Pereira Ribeiro, Andrea Cony Cavalcanti, Claudia Maria Braga de Mello, Amilcar Tanuri, Ana Tereza R Vasconcelos |
| EPI_ISL_2101528, EPI_ISL_2101529, EPI_ISL_2101530, EPI_ISL_2101531, EPI_ISL_2101532, EPI_ISL_2101533, EPI_ISL_2101534, EPI_ISL_2101535, EPI_ISL_2101536, EPI_ISL_2101537, EPI_ISL_2101538, EPI_ISL_2101539, EPI_ISL_2101540, EPI_ISL_2101541, EPI_ISL_2101542, EPI_ISL_2101543, EPI_ISL_2101544, | | | |

| | | | |
|---|---|---|---|
| EPI_ISL_2101545, EPI_ISL_2101546, EPI_ISL_2101547, EPI_ISL_2101548, EPI_ISL_2101549, EPI_ISL_2101550, EPI_ISL_2101551, EPI_ISL_2101552, EPI_ISL_2101553, EPI_ISL_2101554, EPI_ISL_2101555, EPI_ISL_2101556, EPI_ISL_2101557, EPI_ISL_2101558, EPI_ISL_2101559, EPI_ISL_2101560, EPI_ISL_2101561, EPI_ISL_2101562, EPI_ISL_2101563, EPI_ISL_2101564, EPI_ISL_2101565 | | | |
| see above | Laboratorio Central Noel Nutels | Bioinformatics Laboratory / LNCC | Luiz G P de Almeida, Alessandra P Lamarca, Ronaldo da Silva F Jr, Liliane Cavalcante, Alexandra L Gerber, Ana Paula de C Guimaraes, Douglas Terra Machado, Cassia Alves, Diana Mariani, Cintia Policarpo, Gleidson da Silva de Oliveira, Mario Sergio Ribeiro, Silvia Carvalho, Flavio Dias da Silva, Marcio Henrique de Oliveira Garcia, Leandro Magalhaes de Souza, Cristiane Gomes da Silva, Caio Luiz Pereira Ribeiro, Andrea Cony Cavalcanti, Claudia Maria Braga de Mello, Amilcar Tanuri, Ana Tereza R Vasconcelos |
| EPI_ISL_2101566, EPI_ISL_2101567, EPI_ISL_2101568, EPI_ISL_2101569, EPI_ISL_2101570, EPI_ISL_2101571, EPI_ISL_2101572 | Unidade de apoio ao diagnóstico da COVID - UNADIG | Bioinformatics Laboratory / LNCC | Luiz G P de Almeida, Alessandra P Lamarca, Ronaldo da Silva F Jr, Liliane Cavalcante, Alexandra L Gerber, Ana Paula de C Guimaraes, Douglas Terra Machado, Cassia Alves, Diana Mariani, Cintia Policarpo, Gleidson da Silva de Oliveira, Mario Sergio Ribeiro, Silvia Carvalho, Flavio Dias da Silva, Marcio Henrique de Oliveira Garcia, Leandro Magalhaes de Souza, Cristiane Gomes da Silva, Caio Luiz Pereira Ribeiro, Andrea Cony Cavalcanti, Claudia Maria Braga de Mello, Amilcar Tanuri, Ana Tereza R Vasconcelos |
| EPI_ISL_2101573, EPI_ISL_2101574, EPI_ISL_2101575, EPI_ISL_2101576, EPI_ISL_2101577, EPI_ISL_2101578, EPI_ISL_2101579, EPI_ISL_2101580, EPI_ISL_2101581, EPI_ISL_2101582, EPI_ISL_2101583, EPI_ISL_2101584, EPI_ISL_2101585, EPI_ISL_2101586, EPI_ISL_2101587, EPI_ISL_2101588, EPI_ISL_2101589, EPI_ISL_2101590, EPI_ISL_2101591, EPI_ISL_2101592, EPI_ISL_2101593, EPI_ISL_2101594, EPI_ISL_2101595, EPI_ISL_2101596, EPI_ISL_2101597, EPI_ISL_2101598, EPI_ISL_2101599, EPI_ISL_2101600, EPI_ISL_2101601, EPI_ISL_2101602, EPI_ISL_2101603, EPI_ISL_2101604, EPI_ISL_2101605, EPI_ISL_2101606, EPI_ISL_2101607, EPI_ISL_2101608, EPI_ISL_2101609, EPI_ISL_2101610, EPI_ISL_2101611 | | | |
| see above | Laboratorio Central Noel Nutels | Bioinformatics Laboratory / LNCC | Luiz G P de Almeida, Alessandra P Lamarca, Ronaldo da Silva F Jr, Liliane Cavalcante, Alexandra L Gerber, Ana Paula de C Guimaraes, Douglas Terra Machado, Cassia Alves, Diana Mariani, Cintia Policarpo, Gleidson da Silva de Oliveira, Mario Sergio Ribeiro, Silvia Carvalho, Flavio Dias da Silva, Marcio Henrique de Oliveira Garcia, Leandro Magalhaes de Souza, Cristiane Gomes da Silva, Caio Luiz Pereira Ribeiro, Andrea Cony Cavalcanti, Claudia Maria Braga de Mello, Amilcar Tanuri, Ana Tereza R Vasconcelos |
| EPI_ISL_2101612 | Unidade de apoio ao diagnóstico da COVID - UNADIG | Bioinformatics Laboratory / LNCC | Luiz G P de Almeida, Alessandra P Lamarca, Ronaldo da Silva F Jr, Liliane Cavalcante, Alexandra L Gerber, Ana Paula de C Guimaraes, Douglas Terra Machado, Cassia Alves, Diana Mariani, Cintia Policarpo, Gleidson da Silva de Oliveira, Mario Sergio Ribeiro, Silvia Carvalho, Flavio Dias da Silva, Marcio Henrique de Oliveira Garcia, Leandro Magalhaes de Souza, Cristiane Gomes da Silva, Caio Luiz Pereira Ribeiro, Andrea Cony Cavalcanti, Claudia Maria Braga de Mello, Amilcar Tanuri, Ana Tereza R Vasconcelos |
| EPI_ISL_2101613, EPI_ISL_2101615, EPI_ISL_2101616, EPI_ISL_2101617, EPI_ISL_2101618, EPI_ISL_2101619, EPI_ISL_2101620, EPI_ISL_2101621, EPI_ISL_2101622, EPI_ISL_2101623, EPI_ISL_2101624, EPI_ISL_2101625, EPI_ISL_2101626, EPI_ISL_2101627, EPI_ISL_2101628, EPI_ISL_2101629, EPI_ISL_2101630, EPI_ISL_2101631, EPI_ISL_2101632, EPI_ISL_2101633, EPI_ISL_2101634, EPI_ISL_2101635, EPI_ISL_2101636, EPI_ISL_2101637 | | | |
| see above | Laboratorio Central Noel Nutels | Bioinformatics Laboratory / LNCC | Luiz G P de Almeida, Alessandra P Lamarca, Ronaldo da Silva F Jr, Liliane Cavalcante, Alexandra L Gerber, Ana Paula de C Guimaraes, Douglas Terra Machado, Cassia Alves, Diana Mariani, Cintia Policarpo, Gleidson da Silva de Oliveira, Mario Sergio Ribeiro, Silvia Carvalho, Flavio Dias da Silva, Marcio Henrique de Oliveira Garcia, Leandro Magalhaes de Souza, Cristiane Gomes da Silva, Caio Luiz Pereira Ribeiro, Andrea Cony Cavalcanti, Claudia Maria Braga de Mello, Amilcar Tanuri, Ana Tereza R Vasconcelos |
| EPI_ISL_2101638, EPI_ISL_2101639, EPI_ISL_2101640, EPI_ISL_2101641, EPI_ISL_2101642, EPI_ISL_2101643, EPI_ISL_2101644, EPI_ISL_2101645, EPI_ISL_2101646, EPI_ISL_2101647, EPI_ISL_2101648, EPI_ISL_2101649, EPI_ISL_2101650, EPI_ISL_2101651, EPI_ISL_2101652, EPI_ISL_2101653, EPI_ISL_2101654, EPI_ISL_2101655, EPI_ISL_2101656, EPI_ISL_2101657, EPI_ISL_2101658, EPI_ISL_2101659, EPI_ISL_2101660, EPI_ISL_2101661, EPI_ISL_2101662, EPI_ISL_2101663, EPI_ISL_2101664, EPI_ISL_2101665, EPI_ISL_2101666, EPI_ISL_2101667, EPI_ISL_2101668, EPI_ISL_2101669, EPI_ISL_2101670, EPI_ISL_2101671 | | | |
| see above | Unidade de apoio ao diagnóstico da COVID - UNADIG | Bioinformatics Laboratory / LNCC | Luiz G P de Almeida, Alessandra P Lamarca, Ronaldo da Silva F Jr, Liliane Cavalcante, Alexandra L Gerber, Ana Paula de C Guimaraes, Douglas Terra Machado, Cassia Alves, Diana Mariani, Cintia Policarpo, Gleidson da Silva de Oliveira, Mario Sergio Ribeiro, Silvia Carvalho, Flavio Dias da Silva, Marcio Henrique de Oliveira Garcia, Leandro Magalhaes de Souza, Cristiane Gomes da Silva, Caio Luiz Pereira Ribeiro, Andrea Cony Cavalcanti, Claudia Maria Braga de Mello, Amilcar Tanuri, Ana Tereza R Vasconcelos |
| EPI_ISL_2101672, EPI_ISL_2101673, EPI_ISL_2101674, EPI_ISL_2101675 | Laboratorio Central Noel Nutels | Bioinformatics Laboratory / LNCC | Luiz G P de Almeida, Alessandra P Lamarca, Ronaldo da Silva F Jr, Liliane Cavalcante, Alexandra L Gerber, Ana Paula de C Guimaraes, Douglas Terra Machado, Cassia Alves, Diana Mariani, Cintia Policarpo, Gleidson da Silva de Oliveira, Mario Sergio Ribeiro, Silvia Carvalho, Flavio Dias da Silva, Marcio Henrique de Oliveira Garcia, Leandro Magalhaes de Souza, Cristiane Gomes da Silva, Caio Luiz Pereira Ribeiro, Andrea Cony Cavalcanti, Claudia Maria Braga de Mello, Amilcar Tanuri, Ana Tereza R Vasconcelos |
| EPI_ISL_2101676, EPI_ISL_2101677, EPI_ISL_2101678, EPI_ISL_2101679, EPI_ISL_2101680, EPI_ISL_2101681, EPI_ISL_2101682, EPI_ISL_2101683, EPI_ISL_2101684, EPI_ISL_2101685, EPI_ISL_2101686, EPI_ISL_2101687, EPI_ISL_2101688 | | | |
| see above | Unidade de apoio ao diagnóstico da COVID - UNADIG | Bioinformatics Laboratory / LNCC | Luiz G P de Almeida, Alessandra P Lamarca, Ronaldo da Silva F Jr, Liliane Cavalcante, Alexandra L Gerber, Ana Paula de C Guimaraes, Douglas Terra Machado, Cassia Alves, Diana Mariani, Cintia Policarpo, Gleidson da Silva de Oliveira, Mario Sergio Ribeiro, Silvia Carvalho, Flavio Dias da Silva, Marcio Henrique de Oliveira Garcia, Leandro Magalhaes de Souza, Cristiane Gomes da Silva, Caio Luiz Pereira Ribeiro, Andrea Cony Cavalcanti, Claudia Maria Braga de Mello, Amilcar Tanuri, Ana Tereza R Vasconcelos |
| EPI_ISL_2101689, EPI_ISL_2101690, EPI_ISL_2101691 | Laboratorio Central Noel Nutels | Bioinformatics Laboratory / LNCC | Luiz G P de Almeida, Alessandra P Lamarca, Ronaldo da Silva F Jr, Liliane Cavalcante, Alexandra L Gerber, Ana Paula de C Guimaraes, Douglas Terra Machado, Cassia Alves, Diana Mariani, Cintia Policarpo, Gleidson da Silva de Oliveira, Mario Sergio Ribeiro, Silvia Carvalho, Flavio Dias da Silva, Marcio Henrique de Oliveira Garcia, Leandro Magalhaes de Souza, Cristiane Gomes da Silva, Caio Luiz Pereira Ribeiro, Andrea Cony Cavalcanti, Claudia Maria Braga de Mello, Amilcar Tanuri, Ana Tereza R Vasconcelos |
| EPI_ISL_2101692, EPI_ISL_2101693, EPI_ISL_2101694, EPI_ISL_2101695, EPI_ISL_2101696 | Unidade de apoio ao diagnóstico da COVID - UNADIG | Bioinformatics Laboratory / LNCC | Luiz G P de Almeida, Alessandra P Lamarca, Ronaldo da Silva F Jr, Liliane Cavalcante, Alexandra L Gerber, Ana Paula de C Guimaraes, Douglas Terra Machado, Cassia Alves, Diana Mariani, Cintia Policarpo, Gleidson da Silva de Oliveira, Mario Sergio Ribeiro, Silvia Carvalho, Flavio Dias da Silva, Marcio Henrique de Oliveira Garcia, Leandro Magalhaes de Souza, Cristiane Gomes da Silva, Caio Luiz Pereira Ribeiro, Andrea Cony Cavalcanti, Claudia Maria Braga de Mello, Amilcar Tanuri, Ana Tereza R Vasconcelos |
| EPI_ISL_2101697, EPI_ISL_2101698, EPI_ISL_2101699, EPI_ISL_2101700, EPI_ISL_2101701, EPI_ISL_2101702, EPI_ISL_2101703, EPI_ISL_2101704, EPI_ISL_2101705, EPI_ISL_2101706, EPI_ISL_2101707, EPI_ISL_2101708, EPI_ISL_2101709, EPI_ISL_2101710, EPI_ISL_2101711, EPI_ISL_2101712, EPI_ISL_2101713 | | | |
| see above | Laboratorio Central Noel Nutels | Bioinformatics Laboratory / LNCC | Luiz G P de Almeida, Alessandra P Lamarca, Ronaldo da Silva F Jr, Liliane Cavalcante, Alexandra L Gerber, Ana Paula de C Guimaraes, Douglas Terra Machado, Cassia Alves, Diana Mariani, Cintia Policarpo, Gleidson da Silva de Oliveira, Mario Sergio Ribeiro, Silvia Carvalho, Flavio Dias da Silva, Marcio Henrique de Oliveira Garcia, Leandro Magalhaes de Souza, Cristiane Gomes da Silva, Caio Luiz Pereira Ribeiro, Andrea Cony Cavalcanti, Claudia Maria Braga de Mello, Amilcar Tanuri, Ana Tereza R Vasconcelos |
| EPI_ISL_2101714, EPI_ISL_2101715 | Unidade de apoio ao diagnóstico da COVID - UNADIG | Bioinformatics Laboratory / LNCC | Luiz G P de Almeida, Alessandra P Lamarca, Ronaldo da Silva F Jr, Liliane Cavalcante, Alexandra L Gerber, Ana Paula de C Guimaraes, Douglas Terra Machado, Cassia Alves, Diana Mariani, Cintia Policarpo, Gleidson da Silva de Oliveira, Mario Sergio Ribeiro, Silvia Carvalho, Flavio Dias da Silva, Marcio Henrique de Oliveira Garcia, Leandro Magalhaes de Souza, Cristiane Gomes da Silva, Caio Luiz Pereira Ribeiro, Andrea Cony Cavalcanti, Claudia Maria Braga de Mello, Amilcar Tanuri, Ana Tereza R Vasconcelos |
| EPI_ISL_2101716, EPI_ISL_2101717, EPI_ISL_2101718, EPI_ISL_2101719, EPI_ISL_2101720, EPI_ISL_2101721 | Laboratorio Central Noel Nutels | Bioinformatics Laboratory / LNCC | Luiz G P de Almeida, Alessandra P Lamarca, Ronaldo da Silva F Jr, Liliane Cavalcante, Alexandra L Gerber, Ana Paula de C Guimaraes, Douglas Terra Machado, Cassia Alves, Diana Mariani, Cintia Policarpo, Gleidson da Silva de Oliveira, Mario Sergio Ribeiro, Silvia Carvalho, Flavio Dias da Silva, Marcio Henrique de Oliveira Garcia, Leandro Magalhaes de Souza, Cristiane Gomes da Silva, Caio Luiz Pereira Ribeiro, Andrea Cony Cavalcanti, Claudia Maria Braga de Mello, Amilcar Tanuri, Ana Tereza R Vasconcelos |
| EPI_ISL_2101722 | Unidade de apoio ao diagnóstico da COVID - UNADIG | Bioinformatics Laboratory / LNCC | Luiz G P de Almeida, Alessandra P Lamarca, Ronaldo da Silva F Jr, Liliane Cavalcante, Alexandra L Gerber, Ana Paula de C Guimaraes, Douglas Terra Machado, Cassia Alves, Diana Mariani, Cintia Policarpo, Gleidson da Silva de Oliveira, Mario Sergio Ribeiro, Silvia Carvalho, Flavio Dias da Silva, Marcio Henrique de Oliveira Garcia, Leandro Magalhaes de Souza, Cristiane Gomes da Silva, Caio Luiz Pereira Ribeiro, Andrea Cony Cavalcanti, Claudia Maria Braga de Mello, Amilcar Tanuri, Ana Tereza R Vasconcelos |
| EPI_ISL_2101730 | Laboratorio Central Noel Nutels | Bioinformatics Laboratory / LNCC | Luiz G P de Almeida, Alessandra P Lamarca, Ronaldo da Silva F Jr, Liliane Cavalcante, Alexandra L Gerber, Ana Paula de C Guimaraes, Douglas Terra Machado, Cassia Alves, Diana Mariani, Cintia Policarpo, Gleidson da Silva de Oliveira, Mario Sergio Ribeiro, Silvia Carvalho, Flavio Dias da Silva, Marcio Henrique de Oliveira Garcia, Leandro Magalhaes de Souza, Cristiane Gomes da Silva, Caio Luiz Pereira Ribeiro, Andrea Cony Cavalcanti, Claudia Maria Braga de Mello, Amilcar Tanuri, Ana Tereza R Vasconcelos |
| EPI_ISL_2101731 | Unidade de apoio ao diagnóstico da COVID - UNADIG | Bioinformatics Laboratory / LNCC | Luiz G P de Almeida, Alessandra P Lamarca, Ronaldo da Silva F Jr, Liliane Cavalcante, Alexandra L Gerber, Ana Paula de C Guimaraes, Douglas Terra Machado, Cassia Alves, Diana Mariani, Cintia Policarpo, Gleidson da Silva de Oliveira, Mario Sergio Ribeiro, Silvia Carvalho, Flavio Dias da Silva, Marcio Henrique de Oliveira Garcia, Leandro Magalhaes de Souza, Cristiane Gomes da Silva, Caio Luiz Pereira Ribeiro, Andrea Cony Cavalcanti, Claudia Maria Braga de Mello, Amilcar Tanuri, Ana Tereza R Vasconcelos |
| EPI_ISL_2101732, EPI_ISL_2101733 | Laboratorio Central Noel Nutels | Bioinformatics Laboratory / LNCC | Luiz G P de Almeida, Alessandra P Lamarca, Ronaldo da Silva F Jr, Liliane Cavalcante, Alexandra L Gerber, Ana Paula de C Guimaraes, Douglas Terra |

| | | | |
|---|---|---|---|
| | | | Machado, Cassia Alves, Diana Mariani, Cintia Policarpo, Gleidson da Silva de Oliveira, Mario Sergio Ribeiro, Silvia Carvalho, Flavio Dias da Silva, Marcio Henrique de Oliveira Garcia, Leandro Magalhaes de Souza, Cristiane Gomes da Silva, Caio Luiz Pereira Ribeiro, Andrea Cony Cavalcanti, Claudia Maria Braga de Mello, Amilcar Tanuri, Ana Tereza R Vasconcelos |
| EPI_ISL_2101734 | Unidade de apoio ao diagnóstico da COVID - UNADIG | Bioinformatics Laboratory / LNCC | Luiz G P de Almeida, Alessandra P Lamarca, Ronaldo da Silva F Jr, Liliane Cavalcante, Alexandra L Gerber, Ana Paula de C Guimaraes, Douglas Terra Machado, Cassia Alves, Diana Mariani, Cintia Policarpo, Gleidson da Silva de Oliveira, Mario Sergio Ribeiro, Silvia Carvalho, Flavio Dias da Silva, Marcio Henrique de Oliveira Garcia, Leandro Magalhaes de Souza, Cristiane Gomes da Silva, Caio Luiz Pereira Ribeiro, Andrea Cony Cavalcanti, Claudia Maria Braga de Mello, Amilcar Tanuri, Ana Tereza R Vasconcelos |
| EPI_ISL_2101736, EPI_ISL_2101737, EPI_ISL_2101738, EPI_ISL_2101739 | Laboratorio Central Noel Nutels | Bioinformatics Laboratory / LNCC | Luiz G P de Almeida, Alessandra P Lamarca, Ronaldo da Silva F Jr, Liliane Cavalcante, Alexandra L Gerber, Ana Paula de C Guimaraes, Douglas Terra Machado, Cassia Alves, Diana Mariani, Cintia Policarpo, Gleidson da Silva de Oliveira, Mario Sergio Ribeiro, Silvia Carvalho, Flavio Dias da Silva, Marcio Henrique de Oliveira Garcia, Leandro Magalhaes de Souza, Cristiane Gomes da Silva, Caio Luiz Pereira Ribeiro, Andrea Cony Cavalcanti, Claudia Maria Braga de Mello, Amilcar Tanuri, Ana Tereza R Vasconcelos |
| EPI_ISL_2101740, EPI_ISL_2101741 | Unidade de apoio ao diagnóstico da COVID - UNADIG | Bioinformatics Laboratory / LNCC | Luiz G P de Almeida, Alessandra P Lamarca, Ronaldo da Silva F Jr, Liliane Cavalcante, Alexandra L Gerber, Ana Paula de C Guimaraes, Douglas Terra Machado, Cassia Alves, Diana Mariani, Cintia Policarpo, Gleidson da Silva de Oliveira, Mario Sergio Ribeiro, Silvia Carvalho, Flavio Dias da Silva, Marcio Henrique de Oliveira Garcia, Leandro Magalhaes de Souza, Cristiane Gomes da Silva, Caio Luiz Pereira Ribeiro, Andrea Cony Cavalcanti, Claudia Maria Braga de Mello, Amilcar Tanuri, Ana Tereza R Vasconcelos |
| EPI_ISL_2101742, EPI_ISL_2101743 | Laboratorio Central Noel Nutels | Bioinformatics Laboratory / LNCC | Luiz G P de Almeida, Alessandra P Lamarca, Ronaldo da Silva F Jr, Liliane Cavalcante, Alexandra L Gerber, Ana Paula de C Guimaraes, Douglas Terra Machado, Cassia Alves, Diana Mariani, Cintia Policarpo, Gleidson da Silva de Oliveira, Mario Sergio Ribeiro, Silvia Carvalho, Flavio Dias da Silva, Marcio Henrique de Oliveira Garcia, Leandro Magalhaes de Souza, Cristiane Gomes da Silva, Caio Luiz Pereira Ribeiro, Andrea Cony Cavalcanti, Claudia Maria Braga de Mello, Amilcar Tanuri, Ana Tereza R Vasconcelos |
| EPI_ISL_2101744 | Unidade de apoio ao diagnóstico da COVID - UNADIG | Bioinformatics Laboratory / LNCC | Luiz G P de Almeida, Alessandra P Lamarca, Ronaldo da Silva F Jr, Liliane Cavalcante, Alexandra L Gerber, Ana Paula de C Guimaraes, Douglas Terra Machado, Cassia Alves, Diana Mariani, Cintia Policarpo, Gleidson da Silva de Oliveira, Mario Sergio Ribeiro, Silvia Carvalho, Flavio Dias da Silva, Marcio Henrique de Oliveira Garcia, Leandro Magalhaes de Souza, Cristiane Gomes da Silva, Caio Luiz Pereira Ribeiro, Andrea Cony Cavalcanti, Claudia Maria Braga de Mello, Amilcar Tanuri, Ana Tereza R Vasconcelos |
| EPI_ISL_2101745, EPI_ISL_2101746, EPI_ISL_2101747 | Laboratorio Central Noel Nutels | Bioinformatics Laboratory / LNCC | Luiz G P de Almeida, Alessandra P Lamarca, Ronaldo da Silva F Jr, Liliane Cavalcante, Alexandra L Gerber, Ana Paula de C Guimaraes, Douglas Terra Machado, Cassia Alves, Diana Mariani, Cintia Policarpo, Gleidson da Silva de Oliveira, Mario Sergio Ribeiro, Silvia Carvalho, Flavio Dias da Silva, Marcio Henrique de Oliveira Garcia, Leandro Magalhaes de Souza, Cristiane Gomes da Silva, Caio Luiz Pereira Ribeiro, Andrea Cony Cavalcanti, Claudia Maria Braga de Mello, Amilcar Tanuri, Ana Tereza R Vasconcelos |
| EPI_ISL_2105574, EPI_ISL_2135717, EPI_ISL_2135718, EPI_ISL_2135719, EPI_ISL_2135720, EPI_ISL_2136060, EPI_ISL_2136097, EPI_ISL_2136133 | Servicio Virosis Respiratorias-Departamento Virología-INEI | Instituto Nacional Enfermedades Infecciosas C.G.Malbran | Baumeister E., Avaro M., Benedetti E., Russo M., Dattero ME, Pontoriero A., Cisterna D., Molina V., Perandones C., Tuduri E., Lorenzo F., Poklepovich T., Campos J. |
| EPI_ISL_2139495, EPI_ISL_2139508, EPI_ISL_2139513, EPI_ISL_2139515, EPI_ISL_2139519, EPI_ISL_2139523, EPI_ISL_2139526, EPI_ISL_2139527, EPI_ISL_2139531, EPI_ISL_2139536 | Laboratorio Exame | Universidade Federal de Ciencias da Saude de Porto Alegre | Vinícius Bonetti Franceschi, Gabriel Dickin Caldana et al. |
| EPI_ISL_2140039, EPI_ISL_2140112 | Servicio Virosis Respiratorias-Departamento Virología-INEI | Instituto Nacional Enfermedades Infecciosas C.G.Malbran | Baumeister E., Avaro M., Benedetti E., Russo M., Dattero ME, Pontoriero A., Cisterna D., Molina V., Perandones C., Tuduri E., Lorenzo F., Poklepovich T., Campos J. |
| EPI_ISL_2157382 | Laboratorio Central de Saude Publica do Estado de Sergipe (LACEN/SE) | Laboratory of Respiratory Viruses and Measles, Oswaldo Cruz Institute, FIOCRUZ | Paola Resende, Luciana Appolinario, Fernando Motta, Anna Carolina Paixao, Ana Carolina Mendonca, Alice Sampaio Rocha, Tainá Moreira Martins Venas, Elisa Cavalcante Pereira, Renata Serrano Lopes, Cliomar Alves dos Santos, Marilda Siqueira on behalf of the Fiocruz COVID-19 Genomic Surveillance Network |
| EPI_ISL_2157396, EPI_ISL_2157399 | Laboratório Central de Saude Publica do Estado de Santa Catarina (LACEN/SC) | Laboratory of Respiratory Viruses and Measles, Oswaldo Cruz Institute, FIOCRUZ | Paola Resende, Luciana Appolinario, Fernando Motta, Anna Carolina Paixao, Ana Carolina Mendonca, Alice Sampaio Rocha, Taina Venas, Elisa Cavalcante Pereira, Renata Serrano Lopes, Darcita Buerger Rovaris, Sandra Bianchini Fernandes, Marilda Siqueira on behalf of the Fiocruz COVID-19 Genomic Surveillance Network |
| EPI_ISL_2157404 | Lboratorio Central de Saude Publica do Estado do Parana (LACEN/PR) | Laboratory of Respiratory Viruses and Measles, Oswaldo Cruz Institute, FIOCRUZ | Paola Resende, Luciana Appolinario, Fernando Motta, Anna Carolina Paixao, Ana Carolina Mendonca, Alice Sampaio Rocha, Taina Venas, Elisa Cavalcante Pereira, Renata Serrano Lopes, Irina Riediger, Marilda Siqueira on behalf of the Fiocruz COVID-19 Genomic Surveillance Network |
| EPI_ISL_2157407, EPI_ISL_2157426 | Laboratório Central de Saude Publica do Estado de Santa Catarina (LACEN/SC) | Laboratory of Respiratory Viruses and Measles, Oswaldo Cruz Institute, FIOCRUZ | Paola Resende, Luciana Appolinario, Fernando Motta, Anna Carolina Paixao, Ana Carolina Mendonca, Alice Sampaio Rocha, Taina Venas, Elisa Cavalcante Pereira, Renata Serrano Lopes, Darcita Buerger Rovaris, Sandra Bianchini Fernandes, Marilda Siqueira on behalf of the Fiocruz COVID-19 Genomic Surveillance Network |
| EPI_ISL_2157428, EPI_ISL_2157429, EPI_ISL_2157435, EPI_ISL_2157442, EPI_ISL_2157444 | Laboratorio Central de Saude Publica do Estado de Sergipe (LACEN/SE) | Laboratory of Respiratory Viruses and Measles, Oswaldo Cruz Institute, FIOCRUZ | Paola Resende, Luciana Appolinario, Fernando Motta, Anna Carolina Paixao, Ana Carolina Mendonca, Alice Sampaio Rocha, Tainá Moreira Martins Venas, Elisa Cavalcante Pereira, Renata Serrano Lopes, Cliomar Alves dos Santos, Marilda Siqueira on behalf of the Fiocruz COVID-19 Genomic Surveillance Network |
| EPI_ISL_2157447 | Laboratório Central de Saude Publica do Estado de Santa Catarina (LACEN/SC) | Laboratory of Respiratory Viruses and Measles, Oswaldo Cruz Institute, FIOCRUZ | Paola Resende, Luciana Appolinario, Fernando Motta, Anna Carolina Paixao, Ana Carolina Mendonca, Alice Sampaio Rocha, Taina Venas, Elisa Cavalcante Pereira, Renata Serrano Lopes, Darcita Buerger Rovaris, Sandra Bianchini Fernandes, Marilda Siqueira on behalf of the Fiocruz COVID-19 Genomic Surveillance Network |
| EPI_ISL_2157449, EPI_ISL_2157453, EPI_ISL_2157471, EPI_ISL_2157473, EPI_ISL_2157474, EPI_ISL_2157478 | Laboratorio Central de Saude Publica do Estado de Sergipe (LACEN/SE) | Laboratory of Respiratory Viruses and Measles, Oswaldo Cruz Institute, FIOCRUZ | Paola Resende, Luciana Appolinario, Fernando Motta, Anna Carolina Paixao, Ana Carolina Mendonca, Alice Sampaio Rocha, Tainá Moreira Martins Venas, Elisa Cavalcante Pereira, Renata Serrano Lopes, Cliomar Alves dos Santos, Marilda Siqueira on behalf of the Fiocruz COVID-19 Genomic Surveillance Network |
| EPI_ISL_2157487 | Lboratorio Central de Saude Publica do Estado do Parana (LACEN/PR) | Laboratory of Respiratory Viruses and Measles, Oswaldo Cruz Institute, FIOCRUZ | Paola Resende, Luciana Appolinario, Fernando Motta, Anna Carolina Paixao, Ana Carolina Mendonca, Alice Sampaio Rocha, Taina Venas, Elisa Cavalcante Pereira, Renata Serrano Lopes, Irina Riediger, Marilda Siqueira on behalf of the Fiocruz COVID-19 Genomic Surveillance Network |
| EPI_ISL_2157497, EPI_ISL_2157518 | Laboratório Central de Saude Publica do Estado de Santa Catarina (LACEN/SC) | Laboratory of Respiratory Viruses and Measles, Oswaldo Cruz Institute, FIOCRUZ | Paola Resende, Luciana Appolinario, Fernando Motta, Anna Carolina Paixao, Ana Carolina Mendonca, Alice Sampaio Rocha, Taina Venas, Elisa Cavalcante Pereira, Renata Serrano Lopes, Darcita Buerger Rovaris, Sandra Bianchini Fernandes, Marilda Siqueira on behalf of the Fiocruz COVID-19 Genomic Surveillance Network |
| EPI_ISL_2157524, EPI_ISL_2157525, EPI_ISL_2157526, EPI_ISL_2157528, EPI_ISL_2157529, EPI_ISL_2157530, EPI_ISL_2157536, EPI_ISL_2157537, EPI_ISL_2157538, EPI_ISL_2157539 | Laboratório Central de Saude Publica do Estado do Rio de Janeiro (LACEN/RJ) | Laboratory of Respiratory Viruses and Measles, Oswaldo Cruz Institute, FIOCRUZ | Paola Resende, Luciana Appolinario, Fernando Motta, Anna Carolina Paixao, Ana Carolina Mendonca, Alice Sampaio Rocha, Taina Venas, Elisa Cavalcante Pereira, Renata Serrano Lopes, Andrea Cony Cavalcanti, Marilda Siqueira on behalf of the Fiocruz COVID-19 Genomic Surveillance Network |
| EPI_ISL_2157540, EPI_ISL_2157545 | Laboratory of Respiratory Viruses and Measles, Oswaldo Cruz Institute, FIOCRUZ | Laboratory of Respiratory Viruses and Measles, Oswaldo Cruz Institute, FIOCRUZ | Paola Resende, Luciana Appolinario, Fernando Motta, Anna Carolina Paixao, Ana Carolina Mendonca, Alice Sampaio Rocha, Taina Venas, Elisa Cavalcante Pereira, Renata Serrano Lopes, Andrea Cony Cavalcanti, Marilda Siqueira on behalf of the Fiocruz COVID-19 Genomic Surveillance Network |
| EPI_ISL_2157549 | Laboratório Central de Saude Publica do Estado do Rio de Janeiro (LACEN/RJ) | Laboratory of Respiratory Viruses and Measles, Oswaldo Cruz Institute, FIOCRUZ | Paola Resende, Luciana Appolinario, Fernando Motta, Anna Carolina Paixao, Ana Carolina Mendonca, Alice Sampaio Rocha, Taina Venas, Elisa Cavalcante Pereira, Renata Serrano Lopes, Andrea Cony Cavalcanti, Marilda Siqueira on behalf of the Fiocruz COVID-19 Genomic Surveillance Network |
| EPI_ISL_2157561 | Laboratorio Central de Saude Publica do Estado do Rio de Janeiro (LACEN/RJ) | Laboratory of Respiratory Viruses and Measles, Oswaldo Cruz Institute, FIOCRUZ | Paola Resende, Luciana Appolinario, Fernando Motta, Anna Carolina Paixao, Ana Carolina Mendonca, Alice Sampaio Rocha, Taina Venas, Elisa Cavalcante Pereira, Renata Serrano Lopes, Andrea Cony Cavalcanti, Marilda Siqueira on behalf of the Fiocruz COVID-19 Genomic Surveillance Network |
| EPI_ISL_2157573, EPI_ISL_2157575 | Laboratory of Respiratory Viruses and Measles, Oswaldo Cruz Institute, FIOCRUZ | Laboratory of Respiratory Viruses and Measles, Oswaldo Cruz Institute, FIOCRUZ | Paola Resende, Luciana Appolinario, Fernando Motta, Anna Carolina Paixao, Ana Carolina Mendonca, Alice Sampaio Rocha, Taina Venas, Elisa Cavalcante Pereira, Renata Serrano Lopes, Andrea Cony Cavalcanti, Marilda Siqueira on behalf of the Fiocruz COVID-19 Genomic Surveillance Network |

| | | | |
|---|---|---|---|
| EPI_ISL_2157599 | Laboratório Central de Saude Publica do Estado do Rio de Janeiro (LACEN/RJ) | Laboratory of Respiratory Viruses and Measles, Oswaldo Cruz Institute, FIOCRUZ | Paola Resende, Luciana Appolinario, Fernando Motta, Anna Carolina Paixao, Ana Carolina Mendonca, Alice Sampaio Rocha, Taina Venas, Elisa Cavalcante Pereira, Renata Serrano Lopes, Andrea Cony Cavalcanti, Marilda Siqueira on behalf of the Fiocruz COVID-19 Genomic Surveillance Network |
| EPI_ISL_2158697, EPI_ISL_2158710, EPI_ISL_2158730, EPI_ISL_2158737, EPI_ISL_2158740, EPI_ISL_2158750, EPI_ISL_2158824 | Servicio Virosis Respiratorias-Departamento Virología-INEI | Instituto Nacional Enfermedades Infecciosas C.G.Malbran | Baumeister E., Avaro M., Benedetti E., Russo M., Dattero ME, Pontoriero A., Cisterna D., Molina V., Perandones C., Tuduri E., Lorenzo F., Poklepovich T., Campos J. |
| EPI_ISL_2187879, EPI_ISL_2187911, EPI_ISL_2187952, EPI_ISL_2187964, EPI_ISL_2188002, EPI_ISL_2188008 | HLAGYN - Laboratorio de Imunologia de Transplantes de Goias | HLAGYN - Laboratorio de Imunologia de Transplantes de Goias | Fernando Antonio Vinhal dos Santos, Erika Lopes Rocha Batista, Alessandro Leonardo Alvares Magalhaes, Frederico Rodrigues Vinhal, Sabrina Sara Moreira Duarte, Lucas Carlos Gomes Pereira, Daniel Ferreira de Sousa |
| EPI_ISL_2196185, EPI_ISL_2196186, EPI_ISL_2196187 | Laboratory of Respiratory Viruses and Measles, Oswaldo Cruz Institute, FIOCRUZ | Laboratory of Respiratory Viruses and Measles, Oswaldo Cruz Institute, FIOCRUZ | Paola Resende, Luciana Appolinario, Fernando Motta, Anna Carolina Paixao, Ana Carolina Mendonca, Alice Sampaio Rocha, Taina Venas, Elisa Cavalcante Pereira, Renata Serrano Lopes, Marilda Siqueira on behalf of the Fiocruz COVID-19 Genomic Surveillance Network |
| EPI_ISL_2196189 | Laboratorio Central de Saude Publica do Estado do Rio de Janeiro (LACEN-RJ) | Laboratory of Respiratory Viruses and Measles, Oswaldo Cruz Institute, FIOCRUZ | Paola Resende, Luciana Appolinario, Fernando Motta, Anna Carolina Paixao, Ana Carolina Mendonca, Alice Sampaio Rocha, Taina Venas, Elisa Cavalcante Pereira, Renata Serrano Lopes, Andrea Cony Cavalcanti, Marilda Siqueira on behalf of the Fiocruz COVID-19 Genomic Surveillance Network |
| EPI_ISL_2196190, EPI_ISL_2196191, EPI_ISL_2196192, EPI_ISL_2196193, EPI_ISL_2196194, EPI_ISL_2196195, EPI_ISL_2196196 | Laboratory of Respiratory Viruses and Measles, Oswaldo Cruz Institute, FIOCRUZ | Laboratory of Respiratory Viruses and Measles, Oswaldo Cruz Institute, FIOCRUZ | Paola Resende, Luciana Appolinario, Fernando Motta, Anna Carolina Paixao, Ana Carolina Mendonca, Alice Sampaio Rocha, Taina Venas, Elisa Cavalcante Pereira, Renata Serrano Lopes, Marilda Siqueira on behalf of the Fiocruz COVID-19 Genomic Surveillance Network |
| EPI_ISL_2196197, EPI_ISL_2196198, EPI_ISL_2196199, EPI_ISL_2196200, EPI_ISL_2196201, EPI_ISL_2196202, EPI_ISL_2196203, EPI_ISL_2196204, EPI_ISL_2196205, EPI_ISL_2196206, EPI_ISL_2196207, EPI_ISL_2196208, EPI_ISL_2196209 | | | |
| see above | Laboratorio Central de Saude Publica do Estado do Rio de Janeiro (LACEN-RJ) | Laboratory of Respiratory Viruses and Measles, Oswaldo Cruz Institute, FIOCRUZ | Paola Resende, Luciana Appolinario, Fernando Motta, Anna Carolina Paixao, Ana Carolina Mendonca, Alice Sampaio Rocha, Taina Venas, Elisa Cavalcante Pereira, Renata Serrano Lopes, Andrea Cony Cavalcanti, Marilda Siqueira on behalf of the Fiocruz COVID-19 Genomic Surveillance Network |
| EPI_ISL_2196211, EPI_ISL_2196213, EPI_ISL_2196215, EPI_ISL_2196220 | Laboratorio Central de Saude Publica do Estado de Sergipe (LACEN/SE) | Laboratory of Respiratory Viruses and Measles, Oswaldo Cruz Institute, FIOCRUZ | Paola Resende, Luciana Appolinario, Fernando Motta, Anna Carolina Paixao, Ana Carolina Mendonca, Alice Sampaio Rocha, Tainá Moreira Martins Venas, Elisa Cavalcante Pereira, Renata Serrano Lopes, Cliomar Alves dos Santos, Marilda Siqueira on behalf of the Fiocruz COVID-19 Genomic Surveillance Network |
| EPI_ISL_2196236 | Laboratorio Central de Saude Publica do Estado do Rio de Janeiro (LACEN-RJ) | Laboratory of Respiratory Viruses and Measles, Oswaldo Cruz Institute, FIOCRUZ | Paola Resende, Luciana Appolinario, Fernando Motta, Anna Carolina Paixao, Ana Carolina Mendonca, Alice Sampaio Rocha, Taina Venas, Elisa Cavalcante Pereira, Renata Serrano Lopes, Andrea Cony Cavalcanti, Marilda Siqueira on behalf of the Fiocruz COVID-19 Genomic Surveillance Network |
| EPI_ISL_2196242, EPI_ISL_2196245 | Labortorio Central de Saude Publica do Estado do Rio de Janeiro (LACEN/RJ) | Laboratory of Respiratory Viruses and Measles, Oswaldo Cruz Institute, FIOCRUZ | Paola Resende, Luciana Appolinario, Fernando Motta, Anna Carolina Paixao, Ana Carolina Mendonca, Alice Sampaio Rocha, Taina Venas, Elisa Cavalcante Pereira, Renata Serrano Lopes, Andrea Cony Cavalcanti, Marilda Siqueira on behalf of the Fiocruz COVID-19 Genomic Surveillance Network |
| EPI_ISL_2196251 | Laboratory of Respiratory Viruses and Measles, Oswaldo Cruz Institute, FIOCRUZ | Laboratory of Respiratory Viruses and Measles, Oswaldo Cruz Institute, FIOCRUZ | Paola Resende, Luciana Appolinario, Fernando Motta, Anna Carolina Paixao, Ana Carolina Mendonca, Alice Sampaio Rocha, Taina Venas, Elisa Cavalcante Pereira, Renata Serrano Lopes, Marilda Siqueira on behalf of the Fiocruz COVID-19 Genomic Surveillance Network |
| EPI_ISL_2196267 | Laboratorio Central de Saude Publica do Estado Maranhao (LACEN-MA) | Laboratory of Respiratory Viruses and Measles, Oswaldo Cruz Institute, FIOCRUZ | Paola Resende, Luciana Appolinario, Fernando Motta, Anna Carolina Paixao, Ana Carolina Mendonca, Alice Sampaio Rocha, Taina Venas, Elisa Cavalcante Pereira, Renata Serrano Lopes, Lidio Gonçalves Lima Neto, Marilda Siqueira on behalf of the Fiocruz COVID-19 Genomic Surveillance Network |
| EPI_ISL_2196313, EPI_ISL_2196314, EPI_ISL_2196315, EPI_ISL_2196316, EPI_ISL_2196317, EPI_ISL_2196318, EPI_ISL_2196319, EPI_ISL_2196320, EPI_ISL_2196321, EPI_ISL_2196322, EPI_ISL_2196323, EPI_ISL_2196324, EPI_ISL_2196325, EPI_ISL_2196326, EPI_ISL_2196327, EPI_ISL_2196328, EPI_ISL_2196329, EPI_ISL_2196330, EPI_ISL_2196331, EPI_ISL_2196332, EPI_ISL_2196333, EPI_ISL_2196334, EPI_ISL_2196356 | | | |
| see above | Labortorio Central de Saude Publica do Estado do Rio de Janeiro (LACEN/RJ) | Laboratory of Respiratory Viruses and Measles, Oswaldo Cruz Institute, FIOCRUZ | Paola Resende, Luciana Appolinario, Fernando Motta, Anna Carolina Paixao, Ana Carolina Mendonca, Alice Sampaio Rocha, Taina Venas, Elisa Cavalcante Pereira, Renata Serrano Lopes, Andrea Cony Cavalcanti, Marilda Siqueira on behalf of the Fiocruz COVID-19 Genomic Surveillance Network |
| EPI_ISL_2196361 | Laboratory of Respiratory Viruses and Measles, Oswaldo Cruz Institute, FIOCRUZ | Laboratory of Respiratory Viruses and Measles, Oswaldo Cruz Institute, FIOCRUZ | Paola Resende, Luciana Appolinario, Fernando Motta, Anna Carolina Paixao, Ana Carolina Mendonca, Alice Sampaio Rocha, Taina Venas, Elisa Cavalcante Pereira, Renata Serrano Lopes, Marilda Siqueira on behalf of the Fiocruz COVID-19 Genomic Surveillance Network |
| EPI_ISL_2209955 | SECRETARIA DE SAUDE DE SAO PEDRO | Instituto Butantan / FZEA-USP-Pirassununga | Dimas Tadeu Covas, Antonio Jorge Martins, Claudia Renata dos Santos Barros, David Schlesinger, Debora Botequio Moretti, Elaine Cristina Marqueze, Elaine Vieira Santos, Evandra Strazza Rodrigues, Heidge Fukumasu, Jayme Augusto de Souza-Neto, José Salvatore Leister Patané, Luiz Alcantara, Luiz Lehmann Coutinho, Maria Carolina Elias, Maurício Lacerda Nogueira, Rafael dos Santos Bezerra, Raul Machado Neto, Rejane Maria Tommasini Grotto, Ricardo Haddad, Sandra Coccuzzo Sampaio Vessoni, Simone Kashima, Svetoslav Nanev Slavov, Vincent Louis Viala |
| EPI_ISL_2225349 | Houston Methodist Hospital | Houston Methodist Hospital | Randall J. Olsen, Paul A. Christensen, S. Wesley Long, Sishir Subedi, Robert Olson, Marcus Nguyen, James J. Davis, Matthew Ojeda Saavedra, Prasanti Yerramilli, Layne Pruitt, Kristina Reppond, Madison N. Shyer, Jessica Cambric, Ryan Gadd, Ilya J. Finkelstein, Jimmy Gollihar, and James M. Musser |
| EPI_ISL_2241579 | Laboratório Central de Saúde Pública de Sergipe | Coordenação Geral de Laboratórios de Saúde Pública (CGLAB/DAEVS/SVS/MS) | Vagner Fonseca, et al. |
| EPI_ISL_2241581 | Laboratório Central de Saúde Pública da Paraiba | Coordenação Geral de Laboratórios de Saúde Pública (CGLAB/DAEVS/SVS/MS) | Vagner Fonseca, et al. |
| EPI_ISL_2241589 | Laboratório Central de Saúde Pública de Sergipe | Coordenação Geral de Laboratórios de Saúde Pública (CGLAB/DAEVS/SVS/MS) | Vagner Fonseca, et al. |
| EPI_ISL_2249357, EPI_ISL_2249361, EPI_ISL_2249365 | Laboratório Central de Saúde Pública do Rio Grande do Sul | Coordenação Geral de Laboratórios de Saúde Pública (CGLAB/DAEVS/SVS/MS) | Vagner Fonseca, et al. |
| EPI_ISL_2249404, EPI_ISL_2249405, EPI_ISL_2249406, EPI_ISL_2249407, EPI_ISL_2249408, EPI_ISL_2249409, EPI_ISL_2249410, EPI_ISL_2249411, EPI_ISL_2249412, EPI_ISL_2249413, EPI_ISL_2249414, EPI_ISL_2249415, EPI_ISL_2249416, EPI_ISL_2249417, EPI_ISL_2249418, EPI_ISL_2249419, EPI_ISL_2249420, EPI_ISL_2249421, EPI_ISL_2249422, EPI_ISL_2249423, EPI_ISL_2249424, EPI_ISL_2249425, EPI_ISL_2249426, EPI_ISL_2249427, EPI_ISL_2249428, EPI_ISL_2249429, EPI_ISL_2249430, EPI_ISL_2249431, EPI_ISL_2249432, EPI_ISL_2249433, EPI_ISL_2249434, EPI_ISL_2249440, EPI_ISL_2249441, EPI_ISL_2249442, EPI_ISL_2249443, EPI_ISL_2249444, EPI_ISL_2249445 | | | |
| see above | Laboratório Central de Saúde Pública do Rio de Janeiro | Coordenação Geral de Laboratórios de Saúde Pública (CGLAB/DAEVS/SVS/MS) | Vagner Fonseca, et al. |
| EPI_ISL_2269371 | Fulgent Genetics | Centers for Disease Control and Prevention Division of Viral Diseases, Pathogen Discovery | Dakota Howard, Dhwani Batra, Peter W. Cook, Kara Moser, Adrian Paskey, Jason Caravas, Benjamin Rambo-Martin, Shatavia Morrison, Christopher Gulvick, Scott Sammons, Yvette Unoarumhi, Darlene Wagner, Matthew Schmerer, Harry Gao, Mickey Li, John Gao, Joseph Fierro, Benafsh Sapra, Becky Tsai, Yan Meng, Doreen Ng, James Xie, Clinton R. Paden, Duncan MacCannell |
| EPI_ISL_2274080 | Laboratorio Central de Saude Publica do Estado Maranhao (LACEN-MA) | Laboratory of Respiratory Viruses and Measles, Oswaldo Cruz Institute, FIOCRUZ | Paola Resende, Luciana Appolinario, Fernando Motta, Anna Carolina Paixao, Ana Carolina Mendonca, Alice Sampaio Rocha, Taina Venas, Elisa Cavalcante Pereira, Renata Serrano Lopes, Lidio Gonçalves Lima Neto, Marilda Siqueira on behalf of the Fiocruz COVID-19 Genomic Surveillance Network |
| EPI_ISL_2274096 | Lboratorio Central de Saude Publica do Estado do Parana (LACEN/PR) | Laboratory of Respiratory Viruses and Measles, Oswaldo Cruz Institute, FIOCRUZ | Paola Resende, Luciana Appolinario, Fernando Motta, Anna Carolina Paixao, Ana Carolina Mendonca, Alice Sampaio Rocha, Taina Venas, Elisa Cavalcante Pereira, Renata Serrano Lopes, Irina Riediger, Marilda Siqueira on behalf of the Fiocruz COVID-19 Genomic Surveillance Network |
| EPI_ISL_2274097 | Laboratory of Respiratory Viruses and Measles, Oswaldo Cruz Institute, FIOCRUZ | Laboratory of Respiratory Viruses and Measles, Oswaldo Cruz Institute, FIOCRUZ | Paola Resende, Luciana Appolinario, Fernando Motta, Anna Carolina Paixao, Ana Carolina Mendonca, Alice Sampaio Rocha, Taina Venas, Elisa Cavalcante Pereira, Renata Serrano Lopes, Marilda Siqueira on behalf of the Fiocruz COVID-19 Genomic Surveillance Network |
| EPI_ISL_2274098, EPI_ISL_2274099 | Labortorio Central de Saude Publica do Estado do Rio de Janeiro (LACEN/RJ) | Laboratory of Respiratory Viruses and Measles, Oswaldo Cruz Institute, FIOCRUZ | Paola Resende, Luciana Appolinario, Fernando Motta, Anna Carolina Paixao, Ana Carolina Mendonca, Alice Sampaio Rocha, Taina Venas, Elisa Cavalcante Pereira, Renata Serrano Lopes, Andrea Cony Cavalcanti, Marilda Siqueira on behalf of the Fiocruz COVID-19 Genomic Surveillance Network |
| EPI_ISL_2274100, EPI_ISL_2274101, EPI_ISL_2274102, EPI_ISL_2274103, EPI_ISL_2274104, EPI_ISL_2274105, EPI_ISL_2274106, EPI_ISL_2274107 | Laboratory of Respiratory Viruses and Measles, Oswaldo Cruz Institute, FIOCRUZ | Laboratory of Respiratory Viruses and Measles, Oswaldo Cruz Institute, FIOCRUZ | Paola Resende, Luciana Appolinario, Fernando Motta, Anna Carolina Paixao, Ana Carolina Mendonca, Alice Sampaio Rocha, Taina Venas, Elisa Cavalcante Pereira, Renata Serrano Lopes, Marilda Siqueira on behalf of the Fiocruz COVID-19 Genomic Surveillance Network |

| | | | |
|---|---|---|---|
| EPI_ISL_2274108, EPI_ISL_2274109 | Laboratorio Central de Saude Publica do Estado do Rio Grande do Sul (LACEN-RS) | Laboratory of Respiratory Viruses and Measles, Oswaldo Cruz Institute, FIOCRUZ | Paola Resende, Luciana Appolinario, Fernando Motta, Anna Carolina Paixao, Ana Carolina Mendonca, Alice Sampaio Rocha, Taina Venas, Elisa Cavalcante Pereira, Renata Serrano Lopes, Tatiana Schaffer Gregianini, Richard Salvato, Marilda Siqueira on behalf of the Fiocruz COVID-19 Genomic Surveillance Network |
| EPI_ISL_2308456, EPI_ISL_2308471, EPI_ISL_2308473, EPI_ISL_2308475, EPI_ISL_2308478 | Laboratório Central de Saúde Pública de Sergipe | Coordenação Geral de Laboratórios de Saúde Pública (CGLAB/DAEVS/SVS/MS) | Vagner Fonseca, et al. |
| EPI_ISL_2344442 | Instituto Butantan | Instituto de Medicina Tropical de Sao Paulo | Brazil-UK Centre for Arbovirus Discovery Diagnosis Genomics and Epidemiology (CADDE) Genomic Network - Instituto de Medicina Tropical |
| EPI_ISL_2344552 | HOSPITAL DOS FORNCEDORES | Instituto Butantan / ESALQ-Piracicaba | Dimas Tadeu Covas, Antonio Jorge Martins, Claudia Renata dos Santos Barros, David Schlesinger, Debora Botequio Moretti, Elaine Cristina Marqueze, Elaine Vieira Santos, Evandra Strazza Rodrigues, Heidge Fukumasu, Jayme Augusto de Souza-Neto, José Salvatore Leister Patané, Luiz Alcantara, Luiz Lehmann Coutinho, Maria Carolina Elias, Maurício Lacerda Nogueira, Rafael dos Santos Bezerra, Raul Machado Neto, Rejane Maria Tommasini Grotto, Ricardo Haddad, Sandra Coccuzzo Sampaio Vessoni, Simone Kashima, Svetoslav Nanev Slavov, Vincent Louis Viala |
| EPI_ISL_2344553, EPI_ISL_2344554 | LABORATORIO MUNICIPAL DE PIRACICABA | Instituto Butantan / ESALQ-Piracicaba | Dimas Tadeu Covas, Antonio Jorge Martins, Claudia Renata dos Santos Barros, David Schlesinger, Debora Botequio Moretti, Elaine Cristina Marqueze, Elaine Vieira Santos, Evandra Strazza Rodrigues, Heidge Fukumasu, Jayme Augusto de Souza-Neto, José Salvatore Leister Patané, Luiz Alcantara, Luiz Lehmann Coutinho, Maria Carolina Elias, Maurício Lacerda Nogueira, Rafael dos Santos Bezerra, Raul Machado Neto, Rejane Maria Tommasini Grotto, Ricardo Haddad, Sandra Coccuzzo Sampaio Vessoni, Simone Kashima, Svetoslav Nanev Slavov, Vincent Louis Viala |
| EPI_ISL_2344662 | SECRETARIA DE SAUDE DE SAO PEDRO | Instituto Butantan / ESALQ-Piracicaba | Dimas Tadeu Covas, Antonio Jorge Martins, Claudia Renata dos Santos Barros, David Schlesinger, Debora Botequio Moretti, Elaine Cristina Marqueze, Elaine Vieira Santos, Evandra Strazza Rodrigues, Heidge Fukumasu, Jayme Augusto de Souza-Neto, José Salvatore Leister Patané, Luiz Alcantara, Luiz Lehmann Coutinho, Maria Carolina Elias, Maurício Lacerda Nogueira, Rafael dos Santos Bezerra, Raul Machado Neto, Rejane Maria Tommasini Grotto, Ricardo Haddad, Sandra Coccuzzo Sampaio Vessoni, Simone Kashima, Svetoslav Nanev Slavov, Vincent Louis Viala |
| EPI_ISL_2344851 | LABORATORIO MUNICIPAL DE PIRACICABA | Instituto Butantan / ESALQ-Piracicaba | Dimas Tadeu Covas, Antonio Jorge Martins, Claudia Renata dos Santos Barros, David Schlesinger, Debora Botequio Moretti, Elaine Cristina Marqueze, Elaine Vieira Santos, Evandra Strazza Rodrigues, Heidge Fukumasu, Jayme Augusto de Souza-Neto, José Salvatore Leister Patané, Luiz Alcantara, Luiz Lehmann Coutinho, Maria Carolina Elias, Maurício Lacerda Nogueira, Rafael dos Santos Bezerra, Raul Machado Neto, Rejane Maria Tommasini Grotto, Ricardo Haddad, Sandra Coccuzzo Sampaio Vessoni, Simone Kashima, Svetoslav Nanev Slavov, Vincent Louis Viala |
| EPI_ISL_2344900 | HOSPITAL E PRONTO SOCORRO PORTINARI | Instituto Butantan | Dimas Tadeu Covas, Antonio Jorge Martins, Claudia Renata dos Santos Barros, David Schlesinger, Debora Botequio Moretti, Elaine Cristina Marqueze, Elaine Vieira Santos, Evandra Strazza Rodrigues, Heidge Fukumasu, Jayme Augusto de Souza-Neto, José Salvatore Leister Patané, Luiz Alcantara, Luiz Lehmann Coutinho, Maria Carolina Elias, Maurício Lacerda Nogueira, Rafael dos Santos Bezerra, Raul Machado Neto, Rejane Maria Tommasini Grotto, Ricardo Haddad, Sandra Coccuzzo Sampaio Vessoni, Simone Kashima, Svetoslav Nanev Slavov, Vincent Louis Viala |
| EPI_ISL_2344915 | LABORATORIO MUNICIPAL DE PIRACICABA | Instituto Butantan / ESALQ-Piracicaba | Dimas Tadeu Covas, Antonio Jorge Martins, Claudia Renata dos Santos Barros, David Schlesinger, Debora Botequio Moretti, Elaine Cristina Marqueze, Elaine Vieira Santos, Evandra Strazza Rodrigues, Heidge Fukumasu, Jayme Augusto de Souza-Neto, José Salvatore Leister Patané, Luiz Alcantara, Luiz Lehmann Coutinho, Maria Carolina Elias, Maurício Lacerda Nogueira, Rafael dos Santos Bezerra, Raul Machado Neto, Rejane Maria Tommasini Grotto, Ricardo Haddad, Sandra Coccuzzo Sampaio Vessoni, Simone Kashima, Svetoslav Nanev Slavov, Vincent Louis Viala |
| EPI_ISL_2345181 | NUCLEO SAUDE III N HABITACIONAL PRESIDENTE GEISEL BAURU | Instituto Butantan / ESALQ-Piracicaba | Dimas Tadeu Covas, Antonio Jorge Martins, Claudia Renata dos Santos Barros, David Schlesinger, Debora Botequio Moretti, Elaine Cristina Marqueze, Elaine Vieira Santos, Evandra Strazza Rodrigues, Heidge Fukumasu, Jayme Augusto de Souza-Neto, José Salvatore Leister Patané, Luiz Alcantara, Luiz Lehmann Coutinho, Maria Carolina Elias, Maurício Lacerda Nogueira, Rafael dos Santos Bezerra, Raul Machado Neto, Rejane Maria Tommasini Grotto, Ricardo Haddad, Sandra Coccuzzo Sampaio Vessoni, Simone Kashima, Svetoslav Nanev Slavov, Vincent Louis Viala |
| EPI_ISL_2345207 | NUCLEO DE SAUDE VILA FALCAO DE BAURU | Instituto Butantan / ESALQ-Piracicaba | Dimas Tadeu Covas, Antonio Jorge Martins, Claudia Renata dos Santos Barros, David Schlesinger, Debora Botequio Moretti, Elaine Cristina Marqueze, Elaine Vieira Santos, Evandra Strazza Rodrigues, Heidge Fukumasu, Jayme Augusto de Souza-Neto, José Salvatore Leister Patané, Luiz Alcantara, Luiz Lehmann Coutinho, Maria Carolina Elias, Maurício Lacerda Nogueira, Rafael dos Santos Bezerra, Raul Machado Neto, Rejane Maria Tommasini Grotto, Ricardo Haddad, Sandra Coccuzzo Sampaio Vessoni, Simone Kashima, Svetoslav Nanev Slavov, Vincent Louis Viala |
| EPI_ISL_2345267 | LABORATORIO MUNICIPAL DE PIRACICABA | Instituto Butantan / ESALQ-Piracicaba | Dimas Tadeu Covas, Antonio Jorge Martins, Claudia Renata dos Santos Barros, David Schlesinger, Debora Botequio Moretti, Elaine Cristina Marqueze, Elaine Vieira Santos, Evandra Strazza Rodrigues, Heidge Fukumasu, Jayme Augusto de Souza-Neto, José Salvatore Leister Patané, Luiz Alcantara, Luiz Lehmann Coutinho, Maria Carolina Elias, Maurício Lacerda Nogueira, Rafael dos Santos Bezerra, Raul Machado Neto, Rejane Maria Tommasini Grotto, Ricardo Haddad, Sandra Coccuzzo Sampaio Vessoni, Simone Kashima, Svetoslav Nanev Slavov, Vincent Louis Viala |
| EPI_ISL_2345268 | UPA IPIRANGA | Instituto Butantan / ESALQ-Piracicaba | Dimas Tadeu Covas, Antonio Jorge Martins, Claudia Renata dos Santos Barros, David Schlesinger, Debora Botequio Moretti, Elaine Cristina Marqueze, Elaine Vieira Santos, Evandra Strazza Rodrigues, Heidge Fukumasu, Jayme Augusto de Souza-Neto, José Salvatore Leister Patané, Luiz Alcantara, Luiz Lehmann Coutinho, Maria Carolina Elias, Maurício Lacerda Nogueira, Rafael dos Santos Bezerra, Raul Machado Neto, Rejane Maria Tommasini Grotto, Ricardo Haddad, Sandra Coccuzzo Sampaio Vessoni, Simone Kashima, Svetoslav Nanev Slavov, Vincent Louis Viala |
| EPI_ISL_2345453 | HOSPITAL REGIONAL DE ITAPETININGA | Instituto Butantan / ESALQ-Piracicaba | Dimas Tadeu Covas, Antonio Jorge Martins, Claudia Renata dos Santos Barros, David Schlesinger, Debora Botequio Moretti, Elaine Cristina Marqueze, Elaine Vieira Santos, Evandra Strazza Rodrigues, Heidge Fukumasu, Jayme Augusto de Souza-Neto, José Salvatore Leister Patané, Luiz Alcantara, Luiz Lehmann Coutinho, Maria Carolina Elias, Maurício Lacerda Nogueira, Rafael dos Santos Bezerra, Raul Machado Neto, Rejane Maria Tommasini Grotto, Ricardo Haddad, Sandra Coccuzzo Sampaio Vessoni, Simone Kashima, Svetoslav Nanev Slavov, Vincent Louis Viala |
| EPI_ISL_2345590 | CENTRO DE ESPECIALIDADES DE PRIMAVERA | Instituto Butantan / ESALQ-Piracicaba | Dimas Tadeu Covas, Antonio Jorge Martins, Claudia Renata dos Santos Barros, David Schlesinger, Debora Botequio Moretti, Elaine Cristina Marqueze, Elaine Vieira Santos, Evandra Strazza Rodrigues, Heidge Fukumasu, Jayme Augusto de Souza-Neto, José Salvatore Leister Patané, Luiz Alcantara, Luiz Lehmann Coutinho, Maria Carolina Elias, Maurício Lacerda Nogueira, Rafael dos Santos Bezerra, Raul Machado Neto, Rejane Maria Tommasini Grotto, Ricardo Haddad, Sandra Coccuzzo Sampaio Vessoni, Simone Kashima, Svetoslav Nanev Slavov, Vincent Louis Viala |
| EPI_ISL_2345715 | CENTRO DE ESPECIALIDADES DE PRIMAVERA | Instituto Butantan / Mendelics | Dimas Tadeu Covas, Antonio Jorge Martins, Claudia Renata dos Santos Barros, David Schlesinger, Debora Botequio Moretti, Elaine Cristina Marqueze, Elaine Vieira Santos, Evandra Strazza Rodrigues, Heidge Fukumasu, Jayme Augusto de Souza-Neto, José Salvatore Leister Patané, Luiz Alcantara, Luiz Lehmann Coutinho, Maria Carolina Elias, Maurício Lacerda Nogueira, Rafael dos Santos Bezerra, Raul Machado Neto, Rejane Maria Tommasini Grotto, Ricardo Haddad, Sandra Coccuzzo Sampaio Vessoni, Simone Kashima, Svetoslav Nanev Slavov, Vincent Louis Viala |
| EPI_ISL_2345723 | UNIDADE BASICA DE SAUDE JARDIM DO LAGO | Instituto Butantan / Mendelics | Dimas Tadeu Covas, Antonio Jorge Martins, Claudia Renata dos Santos Barros, David Schlesinger, Debora Botequio Moretti, Elaine Cristina Marqueze, Elaine Vieira Santos, Evandra Strazza Rodrigues, Heidge Fukumasu, Jayme Augusto de Souza-Neto, José Salvatore Leister Patané, Luiz Alcantara, Luiz Lehmann Coutinho, Maria Carolina Elias, Maurício Lacerda Nogueira, Rafael dos Santos Bezerra, Raul Machado Neto, Rejane Maria Tommasini Grotto, Ricardo Haddad, Sandra Coccuzzo Sampaio Vessoni, Simone Kashima, Svetoslav Nanev Slavov, Vincent Louis Viala |
| EPI_ISL_2345755 | SECRETARIA MUNICIPAL DE SAUDE SOROCABA | Instituto Butantan / Mendelics | Dimas Tadeu Covas, Antonio Jorge Martins, Claudia Renata dos Santos Barros, David Schlesinger, Debora Botequio Moretti, Elaine Cristina Marqueze, Elaine Vieira Santos, Evandra Strazza Rodrigues, Heidge Fukumasu, Jayme Augusto de Souza-Neto, José Salvatore Leister Patané, Luiz Alcantara, Luiz Lehmann Coutinho, Maria Carolina Elias, Maurício Lacerda Nogueira, Rafael dos Santos Bezerra, Raul Machado Neto, Rejane Maria Tommasini Grotto, Ricardo Haddad, Sandra Coccuzzo Sampaio Vessoni, Simone Kashima, Svetoslav Nanev Slavov, Vincent Louis Viala |
| EPI_ISL_2345811 | CENTRO DE ESPECIALIDADES DE PRIMAVERA | Instituto Butantan / Mendelics | Dimas Tadeu Covas, Antonio Jorge Martins, Claudia Renata dos Santos Barros, David Schlesinger, Debora Botequio Moretti, Elaine Cristina Marqueze, Elaine Vieira Santos, Evandra Strazza Rodrigues, Heidge Fukumasu, Jayme Augusto de Souza-Neto, José Salvatore Leister Patané, Luiz Alcantara, Luiz Lehmann Coutinho, Maria Carolina Elias, Maurício Lacerda Nogueira, Rafael dos Santos Bezerra, Raul Machado Neto, Rejane Maria Tommasini Grotto, Ricardo Haddad, Sandra Coccuzzo Sampaio Vessoni, Simone Kashima, Svetoslav Nanev Slavov, Vincent Louis Viala |
| EPI_ISL_2345874 | CENTRO DE APOIO EPIDEMIOLOGICO | Instituto Butantan / Mendelics | Dimas Tadeu Covas, Antonio Jorge Martins, Claudia Renata dos Santos Barros, David Schlesinger, Debora Botequio Moretti, Elaine Cristina Marqueze, Elaine Vieira Santos, Evandra Strazza Rodrigues, Heidge Fukumasu, Jayme Augusto de Souza-Neto, José Salvatore Leister Patané, Luiz Alcantara, Luiz Lehmann Coutinho, Maria Carolina Elias, Maurício Lacerda Nogueira, Rafael dos Santos Bezerra, Raul Machado Neto, Rejane Maria Tommasini Grotto, Ricardo Haddad, Sandra Coccuzzo Sampaio Vessoni, Simone Kashima, Svetoslav Nanev Slavov, Vincent Louis Viala |
| EPI_ISL_2346088, EPI_ISL_2346090 | Prefeirtura de SP | Instituto Butantan | Dimas Tadeu Covas, Antonio Jorge Martins, Claudia Renata dos Santos Barros, David Schlesinger, Debora Botequio Moretti, Elaine Cristina Marqueze, Elaine Vieira Santos, Evandra Strazza Rodrigues, Heidge Fukumasu, Jayme Augusto de Souza-Neto, José Salvatore Leister Patané, Luiz Alcantara, Luiz |

| | | | |
|---|---|---|---|
| | | | Lehmann Coutinho, Maria Carolina Elias, Maurício Lacerda Nogueira, Rafael dos Santos Bezerra, Raul Machado Neto, Rejane Maria Tommasini Grotto, Ricardo Haddad, Sandra Coccuzzo Sampaio Vessoni, Simone Kashima, Svetoslav Nanev Slavov, Vincent Louis Viala |
| EPI_ISL_2378653, EPI_ISL_2378681 | Instituto de Biotecnologia - UNESP-Botucatu-SP | Instituto de Biotecnologia - UNESP-Botucatu-SP | Fábio Sossai Possebon; Leila Sabrina Ullmann; Cecília Artico Banho; Cíntia Bittar; Guilherme Campos; Helena Lage Ferreira; Jorge A. Petroli Marchesi; Lívia Sacchetto; Maisa C. Pereira Parra; Marília Moraes; Maurício L. Nogueira; Paula Rahal; Paulo Inacio da Costa; João Pessoa Araújo Jr. |
| EPI_ISL_2378745 | HOSPITAL DO RIM E HIPERTENSAO | Instituto Butantan | Dimas Tadeu Covas, Antonio Jorge Martins, Claudia Renata dos Santos Barros, David Schlesinger, Debora Botequio Moretti, Elaine Cristina Marqueze, Elaine Vieira Santos, Evandra Strazza Rodrigues, Heidge Fukumasu, Jayme Augusto de Souza-Neto, José Salvatore Leister Patané, Luiz Alcantara, Luiz Lehmann Coutinho, Maria Carolina Elias, Maurício Lacerda Nogueira, Rafael dos Santos Bezerra, Raul Machado Neto, Rejane Maria Tommasini Grotto, Ricardo Haddad, Sandra Coccuzzo Sampaio Vessoni, Simone Kashima, Svetoslav Nanev Slavov, Vincent Louis Viala |
| EPI_ISL_2383033 | LABORATORIO MUNICIPAL DE PIRACICABA | Instituto Butantan | Dimas Tadeu Covas, Antonio Jorge Martins, Claudia Renata dos Santos Barros, David Schlesinger, Debora Botequio Moretti, Elaine Cristina Marqueze, Elaine Vieira Santos, Evandra Strazza Rodrigues, Heidge Fukumasu, Jayme Augusto de Souza-Neto, José Salvatore Leister Patané, Luiz Alcantara, Luiz Lehmann Coutinho, Maria Carolina Elias, Maurício Lacerda Nogueira, Rafael dos Santos Bezerra, Raul Machado Neto, Rejane Maria Tommasini Grotto, Ricardo Haddad, Sandra Coccuzzo Sampaio Vessoni, Simone Kashima, Svetoslav Nanev Slavov, Vincent Louis Viala |
| EPI_ISL_2383035 | UNIDADE BASICA DE SAUDE DR MATHEUS GABRIEL BONASSA | Instituto Butantan | Dimas Tadeu Covas, Antonio Jorge Martins, Claudia Renata dos Santos Barros, David Schlesinger, Debora Botequio Moretti, Elaine Cristina Marqueze, Elaine Vieira Santos, Evandra Strazza Rodrigues, Heidge Fukumasu, Jayme Augusto de Souza-Neto, José Salvatore Leister Patané, Luiz Alcantara, Luiz Lehmann Coutinho, Maria Carolina Elias, Maurício Lacerda Nogueira, Rafael dos Santos Bezerra, Raul Machado Neto, Rejane Maria Tommasini Grotto, Ricardo Haddad, Sandra Coccuzzo Sampaio Vessoni, Simone Kashima, Svetoslav Nanev Slavov, Vincent Louis Viala |
| EPI_ISL_2385432, EPI_ISL_2385433, EPI_ISL_2385434, EPI_ISL_2385435, EPI_ISL_2385436, EPI_ISL_2385437, EPI_ISL_2385438, EPI_ISL_2385439, EPI_ISL_2385440, EPI_ISL_2385441, EPI_ISL_2385442, EPI_ISL_2385443, EPI_ISL_2385444, EPI_ISL_2385445, EPI_ISL_2385446, EPI_ISL_2385447, EPI_ISL_2385448, EPI_ISL_2385449, EPI_ISL_2385450, EPI_ISL_2385451, EPI_ISL_2385452, EPI_ISL_2385453, EPI_ISL_2385454, EPI_ISL_2385455, EPI_ISL_2385456, EPI_ISL_2385457, EPI_ISL_2385458, EPI_ISL_2385459, EPI_ISL_2385460, EPI_ISL_2385461, EPI_ISL_2385462, EPI_ISL_2385463, EPI_ISL_2385464, EPI_ISL_2385465, EPI_ISL_2385466, EPI_ISL_2385467, EPI_ISL_2385468, EPI_ISL_2385469, EPI_ISL_2385470, EPI_ISL_2385471, EPI_ISL_2385472, EPI_ISL_2385474, EPI_ISL_2385475, EPI_ISL_2385476, EPI_ISL_2385477, EPI_ISL_2385478, EPI_ISL_2385479, EPI_ISL_2385480, EPI_ISL_2385481, EPI_ISL_2385482, EPI_ISL_2385483, EPI_ISL_2385484, EPI_ISL_2385485, EPI_ISL_2385486, EPI_ISL_2385487, EPI_ISL_2385488, EPI_ISL_2385489, EPI_ISL_2385490, EPI_ISL_2385491, EPI_ISL_2385492, EPI_ISL_2385493, EPI_ISL_2385494, EPI_ISL_2385495, EPI_ISL_2385496, EPI_ISL_2385498, EPI_ISL_2385499, EPI_ISL_2385500, EPI_ISL_2385501, EPI_ISL_2385502, EPI_ISL_2385503, EPI_ISL_2385504, EPI_ISL_2385505, EPI_ISL_2385506, EPI_ISL_2385507, EPI_ISL_2385508, EPI_ISL_2385510, EPI_ISL_2385511, EPI_ISL_2385512, EPI_ISL_2385513, EPI_ISL_2385514, EPI_ISL_2385515, EPI_ISL_2385516, EPI_ISL_2385517, EPI_ISL_2385518, EPI_ISL_2385519, EPI_ISL_2385520, EPI_ISL_2385521, EPI_ISL_2385522, EPI_ISL_2385523, EPI_ISL_2385524, EPI_ISL_2385525, EPI_ISL_2385526, EPI_ISL_2385527, EPI_ISL_2385529, EPI_ISL_2385530, EPI_ISL_2385531, EPI_ISL_2385532, EPI_ISL_2385533, EPI_ISL_2385534, EPI_ISL_2385535, EPI_ISL_2385536, EPI_ISL_2385537, EPI_ISL_2385538, EPI_ISL_2385539, EPI_ISL_2385541, EPI_ISL_2385542, EPI_ISL_2385543, EPI_ISL_2385544, EPI_ISL_2385545, EPI_ISL_2385546, EPI_ISL_2385547, EPI_ISL_2385548, EPI_ISL_2385549, EPI_ISL_2385550, EPI_ISL_2385551, EPI_ISL_2385552, EPI_ISL_2385553, EPI_ISL_2385554, EPI_ISL_2385555, EPI_ISL_2385556, EPI_ISL_2385558, EPI_ISL_2385559, EPI_ISL_2385561, EPI_ISL_2385562, EPI_ISL_2385563, EPI_ISL_2385564, EPI_ISL_2385565, EPI_ISL_2385566, EPI_ISL_2385567, EPI_ISL_2385568, EPI_ISL_2385570, EPI_ISL_2385571, EPI_ISL_2385572, EPI_ISL_2385573, EPI_ISL_2385574, EPI_ISL_2385575, EPI_ISL_2385576, EPI_ISL_2385577, EPI_ISL_2385578, EPI_ISL_2385579, EPI_ISL_2385580, EPI_ISL_2385581, EPI_ISL_2385582, EPI_ISL_2385583, EPI_ISL_2385584, EPI_ISL_2385586, EPI_ISL_2385587, EPI_ISL_2385588, EPI_ISL_2385589, EPI_ISL_2385590, EPI_ISL_2385591, EPI_ISL_2385592, EPI_ISL_2385593, EPI_ISL_2385594, EPI_ISL_2385595, EPI_ISL_2385596, EPI_ISL_2385597, EPI_ISL_2385598, EPI_ISL_2385599, EPI_ISL_2385600, EPI_ISL_2385601, EPI_ISL_2385602, EPI_ISL_2385603, EPI_ISL_2385604, EPI_ISL_2385605, EPI_ISL_2385606, EPI_ISL_2385607, EPI_ISL_2385608, EPI_ISL_2385609, EPI_ISL_2385610, EPI_ISL_2385611, EPI_ISL_2385612, EPI_ISL_2385613, EPI_ISL_2385616, EPI_ISL_2385620, EPI_ISL_2385624, EPI_ISL_2385625, EPI_ISL_2385627, EPI_ISL_2385629, EPI_ISL_2385630, EPI_ISL_2385631, EPI_ISL_2385632, EPI_ISL_2385633, EPI_ISL_2385634, EPI_ISL_2385636, EPI_ISL_2385637, EPI_ISL_2385639, EPI_ISL_2385641, EPI_ISL_2385642, EPI_ISL_2385644, EPI_ISL_2385647, EPI_ISL_2385648, EPI_ISL_2385649, EPI_ISL_2385650, EPI_ISL_2385651, EPI_ISL_2385652, EPI_ISL_2385653, EPI_ISL_2385654, EPI_ISL_2385655, EPI_ISL_2385656, EPI_ISL_2385657, EPI_ISL_2385658, EPI_ISL_2385659, EPI_ISL_2385660, EPI_ISL_2385661, EPI_ISL_2385662, EPI_ISL_2385663, EPI_ISL_2385664, EPI_ISL_2385665, EPI_ISL_2385666, EPI_ISL_2385667, EPI_ISL_2385668 | | | |
| see above | Laboratorio Central Noel Nutels | Bioinformatics Laboratory / LNCC | Luiz G P de Almeida, Alessandra P Lamarca, Ronaldo da Silva F Jr, Liliane Cavalcante, Alexandra L Gerber, Ana Paula de C Guimaraes, Douglas Terra Machado, Cassia Alves, Diana Mariani, Cintia Policarpo, Gleidson da Silva de Oliveira, Mario Sergio Ribeiro, Silvia Carvalho, Flavio Dias da Silva, Marcio Henrique de Oliveira Garcia, Leandro Magalhaes de Souza, Cristiane Gomes da Silva, Caio Luiz Pereira Ribeiro, Andrea Cony Cavalcanti, Claudia Maria Braga de Mello, Amilcar Tanuri, Ana Tereza R Vasconcelos |
| EPI_ISL_2385669, EPI_ISL_2385670, EPI_ISL_2385671, EPI_ISL_2385674, EPI_ISL_2385675, EPI_ISL_2385676, EPI_ISL_2385677, EPI_ISL_2385678, EPI_ISL_2385679, EPI_ISL_2385680, EPI_ISL_2385681, EPI_ISL_2385682, EPI_ISL_2385683, EPI_ISL_2385684, EPI_ISL_2385685, EPI_ISL_2385686, EPI_ISL_2385689, EPI_ISL_2385690, EPI_ISL_2385691, EPI_ISL_2385692, EPI_ISL_2385693, EPI_ISL_2385694, EPI_ISL_2385695, EPI_ISL_2385696, EPI_ISL_2385697, EPI_ISL_2385698, EPI_ISL_2385699, EPI_ISL_2385700, EPI_ISL_2385701, EPI_ISL_2385702, EPI_ISL_2385704, EPI_ISL_2385705, EPI_ISL_2385707, EPI_ISL_2385708, EPI_ISL_2385709, EPI_ISL_2385711, EPI_ISL_2385712, EPI_ISL_2385713, EPI_ISL_2385714, EPI_ISL_2385717, EPI_ISL_2385718, EPI_ISL_2385719, EPI_ISL_2385720, EPI_ISL_2385723, EPI_ISL_2385724, EPI_ISL_2385725, EPI_ISL_2385726, EPI_ISL_2385727, EPI_ISL_2385728, EPI_ISL_2385729, EPI_ISL_2385730, EPI_ISL_2385731, EPI_ISL_2385732, EPI_ISL_2385733, EPI_ISL_2385734, EPI_ISL_2385735, EPI_ISL_2385736, EPI_ISL_2385737, EPI_ISL_2385739, EPI_ISL_2385740, EPI_ISL_2385741, EPI_ISL_2385742, EPI_ISL_2385743, EPI_ISL_2385745, EPI_ISL_2385748, EPI_ISL_2385749, EPI_ISL_2385750, EPI_ISL_2385751, EPI_ISL_2385752, EPI_ISL_2385753, EPI_ISL_2385754, EPI_ISL_2385755, EPI_ISL_2385756, EPI_ISL_2385757, EPI_ISL_2385758, EPI_ISL_2385759, EPI_ISL_2385760, EPI_ISL_2385761, EPI_ISL_2385764, EPI_ISL_2385765, EPI_ISL_2385766, EPI_ISL_2385768, EPI_ISL_2385769, EPI_ISL_2385770, EPI_ISL_2385771, EPI_ISL_2385772, EPI_ISL_2385773, EPI_ISL_2385774, EPI_ISL_2385775, EPI_ISL_2385776, EPI_ISL_2385777, EPI_ISL_2385778, EPI_ISL_2385779, EPI_ISL_2385780, EPI_ISL_2385781, EPI_ISL_2385782, EPI_ISL_2385784, EPI_ISL_2385785, EPI_ISL_2385786 | | | |
| see above | Unidade de apoio ao diagnóstico da COVID - UNADIG | Bioinformatics Laboratory / LNCC | Luiz G P de Almeida, Alessandra P Lamarca, Ronaldo da Silva F Jr, Liliane Cavalcante, Alexandra L Gerber, Ana Paula de C Guimaraes, Douglas Terra Machado, Cassia Alves, Diana Mariani, Cintia Policarpo, Gleidson da Silva de Oliveira, Mario Sergio Ribeiro, Silvia Carvalho, Flavio Dias da Silva, Marcio Henrique de Oliveira Garcia, Leandro Magalhaes de Souza, Cristiane Gomes da Silva, Caio Luiz Pereira Ribeiro, Andrea Cony Cavalcanti, Claudia Maria Braga de Mello, Amilcar Tanuri, Ana Tereza R Vasconcelos |
| EPI_ISL_2431437 | Laboratório de Microbiologia Molecular - Universidade FEEVALE | Molecular Microbiology Laboratory | Alana Witt Hansen, Fágner Henrique Heldt, Fernando Rosado Spilki, Flávio Silveira, Juliana Schons Gularte, Juliane Deise Fleck, Mariana Soares da Silva, Meriane Demoliner, Matheus Nunes Weber, Paula Rodrigues de Almeida, Micheli Filippi. |
| EPI_ISL_2443542 | Lboratorio Central de Saude Publica do Estado do Parana (LACEN/PR) | Laboratory of Respiratory Viruses and Measles, Oswaldo Cruz Institute, FIOCRUZ | Paola Resende, Luciana Appolinario, Fernando Motta, Anna Carolina Paixao, Ana Carolina Mendonca, Alice Sampaio Rocha, Taina Venas, Elisa Cavalcante Pereira, Renata Serrano Lopes, Irina Riediger, Marilda Siqueira on behalf of the Fiocruz COVID-19 Genomic Surveillance Network |
| EPI_ISL_2443578, EPI_ISL_2443579, EPI_ISL_2443580, EPI_ISL_2443581, EPI_ISL_2443582, EPI_ISL_2443583, EPI_ISL_2443584, EPI_ISL_2443585, EPI_ISL_2443586, EPI_ISL_2443587, EPI_ISL_2443588, EPI_ISL_2443589, EPI_ISL_2443590, EPI_ISL_2443591, EPI_ISL_2443592, EPI_ISL_2443593, EPI_ISL_2443594, EPI_ISL_2443595, EPI_ISL_2443596, EPI_ISL_2443597, EPI_ISL_2443598, EPI_ISL_2443599, EPI_ISL_2443600, EPI_ISL_2443601, EPI_ISL_2443602, EPI_ISL_2443603, EPI_ISL_2443604, EPI_ISL_2443605, EPI_ISL_2443606, EPI_ISL_2443607, EPI_ISL_2443608, EPI_ISL_2443609, EPI_ISL_2443610, EPI_ISL_2443611, EPI_ISL_2443612, EPI_ISL_2443613 | | | |
| see above | Laboratory of Respiratory Viruses and Measles, Oswaldo Cruz Institute, FIOCRUZ | Laboratory of Respiratory Viruses and Measles, Oswaldo Cruz Institute, FIOCRUZ | Paola Resende, Luciana Appolinario, Fernando Motta, Anna Carolina Paixao, Ana Carolina Mendonca, Alice Sampaio Rocha, Taina Venas, Elisa Cavalcante Pereira, Renata Serrano Lopes, Marilda Siqueira on behalf of the Fiocruz COVID-19 Genomic Surveillance Network |
| EPI_ISL_2443614, EPI_ISL_2443615, EPI_ISL_2443616, EPI_ISL_2443617, EPI_ISL_2443618, EPI_ISL_2443619, EPI_ISL_2443620, EPI_ISL_2443621, EPI_ISL_2443622, EPI_ISL_2443623, EPI_ISL_2443624, EPI_ISL_2443625, EPI_ISL_2443626, EPI_ISL_2443627, EPI_ISL_2443628, EPI_ISL_2443629, EPI_ISL_2443630, EPI_ISL_2443631, EPI_ISL_2443632 | | | |
| see above | Labortorio Central de Saude Publica do Estado do Rio de Janeiro (LACEN/RJ) | Laboratory of Respiratory Viruses and Measles, Oswaldo Cruz Institute, FIOCRUZ | Paola Resende, Luciana Appolinario, Fernando Motta, Anna Carolina Paixao, Ana Carolina Mendonca, Alice Sampaio Rocha, Taina Venas, Elisa Cavalcante Pereira, Renata Serrano Lopes, Andrea Cony Cavalcanti, Marilda Siqueira on behalf of the Fiocruz COVID-19 Genomic Surveillance Network |
| EPI_ISL_2443633, EPI_ISL_2443634, EPI_ISL_2443635, EPI_ISL_2443636 | Laboratory of Respiratory Viruses and Measles, Oswaldo Cruz Institute, FIOCRUZ | Laboratory of Respiratory Viruses and Measles, Oswaldo Cruz Institute, FIOCRUZ | Paola Resende, Luciana Appolinario, Fernando Motta, Anna Carolina Paixao, Ana Carolina Mendonca, Alice Sampaio Rocha, Taina Venas, Elisa Cavalcante Pereira, Renata Serrano Lopes, Marilda Siqueira on behalf of the Fiocruz COVID-19 Genomic Surveillance Network |
| EPI_ISL_2443637, EPI_ISL_2443638, EPI_ISL_2443639, EPI_ISL_2443640, EPI_ISL_2443641, EPI_ISL_2443642, EPI_ISL_2443643, EPI_ISL_2443644, EPI_ISL_2443645, EPI_ISL_2443646, EPI_ISL_2443647, EPI_ISL_2443648, EPI_ISL_2443649, EPI_ISL_2443650, EPI_ISL_2443651, EPI_ISL_2443652, EPI_ISL_2443653, EPI_ISL_2443654, EPI_ISL_2443655, EPI_ISL_2443656, EPI_ISL_2443657, EPI_ISL_2443658 | | | |
| see above | Labortorio Central de Saude Publica do Estado do Rio de Janeiro (LACEN/RJ) | Laboratory of Respiratory Viruses and Measles, Oswaldo Cruz Institute, FIOCRUZ | Paola Resende, Luciana Appolinario, Fernando Motta, Anna Carolina Paixao, Ana Carolina Mendonca, Alice Sampaio Rocha, Taina Venas, Elisa Cavalcante Pereira, Renata Serrano Lopes, Andrea Cony Cavalcanti, Marilda Siqueira on behalf of the Fiocruz COVID-19 Genomic Surveillance Network |
| EPI_ISL_2443659, EPI_ISL_2443660, EPI_ISL_2443661, EPI_ISL_2443662, EPI_ISL_2443663, EPI_ISL_2443664, EPI_ISL_2443665, EPI_ISL_2443666, EPI_ISL_2443667, EPI_ISL_2443668, EPI_ISL_2443669, EPI_ISL_2443670, EPI_ISL_2443671 | | | |
| see above | Laboratory of Respiratory Viruses and Measles, Oswaldo Cruz Institute, FIOCRUZ | Laboratory of Respiratory Viruses and Measles, Oswaldo Cruz Institute, FIOCRUZ | Paola Resende, Luciana Appolinario, Fernando Motta, Anna Carolina Paixao, Ana Carolina Mendonca, Alice Sampaio Rocha, Taina Venas, Elisa Cavalcante Pereira, Renata Serrano Lopes, Marilda Siqueira on behalf of the Fiocruz COVID-19 Genomic Surveillance Network |
| EPI_ISL_2443675, EPI_ISL_2443712 | Labortorio Central de Saude Publica do Estado de Santa Catarina (LACEN/SC) | Laboratory of Respiratory Viruses and Measles, Oswaldo Cruz Institute, FIOCRUZ | Paola Resende, Luciana Appolinario, Fernando Motta, Anna Carolina Paixao, Ana Carolina Mendonca, Alice Sampaio Rocha, Taina Venas, Elisa Cavalcante Pereira, Renata Serrano Lopes, Darcita Buerger Rovaris, Sandra Bianchini Fernandes, Marilda Siqueira on behalf of the Fiocruz COVID-19 Genomic Surveillance Network |
| EPI_ISL_2444817 | IICS-UNA | IICS-UNA | Magaly Martinez, Adriana Valenzuela, Alejandra Rojas, Chyntia Diaz, Eva Nara, Fatima Cardozo, Florencia del Puerto, Joel Ortiz, Jonas Fernandez, Laura Franco, Laura Mendoza, Leticia Rojas, Maria Eugenia Galeano. |
| EPI_ISL_2445168 | UNIDADE REFERENCIAL SUDOESTE | Instituto Butantan | Dimas Tadeu Covas, Antonio Jorge Martins, Claudia Renata dos Santos Barros, David Schlesinger, Debora Botequio Moretti, Elaine Cristina Marqueze, Elaine Vieira Santos, Evandra Strazza Rodrigues, Heidge Fukumasu, Jayme Augusto de Souza-Neto, José Salvatore Leister Patané, Luiz Alcantara, Luiz Lehmann Coutinho, Maria Carolina Elias, Maurício Lacerda Nogueira, Rafael dos Santos Bezerra, Raul Machado Neto, Rejane Maria Tommasini Grotto, Ricardo Haddad, Sandra Coccuzzo Sampaio Vessoni, Simone Kashima, Svetoslav Nanev Slavov, Vincent Louis Viala |
| EPI_ISL_2445275 | UPA VILA SANTA CATARINA | Instituto Butantan | Dimas Tadeu Covas, Antonio Jorge Martins, Claudia Renata dos Santos Barros, David Schlesinger, Debora Botequio Moretti, Elaine Cristina Marqueze, Elaine Vieira Santos, Evandra Strazza Rodrigues, Heidge Fukumasu, Jayme Augusto de Souza-Neto, José Salvatore Leister Patané, Luiz Alcantara, Luiz |

| | | | |
|---|---|---|---|
| | | | Lehmann Coutinho, Maria Carolina Elias, Maurício Lacerda Nogueira, Rafael dos Santos Bezerra, Raul Machado Neto, Rejane Maria Tommasini Grotto, Ricardo Haddad, Sandra Coccuzzo Sampaio Vessoni, Simone Kashima, Svetoslav Nanev Slavov, Vincent Louis Viala |
| EPI_ISL_2445406 | CS II DR MIGUEL VITALIANO ORLANDIA | Instituto Butantan | Dimas Tadeu Covas, Antonio Jorge Martins, Claudia Renata dos Santos Barros, David Schlesinger, Debora Botequio Moretti, Elaine Cristina Marqueze, Elaine Vieira Santos, Evandra Strazza Rodrigues, Heidge Fukumasu, Jayme Augusto de Souza-Neto, José Salvatore Leister Patané, Luiz Alcantara, Luiz Lehmann Coutinho, Maria Carolina Elias, Maurício Lacerda Nogueira, Rafael dos Santos Bezerra, Raul Machado Neto, Rejane Maria Tommasini Grotto, Ricardo Haddad, Sandra Coccuzzo Sampaio Vessoni, Simone Kashima, Svetoslav Nanev Slavov, Vincent Louis Viala |
| EPI_ISL_2445499 | UNIDADE DE SAUDE DR PHEBO DE OLIVEIRA ROGE FERREIRA | Instituto Butantan | Dimas Tadeu Covas, Antonio Jorge Martins, Claudia Renata dos Santos Barros, David Schlesinger, Debora Botequio Moretti, Elaine Cristina Marqueze, Elaine Vieira Santos, Evandra Strazza Rodrigues, Heidge Fukumasu, Jayme Augusto de Souza-Neto, José Salvatore Leister Patané, Luiz Alcantara, Luiz Lehmann Coutinho, Maria Carolina Elias, Maurício Lacerda Nogueira, Rafael dos Santos Bezerra, Raul Machado Neto, Rejane Maria Tommasini Grotto, Ricardo Haddad, Sandra Coccuzzo Sampaio Vessoni, Simone Kashima, Svetoslav Nanev Slavov, Vincent Louis Viala |
| EPI_ISL_2445564, EPI_ISL_2445570 | IRMANDADE DA SANTA CASA DE MISERICORDIA LORENA | Instituto Butantan | Dimas Tadeu Covas, Antonio Jorge Martins, Claudia Renata dos Santos Barros, David Schlesinger, Debora Botequio Moretti, Elaine Cristina Marqueze, Elaine Vieira Santos, Evandra Strazza Rodrigues, Heidge Fukumasu, Jayme Augusto de Souza-Neto, José Salvatore Leister Patané, Luiz Alcantara, Luiz Lehmann Coutinho, Maria Carolina Elias, Maurício Lacerda Nogueira, Rafael dos Santos Bezerra, Raul Machado Neto, Rejane Maria Tommasini Grotto, Ricardo Haddad, Sandra Coccuzzo Sampaio Vessoni, Simone Kashima, Svetoslav Nanev Slavov, Vincent Louis Viala |
| EPI_ISL_2445581 | HOSPITAL DE CAMPANHA COVID 19 MUNICIPIO DE TAUBATE | Instituto Butantan | Dimas Tadeu Covas, Antonio Jorge Martins, Claudia Renata dos Santos Barros, David Schlesinger, Debora Botequio Moretti, Elaine Cristina Marqueze, Elaine Vieira Santos, Evandra Strazza Rodrigues, Heidge Fukumasu, Jayme Augusto de Souza-Neto, José Salvatore Leister Patané, Luiz Alcantara, Luiz Lehmann Coutinho, Maria Carolina Elias, Maurício Lacerda Nogueira, Rafael dos Santos Bezerra, Raul Machado Neto, Rejane Maria Tommasini Grotto, Ricardo Haddad, Sandra Coccuzzo Sampaio Vessoni, Simone Kashima, Svetoslav Nanev Slavov, Vincent Louis Viala |
| EPI_ISL_2466122, EPI_ISL_2466123, EPI_ISL_2466124 | Laboratory of Respiratory Viruses and Measles, Oswaldo Cruz Institute, FIOCRUZ | Laboratory of Respiratory Viruses and Measles, Oswaldo Cruz Institute, FIOCRUZ | Paola Resende, Luciana Appolinario, Fernando Motta, Anna Carolina Paixao, Ana Carolina Mendonca, Alice Sampaio Rocha, Taina Venas, Elisa Cavalcante Pereira, Renata Serrano Lopes, Marilda Siqueira on behalf of the Fiocruz COVID-19 Genomic Surveillance Network |
| EPI_ISL_2466230 | Labortorio Central de Saude Publica do Estado do Rio de Janeiro (LACEN/RJ) | Laboratory of Respiratory Viruses and Measles, Oswaldo Cruz Institute, FIOCRUZ | Paola Resende, Luciana Appolinario, Fernando Motta, Anna Carolina Paixao, Ana Carolina Mendonca, Alice Sampaio Rocha, Taina Venas, Elisa Cavalcante Pereira, Renata Serrano Lopes, Andrea Cony Cavalcanti, Marilda Siqueira on behalf of the Fiocruz COVID-19 Genomic Surveillance Network |
| EPI_ISL_2466257 | Laboratorio Central de Saude Publica do Estado de Alagoas (LACEN/AL) | Laboratory of Respiratory Viruses and Measles, Oswaldo Cruz Institute, FIOCRUZ | Paola Resende, Luciana Appolinario, Fernando Motta, Anna Carolina Paixao, Ana Carolina Mendonca, Alice Sampaio Rocha, Taina Venas, Elisa Cavalcante Pereira, Renata Serrano Lopes, Anderson Brandao Leite, Marilda Siqueira on behalf of the Fiocruz COVID-19 Genomic Surveillance Network |
| EPI_ISL_2466264 | Laboratorio Central de Saude Publica do Estado da Bahia (LACEN/BA) | Laboratory of Respiratory Viruses and Measles, Oswaldo Cruz Institute, FIOCRUZ | Paola Resende, Luciana Appolinario, Fernando Motta, Anna Carolina Paixao, Ana Carolina Mendonca, Alice Sampaio Rocha, Taina Venas, Elisa Cavalcante Pereira, Renata Serrano Lopes,Felicidade Pereira, Marilda Siqueira on behalf of the Fiocruz COVID-19 Genomic Surveillance Network |
| EPI_ISL_2466268 | Laboratorio Central de Saude Publica do Estado do Rio de Janeiro (LACEN-RJ) | Laboratory of Respiratory Viruses and Measles, Oswaldo Cruz Institute, FIOCRUZ | Paola Resende, Luciana Appolinario, Fernando Motta, Anna Carolina Paixao, Ana Carolina Mendonca, Alice Sampaio Rocha, Taina Venas, Elisa Cavalcante Pereira, Renata Serrano Lopes, Andrea Cony Cavalcanti, Marilda Siqueira on behalf of the Fiocruz COVID-19 Genomic Surveillance Network |
| EPI_ISL_2466447, EPI_ISL_2466459 | HLAGYN - Laboratorio de Imunologia de Transplantes de Goias | HLAGYN - Laboratorio de Imunologia de Transplantes de Goias | Fernando Antonio Vinhal dos Santos, Erika Lopes Rocha Batista, Alessandro Leonardo Alvares Magalhaes, Frederico Rodrigues Vinhal, Sabrina Sara Moreira Duarte, Lucas Carlos Gomes Pereira, Daniel Ferreira de Sousa |
| EPI_ISL_2473714 | UBS PASTOR MARCELINO DEUNGARO RIO GRANDE | Instituto Butantan | Dimas Tadeu Covas, Antonio Jorge Martins, Claudia Renata dos Santos Barros, David Schlesinger, Debora Botequio Moretti, Elaine Cristina Marqueze, Elaine Vieira Santos, Evandra Strazza Rodrigues, Heidge Fukumasu, Jayme Augusto de Souza-Neto, José Salvatore Leister Patané, Luiz Alcantara, Luiz Lehmann Coutinho, Maria Carolina Elias, Maurício Lacerda Nogueira, Rafael dos Santos Bezerra, Raul Machado Neto, Rejane Maria Tommasini Grotto, Ricardo Haddad, Sandra Coccuzzo Sampaio Vessoni, Simone Kashima, Svetoslav Nanev Slavov, Vincent Louis Viala |
| EPI_ISL_2473734 | HOSP GERAL DE SAO PAULO | Instituto Butantan | Dimas Tadeu Covas, Antonio Jorge Martins, Claudia Renata dos Santos Barros, David Schlesinger, Debora Botequio Moretti, Elaine Cristina Marqueze, Elaine Vieira Santos, Evandra Strazza Rodrigues, Heidge Fukumasu, Jayme Augusto de Souza-Neto, José Salvatore Leister Patané, Luiz Alcantara, Luiz Lehmann Coutinho, Maria Carolina Elias, Maurício Lacerda Nogueira, Rafael dos Santos Bezerra, Raul Machado Neto, Rejane Maria Tommasini Grotto, Ricardo Haddad, Sandra Coccuzzo Sampaio Vessoni, Simone Kashima, Svetoslav Nanev Slavov, Vincent Louis Viala |
| EPI_ISL_2473786 | CADIP VILA REGINA FERNANDOPOLIS | Instituto Butantan | Dimas Tadeu Covas, Antonio Jorge Martins, Claudia Renata dos Santos Barros, David Schlesinger, Debora Botequio Moretti, Elaine Cristina Marqueze, Elaine Vieira Santos, Evandra Strazza Rodrigues, Heidge Fukumasu, Jayme Augusto de Souza-Neto, José Salvatore Leister Patané, Luiz Alcantara, Luiz Lehmann Coutinho, Maria Carolina Elias, Maurício Lacerda Nogueira, Rafael dos Santos Bezerra, Raul Machado Neto, Rejane Maria Tommasini Grotto, Ricardo Haddad, Sandra Coccuzzo Sampaio Vessoni, Simone Kashima, Svetoslav Nanev Slavov, Vincent Louis Viala |
| EPI_ISL_2493388 | SECRETARIA DE SAUDE DE SAO PEDRO | Instituto Butantan | Dimas Tadeu Covas, Antonio Jorge Martins, Claudia Renata dos Santos Barros, David Schlesinger, Debora Botequio Moretti, Elaine Cristina Marqueze, Elaine Vieira Santos, Evandra Strazza Rodrigues, Heidge Fukumasu, Jayme Augusto de Souza-Neto, José Salvatore Leister Patané, Luiz Alcantara, Luiz Lehmann Coutinho, Maria Carolina Elias, Maurício Lacerda Nogueira, Rafael dos Santos Bezerra, Raul Machado Neto, Rejane Maria Tommasini Grotto, Ricardo Haddad, Sandra Coccuzzo Sampaio Vessoni, Simone Kashima, Svetoslav Nanev Slavov, Vincent Louis Viala |
| EPI_ISL_2493750 | AFIP | Instituto Butantan | Dimas Tadeu Covas, Antonio Jorge Martins, Claudia Renata dos Santos Barros, David Schlesinger, Debora Botequio Moretti, Elaine Cristina Marqueze, Elaine Vieira Santos, Evandra Strazza Rodrigues, Heidge Fukumasu, Jayme Augusto de Souza-Neto, José Salvatore Leister Patané, Luiz Alcantara, Luiz Lehmann Coutinho, Maria Carolina Elias, Maurício Lacerda Nogueira, Rafael dos Santos Bezerra, Raul Machado Neto, Rejane Maria Tommasini Grotto, Ricardo Haddad, Sandra Coccuzzo Sampaio Vessoni, Simone Kashima, Svetoslav Nanev Slavov, Vincent Louis Viala |
| EPI_ISL_2493820 | UBS JARDIM CAIUBY | Instituto Butantan | Dimas Tadeu Covas, Antonio Jorge Martins, Claudia Renata dos Santos Barros, David Schlesinger, Debora Botequio Moretti, Elaine Cristina Marqueze, Elaine Vieira Santos, Evandra Strazza Rodrigues, Heidge Fukumasu, Jayme Augusto de Souza-Neto, José Salvatore Leister Patané, Luiz Alcantara, Luiz Lehmann Coutinho, Maria Carolina Elias, Maurício Lacerda Nogueira, Rafael dos Santos Bezerra, Raul Machado Neto, Rejane Maria Tommasini Grotto, Ricardo Haddad, Sandra Coccuzzo Sampaio Vessoni, Simone Kashima, Svetoslav Nanev Slavov, Vincent Louis Viala |
| EPI_ISL_2493831 | USAFA SAO JORGE | Instituto Butantan | Dimas Tadeu Covas, Antonio Jorge Martins, Claudia Renata dos Santos Barros, David Schlesinger, Debora Botequio Moretti, Elaine Cristina Marqueze, Elaine Vieira Santos, Evandra Strazza Rodrigues, Heidge Fukumasu, Jayme Augusto de Souza-Neto, José Salvatore Leister Patané, Luiz Alcantara, Luiz Lehmann Coutinho, Maria Carolina Elias, Maurício Lacerda Nogueira, Rafael dos Santos Bezerra, Raul Machado Neto, Rejane Maria Tommasini Grotto, Ricardo Haddad, Sandra Coccuzzo Sampaio Vessoni, Simone Kashima, Svetoslav Nanev Slavov, Vincent Louis Viala |
| EPI_ISL_2493891 | PRONTO ATENDIMENTO DE VARGEM GRANDE PAULISTA | Instituto Butantan | Dimas Tadeu Covas, Antonio Jorge Martins, Claudia Renata dos Santos Barros, David Schlesinger, Debora Botequio Moretti, Elaine Cristina Marqueze, Elaine Vieira Santos, Evandra Strazza Rodrigues, Heidge Fukumasu, Jayme Augusto de Souza-Neto, José Salvatore Leister Patané, Luiz Alcantara, Luiz Lehmann Coutinho, Maria Carolina Elias, Maurício Lacerda Nogueira, Rafael dos Santos Bezerra, Raul Machado Neto, Rejane Maria Tommasini Grotto, Ricardo Haddad, Sandra Coccuzzo Sampaio Vessoni, Simone Kashima, Svetoslav Nanev Slavov, Vincent Louis Viala |
| EPI_ISL_2494003 | INSIDE CRSN PIRITUBA | Instituto Butantan | Dimas Tadeu Covas, Antonio Jorge Martins, Claudia Renata dos Santos Barros, David Schlesinger, Debora Botequio Moretti, Elaine Cristina Marqueze, Elaine Vieira Santos, Evandra Strazza Rodrigues, Heidge Fukumasu, Jayme Augusto de Souza-Neto, José Salvatore Leister Patané, Luiz Alcantara, Luiz Lehmann Coutinho, Maria Carolina Elias, Maurício Lacerda Nogueira, Rafael dos Santos Bezerra, Raul Machado Neto, Rejane Maria Tommasini Grotto, Ricardo Haddad, Sandra Coccuzzo Sampaio Vessoni, Simone Kashima, Svetoslav Nanev Slavov, Vincent Louis Viala |
| EPI_ISL_2494025 | INSIDE CRSSU IPIRANGA | Instituto Butantan | Dimas Tadeu Covas, Antonio Jorge Martins, Claudia Renata dos Santos Barros, David Schlesinger, Debora Botequio Moretti, Elaine Cristina Marqueze, Elaine Vieira Santos, Evandra Strazza Rodrigues, Heidge Fukumasu, Jayme Augusto de Souza-Neto, José Salvatore Leister Patané, Luiz Alcantara, Luiz Lehmann Coutinho, Maria Carolina Elias, Maurício Lacerda Nogueira, Rafael dos Santos Bezerra, Raul Machado Neto, Rejane Maria Tommasini Grotto, Ricardo Haddad, Sandra Coccuzzo Sampaio Vessoni, Simone Kashima, Svetoslav Nanev Slavov, Vincent Louis Viala |
| EPI_ISL_2494091 | AFIP SUL | Instituto Butantan | Dimas Tadeu Covas, Antonio Jorge Martins, Claudia Renata dos Santos Barros, David Schlesinger, Debora Botequio Moretti, Elaine Cristina Marqueze, Elaine Vieira Santos, Evandra Strazza Rodrigues, Heidge Fukumasu, Jayme Augusto de Souza-Neto, José Salvatore Leister Patané, Luiz Alcantara, Luiz Lehmann Coutinho, Maria Carolina Elias, Maurício Lacerda Nogueira, Rafael dos Santos Bezerra, Raul Machado Neto, Rejane Maria Tommasini Grotto, Ricardo Haddad, Sandra Coccuzzo Sampaio Vessoni, Simone Kashima, Svetoslav Nanev Slavov, Vincent Louis Viala |
| EPI_ISL_2494197 | VIGILANCIA EPIDEMIOLOGICA E CONTROLE DE VETORES PIRASSUNUN | Instituto Butantan | Dimas Tadeu Covas, Antonio Jorge Martins, Claudia Renata dos Santos Barros, David Schlesinger, Debora Botequio Moretti, Elaine Cristina Marqueze, Elaine Vieira Santos, Evandra Strazza Rodrigues, Heidge Fukumasu, Jayme Augusto de Souza-Neto, José Salvatore Leister Patané, Luiz Alcantara, Luiz |

| | | | |
|---|---|---|---|
| | | | Lehmann Coutinho, Maria Carolina Elias, Maurício Lacerda Nogueira, Rafael dos Santos Bezerra, Raul Machado Neto, Rejane Maria Tommasini Grotto, Ricardo Haddad, Sandra Coccuzzo Sampaio Vessoni, Simone Kashima, Svetoslav Nanev Slavov, Vincent Louis Viala |
| EPI_ISL_2494225 | BIOFAST LESTE | Instituto Butantan | Dimas Tadeu Covas, Antonio Jorge Martins, Claudia Renata dos Santos Barros, David Schlesinger, Debora Botequio Moretti, Elaine Cristina Marqueze, Elaine Vieira Santos, Evandra Strazza Rodrigues, Heidge Fukumasu, Jayme Augusto de Souza-Neto, José Salvatore Leister Patané, Luiz Alcantara, Luiz Lehmann Coutinho, Maria Carolina Elias, Maurício Lacerda Nogueira, Rafael dos Santos Bezerra, Raul Machado Neto, Rejane Maria Tommasini Grotto, Ricardo Haddad, Sandra Coccuzzo Sampaio Vessoni, Simone Kashima, Svetoslav Nanev Slavov, Vincent Louis Viala |
| EPI_ISL_2497447, EPI_ISL_2497463 | HLAGYN - Laboratorio de Imunologia de Transplantes de Goias | HLAGYN - Laboratorio de Imunologia de Transplantes de Goias | Fernando Antonio Vinhal dos Santos, Erika Lopes Rocha Batista, Alessandro Leonardo Alvares Magalhaes, Frederico Rodrigues Vinhal, Sabrina Sara Moreira Duarte, Lucas Carlos Gomes Pereira, Daniel Ferreira de Sousa |
| EPI_ISL_2534862, EPI_ISL_2534863, EPI_ISL_2534864, EPI_ISL_2534865, EPI_ISL_2534866, EPI_ISL_2534867, EPI_ISL_2534868, EPI_ISL_2534869, EPI_ISL_2534870, EPI_ISL_2534871, EPI_ISL_2534872, EPI_ISL_2534873, EPI_ISL_2534874, EPI_ISL_2534875, EPI_ISL_2534876, EPI_ISL_2534877, EPI_ISL_2534878, EPI_ISL_2534879, EPI_ISL_2534880, EPI_ISL_2534881, EPI_ISL_2534882, EPI_ISL_2534883, EPI_ISL_2534884, EPI_ISL_2534885, EPI_ISL_2534886, EPI_ISL_2534887, EPI_ISL_2534888, EPI_ISL_2534889, EPI_ISL_2534890, EPI_ISL_2534891, EPI_ISL_2534892, EPI_ISL_2534893, EPI_ISL_2534894, EPI_ISL_2534895, EPI_ISL_2534896, EPI_ISL_2534897, EPI_ISL_2534898, EPI_ISL_2534899, EPI_ISL_2534900, EPI_ISL_2534901, EPI_ISL_2534902, EPI_ISL_2534903, EPI_ISL_2534904, EPI_ISL_2534905, EPI_ISL_2534906, EPI_ISL_2534907, EPI_ISL_2534908, EPI_ISL_2534909, EPI_ISL_2534910, EPI_ISL_2534911, EPI_ISL_2534912, EPI_ISL_2534913, EPI_ISL_2534914, EPI_ISL_2534915, EPI_ISL_2534916, EPI_ISL_2534917, EPI_ISL_2534918, EPI_ISL_2534919, EPI_ISL_2534920, EPI_ISL_2534921, EPI_ISL_2534922, EPI_ISL_2534924, EPI_ISL_2534925, EPI_ISL_2534926, EPI_ISL_2534927, EPI_ISL_2534928, EPI_ISL_2534929, EPI_ISL_2534930, EPI_ISL_2534931, EPI_ISL_2534933, EPI_ISL_2534934, EPI_ISL_2534935, EPI_ISL_2534936, EPI_ISL_2534937, EPI_ISL_2534938, EPI_ISL_2534939, EPI_ISL_2534940, EPI_ISL_2534941, EPI_ISL_2534942, EPI_ISL_2534943, EPI_ISL_2534944, EPI_ISL_2534946, EPI_ISL_2534947, EPI_ISL_2534948, EPI_ISL_2534949, EPI_ISL_2534950, EPI_ISL_2534951, EPI_ISL_2534952, EPI_ISL_2534955, EPI_ISL_2534956, EPI_ISL_2534957, EPI_ISL_2534958, EPI_ISL_2534959, EPI_ISL_2534960, EPI_ISL_2534961, EPI_ISL_2534962, EPI_ISL_2534963, EPI_ISL_2534964, EPI_ISL_2534965, EPI_ISL_2534966, EPI_ISL_2534967, EPI_ISL_2534968, EPI_ISL_2534969, EPI_ISL_2534970, EPI_ISL_2534971, EPI_ISL_2534972, EPI_ISL_2534973, EPI_ISL_2534974, EPI_ISL_2534975, EPI_ISL_2534976, EPI_ISL_2534977, EPI_ISL_2534978, EPI_ISL_2534979, EPI_ISL_2534980, EPI_ISL_2534981, EPI_ISL_2534982, EPI_ISL_2534983, EPI_ISL_2534984, EPI_ISL_2534985, EPI_ISL_2534986, EPI_ISL_2534987, EPI_ISL_2534988, EPI_ISL_2534989, EPI_ISL_2534990, EPI_ISL_2534991, EPI_ISL_2534992, EPI_ISL_2534993, EPI_ISL_2534994, EPI_ISL_2534996, EPI_ISL_2534997, EPI_ISL_2534999, EPI_ISL_2535000, EPI_ISL_2535001, EPI_ISL_2535002, EPI_ISL_2535003, EPI_ISL_2535004, EPI_ISL_2535005, EPI_ISL_2535006, EPI_ISL_2535007, EPI_ISL_2535009, EPI_ISL_2535010, EPI_ISL_2535011, EPI_ISL_2535012, EPI_ISL_2535013, EPI_ISL_2535014, EPI_ISL_2535015, EPI_ISL_2535016, EPI_ISL_2535018, EPI_ISL_2535019, EPI_ISL_2535021, EPI_ISL_2535022, EPI_ISL_2535023, EPI_ISL_2535024, EPI_ISL_2535025, EPI_ISL_2535026, EPI_ISL_2535027, EPI_ISL_2535028, EPI_ISL_2535029, EPI_ISL_2535030, EPI_ISL_2535031, EPI_ISL_2535032, EPI_ISL_2535033, EPI_ISL_2535034, EPI_ISL_2535035, EPI_ISL_2535036, EPI_ISL_2535037, EPI_ISL_2535038, EPI_ISL_2535039, EPI_ISL_2535040, EPI_ISL_2535041, EPI_ISL_2535043, EPI_ISL_2535044, EPI_ISL_2535045, EPI_ISL_2535046, EPI_ISL_2535047, EPI_ISL_2535048, EPI_ISL_2535049, EPI_ISL_2535050, EPI_ISL_2535051, EPI_ISL_2535052, EPI_ISL_2535053, EPI_ISL_2535054, EPI_ISL_2535055, EPI_ISL_2535056, EPI_ISL_2535057, EPI_ISL_2535058, EPI_ISL_2535059, EPI_ISL_2535060, EPI_ISL_2535061, EPI_ISL_2535062, EPI_ISL_2535063, EPI_ISL_2535064, EPI_ISL_2535065, EPI_ISL_2535066, EPI_ISL_2535067, EPI_ISL_2535068, EPI_ISL_2535069, EPI_ISL_2535070, EPI_ISL_2535071, EPI_ISL_2535072, EPI_ISL_2535073, EPI_ISL_2535074, EPI_ISL_2535075, EPI_ISL_2535076, EPI_ISL_2535077, EPI_ISL_2535078, EPI_ISL_2535080, EPI_ISL_2535081, EPI_ISL_2535082, EPI_ISL_2535083, EPI_ISL_2535084, EPI_ISL_2535085, EPI_ISL_2535086, EPI_ISL_2535087, EPI_ISL_2535088, EPI_ISL_2535089, EPI_ISL_2535090, EPI_ISL_2535091, EPI_ISL_2535092, EPI_ISL_2535093, EPI_ISL_2535094, EPI_ISL_2535095, EPI_ISL_2535096, EPI_ISL_2535097, EPI_ISL_2535098, EPI_ISL_2535099, EPI_ISL_2535100, EPI_ISL_2535101, EPI_ISL_2535102, EPI_ISL_2535103, EPI_ISL_2535104, EPI_ISL_2535105, EPI_ISL_2535106, EPI_ISL_2535107, EPI_ISL_2535108, EPI_ISL_2535109, EPI_ISL_2535110 | | | |
| see above | Laboratorio Central Noel Nutels | Bioinformatics Laboratory / LNCC | Luiz G P de Almeida, Alessandra P Lamarca, Ronaldo da Silva F Jr, Liliane Cavalcante, Alexandra L Gerber, Ana Paula de C Guimaraes, Douglas Terra Machado, Cassia Alves, Diana Mariani, Cintia Policarpo, Gleidson da Silva de Oliveira, Mario Sergio Ribeiro, Silvia Carvalho, Flavio Dias da Silva, Marcio Henrique de Oliveira Garcia, Leandro Magalhaes de Souza, Cristiane Gomes da Silva, Caio Luiz Pereira Ribeiro, Andrea Cony Cavalcanti, Claudia Maria Braga de Mello, Amilcar Tanuri, Ana Tereza R Vasconcelos |
| EPI_ISL_2535111, EPI_ISL_2535112, EPI_ISL_2535113, EPI_ISL_2535114, EPI_ISL_2535115, EPI_ISL_2535117, EPI_ISL_2535118, EPI_ISL_2535119, EPI_ISL_2535120, EPI_ISL_2535121, EPI_ISL_2535123, EPI_ISL_2535125, EPI_ISL_2535126, EPI_ISL_2535127, EPI_ISL_2535128, EPI_ISL_2535131, EPI_ISL_2535132, EPI_ISL_2535133, EPI_ISL_2535134, EPI_ISL_2535135, EPI_ISL_2535136, EPI_ISL_2535137, EPI_ISL_2535138, EPI_ISL_2535140, EPI_ISL_2535141, EPI_ISL_2535142, EPI_ISL_2535143, EPI_ISL_2535144, EPI_ISL_2535145, EPI_ISL_2535146, EPI_ISL_2535147, EPI_ISL_2535148, EPI_ISL_2535149, EPI_ISL_2535151, EPI_ISL_2535152, EPI_ISL_2535153, EPI_ISL_2535154, EPI_ISL_2535155, EPI_ISL_2535156, EPI_ISL_2535157, EPI_ISL_2535158, EPI_ISL_2535160, EPI_ISL_2535161, EPI_ISL_2535162, EPI_ISL_2535163, EPI_ISL_2535164, EPI_ISL_2535165, EPI_ISL_2535166, EPI_ISL_2535167, EPI_ISL_2535168, EPI_ISL_2535169, EPI_ISL_2535170, EPI_ISL_2535171, EPI_ISL_2535172, EPI_ISL_2535173, EPI_ISL_2535174, EPI_ISL_2535175, EPI_ISL_2535176, EPI_ISL_2535177, EPI_ISL_2535178, EPI_ISL_2535179, EPI_ISL_2535180, EPI_ISL_2535181, EPI_ISL_2535182, EPI_ISL_2535184, EPI_ISL_2535185, EPI_ISL_2535187, EPI_ISL_2535188, EPI_ISL_2535189, EPI_ISL_2535190, EPI_ISL_2535191, EPI_ISL_2535192, EPI_ISL_2535193, EPI_ISL_2535194, EPI_ISL_2535195, EPI_ISL_2535196, EPI_ISL_2535197, EPI_ISL_2535198, EPI_ISL_2535199, EPI_ISL_2535200, EPI_ISL_2535201, EPI_ISL_2535202, EPI_ISL_2535203, EPI_ISL_2535204, EPI_ISL_2535205, EPI_ISL_2535206, EPI_ISL_2535207, EPI_ISL_2535208, EPI_ISL_2535209, EPI_ISL_2535210, EPI_ISL_2535211, EPI_ISL_2535212, EPI_ISL_2535213, EPI_ISL_2535214, EPI_ISL_2535216, EPI_ISL_2535217, EPI_ISL_2535218, EPI_ISL_2535219, EPI_ISL_2535220, EPI_ISL_2535221, EPI_ISL_2535222, EPI_ISL_2535224, EPI_ISL_2535225, EPI_ISL_2535226, EPI_ISL_2535227, EPI_ISL_2535228, EPI_ISL_2535229, EPI_ISL_2535230, EPI_ISL_2535231, EPI_ISL_2535232, EPI_ISL_2535233, EPI_ISL_2535234, EPI_ISL_2535235 | | | |
| see above | Unidade de apoio ao diagnostico da COVID - UNADIG | Bioinformatics Laboratory / LNCC | Luiz G P de Almeida, Alessandra P Lamarca, Ronaldo da Silva F Jr, Liliane Cavalcante, Alexandra L Gerber, Ana Paula de C Guimaraes, Douglas Terra Machado, Cassia Alves, Diana Mariani, Cintia Policarpo, Gleidson da Silva de Oliveira, Mario Sergio Ribeiro, Silvia Carvalho, Flavio Dias da Silva, Marcio Henrique de Oliveira Garcia, Leandro Magalhaes de Souza, Cristiane Gomes da Silva, Caio Luiz Pereira Ribeiro, Andrea Cony Cavalcanti, Claudia Maria Braga de Mello, Amilcar Tanuri, Ana Tereza R Vasconcelos |
| EPI_ISL_2536228 | Labortorio Central de Saude Publica do Estado de Santa Catarina (LACEN/SC) | Laboratory of Respiratory Viruses and Measles, Oswaldo Cruz Institute, FIOCRUZ | Paola Resende, Luciana Appolinario, Fernando Motta, Anna Carolina Paixao, Ana Carolina Mendonca, Alice Sampaio Rocha, Taina Venas, Elisa Cavalcante Pereira, Renata Serrano Lopes, Darcita Buerger Rovaris, Sandra Bianchini Fernandes, Marilda Siqueira on behalf of the Fiocruz COVID-19 Genomic Surveillance Network |
| EPI_ISL_2536269, EPI_ISL_2536270, EPI_ISL_2536271, EPI_ISL_2536272, EPI_ISL_2536273, EPI_ISL_2536274 | Laboratory of Respiratory Viruses and Measles, Oswaldo Cruz Institute, FIOCRUZ | Laboratory of Respiratory Viruses and Measles, Oswaldo Cruz Institute, FIOCRUZ | Paola Resende, Patricia Brasil, Luciana Appolinario, Fernando Motta, Anna Carolina Paixao, Ana Carolina Mendonca, Alice Sampaio Rocha, Taina Venas, Elisa Cavalcante Pereira, Renata Serrano Lopes, Marilda Siqueira on behalf of the Fiocruz COVID-19 Genomic Surveillance Network |
| EPI_ISL_2536275, EPI_ISL_2536276, EPI_ISL_2536277, EPI_ISL_2536278, EPI_ISL_2536279, EPI_ISL_2536281, EPI_ISL_2536282, EPI_ISL_2536283, EPI_ISL_2536284, EPI_ISL_2536285, EPI_ISL_2536286, EPI_ISL_2536287, EPI_ISL_2536288, EPI_ISL_2536289, EPI_ISL_2536290, EPI_ISL_2536291, EPI_ISL_2536292, EPI_ISL_2536293, EPI_ISL_2536294, EPI_ISL_2536295, EPI_ISL_2536296, EPI_ISL_2536297, EPI_ISL_2536298, EPI_ISL_2536299, EPI_ISL_2536300, EPI_ISL_2536301, EPI_ISL_2536302 | | | |
| see above | Labortorio Central de Saude Publica do Estado do Rio de Janeiro (LACEN/RJ) | Laboratory of Respiratory Viruses and Measles, Oswaldo Cruz Institute, FIOCRUZ | Paola Resende, Luciana Appolinario, Fernando Motta, Anna Carolina Paixao, Ana Carolina Mendonca, Alice Sampaio Rocha, Taina Venas, Elisa Cavalcante Pereira, Renata Serrano Lopes, Andrea Cony Cavalcanti, Marilda Siqueira on behalf of the Fiocruz COVID-19 Genomic Surveillance Network |
| EPI_ISL_2543484 | Genetica Molecular and Subdepartamento de Virologia ISP Chile | Instituto de Salud Publica de Chile | Karen Orostica, Constanza Campano, Barbara Parra, Loredana Arata, Gisselle Barra, Patricia Bustos, Rodrigo Fasce, Javier Tognarelli, Andres Castillo, Soledad Ulloa, Jorge Fernandez |
| EPI_ISL_2557376, EPI_ISL_2557377, EPI_ISL_2557378, EPI_ISL_2557379, EPI_ISL_2557380, EPI_ISL_2557381, EPI_ISL_2557382, EPI_ISL_2557383, EPI_ISL_2557384 | Labortorio Central de Saude Publica do Estado do Rio de Janeiro (LACEN/RJ) | Laboratory of Respiratory Viruses and Measles, Oswaldo Cruz Institute, FIOCRUZ | Paola Resende, Luciana Appolinario, Fernando Motta, Anna Carolina Paixao, Ana Carolina Mendonca, Alice Sampaio Rocha, Taina Venas, Elisa Cavalcante Pereira, Renata Serrano Lopes, Andrea Cony Cavalcanti, Marilda Siqueira on behalf of the Fiocruz COVID-19 Genomic Surveillance Network |
| EPI_ISL_2557385, EPI_ISL_2557386, EPI_ISL_2557387, EPI_ISL_2557388, EPI_ISL_2557389, EPI_ISL_2557390, EPI_ISL_2557391, EPI_ISL_2557392, EPI_ISL_2557393, EPI_ISL_2557394, EPI_ISL_2557395, EPI_ISL_2557396, EPI_ISL_2557397, EPI_ISL_2557398, EPI_ISL_2557399, EPI_ISL_2557400, EPI_ISL_2557401, EPI_ISL_2557402, EPI_ISL_2557403 | | | |
| see above | Laboratory of Respiratory Viruses and Measles, Oswaldo Cruz Institute, FIOCRUZ | Laboratory of Respiratory Viruses and Measles, Oswaldo Cruz Institute, FIOCRUZ | Paola Resende, Luciana Appolinario, Fernando Motta, Anna Carolina Paixao, Ana Carolina Mendonca, Alice Sampaio Rocha, Taina Venas, Elisa Cavalcante Pereira, Renata Serrano Lopes, Marilda Siqueira on behalf of the Fiocruz COVID-19 Genomic Surveillance Network |
| EPI_ISL_2557404 | Labortorio Central de Saude Publica do Estado do Rio de Janeiro (LACEN/RJ) | Laboratory of Respiratory Viruses and Measles, Oswaldo Cruz Institute, FIOCRUZ | Paola Resende, Luciana Appolinario, Fernando Motta, Anna Carolina Paixao, Ana Carolina Mendonca, Alice Sampaio Rocha, Taina Venas, Elisa Cavalcante Pereira, Renata Serrano Lopes, Andrea Cony Cavalcanti, Marilda Siqueira on behalf of the Fiocruz COVID-19 Genomic Surveillance Network |
| EPI_ISL_2557405 | Laboratory of Respiratory Viruses and Measles, Oswaldo Cruz Institute, FIOCRUZ | Laboratory of Respiratory Viruses and Measles, Oswaldo Cruz Institute, FIOCRUZ | Paola Resende, Luciana Appolinario, Fernando Motta, Anna Carolina Paixao, Ana Carolina Mendonca, Alice Sampaio Rocha, Taina Venas, Elisa Cavalcante Pereira, Renata Serrano Lopes, Marilda Siqueira on behalf of the Fiocruz COVID-19 Genomic Surveillance Network |
| EPI_ISL_2557406 | Labortorio Central de Saude Publica do Estado do Rio de Janeiro (LACEN/RJ) | Laboratory of Respiratory Viruses and Measles, Oswaldo Cruz Institute, FIOCRUZ | Paola Resende, Luciana Appolinario, Fernando Motta, Anna Carolina Paixao, Ana Carolina Mendonca, Alice Sampaio Rocha, Taina Venas, Elisa Cavalcante Pereira, Renata Serrano Lopes, Andrea Cony Cavalcanti, Marilda Siqueira on behalf of the Fiocruz COVID-19 Genomic Surveillance Network |
| EPI_ISL_2557407, EPI_ISL_2557408, EPI_ISL_2557409, EPI_ISL_2557410 | Laboratory of Respiratory Viruses and Measles, Oswaldo Cruz Institute, FIOCRUZ | Laboratory of Respiratory Viruses and Measles, Oswaldo Cruz Institute, FIOCRUZ | Paola Resende, Luciana Appolinario, Fernando Motta, Anna Carolina Paixao, Ana Carolina Mendonca, Alice Sampaio Rocha, Taina Venas, Elisa Cavalcante Pereira, Renata Serrano Lopes, Marilda Siqueira on behalf of the Fiocruz COVID-19 Genomic Surveillance Network |
| EPI_ISL_2601693 | Hospital General Universitario Gregorio Marañón | Hospital General Universitario Gregorio Marañón | Sergio Buenestado Serrano, Pedro Sola Campoy, Laura Pérez-Lago, Cristina Rodriguez-Grande, Marta Herranz Martin, Victor Manuel de la Cueva, Julia Suárez, Pilar Catalán, Patricia Muñoz, Darío García de Viedma |
| EPI_ISL_2603449 | Laboratorio Central de Saude Publica do Estado do Rio Grande do Sul (LACEN-RS) | Laboratory of Respiratory Viruses and Measles, Oswaldo Cruz Institute, FIOCRUZ | Paola Resende, Luciana Appolinario, Fernando Motta, Anna Carolina Paixao, Ana Carolina Mendonca, Alice Sampaio Rocha, Taina Venas, Elisa Cavalcante Pereira, Renata Serrano Lopes, Anderson Brandao Leite, Marilda Siqueira on behalf of the Fiocruz COVID-19 Genomic Surveillance Network |
| EPI_ISL_2603472, EPI_ISL_2603473, EPI_ISL_2603474, EPI_ISL_2603475, EPI_ISL_2603476, EPI_ISL_2603477, EPI_ISL_2603478, EPI_ISL_2603479, EPI_ISL_2603480, EPI_ISL_2603482, EPI_ISL_2603483, EPI_ISL_2603484, EPI_ISL_2603485, EPI_ISL_2603486, EPI_ISL_2603487, EPI_ISL_2603488, EPI_ISL_2603489, EPI_ISL_2603490, EPI_ISL_2603491, EPI_ISL_2603492, EPI_ISL_2603493, EPI_ISL_2603494, EPI_ISL_2603495, EPI_ISL_2603496, EPI_ISL_2603497, EPI_ISL_2603498, EPI_ISL_2603499, EPI_ISL_2603500, EPI_ISL_2603501, EPI_ISL_2603502, EPI_ISL_2603503, EPI_ISL_2603504, EPI_ISL_2603505, EPI_ISL_2603506, EPI_ISL_2603507, EPI_ISL_2603508, EPI_ISL_2603509, EPI_ISL_2603510, EPI_ISL_2603511, EPI_ISL_2603512, EPI_ISL_2603513, EPI_ISL_2603514, EPI_ISL_2603515, EPI_ISL_2603516, EPI_ISL_2603517, EPI_ISL_2603518, EPI_ISL_2603519, EPI_ISL_2603520 | | | |
| see above | Laboratory of Respiratory Viruses and Measles, Oswaldo Cruz Institute, FIOCRUZ | Laboratory of Respiratory Viruses and Measles, Oswaldo Cruz Institute, FIOCRUZ | Paola Resende, Luciana Appolinario, Fernando Motta, Anna Carolina Paixao, Ana Carolina Mendonca, Alice Sampaio Rocha, Taina Venas, Elisa Cavalcante Pereira, Renata Serrano Lopes, Marilda Siqueira on behalf of the Fiocruz COVID-19 Genomic Surveillance Network |
| EPI_ISL_2603539, EPI_ISL_2603549, | Laboratorio Central de Saude Publica do Estado do Rio | Laboratory of Respiratory Viruses and Measles, Oswaldo Cruz | Paola Resende, Luciana Appolinario, Fernando Motta, Anna Carolina Paixao, Ana Carolina Mendonca, Alice Sampaio Rocha, Taina Venas, Elisa |

| | | | |
|---|---|---|---|
| EPI_ISL_2603558, EPI_ISL_2603584 | Grande do Sul (LACEN-RS) | Institute, FIOCRUZ | Cavalcante Pereira, Renata Serrano Lopes, Anderson Brandao Leite, Marilda Siqueira on behalf of the Fiocruz COVID-19 Genomic Surveillance Network |
| EPI_ISL_2614070, EPI_ISL_2614071, EPI_ISL_2614072, EPI_ISL_2614073, EPI_ISL_2614074, EPI_ISL_2614075, EPI_ISL_2614076, EPI_ISL_2614077, EPI_ISL_2614078, EPI_ISL_2614079, EPI_ISL_2614080, EPI_ISL_2614081, EPI_ISL_2614082, EPI_ISL_2614083, EPI_ISL_2614084, EPI_ISL_2614085, EPI_ISL_2614086, EPI_ISL_2614087, EPI_ISL_2614088, EPI_ISL_2614089, EPI_ISL_2614090, EPI_ISL_2614091 | | | |
| see above | Laboratory of Molecular Virology, Federal University of Rio de Janeito, UFRJ | Laboratory of Respiratory Viruses and Measles, Oswaldo Cruz Institute, FIOCRUZ | Paola Resende, Amilcar Tanuri, Luciana Appolinario, Fernando Motta, Anna Carolina Paixao, Ana Carolina Mendonca, Alice Sampaio Rocha, Taina Venas, Elisa Cavalcante Pereira, Renata Serrano Lopes, Marilda Siqueira on behalf of the Fiocruz COVID-19 Genomic Surveillance Network |
| EPI_ISL_2614092, EPI_ISL_2614093, EPI_ISL_2614094, EPI_ISL_2614095, EPI_ISL_2614096, EPI_ISL_2614097, EPI_ISL_2614098, EPI_ISL_2614099, EPI_ISL_2614100, EPI_ISL_2614101, EPI_ISL_2614102, EPI_ISL_2614103, EPI_ISL_2614104, EPI_ISL_2614105, EPI_ISL_2614106, EPI_ISL_2614107, EPI_ISL_2614108, EPI_ISL_2614109, EPI_ISL_2614110, EPI_ISL_2614111, EPI_ISL_2614112, EPI_ISL_2614113, EPI_ISL_2614114 | | | |
| see above | Laboratory of Respiratory Viruses and Measles, Oswaldo Cruz Institute, FIOCRUZ | Laboratory of Respiratory Viruses and Measles, Oswaldo Cruz Institute, FIOCRUZ | Paola Resende, Luciana Appolinario, Fernando Motta, Anna Carolina Paixao, Ana Carolina Mendonca, Alice Sampaio Rocha, Taina Venas, Elisa Cavalcante Pereira, Renata Serrano Lopes, Marilda Siqueira on behalf of the Fiocruz COVID-19 Genomic Surveillance Network |
| EPI_ISL_2614115 | Laboratory of Respiratory Viruses and Measles, Oswaldo Cruz Institute, FIOCRUZ | Laboratory of Respiratory Viruses and Measles, Oswaldo Cruz Institute, FIOCRUZ | Paola Resende, Amilcar Tanuri, Luciana Appolinario, Fernando Motta, Anna Carolina Paixao, Ana Carolina Mendonca, Alice Sampaio Rocha, Taina Venas, Elisa Cavalcante Pereira, Renata Serrano Lopes, Marilda Siqueira on behalf of the Fiocruz COVID-19 Genomic Surveillance Network |
| EPI_ISL_2614116, EPI_ISL_2614117, EPI_ISL_2614118, EPI_ISL_2614119, EPI_ISL_2614120, EPI_ISL_2614121, EPI_ISL_2614122, EPI_ISL_2614123, EPI_ISL_2614124, EPI_ISL_2614125, EPI_ISL_2614126, EPI_ISL_2614127, EPI_ISL_2614128, EPI_ISL_2614129, EPI_ISL_2614130, EPI_ISL_2614131, EPI_ISL_2614132, EPI_ISL_2614133, EPI_ISL_2614134, EPI_ISL_2614135, EPI_ISL_2614136, EPI_ISL_2614137, EPI_ISL_2614138, EPI_ISL_2614139, EPI_ISL_2614140, EPI_ISL_2614141, EPI_ISL_2614142, EPI_ISL_2614143, EPI_ISL_2614144, EPI_ISL_2614145, EPI_ISL_2614146, EPI_ISL_2614147, EPI_ISL_2614148, EPI_ISL_2614149, EPI_ISL_2614150, EPI_ISL_2614151, EPI_ISL_2614152, EPI_ISL_2614153, EPI_ISL_2614154, EPI_ISL_2614155, EPI_ISL_2614156, EPI_ISL_2614157, EPI_ISL_2614158, EPI_ISL_2614159, EPI_ISL_2614160, EPI_ISL_2614161, EPI_ISL_2614162, EPI_ISL_2614163, EPI_ISL_2614164, EPI_ISL_2614165, EPI_ISL_2614166, EPI_ISL_2614167, EPI_ISL_2614168, EPI_ISL_2614169, EPI_ISL_2614170, EPI_ISL_2614171, EPI_ISL_2614172, EPI_ISL_2614173, EPI_ISL_2614174, EPI_ISL_2614175, EPI_ISL_2614176, EPI_ISL_2614177, EPI_ISL_2614178, EPI_ISL_2614181, EPI_ISL_2614182, EPI_ISL_2614183, EPI_ISL_2614184, EPI_ISL_2614185, EPI_ISL_2614186, EPI_ISL_2614187, EPI_ISL_2614188, EPI_ISL_2614189, EPI_ISL_2614190, EPI_ISL_2614191, EPI_ISL_2614192, EPI_ISL_2614193, EPI_ISL_2614194, EPI_ISL_2614195, EPI_ISL_2614196, EPI_ISL_2614197, EPI_ISL_2614198, EPI_ISL_2614199, EPI_ISL_2614200, EPI_ISL_2614201, EPI_ISL_2614202, EPI_ISL_2614203, EPI_ISL_2614204, EPI_ISL_2614205, EPI_ISL_2614206, EPI_ISL_2614207, EPI_ISL_2614209, EPI_ISL_2614210, EPI_ISL_2614211, EPI_ISL_2614212, EPI_ISL_2614213, EPI_ISL_2614214, EPI_ISL_2614215, EPI_ISL_2614216, EPI_ISL_2614217, EPI_ISL_2614218, EPI_ISL_2614219, EPI_ISL_2614220, EPI_ISL_2614221, EPI_ISL_2614222, EPI_ISL_2614223, EPI_ISL_2614224, EPI_ISL_2614225, EPI_ISL_2614226, EPI_ISL_2614227, EPI_ISL_2614228, EPI_ISL_2614229, EPI_ISL_2614230, EPI_ISL_2614231, EPI_ISL_2614232, EPI_ISL_2614233, EPI_ISL_2614234, EPI_ISL_2614235, EPI_ISL_2614236, EPI_ISL_2614237, EPI_ISL_2614238, EPI_ISL_2614239, EPI_ISL_2614240, EPI_ISL_2614241, EPI_ISL_2614242, EPI_ISL_2614243, EPI_ISL_2614244, EPI_ISL_2614245, EPI_ISL_2614246, EPI_ISL_2614247, EPI_ISL_2614248, EPI_ISL_2614249, EPI_ISL_2614250, EPI_ISL_2614251, EPI_ISL_2614252, EPI_ISL_2614253, EPI_ISL_2614254, EPI_ISL_2614255, EPI_ISL_2614256, EPI_ISL_2614257, EPI_ISL_2614258, EPI_ISL_2614259, EPI_ISL_2614260, EPI_ISL_2614261, EPI_ISL_2614262, EPI_ISL_2614263, EPI_ISL_2614264, EPI_ISL_2614265, EPI_ISL_2614266, EPI_ISL_2614267, EPI_ISL_2614268, EPI_ISL_2614269, EPI_ISL_2614270, EPI_ISL_2614271, EPI_ISL_2614272, EPI_ISL_2614273, EPI_ISL_2614274, EPI_ISL_2614275, EPI_ISL_2614276, EPI_ISL_2614277, EPI_ISL_2614278, EPI_ISL_2614279, EPI_ISL_2614280, EPI_ISL_2614281, EPI_ISL_2614282, EPI_ISL_2614283, EPI_ISL_2614284, EPI_ISL_2614285, EPI_ISL_2614286, EPI_ISL_2614287, EPI_ISL_2614288, EPI_ISL_2614289, EPI_ISL_2614290, EPI_ISL_2614291, EPI_ISL_2614292, EPI_ISL_2614293, EPI_ISL_2614294, EPI_ISL_2614295, EPI_ISL_2614296, EPI_ISL_2614297, EPI_ISL_2614298, EPI_ISL_2614299, EPI_ISL_2614300, EPI_ISL_2614301, EPI_ISL_2614302, EPI_ISL_2614303, EPI_ISL_2614304, EPI_ISL_2614305, EPI_ISL_2614306, EPI_ISL_2614307, EPI_ISL_2614308, EPI_ISL_2614309, EPI_ISL_2614310, EPI_ISL_2614311, EPI_ISL_2614312, EPI_ISL_2614313, EPI_ISL_2614314, EPI_ISL_2614315, EPI_ISL_2614316, EPI_ISL_2614317, EPI_ISL_2614318, EPI_ISL_2614319, EPI_ISL_2614320, EPI_ISL_2614321, EPI_ISL_2614322, EPI_ISL_2614323, EPI_ISL_2614324, EPI_ISL_2614325, EPI_ISL_2614326, EPI_ISL_2614327, EPI_ISL_2614328, EPI_ISL_2614329, EPI_ISL_2614330, EPI_ISL_2614331, EPI_ISL_2614332, EPI_ISL_2614333, EPI_ISL_2614334, EPI_ISL_2614335, EPI_ISL_2614336, EPI_ISL_2614337, EPI_ISL_2614338, EPI_ISL_2614339, EPI_ISL_2614340, EPI_ISL_2614341, EPI_ISL_2614342, EPI_ISL_2614343, EPI_ISL_2614344, EPI_ISL_2614345, EPI_ISL_2614346, EPI_ISL_2614347, EPI_ISL_2614348, EPI_ISL_2614349, EPI_ISL_2614350 | | | |
| see above | Laboratory of Respiratory Viruses and Measles, Oswaldo Cruz Institute, FIOCRUZ | Laboratory of Respiratory Viruses and Measles, Oswaldo Cruz Institute, FIOCRUZ | Paola Resende, Luciana Appolinario, Fernando Motta, Anna Carolina Paixao, Ana Carolina Mendonca, Alice Sampaio Rocha, Taina Venas, Elisa Cavalcante Pereira, Renata Serrano Lopes, Marilda Siqueira on behalf of the Fiocruz COVID-19 Genomic Surveillance Network |
| EPI_ISL_2614351, EPI_ISL_2614352, EPI_ISL_2614353, EPI_ISL_2614354, EPI_ISL_2614355, EPI_ISL_2614356, EPI_ISL_2614357, EPI_ISL_2614358, EPI_ISL_2614359, EPI_ISL_2614360, EPI_ISL_2614361, EPI_ISL_2614362, EPI_ISL_2614363, EPI_ISL_2614364, EPI_ISL_2614365, EPI_ISL_2614366, EPI_ISL_2614367, EPI_ISL_2614368, EPI_ISL_2614369, EPI_ISL_2614370, EPI_ISL_2614371, EPI_ISL_2614372, EPI_ISL_2614373, EPI_ISL_2614374, EPI_ISL_2614375, EPI_ISL_2614376, EPI_ISL_2614377, EPI_ISL_2614378, EPI_ISL_2614379, EPI_ISL_2614380 | | | |
| see above | Labortorio Central de Saude Publica do Estado do Rio de Janeiro (LACEN/RJ) | Laboratory of Respiratory Viruses and Measles, Oswaldo Cruz Institute, FIOCRUZ | Paola Resende, Luciana Appolinario, Fernando Motta, Anna Carolina Paixao, Ana Carolina Mendonca, Alice Sampaio Rocha, Taina Venas, Elisa Cavalcante Pereira, Renata Serrano Lopes, Andrea Cony Cavalcanti, Marilda Siqueira on behalf of the Fiocruz COVID-19 Genomic Surveillance Network |
| EPI_ISL_2614381, EPI_ISL_2614382, EPI_ISL_2614383 | Instituto Estadual do Cerebro Paulo Niemeyer | Laboratory of Respiratory Viruses and Measles, Oswaldo Cruz Institute, FIOCRUZ | Paola Resende,Carlos Azevedo, Luciana Appolinario, Fernando Motta, Anna Carolina Paixao, Ana Carolina Mendonca, Alice Sampaio Rocha, Taina Venas, Elisa Cavalcante Pereira, Renata Serrano Lopes, Marilda Siqueira on behalf of the Fiocruz COVID-19 Genomic Surveillance Network |
| EPI_ISL_406798 | General Hospital of Central Theater Command of People's Liberation Army of China | BGI & Institute of Microbiology, Chinese Academy of Sciences & Shandong First Medical University & Shandong Academy of Medical Sciences & General Hospital of Central Theater Command of People's Liberation Army of China | Weijun Chen, Yuhai Bi, Weifeng Shi and Zhenhong Hu |
| EPI_ISL_414045 | Laboratório Central de Saúde Pública Noel Nutels (LACEN-RJ) | Laboratory of Respiratory Viruses and Measles, Oswaldo Cruz Institute, FIOCRUZ | Paola Resende, Alisson Fabri, Joilson Xavier, Sunando Roy, Fernando Motta, Aline Mattos, Milene Miranda, Cristiana Garcia, Braulia Caetano, Maria Ogrzewalska, Jonathan Lopes, Luciana Appolinario, Maria Nóbrega, Marilda SiqueiraFiocruz COVID-19 Genomic Surveillance Network |
| EPI_ISL_427299, EPI_ISL_427300, EPI_ISL_427301, EPI_ISL_427302, EPI_ISL_427303, EPI_ISL_427304 | Laboratory of Respiratory Viruses and Measles, Oswaldo Cruz Institute, FIOCRUZ | Laboratory of Respiratory Viruses and Measles, Oswaldo Cruz Institute, FIOCRUZ | Paola Resende, Fernando Motta, Luciana Appolinario, Sunando Roy, Aline Mattos, Milene Miranda, Cristiana Garcia, Braulia Caetano, Maria Ogrzewalska, Priscila Born, Jonathan Lopes, Marilda Siqueira on behalf of the Fiocruz COVID-19 Genomic Surveillance Network |
| EPI_ISL_456071, EPI_ISL_456072, EPI_ISL_456073, EPI_ISL_456074, EPI_ISL_456075 | Laboratory of Respiratory Viruses and Measles, Oswaldo Cruz Institute, FIOCRUZ | Laboratory of Respiratory Viruses and Measles, Oswaldo Cruz Institute, FIOCRUZ | Paola Resende, Luciana Appolinario, Fernando Motta, Aline Mattos, Milene Miranda, Cristiana Garcia, Braulia Caetano, Maria Ogrzewalska, Jonathan Lopes, Marilda Siqueira on behalf of the Fiocruz COVID-19 Genomic Surveillance Network |
| EPI_ISL_456076, EPI_ISL_456077 | Laboratório Central de Saúde Pública Noel Nutels (LACEN-RJ) | Laboratory of Respiratory Viruses and Measles, Oswaldo Cruz Institute, FIOCRUZ | Paola Resende, Luciana Appolinario, Fernando Motta, Aline Mattos, Milene Miranda, Cristiana Garcia, Braulia Caetano, Maria Ogrzewalska, Jonathan Lopes, Marilda Siqueira on behalf of the Fiocruz COVID-19 Genomic Surveillance Network |
| EPI_ISL_456079, EPI_ISL_456080, EPI_ISL_456081 | Laboratory of Respiratory Viruses and Measles, Oswaldo Cruz Institute, FIOCRUZ | Laboratory of Respiratory Viruses and Measles, Oswaldo Cruz Institute, FIOCRUZ | Paola Resende, Luciana Appolinario, Fernando Motta, Aline Mattos, Milene Miranda, Cristiana Garcia, Braulia Caetano, Maria Ogrzewalska, Jonathan Lopes, Marilda Siqueira on behalf of the Fiocruz COVID-19 Genomic Surveillance Network |
| EPI_ISL_456082, EPI_ISL_456083 | Laboratório Central de Saúde Pública Noel Nutels (LACEN-RJ) | Laboratory of Respiratory Viruses and Measles, Oswaldo Cruz Institute, FIOCRUZ | Paola Resende, Luciana Appolinario, Fernando Motta, Aline Mattos, Milene Miranda, Cristiana Garcia, Braulia Caetano, Maria Ogrzewalska, Jonathan Lopes, Marilda Siqueira on behalf of the Fiocruz COVID-19 Genomic Surveillance Network |
| EPI_ISL_456084, EPI_ISL_456085, EPI_ISL_456086, EPI_ISL_456087 | Laboratory of Respiratory Viruses and Measles, Oswaldo Cruz Institute, FIOCRUZ | Laboratory of Respiratory Viruses and Measles, Oswaldo Cruz Institute, FIOCRUZ | Paola Resende, Luciana Appolinario, Fernando Motta, Aline Mattos, Milene Miranda, Cristiana Garcia, Braulia Caetano, Maria Ogrzewalska, Jonathan Lopes, Marilda Siqueira on behalf of the Fiocruz COVID-19 Genomic Surveillance Network |
| EPI_ISL_456088 | Laboratório Central de Saúde Pública Noel Nutels (LACEN-RJ) | Laboratory of Respiratory Viruses and Measles, Oswaldo Cruz Institute, FIOCRUZ | Paola Resende, Luciana Appolinario, Fernando Motta, Aline Mattos, Milene Miranda, Cristiana Garcia, Braulia Caetano, Maria Ogrzewalska, Jonathan Lopes, Marilda Siqueira on behalf of the Fiocruz COVID-19 Genomic Surveillance Network |
| EPI_ISL_456089, EPI_ISL_456090, EPI_ISL_456091, EPI_ISL_456092, EPI_ISL_456093, EPI_ISL_456094, EPI_ISL_456095, EPI_ISL_456096, EPI_ISL_456097, EPI_ISL_456098, EPI_ISL_456099, EPI_ISL_456100, EPI_ISL_456101, EPI_ISL_456102, EPI_ISL_456103, EPI_ISL_456104, EPI_ISL_456105, EPI_ISL_456106 | | | |
| see above | Laboratory of Respiratory Viruses and Measles, Oswaldo Cruz Institute, FIOCRUZ | Laboratory of Respiratory Viruses and Measles, Oswaldo Cruz Institute, FIOCRUZ | Paola Resende, Luciana Appolinario, Fernando Motta, Aline Mattos, Milene Miranda, Cristiana Garcia, Braulia Caetano, Maria Ogrzewalska, Jonathan Lopes, Marilda Siqueira on behalf of the Fiocruz COVID-19 Genomic Surveillance Network |
| EPI_ISL_467344, EPI_ISL_467345, EPI_ISL_467346, EPI_ISL_467347, EPI_ISL_467348, EPI_ISL_467349, EPI_ISL_467350, EPI_ISL_467351, EPI_ISL_467352, EPI_ISL_467353, EPI_ISL_467354, EPI_ISL_467355, EPI_ISL_467356, EPI_ISL_467357, EPI_ISL_467358, EPI_ISL_467359, EPI_ISL_467360, EPI_ISL_467362, EPI_ISL_467363, EPI_ISL_467364, EPI_ISL_467365, EPI_ISL_467366, EPI_ISL_467367, EPI_ISL_467368, EPI_ISL_467369, EPI_ISL_467370, EPI_ISL_467371 | | | |
| see above | Laboratory of Respiratory Viruses and Measles, Oswaldo Cruz Institute, FIOCRUZ | Laboratory of Respiratory Viruses and Measles, Oswaldo Cruz Institute, FIOCRUZ | Paola Resende, Luciana Appolinario, Fernando Motta, Anna Carolina Paixão, Ana Carolina Mendonça, Aline Mattos, Milene Miranda, Cristiana Garcia, Braulia Caetano, Maria Ogrzewalska, Jonathan Lopes, Marilda Siqueira on behalf of the Fiocruz COVID-19 Genomic Surveillance Network |
| EPI_ISL_483065 | Centro de Desenvolvimento Tecnologico em Saude, Fundacao Oswaldo Cruz | Centro de Desenvolvimento Tecnologico em Saude, Fundacao Oswaldo Cruz | Souza,T.M., Fintelman-Rodrigues,N., De Paula,A.D., Tschoeke,D., Barroso,S.P., Gregorio,M.L., Oliveira,J.S., Saraiva,F.B., Ferreira,M.A., Sacramento,C.Q. |
| EPI_ISL_492032 | Instituto de Biologia do Exército | Laboratório Metabolismo Macromolecular FirminoTorres de Castro, Instituto de Biofísica Carlos Chagas Filho, Universidade Federal do Rio de Janeiro | Bianca Catarina Azevedo Cabral, Aline Rosa Vianna de Souza , Marcos Dornelas-Ribeiro, Tatiana LS Nogueira, Nádia Vaez Gonçalves da Cruz, Caleb GM Santos, Elizabeth Valentin, Marcio da Costa Cipitelli, Virginia Sara Grancieri do Amaral, Rodrigo Soares de Moura Neto, Clarissa Damaso, Rosane Silva |
| EPI_ISL_492033 | Instituto de Biologia do Exército | Laboratório Metabolismo Macromolecular FirminoTorres de Castro, Instituto de Biofísica Carlos Chagas Filho, Universidade Federal do Rio de Janeiro | Bianca Catarina Azevedo Cabral, Aline Rosa Vianna de Souza, Caleb GM Santos, Marcos Dornelas-Ribeiro, Tatiana LS Nogueira, Nádia Vaez Gonçalves da Cruz, Elizabeth Valentin, Marcio da Costa Cipitelli, Virginia Sara Grancieri do Amaral, Rodrigo Soares de Moura Neto, Clarissa Damaso, Rosane Silva |
| EPI_ISL_492034 | Instituto de Biologia do Exército | Laboratório Metabolismo Macromolecular FirminoTorres de Castro, Instituto de Biofísica Carlos Chagas Filho, Universidade Federal do Rio de Janeiro | Bianca Catarina Azevedo Cabral, Aline Rosa Vianna de Souza, Nádia Vaez Gonçalves da Cruz, Caleb GM Santos, Marcos Dornelas-Ribeiro, Tatiana LS Nogueira, Elizabeth Valentin, Marcio da Costa Cipitelli, Virginia Sara Grancieri do Amaral, Rodrigo Soares de Moura Neto, Clarissa Damaso, Rosane Silva |

| | | | |
|---|---|---|---|
| EPI_ISL_492035 | Instituto de Biologia do Exército | Laboratório Metabolismo Macromolecular FirminoTorres de Castro, Instituto de Biofísica Carlos Chagas Filho, Universidade Federal do Rio de Janeiro | Bianca Catarina Azevedo Cabral, Aline Rosa Vianna de Souza, Tatiana LS Nogueira, Nádia Vaez Gonçalves da Cruz, Caleb GM Santos, Marcos Dornelas-Ribeiro, Elizabeth Valentin, Marcio da Costa Cipitelli, Virginia Sara Grancieri do Amaral, Rodrigo Soares de Moura Neto, Clarissa Damaso, Rosane Silva |
| EPI_ISL_492036 | Instituto de Biologia do Exército | Laboratório Metabolismo Macromolecular FirminoTorres de Castro, Instituto de Biofísica Carlos Chagas Filho, Universidade Federal do Rio de Janeiro | Bianca Catarina Azevedo Cabral, Aline Rosa Vianna de Souza , Marcos Dornelas-Ribeiro, Tatiana LS Nogueira, Nádia Vaez Gonçalves da Cruz, Caleb GM Santos, Elizabeth Valentin, Marcio da Costa Cipitelli, Virginia Sara Grancieri do Amaral, Rodrigo Soares de Moura Neto, Clarissa Damaso, Rosane Silva |
| EPI_ISL_492037 | Instituto de Biologia do Exército | Laboratório Metabolismo Macromolecular FirminoTorres de Castro, Instituto de Biofísica Carlos Chagas Filho, Universidade Federal do Rio de Janeiro | Bianca Catarina Azevedo Cabral, Aline Rosa Vianna de Souza, Caleb GM Santos, Marcos Dornelas-Ribeiro, Tatiana LS Nogueira, Nádia Vaez Gonçalves da Cruz, Elizabeth Valentin, Marcio da Costa Cipitelli, Virginia Sara Grancieri do Amaral, Rodrigo Soares de Moura Neto, Clarissa Damaso, Rosane Silva |
| EPI_ISL_492038 | Instituto de Biologia do Exército | Laboratório Metabolismo Macromolecular FirminoTorres de Castro, Instituto de Biofísica Carlos Chagas Filho, Universidade Federal do Rio de Janeiro | Bianca Catarina Azevedo Cabral, Aline Rosa Vianna de Souza, Nádia Vaez Gonçalves da Cruz, Caleb GM Santos, Marcos Dornelas-Ribeiro, Tatiana LS Nogueira, Elizabeth Valentin, Marcio da Costa Cipitelli, Virginia Sara Grancieri do Amaral, Rodrigo Soares de Moura Neto, Clarissa Damaso, Rosane Silva |
| EPI_ISL_492039 | Instituto de Biologia do Exército | Laboratório Metabolismo Macromolecular FirminoTorres de Castro, Instituto de Biofísica Carlos Chagas Filho, Universidade Federal do Rio de Janeiro | Bianca Catarina Azevedo Cabral, Aline Rosa Vianna de Souza, Tatiana LS Nogueira, Nádia Vaez Gonçalves da Cruz, Caleb GM Santos, Marcos Dornelas-Ribeiro, Elizabeth Valentin, Marcio da Costa Cipitelli, Virginia Sara Grancieri do Amaral, Rodrigo Soares de Moura Neto, Clarissa Damaso, Rosane Silva |
| EPI_ISL_492040 | Instituto de Biologia do Exército | Laboratório Metabolismo Macromolecular FirminoTorres de Castro, Instituto de Biofísica Carlos Chagas Filho, Universidade Federal do Rio de Janeiro | Bianca Catarina Azevedo Cabral, Aline Rosa Vianna de Souza , Marcos Dornelas-Ribeiro, Tatiana LS Nogueira, Nádia Vaez Gonçalves da Cruz, Caleb GM Santos, Elizabeth Valentin, Marcio da Costa Cipitelli, Virginia Sara Grancieri do Amaral, Rodrigo Soares de Moura Neto, Clarissa Damaso, Rosane Silva |
| EPI_ISL_492041 | Instituto de Biologia do Exército | Laboratório Metabolismo Macromolecular FirminoTorres de Castro, Instituto de Biofísica Carlos Chagas Filho, Universidade Federal do Rio de Janeiro | Bianca Catarina Azevedo Cabral, Aline Rosa Vianna de Souza, Caleb GM Santos, Marcos Dornelas-Ribeiro, Tatiana LS Nogueira, Nádia Vaez Gonçalves da Cruz, Elizabeth Valentin, Marcio da Costa Cipitelli, Virginia Sara Grancieri do Amaral, Rodrigo Soares de Moura Neto, Clarissa Damaso, Rosane Silva |
| EPI_ISL_492042 | Instituto de Biologia do Exército | Laboratório Metabolismo Macromolecular FirminoTorres de Castro, Instituto de Biofísica Carlos Chagas Filho, Universidade Federal do Rio de Janeiro | Bianca Catarina Azevedo Cabral, Aline Rosa Vianna de Souza, Nádia Vaez Gonçalves da Cruz, Caleb GM Santos, Marcos Dornelas-Ribeiro, Tatiana LS Nogueira, Elizabeth Valentin, Marcio da Costa Cipitelli, Virginia Sara Grancieri do Amaral, Rodrigo Soares de Moura Neto, Clarissa Damaso, Rosane Silva |
| EPI_ISL_492043 | Instituto de Biologia do Exército | Laboratório Metabolismo Macromolecular FirminoTorres de Castro, Instituto de Biofísica Carlos Chagas Filho, Universidade Federal do Rio de Janeiro | Bianca Catarina Azevedo Cabral, Aline Rosa Vianna de Souza, Tatiana LS Nogueira, Nádia Vaez Gonçalves da Cruz, Caleb GM Santos, Marcos Dornelas-Ribeiro, Elizabeth Valentin, Marcio da Costa Cipitelli, Virginia Sara Grancieri do Amaral, Rodrigo Soares de Moura Neto, Clarissa Damaso, Rosane Silva |
| EPI_ISL_492044 | Instituto de Biologia do Exército | Laboratório Metabolismo Macromolecular FirminoTorres de Castro, Instituto de Biofísica Carlos Chagas Filho, Universidade Federal do Rio de Janeiro | Bianca Catarina Azevedo Cabral, Aline Rosa Vianna de Souza , Marcos Dornelas-Ribeiro, Tatiana LS Nogueira, Nádia Vaez Gonçalves da Cruz, Caleb GM Santos, Elizabeth Valentin, Marcio da Costa Cipitelli, Virginia Sara Grancieri do Amaral, Rodrigo Soares de Moura Neto, Clarissa Damaso, Rosane Silva |
| EPI_ISL_492045 | Instituto de Biologia do Exército | Laboratório Metabolismo Macromolecular FirminoTorres de Castro, Instituto de Biofísica Carlos Chagas Filho, Universidade Federal do Rio de Janeiro | Bianca Catarina Azevedo Cabral, Aline Rosa Vianna de Souza, Caleb GM Santos, Marcos Dornelas-Ribeiro, Tatiana LS Nogueira, Nádia Vaez Gonçalves da Cruz, Elizabeth Valentin, Marcio da Costa Cipitelli, Virginia Sara Grancieri do Amaral, Rodrigo Soares de Moura Neto, Clarissa Damaso, Rosane Silva |
| EPI_ISL_492048 | Instituto de Biologia do Exército | Laboratório Metabolismo Macromolecular FirminoTorres de Castro, Instituto de Biofísica Carlos Chagas Filho, Universidade Federal do Rio de Janeiro | Bianca Catarina Azevedo Cabral, Aline Rosa Vianna de Souza , Marcos Dornelas-Ribeiro, Tatiana LS Nogueira, Nádia Vaez Gonçalves da Cruz, Caleb GM Santos, Elizabeth Valentin, Marcio da Costa Cipitelli, Virginia Sara Grancieri do Amaral, Rodrigo Soares de Moura Neto, Clarissa Damaso, Rosane Silva |
| EPI_ISL_509430, EPI_ISL_509431, EPI_ISL_509432, EPI_ISL_509433, EPI_ISL_509434, EPI_ISL_509435 | Centro de Desenvolvimento Tecnologico em Saude, Fundacao Oswaldo Cruz | Centro de Desenvolvimento Tecnologico em Saude, Fundacao Oswaldo Cruz | Souza,T.M., Fintelman-Rodrigues,N., De Paula,A.D., Saraiva,F.B., Ferreira,M.A., Sacramento,C.Q., Medeiros,M.A. |
| EPI_ISL_510536 | Centro de Desenvolvimento Tecnologico em Saude, Fundacao Oswaldo Cruz | Centro de Desenvolvimento Tecnologico em Saude, Fundacao Oswaldo Cruz | Souza,T.M., Fintelman-Rodrigues,N., De Paula,A.D., Saraiva,F.B., Ferreira,M.A., Sacramento,C.Q. and Medeiros,M.A. |
| EPI_ISL_510541 | Centro de Desenvolvimento Tecnologico em Saude, Fundacao Oswaldo Cruz | Centro de Desenvolvimento Tecnologico em Saude, Fundacao Oswaldo Cruz | Souza,T.M., Fintelman-Rodrigues,N., De Paula,A.D., Saraiva,F.B., Ferreira,M.A., Sacramento,C.Q. and Medeiros,M.A. |
| EPI_ISL_513513, EPI_ISL_513514, EPI_ISL_513515, EPI_ISL_513516, EPI_ISL_513518, EPI_ISL_513519, EPI_ISL_513520, EPI_ISL_513521, EPI_ISL_513522, EPI_ISL_513524, EPI_ISL_513525, EPI_ISL_513526, EPI_ISL_513527, EPI_ISL_513529, EPI_ISL_513530, EPI_ISL_513531, EPI_ISL_513532, EPI_ISL_513533, EPI_ISL_513534, EPI_ISL_513535, EPI_ISL_513536, EPI_ISL_513537, EPI_ISL_513538, EPI_ISL_513539, EPI_ISL_513540, EPI_ISL_513541, EPI_ISL_513542, EPI_ISL_513543, EPI_ISL_513544, EPI_ISL_513545, EPI_ISL_513546, EPI_ISL_513547, EPI_ISL_513548, EPI_ISL_513549, EPI_ISL_513550, EPI_ISL_513551, EPI_ISL_513553, EPI_ISL_513556, EPI_ISL_513557, EPI_ISL_513558, EPI_ISL_513559, EPI_ISL_513560, EPI_ISL_513561, EPI_ISL_513562, EPI_ISL_513563, EPI_ISL_513564, EPI_ISL_513567, EPI_ISL_513568, EPI_ISL_513571, EPI_ISL_513572, EPI_ISL_513573, EPI_ISL_513574, EPI_ISL_513575, EPI_ISL_513576, EPI_ISL_513577, EPI_ISL_513578, EPI_ISL_513579, EPI_ISL_513580, EPI_ISL_513581, EPI_ISL_513582, EPI_ISL_513583 | | | |
| see above | Programa de Oncovirologia, Instituto Nacional de Câncer | Programa de Oncovirologia, Instituto Nacional de Câncer | Juliana D. Siqueira, Livia R. Goes, Brunna M. Alves, Claudia Cicala,James Arthos, João P.B. Viola, Andreia C. de Melo, Marcelo A. Soares |
| EPI_ISL_528539 | LVM/UFRJ | LNCC | Gustavo M. Romário M. de Souza; Bruno B. Bezerra; Lucio A. Caldas; Fabio Limonte; Elena Cobos; Sharton V. A. Coelho; Luiz Almeida; Luiza Higga; Isadora A. Correa; Diana Marianni; Luciana B. Arruda; Marcelo Bozza; Orlando Ferreira; Wanderley de Souza; Ana Teresa R. Vasconcelos; Terezinha M. Castineiras; Amilcar Tanuri; Luciana J. Costa. |
| EPI_ISL_528637, EPI_ISL_528638 | LVM/UFRJ | Bioinformatics Laboratory / LNCC | Gustavo D. P. Silva; M. Romário M. de Souza; Bruno B. Bezerra; Lucio A. Caldas; Fabio Limonte; Elena Cobos; Sharton V. A. Coelho; Luiz Almeida; Luiza Higga; Isadora A. Correa; Diana Marianni; Luciana B. Arruda; Marcelo Bozza; Orlando Ferreira; Wanderley de Souza; Ana Teresa R. Vasconcelos; Terezinha M. Castineiras; Amilcar Tanuri; Luciana J. Costa |
| EPI_ISL_529139 | Centro de Desenvolvimento Tecnologico em Saude, Fundacao Oswaldo Cruz | Centro de Desenvolvimento Tecnologico em Saude, Fundacao Oswaldo Cruz | Souza,T.M., Fintelman-Rodrigues,N., De Paula,A.D., Saraiva,F.B., Ferreira,M.A., Sacramento,C.Q., Medeiros,M.A. |
| EPI_ISL_529140 | Centro de Desenvolvimento Tecnologico em Saude, Fundacao Oswaldo Cruz | Centro de Desenvolvimento Tecnologico em Saude, Fundacao Oswaldo Cruz | Souza,T.M., Fintelman-Rodrigues,N., De Paula,A.D., Saraiva,F.B., Ferreira,M.A., Sacramento,C.Q., Medeiros,M.A. |
| EPI_ISL_541347, EPI_ISL_541348, EPI_ISL_541349, EPI_ISL_541350, EPI_ISL_541351 | Laboratory of Respiratory Viruses and Measles, Oswaldo Cruz Institute, FIOCRUZ | Laboratory of Respiratory Viruses and Measles, Oswaldo Cruz Institute, FIOCRUZ | Paola Resende, Luciana Appolinario, Fernando Motta, Anna Carolina Paixão, Ana Carolina Mendonça, Jonathan Lopes, Marilda Siqueira on behalf of the Fiocruz COVID-19 Genomic Surveillance Network |
| EPI_ISL_541352 | Laboratory of Respiratory Viruses and Measles, Oswaldo Cruz Institute, FIOCRUZ | Laboratory of Respiratory Viruses and Measles, Oswaldo Cruz Institute, FIOCRUZ | Paola Resende, Roxana Loayza, Cinthia Avila, Luciana Appolinario, Fernando Motta, Anna Carolina Paixao, Ana Carolina Mendonca, Marilda Siqueira on behalf of the Fiocruz COVID-19 Genomic Surveillance Network |
| EPI_ISL_541353, EPI_ISL_541354, EPI_ISL_541355 | Laboratory of Respiratory Viruses and Measles, Oswaldo Cruz Institute, FIOCRUZ | Laboratory of Respiratory Viruses and Measles, Oswaldo Cruz Institute, FIOCRUZ | Paola Resende, Luciana Appolinario, Fernando Motta, Anna Carolina Paixão, Ana Carolina Mendonça, Jonathan Lopes, Marilda Siqueira on behalf of the Fiocruz COVID-19 Genomic Surveillance Network |
| EPI_ISL_541356 | Laboratory of Respiratory Viruses and Measles, Oswaldo Cruz Institute, FIOCRUZ | Laboratory of Respiratory Viruses and Measles, Oswaldo Cruz Institute, FIOCRUZ | Paola Resende, Roxana Loayza, Cinthia Avila, Luciana Appolinario, Fernando Motta, Anna Carolina Paixao, Ana Carolina Mendonca, Marilda Siqueira on behalf of the Fiocruz COVID-19 Genomic Surveillance Network |
| EPI_ISL_541357, EPI_ISL_541358 | Laboratory of Respiratory Viruses and Measles, Oswaldo Cruz Institute, FIOCRUZ | Laboratory of Respiratory Viruses and Measles, Oswaldo Cruz Institute, FIOCRUZ | Paola Resende, Luciana Appolinario, Fernando Motta, Anna Carolina Paixão, Ana Carolina Mendonça, Jonathan Lopes, Marilda Siqueira on behalf of the Fiocruz COVID-19 Genomic Surveillance Network |
| EPI_ISL_541359, EPI_ISL_541360, EPI_ISL_541361 | Laboratory of Respiratory Viruses and Measles, Oswaldo Cruz Institute, FIOCRUZ | Laboratory of Respiratory Viruses and Measles, Oswaldo Cruz Institute, FIOCRUZ | Paola Resende, Roxana Loayza, Cinthia Avila, Luciana Appolinario, Fernando Motta, Anna Carolina Paixao, Ana Carolina Mendonca, Marilda Siqueira on behalf of the Fiocruz COVID-19 Genomic Surveillance Network |
| EPI_ISL_541362, EPI_ISL_541363 | Laboratory of Respiratory Viruses and Measles, Oswaldo Cruz Institute, FIOCRUZ | Laboratory of Respiratory Viruses and Measles, Oswaldo Cruz Institute, FIOCRUZ | Paola Resende, Luciana Appolinario, Fernando Motta, Anna Carolina Paixão, Ana Carolina Mendonça, Jonathan Lopes, Marilda Siqueira on behalf of the Fiocruz COVID-19 Genomic Surveillance Network |

| | | | |
|---|---|---|---|
| EPI_ISL_541364, EPI_ISL_541365, EPI_ISL_541366 | Laboratory of Respiratory Viruses and Measles, Oswaldo Cruz Institute, FIOCRUZ | Laboratory of Respiratory Viruses and Measles, Oswaldo Cruz Institute, FIOCRUZ | Paola Resende, Roxana Loayza, Cinthia Avila, Luciana Appolinario, Fernando Motta, Anna Carolina Paixao, Ana Carolina Mendonca, Marilda Siqueira on behalf of the Fiocruz COVID-19 Genomic Surveillance Network |
| EPI_ISL_541367 | Laboratory of Respiratory Viruses and Measles, Oswaldo Cruz Institute, FIOCRUZ | Laboratory of Respiratory Viruses and Measles, Oswaldo Cruz Institute, FIOCRUZ | Paola Resende, Luciana Appolinario, Fernando Motta, Anna Carolina Paixão, Ana Carolina Mendonça, Jonathan Lopes, Marilda Siqueira on behalf of the Fiocruz COVID-19 Genomic Surveillance Network |
| EPI_ISL_541368, EPI_ISL_541369 | Laboratory of Respiratory Viruses and Measles, Oswaldo Cruz Institute, FIOCRUZ | Laboratory of Respiratory Viruses and Measles, Oswaldo Cruz Institute, FIOCRUZ | Paola Resende, Roxana Loayza, Cinthia Avila, Luciana Appolinario, Fernando Motta, Anna Carolina Paixao, Ana Carolina Mendonca, Marilda Siqueira on behalf of the Fiocruz COVID-19 Genomic Surveillance Network |
| EPI_ISL_623108, EPI_ISL_623110, EPI_ISL_623112, EPI_ISL_623114, EPI_ISL_623116, EPI_ISL_623118, EPI_ISL_623119, EPI_ISL_623120, EPI_ISL_623124, EPI_ISL_623126, EPI_ISL_623129, EPI_ISL_623130, EPI_ISL_623132, EPI_ISL_623134, EPI_ISL_623136, EPI_ISL_623138, EPI_ISL_623140, EPI_ISL_623142, EPI_ISL_623143, EPI_ISL_623144, EPI_ISL_623145, EPI_ISL_623146, EPI_ISL_623147, EPI_ISL_623148, EPI_ISL_623149, EPI_ISL_623152, EPI_ISL_623154, EPI_ISL_623157, EPI_ISL_623158, EPI_ISL_623160, EPI_ISL_623161, EPI_ISL_623162, EPI_ISL_623164, EPI_ISL_623165, EPI_ISL_623166, EPI_ISL_623168 | | | |
| see above | Laboratorio de Virologia Molecular / UFRJ | Bioinformatics Laboratory / LNCC | Carolina M Voloch, Ronaldo S Francisco Jr, Luiz G P de Almeida, Otavio J. Brustolini, Cynthia C Cardoso, Alexandra L Gerber, Ana Paula de C Guimarães, Diana Mariani, Covid19-UFRJ Workgroup, Luís Cristóvão Pôrto, Renato S Aguiar, Terezinha M P P Castiñeiras, Orlando C. Ferreira, Amilcar Tanuri, Ana Tereza R de Vasconcelos |
| EPI_ISL_636737, EPI_ISL_636834, EPI_ISL_636835, EPI_ISL_636837 | Laboratório de Imunofarmacologia - Instituto Oswaldo Cruz | Laboratório de Imunofarmacologia - Instituto Oswaldo Cruz | Souza,T.M., Fintelman-Rodrigues,N., De Paula,A.D., Saraiva,F.B., Ferreira,M.A. and Sacramento,C.Q. |
| EPI_ISL_717806, EPI_ISL_717807, EPI_ISL_717808 | Laboratorio de Virologia Molecular / UFRJ | Bioinformatics Laboratory / LNCC | Carolina M Voloch, Ronaldo da Silva F Jr, Luiz G P de Almeida, Cynthia C Cardoso, Otavio Bustrolini, Alexandra L Gerber, Ana Paula de C Guimarães, Diana Mariani, Andréa Cony Cavalcanti, Claudia dos Santos Rodrigues, Terezinha M P P Castiñeira, Amilcar Tanuri, Ana Tereza R de Vasconcelos |
| EPI_ISL_717809 | LACEN Dr. Francisco Rimolo Neto | Bioinformatics Laboratory / LNCC | Carolina M Voloch, Ronaldo da Silva F Jr, Luiz G P de Almeida, Cynthia C Cardoso, Otavio Bustrolini, Alexandra L Gerber, Ana Paula de C Guimarães, Diana Mariani, Andréa Cony Cavalcanti, Claudia dos Santos Rodrigues, Terezinha M P P Castiñeira, Amilcar Tanuri, Ana Tereza R de Vasconcelos |
| EPI_ISL_717810, EPI_ISL_717811, EPI_ISL_717812, EPI_ISL_717813, EPI_ISL_717814, EPI_ISL_717815, EPI_ISL_717816, EPI_ISL_717817, EPI_ISL_717818, EPI_ISL_717819, EPI_ISL_717820, EPI_ISL_717821, EPI_ISL_717822, EPI_ISL_717823, EPI_ISL_717824, EPI_ISL_717825, EPI_ISL_717826, EPI_ISL_717827, EPI_ISL_717828, EPI_ISL_717829, EPI_ISL_717830, EPI_ISL_717831 | | | |
| see above | Laboratorio de Virologia Molecular / UFRJ | Bioinformatics Laboratory / LNCC | Carolina M Voloch, Ronaldo da Silva F Jr, Luiz G P de Almeida, Cynthia C Cardoso, Otavio Bustrolini, Alexandra L Gerber, Ana Paula de C Guimarães, Diana Mariani, Andréa Cony Cavalcanti, Claudia dos Santos Rodrigues, Terezinha M P P Castiñeira, Amilcar Tanuri, Ana Tereza R de Vasconcelos |
| EPI_ISL_717832, EPI_ISL_717833, EPI_ISL_717834, EPI_ISL_717835, EPI_ISL_717836 | LACEN Dr. Francisco Rimolo Neto | Bioinformatics Laboratory / LNCC | Carolina M Voloch, Ronaldo da Silva F Jr, Luiz G P de Almeida, Cynthia C Cardoso, Otavio Bustrolini, Alexandra L Gerber, Ana Paula de C Guimarães, Diana Mariani, Andréa Cony Cavalcanti, Claudia dos Santos Rodrigues, Terezinha M P P Castiñeira, Amilcar Tanuri, Ana Tereza R de Vasconcelos |
| EPI_ISL_717837, EPI_ISL_717838, EPI_ISL_717839, EPI_ISL_717840 | Laboratorio de Virologia Molecular / UFRJ | Bioinformatics Laboratory / LNCC | Carolina M Voloch, Ronaldo da Silva F Jr, Luiz G P de Almeida, Cynthia C Cardoso, Otavio Bustrolini, Alexandra L Gerber, Ana Paula de C Guimarães, Diana Mariani, Andréa Cony Cavalcanti, Claudia dos Santos Rodrigues, Terezinha M P P Castiñeira, Amilcar Tanuri, Ana Tereza R de Vasconcelos |
| EPI_ISL_717841 | LACEN Dr. Francisco Rimolo Neto | Bioinformatics Laboratory / LNCC | Carolina M Voloch, Ronaldo da Silva F Jr, Luiz G P de Almeida, Cynthia C Cardoso, Otavio Bustrolini, Alexandra L Gerber, Ana Paula de C Guimarães, Diana Mariani, Andréa Cony Cavalcanti, Claudia dos Santos Rodrigues, Terezinha M P P Castiñeira, Amilcar Tanuri, Ana Tereza R de Vasconcelos |
| EPI_ISL_717842, EPI_ISL_717843, EPI_ISL_717844, EPI_ISL_717845, EPI_ISL_717846, EPI_ISL_717847, EPI_ISL_717848, EPI_ISL_717849, EPI_ISL_717850, EPI_ISL_717851, EPI_ISL_717852, EPI_ISL_717853, EPI_ISL_717854, EPI_ISL_717855, EPI_ISL_717856, EPI_ISL_717857, EPI_ISL_717858, EPI_ISL_717859, EPI_ISL_717860, EPI_ISL_717861, EPI_ISL_717862, EPI_ISL_717863, EPI_ISL_717864, EPI_ISL_717865, EPI_ISL_717866, EPI_ISL_717867, EPI_ISL_717868, EPI_ISL_717869, EPI_ISL_717870, EPI_ISL_717871, EPI_ISL_717872, EPI_ISL_717873, EPI_ISL_717874, EPI_ISL_717875, EPI_ISL_717876, EPI_ISL_717877, EPI_ISL_717878, EPI_ISL_717879, EPI_ISL_717880, EPI_ISL_717881, EPI_ISL_717882, EPI_ISL_717883, EPI_ISL_717884, EPI_ISL_717885, EPI_ISL_717886, EPI_ISL_717887, EPI_ISL_717888, EPI_ISL_717889, EPI_ISL_717890, EPI_ISL_717891, EPI_ISL_717892, EPI_ISL_717893, EPI_ISL_717894, EPI_ISL_717895, EPI_ISL_717896, EPI_ISL_717897, EPI_ISL_717898 | | | |
| see above | Laboratorio de Virologia Molecular / UFRJ | Bioinformatics Laboratory / LNCC | Carolina M Voloch, Ronaldo da Silva F Jr, Luiz G P de Almeida, Cynthia C Cardoso, Otavio Bustrolini, Alexandra L Gerber, Ana Paula de C Guimarães, Diana Mariani, Andréa Cony Cavalcanti, Claudia dos Santos Rodrigues, Terezinha M P P Castiñeira, Amilcar Tanuri, Ana Tereza R de Vasconcelos |
| EPI_ISL_717899, EPI_ISL_717900, EPI_ISL_717901, EPI_ISL_717902, EPI_ISL_717903, EPI_ISL_717904, EPI_ISL_717905, EPI_ISL_717906, EPI_ISL_717907, EPI_ISL_717908, EPI_ISL_717909 | | | |
| see above | LACEN RJ - Noel Nutels | Bioinformatics Laboratory / LNCC | Carolina M Voloch, Ronaldo da Silva F Jr, Luiz G P de Almeida, Cynthia C Cardoso, Otavio Bustrolini, Alexandra L Gerber, Ana Paula de C Guimarães, Diana Mariani, Andréa Cony Cavalcanti, Claudia dos Santos Rodrigues, Terezinha M P P Castiñeira, Amilcar Tanuri, Ana Tereza R de Vasconcelos |
| EPI_ISL_717910, EPI_ISL_717911, EPI_ISL_717912, EPI_ISL_717913, EPI_ISL_717914, EPI_ISL_717915, EPI_ISL_717916, EPI_ISL_717917, EPI_ISL_717918, EPI_ISL_717919 | LACEN Dr. Francisco Rimolo Neto | Bioinformatics Laboratory / LNCC | Carolina M Voloch, Ronaldo da Silva F Jr, Luiz G P de Almeida, Cynthia C Cardoso, Otavio Bustrolini, Alexandra L Gerber, Ana Paula de C Guimarães, Diana Mariani, Andréa Cony Cavalcanti, Claudia dos Santos Rodrigues, Terezinha M P P Castiñeira, Amilcar Tanuri, Ana Tereza R de Vasconcelos |
| EPI_ISL_717920, EPI_ISL_717921, EPI_ISL_717922, EPI_ISL_717923, EPI_ISL_717924, EPI_ISL_717925, EPI_ISL_717926, EPI_ISL_717927, EPI_ISL_717928, EPI_ISL_717929, EPI_ISL_717930, EPI_ISL_717931, EPI_ISL_717932, EPI_ISL_717933, EPI_ISL_717934, EPI_ISL_717935, EPI_ISL_717936, EPI_ISL_717937, EPI_ISL_717938, EPI_ISL_717939, EPI_ISL_717940, EPI_ISL_717941, EPI_ISL_717942, EPI_ISL_717943, EPI_ISL_717944, EPI_ISL_717945, EPI_ISL_717946, EPI_ISL_717947, EPI_ISL_717948, EPI_ISL_717949, EPI_ISL_717950, EPI_ISL_717951, EPI_ISL_717952, EPI_ISL_717953, EPI_ISL_717954, EPI_ISL_717955, EPI_ISL_717956, EPI_ISL_717957 | | | |
| see above | Laboratorio de Virologia Molecular / UFRJ | Bioinformatics Laboratory / LNCC | Carolina M Voloch, Ronaldo da Silva F Jr, Luiz G P de Almeida, Cynthia C Cardoso, Otavio Bustrolini, Alexandra L Gerber, Ana Paula de C Guimarães, Diana Mariani, Andréa Cony Cavalcanti, Claudia dos Santos Rodrigues, Terezinha M P P Castiñeira, Amilcar Tanuri, Ana Tereza R de Vasconcelos |
| EPI_ISL_717958 | LACEN Dr. Francisco Rimolo Neto | Bioinformatics Laboratory / LNCC | Carolina M Voloch, Ronaldo da Silva F Jr, Luiz G P de Almeida, Cynthia C Cardoso, Otavio Bustrolini, Alexandra L Gerber, Ana Paula de C Guimarães, Diana Mariani, Andréa Cony Cavalcanti, Claudia dos Santos Rodrigues, Terezinha M P P Castiñeira, Amilcar Tanuri, Ana Tereza R de Vasconcelos |
| EPI_ISL_717959, EPI_ISL_717960, EPI_ISL_717961 | Laboratorio de Virologia Molecular / UFRJ | Bioinformatics Laboratory / LNCC | Carolina M Voloch, Ronaldo da Silva F Jr, Luiz G P de Almeida, Cynthia C Cardoso, Otavio Bustrolini, Alexandra L Gerber, Ana Paula de C Guimarães, Diana Mariani, Andréa Cony Cavalcanti, Claudia dos Santos Rodrigues, Terezinha M P P Castiñeira, Amilcar Tanuri, Ana Tereza R de Vasconcelos |
| EPI_ISL_717962 | LACEN RJ - Noel Nutels | Bioinformatics Laboratory / LNCC | Carolina M Voloch, Ronaldo da Silva F Jr, Luiz G P de Almeida, Cynthia C Cardoso, Otavio Bustrolini, Alexandra L Gerber, Ana Paula de C Guimarães, Diana Mariani, Andréa Cony Cavalcanti, Claudia dos Santos Rodrigues, Terezinha M P P Castiñeira, Amilcar Tanuri, Ana Tereza R de Vasconcelos |
| EPI_ISL_717963, EPI_ISL_717964 | LACEN Dr. Francisco Rimolo Neto | Bioinformatics Laboratory / LNCC | Carolina M Voloch, Ronaldo da Silva F Jr, Luiz G P de Almeida, Cynthia C Cardoso, Otavio Bustrolini, Alexandra L Gerber, Ana Paula de C Guimarães, Diana Mariani, Andréa Cony Cavalcanti, Claudia dos Santos Rodrigues, Terezinha M P P Castiñeira, Amilcar Tanuri, Ana Tereza R de Vasconcelos |
| EPI_ISL_983865 | Central Laboratory of Public Health of Rio Grande do Sul (Lacen-RS) | State Center for Health Surveillance of the Health Department of the State of Rio Grande do Sul (CEVS/SES-RS) | Aline Campos, Cynthia Molina, Lara Crescente, Leticia Garay, Ludmila Fiorenzano Baethgen, Richard Salvato, Tatiana Gregianini |

We gratefully acknowledge the following Authors from the Originating laboratories responsible for obtaining the specimens, as well as the Submitting laboratories where the genome data were generated and shared via GISAID, on which this research is based.

All Submitters of data may be contacted directly via www.gisaid.org

Authors are sorted alphabetically.

| Accession ID | Originating Laboratory | Submitting Laboratory | Authors |
|---|---|---|---|
| EPI_ISL_1023841 | Centro de Desenvolvimento Tecnológico em Saúde - CDTS | Centro de Desenvolvimento Tecnológico em Saúde - CDTS | Souza,T.M., Fintelman-Rodrigues,N., De Paula,A.D., Saraiva,F.B., Ferreira,M.A. and Sacramento,C.Q. |
| EPI_ISL_1133266, EPI_ISL_1133268 | Laboratório Hermes Pardini | Laboratório de Biologia Integrativa, Instituto de Ciências Biológicas, Universidade Federal de Minas Gerais | Filipe Romero Rebello Moreira, Diego Menezes Bonfim, Danielle Alves Gomes Zauli, Joice do Prado Silva, Aline Brito de Lima, Frederico Scott Varella Malta, Alessandro Clayton de Souza Ferreira, Victor Cavalcanti Pardini, Daniel Costa Queiroz, Rafael Marques de Souza, Victor Emmanuel Viana Geddes, Walyson Coelho Costa, Wagner Carlos Santos Magalhaes, Rennan Garcias Moreira, Carolina Moreira Voloch, Renan Pedra de Souza, Renato Santana Aguiar. |
| EPI_ISL_1181360, EPI_ISL_1181361, EPI_ISL_1181363, EPI_ISL_1181367, EPI_ISL_1181369, EPI_ISL_1181381, EPI_ISL_1181382, EPI_ISL_1181383, EPI_ISL_1181384, EPI_ISL_1181385, EPI_ISL_1181386, EPI_ISL_1181387, EPI_ISL_1181388, EPI_ISL_1181389, EPI_ISL_1181391, EPI_ISL_1181393, EPI_ISL_1181398, EPI_ISL_1181410, EPI_ISL_1181415 | | | |
| see above | Laboratory of Respiratory Viruses and Measles, Oswaldo Cruz Institute, FIOCRUZ | Laboratory of Respiratory Viruses and Measles, Oswaldo Cruz Institute, FIOCRUZ | Paola Resende, Luciana Appolinario, Fernando Motta, Anna Carolina Paixao, Ana Carolina Mendonca, Alice Sampaio Rocha, Renata Serrano Lopes, Marilda Siqueira on behalf of the Fiocruz COVID-19 Genomic Surveillance Network |
| EPI_ISL_1181425 | Laboratorio Central de Saude Publica do Estado do Rio de Janeiro (LACEN-RJ) | Laboratory of Respiratory Viruses and Measles, Oswaldo Cruz Institute, FIOCRUZ | Paola Resende, Luciana Appolinario, Fernando Motta, Anna Carolina Paixao, Ana Carolina Mendonca, Alice Sampaio Rocha, Renata Serrano Lopes, Andrea Cony Cavalcanti, Marilda Siqueira on behalf of the Fiocruz COVID-19 Genomic Surveillance Network |
| EPI_ISL_1181430 | Laboratory of Respiratory Viruses and Measles, Oswaldo Cruz Institute, FIOCRUZ | Laboratory of Respiratory Viruses and Measles, Oswaldo Cruz Institute, FIOCRUZ | Paola Resende, Luciana Appolinario, Fernando Motta, Anna Carolina Paixao, Ana Carolina Mendonca, Alice Sampaio Rocha, Renata Serrano Lopes, Marilda Siqueira on behalf of the Fiocruz COVID-19 Genomic Surveillance Network |
| EPI_ISL_1181431 | Laboratorio Central de Saude Publica do Estado do Rio de Janeiro (LACEN-RJ) | Laboratory of Respiratory Viruses and Measles, Oswaldo Cruz Institute, FIOCRUZ | Paola Resende, Luciana Appolinario, Fernando Motta, Anna Carolina Paixao, Ana Carolina Mendonca, Alice Sampaio Rocha, Renata Serrano Lopes, Andrea Cony Cavalcanti, Marilda Siqueira on behalf of the Fiocruz COVID-19 Genomic Surveillance Network |
| EPI_ISL_1181437, EPI_ISL_1181438, EPI_ISL_1181439, EPI_ISL_1181442, EPI_ISL_1181446, EPI_ISL_1181447, EPI_ISL_1181448, EPI_ISL_1181454, EPI_ISL_1181455, EPI_ISL_1181456, EPI_ISL_1181457, EPI_ISL_1181458, EPI_ISL_1181459, EPI_ISL_1181460, EPI_ISL_1181461, EPI_ISL_1181462, EPI_ISL_1181463, EPI_ISL_1181464, EPI_ISL_1181465, EPI_ISL_1181466, EPI_ISL_1181467, EPI_ISL_1181470, EPI_ISL_1181475, EPI_ISL_1181476, EPI_ISL_1181477, EPI_ISL_1181481, EPI_ISL_1181482, EPI_ISL_1181483, EPI_ISL_1181486, EPI_ISL_1181488, EPI_ISL_1181489, EPI_ISL_1181490, EPI_ISL_1181491, EPI_ISL_1181493, EPI_ISL_1181494, EPI_ISL_1181495, EPI_ISL_1181496, EPI_ISL_1181497, EPI_ISL_1181499, EPI_ISL_1181500, EPI_ISL_1181501, EPI_ISL_1181502, EPI_ISL_1181503, EPI_ISL_1181504, EPI_ISL_1181505, EPI_ISL_1181506, EPI_ISL_1181507, EPI_ISL_1181508, EPI_ISL_1181509, EPI_ISL_1181510, EPI_ISL_1181511, EPI_ISL_1181512, EPI_ISL_1181513, EPI_ISL_1181514, EPI_ISL_1181515, EPI_ISL_1181518, EPI_ISL_1181519, EPI_ISL_1181521, EPI_ISL_1181523, EPI_ISL_1181529, EPI_ISL_1181531, EPI_ISL_1181560, EPI_ISL_1181561, EPI_ISL_1181562, EPI_ISL_1181571, EPI_ISL_1181572, EPI_ISL_1181573, EPI_ISL_1181574, EPI_ISL_1181578, EPI_ISL_1181579, EPI_ISL_1181587, EPI_ISL_1181589, EPI_ISL_1181594, EPI_ISL_1181597, EPI_ISL_1181599 | | | |
| see above | Laboratory of Respiratory Viruses and Measles, Oswaldo Cruz Institute, FIOCRUZ | Laboratory of Respiratory Viruses and Measles, Oswaldo Cruz Institute, FIOCRUZ | Paola Resende, Luciana Appolinario, Fernando Motta, Anna Carolina Paixao, Ana Carolina Mendonca, Alice Sampaio Rocha, Renata Serrano Lopes, Marilda Siqueira on behalf of the Fiocruz COVID-19 Genomic Surveillance Network |
| EPI_ISL_1181600, EPI_ISL_1181601, EPI_ISL_1181602, EPI_ISL_1181603, EPI_ISL_1181606, EPI_ISL_1181607 | Laboratorio Central de Saude Publica do Estado do Rio de Janeiro (LACEN-RJ) | Laboratory of Respiratory Viruses and Measles, Oswaldo Cruz Institute, FIOCRUZ | Paola Resende, Luciana Appolinario, Fernando Motta, Anna Carolina Paixao, Ana Carolina Mendonca, Alice Sampaio Rocha, Renata Serrano Lopes, Andrea Cony Cavalcanti, Marilda Siqueira on behalf of the Fiocruz COVID-19 Genomic Surveillance Network |
| EPI_ISL_1181608 | Laboratory of Respiratory Viruses and Measles, Oswaldo Cruz Institute, FIOCRUZ | Laboratory of Respiratory Viruses and Measles, Oswaldo Cruz Institute, FIOCRUZ | Paola Resende, Luciana Appolinario, Fernando Motta, Anna Carolina Paixao, Ana Carolina Mendonca, Alice Sampaio Rocha, Renata Serrano Lopes, Marilda Siqueira on behalf of the Fiocruz COVID-19 Genomic Surveillance Network |
| EPI_ISL_1181611, EPI_ISL_1181612, EPI_ISL_1181613 | Laboratorio Central de Saude Publica do Estado do Rio de Janeiro (LACEN-RJ) | Laboratory of Respiratory Viruses and Measles, Oswaldo Cruz Institute, FIOCRUZ | Paola Resende, Luciana Appolinario, Fernando Motta, Anna Carolina Paixao, Ana Carolina Mendonca, Alice Sampaio Rocha, Renata Serrano Lopes, Andrea Cony Cavalcanti, Marilda Siqueira on behalf of the Fiocruz COVID-19 Genomic Surveillance Network |
| EPI_ISL_1182552, EPI_ISL_1182553, EPI_ISL_1182554, EPI_ISL_1182556, EPI_ISL_1182557, EPI_ISL_1182558 | Laboratório Central do Estado do Rio de Janeiro | Coordenação Geral de Laboratórios de Saúde Pública (CGLAB/DAEVS/SVS/MS) | Vagner Fonseca, et al. |
| EPI_ISL_1213163, EPI_ISL_1213171, EPI_ISL_1213248, EPI_ISL_1213249, EPI_ISL_1213270, EPI_ISL_1213272, EPI_ISL_1213307, EPI_ISL_1213309, EPI_ISL_1213310, EPI_ISL_1213312, EPI_ISL_1213314, EPI_ISL_1213339, EPI_ISL_1213340, EPI_ISL_1213372, EPI_ISL_1213374, EPI_ISL_1213376, EPI_ISL_1213378, EPI_ISL_1213379, EPI_ISL_1213381, EPI_ISL_1213382, EPI_ISL_1213393, EPI_ISL_1213395, EPI_ISL_1213397, EPI_ISL_1213408, EPI_ISL_1213410, EPI_ISL_1213417, EPI_ISL_1213418, EPI_ISL_1213420, EPI_ISL_1213422, EPI_ISL_1213424, EPI_ISL_1213425 | | | |
| see above | Laboratório HLA/UERJ | Bioinformatics Laboratory / LNCC | Alessandra P Lamarca, Luiz G P de Almeida, Ronaldo da Silva Francisco Jr, Lucymara Fassarella Agnez Lima, Kátia Castanho Scortecci, Vinícius Pietta Perez, Otavio J. Brustolini, Eduardo Sérgio Soares Sousa, Danielle Angst Secco, Angela Maria Guimarães Santos, George Rego Albuquerque, Ana Paula Melo Mariano, Bianca Mendes Maciel, Alexandra L Gerber, Ana Paula de C Guimarães, Paulo Ricardo Nascimento, Francisco Paulo Freire Neto, Sandra Rocha Gadelha, Luís Cristóvão Porto, Eloiza Helena Campana, Selma Maria Bezerra Jeronimo, Ana Tereza R Vasconcelos |
| EPI_ISL_1239111, EPI_ISL_1239112, EPI_ISL_1239113 | Laboratório Central de Saúde Pública Noel Nutels | Coordenação Geral de Laboratórios de Saúde Pública (CGLAB) | Vagner Fonseca et al, |
| EPI_ISL_1289218 | Dutch COVID-19 response team | National Institute for Public Health and the Environment (RIVM) | Adam Meijer, Harry Vennema, Dirk Eggink, Jeroen Cremer, Sharon van den Brink, Bas van der Veer, AnneMarie van den Brandt, Florian Zwagemaker, Dennis Schmitz, Chantal Reusken, on behalf of the national COVID-19 response team |
| EPI_ISL_1324142, EPI_ISL_1324145 | UW Virology Lab | UW Virology Lab | Pavitra Roychoudhury, Hong Xie, Lasata Shrestha, Shah Mohamed Bakhash, Michelle Lin, Margaret Mills, Noah Baker, Sean Ellis, Saraswathi Sathees, Meei-Li Huang, Keith R Jerome, Alexander Greninger |
| EPI_ISL_1327150 | Lighthouse Lab in Cambridge | Wellcome Sanger Institute for the COVID-19 Genomics UK (COG-UK) Consortium | Rob Howes, The Lighthouse Lab in Cambridge and Alex Alderton, Roberto Amato, Jeffrey Barrett, Sonia Goncalves, Ewan Harrison, David K. Jackson, Ian Johnston, Dominic Kwiatkowski, Cordelia Langford, John Sillitoe on behalf of the Wellcome Sanger Institute COVID-19 Surveillance Team |
| EPI_ISL_1402430 | Laboratory of Respiratory Viruses and Measles, Oswaldo Cruz Institute, FIOCRUZ | Laboratory of Respiratory Viruses and Measles, Oswaldo Cruz Institute, FIOCRUZ | Paola Resende, Alex Pauvolid-Correa, Mia Ferreira Araujo, Ana Beatriz Machado Lima, Luciana Appolinario, Fernando Motta, Anna Carolina Paixao, Ana Carolina Mendonca, Alice Sampaio Rocha, Renata Serrano Lopes, Marilda Siqueira on behalf of the Fiocruz COVID-19 Genomic Surveillance Network |
| EPI_ISL_1416322 | PathWest Laboratory Medicine WA | PathWest Laboratory Medicine WA Microbial Surveillance Unit | PathWest Laboratory Medicine WA Microbial Surveillance Unit |
| EPI_ISL_1445142 | CS II EGIDIO BRUNHARA MORRO AGUDO | Instituto Butantan / Mendelics | Dimas Tadeu Covas, Sandra Coccuzzo Sampaio, Maria Carolina Elias, José Salvatore Leister Patané, Vincent Louis Viala, Antonio Jorge Martins, Ricardo Haddad, Claudia Renata dos Santos Barros, Elaine Cristina Marqueze, Raul Machado Neto, Debora Botequio Moretti, Bibiana Santos, João Paulo Kitajima, Erika Freitas, David Schlesinger, Simone Kashima, Evandra Strazza Rodrigues, Svetoslav Nanev Slavov, Elaine Vieira dos Santos, Rafael dos Santos Bezerra, Luiz Carlos Junior de Alcantara, Marta Giovanetti, Vagner Fonseca, Flavia Aburjaile, Rodrigo Tocantins Calado. |
| EPI_ISL_1445155 | LABORATORIO DE FRANCA | Instituto Butantan / Mendelics | Dimas Tadeu Covas, Sandra Coccuzzo Sampaio, Maria Carolina Elias, José Salvatore Leister Patané, Vincent Louis Viala, Antonio Jorge Martins, Ricardo Haddad, Claudia Renata dos Santos Barros, Elaine Cristina Marqueze, Raul Machado Neto, Debora Botequio Moretti, Bibiana Santos, João Paulo Kitajima, Erika Freitas, David Schlesinger, Simone Kashima, Evandra Strazza Rodrigues, Svetoslav Nanev Slavov, Elaine Vieira dos Santos, Rafael dos Santos Bezerra, Luiz Carlos Junior de Alcantara, Marta Giovanetti, Vagner Fonseca, Flavia Aburjaile, Rodrigo Tocantins Calado. |
| EPI_ISL_1445184 | UNIDADE BASICA DE SAUDE DA FAMILIA BILAC | Instituto Butantan / Mendelics | Dimas Tadeu Covas, Sandra Coccuzzo Sampaio, Maria Carolina Elias, José Salvatore Leister Patané, Vincent Louis Viala, Antonio Jorge Martins, Ricardo Haddad, Claudia Renata dos Santos Barros, Elaine Cristina Marqueze, Raul Machado Neto, Debora Botequio Moretti, Bibiana Santos, João Paulo Kitajima, Erika Freitas, David Schlesinger, Simone Kashima, Evandra Strazza Rodrigues, Svetoslav Nanev Slavov, Elaine Vieira dos Santos, Rafael dos Santos Bezerra, Luiz Carlos Junior de Alcantara, Marta Giovanetti, Vagner Fonseca, Flavia Aburjaile, Rodrigo Tocantins Calado. |
| EPI_ISL_1446194 | LATE - Laboratório de Técnicas Especiais - Hospital Israelita Albert Einstein | LATE - Laboratório de Técnicas Especiais - Hospital Israelita Albert Einstein | Deyvid Amgarten, Fernanda de Mello Malta, Raquel Riyuzo, Ana Paula Moreira Salles, Pedro Henrique Sebe Rodrigues, João Renato Rebello Pinho |

| | | | |
|---|---|---|---|
| EPI_ISL_1456633, EPI_ISL_1456637, EPI_ISL_1456638, EPI_ISL_1456646, EPI_ISL_1456649, EPI_ISL_1456650, EPI_ISL_1456651, EPI_ISL_1456652, EPI_ISL_1521307, EPI_ISL_1521308, EPI_ISL_1521309, EPI_ISL_1521310, EPI_ISL_1521312, EPI_ISL_1521313, EPI_ISL_1521314, EPI_ISL_1521315 | | | |
| see above | Dutch COVID-19 response team | National Institute for Public Health and the Environment (RIVM) | Adam Meijer, Harry Vennema, Dirk Eggink, Jeroen Cremer, Sharon van den Brink, Bas van der Veer, AnneMarie van den Brandt, Lisa Wijsman, Kim Freriks, Ryanne Jaarsma, Euníce Then, Jolienke Hardeman, Lynn Aarts, Sanne Bos, Melissa van Tuil, Robert Kohl, Linda van de Nes, Sjoerd Kuiling, James Groot, Florian Zwagemaker, Dennis Schmitz, Annelies Kroneman, Karim Hajji, Chantal Reusken, on behalf of the national COVID-19 response team |
| EPI_ISL_1533977 | Laboratorio de Virologia Molecular / UFRJ | Laboratory of Respiratory Viruses and Measles, Oswaldo Cruz Institute, FIOCRUZ | Paola Resende, Carolina M Voloch, Luciana Appolinario, Fernando Motta, Anna Carolina Paixao, Ana Carolina Mendonca, Alice Sampaio Rocha, Renata Serrano Lopes, Amilcar Tanuri, Marilda Siqueira on behalf of the Fiocruz COVID-19 Genomic Surveillance Network |
| EPI_ISL_1533980, EPI_ISL_1533983 | Laboratory of Respiratory Viruses and Measles, Oswaldo Cruz Institute, FIOCRUZ | Laboratory of Respiratory Viruses and Measles, Oswaldo Cruz Institute, FIOCRUZ | Paola Resende, Luciana Appolinario, Fernando Motta, Anna Carolina Paixao, Ana Carolina Mendonca, Alice Sampaio Rocha, Renata Serrano Lopes, Marilda Siqueira on behalf of the Fiocruz COVID-19 Genomic Surveillance Network |
| EPI_ISL_1533991, EPI_ISL_1533995, EPI_ISL_1533998 | Laboratorio de Virologia Molecular / UFRJ | Laboratory of Respiratory Viruses and Measles, Oswaldo Cruz Institute, FIOCRUZ | Paola Resende, Carolina M Voloch, Luciana Appolinario, Fernando Motta, Anna Carolina Paixao, Ana Carolina Mendonca, Alice Sampaio Rocha, Renata Serrano Lopes, Amilcar Tanuri, Marilda Siqueira on behalf of the Fiocruz COVID-19 Genomic Surveillance Network |
| EPI_ISL_1534005, EPI_ISL_1534011 | Laboratory of Respiratory Viruses and Measles, Oswaldo Cruz Institute, FIOCRUZ | Laboratory of Respiratory Viruses and Measles, Oswaldo Cruz Institute, FIOCRUZ | Paola Resende, Luciana Appolinario, Fernando Motta, Anna Carolina Paixao, Ana Carolina Mendonca, Alice Sampaio Rocha, Renata Serrano Lopes, Marilda Siqueira on behalf of the Fiocruz COVID-19 Genomic Surveillance Network |
| EPI_ISL_1534012 | Laboratorio Central de Saude Publica do Estado do Rio de Janeiro (LACEN-RJ) | Laboratory of Respiratory Viruses and Measles, Oswaldo Cruz Institute, FIOCRUZ | Paola Resende, Luciana Appolinario, Fernando Motta, Anna Carolina Paixao, Ana Carolina Mendonca, Alice Sampaio Rocha, Renata Serrano Lopes, Andrea Cony Cavalcanti, Marilda Siqueira on behalf of the Fiocruz COVID-19 Genomic Surveillance Network |
| EPI_ISL_1534014, EPI_ISL_1534015, EPI_ISL_1534016 | Laboratory of Respiratory Viruses and Measles, Oswaldo Cruz Institute, FIOCRUZ | Laboratory of Respiratory Viruses and Measles, Oswaldo Cruz Institute, FIOCRUZ | Paola Resende, Luciana Appolinario, Fernando Motta, Anna Carolina Paixao, Ana Carolina Mendonca, Alice Sampaio Rocha, Renata Serrano Lopes, Marilda Siqueira on behalf of the Fiocruz COVID-19 Genomic Surveillance Network |
| EPI_ISL_1583681 | Central Public Health Laboratory - LACEN -Bahia, Salvador, Brazil | Central Public Health Laboratory - LACEN -Bahia, Salvador, Brazil | Stephane Tosta, Luciana Oliveira, Vanessa Nardy,Patrícia Cajado,Marcela Gómez, Breno Dominguez, Jaqueline Gomes, Vagner Fonseca,Marta Giovanetti,Luiz Alcantara, Felicidade Pereira, Arabela Leal |
| EPI_ISL_1592230 | Laboratório Hermes Pardini | Laboratório de Biologia Integrativa, Instituto de Ciências Biológicas, Universidade Federal de Minas Gerais | Filipe Romero Rebello Moreira, Diego Menezes Bonfim, Danielle Alves Gomes Zauli, Joice do Prado Silva, Aline Brito de Lima, Frederico Scott Varella Malta, Alessandro Clayton de Souza Ferreira, Victor Cavalcanti Pardini, Daniel Costa Queiroz, Rafael Marques de Souza, Victor Emmanuel Viana Geddes, Walyson Coelho Costa, Wagner Carlos Santos Magalhaes, Rennan Garcias Moreira, Carolina Moreira Voloch, Renan Pedra de Souza, Renato Santana Aguiar. |
| EPI_ISL_1596573 | Dutch COVID-19 response team | National Institute for Public Health and the Environment (RIVM) | Adam Meijer, Harry Vennema, Dirk Eggink, Jeroen Cremer, Sharon van den Brink, Bas van der Veer, AnneMarie van den Brandt, Lisa Wijsman, Kim Freriks, Ryanne Jaarsma, Euníce Then, Jolienke Hardeman, Lynn Aarts, Sanne Bos, Melissa van Tuil, Robert Kohl, Linda van de Nes, Sjoerd Kuiling, James Groot, Florian Zwagemaker, Dennis Schmitz, Annelies Kroneman, Karim Hajji, Chantal Reusken, on behalf of the national COVID-19 response team |
| EPI_ISL_1649154 | Aegis Sciences Corporation | Centers for Disease Control and Prevention Division of Viral Diseases, Pathogen Discovery | Dakota Howard, Dhwani Batra, Peter W. Cook, Kara Moser, Adrian Paskey, Jason Caravas, Benjamin Rambo-Martin, Shatavia Morrison, Christopher Gulvick, Scott Sammons, Yvette Unoarumhi, Darlene Wagner, Matthew Schmerer, Cyndi Clark, Patrick Campbell, Rob Case, Vikramsinha Ghorpade, Holly Houdeshell, Ola Kvalvaag, Dillon Nall, Ethan Sanders, Alec Vest, Shaun Westlund, Matthew Hardison, Clinton R. Paden, Duncan MacCannell |
| EPI_ISL_1664113, EPI_ISL_1664114, EPI_ISL_1664115, EPI_ISL_1664116, EPI_ISL_1664117, EPI_ISL_1664118, EPI_ISL_1664119, EPI_ISL_1664120, EPI_ISL_1664121, EPI_ISL_1664122, EPI_ISL_1664123, EPI_ISL_1664124, EPI_ISL_1664125, EPI_ISL_1664126, EPI_ISL_1664127, EPI_ISL_1664128, EPI_ISL_1664129, EPI_ISL_1664130, EPI_ISL_1664131, EPI_ISL_1664132, EPI_ISL_1664133, EPI_ISL_1664134, EPI_ISL_1664135, EPI_ISL_1664136, EPI_ISL_1664137, EPI_ISL_1664138, EPI_ISL_1664139, EPI_ISL_1664140, EPI_ISL_1664141, EPI_ISL_1664142, EPI_ISL_1664143, EPI_ISL_1664144, EPI_ISL_1664145, EPI_ISL_1664146, EPI_ISL_1664147, EPI_ISL_1664148, EPI_ISL_1664149, EPI_ISL_1664150, EPI_ISL_1664151, EPI_ISL_1664152, EPI_ISL_1664153, EPI_ISL_1664154, EPI_ISL_1664155, EPI_ISL_1664156, EPI_ISL_1664157, EPI_ISL_1664158, EPI_ISL_1664159, EPI_ISL_1664160, EPI_ISL_1664161, EPI_ISL_1664162, EPI_ISL_1664163, EPI_ISL_1664164, EPI_ISL_1664165, EPI_ISL_1664166, EPI_ISL_1664167, EPI_ISL_1664168, EPI_ISL_1664169, EPI_ISL_1664170, EPI_ISL_1664171, EPI_ISL_1664172, EPI_ISL_1664173, EPI_ISL_1664174, EPI_ISL_1664175, EPI_ISL_1664176, EPI_ISL_1664177, EPI_ISL_1664178, EPI_ISL_1664179, EPI_ISL_1664180, EPI_ISL_1664181, EPI_ISL_1664182, EPI_ISL_1664183, EPI_ISL_1664184, EPI_ISL_1664185, EPI_ISL_1664186, EPI_ISL_1664187, EPI_ISL_1664188, EPI_ISL_1664189, EPI_ISL_1664190, EPI_ISL_1664191, EPI_ISL_1664192, EPI_ISL_1664193, EPI_ISL_1664194, EPI_ISL_1664195, EPI_ISL_1664196, EPI_ISL_1664197, EPI_ISL_1664198, EPI_ISL_1664199, EPI_ISL_1664200, EPI_ISL_1664201, EPI_ISL_1664202 | | | |
| see above | Laboratorio Central Noel Nutels | Bioinformatics Laboratory / LNCC | Luiz G P de Almeida, Alessandra P Lamarca, Ronaldo da Silva F Jr, Liliane Cavalcante, Alexandra L Gerber, Ana Paula de C Guimarães, Douglas Terra Machado, Cassia Alves, Diana Mariani, Thais Felix Cruz, Mario Sergio Ribeiro, Silvia Carvalho, Flávio Dias da Silva, Marcio Henrique de Oliveira Garcia, Leandro Magalhães de Souza, Cristiane Gomes da Silva, Caio Luiz Pereira Ribeiro, Andréa Cony Cavalcanti, Claudia Maria Braga de Mello, Amilcar Tanuri, Ana Tereza R Vasconcelos |
| EPI_ISL_1678584, EPI_ISL_1690551, EPI_ISL_1694627, EPI_ISL_1694628, EPI_ISL_1694631, EPI_ISL_1694632, EPI_ISL_1694634, EPI_ISL_1695909 | Instituto Estadual do Cérebro Paulo Niemayer (IECPN) | Laboratório de Imunofarmacologia | Thiago Moreno Lopes Souza |
| EPI_ISL_1738831 | unknown | Instituto Nacional de Saude (INSA) | Borges et al |
| EPI_ISL_1752639 | UPA Dr Luis Atilio Losi Viana Ribeirao Preto | Instituto Adolfo Lutz, Interdiciplinary Procedures Center, Strategic Laboratory | Claudio Tavares Sacchi, Claudia Regina Gonçalves, Erica Valessa Ramos Gomes, Karoline Rodrigues Campos, Caio Vinicius Dias Lopes, Leonardo Jose Tadeu de Araujo, Katia Correa de Oliveira Santos |
| EPI_ISL_1795103 | HOSPITAL DOS FORNCEDORES | Instituto Butantan / ESALQ-Piracicaba | Instituto Butantan: Alexander Roberto Precioso, Dimas Tadeu Covas, Sandra Coccuzzo Sampaio, Maria Carolina Elias, José Salvatore Leister Patané, Vincent Louis Viala, Antonio Jorge Martins, Ricardo Haddad, Claudia Renata dos Santos Barros, Elaine Cristina Marqueze, Raul Machado Neto, Debora Botequio Moretti. Centro de Genômica Funcional da ESALQ: Luiz Lehmann Coutinho, Ricardo Augusto Brassaloti, Raquel de Lello Rocha Campos Cassano. NGS Soluções Genômicas: Pilar Drummond Sampaio Corrêa Mariani. FZEA-USP Pirassunumga: Mirele Daiana Poleti, Jessika Cristina Chagas Lesbon, Elisangela Chicaroni Mattos, Heidge Fukumasu. USP-Botucatu: Rejane Maria Tommasini Grotto, Jayme A. Souza-Neto, Guilherme Targino Valente, Patricia Akemi Assato, Felipe Allan da Silva da Costa, Bianca Cechetto Carlos. Mendelics: Bibiana Santos, João Paulo Kitajima, Erika Freitas, David Schlesinger. Hemocentro Ribeirão Preto: Simone Kashima, Evandra Strazza Rodrigues, Svetoslav Nanev Slavov, Elaine Vieira dos Santos, Rafael dos Santos Bezerra, Luiz Carlos Junior de Alcantara, Marta Giovanetti, Vagner Fonseca, Flavia Aburjaile, Rodrigo Tocantins Calado. |
| EPI_ISL_1795104, EPI_ISL_1795105 | LABORATORIO MUNICIPAL DE PIRACICABA | Instituto Butantan / ESALQ-Piracicaba | Instituto Butantan: Alexander Roberto Precioso, Dimas Tadeu Covas, Sandra Coccuzzo Sampaio, Maria Carolina Elias, José Salvatore Leister Patané, Vincent Louis Viala, Antonio Jorge Martins, Ricardo Haddad, Claudia Renata dos Santos Barros, Elaine Cristina Marqueze, Raul Machado Neto, Debora Botequio Moretti. Centro de Genômica Funcional da ESALQ: Luiz Lehmann Coutinho, Ricardo Augusto Brassaloti, Raquel de Lello Rocha Campos Cassano. NGS Soluções Genômicas: Pilar Drummond Sampaio Corrêa Mariani. FZEA-USP Pirassunumga: Mirele Daiana Poleti, Jessika Cristina Chagas Lesbon, Elisangela Chicaroni Mattos, Heidge Fukumasu. USP-Botucatu: Rejane Maria Tommasini Grotto, Jayme A. Souza-Neto, Guilherme Targino Valente, Patricia Akemi Assato, Felipe Allan da Silva da Costa, Bianca Cechetto Carlos. Mendelics: Bibiana Santos, João Paulo Kitajima, Erika Freitas, David Schlesinger. Hemocentro Ribeirão Preto: Simone Kashima, Evandra Strazza Rodrigues, Svetoslav Nanev Slavov, Elaine Vieira dos Santos, Rafael dos Santos Bezerra, Luiz Carlos Junior de Alcantara, Marta Giovanetti, Vagner Fonseca, Flavia Aburjaile, Rodrigo Tocantins Calado. |
| EPI_ISL_1795108 | SECRETARIA DE SAUDE DE SAO PEDRO | Instituto Butantan / ESALQ-Piracicaba | Instituto Butantan: Alexander Roberto Precioso, Dimas Tadeu Covas, Sandra Coccuzzo Sampaio, Maria Carolina Elias, José Salvatore Leister Patané, Vincent Louis Viala, Antonio Jorge Martins, Ricardo Haddad, Claudia Renata dos Santos Barros, Elaine Cristina Marqueze, Raul Machado Neto, Debora Botequio Moretti. Centro de Genômica Funcional da ESALQ: Luiz Lehmann Coutinho, Ricardo Augusto Brassaloti, Raquel de Lello Rocha Campos Cassano. NGS Soluções Genômicas: Pilar Drummond Sampaio Corrêa Mariani. FZEA-USP Pirassunumga: Mirele Daiana Poleti, Jessika Cristina Chagas Lesbon, Elisangela Chicaroni Mattos, Heidge Fukumasu. USP-Botucatu: Rejane Maria Tommasini Grotto, Jayme A. Souza-Neto, Guilherme Targino Valente, Patricia Akemi Assato, Felipe Allan da Silva da Costa, Bianca Cechetto Carlos. Mendelics: Bibiana Santos, João Paulo Kitajima, Erika Freitas, David Schlesinger. Hemocentro Ribeirão Preto: Simone Kashima, Evandra Strazza Rodrigues, Svetoslav Nanev Slavov, Elaine Vieira dos Santos, Rafael dos Santos Bezerra, Luiz Carlos Junior de Alcantara, Marta Giovanetti, Vagner Fonseca, Flavia Aburjaile, Rodrigo Tocantins Calado. |
| EPI_ISL_1795223 | HOSPITAL REGIONAL DE ITAPETININGA | Instituto Butantan / ESALQ-Piracicaba | Instituto Butantan: Alexander Roberto Precioso, Dimas Tadeu Covas, Sandra Coccuzzo Sampaio, Maria Carolina Elias, José Salvatore Leister Patané, Vincent Louis Viala, Antonio Jorge Martins, Ricardo Haddad, Claudia Renata dos Santos Barros, Elaine Cristina Marqueze, Raul Machado Neto, Debora Botequio Moretti. Centro de Genômica Funcional da ESALQ: Luiz Lehmann Coutinho, Ricardo Augusto Brassaloti, Raquel de Lello Rocha Campos Cassano. NGS Soluções Genômicas: Pilar Drummond Sampaio Corrêa Mariani. FZEA-USP Pirassunumga: Mirele Daiana Poleti, Jessika Cristina Chagas Lesbon, Elisangela Chicaroni Mattos, Heidge Fukumasu. USP-Botucatu: Rejane Maria Tommasini Grotto, Jayme A. Souza-Neto, Guilherme Targino Valente, Patricia Akemi Assato, Felipe Allan da Silva da Costa, Bianca Cechetto Carlos. Mendelics: Bibiana Santos, João Paulo Kitajima, Erika Freitas, |

| | | | |
|---|---|---|---|
| | | | David Schlesinger. Hemocentro Ribeirão Preto: Simone Kashima, Evandra Strazza Rodrigues, Svetoslav Nanev Slavov, Elaine Vieira dos Santos, Rafael dos Santos Bezerra, Luiz Carlos Junior de Alcantara, Marta Giovanetti, Vagner Fonseca, Flavia Aburjaile, Rodrigo Tocantins Calado. |
| EPI_ISL_1795329 | CENTRO DE ESPECIALIDADES DE PRIMAVERA | Instituto Butantan / ESALQ-Piracicaba | Instituto Butantan: Alexander Roberto Precioso, Dimas Tadeu Covas, Sandra Coccuzzo Sampaio, Maria Carolina Elias, José Salvatore Leister Patané, Vincent Louis Viala, Antonio Jorge Martins, Ricardo Haddad, Claudia Renata dos Santos Barros, Elaine Cristina Marqueze, Raul Machado Neto, Debora Botequio Moretti. Centro de Genômica Funcional da ESALQ: Luiz Lehmann Coutinho, Ricardo Augusto Brassaloti, Raquel de Lello Rocha Campos Cassano. NGS Soluções Genômicas: Pilar Drummond Sampaio Corrêa Mariani. FZEA-USP Pirassununga: Mirele Daiana Poleti, Jessika Cristina Chagas Lesbon, Elisangela Chicaroni Mattos, Heidge Fukumasu. USP-Botucatu: Rejane Maria Tommasini Grotto, Jayme A. Souza-Neto, Guilherme Targino Valente, Patricia Akemi Assato, Felipe Allan da Silva da Costa, Bianca Cechetto Carlos. Mendelics: Bibiana Santos, João Paulo Kitajima, Erika Freitas, David Schlesinger. Hemocentro Ribeirão Preto: Simone Kashima, Evandra Strazza Rodrigues, Svetoslav Nanev Slavov, Elaine Vieira dos Santos, Rafael dos Santos Bezerra, Luiz Carlos Junior de Alcantara, Marta Giovanetti, Vagner Fonseca, Flavia Aburjaile, Rodrigo Tocantins Calado. |
| EPI_ISL_1795395, EPI_ISL_1795396, EPI_ISL_1795397 | UBS III DE RANCHARIA | Instituto Butantan / ESALQ-Piracicaba | Instituto Butantan: Alexander Roberto Precioso, Dimas Tadeu Covas, Sandra Coccuzzo Sampaio, Maria Carolina Elias, José Salvatore Leister Patané, Vincent Louis Viala, Antonio Jorge Martins, Ricardo Haddad, Claudia Renata dos Santos Barros, Elaine Cristina Marqueze, Raul Machado Neto, Debora Botequio Moretti. Centro de Genômica Funcional da ESALQ: Luiz Lehmann Coutinho, Ricardo Augusto Brassaloti, Raquel de Lello Rocha Campos Cassano. NGS Soluções Genômicas: Pilar Drummond Sampaio Corrêa Mariani. FZEA-USP Pirassununga: Mirele Daiana Poleti, Jessika Cristina Chagas Lesbon, Elisangela Chicaroni Mattos, Heidge Fukumasu. USP-Botucatu: Rejane Maria Tommasini Grotto, Jayme A. Souza-Neto, Guilherme Targino Valente, Patricia Akemi Assato, Felipe Allan da Silva da Costa, Bianca Cechetto Carlos. Mendelics: Bibiana Santos, João Paulo Kitajima, Erika Freitas, David Schlesinger. Hemocentro Ribeirão Preto: Simone Kashima, Evandra Strazza Rodrigues, Svetoslav Nanev Slavov, Elaine Vieira dos Santos, Rafael dos Santos Bezerra, Luiz Carlos Junior de Alcantara, Marta Giovanetti, Vagner Fonseca, Flavia Aburjaile, Rodrigo Tocantins Calado. |
| EPI_ISL_1858267, EPI_ISL_1858269, EPI_ISL_1858271, EPI_ISL_1858273, EPI_ISL_1858275, EPI_ISL_1858277, EPI_ISL_1858279, EPI_ISL_1858281, EPI_ISL_1858282, EPI_ISL_1858284, EPI_ISL_1858286, EPI_ISL_1858288, EPI_ISL_1858290, EPI_ISL_1858292, EPI_ISL_1858294, EPI_ISL_1858295, EPI_ISL_1858297, EPI_ISL_1858299, EPI_ISL_1858301, EPI_ISL_1858303, EPI_ISL_1858304, EPI_ISL_1858306, EPI_ISL_1858308, EPI_ISL_1858310, EPI_ISL_1858312, EPI_ISL_1858314, EPI_ISL_1858315, EPI_ISL_1858317, EPI_ISL_1858319, EPI_ISL_1858321, EPI_ISL_1858323, EPI_ISL_1858325, EPI_ISL_1858327, EPI_ISL_1858329, EPI_ISL_1858331, EPI_ISL_1858333, EPI_ISL_1858334, EPI_ISL_1858336, EPI_ISL_1858338, EPI_ISL_1858340, EPI_ISL_1858342, EPI_ISL_1858344, EPI_ISL_1858346, EPI_ISL_1858348, EPI_ISL_1858350, EPI_ISL_1858352, EPI_ISL_1858354, EPI_ISL_1858356, EPI_ISL_1858357, EPI_ISL_1858359, EPI_ISL_1858361, EPI_ISL_1858363, EPI_ISL_1858365, EPI_ISL_1858367, EPI_ISL_1858369, EPI_ISL_1858371, EPI_ISL_1858373, EPI_ISL_1858375, EPI_ISL_1858377, EPI_ISL_1858379, EPI_ISL_1858380, EPI_ISL_1858383, EPI_ISL_1858384, EPI_ISL_1858387, EPI_ISL_1858389, EPI_ISL_1858391, EPI_ISL_1858392, EPI_ISL_1858395, EPI_ISL_1858397, EPI_ISL_1858399, EPI_ISL_1858401, EPI_ISL_1858403, EPI_ISL_1858409, EPI_ISL_1858411, EPI_ISL_1858414, EPI_ISL_1858416, EPI_ISL_1858418, EPI_ISL_1858420, EPI_ISL_1858422, EPI_ISL_1858423, EPI_ISL_1858426, EPI_ISL_1858428, EPI_ISL_1858430, EPI_ISL_1858432, EPI_ISL_1858434, EPI_ISL_1858436, EPI_ISL_1858438, EPI_ISL_1858440, EPI_ISL_1858442, EPI_ISL_1858444, EPI_ISL_1858446, EPI_ISL_1858449, EPI_ISL_1858451, EPI_ISL_1858453, EPI_ISL_1858454, EPI_ISL_1858456, EPI_ISL_1858458, EPI_ISL_1858460, EPI_ISL_1858462, EPI_ISL_1858464, EPI_ISL_1858466, EPI_ISL_1858468, EPI_ISL_1858470, EPI_ISL_1858473, EPI_ISL_1858475, EPI_ISL_1858477, EPI_ISL_1858479, EPI_ISL_1858481, EPI_ISL_1858483, EPI_ISL_1858486, EPI_ISL_1858488, EPI_ISL_1858490, EPI_ISL_1858492, EPI_ISL_1858494, EPI_ISL_1858496, EPI_ISL_1858498, EPI_ISL_1858500, EPI_ISL_1858502, EPI_ISL_1858504, EPI_ISL_1858506, EPI_ISL_1858508, EPI_ISL_1858512, EPI_ISL_1858514, EPI_ISL_1858516, EPI_ISL_1858518, EPI_ISL_1858520, EPI_ISL_1858522, EPI_ISL_1858526, EPI_ISL_1858528, EPI_ISL_1858530, EPI_ISL_1858532, EPI_ISL_1858536, EPI_ISL_1858538, EPI_ISL_1858540, EPI_ISL_1858542, EPI_ISL_1858544, EPI_ISL_1858546, EPI_ISL_1858548, EPI_ISL_1858549, EPI_ISL_1858551, EPI_ISL_1858554, EPI_ISL_1858555, EPI_ISL_1858557, EPI_ISL_1858559, EPI_ISL_1858561, EPI_ISL_1858563, EPI_ISL_1858565, EPI_ISL_1858567, EPI_ISL_1858571, EPI_ISL_1858572, EPI_ISL_1858575, EPI_ISL_1858577, EPI_ISL_1858578, EPI_ISL_1858580, EPI_ISL_1858582, EPI_ISL_1858584, EPI_ISL_1858586, EPI_ISL_1858588, EPI_ISL_1858590, EPI_ISL_1858592, EPI_ISL_1858594, EPI_ISL_1858596, EPI_ISL_1858598, EPI_ISL_1858599, EPI_ISL_1858603, EPI_ISL_1858606, EPI_ISL_1858607, EPI_ISL_1858610, EPI_ISL_1858611, EPI_ISL_1858613, EPI_ISL_1858615, EPI_ISL_1858617, EPI_ISL_1858619, EPI_ISL_1858621, EPI_ISL_1858623, EPI_ISL_1858625, EPI_ISL_1858627, EPI_ISL_1858629, EPI_ISL_1858631, EPI_ISL_1858633, EPI_ISL_1858636, EPI_ISL_1858637, EPI_ISL_1858639, EPI_ISL_1858641, EPI_ISL_1858643, EPI_ISL_1858645, EPI_ISL_1858648, EPI_ISL_1858650, EPI_ISL_1858651, EPI_ISL_1858655, EPI_ISL_1858656, EPI_ISL_1858658, EPI_ISL_1858660, EPI_ISL_1858662, EPI_ISL_1858664, EPI_ISL_1858666, EPI_ISL_1858668, EPI_ISL_1858670, EPI_ISL_1858672, EPI_ISL_1858674, EPI_ISL_1858675, EPI_ISL_1858677, EPI_ISL_1858680, EPI_ISL_1858682, EPI_ISL_1858684, EPI_ISL_1858686, EPI_ISL_1858688, EPI_ISL_1858691, EPI_ISL_1858693, EPI_ISL_1858694, EPI_ISL_1858697, EPI_ISL_1858701, EPI_ISL_1858703, EPI_ISL_1858705, EPI_ISL_1858707, EPI_ISL_1858709, EPI_ISL_1858711, EPI_ISL_1858713, EPI_ISL_1858715, EPI_ISL_1858717, EPI_ISL_1858719, EPI_ISL_1858721, EPI_ISL_1858722, EPI_ISL_1858724, EPI_ISL_1858726, EPI_ISL_1858728, EPI_ISL_1858730, EPI_ISL_1858732, EPI_ISL_1858734, EPI_ISL_1858736, EPI_ISL_1858738, EPI_ISL_1858740, EPI_ISL_1858741, EPI_ISL_1858743, EPI_ISL_1858745, EPI_ISL_1858747, EPI_ISL_1858751, EPI_ISL_1858752, EPI_ISL_1858754, EPI_ISL_1858756 | | | |
| see above | Laboratorio Central Noel Nutels | Bioinformatics Laboratory / LNCC | Luiz G P de Almeida, Alessandra P Lamarca, Ronaldo da Silva F Jr, Liliane Cavalcante, Alexandra L Gerber, Ana Paula de C Guimarães, Douglas Terra Machado, Cassia Alves, Diana Mariani, Thais Felix Cruz, Mario Sergio Ribeiro, Silvia Carvalho, Flávio Dias da Silva, Marcio Henrique de Oliveira Garcia, Leandro Magalhães de Souza, Cristiane Gomes da Silva, Caio Luiz Pereira Ribeiro, Andréa Cony Cavalcanti, Claudia Maria Braga de Mello, Amilcar Tanuri, Ana Tereza R Vasconcelos |
| EPI_ISL_1858758, EPI_ISL_1858761, EPI_ISL_1858763, EPI_ISL_1858765, EPI_ISL_1858767, EPI_ISL_1858771, EPI_ISL_1858775, EPI_ISL_1858777, EPI_ISL_1858779, EPI_ISL_1858781, EPI_ISL_1858783, EPI_ISL_1858786, EPI_ISL_1858788, EPI_ISL_1858790, EPI_ISL_1858792, EPI_ISL_1858794, EPI_ISL_1858796, EPI_ISL_1858798, EPI_ISL_1858800, EPI_ISL_1858802, EPI_ISL_1858803, EPI_ISL_1858805, EPI_ISL_1858807, EPI_ISL_1858809, EPI_ISL_1858811, EPI_ISL_1858814, EPI_ISL_1858816, EPI_ISL_1858818, EPI_ISL_1858819, EPI_ISL_1858822, EPI_ISL_1858824, EPI_ISL_1858826, EPI_ISL_1858828, EPI_ISL_1858830, EPI_ISL_1858832, EPI_ISL_1858834, EPI_ISL_1858836, EPI_ISL_1858838, EPI_ISL_1858840, EPI_ISL_1858842, EPI_ISL_1858843, EPI_ISL_1858846, EPI_ISL_1858847, EPI_ISL_1858850, EPI_ISL_1858852, EPI_ISL_1858853, EPI_ISL_1858856, EPI_ISL_1858858, EPI_ISL_1858860, EPI_ISL_1858862, EPI_ISL_1858864, EPI_ISL_1858866, EPI_ISL_1858868, EPI_ISL_1858870, EPI_ISL_1858872, EPI_ISL_1858878, EPI_ISL_1858880, EPI_ISL_1858882, EPI_ISL_1858884, EPI_ISL_1858886, EPI_ISL_1858888, EPI_ISL_1858890, EPI_ISL_1858892, EPI_ISL_1858894, EPI_ISL_1858896, EPI_ISL_1858898, EPI_ISL_1858900, EPI_ISL_1858902, EPI_ISL_1858904, EPI_ISL_1858906, EPI_ISL_1858908, EPI_ISL_1858910, EPI_ISL_1858912, EPI_ISL_1858914, EPI_ISL_1858916, EPI_ISL_1858918, EPI_ISL_1858920, EPI_ISL_1858922, EPI_ISL_1858924, EPI_ISL_1858926, EPI_ISL_1858928, EPI_ISL_1858930, EPI_ISL_1858933, EPI_ISL_1858934, EPI_ISL_1858936, EPI_ISL_1858938, EPI_ISL_1858940, EPI_ISL_1858942, EPI_ISL_1858944, EPI_ISL_1858946, EPI_ISL_1858948, EPI_ISL_1858950, EPI_ISL_1858952, EPI_ISL_1858954, EPI_ISL_1858957, EPI_ISL_1858960, EPI_ISL_1858963, EPI_ISL_1858964, EPI_ISL_1858966, EPI_ISL_1858968, EPI_ISL_1858969, EPI_ISL_1858970, EPI_ISL_1858971, EPI_ISL_1858973, EPI_ISL_1858975, EPI_ISL_1858977, EPI_ISL_1858979, EPI_ISL_1858981, EPI_ISL_1858984, EPI_ISL_1858986, EPI_ISL_1858988, EPI_ISL_1858990, EPI_ISL_1858992, EPI_ISL_1858994, EPI_ISL_1858996, EPI_ISL_1858998, EPI_ISL_1859000, EPI_ISL_1859002, EPI_ISL_1859005, EPI_ISL_1859007, EPI_ISL_1859008 | | | |
| see above | Unidade de apoio ao diagnóstico da COVID - UNADIG | Bioinformatics Laboratory / LNCC | Luiz G P de Almeida, Alessandra P Lamarca, Ronaldo da Silva F Jr, Liliane Cavalcante, Alexandra L Gerber, Ana Paula de C Guimarães, Douglas Terra Machado, Cassia Alves, Diana Mariani, Thais Felix Cruz, Mario Sergio Ribeiro, Silvia Carvalho, Flávio Dias da Silva, Marcio Henrique de Oliveira Garcia, Leandro Magalhães de Souza, Cristiane Gomes da Silva, Caio Luiz Pereira Ribeiro, Andréa Cony Cavalcanti, Claudia Maria Braga de Mello, Amilcar Tanuri, Ana Tereza R Vasconcelos |
| EPI_ISL_1966113 | CS II EGIDIO BRUNHARA MORRO AGUDO | Instituto Butantan / Mendelics | Instituto Butantan: Dimas Tadeu Covas, Sandra Coccuzzo Sampaio, Maria Carolina Elias, José Salvatore Leister Patané, Vincent Louis Viala, Antonio Jorge Martins, Ricardo Haddad, Claudia Renata dos Santos Barros, Elaine Cristina Marqueze, Raul Machado Neto, Debora Botequio Moretti, Jardelina de Souza Todao Bernardino, Loyze Paola Oliveira de Lima, Luiz Aurelio de Campos Crispin. Centro de Genômica Funcional da ESALQ: Luiz Lehmann Coutinho, Ricardo Augusto Brassaloti, Raquel de Lello Rocha Campos Cassano. NGS Soluções Genômicas: Pilar Drummond Sampaio Corrêa Mariani. FZEA-USP Pirassununga: Mirele Daiana Poleti, Jessika Cristina Chagas Lesbon, Elisangela Chicaroni Mattos, Heidge Fukumasu. USP-Botucatu: Rejane Maria Tommasini Grotto, Jayme A. Souza-Neto, Guilherme Targino Valente, Patricia Akemi Assato, Felipe Allan da Silva da Costa, Bianca Cechetto Carlos. Mendelics: Bibiana Santos, João Paulo Kitajima, Erika Freitas, David Schlesinger. Hemocentro Ribeirão Preto: Simone Kashima, Evandra Strazza Rodrigues, Svetoslav Nanev Slavov, Elaine Vieira dos Santos, Rafael dos Santos Bezerra, Luiz Carlos Junior de Alcantara, Marta Giovanetti, Vagner Fonseca, Flavia Aburjaile, Rodrigo Tocantins Calado. FAMERP-SJRP: Cecília Artico Banho, Lívia Sacchetto, Fábio Sossai Possebon, Leila Sabrina Ullmann, Cíntia Bittar, Guilherme Campos, Helena Lage Ferreira, Jorge A. Petroli Marchesi, Maisa C. Pereira Parra, Marília Moraes, Paula Rahal, Paulo Inacio da Costa, João Pessoa Araújo Jr., Maurício Lacerda Nogueira. Prefeitura de Sao Paulo: Melissa Palmieri. |
| EPI_ISL_1966358 | SMS SECRETARIA MUNICIPAL DE SAUDE DE BOITUVA | Instituto Butantan / Mendelics | Instituto Butantan: Dimas Tadeu Covas, Sandra Coccuzzo Sampaio, Maria Carolina Elias, José Salvatore Leister Patané, Vincent Louis Viala, Antonio Jorge Martins, Ricardo Haddad, Claudia Renata dos Santos Barros, Elaine Cristina Marqueze, Raul Machado Neto, Debora Botequio Moretti, Jardelina de Souza Todao Bernardino, Loyze Paola Oliveira de Lima, Luiz Aurelio de Campos Crispin. Centro de Genômica Funcional da ESALQ: Luiz Lehmann Coutinho, Ricardo Augusto Brassaloti, Raquel de Lello Rocha Campos Cassano. NGS Soluções Genômicas: Pilar Drummond Sampaio Corrêa Mariani. FZEA-USP Pirassununga: Mirele Daiana Poleti, Jessika Cristina Chagas Lesbon, Elisangela Chicaroni Mattos, Heidge Fukumasu. USP-Botucatu: Rejane Maria Tommasini Grotto, Jayme A. Souza-Neto, Guilherme Targino Valente, Patricia Akemi Assato, Felipe Allan da Silva da Costa, Bianca Cechetto Carlos. Mendelics: Bibiana Santos, João Paulo Kitajima, Erika Freitas, David Schlesinger. Hemocentro Ribeirão Preto: Simone Kashima, Evandra Strazza Rodrigues, Svetoslav Nanev Slavov, Elaine Vieira dos Santos, Rafael dos Santos Bezerra, Luiz Carlos Junior de Alcantara, Marta Giovanetti, Vagner Fonseca, Flavia Aburjaile, Rodrigo Tocantins Calado. FAMERP-SJRP: Cecília Artico Banho, Lívia Sacchetto, Fábio Sossai Possebon, Leila Sabrina Ullmann, Cíntia Bittar, Guilherme Campos, Helena Lage Ferreira, Jorge A. Petroli Marchesi, Maisa C. Pereira Parra, Marília Moraes, Paula Rahal, Paulo Inacio da Costa, João Pessoa Araújo Jr., Maurício Lacerda Nogueira. Prefeitura de Sao Paulo: Melissa Palmieri. |
| EPI_ISL_1966448 | NUCLEO DE SAUDE VILA FALCAO DE BAURU | Instituto Butantan / Mendelics | Instituto Butantan: Dimas Tadeu Covas, Sandra Coccuzzo Sampaio, Maria Carolina Elias, José Salvatore Leister Patané, Vincent Louis Viala, Antonio Jorge Martins, Ricardo Haddad, Claudia Renata dos Santos Barros, Elaine Cristina Marqueze, Raul Machado Neto, Debora Botequio Moretti, Jardelina de Souza Todao Bernardino, Loyze Paola Oliveira de Lima, Luiz Aurelio de Campos Crispin. Centro de Genômica Funcional da ESALQ: Luiz Lehmann Coutinho, Ricardo Augusto Brassaloti, Raquel de Lello Rocha Campos Cassano. NGS Soluções Genômicas: Pilar Drummond Sampaio Corrêa Mariani. FZEA-USP Pirassununga: Mirele Daiana Poleti, Jessika Cristina Chagas Lesbon, Elisangela Chicaroni Mattos, Heidge Fukumasu. USP-Botucatu: Rejane Maria Tommasini Grotto, Jayme A. Souza-Neto, Guilherme Targino Valente, Patricia Akemi Assato, Felipe Allan da Silva da Costa, Bianca Cechetto Carlos. Mendelics: Bibiana Santos, João Paulo Kitajima, Erika Freitas, David Schlesinger. Hemocentro Ribeirão Preto: Simone Kashima, Evandra Strazza Rodrigues, Svetoslav Nanev Slavov, Elaine Vieira dos Santos, Rafael dos Santos Bezerra, Luiz Carlos Junior de Alcantara, Marta Giovanetti, Vagner Fonseca, Flavia Aburjaile, Rodrigo Tocantins Calado. FAMERP-SJRP: Cecília Artico Banho, Lívia Sacchetto, Fábio Sossai Possebon, Leila Sabrina Ullmann, Cíntia Bittar, Guilherme Campos, Helena Lage Ferreira, Jorge A. Petroli Marchesi, Maisa C. Pereira Parra, Marília Moraes, Paula Rahal, Paulo Inacio da Costa, João Pessoa Araújo Jr., Maurício Lacerda Nogueira. Prefeitura de Sao Paulo: Melissa Palmieri. |

| | | | |
|---|---|---|---|
| EPI_ISL_1966526 | USF PAULISTA FERNANDOPOLIS ANTONIO PIVATO | Instituto Butantan / FZEA-USP (Pirassununga) | Instituto Butantan: Dimas Tadeu Covas, Sandra Coccuzzo Sampaio, Maria Carolina Elias, José Salvatore Leister Patané, Vincent Louis Viala, Antonio Jorge Martins, Ricardo Haddad, Claudia Renata dos Santos Barros, Elaine Cristina Marqueze, Raul Machado Neto, Debora Botequio Moretti, Jardelina de Souza Todao Bernardino, Loyze Paola Oliveira de Lima, Luiz Aurelio de Campos Crispin. Centro de Genômica Funcional da ESALQ: Luiz Lehmann Coutinho, Ricardo Augusto Brassaloti, Raquel de Lello Rocha Campos Cassano. NGS Soluções Genômicas: Pilar Drummond Sampaio Corrêa Mariani. FZEA-USP Pirassununga: Mirele Daiana Poleti, Jessika Cristina Chagas Lesbon, Elisangela Chicaroni Mattos, Heidge Fukumasu. USP-Botucatu: Rejane Maria Tommasini Grotto, Jayme A. Souza-Neto, Guilherme Targino Valente, Patricia Akemi Assato, Felipe Allan da Silva da Costa, Bianca Cechetto Carlos. Mendelics: Bibiana Santos, João Paulo Kitajima, Erika Freitas, David Schlesinger. Hemocentro Ribeirão Preto: Simone Kashima, Evandra Strazza Rodrigues, Svetoslav Nanev Slavov, Elaine Vieira dos Santos, Rafael dos Santos Bezerra, Luiz Carlos Junior de Alcantara, Marta Giovanetti, Vagner Fonseca, Flavia Aburjaile, Rodrigo Tocantins Calado. FAMERP-SJRP: Cecília Artico Banho, Lívia Sacchetto, Fábio Sossai Possebon, Leila Sabrina Ullmann, Cíntia Bittar, Guilherme Campos, Helena Lage Ferreira, Jorge A. Petroli Marchesi, Maisa C. Pereira Parra, Marília Moraes, Paula Rahal, Paulo Inacio da Costa, João Pessoa Araújo Jr., Maurício Lacerda Nogueira. Prefeitura de Sao Paulo: Melissa Palmieri. |
| EPI_ISL_1966567, EPI_ISL_1966578, EPI_ISL_1966580, EPI_ISL_1966581, EPI_ISL_1966582, EPI_ISL_1966585 | NUCLEO DE SAUDE VILA FALCAO DE BAURU | Instituto Butantan / Mendelics | Instituto Butantan: Dimas Tadeu Covas, Sandra Coccuzzo Sampaio, Maria Carolina Elias, José Salvatore Leister Patané, Vincent Louis Viala, Antonio Jorge Martins, Ricardo Haddad, Claudia Renata dos Santos Barros, Elaine Cristina Marqueze, Raul Machado Neto, Debora Botequio Moretti, Jardelina de Souza Todao Bernardino, Loyze Paola Oliveira de Lima, Luiz Aurelio de Campos Crispin. Centro de Genômica Funcional da ESALQ: Luiz Lehmann Coutinho, Ricardo Augusto Brassaloti, Raquel de Lello Rocha Campos Cassano. NGS Soluções Genômicas: Pilar Drummond Sampaio Corrêa Mariani. FZEA-USP Pirassununga: Mirele Daiana Poleti, Jessika Cristina Chagas Lesbon, Elisangela Chicaroni Mattos, Heidge Fukumasu. USP-Botucatu: Rejane Maria Tommasini Grotto, Jayme A. Souza-Neto, Guilherme Targino Valente, Patricia Akemi Assato, Felipe Allan da Silva da Costa, Bianca Cechetto Carlos. Mendelics: Bibiana Santos, João Paulo Kitajima, Erika Freitas, David Schlesinger. Hemocentro Ribeirão Preto: Simone Kashima, Evandra Strazza Rodrigues, Svetoslav Nanev Slavov, Elaine Vieira dos Santos, Rafael dos Santos Bezerra, Luiz Carlos Junior de Alcantara, Marta Giovanetti, Vagner Fonseca, Flavia Aburjaile, Rodrigo Tocantins Calado. FAMERP-SJRP: Cecília Artico Banho, Lívia Sacchetto, Fábio Sossai Possebon, Leila Sabrina Ullmann, Cíntia Bittar, Guilherme Campos, Helena Lage Ferreira, Jorge A. Petroli Marchesi, Maisa C. Pereira Parra, Marília Moraes, Paula Rahal, Paulo Inacio da Costa, João Pessoa Araújo Jr., Maurício Lacerda Nogueira. Prefeitura de Sao Paulo: Melissa Palmieri. |
| EPI_ISL_1966658 | UPA UNIDADE DE PRONTO ATENDIMENTO 24 HORAS BOM JESUS | Instituto Butantan / Mendelics | Instituto Butantan: Dimas Tadeu Covas, Sandra Coccuzzo Sampaio, Maria Carolina Elias, José Salvatore Leister Patané, Vincent Louis Viala, Antonio Jorge Martins, Ricardo Haddad, Claudia Renata dos Santos Barros, Elaine Cristina Marqueze, Raul Machado Neto, Debora Botequio Moretti, Jardelina de Souza Todao Bernardino, Loyze Paola Oliveira de Lima, Luiz Aurelio de Campos Crispin. Centro de Genômica Funcional da ESALQ: Luiz Lehmann Coutinho, Ricardo Augusto Brassaloti, Raquel de Lello Rocha Campos Cassano. NGS Soluções Genômicas: Pilar Drummond Sampaio Corrêa Mariani. FZEA-USP Pirassununga: Mirele Daiana Poleti, Jessika Cristina Chagas Lesbon, Elisangela Chicaroni Mattos, Heidge Fukumasu. USP-Botucatu: Rejane Maria Tommasini Grotto, Jayme A. Souza-Neto, Guilherme Targino Valente, Patricia Akemi Assato, Felipe Allan da Silva da Costa, Bianca Cechetto Carlos. Mendelics: Bibiana Santos, João Paulo Kitajima, Erika Freitas, David Schlesinger. Hemocentro Ribeirão Preto: Simone Kashima, Evandra Strazza Rodrigues, Svetoslav Nanev Slavov, Elaine Vieira dos Santos, Rafael dos Santos Bezerra, Luiz Carlos Junior de Alcantara, Marta Giovanetti, Vagner Fonseca, Flavia Aburjaile, Rodrigo Tocantins Calado. FAMERP-SJRP: Cecília Artico Banho, Lívia Sacchetto, Fábio Sossai Possebon, Leila Sabrina Ullmann, Cíntia Bittar, Guilherme Campos, Helena Lage Ferreira, Jorge A. Petroli Marchesi, Maisa C. Pereira Parra, Marília Moraes, Paula Rahal, Paulo Inacio da Costa, João Pessoa Araújo Jr., Maurício Lacerda Nogueira. Prefeitura de Sao Paulo: Melissa Palmieri. |
| EPI_ISL_1966832 | ESTRATEGIA DE SAUDE DA FAMILIA DE ARCO IRIS | Instituto Butantan / ESALQ-USP (Piracicaba) | Instituto Butantan: Dimas Tadeu Covas, Sandra Coccuzzo Sampaio, Maria Carolina Elias, José Salvatore Leister Patané, Vincent Louis Viala, Antonio Jorge Martins, Ricardo Haddad, Claudia Renata dos Santos Barros, Elaine Cristina Marqueze, Raul Machado Neto, Debora Botequio Moretti, Jardelina de Souza Todao Bernardino, Loyze Paola Oliveira de Lima, Luiz Aurelio de Campos Crispin. Centro de Genômica Funcional da ESALQ: Luiz Lehmann Coutinho, Ricardo Augusto Brassaloti, Raquel de Lello Rocha Campos Cassano. NGS Soluções Genômicas: Pilar Drummond Sampaio Corrêa Mariani. FZEA-USP Pirassununga: Mirele Daiana Poleti, Jessika Cristina Chagas Lesbon, Elisangela Chicaroni Mattos, Heidge Fukumasu. USP-Botucatu: Rejane Maria Tommasini Grotto, Jayme A. Souza-Neto, Guilherme Targino Valente, Patricia Akemi Assato, Felipe Allan da Silva da Costa, Bianca Cechetto Carlos. Mendelics: Bibiana Santos, João Paulo Kitajima, Erika Freitas, David Schlesinger. Hemocentro Ribeirão Preto: Simone Kashima, Evandra Strazza Rodrigues, Svetoslav Nanev Slavov, Elaine Vieira dos Santos, Rafael dos Santos Bezerra, Luiz Carlos Junior de Alcantara, Marta Giovanetti, Vagner Fonseca, Flavia Aburjaile, Rodrigo Tocantins Calado. FAMERP-SJRP: Cecília Artico Banho, Lívia Sacchetto, Fábio Sossai Possebon, Leila Sabrina Ullmann, Cíntia Bittar, Guilherme Campos, Helena Lage Ferreira, Jorge A. Petroli Marchesi, Maisa C. Pereira Parra, Marília Moraes, Paula Rahal, Paulo Inacio da Costa, João Pessoa Araújo Jr., Maurício Lacerda Nogueira. Prefeitura de Sao Paulo: Melissa Palmieri. |
| EPI_ISL_1966901 | CENTRO DE APOIO EPIDEMIOLOGICO | Instituto Butantan / ESALQ-USP (Piracicaba) | Instituto Butantan: Dimas Tadeu Covas, Sandra Coccuzzo Sampaio, Maria Carolina Elias, José Salvatore Leister Patané, Vincent Louis Viala, Antonio Jorge Martins, Ricardo Haddad, Claudia Renata dos Santos Barros, Elaine Cristina Marqueze, Raul Machado Neto, Debora Botequio Moretti, Jardelina de Souza Todao Bernardino, Loyze Paola Oliveira de Lima, Luiz Aurelio de Campos Crispin. Centro de Genômica Funcional da ESALQ: Luiz Lehmann Coutinho, Ricardo Augusto Brassaloti, Raquel de Lello Rocha Campos Cassano. NGS Soluções Genômicas: Pilar Drummond Sampaio Corrêa Mariani. FZEA-USP Pirassununga: Mirele Daiana Poleti, Jessika Cristina Chagas Lesbon, Elisangela Chicaroni Mattos, Heidge Fukumasu. USP-Botucatu: Rejane Maria Tommasini Grotto, Jayme A. Souza-Neto, Guilherme Targino Valente, Patricia Akemi Assato, Felipe Allan da Silva da Costa, Bianca Cechetto Carlos. Mendelics: Bibiana Santos, João Paulo Kitajima, Erika Freitas, David Schlesinger. Hemocentro Ribeirão Preto: Simone Kashima, Evandra Strazza Rodrigues, Svetoslav Nanev Slavov, Elaine Vieira dos Santos, Rafael dos Santos Bezerra, Luiz Carlos Junior de Alcantara, Marta Giovanetti, Vagner Fonseca, Flavia Aburjaile, Rodrigo Tocantins Calado. FAMERP-SJRP: Cecília Artico Banho, Lívia Sacchetto, Fábio Sossai Possebon, Leila Sabrina Ullmann, Cíntia Bittar, Guilherme Campos, Helena Lage Ferreira, Jorge A. Petroli Marchesi, Maisa C. Pereira Parra, Marília Moraes, Paula Rahal, Paulo Inacio da Costa, João Pessoa Araújo Jr., Maurício Lacerda Nogueira. Prefeitura de Sao Paulo: Melissa Palmieri. |
| EPI_ISL_1966926 | HOSPITAL REGIONAL DE ITAPETININGA | Instituto Butantan / ESALQ-USP (Piracicaba) | Instituto Butantan: Dimas Tadeu Covas, Sandra Coccuzzo Sampaio, Maria Carolina Elias, José Salvatore Leister Patané, Vincent Louis Viala, Antonio Jorge Martins, Ricardo Haddad, Claudia Renata dos Santos Barros, Elaine Cristina Marqueze, Raul Machado Neto, Debora Botequio Moretti, Jardelina de Souza Todao Bernardino, Loyze Paola Oliveira de Lima, Luiz Aurelio de Campos Crispin. Centro de Genômica Funcional da ESALQ: Luiz Lehmann Coutinho, Ricardo Augusto Brassaloti, Raquel de Lello Rocha Campos Cassano. NGS Soluções Genômicas: Pilar Drummond Sampaio Corrêa Mariani. FZEA-USP Pirassununga: Mirele Daiana Poleti, Jessika Cristina Chagas Lesbon, Elisangela Chicaroni Mattos, Heidge Fukumasu. USP-Botucatu: Rejane Maria Tommasini Grotto, Jayme A. Souza-Neto, Guilherme Targino Valente, Patricia Akemi Assato, Felipe Allan da Silva da Costa, Bianca Cechetto Carlos. Mendelics: Bibiana Santos, João Paulo Kitajima, Erika Freitas, David Schlesinger. Hemocentro Ribeirão Preto: Simone Kashima, Evandra Strazza Rodrigues, Svetoslav Nanev Slavov, Elaine Vieira dos Santos, Rafael dos Santos Bezerra, Luiz Carlos Junior de Alcantara, Marta Giovanetti, Vagner Fonseca, Flavia Aburjaile, Rodrigo Tocantins Calado. FAMERP-SJRP: Cecília Artico Banho, Lívia Sacchetto, Fábio Sossai Possebon, Leila Sabrina Ullmann, Cíntia Bittar, Guilherme Campos, Helena Lage Ferreira, Jorge A. Petroli Marchesi, Maisa C. Pereira Parra, Marília Moraes, Paula Rahal, Paulo Inacio da Costa, João Pessoa Araújo Jr., Maurício Lacerda Nogueira. Prefeitura de Sao Paulo: Melissa Palmieri. |
| EPI_ISL_1967027 | CENTRO DE SAUDE II | Instituto Butantan / ESALQ-USP (Piracicaba) | Instituto Butantan: Dimas Tadeu Covas, Sandra Coccuzzo Sampaio, Maria Carolina Elias, José Salvatore Leister Patané, Vincent Louis Viala, Antonio Jorge Martins, Ricardo Haddad, Claudia Renata dos Santos Barros, Elaine Cristina Marqueze, Raul Machado Neto, Debora Botequio Moretti, Jardelina de Souza Todao Bernardino, Loyze Paola Oliveira de Lima, Luiz Aurelio de Campos Crispin. Centro de Genômica Funcional da ESALQ: Luiz Lehmann Coutinho, Ricardo Augusto Brassaloti, Raquel de Lello Rocha Campos Cassano. NGS Soluções Genômicas: Pilar Drummond Sampaio Corrêa Mariani. FZEA-USP Pirassununga: Mirele Daiana Poleti, Jessika Cristina Chagas Lesbon, Elisangela Chicaroni Mattos, Heidge Fukumasu. USP-Botucatu: Rejane Maria Tommasini Grotto, Jayme A. Souza-Neto, Guilherme Targino Valente, Patricia Akemi Assato, Felipe Allan da Silva da Costa, Bianca Cechetto Carlos. Mendelics: Bibiana Santos, João Paulo Kitajima, Erika Freitas, David Schlesinger. Hemocentro Ribeirão Preto: Simone Kashima, Evandra Strazza Rodrigues, Svetoslav Nanev Slavov, Elaine Vieira dos Santos, Rafael dos Santos Bezerra, Luiz Carlos Junior de Alcantara, Marta Giovanetti, Vagner Fonseca, Flavia Aburjaile, Rodrigo Tocantins Calado. FAMERP-SJRP: Cecília Artico Banho, Lívia Sacchetto, Fábio Sossai Possebon, Leila Sabrina Ullmann, Cíntia Bittar, Guilherme Campos, Helena Lage Ferreira, Jorge A. Petroli Marchesi, Maisa C. Pereira Parra, Marília Moraes, Paula Rahal, Paulo Inacio da Costa, João Pessoa Araújo Jr., Maurício Lacerda Nogueira. Prefeitura de Sao Paulo: Melissa Palmieri. |
| EPI_ISL_1967164 | UBS ENRIQUE GUILHERME ERNESTO MARZEUSKI | Instituto Butantan / ESALQ-USP (Piracicaba) | Instituto Butantan: Dimas Tadeu Covas, Sandra Coccuzzo Sampaio, Maria Carolina Elias, José Salvatore Leister Patané, Vincent Louis Viala, Antonio Jorge Martins, Ricardo Haddad, Claudia Renata dos Santos Barros, Elaine Cristina Marqueze, Raul Machado Neto, Debora Botequio Moretti, Jardelina de Souza Todao Bernardino, Loyze Paola Oliveira de Lima, Luiz Aurelio de Campos Crispin. Centro de Genômica Funcional da ESALQ: Luiz Lehmann Coutinho, Ricardo Augusto Brassaloti, Raquel de Lello Rocha Campos Cassano. NGS Soluções Genômicas: Pilar Drummond Sampaio Corrêa Mariani. FZEA-USP Pirassununga: Mirele Daiana Poleti, Jessika Cristina Chagas Lesbon, Elisangela Chicaroni Mattos, Heidge Fukumasu. USP-Botucatu: Rejane Maria Tommasini Grotto, Jayme A. Souza-Neto, Guilherme Targino Valente, Patricia Akemi Assato, Felipe Allan da Silva da Costa, Bianca Cechetto Carlos. Mendelics: Bibiana Santos, João Paulo Kitajima, Erika Freitas, David Schlesinger. Hemocentro Ribeirão Preto: Simone Kashima, Evandra Strazza |

| | | | |
|---|---|---|---|
| | | | Rodrigues, Svetoslav Nanev Slavov, Elaine Vieira dos Santos, Rafael dos Santos Bezerra, Luiz Carlos Junior de Alcantara, Marta Giovanetti, Vagner Fonseca, Flavia Aburjaile, Rodrigo Tocantins Calado. FAMERP-SJRP: Cecília Artico Banho, Lívia Sacchetto, Fábio Sossai Possebon, Leila Sabrina Ullmann, Cíntia Bittar, Guilherme Campos, Helena Lage Ferreira, Jorge A. Petroli Marchesi, Maisa C. Pereira Parra, Marília Moraes, Paula Rahal, Paulo Inacio da Costa, João Pessoa Araújo Jr., Maurício Lacerda Nogueira. Prefeitura de Sao Paulo: Melissa Palmieri. |
| EPI_ISL_1967284 | UBS RECREIO SAO JORGE | Instituto Butantan / Mendelics | Instituto Butantan: Dimas Tadeu Covas, Sandra Coccuzzo Sampaio, Maria Carolina Elias, José Salvatore Leister Patané, Vincent Louis Viala, Antonio Jorge Martins, Ricardo Haddad, Claudia Renata dos Santos Barros, Elaine Cristina Marqueze, Raul Machado Neto, Debora Botequio Moretti, Jardelina de Souza Todao Bernardino, Loyze Paola Oliveira de Lima, Luiz Aurelio de Campos Crispin. Centro de Genômica Funcional da ESALQ: Luiz Lehmann Coutinho, Ricardo Augusto Brassaloti, Raquel de Lello Rocha Campos Cassano. NGS Soluções Genômicas: Pilar Drummond Sampaio Corrêa Mariani. FZEA-USP Pirassununga: Mirele Daiana Poleti, Jessika Cristina Chagas Lesbon, Elisangela Chicaroni Mattos, Heidge Fukumasu. USP-Botucatu: Rejane Maria Tommasini Grotto, Jayme A. Souza-Neto, Guilherme Targino Valente, Patricia Akemi Assato, Felipe Allan da Silva da Costa, Bianca Cechetto Carlos. Mendelics: Bibiana Santos, João Paulo Kitajima, Erika Freitas, David Schlesinger. Hemocentro Ribeirão Preto: Simone Kashima, Evandra Strazza Rodrigues, Svetoslav Nanev Slavov, Elaine Vieira dos Santos, Rafael dos Santos Bezerra, Luiz Carlos Junior de Alcantara, Marta Giovanetti, Vagner Fonseca, Flavia Aburjaile, Rodrigo Tocantins Calado. FAMERP-SJRP: Cecília Artico Banho, Lívia Sacchetto, Fábio Sossai Possebon, Leila Sabrina Ullmann, Cíntia Bittar, Guilherme Campos, Helena Lage Ferreira, Jorge A. Petroli Marchesi, Maisa C. Pereira Parra, Marília Moraes, Paula Rahal, Paulo Inacio da Costa, João Pessoa Araújo Jr., Maurício Lacerda Nogueira. Prefeitura de Sao Paulo: Melissa Palmieri. |
| EPI_ISL_2003120, EPI_ISL_2003123 | Instituto Adolfo Lutz Central | Instituto Adolfo Lutz, Interdiciplinary Procedures Center, Strategic Laboratory | Claudio Tavares Sacchi, Claudia Regina Gonçalves, Erica Valessa Ramos Gomes, Karoline Rodrigues Campos, Caio Vinicius Dias Lopes, Leonardo Jose Tadeu de Araujo |
| EPI_ISL_2020090, EPI_ISL_2020097, EPI_ISL_2020105 | HOSPITAL UNIVERSITARIO SON ESPASES | HOSPITAL UNIVERSITARIO SON ESPASES | Carla López-Causapé, Pablo Fraile-Ribot, Antonio Oliver, SeqCovid |
| EPI_ISL_2031726 | Centro de Innovación en Vigilancia Epidemiológica (CiVE), Institut Pasteur Montevideo, Uruguay | Centro de Innovación en Vigilancia Epidemiológica (CiVE), Institut Pasteur Montevideo, Uruguay | Natalia Rego, Alicia Costábile, Mercedes Paz, Cecilia Salazar, Paula Perbolianachis, Tamara Fernández, Ignacio Ferrés, Rodrigo Arce, Alvaro Fajardo, Mailen Arleo, Tania Possi, Inés Bellini, Lucia Bilbao, Natalia Reyes, Ma Noel Bentancor, Andrés Lizosaín, María José Benítez, Odhille Chappos, Melissa Duquía, Belén González, Luciana Griffero, Mauricio Méndez, Ma Pía Techera, Juan Zanetti, Bernardina Rivera, Matías Maidana, Martina Alonso, Cecilia Alonso, Julio Medina, Henry Albornoz, Rodney Colina, Gregorio Iraola, Lucía Spangenberg, Gonzalo Moratorio, Pilar Moreno |
| EPI_ISL_2037442 | Instituto de Virologia "Dr. J. M. Vanella", Facultad de Ciencias Médicas, Universidad Nacional de Córdoba. | Centro de Investigaciones Agropecuarias (CIAP), Instituto Nacional de Tecnologia Agropecuaria (INTA) Cordoba, Argentina, on behalf of 'Proyecto Argentino Interinstitucional de Genomica de SARS-CoV-2 (PAIS Consortium) | Brenda Konigheim, Lorena Spinsanti, Adrian Diaz, Javier Aguilar, Sebastian Blanco, Mauricio Beranek, Maria Elisa Rivarola, Maria Belen Pisano, Viviana Re, Gonzalo Castro, Gabriela Barbas, Franco Fernandez, Humberto Debat, Nathalie Marquez, Sandra Gallego. |
| EPI_ISL_2038956 | Laboratorio Central de Saude Publica do Estado do Rio de Janeiro (LACEN-RJ) | Laboratory of Respiratory Viruses and Measles, Oswaldo Cruz Institute, FIOCRUZ | Paola Resende, Luciana Appolinario, Fernando Motta, Anna Carolina Paixao, Ana Carolina Mendonca, Alice Sampaio Rocha, Taina Venas, Elisa Cavalcante Pereira, Renata Serrano Lopes, Andrea Cony Cavalcanti, Marilda Siqueira on behalf of the Fiocruz COVID-19 Genomic Surveillance Network |
| EPI_ISL_2038961, EPI_ISL_2038962, EPI_ISL_2038963 | Laboratory of Respiratory Viruses and Measles, Oswaldo Cruz Institute, FIOCRUZ | Laboratory of Respiratory Viruses and Measles, Oswaldo Cruz Institute, FIOCRUZ | Paola Resende, Luciana Appolinario, Fernando Motta, Anna Carolina Paixao, Ana Carolina Mendonca, Alice Sampaio Rocha, Taina Venas, Elisa Cavalcante Pereira, Renata Serrano Lopes, Marilda Siqueira on behalf of the Fiocruz COVID-19 Genomic Surveillance Network |
| EPI_ISL_2086268 | Laboratory of Molecular Biology of Flavivirus, Oswaldo Cruz Institute, FIOCRUZ | Laboratory of Molecular Biology of Flavivirus, Oswaldo Cruz Institute, FIOCRUZ | Fernando Lopez Tort, Ieda Ribeiro, Lidiane Menezes, Alexandre dos Santos, Marta Pereira, Myrna Bonaldo |
| EPI_ISL_2093854, EPI_ISL_2093855, EPI_ISL_2093871 | Dutch COVID-19 response team | National Institute for Public Health and the Environment (RIVM) | Adam Meijer, Harry Vennema, Dirk Eggink, Jeroen Cremer, Sharon van den Brink, Bas van der Veer, AnneMarie van den Brandt, Lisa Wijsman, Kim Freriks, Ryanne Jaarsma, Euníce Then, Lynn Aarts, Sanne Bos, Melissa van Tuil, Linda van de Nes, Sjoerd Kuiling, James Groot, Florian Zwagemaker, Dennis Schmitz, Annelies Kroneman, Karim Hajji, Chantal Reusken, on behalf of the national COVID-19 response team |
| EPI_ISL_2101392 | Unidade de apoio ao diagnóstico da COVID - UNADIG | Bioinformatics Laboratory / LNCC | Luiz G P de Almeida, Alessandra P Lamarca, Ronaldo da Silva F Jr, Liliane Cavalcante, Alexandra L Gerber, Ana Paula de C Guimaraes, Douglas Terra Machado, Cassia Alves, Diana Mariani, Cintia Policarpo, Gleidson da Silva de Oliveira, Mario Sergio Ribeiro, Silvia Carvalho, Flavio Dias da Silva, Marcio Henrique de Oliveira Garcia, Leandro Magalhaes de Souza, Cristiane Gomes da Silva, Caio Luiz Pereira Ribeiro, Andrea Cony Cavalcanti, Claudia Maria Braga de Mello, Amilcar Tanuri, Ana Tereza R Vasconcelos |
| EPI_ISL_2101393 | Laboratorio Central Noel Nutels | Bioinformatics Laboratory / LNCC | Luiz G P de Almeida, Alessandra P Lamarca, Ronaldo da Silva F Jr, Liliane Cavalcante, Alexandra L Gerber, Ana Paula de C Guimaraes, Douglas Terra Machado, Cassia Alves, Diana Mariani, Cintia Policarpo, Gleidson da Silva de Oliveira, Mario Sergio Ribeiro, Silvia Carvalho, Flavio Dias da Silva, Marcio Henrique de Oliveira Garcia, Leandro Magalhaes de Souza, Cristiane Gomes da Silva, Caio Luiz Pereira Ribeiro, Andrea Cony Cavalcanti, Claudia Maria Braga de Mello, Amilcar Tanuri, Ana Tereza R Vasconcelos |
| EPI_ISL_2101394 | Unidade de apoio ao diagnóstico da COVID - UNADIG | Bioinformatics Laboratory / LNCC | Luiz G P de Almeida, Alessandra P Lamarca, Ronaldo da Silva F Jr, Liliane Cavalcante, Alexandra L Gerber, Ana Paula de C Guimaraes, Douglas Terra Machado, Cassia Alves, Diana Mariani, Cintia Policarpo, Gleidson da Silva de Oliveira, Mario Sergio Ribeiro, Silvia Carvalho, Flavio Dias da Silva, Marcio Henrique de Oliveira Garcia, Leandro Magalhaes de Souza, Cristiane Gomes da Silva, Caio Luiz Pereira Ribeiro, Andrea Cony Cavalcanti, Claudia Maria Braga de Mello, Amilcar Tanuri, Ana Tereza R Vasconcelos |
| EPI_ISL_2101395, EPI_ISL_2101396, EPI_ISL_2101397, EPI_ISL_2101398, EPI_ISL_2101399, EPI_ISL_2101400, EPI_ISL_2101401, EPI_ISL_2101403, EPI_ISL_2101404, EPI_ISL_2101405, EPI_ISL_2101406, EPI_ISL_2101410, EPI_ISL_2101411, EPI_ISL_2101412, EPI_ISL_2101413, EPI_ISL_2101414, EPI_ISL_2101415, EPI_ISL_2101416, EPI_ISL_2101417, EPI_ISL_2101418, EPI_ISL_2101419, EPI_ISL_2101420, EPI_ISL_2101421, EPI_ISL_2101422, EPI_ISL_2101423, EPI_ISL_2101424, EPI_ISL_2101425, EPI_ISL_2101426, EPI_ISL_2101427, EPI_ISL_2101428 | | | |
| see above | Laboratorio Central Noel Nutels | Bioinformatics Laboratory / LNCC | Luiz G P de Almeida, Alessandra P Lamarca, Ronaldo da Silva F Jr, Liliane Cavalcante, Alexandra L Gerber, Ana Paula de C Guimaraes, Douglas Terra Machado, Cassia Alves, Diana Mariani, Cintia Policarpo, Gleidson da Silva de Oliveira, Mario Sergio Ribeiro, Silvia Carvalho, Flavio Dias da Silva, Marcio Henrique de Oliveira Garcia, Leandro Magalhaes de Souza, Cristiane Gomes da Silva, Caio Luiz Pereira Ribeiro, Andrea Cony Cavalcanti, Claudia Maria Braga de Mello, Amilcar Tanuri, Ana Tereza R Vasconcelos |
| EPI_ISL_2101429, EPI_ISL_2101430, EPI_ISL_2101431, EPI_ISL_2101432, EPI_ISL_2101433 | Unidade de apoio ao diagnóstico da COVID - UNADIG | Bioinformatics Laboratory / LNCC | Luiz G P de Almeida, Alessandra P Lamarca, Ronaldo da Silva F Jr, Liliane Cavalcante, Alexandra L Gerber, Ana Paula de C Guimaraes, Douglas Terra Machado, Cassia Alves, Diana Mariani, Cintia Policarpo, Gleidson da Silva de Oliveira, Mario Sergio Ribeiro, Silvia Carvalho, Flavio Dias da Silva, Marcio Henrique de Oliveira Garcia, Leandro Magalhaes de Souza, Cristiane Gomes da Silva, Caio Luiz Pereira Ribeiro, Andrea Cony Cavalcanti, Claudia Maria Braga de Mello, Amilcar Tanuri, Ana Tereza R Vasconcelos |
| EPI_ISL_2101434, EPI_ISL_2101435, EPI_ISL_2101436, EPI_ISL_2101437, EPI_ISL_2101438, EPI_ISL_2101439, EPI_ISL_2101440, EPI_ISL_2101441, EPI_ISL_2101442 | Laboratorio Central Noel Nutels | Bioinformatics Laboratory / LNCC | Luiz G P de Almeida, Alessandra P Lamarca, Ronaldo da Silva F Jr, Liliane Cavalcante, Alexandra L Gerber, Ana Paula de C Guimaraes, Douglas Terra Machado, Cassia Alves, Diana Mariani, Cintia Policarpo, Gleidson da Silva de Oliveira, Mario Sergio Ribeiro, Silvia Carvalho, Flavio Dias da Silva, Marcio Henrique de Oliveira Garcia, Leandro Magalhaes de Souza, Cristiane Gomes da Silva, Caio Luiz Pereira Ribeiro, Andrea Cony Cavalcanti, Claudia Maria Braga de Mello, Amilcar Tanuri, Ana Tereza R Vasconcelos |
| EPI_ISL_2101443 | Unidade de apoio ao diagnóstico da COVID - UNADIG | Bioinformatics Laboratory / LNCC | Luiz G P de Almeida, Alessandra P Lamarca, Ronaldo da Silva F Jr, Liliane Cavalcante, Alexandra L Gerber, Ana Paula de C Guimaraes, Douglas Terra Machado, Cassia Alves, Diana Mariani, Cintia Policarpo, Gleidson da Silva de Oliveira, Mario Sergio Ribeiro, Silvia Carvalho, Flavio Dias da Silva, Marcio Henrique de Oliveira Garcia, Leandro Magalhaes de Souza, Cristiane Gomes da Silva, Caio Luiz Pereira Ribeiro, Andrea Cony Cavalcanti, Claudia Maria Braga de Mello, Amilcar Tanuri, Ana Tereza R Vasconcelos |
| EPI_ISL_2101444, EPI_ISL_2101445, EPI_ISL_2101446, EPI_ISL_2101447, EPI_ISL_2101448, EPI_ISL_2101449, EPI_ISL_2101450, EPI_ISL_2101451, EPI_ISL_2101452, EPI_ISL_2101453, EPI_ISL_2101454 | | | |
| see above | Laboratorio Central Noel Nutels | Bioinformatics Laboratory / LNCC | Luiz G P de Almeida, Alessandra P Lamarca, Ronaldo da Silva F Jr, Liliane Cavalcante, Alexandra L Gerber, Ana Paula de C Guimaraes, Douglas Terra Machado, Cassia Alves, Diana Mariani, Cintia Policarpo, Gleidson da Silva de Oliveira, Mario Sergio Ribeiro, Silvia Carvalho, Flavio Dias da Silva, Marcio Henrique de Oliveira Garcia, Leandro Magalhaes de Souza, Cristiane Gomes da Silva, Caio Luiz Pereira Ribeiro, Andrea Cony Cavalcanti, Claudia Maria Braga de Mello, Amilcar Tanuri, Ana Tereza R Vasconcelos |
| EPI_ISL_2101455, EPI_ISL_2101456, EPI_ISL_2101457, EPI_ISL_2101458, EPI_ISL_2101459, EPI_ISL_2101460, EPI_ISL_2101461, EPI_ISL_2101462, EPI_ISL_2101463 | Unidade de apoio ao diagnóstico da COVID - UNADIG | Bioinformatics Laboratory / LNCC | Luiz G P de Almeida, Alessandra P Lamarca, Ronaldo da Silva F Jr, Liliane Cavalcante, Alexandra L Gerber, Ana Paula de C Guimaraes, Douglas Terra Machado, Cassia Alves, Diana Mariani, Cintia Policarpo, Gleidson da Silva de Oliveira, Mario Sergio Ribeiro, Silvia Carvalho, Flavio Dias da Silva, Marcio Henrique de Oliveira Garcia, Leandro Magalhaes de Souza, Cristiane Gomes da Silva, Caio Luiz Pereira Ribeiro, Andrea Cony Cavalcanti, Claudia Maria Braga de Mello, Amilcar Tanuri, Ana Tereza R Vasconcelos |

| | | | |
|---|---|---|---|
| EPI_ISL_2101464, EPI_ISL_2101465, EPI_ISL_2101466, EPI_ISL_2101467, EPI_ISL_2101468, EPI_ISL_2101469 | Laboratorio Central Noel Nutels | Bioinformatics Laboratory / LNCC | Luiz G P de Almeida, Alessandra P Lamarca, Ronaldo da Silva F Jr, Liliane Cavalcante, Alexandra L Gerber, Ana Paula de C Guimaraes, Douglas Terra Machado, Cassia Alves, Diana Mariani, Cintia Policarpo, Gleidson da Silva de Oliveira, Mario Sergio Ribeiro, Silvia Carvalho, Flavio Dias da Silva, Marcio Henrique de Oliveira Garcia, Leandro Magalhaes de Souza, Cristiane Gomes da Silva, Caio Luiz Pereira Ribeiro, Andrea Cony Cavalcanti, Claudia Maria Braga de Mello, Amilcar Tanuri, Ana Tereza R Vasconcelos |
| EPI_ISL_2101470, EPI_ISL_2101471, EPI_ISL_2101472, EPI_ISL_2101473 | Unidade de apoio ao diagnóstico da COVID - UNADIG | Bioinformatics Laboratory / LNCC | Luiz G P de Almeida, Alessandra P Lamarca, Ronaldo da Silva F Jr, Liliane Cavalcante, Alexandra L Gerber, Ana Paula de C Guimaraes, Douglas Terra Machado, Cassia Alves, Diana Mariani, Cintia Policarpo, Gleidson da Silva de Oliveira, Mario Sergio Ribeiro, Silvia Carvalho, Flavio Dias da Silva, Marcio Henrique de Oliveira Garcia, Leandro Magalhaes de Souza, Cristiane Gomes da Silva, Caio Luiz Pereira Ribeiro, Andrea Cony Cavalcanti, Claudia Maria Braga de Mello, Amilcar Tanuri, Ana Tereza R Vasconcelos |
| EPI_ISL_2101474 | Laboratorio Central Noel Nutels | Bioinformatics Laboratory / LNCC | Luiz G P de Almeida, Alessandra P Lamarca, Ronaldo da Silva F Jr, Liliane Cavalcante, Alexandra L Gerber, Ana Paula de C Guimaraes, Douglas Terra Machado, Cassia Alves, Diana Mariani, Cintia Policarpo, Gleidson da Silva de Oliveira, Mario Sergio Ribeiro, Silvia Carvalho, Flavio Dias da Silva, Marcio Henrique de Oliveira Garcia, Leandro Magalhaes de Souza, Cristiane Gomes da Silva, Caio Luiz Pereira Ribeiro, Andrea Cony Cavalcanti, Claudia Maria Braga de Mello, Amilcar Tanuri, Ana Tereza R Vasconcelos |
| EPI_ISL_2101475 | Unidade de apoio ao diagnóstico da COVID - UNADIG | Bioinformatics Laboratory / LNCC | Luiz G P de Almeida, Alessandra P Lamarca, Ronaldo da Silva F Jr, Liliane Cavalcante, Alexandra L Gerber, Ana Paula de C Guimaraes, Douglas Terra Machado, Cassia Alves, Diana Mariani, Cintia Policarpo, Gleidson da Silva de Oliveira, Mario Sergio Ribeiro, Silvia Carvalho, Flavio Dias da Silva, Marcio Henrique de Oliveira Garcia, Leandro Magalhaes de Souza, Cristiane Gomes da Silva, Caio Luiz Pereira Ribeiro, Andrea Cony Cavalcanti, Claudia Maria Braga de Mello, Amilcar Tanuri, Ana Tereza R Vasconcelos |
| EPI_ISL_2101476, EPI_ISL_2101477, EPI_ISL_2101478, EPI_ISL_2101479, EPI_ISL_2101480, EPI_ISL_2101481, EPI_ISL_2101482 | Laboratorio Central Noel Nutels | Bioinformatics Laboratory / LNCC | Luiz G P de Almeida, Alessandra P Lamarca, Ronaldo da Silva F Jr, Liliane Cavalcante, Alexandra L Gerber, Ana Paula de C Guimaraes, Douglas Terra Machado, Cassia Alves, Diana Mariani, Cintia Policarpo, Gleidson da Silva de Oliveira, Mario Sergio Ribeiro, Silvia Carvalho, Flavio Dias da Silva, Marcio Henrique de Oliveira Garcia, Leandro Magalhaes de Souza, Cristiane Gomes da Silva, Caio Luiz Pereira Ribeiro, Andrea Cony Cavalcanti, Claudia Maria Braga de Mello, Amilcar Tanuri, Ana Tereza R Vasconcelos |
| EPI_ISL_2101483, EPI_ISL_2101484 | Unidade de apoio ao diagnóstico da COVID - UNADIG | Bioinformatics Laboratory / LNCC | Luiz G P de Almeida, Alessandra P Lamarca, Ronaldo da Silva F Jr, Liliane Cavalcante, Alexandra L Gerber, Ana Paula de C Guimaraes, Douglas Terra Machado, Cassia Alves, Diana Mariani, Cintia Policarpo, Gleidson da Silva de Oliveira, Mario Sergio Ribeiro, Silvia Carvalho, Flavio Dias da Silva, Marcio Henrique de Oliveira Garcia, Leandro Magalhaes de Souza, Cristiane Gomes da Silva, Caio Luiz Pereira Ribeiro, Andrea Cony Cavalcanti, Claudia Maria Braga de Mello, Amilcar Tanuri, Ana Tereza R Vasconcelos |
| EPI_ISL_2101485, EPI_ISL_2101486, EPI_ISL_2101487, EPI_ISL_2101488, EPI_ISL_2101489, EPI_ISL_2101490, EPI_ISL_2101491, EPI_ISL_2101492, EPI_ISL_2101493 | Laboratorio Central Noel Nutels | Bioinformatics Laboratory / LNCC | Luiz G P de Almeida, Alessandra P Lamarca, Ronaldo da Silva F Jr, Liliane Cavalcante, Alexandra L Gerber, Ana Paula de C Guimaraes, Douglas Terra Machado, Cassia Alves, Diana Mariani, Cintia Policarpo, Gleidson da Silva de Oliveira, Mario Sergio Ribeiro, Silvia Carvalho, Flavio Dias da Silva, Marcio Henrique de Oliveira Garcia, Leandro Magalhaes de Souza, Cristiane Gomes da Silva, Caio Luiz Pereira Ribeiro, Andrea Cony Cavalcanti, Claudia Maria Braga de Mello, Amilcar Tanuri, Ana Tereza R Vasconcelos |
| EPI_ISL_2101494, EPI_ISL_2101495 | Unidade de apoio ao diagnóstico da COVID - UNADIG | Bioinformatics Laboratory / LNCC | Luiz G P de Almeida, Alessandra P Lamarca, Ronaldo da Silva F Jr, Liliane Cavalcante, Alexandra L Gerber, Ana Paula de C Guimaraes, Douglas Terra Machado, Cassia Alves, Diana Mariani, Cintia Policarpo, Gleidson da Silva de Oliveira, Mario Sergio Ribeiro, Silvia Carvalho, Flavio Dias da Silva, Marcio Henrique de Oliveira Garcia, Leandro Magalhaes de Souza, Cristiane Gomes da Silva, Caio Luiz Pereira Ribeiro, Andrea Cony Cavalcanti, Claudia Maria Braga de Mello, Amilcar Tanuri, Ana Tereza R Vasconcelos |
| EPI_ISL_2101496, EPI_ISL_2101497, EPI_ISL_2101498, EPI_ISL_2101499, EPI_ISL_2101500, EPI_ISL_2101501, EPI_ISL_2101502, EPI_ISL_2101503, EPI_ISL_2101504 | Laboratorio Central Noel Nutels | Bioinformatics Laboratory / LNCC | Luiz G P de Almeida, Alessandra P Lamarca, Ronaldo da Silva F Jr, Liliane Cavalcante, Alexandra L Gerber, Ana Paula de C Guimaraes, Douglas Terra Machado, Cassia Alves, Diana Mariani, Cintia Policarpo, Gleidson da Silva de Oliveira, Mario Sergio Ribeiro, Silvia Carvalho, Flavio Dias da Silva, Marcio Henrique de Oliveira Garcia, Leandro Magalhaes de Souza, Cristiane Gomes da Silva, Caio Luiz Pereira Ribeiro, Andrea Cony Cavalcanti, Claudia Maria Braga de Mello, Amilcar Tanuri, Ana Tereza R Vasconcelos |
| EPI_ISL_2101505, EPI_ISL_2101506 | Unidade de apoio ao diagnóstico da COVID - UNADIG | Bioinformatics Laboratory / LNCC | Luiz G P de Almeida, Alessandra P Lamarca, Ronaldo da Silva F Jr, Liliane Cavalcante, Alexandra L Gerber, Ana Paula de C Guimaraes, Douglas Terra Machado, Cassia Alves, Diana Mariani, Cintia Policarpo, Gleidson da Silva de Oliveira, Mario Sergio Ribeiro, Silvia Carvalho, Flavio Dias da Silva, Marcio Henrique de Oliveira Garcia, Leandro Magalhaes de Souza, Cristiane Gomes da Silva, Caio Luiz Pereira Ribeiro, Andrea Cony Cavalcanti, Claudia Maria Braga de Mello, Amilcar Tanuri, Ana Tereza R Vasconcelos |
| EPI_ISL_2101507 | Laboratorio Central Noel Nutels | Bioinformatics Laboratory / LNCC | Luiz G P de Almeida, Alessandra P Lamarca, Ronaldo da Silva F Jr, Liliane Cavalcante, Alexandra L Gerber, Ana Paula de C Guimaraes, Douglas Terra Machado, Cassia Alves, Diana Mariani, Cintia Policarpo, Gleidson da Silva de Oliveira, Mario Sergio Ribeiro, Silvia Carvalho, Flavio Dias da Silva, Marcio Henrique de Oliveira Garcia, Leandro Magalhaes de Souza, Cristiane Gomes da Silva, Caio Luiz Pereira Ribeiro, Andrea Cony Cavalcanti, Claudia Maria Braga de Mello, Amilcar Tanuri, Ana Tereza R Vasconcelos |
| EPI_ISL_2101508, EPI_ISL_2101509, EPI_ISL_2101510, EPI_ISL_2101511, EPI_ISL_2101512, EPI_ISL_2101513 | Unidade de apoio ao diagnóstico da COVID - UNADIG | Bioinformatics Laboratory / LNCC | Luiz G P de Almeida, Alessandra P Lamarca, Ronaldo da Silva F Jr, Liliane Cavalcante, Alexandra L Gerber, Ana Paula de C Guimaraes, Douglas Terra Machado, Cassia Alves, Diana Mariani, Cintia Policarpo, Gleidson da Silva de Oliveira, Mario Sergio Ribeiro, Silvia Carvalho, Flavio Dias da Silva, Marcio Henrique de Oliveira Garcia, Leandro Magalhaes de Souza, Cristiane Gomes da Silva, Caio Luiz Pereira Ribeiro, Andrea Cony Cavalcanti, Claudia Maria Braga de Mello, Amilcar Tanuri, Ana Tereza R Vasconcelos |
| EPI_ISL_2101514, EPI_ISL_2101515, EPI_ISL_2101516, EPI_ISL_2101517, EPI_ISL_2101518, EPI_ISL_2101519 | Laboratorio Central Noel Nutels | Bioinformatics Laboratory / LNCC | Luiz G P de Almeida, Alessandra P Lamarca, Ronaldo da Silva F Jr, Liliane Cavalcante, Alexandra L Gerber, Ana Paula de C Guimaraes, Douglas Terra Machado, Cassia Alves, Diana Mariani, Cintia Policarpo, Gleidson da Silva de Oliveira, Mario Sergio Ribeiro, Silvia Carvalho, Flavio Dias da Silva, Marcio Henrique de Oliveira Garcia, Leandro Magalhaes de Souza, Cristiane Gomes da Silva, Caio Luiz Pereira Ribeiro, Andrea Cony Cavalcanti, Claudia Maria Braga de Mello, Amilcar Tanuri, Ana Tereza R Vasconcelos |
| EPI_ISL_2101520, EPI_ISL_2101521, EPI_ISL_2101522, EPI_ISL_2101523, EPI_ISL_2101524, EPI_ISL_2101525, EPI_ISL_2101526, EPI_ISL_2101527 | Unidade de apoio ao diagnóstico da COVID - UNADIG | Bioinformatics Laboratory / LNCC | Luiz G P de Almeida, Alessandra P Lamarca, Ronaldo da Silva F Jr, Liliane Cavalcante, Alexandra L Gerber, Ana Paula de C Guimaraes, Douglas Terra Machado, Cassia Alves, Diana Mariani, Cintia Policarpo, Gleidson da Silva de Oliveira, Mario Sergio Ribeiro, Silvia Carvalho, Flavio Dias da Silva, Marcio Henrique de Oliveira Garcia, Leandro Magalhaes de Souza, Cristiane Gomes da Silva, Caio Luiz Pereira Ribeiro, Andrea Cony Cavalcanti, Claudia Maria Braga de Mello, Amilcar Tanuri, Ana Tereza R Vasconcelos |
| EPI_ISL_2101528, EPI_ISL_2101529, EPI_ISL_2101530, EPI_ISL_2101531, EPI_ISL_2101532, EPI_ISL_2101533, EPI_ISL_2101534, EPI_ISL_2101535, EPI_ISL_2101536, EPI_ISL_2101537, EPI_ISL_2101538, EPI_ISL_2101539, EPI_ISL_2101540, EPI_ISL_2101541, EPI_ISL_2101542, EPI_ISL_2101543, EPI_ISL_2101544, EPI_ISL_2101545, EPI_ISL_2101546, EPI_ISL_2101547, EPI_ISL_2101548, EPI_ISL_2101549, EPI_ISL_2101550, EPI_ISL_2101551, EPI_ISL_2101552, EPI_ISL_2101553, EPI_ISL_2101554, EPI_ISL_2101555, EPI_ISL_2101556, EPI_ISL_2101557, EPI_ISL_2101558, EPI_ISL_2101559, EPI_ISL_2101560, EPI_ISL_2101561, EPI_ISL_2101562, EPI_ISL_2101563, EPI_ISL_2101564, EPI_ISL_2101565 | | | |
| see above | Laboratorio Central Noel Nutels | Bioinformatics Laboratory / LNCC | Luiz G P de Almeida, Alessandra P Lamarca, Ronaldo da Silva F Jr, Liliane Cavalcante, Alexandra L Gerber, Ana Paula de C Guimaraes, Douglas Terra Machado, Cassia Alves, Diana Mariani, Cintia Policarpo, Gleidson da Silva de Oliveira, Mario Sergio Ribeiro, Silvia Carvalho, Flavio Dias da Silva, Marcio Henrique de Oliveira Garcia, Leandro Magalhaes de Souza, Cristiane Gomes da Silva, Caio Luiz Pereira Ribeiro, Andrea Cony Cavalcanti, Claudia Maria Braga de Mello, Amilcar Tanuri, Ana Tereza R Vasconcelos |
| EPI_ISL_2101566, EPI_ISL_2101567, EPI_ISL_2101568, EPI_ISL_2101569, EPI_ISL_2101570, EPI_ISL_2101571, EPI_ISL_2101572 | Unidade de apoio ao diagnóstico da COVID - UNADIG | Bioinformatics Laboratory / LNCC | Luiz G P de Almeida, Alessandra P Lamarca, Ronaldo da Silva F Jr, Liliane Cavalcante, Alexandra L Gerber, Ana Paula de C Guimaraes, Douglas Terra Machado, Cassia Alves, Diana Mariani, Cintia Policarpo, Gleidson da Silva de Oliveira, Mario Sergio Ribeiro, Silvia Carvalho, Flavio Dias da Silva, Marcio Henrique de Oliveira Garcia, Leandro Magalhaes de Souza, Cristiane Gomes da Silva, Caio Luiz Pereira Ribeiro, Andrea Cony Cavalcanti, Claudia Maria Braga de Mello, Amilcar Tanuri, Ana Tereza R Vasconcelos |
| EPI_ISL_2101573, EPI_ISL_2101574, EPI_ISL_2101575, EPI_ISL_2101576, EPI_ISL_2101577, EPI_ISL_2101578, EPI_ISL_2101579, EPI_ISL_2101580, EPI_ISL_2101581, EPI_ISL_2101582, EPI_ISL_2101583, EPI_ISL_2101584, EPI_ISL_2101585, EPI_ISL_2101586, EPI_ISL_2101587, EPI_ISL_2101588, EPI_ISL_2101589, EPI_ISL_2101590, EPI_ISL_2101591, EPI_ISL_2101592, EPI_ISL_2101593, EPI_ISL_2101594, EPI_ISL_2101595, EPI_ISL_2101596, EPI_ISL_2101597, EPI_ISL_2101598, EPI_ISL_2101599, EPI_ISL_2101600, EPI_ISL_2101601, EPI_ISL_2101602, EPI_ISL_2101603, EPI_ISL_2101604, EPI_ISL_2101605, EPI_ISL_2101606, EPI_ISL_2101607, EPI_ISL_2101608, EPI_ISL_2101609, EPI_ISL_2101610, EPI_ISL_2101611 | | | |
| see above | Laboratorio Central Noel Nutels | Bioinformatics Laboratory / LNCC | Luiz G P de Almeida, Alessandra P Lamarca, Ronaldo da Silva F Jr, Liliane Cavalcante, Alexandra L Gerber, Ana Paula de C Guimaraes, Douglas Terra Machado, Cassia Alves, Diana Mariani, Cintia Policarpo, Gleidson da Silva de Oliveira, Mario Sergio Ribeiro, Silvia Carvalho, Flavio Dias da Silva, Marcio Henrique de Oliveira Garcia, Leandro Magalhaes de Souza, Cristiane Gomes da Silva, Caio Luiz Pereira Ribeiro, Andrea Cony Cavalcanti, Claudia Maria Braga de Mello, Amilcar Tanuri, Ana Tereza R Vasconcelos |
| EPI_ISL_2101612 | Unidade de apoio ao diagnóstico da COVID - UNADIG | Bioinformatics Laboratory / LNCC | Luiz G P de Almeida, Alessandra P Lamarca, Ronaldo da Silva F Jr, Liliane Cavalcante, Alexandra L Gerber, Ana Paula de C Guimaraes, Douglas Terra Machado, Cassia Alves, Diana Mariani, Cintia Policarpo, Gleidson da Silva de Oliveira, Mario Sergio Ribeiro, Silvia Carvalho, Flavio Dias da Silva, Marcio |

| | | | |
|---|---|---|---|
| | | | Henrique de Oliveira Garcia, Leandro Magalhaes de Souza, Cristiane Gomes da Silva, Caio Luiz Pereira Ribeiro, Andrea Cony Cavalcanti, Claudia Maria Braga de Mello, Amilcar Tanuri, Ana Tereza R Vasconcelos |
| EPI_ISL_2101613, EPI_ISL_2101615, EPI_ISL_2101616, EPI_ISL_2101617, EPI_ISL_2101618, EPI_ISL_2101619, EPI_ISL_2101620, EPI_ISL_2101621, EPI_ISL_2101622, EPI_ISL_2101623, EPI_ISL_2101624, EPI_ISL_2101625, EPI_ISL_2101626, EPI_ISL_2101627, EPI_ISL_2101628, EPI_ISL_2101629, EPI_ISL_2101630, EPI_ISL_2101631, EPI_ISL_2101632, EPI_ISL_2101633, EPI_ISL_2101634, EPI_ISL_2101635, EPI_ISL_2101636, EPI_ISL_2101637 | | | |
| see above | Laboratorio Central Noel Nutels | Bioinformatics Laboratory / LNCC | Luiz G P de Almeida, Alessandra P Lamarca, Ronaldo da Silva F Jr, Liliane Cavalcante, Alexandra L Gerber, Ana Paula de C Guimaraes, Douglas Terra Machado, Cassia Alves, Diana Mariani, Cintia Policarpo, Gleidson da Silva de Oliveira, Mario Sergio Ribeiro, Silvia Carvalho, Flavio Dias da Silva, Marcio Henrique de Oliveira Garcia, Leandro Magalhaes de Souza, Cristiane Gomes da Silva, Caio Luiz Pereira Ribeiro, Andrea Cony Cavalcanti, Claudia Maria Braga de Mello, Amilcar Tanuri, Ana Tereza R Vasconcelos |
| EPI_ISL_2101638, EPI_ISL_2101639, EPI_ISL_2101640, EPI_ISL_2101641, EPI_ISL_2101642, EPI_ISL_2101643, EPI_ISL_2101644, EPI_ISL_2101645, EPI_ISL_2101646, EPI_ISL_2101647, EPI_ISL_2101648, EPI_ISL_2101649, EPI_ISL_2101650, EPI_ISL_2101651, EPI_ISL_2101652, EPI_ISL_2101653, EPI_ISL_2101654, EPI_ISL_2101655, EPI_ISL_2101656, EPI_ISL_2101657, EPI_ISL_2101658, EPI_ISL_2101659, EPI_ISL_2101660, EPI_ISL_2101661, EPI_ISL_2101662, EPI_ISL_2101663, EPI_ISL_2101664, EPI_ISL_2101665, EPI_ISL_2101666, EPI_ISL_2101667, EPI_ISL_2101668, EPI_ISL_2101669, EPI_ISL_2101670, EPI_ISL_2101671 | | | |
| see above | Unidade de apoio ao diagnóstico da COVID - UNADIG | Bioinformatics Laboratory / LNCC | Luiz G P de Almeida, Alessandra P Lamarca, Ronaldo da Silva F Jr, Liliane Cavalcante, Alexandra L Gerber, Ana Paula de C Guimaraes, Douglas Terra Machado, Cassia Alves, Diana Mariani, Cintia Policarpo, Gleidson da Silva de Oliveira, Mario Sergio Ribeiro, Silvia Carvalho, Flavio Dias da Silva, Marcio Henrique de Oliveira Garcia, Leandro Magalhaes de Souza, Cristiane Gomes da Silva, Caio Luiz Pereira Ribeiro, Andrea Cony Cavalcanti, Claudia Maria Braga de Mello, Amilcar Tanuri, Ana Tereza R Vasconcelos |
| EPI_ISL_2101672, EPI_ISL_2101673, EPI_ISL_2101674, EPI_ISL_2101675 | Laboratorio Central Noel Nutels | Bioinformatics Laboratory / LNCC | Luiz G P de Almeida, Alessandra P Lamarca, Ronaldo da Silva F Jr, Liliane Cavalcante, Alexandra L Gerber, Ana Paula de C Guimaraes, Douglas Terra Machado, Cassia Alves, Diana Mariani, Cintia Policarpo, Gleidson da Silva de Oliveira, Mario Sergio Ribeiro, Silvia Carvalho, Flavio Dias da Silva, Marcio Henrique de Oliveira Garcia, Leandro Magalhaes de Souza, Cristiane Gomes da Silva, Caio Luiz Pereira Ribeiro, Andrea Cony Cavalcanti, Claudia Maria Braga de Mello, Amilcar Tanuri, Ana Tereza R Vasconcelos |
| EPI_ISL_2101676, EPI_ISL_2101677, EPI_ISL_2101678, EPI_ISL_2101679, EPI_ISL_2101680, EPI_ISL_2101681, EPI_ISL_2101682, EPI_ISL_2101683, EPI_ISL_2101684, EPI_ISL_2101685, EPI_ISL_2101686, EPI_ISL_2101687, EPI_ISL_2101688 | | | |
| see above | Unidade de apoio ao diagnóstico da COVID - UNADIG | Bioinformatics Laboratory / LNCC | Luiz G P de Almeida, Alessandra P Lamarca, Ronaldo da Silva F Jr, Liliane Cavalcante, Alexandra L Gerber, Ana Paula de C Guimaraes, Douglas Terra Machado, Cassia Alves, Diana Mariani, Cintia Policarpo, Gleidson da Silva de Oliveira, Mario Sergio Ribeiro, Silvia Carvalho, Flavio Dias da Silva, Marcio Henrique de Oliveira Garcia, Leandro Magalhaes de Souza, Cristiane Gomes da Silva, Caio Luiz Pereira Ribeiro, Andrea Cony Cavalcanti, Claudia Maria Braga de Mello, Amilcar Tanuri, Ana Tereza R Vasconcelos |
| EPI_ISL_2101689, EPI_ISL_2101690, EPI_ISL_2101691 | Laboratorio Central Noel Nutels | Bioinformatics Laboratory / LNCC | Luiz G P de Almeida, Alessandra P Lamarca, Ronaldo da Silva F Jr, Liliane Cavalcante, Alexandra L Gerber, Ana Paula de C Guimaraes, Douglas Terra Machado, Cassia Alves, Diana Mariani, Cintia Policarpo, Gleidson da Silva de Oliveira, Mario Sergio Ribeiro, Silvia Carvalho, Flavio Dias da Silva, Marcio Henrique de Oliveira Garcia, Leandro Magalhaes de Souza, Cristiane Gomes da Silva, Caio Luiz Pereira Ribeiro, Andrea Cony Cavalcanti, Claudia Maria Braga de Mello, Amilcar Tanuri, Ana Tereza R Vasconcelos |
| EPI_ISL_2101692, EPI_ISL_2101693, EPI_ISL_2101694, EPI_ISL_2101695, EPI_ISL_2101696 | Unidade de apoio ao diagnóstico da COVID - UNADIG | Bioinformatics Laboratory / LNCC | Luiz G P de Almeida, Alessandra P Lamarca, Ronaldo da Silva F Jr, Liliane Cavalcante, Alexandra L Gerber, Ana Paula de C Guimaraes, Douglas Terra Machado, Cassia Alves, Diana Mariani, Cintia Policarpo, Gleidson da Silva de Oliveira, Mario Sergio Ribeiro, Silvia Carvalho, Flavio Dias da Silva, Marcio Henrique de Oliveira Garcia, Leandro Magalhaes de Souza, Cristiane Gomes da Silva, Caio Luiz Pereira Ribeiro, Andrea Cony Cavalcanti, Claudia Maria Braga de Mello, Amilcar Tanuri, Ana Tereza R Vasconcelos |
| EPI_ISL_2101697, EPI_ISL_2101698, EPI_ISL_2101699, EPI_ISL_2101700, EPI_ISL_2101701, EPI_ISL_2101702, EPI_ISL_2101703, EPI_ISL_2101704, EPI_ISL_2101705, EPI_ISL_2101706, EPI_ISL_2101707, EPI_ISL_2101708, EPI_ISL_2101709, EPI_ISL_2101710, EPI_ISL_2101711, EPI_ISL_2101712, EPI_ISL_2101713 | | | |
| see above | Laboratorio Central Noel Nutels | Bioinformatics Laboratory / LNCC | Luiz G P de Almeida, Alessandra P Lamarca, Ronaldo da Silva F Jr, Liliane Cavalcante, Alexandra L Gerber, Ana Paula de C Guimaraes, Douglas Terra Machado, Cassia Alves, Diana Mariani, Cintia Policarpo, Gleidson da Silva de Oliveira, Mario Sergio Ribeiro, Silvia Carvalho, Flavio Dias da Silva, Marcio Henrique de Oliveira Garcia, Leandro Magalhaes de Souza, Cristiane Gomes da Silva, Caio Luiz Pereira Ribeiro, Andrea Cony Cavalcanti, Claudia Maria Braga de Mello, Amilcar Tanuri, Ana Tereza R Vasconcelos |
| EPI_ISL_2101714, EPI_ISL_2101715 | Unidade de apoio ao diagnóstico da COVID - UNADIG | Bioinformatics Laboratory / LNCC | Luiz G P de Almeida, Alessandra P Lamarca, Ronaldo da Silva F Jr, Liliane Cavalcante, Alexandra L Gerber, Ana Paula de C Guimaraes, Douglas Terra Machado, Cassia Alves, Diana Mariani, Cintia Policarpo, Gleidson da Silva de Oliveira, Mario Sergio Ribeiro, Silvia Carvalho, Flavio Dias da Silva, Marcio Henrique de Oliveira Garcia, Leandro Magalhaes de Souza, Cristiane Gomes da Silva, Caio Luiz Pereira Ribeiro, Andrea Cony Cavalcanti, Claudia Maria Braga de Mello, Amilcar Tanuri, Ana Tereza R Vasconcelos |
| EPI_ISL_2101716, EPI_ISL_2101717, EPI_ISL_2101718, EPI_ISL_2101719, EPI_ISL_2101720, EPI_ISL_2101721 | Laboratorio Central Noel Nutels | Bioinformatics Laboratory / LNCC | Luiz G P de Almeida, Alessandra P Lamarca, Ronaldo da Silva F Jr, Liliane Cavalcante, Alexandra L Gerber, Ana Paula de C Guimaraes, Douglas Terra Machado, Cassia Alves, Diana Mariani, Cintia Policarpo, Gleidson da Silva de Oliveira, Mario Sergio Ribeiro, Silvia Carvalho, Flavio Dias da Silva, Marcio Henrique de Oliveira Garcia, Leandro Magalhaes de Souza, Cristiane Gomes da Silva, Caio Luiz Pereira Ribeiro, Andrea Cony Cavalcanti, Claudia Maria Braga de Mello, Amilcar Tanuri, Ana Tereza R Vasconcelos |
| EPI_ISL_2101722 | Unidade de apoio ao diagnóstico da COVID - UNADIG | Bioinformatics Laboratory / LNCC | Luiz G P de Almeida, Alessandra P Lamarca, Ronaldo da Silva F Jr, Liliane Cavalcante, Alexandra L Gerber, Ana Paula de C Guimaraes, Douglas Terra Machado, Cassia Alves, Diana Mariani, Cintia Policarpo, Gleidson da Silva de Oliveira, Mario Sergio Ribeiro, Silvia Carvalho, Flavio Dias da Silva, Marcio Henrique de Oliveira Garcia, Leandro Magalhaes de Souza, Cristiane Gomes da Silva, Caio Luiz Pereira Ribeiro, Andrea Cony Cavalcanti, Claudia Maria Braga de Mello, Amilcar Tanuri, Ana Tereza R Vasconcelos |
| EPI_ISL_2101730 | Laboratorio Central Noel Nutels | Bioinformatics Laboratory / LNCC | Luiz G P de Almeida, Alessandra P Lamarca, Ronaldo da Silva F Jr, Liliane Cavalcante, Alexandra L Gerber, Ana Paula de C Guimaraes, Douglas Terra Machado, Cassia Alves, Diana Mariani, Cintia Policarpo, Gleidson da Silva de Oliveira, Mario Sergio Ribeiro, Silvia Carvalho, Flavio Dias da Silva, Marcio Henrique de Oliveira Garcia, Leandro Magalhaes de Souza, Cristiane Gomes da Silva, Caio Luiz Pereira Ribeiro, Andrea Cony Cavalcanti, Claudia Maria Braga de Mello, Amilcar Tanuri, Ana Tereza R Vasconcelos |
| EPI_ISL_2101731 | Unidade de apoio ao diagnóstico da COVID - UNADIG | Bioinformatics Laboratory / LNCC | Luiz G P de Almeida, Alessandra P Lamarca, Ronaldo da Silva F Jr, Liliane Cavalcante, Alexandra L Gerber, Ana Paula de C Guimaraes, Douglas Terra Machado, Cassia Alves, Diana Mariani, Cintia Policarpo, Gleidson da Silva de Oliveira, Mario Sergio Ribeiro, Silvia Carvalho, Flavio Dias da Silva, Marcio Henrique de Oliveira Garcia, Leandro Magalhaes de Souza, Cristiane Gomes da Silva, Caio Luiz Pereira Ribeiro, Andrea Cony Cavalcanti, Claudia Maria Braga de Mello, Amilcar Tanuri, Ana Tereza R Vasconcelos |
| EPI_ISL_2101732, EPI_ISL_2101733 | Laboratorio Central Noel Nutels | Bioinformatics Laboratory / LNCC | Luiz G P de Almeida, Alessandra P Lamarca, Ronaldo da Silva F Jr, Liliane Cavalcante, Alexandra L Gerber, Ana Paula de C Guimaraes, Douglas Terra Machado, Cassia Alves, Diana Mariani, Cintia Policarpo, Gleidson da Silva de Oliveira, Mario Sergio Ribeiro, Silvia Carvalho, Flavio Dias da Silva, Marcio Henrique de Oliveira Garcia, Leandro Magalhaes de Souza, Cristiane Gomes da Silva, Caio Luiz Pereira Ribeiro, Andrea Cony Cavalcanti, Claudia Maria Braga de Mello, Amilcar Tanuri, Ana Tereza R Vasconcelos |
| EPI_ISL_2101734 | Unidade de apoio ao diagnóstico da COVID - UNADIG | Bioinformatics Laboratory / LNCC | Luiz G P de Almeida, Alessandra P Lamarca, Ronaldo da Silva F Jr, Liliane Cavalcante, Alexandra L Gerber, Ana Paula de C Guimaraes, Douglas Terra Machado, Cassia Alves, Diana Mariani, Cintia Policarpo, Gleidson da Silva de Oliveira, Mario Sergio Ribeiro, Silvia Carvalho, Flavio Dias da Silva, Marcio Henrique de Oliveira Garcia, Leandro Magalhaes de Souza, Cristiane Gomes da Silva, Caio Luiz Pereira Ribeiro, Andrea Cony Cavalcanti, Claudia Maria Braga de Mello, Amilcar Tanuri, Ana Tereza R Vasconcelos |
| EPI_ISL_2101736, EPI_ISL_2101737, EPI_ISL_2101738, EPI_ISL_2101739 | Laboratorio Central Noel Nutels | Bioinformatics Laboratory / LNCC | Luiz G P de Almeida, Alessandra P Lamarca, Ronaldo da Silva F Jr, Liliane Cavalcante, Alexandra L Gerber, Ana Paula de C Guimaraes, Douglas Terra Machado, Cassia Alves, Diana Mariani, Cintia Policarpo, Gleidson da Silva de Oliveira, Mario Sergio Ribeiro, Silvia Carvalho, Flavio Dias da Silva, Marcio Henrique de Oliveira Garcia, Leandro Magalhaes de Souza, Cristiane Gomes da Silva, Caio Luiz Pereira Ribeiro, Andrea Cony Cavalcanti, Claudia Maria Braga de Mello, Amilcar Tanuri, Ana Tereza R Vasconcelos |
| EPI_ISL_2101740, EPI_ISL_2101741 | Unidade de apoio ao diagnóstico da COVID - UNADIG | Bioinformatics Laboratory / LNCC | Luiz G P de Almeida, Alessandra P Lamarca, Ronaldo da Silva F Jr, Liliane Cavalcante, Alexandra L Gerber, Ana Paula de C Guimaraes, Douglas Terra Machado, Cassia Alves, Diana Mariani, Cintia Policarpo, Gleidson da Silva de Oliveira, Mario Sergio Ribeiro, Silvia Carvalho, Flavio Dias da Silva, Marcio Henrique de Oliveira Garcia, Leandro Magalhaes de Souza, Cristiane Gomes da Silva, Caio Luiz Pereira Ribeiro, Andrea Cony Cavalcanti, Claudia Maria Braga de Mello, Amilcar Tanuri, Ana Tereza R Vasconcelos |
| EPI_ISL_2101742, EPI_ISL_2101743 | Laboratorio Central Noel Nutels | Bioinformatics Laboratory / LNCC | Luiz G P de Almeida, Alessandra P Lamarca, Ronaldo da Silva F Jr, Liliane Cavalcante, Alexandra L Gerber, Ana Paula de C Guimaraes, Douglas Terra Machado, Cassia Alves, Diana Mariani, Cintia Policarpo, Gleidson da Silva de Oliveira, Mario Sergio Ribeiro, Silvia Carvalho, Flavio Dias da Silva, Marcio Henrique de Oliveira Garcia, Leandro Magalhaes de Souza, Cristiane Gomes da Silva, Caio Luiz Pereira Ribeiro, Andrea Cony Cavalcanti, Claudia Maria Braga de Mello, Amilcar Tanuri, Ana Tereza R Vasconcelos |

| | | | |
|---|---|---|---|
| EPI_ISL_2101744 | Unidade de apoio ao diagnóstico da COVID - UNADIG | Bioinformatics Laboratory / LNCC | Luiz G P de Almeida, Alessandra P Lamarca, Ronaldo da Silva F Jr, Liliane Cavalcante, Alexandra L Gerber, Ana Paula de C Guimaraes, Douglas Terra Machado, Cassia Alves, Diana Mariani, Cintia Policarpo, Gleidson da Silva de Oliveira, Mario Sergio Ribeiro, Silvia Carvalho, Flavio Dias da Silva, Marcio Henrique de Oliveira Garcia, Leandro Magalhaes de Souza, Cristiane Gomes da Silva, Caio Luiz Pereira Ribeiro, Andrea Cony Cavalcanti, Claudia Maria Braga de Mello, Amilcar Tanuri, Ana Tereza R Vasconcelos |
| EPI_ISL_2101745, EPI_ISL_2101746, EPI_ISL_2101747 | Laboratorio Central Noel Nutels | Bioinformatics Laboratory / LNCC | Luiz G P de Almeida, Alessandra P Lamarca, Ronaldo da Silva F Jr, Liliane Cavalcante, Alexandra L Gerber, Ana Paula de C Guimaraes, Douglas Terra Machado, Cassia Alves, Diana Mariani, Cintia Policarpo, Gleidson da Silva de Oliveira, Mario Sergio Ribeiro, Silvia Carvalho, Flavio Dias da Silva, Marcio Henrique de Oliveira Garcia, Leandro Magalhaes de Souza, Cristiane Gomes da Silva, Caio Luiz Pereira Ribeiro, Andrea Cony Cavalcanti, Claudia Maria Braga de Mello, Amilcar Tanuri, Ana Tereza R Vasconcelos |
| EPI_ISL_2105574, EPI_ISL_2135717, EPI_ISL_2135718, EPI_ISL_2135719, EPI_ISL_2135720, EPI_ISL_2136060, EPI_ISL_2136097, EPI_ISL_2136133 | Servicio Virosis Respiratorias-Departamento Virología-INEI | Instituto Nacional Enfermedades Infecciosas C.G.Malbran | Baumeister E., Avaro M., Benedetti E., Russo M., Dattero ME, Pontoriero A., Cisterna D., Molina V., Perandones C., Tuduri E., Lorenzo F., Poklepovich T., Campos J. |
| EPI_ISL_2139495, EPI_ISL_2139508, EPI_ISL_2139513, EPI_ISL_2139515, EPI_ISL_2139519, EPI_ISL_2139523, EPI_ISL_2139526, EPI_ISL_2139527, EPI_ISL_2139531, EPI_ISL_2139536 | Laboratorio Exame | Universidade Federal de Ciencias da Saude de Porto Alegre | Vinícius Bonetti Franceschi, Gabriel Dickin Caldana et al. |
| EPI_ISL_2140039, EPI_ISL_2140112 | Servicio Virosis Respiratorias-Departamento Virología-INEI | Instituto Nacional Enfermedades Infecciosas C.G.Malbran | Baumeister E., Avaro M., Benedetti E., Russo M., Dattero ME, Pontoriero A., Cisterna D., Molina V., Perandones C., Tuduri E., Lorenzo F., Poklepovich T., Campos J. |
| EPI_ISL_2157382 | Laboratorio Central de Saude Publica do Estado de Sergipe (LACEN/SE) | Laboratory of Respiratory Viruses and Measles, Oswaldo Cruz Institute, FIOCRUZ | Paola Resende, Luciana Appolinario, Fernando Motta, Anna Carolina Paixao, Ana Carolina Mendonca, Alice Sampaio Rocha, Tainá Moreira Martins Venas, Elisa Cavalcante Pereira, Renata Serrano Lopes, Cliomar Alves dos Santos, Marilda Siqueira on behalf of the Fiocruz COVID-19 Genomic Surveillance Network |
| EPI_ISL_2157396, EPI_ISL_2157399 | Laboratório Central de Saude Publica do Estado de Santa Catarina (LACEN/SC) | Laboratory of Respiratory Viruses and Measles, Oswaldo Cruz Institute, FIOCRUZ | Paola Resende, Luciana Appolinario, Fernando Motta, Anna Carolina Paixao, Ana Carolina Mendonca, Alice Sampaio Rocha, Taina Venas, Elisa Cavalcante Pereira, Renata Serrano Lopes, Darcita Buerger Rovaris, Sandra Bianchini Fernandes, Marilda Siqueira on behalf of the Fiocruz COVID-19 Genomic Surveillance Network |
| EPI_ISL_2157404 | Lboratorio Central de Saude Publica do Estado do Parana (LACEN/PR) | Laboratory of Respiratory Viruses and Measles, Oswaldo Cruz Institute, FIOCRUZ | Paola Resende, Luciana Appolinario, Fernando Motta, Anna Carolina Paixao, Ana Carolina Mendonca, Alice Sampaio Rocha, Taina Venas, Elisa Cavalcante Pereira, Renata Serrano Lopes, Irina Riediger, Marilda Siqueira on behalf of the Fiocruz COVID-19 Genomic Surveillance Network |
| EPI_ISL_2157407, EPI_ISL_2157426 | Laboratório Central de Saude Publica do Estado de Santa Catarina (LACEN/SC) | Laboratory of Respiratory Viruses and Measles, Oswaldo Cruz Institute, FIOCRUZ | Paola Resende, Luciana Appolinario, Fernando Motta, Anna Carolina Paixao, Ana Carolina Mendonca, Alice Sampaio Rocha, Taina Venas, Elisa Cavalcante Pereira, Renata Serrano Lopes, Darcita Buerger Rovaris, Sandra Bianchini Fernandes, Marilda Siqueira on behalf of the Fiocruz COVID-19 Genomic Surveillance Network |
| EPI_ISL_2157428, EPI_ISL_2157429, EPI_ISL_2157435, EPI_ISL_2157442, EPI_ISL_2157444 | Laboratorio Central de Saude Publica do Estado de Sergipe (LACEN/SE) | Laboratory of Respiratory Viruses and Measles, Oswaldo Cruz Institute, FIOCRUZ | Paola Resende, Luciana Appolinario, Fernando Motta, Anna Carolina Paixao, Ana Carolina Mendonca, Alice Sampaio Rocha, Tainá Moreira Martins Venas, Elisa Cavalcante Pereira, Renata Serrano Lopes, Cliomar Alves dos Santos, Marilda Siqueira on behalf of the Fiocruz COVID-19 Genomic Surveillance Network |
| EPI_ISL_2157447 | Laboratório Central de Saude Publica do Estado de Santa Catarina (LACEN/SC) | Laboratory of Respiratory Viruses and Measles, Oswaldo Cruz Institute, FIOCRUZ | Paola Resende, Luciana Appolinario, Fernando Motta, Anna Carolina Paixao, Ana Carolina Mendonca, Alice Sampaio Rocha, Taina Venas, Elisa Cavalcante Pereira, Renata Serrano Lopes, Darcita Buerger Rovaris, Sandra Bianchini Fernandes, Marilda Siqueira on behalf of the Fiocruz COVID-19 Genomic Surveillance Network |
| EPI_ISL_2157449, EPI_ISL_2157453, EPI_ISL_2157471, EPI_ISL_2157473, EPI_ISL_2157474, EPI_ISL_2157478 | Laboratorio Central de Saude Publica do Estado de Sergipe (LACEN/SE) | Laboratory of Respiratory Viruses and Measles, Oswaldo Cruz Institute, FIOCRUZ | Paola Resende, Luciana Appolinario, Fernando Motta, Anna Carolina Paixao, Ana Carolina Mendonca, Alice Sampaio Rocha, Tainá Moreira Martins Venas, Elisa Cavalcante Pereira, Renata Serrano Lopes, Cliomar Alves dos Santos, Marilda Siqueira on behalf of the Fiocruz COVID-19 Genomic Surveillance Network |
| EPI_ISL_2157487 | Lboratorio Central de Saude Publica do Estado do Parana (LACEN/PR) | Laboratory of Respiratory Viruses and Measles, Oswaldo Cruz Institute, FIOCRUZ | Paola Resende, Luciana Appolinario, Fernando Motta, Anna Carolina Paixao, Ana Carolina Mendonca, Alice Sampaio Rocha, Taina Venas, Elisa Cavalcante Pereira, Renata Serrano Lopes, Irina Riediger, Marilda Siqueira on behalf of the Fiocruz COVID-19 Genomic Surveillance Network |
| EPI_ISL_2157497, EPI_ISL_2157518 | Laboratório Central de Saude Publica do Estado de Santa Catarina (LACEN/SC) | Laboratory of Respiratory Viruses and Measles, Oswaldo Cruz Institute, FIOCRUZ | Paola Resende, Luciana Appolinario, Fernando Motta, Anna Carolina Paixao, Ana Carolina Mendonca, Alice Sampaio Rocha, Taina Venas, Elisa Cavalcante Pereira, Renata Serrano Lopes, Darcita Buerger Rovaris, Sandra Bianchini Fernandes, Marilda Siqueira on behalf of the Fiocruz COVID-19 Genomic Surveillance Network |
| EPI_ISL_2157524, EPI_ISL_2157525, EPI_ISL_2157526, EPI_ISL_2157528, EPI_ISL_2157529, EPI_ISL_2157530, EPI_ISL_2157536, EPI_ISL_2157537, EPI_ISL_2157538, EPI_ISL_2157539 | Laboratório Central de Saude Publica do Estado do Rio de Janeiro (LACEN/RJ) | Laboratory of Respiratory Viruses and Measles, Oswaldo Cruz Institute, FIOCRUZ | Paola Resende, Luciana Appolinario, Fernando Motta, Anna Carolina Paixao, Ana Carolina Mendonca, Alice Sampaio Rocha, Taina Venas, Elisa Cavalcante Pereira, Renata Serrano Lopes, Andrea Cony Cavalcanti, Marilda Siqueira on behalf of the Fiocruz COVID-19 Genomic Surveillance Network |
| EPI_ISL_2157540, EPI_ISL_2157545 | Laboratory of Respiratory Viruses and Measles, Oswaldo Cruz Institute, FIOCRUZ | Laboratory of Respiratory Viruses and Measles, Oswaldo Cruz Institute, FIOCRUZ | Paola Resende, Luciana Appolinario, Fernando Motta, Anna Carolina Paixao, Ana Carolina Mendonca, Alice Sampaio Rocha, Taina Venas, Elisa Cavalcante Pereira, Renata Serrano Lopes, Andrea Cony Cavalcanti, Marilda Siqueira on behalf of the Fiocruz COVID-19 Genomic Surveillance Network |
| EPI_ISL_2157549 | Laboratório Central de Saude Publica do Estado do Rio de Janeiro (LACEN/RJ) | Laboratory of Respiratory Viruses and Measles, Oswaldo Cruz Institute, FIOCRUZ | Paola Resende, Luciana Appolinario, Fernando Motta, Anna Carolina Paixao, Ana Carolina Mendonca, Alice Sampaio Rocha, Taina Venas, Elisa Cavalcante Pereira, Renata Serrano Lopes, Andrea Cony Cavalcanti, Marilda Siqueira on behalf of the Fiocruz COVID-19 Genomic Surveillance Network |
| EPI_ISL_2157561 | Laboratorio Central de Saude Publica do Estado do Rio de Janeiro (LACEN/RJ) | Laboratory of Respiratory Viruses and Measles, Oswaldo Cruz Institute, FIOCRUZ | Paola Resende, Luciana Appolinario, Fernando Motta, Anna Carolina Paixao, Ana Carolina Mendonca, Alice Sampaio Rocha, Taina Venas, Elisa Cavalcante Pereira, Renata Serrano Lopes, Andrea Cony Cavalcanti, Marilda Siqueira on behalf of the Fiocruz COVID-19 Genomic Surveillance Network |
| EPI_ISL_2157573, EPI_ISL_2157575 | Laboratory of Respiratory Viruses and Measles, Oswaldo Cruz Institute, FIOCRUZ | Laboratory of Respiratory Viruses and Measles, Oswaldo Cruz Institute, FIOCRUZ | Paola Resende, Luciana Appolinario, Fernando Motta, Anna Carolina Paixao, Ana Carolina Mendonca, Alice Sampaio Rocha, Taina Venas, Elisa Cavalcante Pereira, Renata Serrano Lopes, Andrea Cony Cavalcanti, Marilda Siqueira on behalf of the Fiocruz COVID-19 Genomic Surveillance Network |
| EPI_ISL_2157599 | Laboratório Central de Saude Publica do Estado do Rio de Janeiro (LACEN/RJ) | Laboratory of Respiratory Viruses and Measles, Oswaldo Cruz Institute, FIOCRUZ | Paola Resende, Luciana Appolinario, Fernando Motta, Anna Carolina Paixao, Ana Carolina Mendonca, Alice Sampaio Rocha, Taina Venas, Elisa Cavalcante Pereira, Renata Serrano Lopes, Andrea Cony Cavalcanti, Marilda Siqueira on behalf of the Fiocruz COVID-19 Genomic Surveillance Network |
| EPI_ISL_2158697, EPI_ISL_2158710, EPI_ISL_2158730, EPI_ISL_2158737, EPI_ISL_2158740, EPI_ISL_2158750, EPI_ISL_2158824 | Servicio Virosis Respiratorias-Departamento Virología-INEI | Instituto Nacional Enfermedades Infecciosas C.G.Malbran | Baumeister E., Avaro M., Benedetti E., Russo M., Dattero ME, Pontoriero A., Cisterna D., Molina V., Perandones C., Tuduri E., Lorenzo F., Poklepovich T., Campos J. |
| EPI_ISL_2170908 | LABORATORIO DE FRANCA | Instituto Butantan / Mendelics | Instituto Butantan: Dimas Tadeu Covas, Sandra Coccuzzo Sampaio, Maria Carolina Elias, José Salvatore Leister Patané, Vincent Louis Viala, Antonio Jorge Martins, Ricardo Haddad, Claudia Renata dos Santos Barros, Elaine Cristina Marqueze, Raul Machado Neto, Debora Botequio Moretti, Jardelina de Souza Todao Bernardino, Loyze Paola Oliveira de Lima, Luiz Aurelio de Campos Crispin. Centro de Genômica Funcional da ESALQ: Luiz Lehmann Coutinho, Ricardo Augusto Brassaloti, Raquel de Lello Rocha Campos Cassano. NGS Soluções Genômicas: Pilar Drummond Sampaio Corrêa Mariani. FZEA-USP Pirassununga: Mirele Daiana Poleti, Jessika Cristina Chagas Lesbon, Elisangela Chicaroni Mattos, Heidge Fukumasu. USP-Botucatu: Rejane Maria Tommasini Grotto, Jayme A. Souza-Neto, Guilherme Targino Valente, Patricia Akemi Assato, Felipe Allan da Silva da Costa, Bianca Cechetto Carlos. Mendelics: Bibiana Santos, João Paulo Kitajima, Erika Freitas, David Schlesinger. Hemocentro Ribeirão Preto: Simone Kashima, Evandra Strazza Rodrigues, Svetoslav Nanev Slavov, Elaine Vieira dos Santos, Rafael dos Santos Bezerra, Luiz Carlos Junior de Alcantara, Marta Giovanetti, Vagner Fonseca, Flavia Aburjaile, Rodrigo Tocantins Calado. FAMERP-SJRP: Cecília Artico Banho, Lívia Sacchetto, Fábio Sossai Possebon, Leila Sabrina Ullmann, Cíntia Bittar, Guilherme Campos, Helena Lage Ferreira, Jorge A. Petroli Marchesi, Maisa C. Pereira Parra, Marília Moraes, Paula Rahal, Paulo Inacio da Costa, João Pessoa Araújo Jr., Maurício Lacerda Nogueira. Prefeitura de Sao Paulo: Melissa Palmieri. |
| EPI_ISL_2170911 | UNIDADE BASICA DE SAUDE DA FAMILIA BILAC | Instituto Butantan / Mendelics | Instituto Butantan: Dimas Tadeu Covas, Sandra Coccuzzo Sampaio, Maria Carolina Elias, José Salvatore Leister Patané, Vincent Louis Viala, Antonio Jorge Martins, Ricardo Haddad, Claudia Renata dos Santos Barros, Elaine Cristina Marqueze, Raul Machado Neto, Debora Botequio Moretti, Jardelina de |

| | | | |
|---|---|---|---|
| | | | Souza Todao Bernardino, Loyze Paola Oliveira de Lima, Luiz Aurelio de Campos Crispin. Centro de Genômica Funcional da ESALQ: Luiz Lehmann Coutinho, Ricardo Augusto Brassaloti, Raquel de Lello Rocha Campos Cassano. NGS Soluções Genômicas: Pilar Drummond Sampaio Corrêa Mariani. FZEA-USP Pirassununga: Mirele Daiana Poleti, Jessika Cristina Chagas Lesbon, Elisangela Chicaroni Mattos, Heidge Fukumasu. USP-Botucatu: Rejane Maria Tommasini Grotto, Jayme A. Souza-Neto, Guilherme Targino Valente, Patricia Akemi Assato, Felipe Allan da Silva da Costa, Bianca Cechetto Carlos. Mendelics: Bibiana Santos, João Paulo Kitajima, Erika Freitas, David Schlesinger. Hemocentro Ribeirão Preto: Simone Kashima, Evandra Strazza Rodrigues, Svetoslav Nanev Slavov, Elaine Vieira dos Santos, Rafael dos Santos Bezerra, Luiz Carlos Junior de Alcantara, Marta Giovanetti, Vagner Fonseca, Flavia Aburjaile, Rodrigo Tocantins Calado. FAMERP-SJRP: Cecília Artico Banho, Lívia Sacchetto, Fábio Sossai Possebon, Leila Sabrina Ullmann, Cíntia Bittar, Guilherme Campos, Helena Lage Ferreira, Jorge A. Petroli Marchesi, Maisa C. Pereira Parra, Marília Moraes, Paula Rahal, Paulo Inacio da Costa, João Pessoa Araújo Jr., Maurício Lacerda Nogueira. Prefeitura de Sao Paulo: Melissa Palmieri. |
| EPI_ISL_2187879, EPI_ISL_2187911, EPI_ISL_2187952, EPI_ISL_2187964, EPI_ISL_2188002, EPI_ISL_2188008 | HLAGYN - Laboratorio de Imunologia de Transplantes de Goias | HLAGYN - Laboratorio de Imunologia de Transplantes de Goias | Fernando Antonio Vinhal dos Santos, Erika Lopes Rocha Batista, Alessandro Leonardo Alvares Magalhaes, Frederico Rodrigues Vinhal, Sabrina Sara Moreira Duarte, Lucas Carlos Gomes Pereira, Daniel Ferreira de Sousa |
| EPI_ISL_2196185, EPI_ISL_2196186, EPI_ISL_2196187 | Laboratory of Respiratory Viruses and Measles, Oswaldo Cruz Institute, FIOCRUZ | Laboratory of Respiratory Viruses and Measles, Oswaldo Cruz Institute, FIOCRUZ | Paola Resende, Luciana Appolinario, Fernando Motta, Anna Carolina Paixao, Ana Carolina Mendonca, Alice Sampaio Rocha, Taina Venas, Elisa Cavalcante Pereira, Renata Serrano Lopes, Marilda Siqueira on behalf of the Fiocruz COVID-19 Genomic Surveillance Network |
| EPI_ISL_2196189 | Laboratorio Central de Saude Publica do Estado do Rio de Janeiro (LACEN-RJ) | Laboratory of Respiratory Viruses and Measles, Oswaldo Cruz Institute, FIOCRUZ | Paola Resende, Luciana Appolinario, Fernando Motta, Anna Carolina Paixao, Ana Carolina Mendonca, Alice Sampaio Rocha, Taina Venas, Elisa Cavalcante Pereira, Renata Serrano Lopes, Andrea Cony Cavalcanti, Marilda Siqueira on behalf of the Fiocruz COVID-19 Genomic Surveillance Network |
| EPI_ISL_2196190, EPI_ISL_2196191, EPI_ISL_2196192, EPI_ISL_2196193, EPI_ISL_2196194, EPI_ISL_2196195, EPI_ISL_2196196 | Laboratory of Respiratory Viruses and Measles, Oswaldo Cruz Institute, FIOCRUZ | Laboratory of Respiratory Viruses and Measles, Oswaldo Cruz Institute, FIOCRUZ | Paola Resende, Luciana Appolinario, Fernando Motta, Anna Carolina Paixao, Ana Carolina Mendonca, Alice Sampaio Rocha, Taina Venas, Elisa Cavalcante Pereira, Renata Serrano Lopes, Marilda Siqueira on behalf of the Fiocruz COVID-19 Genomic Surveillance Network |
| EPI_ISL_2196197, EPI_ISL_2196198, EPI_ISL_2196199, EPI_ISL_2196200, EPI_ISL_2196201, EPI_ISL_2196202, EPI_ISL_2196203, EPI_ISL_2196204, EPI_ISL_2196205, EPI_ISL_2196206, EPI_ISL_2196207, EPI_ISL_2196208, EPI_ISL_2196209 | | | |
| see above | Laboratorio Central de Saude Publica do Estado do Rio de Janeiro (LACEN-RJ) | Laboratory of Respiratory Viruses and Measles, Oswaldo Cruz Institute, FIOCRUZ | Paola Resende, Luciana Appolinario, Fernando Motta, Anna Carolina Paixao, Ana Carolina Mendonca, Alice Sampaio Rocha, Taina Venas, Elisa Cavalcante Pereira, Renata Serrano Lopes, Andrea Cony Cavalcanti, Marilda Siqueira on behalf of the Fiocruz COVID-19 Genomic Surveillance Network |
| EPI_ISL_2196211, EPI_ISL_2196213, EPI_ISL_2196215, EPI_ISL_2196220 | Laboratorio Central de Saude Publica do Estado de Sergipe (LACEN/SE) | Laboratory of Respiratory Viruses and Measles, Oswaldo Cruz Institute, FIOCRUZ | Paola Resende, Luciana Appolinario, Fernando Motta, Anna Carolina Paixao, Ana Carolina Mendonca, Alice Sampaio Rocha, Tainá Moreira Martins Venas, Elisa Cavalcante Pereira, Renata Serrano Lopes, Cliomar Alves dos Santos, Marilda Siqueira on behalf of the Fiocruz COVID-19 Genomic Surveillance Network |
| EPI_ISL_2196236 | Laboratorio Central de Saude Publica do Estado do Rio de Janeiro (LACEN-RJ) | Laboratory of Respiratory Viruses and Measles, Oswaldo Cruz Institute, FIOCRUZ | Paola Resende, Luciana Appolinario, Fernando Motta, Anna Carolina Paixao, Ana Carolina Mendonca, Alice Sampaio Rocha, Taina Venas, Elisa Cavalcante Pereira, Renata Serrano Lopes, Andrea Cony Cavalcanti, Marilda Siqueira on behalf of the Fiocruz COVID-19 Genomic Surveillance Network |
| EPI_ISL_2196242, EPI_ISL_2196245 | Labortorio Central de Saude Publica do Estado do Rio de Janeiro (LACEN/RJ) | Laboratory of Respiratory Viruses and Measles, Oswaldo Cruz Institute, FIOCRUZ | Paola Resende, Luciana Appolinario, Fernando Motta, Anna Carolina Paixao, Ana Carolina Mendonca, Alice Sampaio Rocha, Taina Venas, Elisa Cavalcante Pereira, Renata Serrano Lopes, Andrea Cony Cavalcanti, Marilda Siqueira on behalf of the Fiocruz COVID-19 Genomic Surveillance Network |
| EPI_ISL_2196251 | Laboratory of Respiratory Viruses and Measles, Oswaldo Cruz Institute, FIOCRUZ | Laboratory of Respiratory Viruses and Measles, Oswaldo Cruz Institute, FIOCRUZ | Paola Resende, Luciana Appolinario, Fernando Motta, Anna Carolina Paixao, Ana Carolina Mendonca, Alice Sampaio Rocha, Taina Venas, Elisa Cavalcante Pereira, Renata Serrano Lopes, Marilda Siqueira on behalf of the Fiocruz COVID-19 Genomic Surveillance Network |
| EPI_ISL_2196267 | Laboratorio Central de Saude Publica do Estado Maranhao (LACEN-MA) | Laboratory of Respiratory Viruses and Measles, Oswaldo Cruz Institute, FIOCRUZ | Paola Resende, Luciana Appolinario, Fernando Motta, Anna Carolina Paixao, Ana Carolina Mendonca, Alice Sampaio Rocha, Taina Venas, Elisa Cavalcante Pereira, Renata Serrano Lopes, Lidio Gonçalves Lima Neto, Marilda Siqueira on behalf of the Fiocruz COVID-19 Genomic Surveillance Network |
| EPI_ISL_2196313, EPI_ISL_2196314, EPI_ISL_2196315, EPI_ISL_2196316, EPI_ISL_2196317, EPI_ISL_2196318, EPI_ISL_2196319, EPI_ISL_2196320, EPI_ISL_2196321, EPI_ISL_2196322, EPI_ISL_2196323, EPI_ISL_2196324, EPI_ISL_2196325, EPI_ISL_2196326, EPI_ISL_2196327, EPI_ISL_2196328, EPI_ISL_2196329, EPI_ISL_2196330, EPI_ISL_2196331, EPI_ISL_2196332, EPI_ISL_2196333, EPI_ISL_2196334, EPI_ISL_2196356 | | | |
| see above | Labortorio Central de Saude Publica do Estado do Rio de Janeiro (LACEN/RJ) | Laboratory of Respiratory Viruses and Measles, Oswaldo Cruz Institute, FIOCRUZ | Paola Resende, Luciana Appolinario, Fernando Motta, Anna Carolina Paixao, Ana Carolina Mendonca, Alice Sampaio Rocha, Taina Venas, Elisa Cavalcante Pereira, Renata Serrano Lopes, Andrea Cony Cavalcanti, Marilda Siqueira on behalf of the Fiocruz COVID-19 Genomic Surveillance Network |
| EPI_ISL_2196361 | Laboratory of Respiratory Viruses and Measles, Oswaldo Cruz Institute, FIOCRUZ | Laboratory of Respiratory Viruses and Measles, Oswaldo Cruz Institute, FIOCRUZ | Paola Resende, Luciana Appolinario, Fernando Motta, Anna Carolina Paixao, Ana Carolina Mendonca, Alice Sampaio Rocha, Taina Venas, Elisa Cavalcante Pereira, Renata Serrano Lopes, Marilda Siqueira on behalf of the Fiocruz COVID-19 Genomic Surveillance Network |
| EPI_ISL_2209955 | SECRETARIA DE SAUDE DE SAO PEDRO | Instituto Butantan / FZEA-USP-Pirassununga | Dimas Tadeu Covas, Antonio Jorge Martins, Claudia Renata dos Santos Barros, David Schlesinger, Debora Botequio Moretti, Elaine Cristina Marqueze, Elaine Vieira Santos, Evandra Strazza Rodrigues, Heidge Fukumasu, Jayme Augusto de Souza-Neto, José Salvatore Leister Patané, Luiz Alcantara, Luiz Lehmann Coutinho, Maria Carolina Elias, Maurício Lacerda Nogueira, Rafael dos Santos Bezerra, Raul Machado Neto, Rejane Maria Tommasini Grotto, Ricardo Haddad, Sandra Coccuzzo Sampaio Vessoni, Simone Kashima, Svetoslav Nanev Slavov, Vincent Louis Viala |
| EPI_ISL_2225349 | Houston Methodist Hospital | Houston Methodist Hospital | Randall J. Olsen, Paul A. Christensen, S. Wesley Long, Sishir Subedi, Robert Olson, Marcus Nguyen, James J. Davis, Matthew Ojeda Saavedra, Prasanti Yerramilli, Layne Pruitt, Kristina Reppond, Madison N. Shyer, Jessica Cambric, Ryan Gadd, Ilya J. Finkelstein, Jimmy Gollihar, and James M. Musser |
| EPI_ISL_2241579 | Laboratório Central de Saúde Pública de Sergipe | Coordenação Geral de Laboratórios de Saúde Pública (CGLAB/DAEVS/SVS/MS) | Vagner Fonseca, et al. |
| EPI_ISL_2241581 | Laboratório Central de Saúde Pública da Paraiba | Coordenação Geral de Laboratórios de Saúde Pública (CGLAB/DAEVS/SVS/MS) | Vagner Fonseca, et al. |
| EPI_ISL_2241589 | Laboratório Central de Saúde Pública de Sergipe | Coordenação Geral de Laboratórios de Saúde Pública (CGLAB/DAEVS/SVS/MS) | Vagner Fonseca, et al. |
| EPI_ISL_2249357, EPI_ISL_2249361, EPI_ISL_2249365 | Laboratório Central de Saúde Pública do Rio Grande do Sul | Coordenação Geral de Laboratórios de Saúde Pública (CGLAB/DAEVS/SVS/MS) | Vagner Fonseca, et al. |
| EPI_ISL_2249404, EPI_ISL_2249405, EPI_ISL_2249406, EPI_ISL_2249407, EPI_ISL_2249408, EPI_ISL_2249409, EPI_ISL_2249410, EPI_ISL_2249411, EPI_ISL_2249412, EPI_ISL_2249413, EPI_ISL_2249414, EPI_ISL_2249415, EPI_ISL_2249416, EPI_ISL_2249417, EPI_ISL_2249418, EPI_ISL_2249419, EPI_ISL_2249420, EPI_ISL_2249421, EPI_ISL_2249422, EPI_ISL_2249423, EPI_ISL_2249424, EPI_ISL_2249425, EPI_ISL_2249426, EPI_ISL_2249427, EPI_ISL_2249428, EPI_ISL_2249429, EPI_ISL_2249430, EPI_ISL_2249431, EPI_ISL_2249432, EPI_ISL_2249433, EPI_ISL_2249434, EPI_ISL_2249440, EPI_ISL_2249441, EPI_ISL_2249442, EPI_ISL_2249443, EPI_ISL_2249444, EPI_ISL_2249445 | | | |
| see above | Laboratório Central de Saúde Pública do Rio de Janeiro | Coordenação Geral de Laboratórios de Saúde Pública (CGLAB/DAEVS/SVS/MS) | Vagner Fonseca, et al. |
| EPI_ISL_2269371 | Fulgent Genetics | Centers for Disease Control and Prevention Division of Viral Diseases, Pathogen Discovery | Dakota Howard, Dhwani Batra, Peter W. Cook, Kara Moser, Adrian Paskey, Jason Caravas, Benjamin Rambo-Martin, Shatavia Morrison, Christopher Gulvick, Scott Sammons, Yvette Unoarumhi, Darlene Wagner, Matthew Schmerer, Harry Gao, Mickey Li, John Gao, Joseph Fierro, Benafsh Sapra, Becky Tsai, Yan Meng, Doreen Ng, James Xie, Clinton R. Paden, Duncan MacCannell |
| EPI_ISL_2274080 | Laboratorio Central de Saude Publica do Estado Maranhao (LACEN-MA) | Laboratory of Respiratory Viruses and Measles, Oswaldo Cruz Institute, FIOCRUZ | Paola Resende, Luciana Appolinario, Fernando Motta, Anna Carolina Paixao, Ana Carolina Mendonca, Alice Sampaio Rocha, Taina Venas, Elisa Cavalcante Pereira, Renata Serrano Lopes, Lidio Gonçalves Lima Neto, Marilda Siqueira on behalf of the Fiocruz COVID-19 Genomic Surveillance Network |
| EPI_ISL_2274096 | Lboratorio Central de Saude Publica do Estado do Parana (LACEN/PR) | Laboratory of Respiratory Viruses and Measles, Oswaldo Cruz Institute, FIOCRUZ | Paola Resende, Luciana Appolinario, Fernando Motta, Anna Carolina Paixao, Ana Carolina Mendonca, Alice Sampaio Rocha, Taina Venas, Elisa Cavalcante Pereira, Renata Serrano Lopes, Irina Riediger, Marilda Siqueira on behalf of the Fiocruz COVID-19 Genomic Surveillance Network |
| EPI_ISL_2274097 | Laboratory of Respiratory Viruses and Measles, Oswaldo Cruz Institute, FIOCRUZ | Laboratory of Respiratory Viruses and Measles, Oswaldo Cruz Institute, FIOCRUZ | Paola Resende, Luciana Appolinario, Fernando Motta, Anna Carolina Paixao, Ana Carolina Mendonca, Alice Sampaio Rocha, Taina Venas, Elisa Cavalcante Pereira, Renata Serrano Lopes, Marilda Siqueira on behalf of the Fiocruz COVID-19 Genomic Surveillance Network |
| EPI_ISL_2274098, EPI_ISL_2274099 | Labortorio Central de Saude Publica do Estado do Rio de Janeiro (LACEN/RJ) | Laboratory of Respiratory Viruses and Measles, Oswaldo Cruz Institute, FIOCRUZ | Paola Resende, Luciana Appolinario, Fernando Motta, Anna Carolina Paixao, Ana Carolina Mendonca, Alice Sampaio Rocha, Taina Venas, Elisa Cavalcante Pereira, Renata Serrano Lopes, Andrea Cony Cavalcanti, Marilda Siqueira on behalf of the Fiocruz COVID-19 Genomic Surveillance Network |
| EPI_ISL_2274100, EPI_ISL_2274101, | Laboratory of Respiratory Viruses and Measles, Oswaldo Cruz | Laboratory of Respiratory Viruses and Measles, Oswaldo Cruz | Paola Resende, Luciana Appolinario, Fernando Motta, Anna Carolina Paixao, Ana Carolina Mendonca, Alice Sampaio Rocha, Taina Venas, Elisa |

| | | | |
|---|---|---|---|
| EPI_ISL_2274102, EPI_ISL_2274103, EPI_ISL_2274104, EPI_ISL_2274105, EPI_ISL_2274106, EPI_ISL_2274107 | Institute, FIOCRUZ | Institute, FIOCRUZ | Cavalcante Pereira, Renata Serrano Lopes, Marilda Siqueira on behalf of the Fiocruz COVID-19 Genomic Surveillance Network |
| EPI_ISL_2274108, EPI_ISL_2274109 | Laboratorio Central de Saude Publica do Estado do Rio Grande do Sul (LACEN-RS) | Laboratory of Respiratory Viruses and Measles, Oswaldo Cruz Institute, FIOCRUZ | Paola Resende, Luciana Appolinario, Fernando Motta, Anna Carolina Paixao, Ana Carolina Mendonca, Alice Sampaio Rocha, Taina Venas, Elisa Cavalcante Pereira, Renata Serrano Lopes, Tatiana Schaffer Gregianini, Richard Salvato, Marilda Siqueira on behalf of the Fiocruz COVID-19 Genomic Surveillance Network |
| EPI_ISL_2308456, EPI_ISL_2308471, EPI_ISL_2308473, EPI_ISL_2308475, EPI_ISL_2308478 | Laboratório Central de Saúde Pública de Sergipe | Coordenação Geral de Laboratórios de Saúde Pública (CGLAB/DAEVS/SVS/MS) | Vagner Fonseca, et al. |
| EPI_ISL_2344442 | Instituto Butantan | Instituto de Medicina Tropical de Sao Paulo | Brazil-UK Centre for Arbovirus Discovery Diagnosis Genomics and Epidemiology (CADDE) Genomic Network - Instituto de Medicina Tropical |
| EPI_ISL_2344552 | HOSPITAL DOS FORNCEDORES | Instituto Butantan / ESALQ-Piracicaba | Dimas Tadeu Covas, Antonio Jorge Martins, Claudia Renata dos Santos Barros, David Schlesinger, Debora Botequio Moretti, Elaine Cristina Marqueze, Elaine Vieira Santos, Evandra Strazza Rodrigues, Heidge Fukumasu, Jayme Augusto de Souza-Neto, José Salvatore Leister Patané, Luiz Alcantara, Luiz Lehmann Coutinho, Maria Carolina Elias, Maurício Lacerda Nogueira, Rafael dos Santos Bezerra, Raul Machado Neto, Rejane Maria Tommasini Grotto, Ricardo Haddad, Sandra Coccuzzo Sampaio Vessoni, Simone Kashima, Svetoslav Nanev Slavov, Vincent Louis Viala |
| EPI_ISL_2344553, EPI_ISL_2344554 | LABORATORIO MUNICIPAL DE PIRACICABA | Instituto Butantan / ESALQ-Piracicaba | Dimas Tadeu Covas, Antonio Jorge Martins, Claudia Renata dos Santos Barros, David Schlesinger, Debora Botequio Moretti, Elaine Cristina Marqueze, Elaine Vieira Santos, Evandra Strazza Rodrigues, Heidge Fukumasu, Jayme Augusto de Souza-Neto, José Salvatore Leister Patané, Luiz Alcantara, Luiz Lehmann Coutinho, Maria Carolina Elias, Maurício Lacerda Nogueira, Rafael dos Santos Bezerra, Raul Machado Neto, Rejane Maria Tommasini Grotto, Ricardo Haddad, Sandra Coccuzzo Sampaio Vessoni, Simone Kashima, Svetoslav Nanev Slavov, Vincent Louis Viala |
| EPI_ISL_2344662 | SECRETARIA DE SAUDE DE SAO PEDRO | Instituto Butantan / ESALQ-Piracicaba | Dimas Tadeu Covas, Antonio Jorge Martins, Claudia Renata dos Santos Barros, David Schlesinger, Debora Botequio Moretti, Elaine Cristina Marqueze, Elaine Vieira Santos, Evandra Strazza Rodrigues, Heidge Fukumasu, Jayme Augusto de Souza-Neto, José Salvatore Leister Patané, Luiz Alcantara, Luiz Lehmann Coutinho, Maria Carolina Elias, Maurício Lacerda Nogueira, Rafael dos Santos Bezerra, Raul Machado Neto, Rejane Maria Tommasini Grotto, Ricardo Haddad, Sandra Coccuzzo Sampaio Vessoni, Simone Kashima, Svetoslav Nanev Slavov, Vincent Louis Viala |
| EPI_ISL_2344814 | ESTRATEGIA DE SAUDE DA FAMILIA DE ARCO IRIS | Instituto Butantan / ESALQ-Piracicaba | Dimas Tadeu Covas, Antonio Jorge Martins, Claudia Renata dos Santos Barros, David Schlesinger, Debora Botequio Moretti, Elaine Cristina Marqueze, Elaine Vieira Santos, Evandra Strazza Rodrigues, Heidge Fukumasu, Jayme Augusto de Souza-Neto, José Salvatore Leister Patané, Luiz Alcantara, Luiz Lehmann Coutinho, Maria Carolina Elias, Maurício Lacerda Nogueira, Rafael dos Santos Bezerra, Raul Machado Neto, Rejane Maria Tommasini Grotto, Ricardo Haddad, Sandra Coccuzzo Sampaio Vessoni, Simone Kashima, Svetoslav Nanev Slavov, Vincent Louis Viala |
| EPI_ISL_2344851 | LABORATORIO MUNICIPAL DE PIRACICABA | Instituto Butantan / ESALQ-Piracicaba | Dimas Tadeu Covas, Antonio Jorge Martins, Claudia Renata dos Santos Barros, David Schlesinger, Debora Botequio Moretti, Elaine Cristina Marqueze, Elaine Vieira Santos, Evandra Strazza Rodrigues, Heidge Fukumasu, Jayme Augusto de Souza-Neto, José Salvatore Leister Patané, Luiz Alcantara, Luiz Lehmann Coutinho, Maria Carolina Elias, Maurício Lacerda Nogueira, Rafael dos Santos Bezerra, Raul Machado Neto, Rejane Maria Tommasini Grotto, Ricardo Haddad, Sandra Coccuzzo Sampaio Vessoni, Simone Kashima, Svetoslav Nanev Slavov, Vincent Louis Viala |
| EPI_ISL_2344900 | HOSPITAL E PRONTO SOCORRO PORTINARI | Instituto Butantan | Dimas Tadeu Covas, Antonio Jorge Martins, Claudia Renata dos Santos Barros, David Schlesinger, Debora Botequio Moretti, Elaine Cristina Marqueze, Elaine Vieira Santos, Evandra Strazza Rodrigues, Heidge Fukumasu, Jayme Augusto de Souza-Neto, José Salvatore Leister Patané, Luiz Alcantara, Luiz Lehmann Coutinho, Maria Carolina Elias, Maurício Lacerda Nogueira, Rafael dos Santos Bezerra, Raul Machado Neto, Rejane Maria Tommasini Grotto, Ricardo Haddad, Sandra Coccuzzo Sampaio Vessoni, Simone Kashima, Svetoslav Nanev Slavov, Vincent Louis Viala |
| EPI_ISL_2344915 | LABORATORIO MUNICIPAL DE PIRACICABA | Instituto Butantan / ESALQ-Piracicaba | Dimas Tadeu Covas, Antonio Jorge Martins, Claudia Renata dos Santos Barros, David Schlesinger, Debora Botequio Moretti, Elaine Cristina Marqueze, Elaine Vieira Santos, Evandra Strazza Rodrigues, Heidge Fukumasu, Jayme Augusto de Souza-Neto, José Salvatore Leister Patané, Luiz Alcantara, Luiz Lehmann Coutinho, Maria Carolina Elias, Maurício Lacerda Nogueira, Rafael dos Santos Bezerra, Raul Machado Neto, Rejane Maria Tommasini Grotto, Ricardo Haddad, Sandra Coccuzzo Sampaio Vessoni, Simone Kashima, Svetoslav Nanev Slavov, Vincent Louis Viala |
| EPI_ISL_2345006 | SANTA CASA DE MISERICORDIA SAO JOSE | Instituto Butantan / ESALQ-Piracicaba | Dimas Tadeu Covas, Antonio Jorge Martins, Claudia Renata dos Santos Barros, David Schlesinger, Debora Botequio Moretti, Elaine Cristina Marqueze, Elaine Vieira Santos, Evandra Strazza Rodrigues, Heidge Fukumasu, Jayme Augusto de Souza-Neto, José Salvatore Leister Patané, Luiz Alcantara, Luiz Lehmann Coutinho, Maria Carolina Elias, Maurício Lacerda Nogueira, Rafael dos Santos Bezerra, Raul Machado Neto, Rejane Maria Tommasini Grotto, Ricardo Haddad, Sandra Coccuzzo Sampaio Vessoni, Simone Kashima, Svetoslav Nanev Slavov, Vincent Louis Viala |
| EPI_ISL_2345181 | NUCLEO SAUDE III N HABITACIONAL PRESIDENTE GEISEL BAURU | Instituto Butantan / ESALQ-Piracicaba | Dimas Tadeu Covas, Antonio Jorge Martins, Claudia Renata dos Santos Barros, David Schlesinger, Debora Botequio Moretti, Elaine Cristina Marqueze, Elaine Vieira Santos, Evandra Strazza Rodrigues, Heidge Fukumasu, Jayme Augusto de Souza-Neto, José Salvatore Leister Patané, Luiz Alcantara, Luiz Lehmann Coutinho, Maria Carolina Elias, Maurício Lacerda Nogueira, Rafael dos Santos Bezerra, Raul Machado Neto, Rejane Maria Tommasini Grotto, Ricardo Haddad, Sandra Coccuzzo Sampaio Vessoni, Simone Kashima, Svetoslav Nanev Slavov, Vincent Louis Viala |
| EPI_ISL_2345207 | NUCLEO DE SAUDE VILA FALCAO DE BAURU | Instituto Butantan / ESALQ-Piracicaba | Dimas Tadeu Covas, Antonio Jorge Martins, Claudia Renata dos Santos Barros, David Schlesinger, Debora Botequio Moretti, Elaine Cristina Marqueze, Elaine Vieira Santos, Evandra Strazza Rodrigues, Heidge Fukumasu, Jayme Augusto de Souza-Neto, José Salvatore Leister Patané, Luiz Alcantara, Luiz Lehmann Coutinho, Maria Carolina Elias, Maurício Lacerda Nogueira, Rafael dos Santos Bezerra, Raul Machado Neto, Rejane Maria Tommasini Grotto, Ricardo Haddad, Sandra Coccuzzo Sampaio Vessoni, Simone Kashima, Svetoslav Nanev Slavov, Vincent Louis Viala |
| EPI_ISL_2345267 | LABORATORIO MUNICIPAL DE PIRACICABA | Instituto Butantan / ESALQ-Piracicaba | Dimas Tadeu Covas, Antonio Jorge Martins, Claudia Renata dos Santos Barros, David Schlesinger, Debora Botequio Moretti, Elaine Cristina Marqueze, Elaine Vieira Santos, Evandra Strazza Rodrigues, Heidge Fukumasu, Jayme Augusto de Souza-Neto, José Salvatore Leister Patané, Luiz Alcantara, Luiz Lehmann Coutinho, Maria Carolina Elias, Maurício Lacerda Nogueira, Rafael dos Santos Bezerra, Raul Machado Neto, Rejane Maria Tommasini Grotto, Ricardo Haddad, Sandra Coccuzzo Sampaio Vessoni, Simone Kashima, Svetoslav Nanev Slavov, Vincent Louis Viala |
| EPI_ISL_2345268 | UPA IPIRANGA | Instituto Butantan / ESALQ-Piracicaba | Dimas Tadeu Covas, Antonio Jorge Martins, Claudia Renata dos Santos Barros, David Schlesinger, Debora Botequio Moretti, Elaine Cristina Marqueze, Elaine Vieira Santos, Evandra Strazza Rodrigues, Heidge Fukumasu, Jayme Augusto de Souza-Neto, José Salvatore Leister Patané, Luiz Alcantara, Luiz Lehmann Coutinho, Maria Carolina Elias, Maurício Lacerda Nogueira, Rafael dos Santos Bezerra, Raul Machado Neto, Rejane Maria Tommasini Grotto, Ricardo Haddad, Sandra Coccuzzo Sampaio Vessoni, Simone Kashima, Svetoslav Nanev Slavov, Vincent Louis Viala |
| EPI_ISL_2345453 | HOSPITAL REGIONAL DE ITAPETININGA | Instituto Butantan / ESALQ-Piracicaba | Dimas Tadeu Covas, Antonio Jorge Martins, Claudia Renata dos Santos Barros, David Schlesinger, Debora Botequio Moretti, Elaine Cristina Marqueze, Elaine Vieira Santos, Evandra Strazza Rodrigues, Heidge Fukumasu, Jayme Augusto de Souza-Neto, José Salvatore Leister Patané, Luiz Alcantara, Luiz Lehmann Coutinho, Maria Carolina Elias, Maurício Lacerda Nogueira, Rafael dos Santos Bezerra, Raul Machado Neto, Rejane Maria Tommasini Grotto, Ricardo Haddad, Sandra Coccuzzo Sampaio Vessoni, Simone Kashima, Svetoslav Nanev Slavov, Vincent Louis Viala |
| EPI_ISL_2345590 | CENTRO DE ESPECIALIDADES DE PRIMAVERA | Instituto Butantan / ESALQ-Piracicaba | Dimas Tadeu Covas, Antonio Jorge Martins, Claudia Renata dos Santos Barros, David Schlesinger, Debora Botequio Moretti, Elaine Cristina Marqueze, Elaine Vieira Santos, Evandra Strazza Rodrigues, Heidge Fukumasu, Jayme Augusto de Souza-Neto, José Salvatore Leister Patané, Luiz Alcantara, Luiz Lehmann Coutinho, Maria Carolina Elias, Maurício Lacerda Nogueira, Rafael dos Santos Bezerra, Raul Machado Neto, Rejane Maria Tommasini Grotto, Ricardo Haddad, Sandra Coccuzzo Sampaio Vessoni, Simone Kashima, Svetoslav Nanev Slavov, Vincent Louis Viala |
| EPI_ISL_2345595, EPI_ISL_2345597, EPI_ISL_2345601 | UBS III DE RANCHARIA | Instituto Butantan / ESALQ-Piracicaba | Dimas Tadeu Covas, Antonio Jorge Martins, Claudia Renata dos Santos Barros, David Schlesinger, Debora Botequio Moretti, Elaine Cristina Marqueze, Elaine Vieira Santos, Evandra Strazza Rodrigues, Heidge Fukumasu, Jayme Augusto de Souza-Neto, José Salvatore Leister Patané, Luiz Alcantara, Luiz Lehmann Coutinho, Maria Carolina Elias, Maurício Lacerda Nogueira, Rafael dos Santos Bezerra, Raul Machado Neto, Rejane Maria Tommasini Grotto, Ricardo Haddad, Sandra Coccuzzo Sampaio Vessoni, Simone Kashima, Svetoslav Nanev Slavov, Vincent Louis Viala |
| EPI_ISL_2345715 | CENTRO DE ESPECIALIDADES DE PRIMAVERA | Instituto Butantan / Mendelics | Dimas Tadeu Covas, Antonio Jorge Martins, Claudia Renata dos Santos Barros, David Schlesinger, Debora Botequio Moretti, Elaine Cristina Marqueze, Elaine Vieira Santos, Evandra Strazza Rodrigues, Heidge Fukumasu, Jayme Augusto de Souza-Neto, José Salvatore Leister Patané, Luiz Alcantara, Luiz Lehmann Coutinho, Maria Carolina Elias, Maurício Lacerda Nogueira, Rafael dos Santos Bezerra, Raul Machado Neto, Rejane Maria Tommasini Grotto, Ricardo Haddad, Sandra Coccuzzo Sampaio Vessoni, Simone Kashima, Svetoslav Nanev Slavov, Vincent Louis Viala |
| EPI_ISL_2345723 | UNIDADE BASICA DE SAUDE JARDIM DO LAGO | Instituto Butantan / Mendelics | Dimas Tadeu Covas, Antonio Jorge Martins, Claudia Renata dos Santos Barros, David Schlesinger, Debora Botequio Moretti, Elaine Cristina Marqueze, Elaine Vieira Santos, Evandra Strazza Rodrigues, Heidge Fukumasu, Jayme Augusto de Souza-Neto, José Salvatore Leister Patané, Luiz Alcantara, Luiz Lehmann Coutinho, Maria Carolina Elias, Maurício Lacerda Nogueira, Rafael dos Santos Bezerra, Raul Machado Neto, Rejane Maria Tommasini Grotto, |

| | | | |
|---|---|---|---|
| | | | Ricardo Haddad, Sandra Coccuzzo Sampaio Vessoni, Simone Kashima, Svetoslav Nanev Slavov, Vincent Louis Viala |
| EPI_ISL_2345755 | SECRETARIA MUNICIPAL DE SAUDE SOROCABA | Instituto Butantan / Mendelics | Dimas Tadeu Covas, Antonio Jorge Martins, Claudia Renata dos Santos Barros, David Schlesinger, Debora Botequio Moretti, Elaine Cristina Marqueze, Elaine Vieira Santos, Evandra Strazza Rodrigues, Heidge Fukumasu, Jayme Augusto de Souza-Neto, José Salvatore Leister Patané, Luiz Alcantara, Luiz Lehmann Coutinho, Maria Carolina Elias, Maurício Lacerda Nogueira, Rafael dos Santos Bezerra, Raul Machado Neto, Rejane Maria Tommasini Grotto, Ricardo Haddad, Sandra Coccuzzo Sampaio Vessoni, Simone Kashima, Svetoslav Nanev Slavov, Vincent Louis Viala |
| EPI_ISL_2345811 | CENTRO DE ESPECIALIDADES DE PRIMAVERA | Instituto Butantan / Mendelics | Dimas Tadeu Covas, Antonio Jorge Martins, Claudia Renata dos Santos Barros, David Schlesinger, Debora Botequio Moretti, Elaine Cristina Marqueze, Elaine Vieira Santos, Evandra Strazza Rodrigues, Heidge Fukumasu, Jayme Augusto de Souza-Neto, José Salvatore Leister Patané, Luiz Alcantara, Luiz Lehmann Coutinho, Maria Carolina Elias, Maurício Lacerda Nogueira, Rafael dos Santos Bezerra, Raul Machado Neto, Rejane Maria Tommasini Grotto, Ricardo Haddad, Sandra Coccuzzo Sampaio Vessoni, Simone Kashima, Svetoslav Nanev Slavov, Vincent Louis Viala |
| EPI_ISL_2345874 | CENTRO DE APOIO EPIDEMIOLOGICO | Instituto Butantan / Mendelics | Dimas Tadeu Covas, Antonio Jorge Martins, Claudia Renata dos Santos Barros, David Schlesinger, Debora Botequio Moretti, Elaine Cristina Marqueze, Elaine Vieira Santos, Evandra Strazza Rodrigues, Heidge Fukumasu, Jayme Augusto de Souza-Neto, José Salvatore Leister Patané, Luiz Alcantara, Luiz Lehmann Coutinho, Maria Carolina Elias, Maurício Lacerda Nogueira, Rafael dos Santos Bezerra, Raul Machado Neto, Rejane Maria Tommasini Grotto, Ricardo Haddad, Sandra Coccuzzo Sampaio Vessoni, Simone Kashima, Svetoslav Nanev Slavov, Vincent Louis Viala |
| EPI_ISL_2346088, EPI_ISL_2346090 | Prefeirtura de SP | Instituto Butantan | Dimas Tadeu Covas, Antonio Jorge Martins, Claudia Renata dos Santos Barros, David Schlesinger, Debora Botequio Moretti, Elaine Cristina Marqueze, Elaine Vieira Santos, Evandra Strazza Rodrigues, Heidge Fukumasu, Jayme Augusto de Souza-Neto, José Salvatore Leister Patané, Luiz Alcantara, Luiz Lehmann Coutinho, Maria Carolina Elias, Maurício Lacerda Nogueira, Rafael dos Santos Bezerra, Raul Machado Neto, Rejane Maria Tommasini Grotto, Ricardo Haddad, Sandra Coccuzzo Sampaio Vessoni, Simone Kashima, Svetoslav Nanev Slavov, Vincent Louis Viala |
| EPI_ISL_2362244 | HOSPITAL DE CAMPANHA COVID 19 MUNICIPIO DE TAUBATE | Instituto Butantan / Mendelics | Dimas Tadeu Covas, Antonio Jorge Martins, Claudia Renata dos Santos Barros, David Schlesinger, Debora Botequio Moretti, Elaine Cristina Marqueze, Elaine Vieira Santos, Evandra Strazza Rodrigues, Heidge Fukumasu, Jayme Augusto de Souza-Neto, José Salvatore Leister Patané, Luiz Alcantara, Luiz Lehmann Coutinho, Maria Carolina Elias, Maurício Lacerda Nogueira, Rafael dos Santos Bezerra, Raul Machado Neto, Rejane Maria Tommasini Grotto, Ricardo Haddad, Sandra Coccuzzo Sampaio Vessoni, Simone Kashima, Svetoslav Nanev Slavov, Vincent Louis Viala |
| EPI_ISL_2362257 | SANTA CASA DE MISERICORDIA SAO JOSE | Instituto Butantan / Mendelics | Dimas Tadeu Covas, Antonio Jorge Martins, Claudia Renata dos Santos Barros, David Schlesinger, Debora Botequio Moretti, Elaine Cristina Marqueze, Elaine Vieira Santos, Evandra Strazza Rodrigues, Heidge Fukumasu, Jayme Augusto de Souza-Neto, José Salvatore Leister Patané, Luiz Alcantara, Luiz Lehmann Coutinho, Maria Carolina Elias, Maurício Lacerda Nogueira, Rafael dos Santos Bezerra, Raul Machado Neto, Rejane Maria Tommasini Grotto, Ricardo Haddad, Sandra Coccuzzo Sampaio Vessoni, Simone Kashima, Svetoslav Nanev Slavov, Vincent Louis Viala |
| EPI_ISL_2378653, EPI_ISL_2378681 | Instituto de Biotecnologia - UNESP-Botucatu-SP | Instituto de Biotecnologia - UNESP-Botucatu-SP | Fábio Sossai Possebon; Leila Sabrina Ullmann; Cecília Artico Banho; Cíntia Bittar; Guilherme Campos; Helena Lage Ferreira; Jorge A. Petroli Marchesi; Lívia Sacchetto; Maisa C. Pereira Parra; Marília Moraes; Maurício L. Nogueira; Paula Rahal; Paulo Inacio da Costa; João Pessoa Araújo Jr. |
| EPI_ISL_2378745 | HOSPITAL DO RIM E HIPERTENSAO | Instituto Butantan | Dimas Tadeu Covas, Antonio Jorge Martins, Claudia Renata dos Santos Barros, David Schlesinger, Debora Botequio Moretti, Elaine Cristina Marqueze, Elaine Vieira Santos, Evandra Strazza Rodrigues, Heidge Fukumasu, Jayme Augusto de Souza-Neto, José Salvatore Leister Patané, Luiz Alcantara, Luiz Lehmann Coutinho, Maria Carolina Elias, Maurício Lacerda Nogueira, Rafael dos Santos Bezerra, Raul Machado Neto, Rejane Maria Tommasini Grotto, Ricardo Haddad, Sandra Coccuzzo Sampaio Vessoni, Simone Kashima, Svetoslav Nanev Slavov, Vincent Louis Viala |
| EPI_ISL_2383033 | LABORATORIO MUNICIPAL DE PIRACICABA | Instituto Butantan | Dimas Tadeu Covas, Antonio Jorge Martins, Claudia Renata dos Santos Barros, David Schlesinger, Debora Botequio Moretti, Elaine Cristina Marqueze, Elaine Vieira Santos, Evandra Strazza Rodrigues, Heidge Fukumasu, Jayme Augusto de Souza-Neto, José Salvatore Leister Patané, Luiz Alcantara, Luiz Lehmann Coutinho, Maria Carolina Elias, Maurício Lacerda Nogueira, Rafael dos Santos Bezerra, Raul Machado Neto, Rejane Maria Tommasini Grotto, Ricardo Haddad, Sandra Coccuzzo Sampaio Vessoni, Simone Kashima, Svetoslav Nanev Slavov, Vincent Louis Viala |
| EPI_ISL_2383035 | UNIDADE BASICA DE SAUDE DR MATHEUS GABRIEL BONASSA | Instituto Butantan | Dimas Tadeu Covas, Antonio Jorge Martins, Claudia Renata dos Santos Barros, David Schlesinger, Debora Botequio Moretti, Elaine Cristina Marqueze, Elaine Vieira Santos, Evandra Strazza Rodrigues, Heidge Fukumasu, Jayme Augusto de Souza-Neto, José Salvatore Leister Patané, Luiz Alcantara, Luiz Lehmann Coutinho, Maria Carolina Elias, Maurício Lacerda Nogueira, Rafael dos Santos Bezerra, Raul Machado Neto, Rejane Maria Tommasini Grotto, Ricardo Haddad, Sandra Coccuzzo Sampaio Vessoni, Simone Kashima, Svetoslav Nanev Slavov, Vincent Louis Viala |
| EPI_ISL_2385432, EPI_ISL_2385433, EPI_ISL_2385434, EPI_ISL_2385435, EPI_ISL_2385436, EPI_ISL_2385437, EPI_ISL_2385438, EPI_ISL_2385439, EPI_ISL_2385440, EPI_ISL_2385441, EPI_ISL_2385442, EPI_ISL_2385443, EPI_ISL_2385444, EPI_ISL_2385445, EPI_ISL_2385446, EPI_ISL_2385447, EPI_ISL_2385448, EPI_ISL_2385449, EPI_ISL_2385450, EPI_ISL_2385451, EPI_ISL_2385452, EPI_ISL_2385453, EPI_ISL_2385454, EPI_ISL_2385455, EPI_ISL_2385456, EPI_ISL_2385457, EPI_ISL_2385458, EPI_ISL_2385459, EPI_ISL_2385460, EPI_ISL_2385461, EPI_ISL_2385462, EPI_ISL_2385463, EPI_ISL_2385464, EPI_ISL_2385465, EPI_ISL_2385466, EPI_ISL_2385467, EPI_ISL_2385468, EPI_ISL_2385469, EPI_ISL_2385470, EPI_ISL_2385471, EPI_ISL_2385472, EPI_ISL_2385474, EPI_ISL_2385475, EPI_ISL_2385476, EPI_ISL_2385477, EPI_ISL_2385478, EPI_ISL_2385479, EPI_ISL_2385480, EPI_ISL_2385481, EPI_ISL_2385482, EPI_ISL_2385483, EPI_ISL_2385484, EPI_ISL_2385485, EPI_ISL_2385486, EPI_ISL_2385487, EPI_ISL_2385488, EPI_ISL_2385489, EPI_ISL_2385490, EPI_ISL_2385491, EPI_ISL_2385492, EPI_ISL_2385493, EPI_ISL_2385494, EPI_ISL_2385495, EPI_ISL_2385496, EPI_ISL_2385498, EPI_ISL_2385499, EPI_ISL_2385500, EPI_ISL_2385501, EPI_ISL_2385502, EPI_ISL_2385503, EPI_ISL_2385504, EPI_ISL_2385505, EPI_ISL_2385506, EPI_ISL_2385507, EPI_ISL_2385508, EPI_ISL_2385510, EPI_ISL_2385511, EPI_ISL_2385512, EPI_ISL_2385513, EPI_ISL_2385514, EPI_ISL_2385515, EPI_ISL_2385516, EPI_ISL_2385517, EPI_ISL_2385518, EPI_ISL_2385519, EPI_ISL_2385520, EPI_ISL_2385521, EPI_ISL_2385522, EPI_ISL_2385523, EPI_ISL_2385524, EPI_ISL_2385525, EPI_ISL_2385526, EPI_ISL_2385527, EPI_ISL_2385529, EPI_ISL_2385530, EPI_ISL_2385531, EPI_ISL_2385532, EPI_ISL_2385533, EPI_ISL_2385534, EPI_ISL_2385535, EPI_ISL_2385536, EPI_ISL_2385537, EPI_ISL_2385538, EPI_ISL_2385539, EPI_ISL_2385541, EPI_ISL_2385542, EPI_ISL_2385543, EPI_ISL_2385544, EPI_ISL_2385545, EPI_ISL_2385546, EPI_ISL_2385547, EPI_ISL_2385548, EPI_ISL_2385549, EPI_ISL_2385550, EPI_ISL_2385551, EPI_ISL_2385552, EPI_ISL_2385553, EPI_ISL_2385554, EPI_ISL_2385555, EPI_ISL_2385556, EPI_ISL_2385558, EPI_ISL_2385559, EPI_ISL_2385561, EPI_ISL_2385562, EPI_ISL_2385563, EPI_ISL_2385564, EPI_ISL_2385565, EPI_ISL_2385566, EPI_ISL_2385567, EPI_ISL_2385568, EPI_ISL_2385570, EPI_ISL_2385571, EPI_ISL_2385572, EPI_ISL_2385573, EPI_ISL_2385574, EPI_ISL_2385575, EPI_ISL_2385576, EPI_ISL_2385577, EPI_ISL_2385578, EPI_ISL_2385579, EPI_ISL_2385580, EPI_ISL_2385581, EPI_ISL_2385582, EPI_ISL_2385583, EPI_ISL_2385584, EPI_ISL_2385586, EPI_ISL_2385587, EPI_ISL_2385588, EPI_ISL_2385589, EPI_ISL_2385590, EPI_ISL_2385591, EPI_ISL_2385592, EPI_ISL_2385593, EPI_ISL_2385594, EPI_ISL_2385595, EPI_ISL_2385596, EPI_ISL_2385597, EPI_ISL_2385598, EPI_ISL_2385599, EPI_ISL_2385600, EPI_ISL_2385601, EPI_ISL_2385602, EPI_ISL_2385603, EPI_ISL_2385604, EPI_ISL_2385605, EPI_ISL_2385606, EPI_ISL_2385607, EPI_ISL_2385608, EPI_ISL_2385609, EPI_ISL_2385610, EPI_ISL_2385611, EPI_ISL_2385612, EPI_ISL_2385613, EPI_ISL_2385616, EPI_ISL_2385620, EPI_ISL_2385624, EPI_ISL_2385625, EPI_ISL_2385627, EPI_ISL_2385629, EPI_ISL_2385630, EPI_ISL_2385631, EPI_ISL_2385632, EPI_ISL_2385633, EPI_ISL_2385634, EPI_ISL_2385636, EPI_ISL_2385637, EPI_ISL_2385639, EPI_ISL_2385641, EPI_ISL_2385642, EPI_ISL_2385644, EPI_ISL_2385647, EPI_ISL_2385648, EPI_ISL_2385649, EPI_ISL_2385650, EPI_ISL_2385651, EPI_ISL_2385652, EPI_ISL_2385653, EPI_ISL_2385654, EPI_ISL_2385655, EPI_ISL_2385656, EPI_ISL_2385657, EPI_ISL_2385658, EPI_ISL_2385659, EPI_ISL_2385660, EPI_ISL_2385661, EPI_ISL_2385662, EPI_ISL_2385663, EPI_ISL_2385664, EPI_ISL_2385665, EPI_ISL_2385666, EPI_ISL_2385667, EPI_ISL_2385668 | | | |
| see above | Laboratorio Central Noel Nutels | Bioinformatics Laboratory / LNCC | Luiz G P de Almeida, Alessandra P Lamarca, Ronaldo da Silva F Jr, Liliane Cavalcante, Alexandra L Gerber, Ana Paula de C Guimaraes, Douglas Terra Machado, Cassia Alves, Diana Mariani, Cintia Policarpo, Gleidson da Silva de Oliveira, Mario Sergio Ribeiro, Silvia Carvalho, Flavio Dias da Silva, Marcio Henrique de Oliveira Garcia, Leandro Magalhaes de Souza, Cristiane Gomes da Silva, Caio Luiz Pereira Ribeiro, Andrea Cony Cavalcanti, Claudia Maria Braga de Mello, Amilcar Tanuri, Ana Tereza R Vasconcelos |
| EPI_ISL_2385669, EPI_ISL_2385670, EPI_ISL_2385671, EPI_ISL_2385674, EPI_ISL_2385675, EPI_ISL_2385676, EPI_ISL_2385677, EPI_ISL_2385678, EPI_ISL_2385679, EPI_ISL_2385680, EPI_ISL_2385681, EPI_ISL_2385682, EPI_ISL_2385683, EPI_ISL_2385684, EPI_ISL_2385685, EPI_ISL_2385686, EPI_ISL_2385689, EPI_ISL_2385690, EPI_ISL_2385691, EPI_ISL_2385692, EPI_ISL_2385693, EPI_ISL_2385694, EPI_ISL_2385695, EPI_ISL_2385696, EPI_ISL_2385697, EPI_ISL_2385698, EPI_ISL_2385699, EPI_ISL_2385700, EPI_ISL_2385701, EPI_ISL_2385702, EPI_ISL_2385704, EPI_ISL_2385705, EPI_ISL_2385707, EPI_ISL_2385708, EPI_ISL_2385709, EPI_ISL_2385711, EPI_ISL_2385712, EPI_ISL_2385713, EPI_ISL_2385714, EPI_ISL_2385717, EPI_ISL_2385718, EPI_ISL_2385719, EPI_ISL_2385720, EPI_ISL_2385723, EPI_ISL_2385724, EPI_ISL_2385725, EPI_ISL_2385726, EPI_ISL_2385727, EPI_ISL_2385728, EPI_ISL_2385729, EPI_ISL_2385730, EPI_ISL_2385731, EPI_ISL_2385732, EPI_ISL_2385733, EPI_ISL_2385734, EPI_ISL_2385735, EPI_ISL_2385736, EPI_ISL_2385737, EPI_ISL_2385739, EPI_ISL_2385740, EPI_ISL_2385741, EPI_ISL_2385742, EPI_ISL_2385743, EPI_ISL_2385745, EPI_ISL_2385748, EPI_ISL_2385749, EPI_ISL_2385750, EPI_ISL_2385751, EPI_ISL_2385752, EPI_ISL_2385753, EPI_ISL_2385754, EPI_ISL_2385755, EPI_ISL_2385756, EPI_ISL_2385757, EPI_ISL_2385758, EPI_ISL_2385759, EPI_ISL_2385760, EPI_ISL_2385761, EPI_ISL_2385764, EPI_ISL_2385765, EPI_ISL_2385766, EPI_ISL_2385768, EPI_ISL_2385769, EPI_ISL_2385770, EPI_ISL_2385771, EPI_ISL_2385772, EPI_ISL_2385773, EPI_ISL_2385774, EPI_ISL_2385775, EPI_ISL_2385776, EPI_ISL_2385777, EPI_ISL_2385778, EPI_ISL_2385779, EPI_ISL_2385780, EPI_ISL_2385781, EPI_ISL_2385782, EPI_ISL_2385784, EPI_ISL_2385785, EPI_ISL_2385786 | | | |
| see above | Unidade de apoio ao diagnóstico da COVID - UNADIG | Bioinformatics Laboratory / LNCC | Luiz G P de Almeida, Alessandra P Lamarca, Ronaldo da Silva F Jr, Liliane Cavalcante, Alexandra L Gerber, Ana Paula de C Guimaraes, Douglas Terra Machado, Cassia Alves, Diana Mariani, Cintia Policarpo, Gleidson da Silva de Oliveira, Mario Sergio Ribeiro, Silvia Carvalho, Flavio Dias da Silva, Marcio Henrique de Oliveira Garcia, Leandro Magalhaes de Souza, Cristiane Gomes da Silva, Caio Luiz Pereira Ribeiro, Andrea Cony Cavalcanti, Claudia Maria Braga de Mello, Amilcar Tanuri, Ana Tereza R Vasconcelos |
| EPI_ISL_2431437 | Laboratório de Microbiologia Molecular - Universidade FEEVALE | Molecular Microbiology Laboratory | Alana Witt Hansen, Fágner Henrique Heldt, Fernando Rosado Spilki, Flávio Silveira, Juliana Schons Gularte, Juliane Deise Fleck, Mariana Soares da Silva, Meriane Demoliner, Matheus Nunes Weber, Paula Rodrigues de Almeida, Micheli Filippi. |
| EPI_ISL_2443542 | Lboratorio Central de Saude Publica do Estado do Parana (LACEN/PR) | Laboratory of Respiratory Viruses and Measles, Oswaldo Cruz Institute, FIOCRUZ | Paola Resende, Luciana Appolinario, Fernando Motta, Anna Carolina Paixao, Ana Carolina Mendonca, Alice Sampaio Rocha, Taina Venas, Elisa Cavalcante Pereira, Renata Serrano Lopes, Irina Riediger, Marilda Siqueira on behalf of the Fiocruz COVID-19 Genomic Surveillance Network |
| EPI_ISL_2443578, EPI_ISL_2443579, EPI_ISL_2443580, EPI_ISL_2443581, EPI_ISL_2443582, EPI_ISL_2443583, EPI_ISL_2443584, EPI_ISL_2443585, EPI_ISL_2443586, EPI_ISL_2443587, EPI_ISL_2443588, EPI_ISL_2443589, EPI_ISL_2443590, EPI_ISL_2443591, EPI_ISL_2443592, EPI_ISL_2443593, EPI_ISL_2443594, EPI_ISL_2443595, EPI_ISL_2443596, EPI_ISL_2443597, EPI_ISL_2443598, EPI_ISL_2443599, EPI_ISL_2443600, EPI_ISL_2443601, EPI_ISL_2443602, EPI_ISL_2443603, EPI_ISL_2443604, EPI_ISL_2443605, EPI_ISL_2443606, EPI_ISL_2443607, EPI_ISL_2443608, EPI_ISL_2443609, EPI_ISL_2443610, EPI_ISL_2443611, EPI_ISL_2443612, EPI_ISL_2443613 | | | |
| see above | Laboratory of Respiratory Viruses and Measles, Oswaldo Cruz Institute, FIOCRUZ | Laboratory of Respiratory Viruses and Measles, Oswaldo Cruz Institute, FIOCRUZ | Paola Resende, Luciana Appolinario, Fernando Motta, Anna Carolina Paixao, Ana Carolina Mendonca, Alice Sampaio Rocha, Taina Venas, Elisa Cavalcante Pereira, Renata Serrano Lopes, Marilda Siqueira on behalf of the Fiocruz COVID-19 Genomic Surveillance Network |
| EPI_ISL_2443614, EPI_ISL_2443615, EPI_ISL_2443616, EPI_ISL_2443617, EPI_ISL_2443618, EPI_ISL_2443619, EPI_ISL_2443620, EPI_ISL_2443621, EPI_ISL_2443622, EPI_ISL_2443623, EPI_ISL_2443624, EPI_ISL_2443625, EPI_ISL_2443626, EPI_ISL_2443627, EPI_ISL_2443628, EPI_ISL_2443629, EPI_ISL_2443630, EPI_ISL_2443631, EPI_ISL_2443632 | | | |

| | | | |
|---|---|---|---|
| see above | Labortorio Central de Saude Publica do Estado do Rio de Janeiro (LACEN/RJ) | Laboratory of Respiratory Viruses and Measles, Oswaldo Cruz Institute, FIOCRUZ | Paola Resende, Luciana Appolinario, Fernando Motta, Anna Carolina Paixao, Ana Carolina Mendonca, Alice Sampaio Rocha, Taina Venas, Elisa Cavalcante Pereira, Renata Serrano Lopes, Andrea Cony Cavalcanti, Marilda Siqueira on behalf of the Fiocruz COVID-19 Genomic Surveillance Network |
| EPI_ISL_2443633, EPI_ISL_2443634, EPI_ISL_2443635, EPI_ISL_2443636 | Laboratory of Respiratory Viruses and Measles, Oswaldo Cruz Institute, FIOCRUZ | Laboratory of Respiratory Viruses and Measles, Oswaldo Cruz Institute, FIOCRUZ | Paola Resende, Luciana Appolinario, Fernando Motta, Anna Carolina Paixao, Ana Carolina Mendonca, Alice Sampaio Rocha, Taina Venas, Elisa Cavalcante Pereira, Renata Serrano Lopes, Marilda Siqueira on behalf of the Fiocruz COVID-19 Genomic Surveillance Network |
| EPI_ISL_2443637, EPI_ISL_2443638, EPI_ISL_2443639, EPI_ISL_2443640, EPI_ISL_2443641, EPI_ISL_2443642, EPI_ISL_2443643, EPI_ISL_2443644, EPI_ISL_2443645, EPI_ISL_2443646, EPI_ISL_2443647, EPI_ISL_2443648, EPI_ISL_2443649, EPI_ISL_2443650, EPI_ISL_2443651, EPI_ISL_2443652, EPI_ISL_2443653, EPI_ISL_2443654, EPI_ISL_2443655, EPI_ISL_2443656, EPI_ISL_2443657, EPI_ISL_2443658 | | | |
| see above | Labortorio Central de Saude Publica do Estado do Rio de Janeiro (LACEN/RJ) | Laboratory of Respiratory Viruses and Measles, Oswaldo Cruz Institute, FIOCRUZ | Paola Resende, Luciana Appolinario, Fernando Motta, Anna Carolina Paixao, Ana Carolina Mendonca, Alice Sampaio Rocha, Taina Venas, Elisa Cavalcante Pereira, Renata Serrano Lopes, Andrea Cony Cavalcanti, Marilda Siqueira on behalf of the Fiocruz COVID-19 Genomic Surveillance Network |
| EPI_ISL_2443659, EPI_ISL_2443660, EPI_ISL_2443661, EPI_ISL_2443662, EPI_ISL_2443663, EPI_ISL_2443664, EPI_ISL_2443665, EPI_ISL_2443666, EPI_ISL_2443667, EPI_ISL_2443668, EPI_ISL_2443669, EPI_ISL_2443670, EPI_ISL_2443671 | | | |
| see above | Laboratory of Respiratory Viruses and Measles, Oswaldo Cruz Institute, FIOCRUZ | Laboratory of Respiratory Viruses and Measles, Oswaldo Cruz Institute, FIOCRUZ | Paola Resende, Luciana Appolinario, Fernando Motta, Anna Carolina Paixao, Ana Carolina Mendonca, Alice Sampaio Rocha, Taina Venas, Elisa Cavalcante Pereira, Renata Serrano Lopes, Marilda Siqueira on behalf of the Fiocruz COVID-19 Genomic Surveillance Network |
| EPI_ISL_2443675, EPI_ISL_2443712 | Labortorio Central de Saude Publica do Estado de Santa Catarina (LACEN/SC) | Laboratory of Respiratory Viruses and Measles, Oswaldo Cruz Institute, FIOCRUZ | Paola Resende, Luciana Appolinario, Fernando Motta, Anna Carolina Paixao, Ana Carolina Mendonca, Alice Sampaio Rocha, Taina Venas, Elisa Cavalcante Pereira, Renata Serrano Lopes, Darcita Buerger Rovaris, Sandra Bianchini Fernandes, Marilda Siqueira on behalf of the Fiocruz COVID-19 Genomic Surveillance Network |
| EPI_ISL_2444817 | IICS-UNA | IICS-UNA | Magaly Martinez, Adriana Valenzuela, Alejandra Rojas, Chyntia Diaz, Eva Nara, Fatima Cardozo, Florencia del Puerto, Joel Ortiz, Jonas Fernandez, Laura Franco, Laura Mendoza, Leticia Rojas, Maria Eugenia Galeano. |
| EPI_ISL_2445168 | UNIDADE REFERENCIAL SUDOESTE | Instituto Butantan | Dimas Tadeu Covas, Antonio Jorge Martins, Claudia Renata dos Santos Barros, David Schlesinger, Debora Botequio Moretti, Elaine Cristina Marqueze, Elaine Vieira Santos, Evandra Strazza Rodrigues, Heidge Fukumasu, Jayme Augusto de Souza-Neto, José Salvatore Leister Patané, Luiz Alcantara, Luiz Lehmann Coutinho, Maria Carolina Elias, Maurício Lacerda Nogueira, Rafael dos Santos Bezerra, Raul Machado Neto, Rejane Maria Tommasini Grotto, Ricardo Haddad, Sandra Coccuzzo Sampaio Vessoni, Simone Kashima, Svetoslav Nanev Slavov, Vincent Louis Viala |
| EPI_ISL_2445275 | UPA VILA SANTA CATARINA | Instituto Butantan | Dimas Tadeu Covas, Antonio Jorge Martins, Claudia Renata dos Santos Barros, David Schlesinger, Debora Botequio Moretti, Elaine Cristina Marqueze, Elaine Vieira Santos, Evandra Strazza Rodrigues, Heidge Fukumasu, Jayme Augusto de Souza-Neto, José Salvatore Leister Patané, Luiz Alcantara, Luiz Lehmann Coutinho, Maria Carolina Elias, Maurício Lacerda Nogueira, Rafael dos Santos Bezerra, Raul Machado Neto, Rejane Maria Tommasini Grotto, Ricardo Haddad, Sandra Coccuzzo Sampaio Vessoni, Simone Kashima, Svetoslav Nanev Slavov, Vincent Louis Viala |
| EPI_ISL_2445406 | CS II DR MIGUEL VITALIANO ORLANDIA | Instituto Butantan | Dimas Tadeu Covas, Antonio Jorge Martins, Claudia Renata dos Santos Barros, David Schlesinger, Debora Botequio Moretti, Elaine Cristina Marqueze, Elaine Vieira Santos, Evandra Strazza Rodrigues, Heidge Fukumasu, Jayme Augusto de Souza-Neto, José Salvatore Leister Patané, Luiz Alcantara, Luiz Lehmann Coutinho, Maria Carolina Elias, Maurício Lacerda Nogueira, Rafael dos Santos Bezerra, Raul Machado Neto, Rejane Maria Tommasini Grotto, Ricardo Haddad, Sandra Coccuzzo Sampaio Vessoni, Simone Kashima, Svetoslav Nanev Slavov, Vincent Louis Viala |
| EPI_ISL_2445499 | UNIDADE DE SAUDE DR PHEBO DE OLIVEIRA ROGE FERREIRA | Instituto Butantan | Dimas Tadeu Covas, Antonio Jorge Martins, Claudia Renata dos Santos Barros, David Schlesinger, Debora Botequio Moretti, Elaine Cristina Marqueze, Elaine Vieira Santos, Evandra Strazza Rodrigues, Heidge Fukumasu, Jayme Augusto de Souza-Neto, José Salvatore Leister Patané, Luiz Alcantara, Luiz Lehmann Coutinho, Maria Carolina Elias, Maurício Lacerda Nogueira, Rafael dos Santos Bezerra, Raul Machado Neto, Rejane Maria Tommasini Grotto, Ricardo Haddad, Sandra Coccuzzo Sampaio Vessoni, Simone Kashima, Svetoslav Nanev Slavov, Vincent Louis Viala |
| EPI_ISL_2445564, EPI_ISL_2445570 | IRMANDADE DA SANTA CASA DE MISERICORDIA LORENA | Instituto Butantan | Dimas Tadeu Covas, Antonio Jorge Martins, Claudia Renata dos Santos Barros, David Schlesinger, Debora Botequio Moretti, Elaine Cristina Marqueze, Elaine Vieira Santos, Evandra Strazza Rodrigues, Heidge Fukumasu, Jayme Augusto de Souza-Neto, José Salvatore Leister Patané, Luiz Alcantara, Luiz Lehmann Coutinho, Maria Carolina Elias, Maurício Lacerda Nogueira, Rafael dos Santos Bezerra, Raul Machado Neto, Rejane Maria Tommasini Grotto, Ricardo Haddad, Sandra Coccuzzo Sampaio Vessoni, Simone Kashima, Svetoslav Nanev Slavov, Vincent Louis Viala |
| EPI_ISL_2445581 | HOSPITAL DE CAMPANHA COVID 19 MUNICIPIO DE TAUBATE | Instituto Butantan | Dimas Tadeu Covas, Antonio Jorge Martins, Claudia Renata dos Santos Barros, David Schlesinger, Debora Botequio Moretti, Elaine Cristina Marqueze, Elaine Vieira Santos, Evandra Strazza Rodrigues, Heidge Fukumasu, Jayme Augusto de Souza-Neto, José Salvatore Leister Patané, Luiz Alcantara, Luiz Lehmann Coutinho, Maria Carolina Elias, Maurício Lacerda Nogueira, Rafael dos Santos Bezerra, Raul Machado Neto, Rejane Maria Tommasini Grotto, Ricardo Haddad, Sandra Coccuzzo Sampaio Vessoni, Simone Kashima, Svetoslav Nanev Slavov, Vincent Louis Viala |
| EPI_ISL_2466122, EPI_ISL_2466123, EPI_ISL_2466124 | Laboratory of Respiratory Viruses and Measles, Oswaldo Cruz Institute, FIOCRUZ | Laboratory of Respiratory Viruses and Measles, Oswaldo Cruz Institute, FIOCRUZ | Paola Resende, Luciana Appolinario, Fernando Motta, Anna Carolina Paixao, Ana Carolina Mendonca, Alice Sampaio Rocha, Taina Venas, Elisa Cavalcante Pereira, Renata Serrano Lopes, Marilda Siqueira on behalf of the Fiocruz COVID-19 Genomic Surveillance Network |
| EPI_ISL_2466230 | Labortorio Central de Saude Publica do Estado do Rio de Janeiro (LACEN/RJ) | Laboratory of Respiratory Viruses and Measles, Oswaldo Cruz Institute, FIOCRUZ | Paola Resende, Luciana Appolinario, Fernando Motta, Anna Carolina Paixao, Ana Carolina Mendonca, Alice Sampaio Rocha, Taina Venas, Elisa Cavalcante Pereira, Renata Serrano Lopes, Andrea Cony Cavalcanti, Marilda Siqueira on behalf of the Fiocruz COVID-19 Genomic Surveillance Network |
| EPI_ISL_2466257 | Laboratorio Central de Saude Publica do Estado de Alagoas (LACEN/AL) | Laboratory of Respiratory Viruses and Measles, Oswaldo Cruz Institute, FIOCRUZ | Paola Resende, Luciana Appolinario, Fernando Motta, Anna Carolina Paixao, Ana Carolina Mendonca, Alice Sampaio Rocha, Taina Venas, Elisa Cavalcante Pereira, Renata Serrano Lopes, Anderson Brandao Leite, Marilda Siqueira on behalf of the Fiocruz COVID-19 Genomic Surveillance Network |
| EPI_ISL_2466264 | Laboratorio Central de Saude Publica do Estado da Bahia (LACEN/BA) | Laboratory of Respiratory Viruses and Measles, Oswaldo Cruz Institute, FIOCRUZ | Paola Resende, Luciana Appolinario, Fernando Motta, Anna Carolina Paixao, Ana Carolina Mendonca, Alice Sampaio Rocha, Taina Venas, Elisa Cavalcante Pereira, Renata Serrano Lopes,Felicidade Pereira, Marilda Siqueira on behalf of the Fiocruz COVID-19 Genomic Surveillance Network |
| EPI_ISL_2466268 | Laboratorio Central de Saude Publica do Estado do Rio de Janeiro (LACEN-RJ) | Laboratory of Respiratory Viruses and Measles, Oswaldo Cruz Institute, FIOCRUZ | Paola Resende, Luciana Appolinario, Fernando Motta, Anna Carolina Paixao, Ana Carolina Mendonca, Alice Sampaio Rocha, Taina Venas, Elisa Cavalcante Pereira, Renata Serrano Lopes, Andrea Cony Cavalcanti, Marilda Siqueira on behalf of the Fiocruz COVID-19 Genomic Surveillance Network |
| EPI_ISL_2466447, EPI_ISL_2466459 | HLAGYN - Laboratorio de Imunologia de Transplantes de Goias | HLAGYN - Laboratorio de Imunologia de Transplantes de Goias | Fernando Antonio Vinhal dos Santos, Erika Lopes Rocha Batista, Alessandro Leonardo Alvares Magalhaes, Frederico Rodrigues Vinhal, Sabrina Sara Moreira Duarte, Lucas Carlos Gomes Pereira, Daniel Ferreira de Sousa |
| EPI_ISL_2473714 | UBS PASTOR MARCELINO DEUNGARO RIO GRANDE | Instituto Butantan | Dimas Tadeu Covas, Antonio Jorge Martins, Claudia Renata dos Santos Barros, David Schlesinger, Debora Botequio Moretti, Elaine Cristina Marqueze, Elaine Vieira Santos, Evandra Strazza Rodrigues, Heidge Fukumasu, Jayme Augusto de Souza-Neto, José Salvatore Leister Patané, Luiz Alcantara, Luiz Lehmann Coutinho, Maria Carolina Elias, Maurício Lacerda Nogueira, Rafael dos Santos Bezerra, Raul Machado Neto, Rejane Maria Tommasini Grotto, Ricardo Haddad, Sandra Coccuzzo Sampaio Vessoni, Simone Kashima, Svetoslav Nanev Slavov, Vincent Louis Viala |
| EPI_ISL_2473734 | HOSP GERAL DE SAO PAULO | Instituto Butantan | Dimas Tadeu Covas, Antonio Jorge Martins, Claudia Renata dos Santos Barros, David Schlesinger, Debora Botequio Moretti, Elaine Cristina Marqueze, Elaine Vieira Santos, Evandra Strazza Rodrigues, Heidge Fukumasu, Jayme Augusto de Souza-Neto, José Salvatore Leister Patané, Luiz Alcantara, Luiz Lehmann Coutinho, Maria Carolina Elias, Maurício Lacerda Nogueira, Rafael dos Santos Bezerra, Raul Machado Neto, Rejane Maria Tommasini Grotto, Ricardo Haddad, Sandra Coccuzzo Sampaio Vessoni, Simone Kashima, Svetoslav Nanev Slavov, Vincent Louis Viala |
| EPI_ISL_2473786 | CADIP VILA REGINA FERNANDOPOLIS | Instituto Butantan | Dimas Tadeu Covas, Antonio Jorge Martins, Claudia Renata dos Santos Barros, David Schlesinger, Debora Botequio Moretti, Elaine Cristina Marqueze, Elaine Vieira Santos, Evandra Strazza Rodrigues, Heidge Fukumasu, Jayme Augusto de Souza-Neto, José Salvatore Leister Patané, Luiz Alcantara, Luiz Lehmann Coutinho, Maria Carolina Elias, Maurício Lacerda Nogueira, Rafael dos Santos Bezerra, Raul Machado Neto, Rejane Maria Tommasini Grotto, Ricardo Haddad, Sandra Coccuzzo Sampaio Vessoni, Simone Kashima, Svetoslav Nanev Slavov, Vincent Louis Viala |
| EPI_ISL_2493388 | SECRETARIA DE SAUDE DE SAO PEDRO | Instituto Butantan | Dimas Tadeu Covas, Antonio Jorge Martins, Claudia Renata dos Santos Barros, David Schlesinger, Debora Botequio Moretti, Elaine Cristina Marqueze, Elaine Vieira Santos, Evandra Strazza Rodrigues, Heidge Fukumasu, Jayme Augusto de Souza-Neto, José Salvatore Leister Patané, Luiz Alcantara, Luiz Lehmann Coutinho, Maria Carolina Elias, Maurício Lacerda Nogueira, Rafael dos Santos Bezerra, Raul Machado Neto, Rejane Maria Tommasini Grotto, Ricardo Haddad, Sandra Coccuzzo Sampaio Vessoni, Simone Kashima, Svetoslav Nanev Slavov, Vincent Louis Viala |
| EPI_ISL_2493750 | AFIP | Instituto Butantan | Dimas Tadeu Covas, Antonio Jorge Martins, Claudia Renata dos Santos Barros, David Schlesinger, Debora Botequio Moretti, Elaine Cristina Marqueze, Elaine Vieira Santos, Evandra Strazza Rodrigues, Heidge Fukumasu, Jayme Augusto de Souza-Neto, José Salvatore Leister Patané, Luiz Alcantara, Luiz Lehmann Coutinho, Maria Carolina Elias, Maurício Lacerda Nogueira, Rafael dos Santos Bezerra, Raul Machado Neto, Rejane Maria Tommasini Grotto, Ricardo Haddad, Sandra Coccuzzo Sampaio Vessoni, Simone Kashima, Svetoslav Nanev Slavov, Vincent Louis Viala |
| EPI_ISL_2493820 | UBS JARDIM CAIUBY | Instituto Butantan | Dimas Tadeu Covas, Antonio Jorge Martins, Claudia Renata dos Santos Barros, David Schlesinger, Debora Botequio Moretti, Elaine Cristina Marqueze, Elaine Vieira Santos, Evandra Strazza Rodrigues, Heidge Fukumasu, Jayme Augusto de Souza-Neto, José Salvatore Leister Patané, Luiz Alcantara, Luiz Lehmann Coutinho, Maria Carolina Elias, Maurício Lacerda Nogueira, Rafael dos Santos Bezerra, Raul Machado Neto, Rejane Maria Tommasini Grotto, |

| | | | |
|---|---|---|---|
| | | | Ricardo Haddad, Sandra Coccuzzo Sampaio Vessoni, Simone Kashima, Svetoslav Nanev Slavov, Vincent Louis Viala |
| EPI_ISL_2493831 | USAFA SAO JORGE | Instituto Butantan | Dimas Tadeu Covas, Antonio Jorge Martins, Claudia Renata dos Santos Barros, David Schlesinger, Debora Botequio Moretti, Elaine Cristina Marqueze, Elaine Vieira Santos, Evandra Strazza Rodrigues, Heidge Fukumasu, Jayme Augusto de Souza-Neto, José Salvatore Leister Patané, Luiz Alcantara, Luiz Lehmann Coutinho, Maria Carolina Elias, Maurício Lacerda Nogueira, Rafael dos Santos Bezerra, Raul Machado Neto, Rejane Maria Tommasini Grotto, Ricardo Haddad, Sandra Coccuzzo Sampaio Vessoni, Simone Kashima, Svetoslav Nanev Slavov, Vincent Louis Viala |
| EPI_ISL_2493891 | PRONTO ATENDIMENTO DE VARGEM GRANDE PAULISTA | Instituto Butantan | Dimas Tadeu Covas, Antonio Jorge Martins, Claudia Renata dos Santos Barros, David Schlesinger, Debora Botequio Moretti, Elaine Cristina Marqueze, Elaine Vieira Santos, Evandra Strazza Rodrigues, Heidge Fukumasu, Jayme Augusto de Souza-Neto, José Salvatore Leister Patané, Luiz Alcantara, Luiz Lehmann Coutinho, Maria Carolina Elias, Maurício Lacerda Nogueira, Rafael dos Santos Bezerra, Raul Machado Neto, Rejane Maria Tommasini Grotto, Ricardo Haddad, Sandra Coccuzzo Sampaio Vessoni, Simone Kashima, Svetoslav Nanev Slavov, Vincent Louis Viala |
| EPI_ISL_2494003 | INSIDE CRSN PIRITUBA | Instituto Butantan | Dimas Tadeu Covas, Antonio Jorge Martins, Claudia Renata dos Santos Barros, David Schlesinger, Debora Botequio Moretti, Elaine Cristina Marqueze, Elaine Vieira Santos, Evandra Strazza Rodrigues, Heidge Fukumasu, Jayme Augusto de Souza-Neto, José Salvatore Leister Patané, Luiz Alcantara, Luiz Lehmann Coutinho, Maria Carolina Elias, Maurício Lacerda Nogueira, Rafael dos Santos Bezerra, Raul Machado Neto, Rejane Maria Tommasini Grotto, Ricardo Haddad, Sandra Coccuzzo Sampaio Vessoni, Simone Kashima, Svetoslav Nanev Slavov, Vincent Louis Viala |
| EPI_ISL_2494025 | INSIDE CRSSU IPIRANGA | Instituto Butantan | Dimas Tadeu Covas, Antonio Jorge Martins, Claudia Renata dos Santos Barros, David Schlesinger, Debora Botequio Moretti, Elaine Cristina Marqueze, Elaine Vieira Santos, Evandra Strazza Rodrigues, Heidge Fukumasu, Jayme Augusto de Souza-Neto, José Salvatore Leister Patané, Luiz Alcantara, Luiz Lehmann Coutinho, Maria Carolina Elias, Maurício Lacerda Nogueira, Rafael dos Santos Bezerra, Raul Machado Neto, Rejane Maria Tommasini Grotto, Ricardo Haddad, Sandra Coccuzzo Sampaio Vessoni, Simone Kashima, Svetoslav Nanev Slavov, Vincent Louis Viala |
| EPI_ISL_2494091 | AFIP SUL | Instituto Butantan | Dimas Tadeu Covas, Antonio Jorge Martins, Claudia Renata dos Santos Barros, David Schlesinger, Debora Botequio Moretti, Elaine Cristina Marqueze, Elaine Vieira Santos, Evandra Strazza Rodrigues, Heidge Fukumasu, Jayme Augusto de Souza-Neto, José Salvatore Leister Patané, Luiz Alcantara, Luiz Lehmann Coutinho, Maria Carolina Elias, Maurício Lacerda Nogueira, Rafael dos Santos Bezerra, Raul Machado Neto, Rejane Maria Tommasini Grotto, Ricardo Haddad, Sandra Coccuzzo Sampaio Vessoni, Simone Kashima, Svetoslav Nanev Slavov, Vincent Louis Viala |
| EPI_ISL_2494197 | VIGILANCIA EPIDEMIOLOGICA E CONTROLE DE VETORES PIRASSUNUN | Instituto Butantan | Dimas Tadeu Covas, Antonio Jorge Martins, Claudia Renata dos Santos Barros, David Schlesinger, Debora Botequio Moretti, Elaine Cristina Marqueze, Elaine Vieira Santos, Evandra Strazza Rodrigues, Heidge Fukumasu, Jayme Augusto de Souza-Neto, José Salvatore Leister Patané, Luiz Alcantara, Luiz Lehmann Coutinho, Maria Carolina Elias, Maurício Lacerda Nogueira, Rafael dos Santos Bezerra, Raul Machado Neto, Rejane Maria Tommasini Grotto, Ricardo Haddad, Sandra Coccuzzo Sampaio Vessoni, Simone Kashima, Svetoslav Nanev Slavov, Vincent Louis Viala |
| EPI_ISL_2494225 | BIOFAST LESTE | Instituto Butantan | Dimas Tadeu Covas, Antonio Jorge Martins, Claudia Renata dos Santos Barros, David Schlesinger, Debora Botequio Moretti, Elaine Cristina Marqueze, Elaine Vieira Santos, Evandra Strazza Rodrigues, Heidge Fukumasu, Jayme Augusto de Souza-Neto, José Salvatore Leister Patané, Luiz Alcantara, Luiz Lehmann Coutinho, Maria Carolina Elias, Maurício Lacerda Nogueira, Rafael dos Santos Bezerra, Raul Machado Neto, Rejane Maria Tommasini Grotto, Ricardo Haddad, Sandra Coccuzzo Sampaio Vessoni, Simone Kashima, Svetoslav Nanev Slavov, Vincent Louis Viala |
| EPI_ISL_2494340 | HOSPITAL E MATERNIDADE NOSSA SENHORA DA AJUDA | Instituto Butantan | Dimas Tadeu Covas, Antonio Jorge Martins, Claudia Renata dos Santos Barros, David Schlesinger, Debora Botequio Moretti, Elaine Cristina Marqueze, Elaine Vieira Santos, Evandra Strazza Rodrigues, Heidge Fukumasu, Jayme Augusto de Souza-Neto, José Salvatore Leister Patané, Luiz Alcantara, Luiz Lehmann Coutinho, Maria Carolina Elias, Maurício Lacerda Nogueira, Rafael dos Santos Bezerra, Raul Machado Neto, Rejane Maria Tommasini Grotto, Ricardo Haddad, Sandra Coccuzzo Sampaio Vessoni, Simone Kashima, Svetoslav Nanev Slavov, Vincent Louis Viala |
| EPI_ISL_2494341 | FACULDADE DE MEDICINA ABC | Instituto Butantan | Dimas Tadeu Covas, Antonio Jorge Martins, Claudia Renata dos Santos Barros, David Schlesinger, Debora Botequio Moretti, Elaine Cristina Marqueze, Elaine Vieira Santos, Evandra Strazza Rodrigues, Heidge Fukumasu, Jayme Augusto de Souza-Neto, José Salvatore Leister Patané, Luiz Alcantara, Luiz Lehmann Coutinho, Maria Carolina Elias, Maurício Lacerda Nogueira, Rafael dos Santos Bezerra, Raul Machado Neto, Rejane Maria Tommasini Grotto, Ricardo Haddad, Sandra Coccuzzo Sampaio Vessoni, Simone Kashima, Svetoslav Nanev Slavov, Vincent Louis Viala |
| EPI_ISL_2497447, EPI_ISL_2497463 | HLAGYN - Laboratorio de Imunologia de Transplantes de Goias | HLAGYN - Laboratorio de Imunologia de Transplantes de Goias | Fernando Antonio Vinhal dos Santos, Erika Lopes Rocha Batista, Alessandro Leonardo Alvares Magalhaes, Frederico Rodrigues Vinhal, Sabrina Sara Moreira Duarte, Lucas Carlos Gomes Pereira, Daniel Ferreira de Sousa |
| EPI_ISL_2534862, EPI_ISL_2534863, EPI_ISL_2534864, EPI_ISL_2534865, EPI_ISL_2534866, EPI_ISL_2534867, EPI_ISL_2534868, EPI_ISL_2534869, EPI_ISL_2534870, EPI_ISL_2534871, EPI_ISL_2534872, EPI_ISL_2534873, EPI_ISL_2534874, EPI_ISL_2534875, EPI_ISL_2534876, EPI_ISL_2534877, EPI_ISL_2534878, EPI_ISL_2534879, EPI_ISL_2534880, EPI_ISL_2534881, EPI_ISL_2534882, EPI_ISL_2534883, EPI_ISL_2534884, EPI_ISL_2534885, EPI_ISL_2534886, EPI_ISL_2534887, EPI_ISL_2534888, EPI_ISL_2534889, EPI_ISL_2534890, EPI_ISL_2534891, EPI_ISL_2534892, EPI_ISL_2534893, EPI_ISL_2534894, EPI_ISL_2534895, EPI_ISL_2534896, EPI_ISL_2534897, EPI_ISL_2534898, EPI_ISL_2534899, EPI_ISL_2534900, EPI_ISL_2534901, EPI_ISL_2534902, EPI_ISL_2534903, EPI_ISL_2534904, EPI_ISL_2534905, EPI_ISL_2534906, EPI_ISL_2534907, EPI_ISL_2534908, EPI_ISL_2534909, EPI_ISL_2534910, EPI_ISL_2534911, EPI_ISL_2534912, EPI_ISL_2534913, EPI_ISL_2534914, EPI_ISL_2534915, EPI_ISL_2534916, EPI_ISL_2534917, EPI_ISL_2534918, EPI_ISL_2534919, EPI_ISL_2534920, EPI_ISL_2534921, EPI_ISL_2534922, EPI_ISL_2534924, EPI_ISL_2534925, EPI_ISL_2534926, EPI_ISL_2534927, EPI_ISL_2534928, EPI_ISL_2534929, EPI_ISL_2534930, EPI_ISL_2534931, EPI_ISL_2534933, EPI_ISL_2534934, EPI_ISL_2534935, EPI_ISL_2534936, EPI_ISL_2534937, EPI_ISL_2534938, EPI_ISL_2534939, EPI_ISL_2534940, EPI_ISL_2534941, EPI_ISL_2534942, EPI_ISL_2534943, EPI_ISL_2534944, EPI_ISL_2534946, EPI_ISL_2534947, EPI_ISL_2534948, EPI_ISL_2534949, EPI_ISL_2534950, EPI_ISL_2534951, EPI_ISL_2534952, EPI_ISL_2534955, EPI_ISL_2534956, EPI_ISL_2534957, EPI_ISL_2534958, EPI_ISL_2534959, EPI_ISL_2534960, EPI_ISL_2534961, EPI_ISL_2534962, EPI_ISL_2534963, EPI_ISL_2534964, EPI_ISL_2534965, EPI_ISL_2534966, EPI_ISL_2534967, EPI_ISL_2534968, EPI_ISL_2534969, EPI_ISL_2534970, EPI_ISL_2534971, EPI_ISL_2534972, EPI_ISL_2534973, EPI_ISL_2534974, EPI_ISL_2534975, EPI_ISL_2534976, EPI_ISL_2534977, EPI_ISL_2534978, EPI_ISL_2534979, EPI_ISL_2534980, EPI_ISL_2534981, EPI_ISL_2534982, EPI_ISL_2534983, EPI_ISL_2534984, EPI_ISL_2534985, EPI_ISL_2534986, EPI_ISL_2534987, EPI_ISL_2534988, EPI_ISL_2534989, EPI_ISL_2534990, EPI_ISL_2534991, EPI_ISL_2534992, EPI_ISL_2534993, EPI_ISL_2534994, EPI_ISL_2534996, EPI_ISL_2534997, EPI_ISL_2534999, EPI_ISL_2535000, EPI_ISL_2535001, EPI_ISL_2535002, EPI_ISL_2535003, EPI_ISL_2535004, EPI_ISL_2535005, EPI_ISL_2535006, EPI_ISL_2535007, EPI_ISL_2535009, EPI_ISL_2535010, EPI_ISL_2535011, EPI_ISL_2535012, EPI_ISL_2535013, EPI_ISL_2535014, EPI_ISL_2535015, EPI_ISL_2535016, EPI_ISL_2535018, EPI_ISL_2535019, EPI_ISL_2535021, EPI_ISL_2535022, EPI_ISL_2535023, EPI_ISL_2535024, EPI_ISL_2535025, EPI_ISL_2535026, EPI_ISL_2535027, EPI_ISL_2535028, EPI_ISL_2535029, EPI_ISL_2535030, EPI_ISL_2535031, EPI_ISL_2535032, EPI_ISL_2535033, EPI_ISL_2535034, EPI_ISL_2535035, EPI_ISL_2535036, EPI_ISL_2535037, EPI_ISL_2535038, EPI_ISL_2535039, EPI_ISL_2535040, EPI_ISL_2535041, EPI_ISL_2535043, EPI_ISL_2535044, EPI_ISL_2535045, EPI_ISL_2535046, EPI_ISL_2535047, EPI_ISL_2535048, EPI_ISL_2535049, EPI_ISL_2535050, EPI_ISL_2535051, EPI_ISL_2535052, EPI_ISL_2535053, EPI_ISL_2535054, EPI_ISL_2535055, EPI_ISL_2535056, EPI_ISL_2535057, EPI_ISL_2535058, EPI_ISL_2535059, EPI_ISL_2535060, EPI_ISL_2535061, EPI_ISL_2535062, EPI_ISL_2535063, EPI_ISL_2535064, EPI_ISL_2535065, EPI_ISL_2535066, EPI_ISL_2535067, EPI_ISL_2535068, EPI_ISL_2535069, EPI_ISL_2535070, EPI_ISL_2535071, EPI_ISL_2535072, EPI_ISL_2535073, EPI_ISL_2535074, EPI_ISL_2535075, EPI_ISL_2535076, EPI_ISL_2535077, EPI_ISL_2535078, EPI_ISL_2535080, EPI_ISL_2535081, EPI_ISL_2535082, EPI_ISL_2535083, EPI_ISL_2535084, EPI_ISL_2535085, EPI_ISL_2535086, EPI_ISL_2535087, EPI_ISL_2535088, EPI_ISL_2535089, EPI_ISL_2535090, EPI_ISL_2535091, EPI_ISL_2535092, EPI_ISL_2535093, EPI_ISL_2535094, EPI_ISL_2535095, EPI_ISL_2535096, EPI_ISL_2535097, EPI_ISL_2535098, EPI_ISL_2535099, EPI_ISL_2535100, EPI_ISL_2535101, EPI_ISL_2535102, EPI_ISL_2535103, EPI_ISL_2535104, EPI_ISL_2535105, EPI_ISL_2535106, EPI_ISL_2535107, EPI_ISL_2535108, EPI_ISL_2535109, EPI_ISL_2535110 | | | |
| see above | Laboratorio Central Noel Nutels | Bioinformatics Laboratory / LNCC | Luiz G P de Almeida, Alessandra P Lamarca, Ronaldo da Silva F Jr, Liliane Cavalcante, Alexandra L Gerber, Ana Paula de C Guimaraes, Douglas Terra Machado, Cassia Alves, Diana Mariani, Cintia Policarpo, Gleidson da Silva de Oliveira, Mario Sergio Ribeiro, Silvia Carvalho, Flavio Dias da Silva, Marcio Henrique de Oliveira Garcia, Leandro Magalhaes de Souza, Cristiane Gomes da Silva, Caio Luiz Pereira Ribeiro, Andrea Cony Cavalcanti, Claudia Maria Braga de Mello, Amilcar Tanuri, Ana Tereza R Vasconcelos |
| EPI_ISL_2535111, EPI_ISL_2535112, EPI_ISL_2535113, EPI_ISL_2535114, EPI_ISL_2535115, EPI_ISL_2535117, EPI_ISL_2535118, EPI_ISL_2535119, EPI_ISL_2535120, EPI_ISL_2535121, EPI_ISL_2535123, EPI_ISL_2535125, EPI_ISL_2535126, EPI_ISL_2535127, EPI_ISL_2535128, EPI_ISL_2535131, EPI_ISL_2535132, EPI_ISL_2535133, EPI_ISL_2535134, EPI_ISL_2535135, EPI_ISL_2535136, EPI_ISL_2535137, EPI_ISL_2535138, EPI_ISL_2535140, EPI_ISL_2535141, EPI_ISL_2535142, EPI_ISL_2535143, EPI_ISL_2535144, EPI_ISL_2535145, EPI_ISL_2535146, EPI_ISL_2535147, EPI_ISL_2535148, EPI_ISL_2535149, EPI_ISL_2535151, EPI_ISL_2535152, EPI_ISL_2535153, EPI_ISL_2535154, EPI_ISL_2535155, EPI_ISL_2535156, EPI_ISL_2535157, EPI_ISL_2535158, EPI_ISL_2535160, EPI_ISL_2535161, EPI_ISL_2535162, EPI_ISL_2535163, EPI_ISL_2535164, EPI_ISL_2535165, EPI_ISL_2535166, EPI_ISL_2535167, EPI_ISL_2535168, EPI_ISL_2535169, EPI_ISL_2535170, EPI_ISL_2535171, EPI_ISL_2535172, EPI_ISL_2535173, EPI_ISL_2535174, EPI_ISL_2535175, EPI_ISL_2535176, EPI_ISL_2535177, EPI_ISL_2535178, EPI_ISL_2535179, EPI_ISL_2535180, EPI_ISL_2535181, EPI_ISL_2535182, EPI_ISL_2535184, EPI_ISL_2535185, EPI_ISL_2535187, EPI_ISL_2535188, EPI_ISL_2535189, EPI_ISL_2535190, EPI_ISL_2535191, EPI_ISL_2535192, EPI_ISL_2535193, EPI_ISL_2535194, EPI_ISL_2535195, EPI_ISL_2535196, EPI_ISL_2535197, EPI_ISL_2535198, EPI_ISL_2535199, EPI_ISL_2535200, EPI_ISL_2535201, EPI_ISL_2535202, EPI_ISL_2535203, EPI_ISL_2535204, EPI_ISL_2535205, EPI_ISL_2535206, EPI_ISL_2535207, EPI_ISL_2535208, EPI_ISL_2535209, EPI_ISL_2535210, EPI_ISL_2535211, EPI_ISL_2535212, EPI_ISL_2535213, EPI_ISL_2535214, EPI_ISL_2535216, EPI_ISL_2535217, EPI_ISL_2535218, EPI_ISL_2535219, EPI_ISL_2535220, EPI_ISL_2535221, EPI_ISL_2535222, EPI_ISL_2535224, EPI_ISL_2535225, EPI_ISL_2535226, EPI_ISL_2535227, EPI_ISL_2535228, EPI_ISL_2535229, EPI_ISL_2535230, EPI_ISL_2535231, EPI_ISL_2535232, EPI_ISL_2535233, EPI_ISL_2535234, EPI_ISL_2535235 | | | |
| see above | Unidade de apoio ao diagnostico da COVID - UNADIG | Bioinformatics Laboratory / LNCC | Luiz G P de Almeida, Alessandra P Lamarca, Ronaldo da Silva F Jr, Liliane Cavalcante, Alexandra L Gerber, Ana Paula de C Guimaraes, Douglas Terra Machado, Cassia Alves, Diana Mariani, Cintia Policarpo, Gleidson da Silva de Oliveira, Mario Sergio Ribeiro, Silvia Carvalho, Flavio Dias da Silva, Marcio Henrique de Oliveira Garcia, Leandro Magalhaes de Souza, Cristiane Gomes da Silva, Caio Luiz Pereira Ribeiro, Andrea Cony Cavalcanti, Claudia Maria Braga de Mello, Amilcar Tanuri, Ana Tereza R Vasconcelos |
| EPI_ISL_2536228 | Labortorio Central de Saude Publica do Estado de Santa Catarina (LACEN/SC) | Laboratory of Respiratory Viruses and Measles, Oswaldo Cruz Institute, FIOCRUZ | Paola Resende, Luciana Appolinario, Fernando Motta, Anna Carolina Paixao, Ana Carolina Mendonca, Alice Sampaio Rocha, Taina Venas, Elisa Cavalcante Pereira, Renata Serrano Lopes, Darcita Buerger Rovaris, Sandra Bianchini Fernandes, Marilda Siqueira on behalf of the Fiocruz COVID-19 Genomic Surveillance Network |
| EPI_ISL_2536269, EPI_ISL_2536270, EPI_ISL_2536271, EPI_ISL_2536272, EPI_ISL_2536273, EPI_ISL_2536274 | Laboratory of Respiratory Viruses and Measles, Oswaldo Cruz Institute, FIOCRUZ | Laboratory of Respiratory Viruses and Measles, Oswaldo Cruz Institute, FIOCRUZ | Paola Resende, Patricia Brasil, Luciana Appolinario, Fernando Motta, Anna Carolina Paixao, Ana Carolina Mendonca, Alice Sampaio Rocha, Taina Venas, Elisa Cavalcante Pereira, Renata Serrano Lopes, Marilda Siqueira on behalf of the Fiocruz COVID-19 Genomic Surveillance Network |
| EPI_ISL_2536275, EPI_ISL_2536276, EPI_ISL_2536277, EPI_ISL_2536278, EPI_ISL_2536279, EPI_ISL_2536281, EPI_ISL_2536282, EPI_ISL_2536283, EPI_ISL_2536284, EPI_ISL_2536285, EPI_ISL_2536286, EPI_ISL_2536287, EPI_ISL_2536288, EPI_ISL_2536289, EPI_ISL_2536290, EPI_ISL_2536291, EPI_ISL_2536292, EPI_ISL_2536293, EPI_ISL_2536294, EPI_ISL_2536295, EPI_ISL_2536296, EPI_ISL_2536297, EPI_ISL_2536298, EPI_ISL_2536299, EPI_ISL_2536300, EPI_ISL_2536301, EPI_ISL_2536302 | | | |
| see above | Labortorio Central de Saude Publica do Estado do Rio de Janeiro (LACEN/RJ) | Laboratory of Respiratory Viruses and Measles, Oswaldo Cruz Institute, FIOCRUZ | Paola Resende, Luciana Appolinario, Fernando Motta, Anna Carolina Paixao, Ana Carolina Mendonca, Alice Sampaio Rocha, Taina Venas, Elisa Cavalcante Pereira, Renata Serrano Lopes, Andrea Cony Cavalcanti, Marilda Siqueira on behalf of the Fiocruz COVID-19 Genomic Surveillance Network |

| | | | |
|---|---|---|---|
| EPI_ISL_2543484 | Genetica Molecular and Subdepartamento de Virologia ISP Chile | Instituto de Salud Publica de Chile | Karen Orostica, Constanza Campano, Barbara Parra, Loredana Arata, Gisselle Barra, Patricia Bustos, Rodrigo Fasce, Javier Tognarelli, Andres Castillo, Soledad Ulloa, Jorge Fernandez |
| EPI_ISL_2557376, EPI_ISL_2557377, EPI_ISL_2557378, EPI_ISL_2557379, EPI_ISL_2557380, EPI_ISL_2557381, EPI_ISL_2557382, EPI_ISL_2557383, EPI_ISL_2557384 | Labortorio Central de Saude Publica do Estado do Rio de Janeiro (LACEN/RJ) | Laboratory of Respiratory Viruses and Measles, Oswaldo Cruz Institute, FIOCRUZ | Paola Resende, Luciana Appolinario, Fernando Motta, Anna Carolina Paixao, Ana Carolina Mendonca, Alice Sampaio Rocha, Taina Venas, Elisa Cavalcante Pereira, Renata Serrano Lopes, Andrea Cony Cavalcanti, Marilda Siqueira on behalf of the Fiocruz COVID-19 Genomic Surveillance Network |
| EPI_ISL_2557385, EPI_ISL_2557386, EPI_ISL_2557387, EPI_ISL_2557388, EPI_ISL_2557389, EPI_ISL_2557390, EPI_ISL_2557391, EPI_ISL_2557392, EPI_ISL_2557393, EPI_ISL_2557394, EPI_ISL_2557395, EPI_ISL_2557396, EPI_ISL_2557397, EPI_ISL_2557398, EPI_ISL_2557399, EPI_ISL_2557400, EPI_ISL_2557401, EPI_ISL_2557402, EPI_ISL_2557403 | | | |
| see above | Laboratory of Respiratory Viruses and Measles, Oswaldo Cruz Institute, FIOCRUZ | Laboratory of Respiratory Viruses and Measles, Oswaldo Cruz Institute, FIOCRUZ | Paola Resende, Luciana Appolinario, Fernando Motta, Anna Carolina Paixao, Ana Carolina Mendonca, Alice Sampaio Rocha, Taina Venas, Elisa Cavalcante Pereira, Renata Serrano Lopes, Marilda Siqueira on behalf of the Fiocruz COVID-19 Genomic Surveillance Network |
| EPI_ISL_2557404 | Labortorio Central de Saude Publica do Estado do Rio de Janeiro (LACEN/RJ) | Laboratory of Respiratory Viruses and Measles, Oswaldo Cruz Institute, FIOCRUZ | Paola Resende, Luciana Appolinario, Fernando Motta, Anna Carolina Paixao, Ana Carolina Mendonca, Alice Sampaio Rocha, Taina Venas, Elisa Cavalcante Pereira, Renata Serrano Lopes, Andrea Cony Cavalcanti, Marilda Siqueira on behalf of the Fiocruz COVID-19 Genomic Surveillance Network |
| EPI_ISL_2557405 | Laboratory of Respiratory Viruses and Measles, Oswaldo Cruz Institute, FIOCRUZ | Laboratory of Respiratory Viruses and Measles, Oswaldo Cruz Institute, FIOCRUZ | Paola Resende, Luciana Appolinario, Fernando Motta, Anna Carolina Paixao, Ana Carolina Mendonca, Alice Sampaio Rocha, Taina Venas, Elisa Cavalcante Pereira, Renata Serrano Lopes, Marilda Siqueira on behalf of the Fiocruz COVID-19 Genomic Surveillance Network |
| EPI_ISL_2557406 | Labortorio Central de Saude Publica do Estado do Rio de Janeiro (LACEN/RJ) | Laboratory of Respiratory Viruses and Measles, Oswaldo Cruz Institute, FIOCRUZ | Paola Resende, Luciana Appolinario, Fernando Motta, Anna Carolina Paixao, Ana Carolina Mendonca, Alice Sampaio Rocha, Taina Venas, Elisa Cavalcante Pereira, Renata Serrano Lopes, Andrea Cony Cavalcanti, Marilda Siqueira on behalf of the Fiocruz COVID-19 Genomic Surveillance Network |
| EPI_ISL_2557407, EPI_ISL_2557408, EPI_ISL_2557409, EPI_ISL_2557410 | Laboratory of Respiratory Viruses and Measles, Oswaldo Cruz Institute, FIOCRUZ | Laboratory of Respiratory Viruses and Measles, Oswaldo Cruz Institute, FIOCRUZ | Paola Resende, Luciana Appolinario, Fernando Motta, Anna Carolina Paixao, Ana Carolina Mendonca, Alice Sampaio Rocha, Taina Venas, Elisa Cavalcante Pereira, Renata Serrano Lopes, Marilda Siqueira on behalf of the Fiocruz COVID-19 Genomic Surveillance Network |
| EPI_ISL_2601693 | Hospital General Universitario Gregorio Marañón | Hospital General Universitario Gregorio Marañón | Sergio Buenestado Serrano, Pedro Sola Campoy, Laura Pérez-Lago, Cristina Rodriguez-Grande, Marta Herranz Martin, Victor Manuel de la Cueva, Julia Suárez, Pilar Catalán, Patricia Muñoz, Darío García de Viedma |
| EPI_ISL_2603449 | Laboratorio Central de Saude Publica do Estado do Rio Grande do Sul (LACEN-RS) | Laboratory of Respiratory Viruses and Measles, Oswaldo Cruz Institute, FIOCRUZ | Paola Resende, Luciana Appolinario, Fernando Motta, Anna Carolina Paixao, Ana Carolina Mendonca, Alice Sampaio Rocha, Taina Venas, Elisa Cavalcante Pereira, Renata Serrano Lopes, Anderson Brandao Leite, Marilda Siqueira on behalf of the Fiocruz COVID-19 Genomic Surveillance Network |
| EPI_ISL_2603472, EPI_ISL_2603473, EPI_ISL_2603474, EPI_ISL_2603475, EPI_ISL_2603476, EPI_ISL_2603477, EPI_ISL_2603478, EPI_ISL_2603479, EPI_ISL_2603480, EPI_ISL_2603482, EPI_ISL_2603483, EPI_ISL_2603484, EPI_ISL_2603485, EPI_ISL_2603486, EPI_ISL_2603487, EPI_ISL_2603488, EPI_ISL_2603489, EPI_ISL_2603490, EPI_ISL_2603491, EPI_ISL_2603492, EPI_ISL_2603493, EPI_ISL_2603494, EPI_ISL_2603495, EPI_ISL_2603496, EPI_ISL_2603497, EPI_ISL_2603498, EPI_ISL_2603499, EPI_ISL_2603500, EPI_ISL_2603501, EPI_ISL_2603502, EPI_ISL_2603503, EPI_ISL_2603504, EPI_ISL_2603505, EPI_ISL_2603506, EPI_ISL_2603507, EPI_ISL_2603508, EPI_ISL_2603509, EPI_ISL_2603510, EPI_ISL_2603511, EPI_ISL_2603512, EPI_ISL_2603513, EPI_ISL_2603514, EPI_ISL_2603515, EPI_ISL_2603516, EPI_ISL_2603517, EPI_ISL_2603518, EPI_ISL_2603519, EPI_ISL_2603520 | | | |
| see above | Laboratory of Respiratory Viruses and Measles, Oswaldo Cruz Institute, FIOCRUZ | Laboratory of Respiratory Viruses and Measles, Oswaldo Cruz Institute, FIOCRUZ | Paola Resende, Luciana Appolinario, Fernando Motta, Anna Carolina Paixao, Ana Carolina Mendonca, Alice Sampaio Rocha, Taina Venas, Elisa Cavalcante Pereira, Renata Serrano Lopes, Marilda Siqueira on behalf of the Fiocruz COVID-19 Genomic Surveillance Network |
| EPI_ISL_2603539, EPI_ISL_2603549, EPI_ISL_2603558, EPI_ISL_2603584 | Laboratorio Central de Saude Publica do Estado do Rio Grande do Sul (LACEN-RS) | Laboratory of Respiratory Viruses and Measles, Oswaldo Cruz Institute, FIOCRUZ | Paola Resende, Luciana Appolinario, Fernando Motta, Anna Carolina Paixao, Ana Carolina Mendonca, Alice Sampaio Rocha, Taina Venas, Elisa Cavalcante Pereira, Renata Serrano Lopes, Anderson Brandao Leite, Marilda Siqueira on behalf of the Fiocruz COVID-19 Genomic Surveillance Network |
| EPI_ISL_2614070, EPI_ISL_2614071, EPI_ISL_2614072, EPI_ISL_2614073, EPI_ISL_2614074, EPI_ISL_2614075, EPI_ISL_2614076, EPI_ISL_2614077, EPI_ISL_2614078, EPI_ISL_2614079, EPI_ISL_2614080, EPI_ISL_2614081, EPI_ISL_2614082, EPI_ISL_2614083, EPI_ISL_2614084, EPI_ISL_2614085, EPI_ISL_2614086, EPI_ISL_2614087, EPI_ISL_2614088, EPI_ISL_2614089, EPI_ISL_2614090, EPI_ISL_2614091 | | | |
| see above | Laboratory of Molecular Virology, Federal University of Rio de Janeito, UFRJ | Laboratory of Respiratory Viruses and Measles, Oswaldo Cruz Institute, FIOCRUZ | Paola Resende, Amilcar Tanuri, Luciana Appolinario, Fernando Motta, Anna Carolina Paixao, Ana Carolina Mendonca, Alice Sampaio Rocha, Taina Venas, Elisa Cavalcante Pereira, Renata Serrano Lopes, Marilda Siqueira on behalf of the Fiocruz COVID-19 Genomic Surveillance Network |
| EPI_ISL_2614092, EPI_ISL_2614093, EPI_ISL_2614094, EPI_ISL_2614095, EPI_ISL_2614096, EPI_ISL_2614097, EPI_ISL_2614098, EPI_ISL_2614099, EPI_ISL_2614100, EPI_ISL_2614101, EPI_ISL_2614102, EPI_ISL_2614103, EPI_ISL_2614104, EPI_ISL_2614105, EPI_ISL_2614106, EPI_ISL_2614107, EPI_ISL_2614108, EPI_ISL_2614109, EPI_ISL_2614110, EPI_ISL_2614111, EPI_ISL_2614112, EPI_ISL_2614113, EPI_ISL_2614114 | | | |
| see above | Laboratory of Respiratory Viruses and Measles, Oswaldo Cruz Institute, FIOCRUZ | Laboratory of Respiratory Viruses and Measles, Oswaldo Cruz Institute, FIOCRUZ | Paola Resende, Luciana Appolinario, Fernando Motta, Anna Carolina Paixao, Ana Carolina Mendonca, Alice Sampaio Rocha, Taina Venas, Elisa Cavalcante Pereira, Renata Serrano Lopes, Marilda Siqueira on behalf of the Fiocruz COVID-19 Genomic Surveillance Network |
| EPI_ISL_2614115 | Laboratory of Respiratory Viruses and Measles, Oswaldo Cruz Institute, FIOCRUZ | Laboratory of Respiratory Viruses and Measles, Oswaldo Cruz Institute, FIOCRUZ | Paola Resende, Amilcar Tanuri, Luciana Appolinario, Fernando Motta, Anna Carolina Paixao, Ana Carolina Mendonca, Alice Sampaio Rocha, Taina Venas, Elisa Cavalcante Pereira, Renata Serrano Lopes, Marilda Siqueira on behalf of the Fiocruz COVID-19 Genomic Surveillance Network |
| EPI_ISL_2614116, EPI_ISL_2614117, EPI_ISL_2614118, EPI_ISL_2614119, EPI_ISL_2614120, EPI_ISL_2614121, EPI_ISL_2614122, EPI_ISL_2614123, EPI_ISL_2614124, EPI_ISL_2614125, EPI_ISL_2614126, EPI_ISL_2614127, EPI_ISL_2614128, EPI_ISL_2614129, EPI_ISL_2614130, EPI_ISL_2614131, EPI_ISL_2614132, EPI_ISL_2614133, EPI_ISL_2614134, EPI_ISL_2614135, EPI_ISL_2614136, EPI_ISL_2614137, EPI_ISL_2614138, EPI_ISL_2614139, EPI_ISL_2614140, EPI_ISL_2614141, EPI_ISL_2614142, EPI_ISL_2614143, EPI_ISL_2614144, EPI_ISL_2614145, EPI_ISL_2614146, EPI_ISL_2614147, EPI_ISL_2614148, EPI_ISL_2614149, EPI_ISL_2614150, EPI_ISL_2614151, EPI_ISL_2614152, EPI_ISL_2614153, EPI_ISL_2614154, EPI_ISL_2614155, EPI_ISL_2614156, EPI_ISL_2614157, EPI_ISL_2614158, EPI_ISL_2614159, EPI_ISL_2614160, EPI_ISL_2614161, EPI_ISL_2614162, EPI_ISL_2614163, EPI_ISL_2614164, EPI_ISL_2614165, EPI_ISL_2614166, EPI_ISL_2614167, EPI_ISL_2614168, EPI_ISL_2614169, EPI_ISL_2614170, EPI_ISL_2614171, EPI_ISL_2614172, EPI_ISL_2614173, EPI_ISL_2614174, EPI_ISL_2614175, EPI_ISL_2614176, EPI_ISL_2614177, EPI_ISL_2614178, EPI_ISL_2614181, EPI_ISL_2614182, EPI_ISL_2614183, EPI_ISL_2614184, EPI_ISL_2614185, EPI_ISL_2614186, EPI_ISL_2614187, EPI_ISL_2614188, EPI_ISL_2614189, EPI_ISL_2614190, EPI_ISL_2614191, EPI_ISL_2614192, EPI_ISL_2614193, EPI_ISL_2614194, EPI_ISL_2614195, EPI_ISL_2614196, EPI_ISL_2614197, EPI_ISL_2614198, EPI_ISL_2614199, EPI_ISL_2614200, EPI_ISL_2614201, EPI_ISL_2614202, EPI_ISL_2614203, EPI_ISL_2614204, EPI_ISL_2614205, EPI_ISL_2614206, EPI_ISL_2614207, EPI_ISL_2614209, EPI_ISL_2614210, EPI_ISL_2614211, EPI_ISL_2614212, EPI_ISL_2614213, EPI_ISL_2614214, EPI_ISL_2614215, EPI_ISL_2614216, EPI_ISL_2614217, EPI_ISL_2614218, EPI_ISL_2614219, EPI_ISL_2614220, EPI_ISL_2614221, EPI_ISL_2614222, EPI_ISL_2614223, EPI_ISL_2614224, EPI_ISL_2614225, EPI_ISL_2614226, EPI_ISL_2614227, EPI_ISL_2614228, EPI_ISL_2614229, EPI_ISL_2614230, EPI_ISL_2614231, EPI_ISL_2614232, EPI_ISL_2614233, EPI_ISL_2614234, EPI_ISL_2614235, EPI_ISL_2614236, EPI_ISL_2614237, EPI_ISL_2614238, EPI_ISL_2614239, EPI_ISL_2614240, EPI_ISL_2614241, EPI_ISL_2614242, EPI_ISL_2614243, EPI_ISL_2614244, EPI_ISL_2614245, EPI_ISL_2614246, EPI_ISL_2614247, EPI_ISL_2614248, EPI_ISL_2614249, EPI_ISL_2614250, EPI_ISL_2614251, EPI_ISL_2614252, EPI_ISL_2614253, EPI_ISL_2614254, EPI_ISL_2614255, EPI_ISL_2614256, EPI_ISL_2614257, EPI_ISL_2614258, EPI_ISL_2614259, EPI_ISL_2614260, EPI_ISL_2614261, EPI_ISL_2614262, EPI_ISL_2614263, EPI_ISL_2614264, EPI_ISL_2614265, EPI_ISL_2614266, EPI_ISL_2614267, EPI_ISL_2614268, EPI_ISL_2614269, EPI_ISL_2614270, EPI_ISL_2614271, EPI_ISL_2614272, EPI_ISL_2614273, EPI_ISL_2614274, EPI_ISL_2614275, EPI_ISL_2614276, EPI_ISL_2614277, EPI_ISL_2614278, EPI_ISL_2614279, EPI_ISL_2614280, EPI_ISL_2614281, EPI_ISL_2614282, EPI_ISL_2614283, EPI_ISL_2614284, EPI_ISL_2614285, EPI_ISL_2614286, EPI_ISL_2614287, EPI_ISL_2614288, EPI_ISL_2614289, EPI_ISL_2614290, EPI_ISL_2614291, EPI_ISL_2614292, EPI_ISL_2614293, EPI_ISL_2614294, EPI_ISL_2614295, EPI_ISL_2614296, EPI_ISL_2614297, EPI_ISL_2614298, EPI_ISL_2614299, EPI_ISL_2614300, EPI_ISL_2614301, EPI_ISL_2614302, EPI_ISL_2614303, EPI_ISL_2614304, EPI_ISL_2614305, EPI_ISL_2614306, EPI_ISL_2614307, EPI_ISL_2614308, EPI_ISL_2614309, EPI_ISL_2614310, EPI_ISL_2614311, EPI_ISL_2614312, EPI_ISL_2614313, EPI_ISL_2614314, EPI_ISL_2614315, EPI_ISL_2614316, EPI_ISL_2614317, EPI_ISL_2614318, EPI_ISL_2614319, EPI_ISL_2614320, EPI_ISL_2614321, EPI_ISL_2614322, EPI_ISL_2614323, EPI_ISL_2614324, EPI_ISL_2614325, EPI_ISL_2614326, EPI_ISL_2614327, EPI_ISL_2614328, EPI_ISL_2614329, EPI_ISL_2614330, EPI_ISL_2614331, EPI_ISL_2614332, EPI_ISL_2614333, EPI_ISL_2614334, EPI_ISL_2614335, EPI_ISL_2614336, EPI_ISL_2614337, EPI_ISL_2614338, EPI_ISL_2614339, EPI_ISL_2614340, EPI_ISL_2614341, EPI_ISL_2614342, EPI_ISL_2614343, EPI_ISL_2614344, EPI_ISL_2614345, EPI_ISL_2614346, EPI_ISL_2614347, EPI_ISL_2614348, EPI_ISL_2614349, EPI_ISL_2614350 | | | |
| see above | Laboratory of Respiratory Viruses and Measles, Oswaldo Cruz Institute, FIOCRUZ | Laboratory of Respiratory Viruses and Measles, Oswaldo Cruz Institute, FIOCRUZ | Paola Resende, Luciana Appolinario, Fernando Motta, Anna Carolina Paixao, Ana Carolina Mendonca, Alice Sampaio Rocha, Taina Venas, Elisa Cavalcante Pereira, Renata Serrano Lopes, Marilda Siqueira on behalf of the Fiocruz COVID-19 Genomic Surveillance Network |
| EPI_ISL_2614351, EPI_ISL_2614352, EPI_ISL_2614353, EPI_ISL_2614354, EPI_ISL_2614355, EPI_ISL_2614356, EPI_ISL_2614357, EPI_ISL_2614358, EPI_ISL_2614359, EPI_ISL_2614360, EPI_ISL_2614361, EPI_ISL_2614362, EPI_ISL_2614363, EPI_ISL_2614364, EPI_ISL_2614365, EPI_ISL_2614366, EPI_ISL_2614367, EPI_ISL_2614368, EPI_ISL_2614369, EPI_ISL_2614370, EPI_ISL_2614371, EPI_ISL_2614372, EPI_ISL_2614373, EPI_ISL_2614374, EPI_ISL_2614375, EPI_ISL_2614376, EPI_ISL_2614377, EPI_ISL_2614378, EPI_ISL_2614379, EPI_ISL_2614380 | | | |
| see above | Labortorio Central de Saude Publica do Estado do Rio de Janeiro (LACEN/RJ) | Laboratory of Respiratory Viruses and Measles, Oswaldo Cruz Institute, FIOCRUZ | Paola Resende, Luciana Appolinario, Fernando Motta, Anna Carolina Paixao, Ana Carolina Mendonca, Alice Sampaio Rocha, Taina Venas, Elisa Cavalcante Pereira, Renata Serrano Lopes, Andrea Cony Cavalcanti, Marilda Siqueira on behalf of the Fiocruz COVID-19 Genomic Surveillance Network |
| EPI_ISL_2614381, EPI_ISL_2614382, EPI_ISL_2614383 | Instituto Estadual do Cerebro Paulo Niemeyer | Laboratory of Respiratory Viruses and Measles, Oswaldo Cruz Institute, FIOCRUZ | Paola Resende,Carlos Azevedo, Luciana Appolinario, Fernando Motta, Anna Carolina Paixao, Ana Carolina Mendonca, Alice Sampaio Rocha, Taina Venas, Elisa Cavalcante Pereira, Renata Serrano Lopes, Marilda Siqueira on behalf of the Fiocruz COVID-19 Genomic Surveillance Network |
| EPI_ISL_2691155 | Instituo Adolfo Lutz Central | Instituto Adolfo Lutz, Interdiciplinary Procedures Center, Strategic Laboratory | Claudio Tavares Sacchi, Claudia Regina Gonçalves, Erica Valessa Ramos Gomes, Karoline Rodrigues Campos, Caio Vinicius Dias Lopes, Leonardo Jose Tadeu de Araujo |
| EPI_ISL_2691532 | Laboratorio Central Noel Nutels | Bioinformatics Laboratory / LNCC | Luiz G P de Almeida, Alessandra P Lamarca, Ronaldo da Silva F Jr, Liliane Cavalcante, Alexandra L Gerber, Ana Paula de C Guimaraes, Douglas Terra Machado, Cassia Alves, Diana Mariani, Cintia Policarpo, Gleidson da Silva de Oliveira, Mario Sergio Ribeiro, Silvia Carvalho, Flavio Dias da Silva, Marcio Henrique de Oliveira Garcia, Leandro Magalhaes de Souza, Cristiane Gomes da Silva, Caio Luiz Pereira Ribeiro, Andrea Cony Cavalcanti, Claudia Maria Braga de Mello, Amilcar Tanuri, Ana Tereza R Vasconcelos |
| EPI_ISL_406798 | General Hospital of Central Theater Command of People's Liberation Army of China | BGI & Institute of Microbiology, Chinese Academy of Sciences & Shandong First Medical University & Shandong Academy of Medical Sciences & General Hospital of Central Theater Command of People's Liberation Army of China | Weijun Chen, Yuhai Bi, Weifeng Shi and Zhenhong Hu |
| EPI_ISL_414045 | Laboratório Central de Saúde Pública Noel Nutels (LACEN-RJ) | Laboratory of Respiratory Viruses and Measles, Oswaldo Cruz Institute, FIOCRUZ | Paola Resende, Alisson Fabri, Joilson Xavier, Sunando Roy, Fernando Motta, Aline Mattos, Milene Miranda, Cristiana Garcia, Braulia Caetano, Maria Ogrzewalska, Jonathan Lopes, Luciana Appolinario, Maria Nóbrega, Marilda SiqueiraFiocruz COVID-19 Genomic Surveillance Network |

| | | | |
|---|---|---|---|
| EPI_ISL_427299, EPI_ISL_427300, EPI_ISL_427301, EPI_ISL_427302, EPI_ISL_427303, EPI_ISL_427304 | Laboratory of Respiratory Viruses and Measles, Oswaldo Cruz Institute, FIOCRUZ | Laboratory of Respiratory Viruses and Measles, Oswaldo Cruz Institute, FIOCRUZ | Paola Resende, Fernando Motta, Luciana Appolinario, Sunando Roy, Aline Mattos, Milene Miranda, Cristiana Garcia, Braulia Caetano, Maria Ogrzewalska, Priscila Born, Jonathan Lopes, Marilda Siqueira on behalf of the Fiocruz COVID-19 Genomic Surveillance Network |
| EPI_ISL_456071, EPI_ISL_456072, EPI_ISL_456073, EPI_ISL_456074, EPI_ISL_456075 | Laboratory of Respiratory Viruses and Measles, Oswaldo Cruz Institute, FIOCRUZ | Laboratory of Respiratory Viruses and Measles, Oswaldo Cruz Institute, FIOCRUZ | Paola Resende, Luciana Appolinario, Fernando Motta, Aline Mattos, Milene Miranda, Cristiana Garcia, Braulia Caetano, Maria Ogrzewalska, Jonathan Lopes, Marilda Siqueira on behalf of the Fiocruz COVID-19 Genomic Surveillance Network |
| EPI_ISL_456076, EPI_ISL_456077 | Laboratório Central de Saúde Pública Noel Nutels (LACEN-RJ) | Laboratory of Respiratory Viruses and Measles, Oswaldo Cruz Institute, FIOCRUZ | Paola Resende, Luciana Appolinario, Fernando Motta, Aline Mattos, Milene Miranda, Cristiana Garcia, Braulia Caetano, Maria Ogrzewalska, Jonathan Lopes, Marilda Siqueira on behalf of the Fiocruz COVID-19 Genomic Surveillance Network |
| EPI_ISL_456079, EPI_ISL_456080, EPI_ISL_456081 | Laboratory of Respiratory Viruses and Measles, Oswaldo Cruz Institute, FIOCRUZ | Laboratory of Respiratory Viruses and Measles, Oswaldo Cruz Institute, FIOCRUZ | Paola Resende, Luciana Appolinario, Fernando Motta, Aline Mattos, Milene Miranda, Cristiana Garcia, Braulia Caetano, Maria Ogrzewalska, Jonathan Lopes, Marilda Siqueira on behalf of the Fiocruz COVID-19 Genomic Surveillance Network |
| EPI_ISL_456082, EPI_ISL_456083 | Laboratório Central de Saúde Pública Noel Nutels (LACEN-RJ) | Laboratory of Respiratory Viruses and Measles, Oswaldo Cruz Institute, FIOCRUZ | Paola Resende, Luciana Appolinario, Fernando Motta, Aline Mattos, Milene Miranda, Cristiana Garcia, Braulia Caetano, Maria Ogrzewalska, Jonathan Lopes, Marilda Siqueira on behalf of the Fiocruz COVID-19 Genomic Surveillance Network |
| EPI_ISL_456084, EPI_ISL_456085, EPI_ISL_456086, EPI_ISL_456087 | Laboratory of Respiratory Viruses and Measles, Oswaldo Cruz Institute, FIOCRUZ | Laboratory of Respiratory Viruses and Measles, Oswaldo Cruz Institute, FIOCRUZ | Paola Resende, Luciana Appolinario, Fernando Motta, Aline Mattos, Milene Miranda, Cristiana Garcia, Braulia Caetano, Maria Ogrzewalska, Jonathan Lopes, Marilda Siqueira on behalf of the Fiocruz COVID-19 Genomic Surveillance Network |
| EPI_ISL_456088 | Laboratório Central de Saúde Pública Noel Nutels (LACEN-RJ) | Laboratory of Respiratory Viruses and Measles, Oswaldo Cruz Institute, FIOCRUZ | Paola Resende, Luciana Appolinario, Fernando Motta, Aline Mattos, Milene Miranda, Cristiana Garcia, Braulia Caetano, Maria Ogrzewalska, Jonathan Lopes, Marilda Siqueira on behalf of the Fiocruz COVID-19 Genomic Surveillance Network |
| EPI_ISL_456089, EPI_ISL_456090, EPI_ISL_456091, EPI_ISL_456092, EPI_ISL_456093, EPI_ISL_456094, EPI_ISL_456095, EPI_ISL_456096, EPI_ISL_456097, EPI_ISL_456098, EPI_ISL_456099, EPI_ISL_456100, EPI_ISL_456101, EPI_ISL_456102, EPI_ISL_456103, EPI_ISL_456104, EPI_ISL_456105, EPI_ISL_456106 | | | |
| see above | Laboratory of Respiratory Viruses and Measles, Oswaldo Cruz Institute, FIOCRUZ | Laboratory of Respiratory Viruses and Measles, Oswaldo Cruz Institute, FIOCRUZ | Paola Resende, Luciana Appolinario, Fernando Motta, Aline Mattos, Milene Miranda, Cristiana Garcia, Braulia Caetano, Maria Ogrzewalska, Jonathan Lopes, Marilda Siqueira on behalf of the Fiocruz COVID-19 Genomic Surveillance Network |
| EPI_ISL_467344, EPI_ISL_467345, EPI_ISL_467346, EPI_ISL_467347, EPI_ISL_467348, EPI_ISL_467349, EPI_ISL_467350, EPI_ISL_467351, EPI_ISL_467352, EPI_ISL_467353, EPI_ISL_467354, EPI_ISL_467355, EPI_ISL_467356, EPI_ISL_467357, EPI_ISL_467358, EPI_ISL_467359, EPI_ISL_467360, EPI_ISL_467362, EPI_ISL_467363, EPI_ISL_467364, EPI_ISL_467365, EPI_ISL_467366, EPI_ISL_467367, EPI_ISL_467368, EPI_ISL_467369, EPI_ISL_467370, EPI_ISL_467371 | | | |
| see above | Laboratory of Respiratory Viruses and Measles, Oswaldo Cruz Institute, FIOCRUZ | Laboratory of Respiratory Viruses and Measles, Oswaldo Cruz Institute, FIOCRUZ | Paola Resende, Luciana Appolinario, Fernando Motta, Anna Carolina Paixão, Ana Carolina Mendonça, Aline Mattos, Milene Miranda, Cristiana Garcia, Braulia Caetano, Maria Ogrzewalska, Jonathan Lopes, Marilda Siqueira on behalf of the Fiocruz COVID-19 Genomic Surveillance Network |
| EPI_ISL_483065 | Centro de Desenvolvimento Tecnologico em Saude, Fundacao Oswaldo Cruz | Centro de Desenvolvimento Tecnologico em Saude, Fundacao Oswaldo Cruz | Souza,T.M., Fintelman-Rodrigues,N., De Paula,A.D., Tschoeke,D., Barroso,S.P., Gregorio,M.L., Oliveira,J.S., Saraiva,F.B., Ferreira,M.A., Sacramento,C.Q. |
| EPI_ISL_492032 | Instituto de Biologia do Exército | Laboratório Metabolismo Macromolecular FirminoTorres de Castro, Instituto de Biofísica Carlos Chagas Filho, Universidade Federal do Rio de Janeiro | Bianca Catarina Azevedo Cabral, Aline Rosa Vianna de Souza , Marcos Dornelas-Ribeiro, Tatiana LS Nogueira, Nádia Vaez Gonçalves da Cruz, Caleb GM Santos, Elizabeth Valentin, Marcio da Costa Cipitelli, Virginia Sara Grancieri do Amaral, Rodrigo Soares de Moura Neto, Clarissa Damaso, Rosane Silva |
| EPI_ISL_492033 | Instituto de Biologia do Exército | Laboratório Metabolismo Macromolecular FirminoTorres de Castro, Instituto de Biofísica Carlos Chagas Filho, Universidade Federal do Rio de Janeiro | Bianca Catarina Azevedo Cabral, Aline Rosa Vianna de Souza, Caleb GM Santos, Marcos Dornelas-Ribeiro, Tatiana LS Nogueira, Nádia Vaez Gonçalves da Cruz, Elizabeth Valentin, Marcio da Costa Cipitelli, Virginia Sara Grancieri do Amaral, Rodrigo Soares de Moura Neto, Clarissa Damaso, Rosane Silva |
| EPI_ISL_492034 | Instituto de Biologia do Exército | Laboratório Metabolismo Macromolecular FirminoTorres de Castro, Instituto de Biofísica Carlos Chagas Filho, Universidade Federal do Rio de Janeiro | Bianca Catarina Azevedo Cabral, Aline Rosa Vianna de Souza, Nádia Vaez Gonçalves da Cruz, Caleb GM Santos, Marcos Dornelas-Ribeiro, Tatiana LS Nogueira, Elizabeth Valentin, Marcio da Costa Cipitelli, Virginia Sara Grancieri do Amaral, Rodrigo Soares de Moura Neto, Clarissa Damaso, Rosane Silva |
| EPI_ISL_492035 | Instituto de Biologia do Exército | Laboratório Metabolismo Macromolecular FirminoTorres de Castro, Instituto de Biofísica Carlos Chagas Filho, Universidade Federal do Rio de Janeiro | Bianca Catarina Azevedo Cabral, Aline Rosa Vianna de Souza, Tatiana LS Nogueira, Nádia Vaez Gonçalves da Cruz, Caleb GM Santos, Marcos Dornelas-Ribeiro, Elizabeth Valentin, Marcio da Costa Cipitelli, Virginia Sara Grancieri do Amaral, Rodrigo Soares de Moura Neto, Clarissa Damaso, Rosane Silva |
| EPI_ISL_492036 | Instituto de Biologia do Exército | Laboratório Metabolismo Macromolecular FirminoTorres de Castro, Instituto de Biofísica Carlos Chagas Filho, Universidade Federal do Rio de Janeiro | Bianca Catarina Azevedo Cabral, Aline Rosa Vianna de Souza , Marcos Dornelas-Ribeiro, Tatiana LS Nogueira, Nádia Vaez Gonçalves da Cruz, Caleb GM Santos, Elizabeth Valentin, Marcio da Costa Cipitelli, Virginia Sara Grancieri do Amaral, Rodrigo Soares de Moura Neto, Clarissa Damaso, Rosane Silva |
| EPI_ISL_492037 | Instituto de Biologia do Exército | Laboratório Metabolismo Macromolecular FirminoTorres de Castro, Instituto de Biofísica Carlos Chagas Filho, Universidade Federal do Rio de Janeiro | Bianca Catarina Azevedo Cabral, Aline Rosa Vianna de Souza, Caleb GM Santos, Marcos Dornelas-Ribeiro, Tatiana LS Nogueira, Nádia Vaez Gonçalves da Cruz, Elizabeth Valentin, Marcio da Costa Cipitelli, Virginia Sara Grancieri do Amaral, Rodrigo Soares de Moura Neto, Clarissa Damaso, Rosane Silva |
| EPI_ISL_492038 | Instituto de Biologia do Exército | Laboratório Metabolismo Macromolecular FirminoTorres de Castro, Instituto de Biofísica Carlos Chagas Filho, Universidade Federal do Rio de Janeiro | Bianca Catarina Azevedo Cabral, Aline Rosa Vianna de Souza, Nádia Vaez Gonçalves da Cruz, Caleb GM Santos, Marcos Dornelas-Ribeiro, Tatiana LS Nogueira, Elizabeth Valentin, Marcio da Costa Cipitelli, Virginia Sara Grancieri do Amaral, Rodrigo Soares de Moura Neto, Clarissa Damaso, Rosane Silva |
| EPI_ISL_492039 | Instituto de Biologia do Exército | Laboratório Metabolismo Macromolecular FirminoTorres de Castro, Instituto de Biofísica Carlos Chagas Filho, Universidade Federal do Rio de Janeiro | Bianca Catarina Azevedo Cabral, Aline Rosa Vianna de Souza, Tatiana LS Nogueira, Nádia Vaez Gonçalves da Cruz, Caleb GM Santos, Marcos Dornelas-Ribeiro, Elizabeth Valentin, Marcio da Costa Cipitelli, Virginia Sara Grancieri do Amaral, Rodrigo Soares de Moura Neto, Clarissa Damaso, Rosane Silva |
| EPI_ISL_492040 | Instituto de Biologia do Exército | Laboratório Metabolismo Macromolecular FirminoTorres de Castro, Instituto de Biofísica Carlos Chagas Filho, Universidade Federal do Rio de Janeiro | Bianca Catarina Azevedo Cabral, Aline Rosa Vianna de Souza , Marcos Dornelas-Ribeiro, Tatiana LS Nogueira, Nádia Vaez Gonçalves da Cruz, Caleb GM Santos, Elizabeth Valentin, Marcio da Costa Cipitelli, Virginia Sara Grancieri do Amaral, Rodrigo Soares de Moura Neto, Clarissa Damaso, Rosane Silva |
| EPI_ISL_492041 | Instituto de Biologia do Exército | Laboratório Metabolismo Macromolecular FirminoTorres de Castro, Instituto de Biofísica Carlos Chagas Filho, Universidade Federal do Rio de Janeiro | Bianca Catarina Azevedo Cabral, Aline Rosa Vianna de Souza, Caleb GM Santos, Marcos Dornelas-Ribeiro, Tatiana LS Nogueira, Nádia Vaez Gonçalves da Cruz, Elizabeth Valentin, Marcio da Costa Cipitelli, Virginia Sara Grancieri do Amaral, Rodrigo Soares de Moura Neto, Clarissa Damaso, Rosane Silva |
| EPI_ISL_492042 | Instituto de Biologia do Exército | Laboratório Metabolismo Macromolecular FirminoTorres de Castro, Instituto de Biofísica Carlos Chagas Filho, Universidade Federal do Rio de Janeiro | Bianca Catarina Azevedo Cabral, Aline Rosa Vianna de Souza, Nádia Vaez Gonçalves da Cruz, Caleb GM Santos, Marcos Dornelas-Ribeiro, Tatiana LS Nogueira, Elizabeth Valentin, Marcio da Costa Cipitelli, Virginia Sara Grancieri do Amaral, Rodrigo Soares de Moura Neto, Clarissa Damaso, Rosane Silva |
| EPI_ISL_492043 | Instituto de Biologia do Exército | Laboratório Metabolismo Macromolecular FirminoTorres de Castro, Instituto de Biofísica Carlos Chagas Filho, Universidade Federal do Rio de Janeiro | Bianca Catarina Azevedo Cabral, Aline Rosa Vianna de Souza, Tatiana LS Nogueira, Nádia Vaez Gonçalves da Cruz, Caleb GM Santos, Marcos Dornelas-Ribeiro, Elizabeth Valentin, Marcio da Costa Cipitelli, Virginia Sara Grancieri do Amaral, Rodrigo Soares de Moura Neto, Clarissa Damaso, Rosane Silva |
| EPI_ISL_492044 | Instituto de Biologia do Exército | Laboratório Metabolismo Macromolecular FirminoTorres de Castro, Instituto de Biofísica Carlos Chagas Filho, Universidade Federal do Rio de Janeiro | Bianca Catarina Azevedo Cabral, Aline Rosa Vianna de Souza , Marcos Dornelas-Ribeiro, Tatiana LS Nogueira, Nádia Vaez Gonçalves da Cruz, Caleb GM Santos, Elizabeth Valentin, Marcio da Costa Cipitelli, Virginia Sara Grancieri do Amaral, Rodrigo Soares de Moura Neto, Clarissa Damaso, Rosane Silva |
| EPI_ISL_492045 | Instituto de Biologia do Exército | Laboratório Metabolismo Macromolecular FirminoTorres de Castro, Instituto de Biofísica Carlos Chagas Filho, Universidade Federal do Rio de Janeiro | Bianca Catarina Azevedo Cabral, Aline Rosa Vianna de Souza, Caleb GM Santos, Marcos Dornelas-Ribeiro, Tatiana LS Nogueira, Nádia Vaez Gonçalves da Cruz, Elizabeth Valentin, Marcio da Costa Cipitelli, Virginia Sara Grancieri do Amaral, Rodrigo Soares de Moura Neto, Clarissa Damaso, Rosane Silva |
| EPI_ISL_492048 | Instituto de Biologia do Exército | Laboratório Metabolismo Macromolecular FirminoTorres de Castro, Instituto de Biofísica Carlos Chagas Filho, Universidade Federal do Rio de Janeiro | Bianca Catarina Azevedo Cabral, Aline Rosa Vianna de Souza , Marcos Dornelas-Ribeiro, Tatiana LS Nogueira, Nádia Vaez Gonçalves da Cruz, Caleb GM Santos, Elizabeth Valentin, Marcio da Costa Cipitelli, Virginia Sara Grancieri do Amaral, Rodrigo Soares de Moura Neto, Clarissa Damaso, Rosane Silva |
| EPI_ISL_509430, EPI_ISL_509431, EPI_ISL_509432, EPI_ISL_509433, EPI_ISL_509434, EPI_ISL_509435 | Centro de Desenvolvimento Tecnologico em Saude, Fundacao Oswaldo Cruz | Centro de Desenvolvimento Tecnologico em Saude, Fundacao Oswaldo Cruz | Souza,T.M., Fintelman-Rodrigues,N., De Paula,A.D., Saraiva,F.B., Ferreira,M.A., Sacramento,C.Q., Medeiros,M.A. |

| | | | |
|---|---|---|---|
| EPI_ISL_510536 | Centro de Desenvolvimento Tecnologico em Saude, Fundacao Oswaldo Cruz | Centro de Desenvolvimento Tecnologico em Saude, Fundacao Oswaldo Cruz | Souza,T.M., Fintelman-Rodrigues,N., De Paula,A.D., Saraiva,F.B., Ferreira,M.A., Sacramento,C.Q. and Medeiros,M.A. |
| EPI_ISL_510541 | Centro de Desenvolvimento Tecnologico em Saude, Fundacao Oswaldo Cruz | Centro de Desenvolvimento Tecnologico em Saude, Fundacao Oswaldo Cruz | Souza,T.M., Fintelman-Rodrigues,N., De Paula,A.D., Saraiva,F.B., Ferreira,M.A., Sacramento,C.Q. and Medeiros,M.A. |
| EPI_ISL_513513, EPI_ISL_513514, EPI_ISL_513515, EPI_ISL_513516, EPI_ISL_513518, EPI_ISL_513519, EPI_ISL_513520, EPI_ISL_513521, EPI_ISL_513522, EPI_ISL_513524, EPI_ISL_513525, EPI_ISL_513526, EPI_ISL_513527, EPI_ISL_513529, EPI_ISL_513530, EPI_ISL_513531, EPI_ISL_513532, EPI_ISL_513533, EPI_ISL_513534, EPI_ISL_513535, EPI_ISL_513536, EPI_ISL_513537, EPI_ISL_513538, EPI_ISL_513539, EPI_ISL_513540, EPI_ISL_513541, EPI_ISL_513542, EPI_ISL_513543, EPI_ISL_513544, EPI_ISL_513545, EPI_ISL_513546, EPI_ISL_513547, EPI_ISL_513548, EPI_ISL_513549, EPI_ISL_513550, EPI_ISL_513551, EPI_ISL_513553, EPI_ISL_513556, EPI_ISL_513557, EPI_ISL_513558, EPI_ISL_513559, EPI_ISL_513560, EPI_ISL_513561, EPI_ISL_513562, EPI_ISL_513563, EPI_ISL_513564, EPI_ISL_513567, EPI_ISL_513568, EPI_ISL_513571, EPI_ISL_513572, EPI_ISL_513573, EPI_ISL_513574, EPI_ISL_513575, EPI_ISL_513576, EPI_ISL_513577, EPI_ISL_513578, EPI_ISL_513579, EPI_ISL_513580, EPI_ISL_513581, EPI_ISL_513582, EPI_ISL_513583 | | | |
| see above | Programa de Oncovirologia, Instituto Nacional de Câncer | Programa de Oncovirologia, Instituto Nacional de Câncer | Juliana D. Siqueira, Livia R. Goes, Brunna M. Alves, Claudia Cicala,James Arthos, João P.B. Viola, Andreia C. de Melo, Marcelo A. Soares |
| EPI_ISL_528539 | LVM/UFRJ | LNCC | Gustavo M. Romário M. de Souza; Bruno B. Bezerra; Lucio A. Caldas; Fabio Limonte; Elena Cobos; Sharton V. A. Coelho; Luiz Almeida; Luiza Higga; Isadora A. Correa; Diana Marianni; Luciana B. Arruda; Marcelo Bozza; Orlando Ferreira; Wanderley de Souza; Ana Teresa R. Vasconcelos; Terezinha M. Castineiras; Amilcar Tanuri; Luciana J. Costa. |
| EPI_ISL_528637, EPI_ISL_528638 | LVM/UFRJ | Bioinformatics Laboratory / LNCC | Gustavo D. P. Silva; M. Romário M. de Souza; Bruno B. Bezerra; Lucio A. Caldas; Fabio Limonte; Elena Cobos; Sharton V. A. Coelho; Luiz Almeida; Luiza Higga; Isadora A. Correa; Diana Marianni; Luciana B. Arruda; Marcelo Bozza; Orlando Ferreira; Wanderley de Souza; Ana Teresa R. Vasconcelos; Terezinha M. Castineiras; Amilcar Tanuri; Luciana J. Costa |
| EPI_ISL_529139 | Centro de Desenvolvimento Tecnologico em Saude, Fundacao Oswaldo Cruz | Centro de Desenvolvimento Tecnologico em Saude, Fundacao Oswaldo Cruz | Souza,T.M., Fintelman-Rodrigues,N., De Paula,A.D., Saraiva,F.B., Ferreira,M.A., Sacramento,C.Q., Medeiros,M.A. |
| EPI_ISL_529140 | Centro de Desenvolvimento Tecnologico em Saude, Fundacao Oswaldo Cruz | Centro de Desenvolvimento Tecnologico em Saude, Fundacao Oswaldo Cruz | Souza,T.M., Fintelman-Rodrigues,N., De Paula,A.D., Saraiva,F.B., Ferreira,M.A., Sacramento,C.Q., Medeiros,M.A. |
| EPI_ISL_541347, EPI_ISL_541348, EPI_ISL_541349, EPI_ISL_541350, EPI_ISL_541351 | Laboratory of Respiratory Viruses and Measles, Oswaldo Cruz Institute, FIOCRUZ | Laboratory of Respiratory Viruses and Measles, Oswaldo Cruz Institute, FIOCRUZ | Paola Resende, Luciana Appolinario, Fernando Motta, Anna Carolina Paixão, Ana Carolina Mendonça, Jonathan Lopes, Marilda Siqueira on behalf of the Fiocruz COVID-19 Genomic Surveillance Network |
| EPI_ISL_541352 | Laboratory of Respiratory Viruses and Measles, Oswaldo Cruz Institute, FIOCRUZ | Laboratory of Respiratory Viruses and Measles, Oswaldo Cruz Institute, FIOCRUZ | Paola Resende, Roxana Loayza, Cinthia Avila, Luciana Appolinario, Fernando Motta, Anna Carolina Paixao, Ana Carolina Mendonca, Marilda Siqueira on behalf of the Fiocruz COVID-19 Genomic Surveillance Network |
| EPI_ISL_541353, EPI_ISL_541354, EPI_ISL_541355 | Laboratory of Respiratory Viruses and Measles, Oswaldo Cruz Institute, FIOCRUZ | Laboratory of Respiratory Viruses and Measles, Oswaldo Cruz Institute, FIOCRUZ | Paola Resende, Luciana Appolinario, Fernando Motta, Anna Carolina Paixão, Ana Carolina Mendonça, Jonathan Lopes, Marilda Siqueira on behalf of the Fiocruz COVID-19 Genomic Surveillance Network |
| EPI_ISL_541356 | Laboratory of Respiratory Viruses and Measles, Oswaldo Cruz Institute, FIOCRUZ | Laboratory of Respiratory Viruses and Measles, Oswaldo Cruz Institute, FIOCRUZ | Paola Resende, Roxana Loayza, Cinthia Avila, Luciana Appolinario, Fernando Motta, Anna Carolina Paixao, Ana Carolina Mendonca, Marilda Siqueira on behalf of the Fiocruz COVID-19 Genomic Surveillance Network |
| EPI_ISL_541357, EPI_ISL_541358 | Laboratory of Respiratory Viruses and Measles, Oswaldo Cruz Institute, FIOCRUZ | Laboratory of Respiratory Viruses and Measles, Oswaldo Cruz Institute, FIOCRUZ | Paola Resende, Luciana Appolinario, Fernando Motta, Anna Carolina Paixão, Ana Carolina Mendonça, Jonathan Lopes, Marilda Siqueira on behalf of the Fiocruz COVID-19 Genomic Surveillance Network |
| EPI_ISL_541359, EPI_ISL_541360, EPI_ISL_541361 | Laboratory of Respiratory Viruses and Measles, Oswaldo Cruz Institute, FIOCRUZ | Laboratory of Respiratory Viruses and Measles, Oswaldo Cruz Institute, FIOCRUZ | Paola Resende, Roxana Loayza, Cinthia Avila, Luciana Appolinario, Fernando Motta, Anna Carolina Paixao, Ana Carolina Mendonca, Marilda Siqueira on behalf of the Fiocruz COVID-19 Genomic Surveillance Network |
| EPI_ISL_541362, EPI_ISL_541363 | Laboratory of Respiratory Viruses and Measles, Oswaldo Cruz Institute, FIOCRUZ | Laboratory of Respiratory Viruses and Measles, Oswaldo Cruz Institute, FIOCRUZ | Paola Resende, Luciana Appolinario, Fernando Motta, Anna Carolina Paixão, Ana Carolina Mendonça, Jonathan Lopes, Marilda Siqueira on behalf of the Fiocruz COVID-19 Genomic Surveillance Network |
| EPI_ISL_541364, EPI_ISL_541365, EPI_ISL_541366 | Laboratory of Respiratory Viruses and Measles, Oswaldo Cruz Institute, FIOCRUZ | Laboratory of Respiratory Viruses and Measles, Oswaldo Cruz Institute, FIOCRUZ | Paola Resende, Roxana Loayza, Cinthia Avila, Luciana Appolinario, Fernando Motta, Anna Carolina Paixao, Ana Carolina Mendonca, Marilda Siqueira on behalf of the Fiocruz COVID-19 Genomic Surveillance Network |
| EPI_ISL_541367 | Laboratory of Respiratory Viruses and Measles, Oswaldo Cruz Institute, FIOCRUZ | Laboratory of Respiratory Viruses and Measles, Oswaldo Cruz Institute, FIOCRUZ | Paola Resende, Luciana Appolinario, Fernando Motta, Anna Carolina Paixão, Ana Carolina Mendonça, Jonathan Lopes, Marilda Siqueira on behalf of the Fiocruz COVID-19 Genomic Surveillance Network |
| EPI_ISL_541368, EPI_ISL_541369 | Laboratory of Respiratory Viruses and Measles, Oswaldo Cruz Institute, FIOCRUZ | Laboratory of Respiratory Viruses and Measles, Oswaldo Cruz Institute, FIOCRUZ | Paola Resende, Roxana Loayza, Cinthia Avila, Luciana Appolinario, Fernando Motta, Anna Carolina Paixao, Ana Carolina Mendonca, Marilda Siqueira on behalf of the Fiocruz COVID-19 Genomic Surveillance Network |
| EPI_ISL_623108, EPI_ISL_623110, EPI_ISL_623112, EPI_ISL_623114, EPI_ISL_623116, EPI_ISL_623118, EPI_ISL_623119, EPI_ISL_623120, EPI_ISL_623124, EPI_ISL_623126, EPI_ISL_623129, EPI_ISL_623130, EPI_ISL_623132, EPI_ISL_623134, EPI_ISL_623136, EPI_ISL_623138, EPI_ISL_623140, EPI_ISL_623142, EPI_ISL_623143, EPI_ISL_623144, EPI_ISL_623145, EPI_ISL_623146, EPI_ISL_623147, EPI_ISL_623148, EPI_ISL_623149, EPI_ISL_623152, EPI_ISL_623154, EPI_ISL_623157, EPI_ISL_623158, EPI_ISL_623160, EPI_ISL_623161, EPI_ISL_623162, EPI_ISL_623164, EPI_ISL_623165, EPI_ISL_623166, EPI_ISL_623168 | | | |
| see above | Laboratorio de Virologia Molecular / UFRJ | Bioinformatics Laboratory / LNCC | Carolina M Voloch, Ronaldo S Francisco Jr, Luiz G P de Almeida, Otavio J. Brustolini, Cynthia C Cardoso, Alexandra L Gerber, Ana Paula de C Guimarães, Diana Mariani, Covid19-UFRJ Workgroup, Luís Cristóvão Pôrto, Renato S Aguiar, Terezinha M P P Castiñeiras, Orlando C. Ferreira, Amilcar Tanuri, Ana Tereza R de Vasconcelos |
| EPI_ISL_636737, EPI_ISL_636834, EPI_ISL_636835, EPI_ISL_636837 | Laboratório de Imunofarmacologia - Instituto Oswaldo Cruz | Laboratório de Imunofarmacologia - Instituto Oswaldo Cruz | Souza,T.M., Fintelman-Rodrigues,N., De Paula,A.D., Saraiva,F.B., Ferreira,M.A. and Sacramento,C.Q. |
| EPI_ISL_717806, EPI_ISL_717807, EPI_ISL_717808 | Laboratorio de Virologia Molecular / UFRJ | Bioinformatics Laboratory / LNCC | Carolina M Voloch, Ronaldo da Silva F Jr, Luiz G P de Almeida, Cynthia C Cardoso, Otavio Bustrolini, Alexandra L Gerber, Ana Paula de C Guimarães, Diana Mariani, Andréa Cony Cavalcanti, Claudia dos Santos Rodrigues, Terezinha M P P Castiñeira, Amilcar Tanuri, Ana Tereza R de Vasconcelos |
| EPI_ISL_717809 | LACEN Dr. Francisco Rimolo Neto | Bioinformatics Laboratory / LNCC | Carolina M Voloch, Ronaldo da Silva F Jr, Luiz G P de Almeida, Cynthia C Cardoso, Otavio Bustrolini, Alexandra L Gerber, Ana Paula de C Guimarães, Diana Mariani, Andréa Cony Cavalcanti, Claudia dos Santos Rodrigues, Terezinha M P P Castiñeira, Amilcar Tanuri, Ana Tereza R de Vasconcelos |
| EPI_ISL_717810, EPI_ISL_717811, EPI_ISL_717812, EPI_ISL_717813, EPI_ISL_717814, EPI_ISL_717815, EPI_ISL_717816, EPI_ISL_717817, EPI_ISL_717818, EPI_ISL_717819, EPI_ISL_717820, EPI_ISL_717821, EPI_ISL_717822, EPI_ISL_717823, EPI_ISL_717824, EPI_ISL_717825, EPI_ISL_717826, EPI_ISL_717827, EPI_ISL_717828, EPI_ISL_717829, EPI_ISL_717830, EPI_ISL_717831 | | | |
| see above | Laboratorio de Virologia Molecular / UFRJ | Bioinformatics Laboratory / LNCC | Carolina M Voloch, Ronaldo da Silva F Jr, Luiz G P de Almeida, Cynthia C Cardoso, Otavio Bustrolini, Alexandra L Gerber, Ana Paula de C Guimarães, Diana Mariani, Andréa Cony Cavalcanti, Claudia dos Santos Rodrigues, Terezinha M P P Castiñeira, Amilcar Tanuri, Ana Tereza R de Vasconcelos |
| EPI_ISL_717832, EPI_ISL_717833, EPI_ISL_717834, EPI_ISL_717835, EPI_ISL_717836 | LACEN Dr. Francisco Rimolo Neto | Bioinformatics Laboratory / LNCC | Carolina M Voloch, Ronaldo da Silva F Jr, Luiz G P de Almeida, Cynthia C Cardoso, Otavio Bustrolini, Alexandra L Gerber, Ana Paula de C Guimarães, Diana Mariani, Andréa Cony Cavalcanti, Claudia dos Santos Rodrigues, Terezinha M P P Castiñeira, Amilcar Tanuri, Ana Tereza R de Vasconcelos |
| EPI_ISL_717837, EPI_ISL_717838, EPI_ISL_717839, EPI_ISL_717840 | Laboratorio de Virologia Molecular / UFRJ | Bioinformatics Laboratory / LNCC | Carolina M Voloch, Ronaldo da Silva F Jr, Luiz G P de Almeida, Cynthia C Cardoso, Otavio Bustrolini, Alexandra L Gerber, Ana Paula de C Guimarães, Diana Mariani, Andréa Cony Cavalcanti, Claudia dos Santos Rodrigues, Terezinha M P P Castiñeira, Amilcar Tanuri, Ana Tereza R de Vasconcelos |
| EPI_ISL_717841 | LACEN Dr. Francisco Rimolo Neto | Bioinformatics Laboratory / LNCC | Carolina M Voloch, Ronaldo da Silva F Jr, Luiz G P de Almeida, Cynthia C Cardoso, Otavio Bustrolini, Alexandra L Gerber, Ana Paula de C Guimarães, Diana Mariani, Andréa Cony Cavalcanti, Claudia dos Santos Rodrigues, Terezinha M P P Castiñeira, Amilcar Tanuri, Ana Tereza R de Vasconcelos |
| EPI_ISL_717842, EPI_ISL_717843, EPI_ISL_717844, EPI_ISL_717845, EPI_ISL_717846, EPI_ISL_717847, EPI_ISL_717848, EPI_ISL_717849, EPI_ISL_717850, EPI_ISL_717851, EPI_ISL_717852, EPI_ISL_717853, EPI_ISL_717854, EPI_ISL_717855, EPI_ISL_717856, EPI_ISL_717857, EPI_ISL_717858, EPI_ISL_717859, EPI_ISL_717860, EPI_ISL_717861, EPI_ISL_717862, EPI_ISL_717863, EPI_ISL_717864, EPI_ISL_717865, EPI_ISL_717866, EPI_ISL_717867, EPI_ISL_717868, EPI_ISL_717869, EPI_ISL_717870, EPI_ISL_717871, EPI_ISL_717872, EPI_ISL_717873, EPI_ISL_717874, EPI_ISL_717875, EPI_ISL_717876, EPI_ISL_717877, EPI_ISL_717878, EPI_ISL_717879, EPI_ISL_717880, EPI_ISL_717881, EPI_ISL_717882, EPI_ISL_717883, EPI_ISL_717884, EPI_ISL_717885, EPI_ISL_717886, EPI_ISL_717887, EPI_ISL_717888, EPI_ISL_717889, EPI_ISL_717890, EPI_ISL_717891, EPI_ISL_717892, EPI_ISL_717893, EPI_ISL_717894, EPI_ISL_717895, EPI_ISL_717896, EPI_ISL_717897, EPI_ISL_717898 | | | |
| see above | Laboratorio de Virologia Molecular / UFRJ | Bioinformatics Laboratory / LNCC | Carolina M Voloch, Ronaldo da Silva F Jr, Luiz G P de Almeida, Cynthia C Cardoso, Otavio Bustrolini, Alexandra L Gerber, Ana Paula de C Guimarães, Diana Mariani, Andréa Cony Cavalcanti, Claudia dos Santos Rodrigues, Terezinha M P P Castiñeira, Amilcar Tanuri, Ana Tereza R de Vasconcelos |
| EPI_ISL_717899, EPI_ISL_717900, EPI_ISL_717901, EPI_ISL_717902, EPI_ISL_717903, EPI_ISL_717904, EPI_ISL_717905, EPI_ISL_717906, EPI_ISL_717907, EPI_ISL_717908, EPI_ISL_717909 | | | |
| see above | LACEN RJ - Noel Nutels | Bioinformatics Laboratory / LNCC | Carolina M Voloch, Ronaldo da Silva F Jr, Luiz G P de Almeida, Cynthia C Cardoso, Otavio Bustrolini, Alexandra L Gerber, Ana Paula de C Guimarães, Diana Mariani, Andréa Cony Cavalcanti, Claudia dos Santos Rodrigues, Terezinha M P P Castiñeira, Amilcar Tanuri, Ana Tereza R de Vasconcelos |
| EPI_ISL_717910, EPI_ISL_717911, | LACEN Dr. Francisco Rimolo Neto | Bioinformatics Laboratory / LNCC | Carolina M Voloch, Ronaldo da Silva F Jr, Luiz G P de Almeida, Cynthia C Cardoso, Otavio Bustrolini, Alexandra L Gerber, Ana Paula de C Guimarães, |

| | | | |
|---|---|---|---|
| EPI_ISL_717912, EPI_ISL_717913, EPI_ISL_717914, EPI_ISL_717915, EPI_ISL_717916, EPI_ISL_717917, EPI_ISL_717918, EPI_ISL_717919 | | | Diana Mariani, Andréa Cony Cavalcanti, Claudia dos Santos Rodrigues, Terezinha M P P Castiñeira, Amilcar Tanuri, Ana Tereza R de Vasconcelos |
| EPI_ISL_717920, EPI_ISL_717921, EPI_ISL_717922, EPI_ISL_717923, EPI_ISL_717924, EPI_ISL_717925, EPI_ISL_717926, EPI_ISL_717927, EPI_ISL_717928, EPI_ISL_717929, EPI_ISL_717930, EPI_ISL_717931, EPI_ISL_717932, EPI_ISL_717933, EPI_ISL_717934, EPI_ISL_717935, EPI_ISL_717936, EPI_ISL_717937, EPI_ISL_717938, EPI_ISL_717939, EPI_ISL_717940, EPI_ISL_717941, EPI_ISL_717942, EPI_ISL_717943, EPI_ISL_717944, EPI_ISL_717945, EPI_ISL_717946, EPI_ISL_717947, EPI_ISL_717948, EPI_ISL_717949, EPI_ISL_717950, EPI_ISL_717951, EPI_ISL_717952, EPI_ISL_717953, EPI_ISL_717954, EPI_ISL_717955, EPI_ISL_717956, EPI_ISL_717957 | | | |
| see above | Laboratorio de Virologia Molecular / UFRJ | Bioinformatics Laboratory / LNCC | Carolina M Voloch, Ronaldo da Silva F Jr, Luiz G P de Almeida, Cynthia C Cardoso, Otavio Bustrolini, Alexandra L Gerber, Ana Paula de C Guimarães, Diana Mariani, Andréa Cony Cavalcanti, Claudia dos Santos Rodrigues, Terezinha M P P Castiñeira, Amilcar Tanuri, Ana Tereza R de Vasconcelos |
| EPI_ISL_717958 | LACEN Dr. Francisco Rimolo Neto | Bioinformatics Laboratory / LNCC | Carolina M Voloch, Ronaldo da Silva F Jr, Luiz G P de Almeida, Cynthia C Cardoso, Otavio Bustrolini, Alexandra L Gerber, Ana Paula de C Guimarães, Diana Mariani, Andréa Cony Cavalcanti, Claudia dos Santos Rodrigues, Terezinha M P P Castiñeira, Amilcar Tanuri, Ana Tereza R de Vasconcelos |
| EPI_ISL_717959, EPI_ISL_717960, EPI_ISL_717961 | Laboratorio de Virologia Molecular / UFRJ | Bioinformatics Laboratory / LNCC | Carolina M Voloch, Ronaldo da Silva F Jr, Luiz G P de Almeida, Cynthia C Cardoso, Otavio Bustrolini, Alexandra L Gerber, Ana Paula de C Guimarães, Diana Mariani, Andréa Cony Cavalcanti, Claudia dos Santos Rodrigues, Terezinha M P P Castiñeira, Amilcar Tanuri, Ana Tereza R de Vasconcelos |
| EPI_ISL_717962 | LACEN RJ - Noel Nutels | Bioinformatics Laboratory / LNCC | Carolina M Voloch, Ronaldo da Silva F Jr, Luiz G P de Almeida, Cynthia C Cardoso, Otavio Bustrolini, Alexandra L Gerber, Ana Paula de C Guimarães, Diana Mariani, Andréa Cony Cavalcanti, Claudia dos Santos Rodrigues, Terezinha M P P Castiñeira, Amilcar Tanuri, Ana Tereza R de Vasconcelos |
| EPI_ISL_717963, EPI_ISL_717964 | LACEN Dr. Francisco Rimolo Neto | Bioinformatics Laboratory / LNCC | Carolina M Voloch, Ronaldo da Silva F Jr, Luiz G P de Almeida, Cynthia C Cardoso, Otavio Bustrolini, Alexandra L Gerber, Ana Paula de C Guimarães, Diana Mariani, Andréa Cony Cavalcanti, Claudia dos Santos Rodrigues, Terezinha M P P Castiñeira, Amilcar Tanuri, Ana Tereza R de Vasconcelos |
| EPI_ISL_983865 | Central Laboratory of Public Health of Rio Grande do Sul (Lacen-RS) | State Center for Health Surveillance of the Health Department of the State of Rio Grande do Sul (CEVS/SES-RS) | Aline Campos, Cynthia Molina, Lara Crescente, Leticia Garay, Ludmila Fiorenzano Baethgen, Richard Salvato, Tatiana Gregianini |

We gratefully acknowledge the following Authors from the Originating laboratories responsible for obtaining the specimens, as well as the Submitting laboratories where the genome data were generated and shared via GISAID, on which this research is based.

All Submitters of data may be contacted directly via www.gisaid.org

Authors are sorted alphabetically.

| Accession ID | Originating Laboratory | Submitting Laboratory | Authors |
|---|---|---|---|
| EPI_ISL_1023841 | Centro de Desenvolvimento Tecnológico em Saúde - CDTS | Centro de Desenvolvimento Tecnológico em Saúde - CDTS | Souza,T.M., Fintelman-Rodrigues,N., De Paula,A.D., Saraiva,F.B., Ferreira,M.A. and Sacramento,C.Q. |
| EPI_ISL_1133266, EPI_ISL_1133268 | Laboratório Hermes Pardini | Laboratório de Biologia Integrativa, Instituto de Ciências Biológicas, Universidade Federal de Minas Gerais | Filipe Romero Rebello Moreira, Diego Menezes Bonfim, Danielle Alves Gomes Zauli, Joice do Prado Silva, Aline Brito de Lima, Frederico Scott Varella Malta, Alessandro Clayton de Souza Ferreira, Victor Cavalcanti Pardini, Daniel Costa Queiroz, Rafael Marques de Souza, Victor Emmanuel Viana Geddes, Walyson Coelho Costa, Wagner Carlos Santos Magalhaes, Rennan Garcias Moreira, Carolina Moreira Voloch, Renan Pedra de Souza, Renato Santana Aguiar. |
| EPI_ISL_1181360, EPI_ISL_1181361, EPI_ISL_1181363, EPI_ISL_1181367, EPI_ISL_1181369, EPI_ISL_1181381, EPI_ISL_1181382, EPI_ISL_1181383, EPI_ISL_1181384, EPI_ISL_1181385, EPI_ISL_1181386, EPI_ISL_1181387, EPI_ISL_1181388, EPI_ISL_1181389, EPI_ISL_1181391, EPI_ISL_1181393, EPI_ISL_1181398, EPI_ISL_1181410, EPI_ISL_1181415 | | | |
| see above | Laboratory of Respiratory Viruses and Measles, Oswaldo Cruz Institute, FIOCRUZ | Laboratory of Respiratory Viruses and Measles, Oswaldo Cruz Institute, FIOCRUZ | Paola Resende, Luciana Appolinario, Fernando Motta, Anna Carolina Paixao, Ana Carolina Mendonca, Alice Sampaio Rocha, Renata Serrano Lopes, Marilda Siqueira on behalf of the Fiocruz COVID-19 Genomic Surveillance Network |
| EPI_ISL_1181425 | Laboratorio Central de Saude Publica do Estado do Rio de Janeiro (LACEN-RJ) | Laboratory of Respiratory Viruses and Measles, Oswaldo Cruz Institute, FIOCRUZ | Paola Resende, Luciana Appolinario, Fernando Motta, Anna Carolina Paixao, Ana Carolina Mendonca, Alice Sampaio Rocha, Renata Serrano Lopes, Andrea Cony Cavalcanti, Marilda Siqueira on behalf of the Fiocruz COVID-19 Genomic Surveillance Network |
| EPI_ISL_1181430 | Laboratory of Respiratory Viruses and Measles, Oswaldo Cruz Institute, FIOCRUZ | Laboratory of Respiratory Viruses and Measles, Oswaldo Cruz Institute, FIOCRUZ | Paola Resende, Luciana Appolinario, Fernando Motta, Anna Carolina Paixao, Ana Carolina Mendonca, Alice Sampaio Rocha, Renata Serrano Lopes, Marilda Siqueira on behalf of the Fiocruz COVID-19 Genomic Surveillance Network |
| EPI_ISL_1181431 | Laboratorio Central de Saude Publica do Estado do Rio de Janeiro (LACEN-RJ) | Laboratory of Respiratory Viruses and Measles, Oswaldo Cruz Institute, FIOCRUZ | Paola Resende, Luciana Appolinario, Fernando Motta, Anna Carolina Paixao, Ana Carolina Mendonca, Alice Sampaio Rocha, Renata Serrano Lopes, Andrea Cony Cavalcanti, Marilda Siqueira on behalf of the Fiocruz COVID-19 Genomic Surveillance Network |
| EPI_ISL_1181437, EPI_ISL_1181438, EPI_ISL_1181439, EPI_ISL_1181442, EPI_ISL_1181446, EPI_ISL_1181447, EPI_ISL_1181448, EPI_ISL_1181454, EPI_ISL_1181455, EPI_ISL_1181456, EPI_ISL_1181457, EPI_ISL_1181458, EPI_ISL_1181459, EPI_ISL_1181460, EPI_ISL_1181461, EPI_ISL_1181462, EPI_ISL_1181463, EPI_ISL_1181464, EPI_ISL_1181465, EPI_ISL_1181466, EPI_ISL_1181467, EPI_ISL_1181470, EPI_ISL_1181475, EPI_ISL_1181476, EPI_ISL_1181477, EPI_ISL_1181481, EPI_ISL_1181482, EPI_ISL_1181483, EPI_ISL_1181486, EPI_ISL_1181488, EPI_ISL_1181489, EPI_ISL_1181490, EPI_ISL_1181491, EPI_ISL_1181493, EPI_ISL_1181494, EPI_ISL_1181495, EPI_ISL_1181496, EPI_ISL_1181497, EPI_ISL_1181499, EPI_ISL_1181500, EPI_ISL_1181501, EPI_ISL_1181502, EPI_ISL_1181503, EPI_ISL_1181504, EPI_ISL_1181505, EPI_ISL_1181506, EPI_ISL_1181507, EPI_ISL_1181508, EPI_ISL_1181509, EPI_ISL_1181510, EPI_ISL_1181511, EPI_ISL_1181512, EPI_ISL_1181513, EPI_ISL_1181514, EPI_ISL_1181515, EPI_ISL_1181518, EPI_ISL_1181519, EPI_ISL_1181521, EPI_ISL_1181523, EPI_ISL_1181529, EPI_ISL_1181531, EPI_ISL_1181560, EPI_ISL_1181561, EPI_ISL_1181562, EPI_ISL_1181571, EPI_ISL_1181572, EPI_ISL_1181573, EPI_ISL_1181574, EPI_ISL_1181578, EPI_ISL_1181579, EPI_ISL_1181587, EPI_ISL_1181589, EPI_ISL_1181594, EPI_ISL_1181597, EPI_ISL_1181599 | | | |
| see above | Laboratory of Respiratory Viruses and Measles, Oswaldo Cruz Institute, FIOCRUZ | Laboratory of Respiratory Viruses and Measles, Oswaldo Cruz Institute, FIOCRUZ | Paola Resende, Luciana Appolinario, Fernando Motta, Anna Carolina Paixao, Ana Carolina Mendonca, Alice Sampaio Rocha, Renata Serrano Lopes, Marilda Siqueira on behalf of the Fiocruz COVID-19 Genomic Surveillance Network |
| EPI_ISL_1181600, EPI_ISL_1181601, EPI_ISL_1181602, EPI_ISL_1181603, EPI_ISL_1181606, EPI_ISL_1181607 | Laboratorio Central de Saude Publica do Estado do Rio de Janeiro (LACEN-RJ) | Laboratory of Respiratory Viruses and Measles, Oswaldo Cruz Institute, FIOCRUZ | Paola Resende, Luciana Appolinario, Fernando Motta, Anna Carolina Paixao, Ana Carolina Mendonca, Alice Sampaio Rocha, Renata Serrano Lopes, Andrea Cony Cavalcanti, Marilda Siqueira on behalf of the Fiocruz COVID-19 Genomic Surveillance Network |
| EPI_ISL_1181608 | Laboratory of Respiratory Viruses and Measles, Oswaldo Cruz Institute, FIOCRUZ | Laboratory of Respiratory Viruses and Measles, Oswaldo Cruz Institute, FIOCRUZ | Paola Resende, Luciana Appolinario, Fernando Motta, Anna Carolina Paixao, Ana Carolina Mendonca, Alice Sampaio Rocha, Renata Serrano Lopes, Marilda Siqueira on behalf of the Fiocruz COVID-19 Genomic Surveillance Network |
| EPI_ISL_1181611, EPI_ISL_1181612, EPI_ISL_1181613 | Laboratorio Central de Saude Publica do Estado do Rio de Janeiro (LACEN-RJ) | Laboratory of Respiratory Viruses and Measles, Oswaldo Cruz Institute, FIOCRUZ | Paola Resende, Luciana Appolinario, Fernando Motta, Anna Carolina Paixao, Ana Carolina Mendonca, Alice Sampaio Rocha, Renata Serrano Lopes, Andrea Cony Cavalcanti, Marilda Siqueira on behalf of the Fiocruz COVID-19 Genomic Surveillance Network |
| EPI_ISL_1182552, EPI_ISL_1182553, EPI_ISL_1182554, EPI_ISL_1182556, EPI_ISL_1182557, EPI_ISL_1182558 | Laboratório Central do Estado do Rio de Janeiro | Coordenação Geral de Laboratórios de Saúde Pública (CGLAB/DAEVS/SVS/MS) | Vagner Fonseca, et al. |
| EPI_ISL_1213163, EPI_ISL_1213171, EPI_ISL_1213248, EPI_ISL_1213249, EPI_ISL_1213270, EPI_ISL_1213272, EPI_ISL_1213307, EPI_ISL_1213309, EPI_ISL_1213310, EPI_ISL_1213312, EPI_ISL_1213314, EPI_ISL_1213339, EPI_ISL_1213340, EPI_ISL_1213372, EPI_ISL_1213374, EPI_ISL_1213376, EPI_ISL_1213378, EPI_ISL_1213379, EPI_ISL_1213381, EPI_ISL_1213382, EPI_ISL_1213393, EPI_ISL_1213395, EPI_ISL_1213397, EPI_ISL_1213408, EPI_ISL_1213410, EPI_ISL_1213417, EPI_ISL_1213418, EPI_ISL_1213420, EPI_ISL_1213422, EPI_ISL_1213424, EPI_ISL_1213425 | | | |
| see above | Laboratório HLA/UERJ | Bioinformatics Laboratory / LNCC | Alessandra P Lamarca, Luiz G P de Almeida, Ronaldo da Silva Francisco Jr, Lucymara Fassarella Agnez Lima, Kátia Castanho Scortecci, Vinícius Pietta Perez, Otavio J. Brustolini, Eduardo Sérgio Soares Sousa, Danielle Angst Secco, Angela Maria Guimarães Santos, George Rego Albuquerque, Ana Paula Melo Mariano, Bianca Mendes Maciel, Alexandra L Gerber, Ana Paula de C Guimarães, Paulo Ricardo Nascimento, Francisco Paulo Freire Neto, Sandra Rocha Gadelha, Luís Cristóvão Porto, Eloiza Helena Campana, Selma Maria Bezerra Jeronimo, Ana Tereza R Vasconcelos |
| EPI_ISL_1239111, EPI_ISL_1239112, EPI_ISL_1239113 | Laboratório Central de Saúde Pública Noel Nutels | Coordenação Geral de Laboratórios de Saúde Pública (CGLAB) | Vagner Fonseca et al, |
| EPI_ISL_1324142, EPI_ISL_1324145 | UW Virology Lab | UW Virology Lab | Pavitra Roychoudhury, Hong Xie, Lasata Shrestha, Shah Mohamed Bakhash, Michelle Lin, Margaret Mills, Noah Baker, Sean Ellis, Saraswathi Sathees, Meei-Li Huang, Keith R Jerome, Alexander Greninger |
| EPI_ISL_1402430 | Laboratory of Respiratory Viruses and Measles, Oswaldo Cruz Institute, FIOCRUZ | Laboratory of Respiratory Viruses and Measles, Oswaldo Cruz Institute, FIOCRUZ | Paola Resende, Alex Pauvolid-Correa, Mia Ferreira Araujo, Ana Beatriz Machado Lima, Luciana Appolinario, Fernando Motta, Anna Carolina Paixao, Ana Carolina Mendonca, Alice Sampaio Rocha, Renata Serrano Lopes, Marilda Siqueira on behalf of the Fiocruz COVID-19 Genomic Surveillance Network |
| EPI_ISL_1533977 | Laboratorio de Virologia Molecular / UFRJ | Laboratory of Respiratory Viruses and Measles, Oswaldo Cruz Institute, FIOCRUZ | Paola Resende, Carolina M Voloch, Luciana Appolinario, Fernando Motta, Anna Carolina Paixao, Ana Carolina Mendonca, Alice Sampaio Rocha, Renata Serrano Lopes, Amilcar Tanuri, Marilda Siqueira on behalf of the Fiocruz COVID-19 Genomic Surveillance Network |
| EPI_ISL_1533980, EPI_ISL_1533983 | Laboratory of Respiratory Viruses and Measles, Oswaldo Cruz Institute, FIOCRUZ | Laboratory of Respiratory Viruses and Measles, Oswaldo Cruz Institute, FIOCRUZ | Paola Resende, Luciana Appolinario, Fernando Motta, Anna Carolina Paixao, Ana Carolina Mendonca, Alice Sampaio Rocha, Renata Serrano Lopes, Marilda Siqueira on behalf of the Fiocruz COVID-19 Genomic Surveillance Network |
| EPI_ISL_1533991, EPI_ISL_1533995, EPI_ISL_1533998 | Laboratorio de Virologia Molecular / UFRJ | Laboratory of Respiratory Viruses and Measles, Oswaldo Cruz Institute, FIOCRUZ | Paola Resende, Carolina M Voloch, Luciana Appolinario, Fernando Motta, Anna Carolina Paixao, Ana Carolina Mendonca, Alice Sampaio Rocha, Renata Serrano Lopes, Amilcar Tanuri, Marilda Siqueira on behalf of the Fiocruz COVID-19 Genomic Surveillance Network |
| EPI_ISL_1534005, EPI_ISL_1534011 | Laboratory of Respiratory Viruses and Measles, Oswaldo Cruz Institute, FIOCRUZ | Laboratory of Respiratory Viruses and Measles, Oswaldo Cruz Institute, FIOCRUZ | Paola Resende, Luciana Appolinario, Fernando Motta, Anna Carolina Paixao, Ana Carolina Mendonca, Alice Sampaio Rocha, Renata Serrano Lopes, Marilda Siqueira on behalf of the Fiocruz COVID-19 Genomic Surveillance Network |
| EPI_ISL_1534012 | Laboratorio Central de Saude Publica do Estado do Rio de Janeiro (LACEN-RJ) | Laboratory of Respiratory Viruses and Measles, Oswaldo Cruz Institute, FIOCRUZ | Paola Resende, Luciana Appolinario, Fernando Motta, Anna Carolina Paixao, Ana Carolina Mendonca, Alice Sampaio Rocha, Renata Serrano Lopes, Andrea Cony Cavalcanti, Marilda Siqueira on behalf of the Fiocruz COVID-19 Genomic Surveillance Network |
| EPI_ISL_1534014, EPI_ISL_1534015, EPI_ISL_1534016 | Laboratory of Respiratory Viruses and Measles, Oswaldo Cruz Institute, FIOCRUZ | Laboratory of Respiratory Viruses and Measles, Oswaldo Cruz Institute, FIOCRUZ | Paola Resende, Luciana Appolinario, Fernando Motta, Anna Carolina Paixao, Ana Carolina Mendonca, Alice Sampaio Rocha, Renata Serrano Lopes, Marilda Siqueira on behalf of the Fiocruz COVID-19 Genomic Surveillance Network |
| EPI_ISL_1592230 | Laboratório Hermes Pardini | Laboratório de Biologia Integrativa, Instituto de Ciências Biológicas, Universidade Federal de Minas Gerais | Filipe Romero Rebello Moreira, Diego Menezes Bonfim, Danielle Alves Gomes Zauli, Joice do Prado Silva, Aline Brito de Lima, Frederico Scott Varella Malta, Alessandro Clayton de Souza Ferreira, Victor Cavalcanti Pardini, Daniel Costa Queiroz, Rafael Marques de Souza, Victor Emmanuel Viana Geddes, Walyson Coelho Costa, Wagner Carlos Santos Magalhaes, Rennan Garcias Moreira, Carolina Moreira Voloch, Renan Pedra de Souza, Renato Santana Aguiar. |
| EPI_ISL_1664113, EPI_ISL_1664114, EPI_ISL_1664115, EPI_ISL_1664116, EPI_ISL_1664117, EPI_ISL_1664118, EPI_ISL_1664119, EPI_ISL_1664120, EPI_ISL_1664121, EPI_ISL_1664122, EPI_ISL_1664123, EPI_ISL_1664124, EPI_ISL_1664125, EPI_ISL_1664126, EPI_ISL_1664127, EPI_ISL_1664128, EPI_ISL_1664129, EPI_ISL_1664130, EPI_ISL_1664131, EPI_ISL_1664132, EPI_ISL_1664133, EPI_ISL_1664134, EPI_ISL_1664135, EPI_ISL_1664136, EPI_ISL_1664137, EPI_ISL_1664138, EPI_ISL_1664139, EPI_ISL_1664140, EPI_ISL_1664141, EPI_ISL_1664142, EPI_ISL_1664143, EPI_ISL_1664144, EPI_ISL_1664145, EPI_ISL_1664146, EPI_ISL_1664147, EPI_ISL_1664148, EPI_ISL_1664149, EPI_ISL_1664150, EPI_ISL_1664151, EPI_ISL_1664152, EPI_ISL_1664153, EPI_ISL_1664154, EPI_ISL_1664155, EPI_ISL_1664156, EPI_ISL_1664157, EPI_ISL_1664158, EPI_ISL_1664159, EPI_ISL_1664160, EPI_ISL_1664161, EPI_ISL_1664162, EPI_ISL_1664163, | | | |

| | | | |
|---|---|---|---|
| EPI_ISL_1664164, EPI_ISL_1664165, EPI_ISL_1664166, EPI_ISL_1664167, EPI_ISL_1664168, EPI_ISL_1664169, EPI_ISL_1664170, EPI_ISL_1664171, EPI_ISL_1664172, EPI_ISL_1664173, EPI_ISL_1664174, EPI_ISL_1664175, EPI_ISL_1664176, EPI_ISL_1664177, EPI_ISL_1664178, EPI_ISL_1664179, EPI_ISL_1664180, EPI_ISL_1664181, EPI_ISL_1664182, EPI_ISL_1664183, EPI_ISL_1664184, EPI_ISL_1664185, EPI_ISL_1664186, EPI_ISL_1664187, EPI_ISL_1664188, EPI_ISL_1664189, EPI_ISL_1664190, EPI_ISL_1664191, EPI_ISL_1664192, EPI_ISL_1664193, EPI_ISL_1664194, EPI_ISL_1664195, EPI_ISL_1664196, EPI_ISL_1664197, EPI_ISL_1664198, EPI_ISL_1664199, EPI_ISL_1664200, EPI_ISL_1664201, EPI_ISL_1664202 | | | |
| see above | Laboratorio Central Noel Nutels | Bioinformatics Laboratory / LNCC | Luiz G P de Almeida, Alessandra P Lamarca, Ronaldo da Silva F Jr, Liliane Cavalcante, Alexandra L Gerber, Ana Paula de C Guimarães, Douglas Terra Machado, Cassia Alves, Diana Mariani, Thais Felix Cruz, Mario Sergio Ribeiro, Silvia Carvalho, Flávio Dias da Silva, Marcio Henrique de Oliveira Garcia, Leandro Magalhães de Souza, Cristiane Gomes da Silva, Caio Luiz Pereira Ribeiro, Andréa Cony Cavalcanti, Claudia Maria Braga de Mello, Amilcar Tanuri, Ana Tereza R Vasconcelos |
| EPI_ISL_1678584, EPI_ISL_1690551, EPI_ISL_1694627, EPI_ISL_1694628, EPI_ISL_1694631, EPI_ISL_1694632, EPI_ISL_1694634, EPI_ISL_1695909 | Instituto Estadual do Cérebro Paulo Niemayer (IECPN) | Laboratório de Imunofarmacologia | Thiago Moreno Lopes Souza |
| EPI_ISL_1858267, EPI_ISL_1858269, EPI_ISL_1858271, EPI_ISL_1858273, EPI_ISL_1858275, EPI_ISL_1858277, EPI_ISL_1858279, EPI_ISL_1858281, EPI_ISL_1858282, EPI_ISL_1858284, EPI_ISL_1858286, EPI_ISL_1858288, EPI_ISL_1858290, EPI_ISL_1858292, EPI_ISL_1858294, EPI_ISL_1858295, EPI_ISL_1858297, EPI_ISL_1858299, EPI_ISL_1858301, EPI_ISL_1858303, EPI_ISL_1858304, EPI_ISL_1858306, EPI_ISL_1858308, EPI_ISL_1858310, EPI_ISL_1858312, EPI_ISL_1858314, EPI_ISL_1858315, EPI_ISL_1858317, EPI_ISL_1858319, EPI_ISL_1858321, EPI_ISL_1858323, EPI_ISL_1858325, EPI_ISL_1858327, EPI_ISL_1858329, EPI_ISL_1858331, EPI_ISL_1858333, EPI_ISL_1858334, EPI_ISL_1858336, EPI_ISL_1858338, EPI_ISL_1858340, EPI_ISL_1858342, EPI_ISL_1858344, EPI_ISL_1858346, EPI_ISL_1858348, EPI_ISL_1858350, EPI_ISL_1858352, EPI_ISL_1858354, EPI_ISL_1858356, EPI_ISL_1858357, EPI_ISL_1858359, EPI_ISL_1858361, EPI_ISL_1858363, EPI_ISL_1858365, EPI_ISL_1858367, EPI_ISL_1858369, EPI_ISL_1858371, EPI_ISL_1858373, EPI_ISL_1858375, EPI_ISL_1858377, EPI_ISL_1858379, EPI_ISL_1858380, EPI_ISL_1858383, EPI_ISL_1858384, EPI_ISL_1858387, EPI_ISL_1858389, EPI_ISL_1858391, EPI_ISL_1858392, EPI_ISL_1858395, EPI_ISL_1858397, EPI_ISL_1858399, EPI_ISL_1858401, EPI_ISL_1858403, EPI_ISL_1858409, EPI_ISL_1858411, EPI_ISL_1858414, EPI_ISL_1858416, EPI_ISL_1858418, EPI_ISL_1858420, EPI_ISL_1858422, EPI_ISL_1858423, EPI_ISL_1858426, EPI_ISL_1858428, EPI_ISL_1858430, EPI_ISL_1858432, EPI_ISL_1858434, EPI_ISL_1858436, EPI_ISL_1858438, EPI_ISL_1858440, EPI_ISL_1858442, EPI_ISL_1858444, EPI_ISL_1858446, EPI_ISL_1858449, EPI_ISL_1858451, EPI_ISL_1858453, EPI_ISL_1858454, EPI_ISL_1858456, EPI_ISL_1858458, EPI_ISL_1858460, EPI_ISL_1858462, EPI_ISL_1858464, EPI_ISL_1858466, EPI_ISL_1858468, EPI_ISL_1858470, EPI_ISL_1858473, EPI_ISL_1858475, EPI_ISL_1858477, EPI_ISL_1858479, EPI_ISL_1858481, EPI_ISL_1858483, EPI_ISL_1858486, EPI_ISL_1858488, EPI_ISL_1858490, EPI_ISL_1858492, EPI_ISL_1858494, EPI_ISL_1858496, EPI_ISL_1858498, EPI_ISL_1858500, EPI_ISL_1858502, EPI_ISL_1858504, EPI_ISL_1858506, EPI_ISL_1858508, EPI_ISL_1858512, EPI_ISL_1858514, EPI_ISL_1858516, EPI_ISL_1858518, EPI_ISL_1858520, EPI_ISL_1858522, EPI_ISL_1858526, EPI_ISL_1858528, EPI_ISL_1858530, EPI_ISL_1858532, EPI_ISL_1858536, EPI_ISL_1858538, EPI_ISL_1858540, EPI_ISL_1858542, EPI_ISL_1858544, EPI_ISL_1858546, EPI_ISL_1858548, EPI_ISL_1858549, EPI_ISL_1858551, EPI_ISL_1858554, EPI_ISL_1858555, EPI_ISL_1858557, EPI_ISL_1858559, EPI_ISL_1858561, EPI_ISL_1858563, EPI_ISL_1858565, EPI_ISL_1858567, EPI_ISL_1858571, EPI_ISL_1858572, EPI_ISL_1858575, EPI_ISL_1858577, EPI_ISL_1858578, EPI_ISL_1858580, EPI_ISL_1858582, EPI_ISL_1858584, EPI_ISL_1858586, EPI_ISL_1858588, EPI_ISL_1858590, EPI_ISL_1858592, EPI_ISL_1858594, EPI_ISL_1858596, EPI_ISL_1858598, EPI_ISL_1858599, EPI_ISL_1858603, EPI_ISL_1858606, EPI_ISL_1858607, EPI_ISL_1858610, EPI_ISL_1858611, EPI_ISL_1858613, EPI_ISL_1858615, EPI_ISL_1858617, EPI_ISL_1858619, EPI_ISL_1858621, EPI_ISL_1858623, EPI_ISL_1858625, EPI_ISL_1858627, EPI_ISL_1858629, EPI_ISL_1858631, EPI_ISL_1858633, EPI_ISL_1858636, EPI_ISL_1858637, EPI_ISL_1858639, EPI_ISL_1858641, EPI_ISL_1858643, EPI_ISL_1858645, EPI_ISL_1858648, EPI_ISL_1858650, EPI_ISL_1858651, EPI_ISL_1858655, EPI_ISL_1858656, EPI_ISL_1858658, EPI_ISL_1858660, EPI_ISL_1858662, EPI_ISL_1858664, EPI_ISL_1858666, EPI_ISL_1858668, EPI_ISL_1858670, EPI_ISL_1858672, EPI_ISL_1858674, EPI_ISL_1858675, EPI_ISL_1858677, EPI_ISL_1858680, EPI_ISL_1858682, EPI_ISL_1858684, EPI_ISL_1858686, EPI_ISL_1858688, EPI_ISL_1858691, EPI_ISL_1858693, EPI_ISL_1858694, EPI_ISL_1858697, EPI_ISL_1858701, EPI_ISL_1858703, EPI_ISL_1858705, EPI_ISL_1858707, EPI_ISL_1858709, EPI_ISL_1858711, EPI_ISL_1858713, EPI_ISL_1858715, EPI_ISL_1858717, EPI_ISL_1858719, EPI_ISL_1858721, EPI_ISL_1858722, EPI_ISL_1858724, EPI_ISL_1858726, EPI_ISL_1858728, EPI_ISL_1858730, EPI_ISL_1858732, EPI_ISL_1858734, EPI_ISL_1858736, EPI_ISL_1858738, EPI_ISL_1858740, EPI_ISL_1858741, EPI_ISL_1858743, EPI_ISL_1858745, EPI_ISL_1858747, EPI_ISL_1858751, EPI_ISL_1858752, EPI_ISL_1858754, EPI_ISL_1858756 | | | |
| see above | Laboratorio Central Noel Nutels | Bioinformatics Laboratory / LNCC | Luiz G P de Almeida, Alessandra P Lamarca, Ronaldo da Silva F Jr, Liliane Cavalcante, Alexandra L Gerber, Ana Paula de C Guimarães, Douglas Terra Machado, Cassia Alves, Diana Mariani, Thais Felix Cruz, Mario Sergio Ribeiro, Silvia Carvalho, Flávio Dias da Silva, Marcio Henrique de Oliveira Garcia, Leandro Magalhães de Souza, Cristiane Gomes da Silva, Caio Luiz Pereira Ribeiro, Andréa Cony Cavalcanti, Claudia Maria Braga de Mello, Amilcar Tanuri, Ana Tereza R Vasconcelos |
| EPI_ISL_1858758, EPI_ISL_1858761, EPI_ISL_1858763, EPI_ISL_1858765, EPI_ISL_1858767, EPI_ISL_1858771, EPI_ISL_1858775, EPI_ISL_1858777, EPI_ISL_1858779, EPI_ISL_1858781, EPI_ISL_1858783, EPI_ISL_1858786, EPI_ISL_1858788, EPI_ISL_1858790, EPI_ISL_1858792, EPI_ISL_1858794, EPI_ISL_1858796, EPI_ISL_1858798, EPI_ISL_1858800, EPI_ISL_1858802, EPI_ISL_1858803, EPI_ISL_1858805, EPI_ISL_1858807, EPI_ISL_1858809, EPI_ISL_1858811, EPI_ISL_1858814, EPI_ISL_1858816, EPI_ISL_1858818, EPI_ISL_1858819, EPI_ISL_1858822, EPI_ISL_1858824, EPI_ISL_1858826, EPI_ISL_1858828, EPI_ISL_1858830, EPI_ISL_1858832, EPI_ISL_1858834, EPI_ISL_1858836, EPI_ISL_1858838, EPI_ISL_1858840, EPI_ISL_1858842, EPI_ISL_1858843, EPI_ISL_1858846, EPI_ISL_1858847, EPI_ISL_1858850, EPI_ISL_1858852, EPI_ISL_1858853, EPI_ISL_1858856, EPI_ISL_1858858, EPI_ISL_1858860, EPI_ISL_1858862, EPI_ISL_1858864, EPI_ISL_1858866, EPI_ISL_1858868, EPI_ISL_1858870, EPI_ISL_1858872, EPI_ISL_1858878, EPI_ISL_1858880, EPI_ISL_1858882, EPI_ISL_1858884, EPI_ISL_1858886, EPI_ISL_1858888, EPI_ISL_1858890, EPI_ISL_1858892, EPI_ISL_1858894, EPI_ISL_1858896, EPI_ISL_1858898, EPI_ISL_1858900, EPI_ISL_1858902, EPI_ISL_1858904, EPI_ISL_1858906, EPI_ISL_1858908, EPI_ISL_1858910, EPI_ISL_1858912, EPI_ISL_1858914, EPI_ISL_1858916, EPI_ISL_1858918, EPI_ISL_1858920, EPI_ISL_1858922, EPI_ISL_1858924, EPI_ISL_1858926, EPI_ISL_1858928, EPI_ISL_1858930, EPI_ISL_1858933, EPI_ISL_1858934, EPI_ISL_1858936, EPI_ISL_1858938, EPI_ISL_1858940, EPI_ISL_1858942, EPI_ISL_1858944, EPI_ISL_1858946, EPI_ISL_1858948, EPI_ISL_1858950, EPI_ISL_1858952, EPI_ISL_1858954, EPI_ISL_1858957, EPI_ISL_1858960, EPI_ISL_1858963, EPI_ISL_1858964, EPI_ISL_1858966, EPI_ISL_1858968, EPI_ISL_1858969, EPI_ISL_1858970, EPI_ISL_1858971, EPI_ISL_1858973, EPI_ISL_1858975, EPI_ISL_1858977, EPI_ISL_1858979, EPI_ISL_1858981, EPI_ISL_1858984, EPI_ISL_1858986, EPI_ISL_1858988, EPI_ISL_1858990, EPI_ISL_1858992, EPI_ISL_1858994, EPI_ISL_1858996, EPI_ISL_1858998, EPI_ISL_1859000, EPI_ISL_1859002, EPI_ISL_1859005, EPI_ISL_1859007, EPI_ISL_1859008 | | | |
| see above | Unidade de apoio ao diagnóstico da COVID - UNADIG | Bioinformatics Laboratory / LNCC | Luiz G P de Almeida, Alessandra P Lamarca, Ronaldo da Silva F Jr, Liliane Cavalcante, Alexandra L Gerber, Ana Paula de C Guimarães, Douglas Terra Machado, Cassia Alves, Diana Mariani, Thais Felix Cruz, Mario Sergio Ribeiro, Silvia Carvalho, Flávio Dias da Silva, Marcio Henrique de Oliveira Garcia, Leandro Magalhães de Souza, Cristiane Gomes da Silva, Caio Luiz Pereira Ribeiro, Andréa Cony Cavalcanti, Claudia Maria Braga de Mello, Amilcar Tanuri, Ana Tereza R Vasconcelos |
| EPI_ISL_2038956 | Laboratorio Central de Saude Publica do Estado do Rio de Janeiro (LACEN-RJ) | Laboratory of Respiratory Viruses and Measles, Oswaldo Cruz Institute, FIOCRUZ | Paola Resende, Luciana Appolinario, Fernando Motta, Anna Carolina Paixao, Ana Carolina Mendonca, Alice Sampaio Rocha, Taina Venas, Elisa Cavalcante Pereira, Renata Serrano Lopes, Andrea Cony Cavalcanti, Marilda Siqueira on behalf of the Fiocruz COVID-19 Genomic Surveillance Network |
| EPI_ISL_2038961, EPI_ISL_2038962, EPI_ISL_2038963 | Laboratory of Respiratory Viruses and Measles, Oswaldo Cruz Institute, FIOCRUZ | Laboratory of Respiratory Viruses and Measles, Oswaldo Cruz Institute, FIOCRUZ | Paola Resende, Luciana Appolinario, Fernando Motta, Anna Carolina Paixao, Ana Carolina Mendonca, Alice Sampaio Rocha, Taina Venas, Elisa Cavalcante Pereira, Renata Serrano Lopes, Marilda Siqueira on behalf of the Fiocruz COVID-19 Genomic Surveillance Network |
| EPI_ISL_2086268 | Laboratory of Molecular Biology of Flavivirus, Oswaldo Cruz Institute, FIOCRUZ | Laboratory of Molecular Biology of Flavivirus, Oswaldo Cruz Institute, FIOCRUZ | Fernando Lopez Tort, Ieda Ribeiro, Lidiane Menezes, Alexandre dos Santos, Marta Pereira, Myrna Bonaldo |
| EPI_ISL_2101392 | Unidade de apoio ao diagnóstico da COVID - UNADIG | Bioinformatics Laboratory / LNCC | Luiz G P de Almeida, Alessandra P Lamarca, Ronaldo da Silva F Jr, Liliane Cavalcante, Alexandra L Gerber, Ana Paula de C Guimaraes, Douglas Terra Machado, Cassia Alves, Diana Mariani, Cintia Policarpo, Gleidson da Silva de Oliveira, Mario Sergio Ribeiro, Silvia Carvalho, Flavio Dias da Silva, Marcio Henrique de Oliveira Garcia, Leandro Magalhaes de Souza, Cristiane Gomes da Silva, Caio Luiz Pereira Ribeiro, Andrea Cony Cavalcanti, Claudia Maria Braga de Mello, Amilcar Tanuri, Ana Tereza R Vasconcelos |
| EPI_ISL_2101393 | Laboratorio Central Noel Nutels | Bioinformatics Laboratory / LNCC | Luiz G P de Almeida, Alessandra P Lamarca, Ronaldo da Silva F Jr, Liliane Cavalcante, Alexandra L Gerber, Ana Paula de C Guimaraes, Douglas Terra Machado, Cassia Alves, Diana Mariani, Cintia Policarpo, Gleidson da Silva de Oliveira, Mario Sergio Ribeiro, Silvia Carvalho, Flavio Dias da Silva, Marcio Henrique de Oliveira Garcia, Leandro Magalhaes de Souza, Cristiane Gomes da Silva, Caio Luiz Pereira Ribeiro, Andrea Cony Cavalcanti, Claudia Maria Braga de Mello, Amilcar Tanuri, Ana Tereza R Vasconcelos |
| EPI_ISL_2101394 | Unidade de apoio ao diagnóstico da COVID - UNADIG | Bioinformatics Laboratory / LNCC | Luiz G P de Almeida, Alessandra P Lamarca, Ronaldo da Silva F Jr, Liliane Cavalcante, Alexandra L Gerber, Ana Paula de C Guimaraes, Douglas Terra Machado, Cassia Alves, Diana Mariani, Cintia Policarpo, Gleidson da Silva de Oliveira, Mario Sergio Ribeiro, Silvia Carvalho, Flavio Dias da Silva, Marcio Henrique de Oliveira Garcia, Leandro Magalhaes de Souza, Cristiane Gomes da Silva, Caio Luiz Pereira Ribeiro, Andrea Cony Cavalcanti, Claudia Maria Braga de Mello, Amilcar Tanuri, Ana Tereza R Vasconcelos |
| EPI_ISL_2101395, EPI_ISL_2101396, EPI_ISL_2101397, EPI_ISL_2101398, EPI_ISL_2101399, EPI_ISL_2101400, EPI_ISL_2101401, EPI_ISL_2101403, EPI_ISL_2101404, EPI_ISL_2101405, EPI_ISL_2101406, EPI_ISL_2101410, EPI_ISL_2101411, EPI_ISL_2101412, EPI_ISL_2101413, EPI_ISL_2101414, EPI_ISL_2101415, EPI_ISL_2101416, EPI_ISL_2101417, EPI_ISL_2101418, EPI_ISL_2101419, EPI_ISL_2101420, EPI_ISL_2101421, EPI_ISL_2101422, EPI_ISL_2101423, EPI_ISL_2101424, EPI_ISL_2101425, EPI_ISL_2101426, EPI_ISL_2101427, EPI_ISL_2101428 | | | |
| see above | Laboratorio Central Noel Nutels | Bioinformatics Laboratory / LNCC | Luiz G P de Almeida, Alessandra P Lamarca, Ronaldo da Silva F Jr, Liliane Cavalcante, Alexandra L Gerber, Ana Paula de C Guimaraes, Douglas Terra Machado, Cassia Alves, Diana Mariani, Cintia Policarpo, Gleidson da Silva de Oliveira, Mario Sergio Ribeiro, Silvia Carvalho, Flavio Dias da Silva, Marcio Henrique de Oliveira Garcia, Leandro Magalhaes de Souza, Cristiane Gomes da Silva, Caio Luiz Pereira Ribeiro, Andrea Cony Cavalcanti, Claudia Maria Braga de Mello, Amilcar Tanuri, Ana Tereza R Vasconcelos |
| EPI_ISL_2101429, EPI_ISL_2101430, EPI_ISL_2101431, EPI_ISL_2101432, EPI_ISL_2101433 | Unidade de apoio ao diagnóstico da COVID - UNADIG | Bioinformatics Laboratory / LNCC | Luiz G P de Almeida, Alessandra P Lamarca, Ronaldo da Silva F Jr, Liliane Cavalcante, Alexandra L Gerber, Ana Paula de C Guimaraes, Douglas Terra Machado, Cassia Alves, Diana Mariani, Cintia Policarpo, Gleidson da Silva de Oliveira, Mario Sergio Ribeiro, Silvia Carvalho, Flavio Dias da Silva, Marcio Henrique de Oliveira Garcia, Leandro Magalhaes de Souza, Cristiane Gomes da Silva, Caio Luiz Pereira Ribeiro, Andrea Cony Cavalcanti, Claudia Maria Braga de Mello, Amilcar Tanuri, Ana Tereza R Vasconcelos |
| EPI_ISL_2101434, EPI_ISL_2101435, EPI_ISL_2101436, EPI_ISL_2101437, EPI_ISL_2101438, EPI_ISL_2101439, EPI_ISL_2101440, EPI_ISL_2101441, EPI_ISL_2101442 | Laboratorio Central Noel Nutels | Bioinformatics Laboratory / LNCC | Luiz G P de Almeida, Alessandra P Lamarca, Ronaldo da Silva F Jr, Liliane Cavalcante, Alexandra L Gerber, Ana Paula de C Guimaraes, Douglas Terra Machado, Cassia Alves, Diana Mariani, Cintia Policarpo, Gleidson da Silva de Oliveira, Mario Sergio Ribeiro, Silvia Carvalho, Flavio Dias da Silva, Marcio Henrique de Oliveira Garcia, Leandro Magalhaes de Souza, Cristiane Gomes da Silva, Caio Luiz Pereira Ribeiro, Andrea Cony Cavalcanti, Claudia Maria Braga de Mello, Amilcar Tanuri, Ana Tereza R Vasconcelos |
| EPI_ISL_2101443 | Unidade de apoio ao diagnóstico da COVID - UNADIG | Bioinformatics Laboratory / LNCC | Luiz G P de Almeida, Alessandra P Lamarca, Ronaldo da Silva F Jr, Liliane Cavalcante, Alexandra L Gerber, Ana Paula de C Guimaraes, Douglas Terra Machado, Cassia Alves, Diana Mariani, Cintia Policarpo, Gleidson da Silva de Oliveira, Mario Sergio Ribeiro, Silvia Carvalho, Flavio Dias da Silva, Marcio Henrique de Oliveira Garcia, Leandro Magalhaes de Souza, Cristiane Gomes da Silva, Caio Luiz Pereira Ribeiro, Andrea Cony Cavalcanti, Claudia Maria Braga de Mello, Amilcar Tanuri, Ana Tereza R Vasconcelos |

| | | | |
|---|---|---|---|
| EPI_ISL_2101444, EPI_ISL_2101445, EPI_ISL_2101446, EPI_ISL_2101447, EPI_ISL_2101448, EPI_ISL_2101449, EPI_ISL_2101450, EPI_ISL_2101451, EPI_ISL_2101452, EPI_ISL_2101453, EPI_ISL_2101454 | | | |
| see above | Laboratorio Central Noel Nutels | Bioinformatics Laboratory / LNCC | Luiz G P de Almeida, Alessandra P Lamarca, Ronaldo da Silva F Jr, Liliane Cavalcante, Alexandra L Gerber, Ana Paula de C Guimaraes, Douglas Terra Machado, Cassia Alves, Diana Mariani, Cintia Policarpo, Gleidson da Silva de Oliveira, Mario Sergio Ribeiro, Silvia Carvalho, Flavio Dias da Silva, Marcio Henrique de Oliveira Garcia, Leandro Magalhaes de Souza, Cristiane Gomes da Silva, Caio Luiz Pereira Ribeiro, Andrea Cony Cavalcanti, Claudia Maria Braga de Mello, Amilcar Tanuri, Ana Tereza R Vasconcelos |
| EPI_ISL_2101455, EPI_ISL_2101456, EPI_ISL_2101457, EPI_ISL_2101458, EPI_ISL_2101459, EPI_ISL_2101460, EPI_ISL_2101461, EPI_ISL_2101462, EPI_ISL_2101463 | Unidade de apoio ao diagnóstico da COVID - UNADIG | Bioinformatics Laboratory / LNCC | Luiz G P de Almeida, Alessandra P Lamarca, Ronaldo da Silva F Jr, Liliane Cavalcante, Alexandra L Gerber, Ana Paula de C Guimaraes, Douglas Terra Machado, Cassia Alves, Diana Mariani, Cintia Policarpo, Gleidson da Silva de Oliveira, Mario Sergio Ribeiro, Silvia Carvalho, Flavio Dias da Silva, Marcio Henrique de Oliveira Garcia, Leandro Magalhaes de Souza, Cristiane Gomes da Silva, Caio Luiz Pereira Ribeiro, Andrea Cony Cavalcanti, Claudia Maria Braga de Mello, Amilcar Tanuri, Ana Tereza R Vasconcelos |
| EPI_ISL_2101464, EPI_ISL_2101465, EPI_ISL_2101466, EPI_ISL_2101467, EPI_ISL_2101468, EPI_ISL_2101469 | Laboratorio Central Noel Nutels | Bioinformatics Laboratory / LNCC | Luiz G P de Almeida, Alessandra P Lamarca, Ronaldo da Silva F Jr, Liliane Cavalcante, Alexandra L Gerber, Ana Paula de C Guimaraes, Douglas Terra Machado, Cassia Alves, Diana Mariani, Cintia Policarpo, Gleidson da Silva de Oliveira, Mario Sergio Ribeiro, Silvia Carvalho, Flavio Dias da Silva, Marcio Henrique de Oliveira Garcia, Leandro Magalhaes de Souza, Cristiane Gomes da Silva, Caio Luiz Pereira Ribeiro, Andrea Cony Cavalcanti, Claudia Maria Braga de Mello, Amilcar Tanuri, Ana Tereza R Vasconcelos |
| EPI_ISL_2101470, EPI_ISL_2101471, EPI_ISL_2101472, EPI_ISL_2101473 | Unidade de apoio ao diagnóstico da COVID - UNADIG | Bioinformatics Laboratory / LNCC | Luiz G P de Almeida, Alessandra P Lamarca, Ronaldo da Silva F Jr, Liliane Cavalcante, Alexandra L Gerber, Ana Paula de C Guimaraes, Douglas Terra Machado, Cassia Alves, Diana Mariani, Cintia Policarpo, Gleidson da Silva de Oliveira, Mario Sergio Ribeiro, Silvia Carvalho, Flavio Dias da Silva, Marcio Henrique de Oliveira Garcia, Leandro Magalhaes de Souza, Cristiane Gomes da Silva, Caio Luiz Pereira Ribeiro, Andrea Cony Cavalcanti, Claudia Maria Braga de Mello, Amilcar Tanuri, Ana Tereza R Vasconcelos |
| EPI_ISL_2101474 | Laboratorio Central Noel Nutels | Bioinformatics Laboratory / LNCC | Luiz G P de Almeida, Alessandra P Lamarca, Ronaldo da Silva F Jr, Liliane Cavalcante, Alexandra L Gerber, Ana Paula de C Guimaraes, Douglas Terra Machado, Cassia Alves, Diana Mariani, Cintia Policarpo, Gleidson da Silva de Oliveira, Mario Sergio Ribeiro, Silvia Carvalho, Flavio Dias da Silva, Marcio Henrique de Oliveira Garcia, Leandro Magalhaes de Souza, Cristiane Gomes da Silva, Caio Luiz Pereira Ribeiro, Andrea Cony Cavalcanti, Claudia Maria Braga de Mello, Amilcar Tanuri, Ana Tereza R Vasconcelos |
| EPI_ISL_2101475 | Unidade de apoio ao diagnóstico da COVID - UNADIG | Bioinformatics Laboratory / LNCC | Luiz G P de Almeida, Alessandra P Lamarca, Ronaldo da Silva F Jr, Liliane Cavalcante, Alexandra L Gerber, Ana Paula de C Guimaraes, Douglas Terra Machado, Cassia Alves, Diana Mariani, Cintia Policarpo, Gleidson da Silva de Oliveira, Mario Sergio Ribeiro, Silvia Carvalho, Flavio Dias da Silva, Marcio Henrique de Oliveira Garcia, Leandro Magalhaes de Souza, Cristiane Gomes da Silva, Caio Luiz Pereira Ribeiro, Andrea Cony Cavalcanti, Claudia Maria Braga de Mello, Amilcar Tanuri, Ana Tereza R Vasconcelos |
| EPI_ISL_2101476, EPI_ISL_2101477, EPI_ISL_2101478, EPI_ISL_2101479, EPI_ISL_2101480, EPI_ISL_2101481, EPI_ISL_2101482 | Laboratorio Central Noel Nutels | Bioinformatics Laboratory / LNCC | Luiz G P de Almeida, Alessandra P Lamarca, Ronaldo da Silva F Jr, Liliane Cavalcante, Alexandra L Gerber, Ana Paula de C Guimaraes, Douglas Terra Machado, Cassia Alves, Diana Mariani, Cintia Policarpo, Gleidson da Silva de Oliveira, Mario Sergio Ribeiro, Silvia Carvalho, Flavio Dias da Silva, Marcio Henrique de Oliveira Garcia, Leandro Magalhaes de Souza, Cristiane Gomes da Silva, Caio Luiz Pereira Ribeiro, Andrea Cony Cavalcanti, Claudia Maria Braga de Mello, Amilcar Tanuri, Ana Tereza R Vasconcelos |
| EPI_ISL_2101483, EPI_ISL_2101484 | Unidade de apoio ao diagnóstico da COVID - UNADIG | Bioinformatics Laboratory / LNCC | Luiz G P de Almeida, Alessandra P Lamarca, Ronaldo da Silva F Jr, Liliane Cavalcante, Alexandra L Gerber, Ana Paula de C Guimaraes, Douglas Terra Machado, Cassia Alves, Diana Mariani, Cintia Policarpo, Gleidson da Silva de Oliveira, Mario Sergio Ribeiro, Silvia Carvalho, Flavio Dias da Silva, Marcio Henrique de Oliveira Garcia, Leandro Magalhaes de Souza, Cristiane Gomes da Silva, Caio Luiz Pereira Ribeiro, Andrea Cony Cavalcanti, Claudia Maria Braga de Mello, Amilcar Tanuri, Ana Tereza R Vasconcelos |
| EPI_ISL_2101485, EPI_ISL_2101486, EPI_ISL_2101487, EPI_ISL_2101488, EPI_ISL_2101489, EPI_ISL_2101490, EPI_ISL_2101491, EPI_ISL_2101492, EPI_ISL_2101493 | Laboratorio Central Noel Nutels | Bioinformatics Laboratory / LNCC | Luiz G P de Almeida, Alessandra P Lamarca, Ronaldo da Silva F Jr, Liliane Cavalcante, Alexandra L Gerber, Ana Paula de C Guimaraes, Douglas Terra Machado, Cassia Alves, Diana Mariani, Cintia Policarpo, Gleidson da Silva de Oliveira, Mario Sergio Ribeiro, Silvia Carvalho, Flavio Dias da Silva, Marcio Henrique de Oliveira Garcia, Leandro Magalhaes de Souza, Cristiane Gomes da Silva, Caio Luiz Pereira Ribeiro, Andrea Cony Cavalcanti, Claudia Maria Braga de Mello, Amilcar Tanuri, Ana Tereza R Vasconcelos |
| EPI_ISL_2101494, EPI_ISL_2101495 | Unidade de apoio ao diagnóstico da COVID - UNADIG | Bioinformatics Laboratory / LNCC | Luiz G P de Almeida, Alessandra P Lamarca, Ronaldo da Silva F Jr, Liliane Cavalcante, Alexandra L Gerber, Ana Paula de C Guimaraes, Douglas Terra Machado, Cassia Alves, Diana Mariani, Cintia Policarpo, Gleidson da Silva de Oliveira, Mario Sergio Ribeiro, Silvia Carvalho, Flavio Dias da Silva, Marcio Henrique de Oliveira Garcia, Leandro Magalhaes de Souza, Cristiane Gomes da Silva, Caio Luiz Pereira Ribeiro, Andrea Cony Cavalcanti, Claudia Maria Braga de Mello, Amilcar Tanuri, Ana Tereza R Vasconcelos |
| EPI_ISL_2101496, EPI_ISL_2101497, EPI_ISL_2101498, EPI_ISL_2101499, EPI_ISL_2101500, EPI_ISL_2101501, EPI_ISL_2101502, EPI_ISL_2101503, EPI_ISL_2101504 | Laboratorio Central Noel Nutels | Bioinformatics Laboratory / LNCC | Luiz G P de Almeida, Alessandra P Lamarca, Ronaldo da Silva F Jr, Liliane Cavalcante, Alexandra L Gerber, Ana Paula de C Guimaraes, Douglas Terra Machado, Cassia Alves, Diana Mariani, Cintia Policarpo, Gleidson da Silva de Oliveira, Mario Sergio Ribeiro, Silvia Carvalho, Flavio Dias da Silva, Marcio Henrique de Oliveira Garcia, Leandro Magalhaes de Souza, Cristiane Gomes da Silva, Caio Luiz Pereira Ribeiro, Andrea Cony Cavalcanti, Claudia Maria Braga de Mello, Amilcar Tanuri, Ana Tereza R Vasconcelos |
| EPI_ISL_2101505, EPI_ISL_2101506 | Unidade de apoio ao diagnóstico da COVID - UNADIG | Bioinformatics Laboratory / LNCC | Luiz G P de Almeida, Alessandra P Lamarca, Ronaldo da Silva F Jr, Liliane Cavalcante, Alexandra L Gerber, Ana Paula de C Guimaraes, Douglas Terra Machado, Cassia Alves, Diana Mariani, Cintia Policarpo, Gleidson da Silva de Oliveira, Mario Sergio Ribeiro, Silvia Carvalho, Flavio Dias da Silva, Marcio Henrique de Oliveira Garcia, Leandro Magalhaes de Souza, Cristiane Gomes da Silva, Caio Luiz Pereira Ribeiro, Andrea Cony Cavalcanti, Claudia Maria Braga de Mello, Amilcar Tanuri, Ana Tereza R Vasconcelos |
| EPI_ISL_2101507 | Laboratorio Central Noel Nutels | Bioinformatics Laboratory / LNCC | Luiz G P de Almeida, Alessandra P Lamarca, Ronaldo da Silva F Jr, Liliane Cavalcante, Alexandra L Gerber, Ana Paula de C Guimaraes, Douglas Terra Machado, Cassia Alves, Diana Mariani, Cintia Policarpo, Gleidson da Silva de Oliveira, Mario Sergio Ribeiro, Silvia Carvalho, Flavio Dias da Silva, Marcio Henrique de Oliveira Garcia, Leandro Magalhaes de Souza, Cristiane Gomes da Silva, Caio Luiz Pereira Ribeiro, Andrea Cony Cavalcanti, Claudia Maria Braga de Mello, Amilcar Tanuri, Ana Tereza R Vasconcelos |
| EPI_ISL_2101508, EPI_ISL_2101509, EPI_ISL_2101510, EPI_ISL_2101511, EPI_ISL_2101512, EPI_ISL_2101513 | Unidade de apoio ao diagnóstico da COVID - UNADIG | Bioinformatics Laboratory / LNCC | Luiz G P de Almeida, Alessandra P Lamarca, Ronaldo da Silva F Jr, Liliane Cavalcante, Alexandra L Gerber, Ana Paula de C Guimaraes, Douglas Terra Machado, Cassia Alves, Diana Mariani, Cintia Policarpo, Gleidson da Silva de Oliveira, Mario Sergio Ribeiro, Silvia Carvalho, Flavio Dias da Silva, Marcio Henrique de Oliveira Garcia, Leandro Magalhaes de Souza, Cristiane Gomes da Silva, Caio Luiz Pereira Ribeiro, Andrea Cony Cavalcanti, Claudia Maria Braga de Mello, Amilcar Tanuri, Ana Tereza R Vasconcelos |
| EPI_ISL_2101514, EPI_ISL_2101515, EPI_ISL_2101516, EPI_ISL_2101517, EPI_ISL_2101518, EPI_ISL_2101519 | Laboratorio Central Noel Nutels | Bioinformatics Laboratory / LNCC | Luiz G P de Almeida, Alessandra P Lamarca, Ronaldo da Silva F Jr, Liliane Cavalcante, Alexandra L Gerber, Ana Paula de C Guimaraes, Douglas Terra Machado, Cassia Alves, Diana Mariani, Cintia Policarpo, Gleidson da Silva de Oliveira, Mario Sergio Ribeiro, Silvia Carvalho, Flavio Dias da Silva, Marcio Henrique de Oliveira Garcia, Leandro Magalhaes de Souza, Cristiane Gomes da Silva, Caio Luiz Pereira Ribeiro, Andrea Cony Cavalcanti, Claudia Maria Braga de Mello, Amilcar Tanuri, Ana Tereza R Vasconcelos |
| EPI_ISL_2101520, EPI_ISL_2101521, EPI_ISL_2101522, EPI_ISL_2101523, EPI_ISL_2101524, EPI_ISL_2101525, EPI_ISL_2101526, EPI_ISL_2101527 | Unidade de apoio ao diagnóstico da COVID - UNADIG | Bioinformatics Laboratory / LNCC | Luiz G P de Almeida, Alessandra P Lamarca, Ronaldo da Silva F Jr, Liliane Cavalcante, Alexandra L Gerber, Ana Paula de C Guimaraes, Douglas Terra Machado, Cassia Alves, Diana Mariani, Cintia Policarpo, Gleidson da Silva de Oliveira, Mario Sergio Ribeiro, Silvia Carvalho, Flavio Dias da Silva, Marcio Henrique de Oliveira Garcia, Leandro Magalhaes de Souza, Cristiane Gomes da Silva, Caio Luiz Pereira Ribeiro, Andrea Cony Cavalcanti, Claudia Maria Braga de Mello, Amilcar Tanuri, Ana Tereza R Vasconcelos |
| EPI_ISL_2101528, EPI_ISL_2101529, EPI_ISL_2101530, EPI_ISL_2101531, EPI_ISL_2101532, EPI_ISL_2101533, EPI_ISL_2101534, EPI_ISL_2101535, EPI_ISL_2101536, EPI_ISL_2101537, EPI_ISL_2101538, EPI_ISL_2101539, EPI_ISL_2101540, EPI_ISL_2101541, EPI_ISL_2101542, EPI_ISL_2101543, EPI_ISL_2101544, EPI_ISL_2101545, EPI_ISL_2101546, EPI_ISL_2101547, EPI_ISL_2101548, EPI_ISL_2101549, EPI_ISL_2101550, EPI_ISL_2101551, EPI_ISL_2101552, EPI_ISL_2101553, EPI_ISL_2101554, EPI_ISL_2101555, EPI_ISL_2101556, EPI_ISL_2101557, EPI_ISL_2101558, EPI_ISL_2101559, EPI_ISL_2101560, EPI_ISL_2101561, EPI_ISL_2101562, EPI_ISL_2101563, EPI_ISL_2101564, EPI_ISL_2101565 | | | |
| see above | Laboratorio Central Noel Nutels | Bioinformatics Laboratory / LNCC | Luiz G P de Almeida, Alessandra P Lamarca, Ronaldo da Silva F Jr, Liliane Cavalcante, Alexandra L Gerber, Ana Paula de C Guimaraes, Douglas Terra Machado, Cassia Alves, Diana Mariani, Cintia Policarpo, Gleidson da Silva de Oliveira, Mario Sergio Ribeiro, Silvia Carvalho, Flavio Dias da Silva, Marcio Henrique de Oliveira Garcia, Leandro Magalhaes de Souza, Cristiane Gomes da Silva, Caio Luiz Pereira Ribeiro, Andrea Cony Cavalcanti, Claudia Maria Braga de Mello, Amilcar Tanuri, Ana Tereza R Vasconcelos |
| EPI_ISL_2101566, EPI_ISL_2101567, EPI_ISL_2101568, EPI_ISL_2101569, EPI_ISL_2101570, EPI_ISL_2101571, | Unidade de apoio ao diagnóstico da COVID - UNADIG | Bioinformatics Laboratory / LNCC | Luiz G P de Almeida, Alessandra P Lamarca, Ronaldo da Silva F Jr, Liliane Cavalcante, Alexandra L Gerber, Ana Paula de C Guimaraes, Douglas Terra Machado, Cassia Alves, Diana Mariani, Cintia Policarpo, Gleidson da Silva de Oliveira, Mario Sergio Ribeiro, Silvia Carvalho, Flavio Dias da Silva, Marcio Henrique de Oliveira Garcia, Leandro Magalhaes de Souza, Cristiane Gomes da Silva, Caio Luiz Pereira Ribeiro, Andrea Cony Cavalcanti, Claudia Maria |

| | | | |
|---|---|---|---|
| EPI_ISL_2101572 | | | Braga de Mello, Amilcar Tanuri, Ana Tereza R Vasconcelos |
| EPI_ISL_2101573, EPI_ISL_2101574, EPI_ISL_2101575, EPI_ISL_2101576, EPI_ISL_2101577, EPI_ISL_2101578, EPI_ISL_2101579, EPI_ISL_2101580, EPI_ISL_2101581, EPI_ISL_2101582, EPI_ISL_2101583, EPI_ISL_2101584, EPI_ISL_2101585, EPI_ISL_2101586, EPI_ISL_2101587, EPI_ISL_2101588, EPI_ISL_2101589, EPI_ISL_2101590, EPI_ISL_2101591, EPI_ISL_2101592, EPI_ISL_2101593, EPI_ISL_2101594, EPI_ISL_2101595, EPI_ISL_2101596, EPI_ISL_2101597, EPI_ISL_2101598, EPI_ISL_2101599, EPI_ISL_2101600, EPI_ISL_2101601, EPI_ISL_2101602, EPI_ISL_2101603, EPI_ISL_2101604, EPI_ISL_2101605, EPI_ISL_2101606, EPI_ISL_2101607, EPI_ISL_2101608, EPI_ISL_2101609, EPI_ISL_2101610, EPI_ISL_2101611 | | | |
| see above | Laboratorio Central Noel Nutels | Bioinformatics Laboratory / LNCC | Luiz G P de Almeida, Alessandra P Lamarca, Ronaldo da Silva F Jr, Liliane Cavalcante, Alexandra L Gerber, Ana Paula de C Guimaraes, Douglas Terra Machado, Cassia Alves, Diana Mariani, Cintia Policarpo, Gleidson da Silva de Oliveira, Mario Sergio Ribeiro, Silvia Carvalho, Flavio Dias da Silva, Marcio Henrique de Oliveira Garcia, Leandro Magalhaes de Souza, Cristiane Gomes da Silva, Caio Luiz Pereira Ribeiro, Andrea Cony Cavalcanti, Claudia Maria Braga de Mello, Amilcar Tanuri, Ana Tereza R Vasconcelos |
| EPI_ISL_2101612 | Unidade de apoio ao diagnóstico da COVID - UNADIG | Bioinformatics Laboratory / LNCC | Luiz G P de Almeida, Alessandra P Lamarca, Ronaldo da Silva F Jr, Liliane Cavalcante, Alexandra L Gerber, Ana Paula de C Guimaraes, Douglas Terra Machado, Cassia Alves, Diana Mariani, Cintia Policarpo, Gleidson da Silva de Oliveira, Mario Sergio Ribeiro, Silvia Carvalho, Flavio Dias da Silva, Marcio Henrique de Oliveira Garcia, Leandro Magalhaes de Souza, Cristiane Gomes da Silva, Caio Luiz Pereira Ribeiro, Andrea Cony Cavalcanti, Claudia Maria Braga de Mello, Amilcar Tanuri, Ana Tereza R Vasconcelos |
| EPI_ISL_2101613, EPI_ISL_2101615, EPI_ISL_2101616, EPI_ISL_2101617, EPI_ISL_2101618, EPI_ISL_2101619, EPI_ISL_2101620, EPI_ISL_2101621, EPI_ISL_2101622, EPI_ISL_2101623, EPI_ISL_2101624, EPI_ISL_2101625, EPI_ISL_2101626, EPI_ISL_2101627, EPI_ISL_2101628, EPI_ISL_2101629, EPI_ISL_2101630, EPI_ISL_2101631, EPI_ISL_2101632, EPI_ISL_2101633, EPI_ISL_2101634, EPI_ISL_2101635, EPI_ISL_2101636, EPI_ISL_2101637 | | | |
| see above | Laboratorio Central Noel Nutels | Bioinformatics Laboratory / LNCC | Luiz G P de Almeida, Alessandra P Lamarca, Ronaldo da Silva F Jr, Liliane Cavalcante, Alexandra L Gerber, Ana Paula de C Guimaraes, Douglas Terra Machado, Cassia Alves, Diana Mariani, Cintia Policarpo, Gleidson da Silva de Oliveira, Mario Sergio Ribeiro, Silvia Carvalho, Flavio Dias da Silva, Marcio Henrique de Oliveira Garcia, Leandro Magalhaes de Souza, Cristiane Gomes da Silva, Caio Luiz Pereira Ribeiro, Andrea Cony Cavalcanti, Claudia Maria Braga de Mello, Amilcar Tanuri, Ana Tereza R Vasconcelos |
| EPI_ISL_2101638, EPI_ISL_2101639, EPI_ISL_2101640, EPI_ISL_2101641, EPI_ISL_2101642, EPI_ISL_2101643, EPI_ISL_2101644, EPI_ISL_2101645, EPI_ISL_2101646, EPI_ISL_2101647, EPI_ISL_2101648, EPI_ISL_2101649, EPI_ISL_2101650, EPI_ISL_2101651, EPI_ISL_2101652, EPI_ISL_2101653, EPI_ISL_2101654, EPI_ISL_2101655, EPI_ISL_2101656, EPI_ISL_2101657, EPI_ISL_2101658, EPI_ISL_2101659, EPI_ISL_2101660, EPI_ISL_2101661, EPI_ISL_2101662, EPI_ISL_2101663, EPI_ISL_2101664, EPI_ISL_2101665, EPI_ISL_2101666, EPI_ISL_2101667, EPI_ISL_2101668, EPI_ISL_2101669, EPI_ISL_2101670, EPI_ISL_2101671 | | | |
| see above | Unidade de apoio ao diagnóstico da COVID - UNADIG | Bioinformatics Laboratory / LNCC | Luiz G P de Almeida, Alessandra P Lamarca, Ronaldo da Silva F Jr, Liliane Cavalcante, Alexandra L Gerber, Ana Paula de C Guimaraes, Douglas Terra Machado, Cassia Alves, Diana Mariani, Cintia Policarpo, Gleidson da Silva de Oliveira, Mario Sergio Ribeiro, Silvia Carvalho, Flavio Dias da Silva, Marcio Henrique de Oliveira Garcia, Leandro Magalhaes de Souza, Cristiane Gomes da Silva, Caio Luiz Pereira Ribeiro, Andrea Cony Cavalcanti, Claudia Maria Braga de Mello, Amilcar Tanuri, Ana Tereza R Vasconcelos |
| EPI_ISL_2101672, EPI_ISL_2101673, EPI_ISL_2101674, EPI_ISL_2101675 | Laboratorio Central Noel Nutels | Bioinformatics Laboratory / LNCC | Luiz G P de Almeida, Alessandra P Lamarca, Ronaldo da Silva F Jr, Liliane Cavalcante, Alexandra L Gerber, Ana Paula de C Guimaraes, Douglas Terra Machado, Cassia Alves, Diana Mariani, Cintia Policarpo, Gleidson da Silva de Oliveira, Mario Sergio Ribeiro, Silvia Carvalho, Flavio Dias da Silva, Marcio Henrique de Oliveira Garcia, Leandro Magalhaes de Souza, Cristiane Gomes da Silva, Caio Luiz Pereira Ribeiro, Andrea Cony Cavalcanti, Claudia Maria Braga de Mello, Amilcar Tanuri, Ana Tereza R Vasconcelos |
| EPI_ISL_2101676, EPI_ISL_2101677, EPI_ISL_2101678, EPI_ISL_2101679, EPI_ISL_2101680, EPI_ISL_2101681, EPI_ISL_2101682, EPI_ISL_2101683, EPI_ISL_2101684, EPI_ISL_2101685, EPI_ISL_2101686, EPI_ISL_2101687, EPI_ISL_2101688 | | | |
| see above | Unidade de apoio ao diagnóstico da COVID - UNADIG | Bioinformatics Laboratory / LNCC | Luiz G P de Almeida, Alessandra P Lamarca, Ronaldo da Silva F Jr, Liliane Cavalcante, Alexandra L Gerber, Ana Paula de C Guimaraes, Douglas Terra Machado, Cassia Alves, Diana Mariani, Cintia Policarpo, Gleidson da Silva de Oliveira, Mario Sergio Ribeiro, Silvia Carvalho, Flavio Dias da Silva, Marcio Henrique de Oliveira Garcia, Leandro Magalhaes de Souza, Cristiane Gomes da Silva, Caio Luiz Pereira Ribeiro, Andrea Cony Cavalcanti, Claudia Maria Braga de Mello, Amilcar Tanuri, Ana Tereza R Vasconcelos |
| EPI_ISL_2101689, EPI_ISL_2101690, EPI_ISL_2101691 | Laboratorio Central Noel Nutels | Bioinformatics Laboratory / LNCC | Luiz G P de Almeida, Alessandra P Lamarca, Ronaldo da Silva F Jr, Liliane Cavalcante, Alexandra L Gerber, Ana Paula de C Guimaraes, Douglas Terra Machado, Cassia Alves, Diana Mariani, Cintia Policarpo, Gleidson da Silva de Oliveira, Mario Sergio Ribeiro, Silvia Carvalho, Flavio Dias da Silva, Marcio Henrique de Oliveira Garcia, Leandro Magalhaes de Souza, Cristiane Gomes da Silva, Caio Luiz Pereira Ribeiro, Andrea Cony Cavalcanti, Claudia Maria Braga de Mello, Amilcar Tanuri, Ana Tereza R Vasconcelos |
| EPI_ISL_2101692, EPI_ISL_2101693, EPI_ISL_2101694, EPI_ISL_2101695, EPI_ISL_2101696 | Unidade de apoio ao diagnóstico da COVID - UNADIG | Bioinformatics Laboratory / LNCC | Luiz G P de Almeida, Alessandra P Lamarca, Ronaldo da Silva F Jr, Liliane Cavalcante, Alexandra L Gerber, Ana Paula de C Guimaraes, Douglas Terra Machado, Cassia Alves, Diana Mariani, Cintia Policarpo, Gleidson da Silva de Oliveira, Mario Sergio Ribeiro, Silvia Carvalho, Flavio Dias da Silva, Marcio Henrique de Oliveira Garcia, Leandro Magalhaes de Souza, Cristiane Gomes da Silva, Caio Luiz Pereira Ribeiro, Andrea Cony Cavalcanti, Claudia Maria Braga de Mello, Amilcar Tanuri, Ana Tereza R Vasconcelos |
| EPI_ISL_2101697, EPI_ISL_2101698, EPI_ISL_2101699, EPI_ISL_2101700, EPI_ISL_2101701, EPI_ISL_2101702, EPI_ISL_2101703, EPI_ISL_2101704, EPI_ISL_2101705, EPI_ISL_2101706, EPI_ISL_2101707, EPI_ISL_2101708, EPI_ISL_2101709, EPI_ISL_2101710, EPI_ISL_2101711, EPI_ISL_2101712, EPI_ISL_2101713 | | | |
| see above | Laboratorio Central Noel Nutels | Bioinformatics Laboratory / LNCC | Luiz G P de Almeida, Alessandra P Lamarca, Ronaldo da Silva F Jr, Liliane Cavalcante, Alexandra L Gerber, Ana Paula de C Guimaraes, Douglas Terra Machado, Cassia Alves, Diana Mariani, Cintia Policarpo, Gleidson da Silva de Oliveira, Mario Sergio Ribeiro, Silvia Carvalho, Flavio Dias da Silva, Marcio Henrique de Oliveira Garcia, Leandro Magalhaes de Souza, Cristiane Gomes da Silva, Caio Luiz Pereira Ribeiro, Andrea Cony Cavalcanti, Claudia Maria Braga de Mello, Amilcar Tanuri, Ana Tereza R Vasconcelos |
| EPI_ISL_2101714, EPI_ISL_2101715 | Unidade de apoio ao diagnóstico da COVID - UNADIG | Bioinformatics Laboratory / LNCC | Luiz G P de Almeida, Alessandra P Lamarca, Ronaldo da Silva F Jr, Liliane Cavalcante, Alexandra L Gerber, Ana Paula de C Guimaraes, Douglas Terra Machado, Cassia Alves, Diana Mariani, Cintia Policarpo, Gleidson da Silva de Oliveira, Mario Sergio Ribeiro, Silvia Carvalho, Flavio Dias da Silva, Marcio Henrique de Oliveira Garcia, Leandro Magalhaes de Souza, Cristiane Gomes da Silva, Caio Luiz Pereira Ribeiro, Andrea Cony Cavalcanti, Claudia Maria Braga de Mello, Amilcar Tanuri, Ana Tereza R Vasconcelos |
| EPI_ISL_2101716, EPI_ISL_2101717, EPI_ISL_2101718, EPI_ISL_2101719, EPI_ISL_2101720, EPI_ISL_2101721 | Laboratorio Central Noel Nutels | Bioinformatics Laboratory / LNCC | Luiz G P de Almeida, Alessandra P Lamarca, Ronaldo da Silva F Jr, Liliane Cavalcante, Alexandra L Gerber, Ana Paula de C Guimaraes, Douglas Terra Machado, Cassia Alves, Diana Mariani, Cintia Policarpo, Gleidson da Silva de Oliveira, Mario Sergio Ribeiro, Silvia Carvalho, Flavio Dias da Silva, Marcio Henrique de Oliveira Garcia, Leandro Magalhaes de Souza, Cristiane Gomes da Silva, Caio Luiz Pereira Ribeiro, Andrea Cony Cavalcanti, Claudia Maria Braga de Mello, Amilcar Tanuri, Ana Tereza R Vasconcelos |
| EPI_ISL_2101722 | Unidade de apoio ao diagnóstico da COVID - UNADIG | Bioinformatics Laboratory / LNCC | Luiz G P de Almeida, Alessandra P Lamarca, Ronaldo da Silva F Jr, Liliane Cavalcante, Alexandra L Gerber, Ana Paula de C Guimaraes, Douglas Terra Machado, Cassia Alves, Diana Mariani, Cintia Policarpo, Gleidson da Silva de Oliveira, Mario Sergio Ribeiro, Silvia Carvalho, Flavio Dias da Silva, Marcio Henrique de Oliveira Garcia, Leandro Magalhaes de Souza, Cristiane Gomes da Silva, Caio Luiz Pereira Ribeiro, Andrea Cony Cavalcanti, Claudia Maria Braga de Mello, Amilcar Tanuri, Ana Tereza R Vasconcelos |
| EPI_ISL_2101730 | Laboratorio Central Noel Nutels | Bioinformatics Laboratory / LNCC | Luiz G P de Almeida, Alessandra P Lamarca, Ronaldo da Silva F Jr, Liliane Cavalcante, Alexandra L Gerber, Ana Paula de C Guimaraes, Douglas Terra Machado, Cassia Alves, Diana Mariani, Cintia Policarpo, Gleidson da Silva de Oliveira, Mario Sergio Ribeiro, Silvia Carvalho, Flavio Dias da Silva, Marcio Henrique de Oliveira Garcia, Leandro Magalhaes de Souza, Cristiane Gomes da Silva, Caio Luiz Pereira Ribeiro, Andrea Cony Cavalcanti, Claudia Maria Braga de Mello, Amilcar Tanuri, Ana Tereza R Vasconcelos |
| EPI_ISL_2101731 | Unidade de apoio ao diagnóstico da COVID - UNADIG | Bioinformatics Laboratory / LNCC | Luiz G P de Almeida, Alessandra P Lamarca, Ronaldo da Silva F Jr, Liliane Cavalcante, Alexandra L Gerber, Ana Paula de C Guimaraes, Douglas Terra Machado, Cassia Alves, Diana Mariani, Cintia Policarpo, Gleidson da Silva de Oliveira, Mario Sergio Ribeiro, Silvia Carvalho, Flavio Dias da Silva, Marcio Henrique de Oliveira Garcia, Leandro Magalhaes de Souza, Cristiane Gomes da Silva, Caio Luiz Pereira Ribeiro, Andrea Cony Cavalcanti, Claudia Maria Braga de Mello, Amilcar Tanuri, Ana Tereza R Vasconcelos |
| EPI_ISL_2101732, EPI_ISL_2101733 | Laboratorio Central Noel Nutels | Bioinformatics Laboratory / LNCC | Luiz G P de Almeida, Alessandra P Lamarca, Ronaldo da Silva F Jr, Liliane Cavalcante, Alexandra L Gerber, Ana Paula de C Guimaraes, Douglas Terra Machado, Cassia Alves, Diana Mariani, Cintia Policarpo, Gleidson da Silva de Oliveira, Mario Sergio Ribeiro, Silvia Carvalho, Flavio Dias da Silva, Marcio Henrique de Oliveira Garcia, Leandro Magalhaes de Souza, Cristiane Gomes da Silva, Caio Luiz Pereira Ribeiro, Andrea Cony Cavalcanti, Claudia Maria Braga de Mello, Amilcar Tanuri, Ana Tereza R Vasconcelos |
| EPI_ISL_2101734 | Unidade de apoio ao diagnóstico da COVID - UNADIG | Bioinformatics Laboratory / LNCC | Luiz G P de Almeida, Alessandra P Lamarca, Ronaldo da Silva F Jr, Liliane Cavalcante, Alexandra L Gerber, Ana Paula de C Guimaraes, Douglas Terra Machado, Cassia Alves, Diana Mariani, Cintia Policarpo, Gleidson da Silva de Oliveira, Mario Sergio Ribeiro, Silvia Carvalho, Flavio Dias da Silva, Marcio Henrique de Oliveira Garcia, Leandro Magalhaes de Souza, Cristiane Gomes da Silva, Caio Luiz Pereira Ribeiro, Andrea Cony Cavalcanti, Claudia Maria Braga de Mello, Amilcar Tanuri, Ana Tereza R Vasconcelos |
| EPI_ISL_2101736, EPI_ISL_2101737, EPI_ISL_2101738, EPI_ISL_2101739 | Laboratorio Central Noel Nutels | Bioinformatics Laboratory / LNCC | Luiz G P de Almeida, Alessandra P Lamarca, Ronaldo da Silva F Jr, Liliane Cavalcante, Alexandra L Gerber, Ana Paula de C Guimaraes, Douglas Terra Machado, Cassia Alves, Diana Mariani, Cintia Policarpo, Gleidson da Silva de Oliveira, Mario Sergio Ribeiro, Silvia Carvalho, Flavio Dias da Silva, Marcio |

| | | | |
|---|---|---|---|
| | | | Henrique de Oliveira Garcia, Leandro Magalhaes de Souza, Cristiane Gomes da Silva, Caio Luiz Pereira Ribeiro, Andrea Cony Cavalcanti, Claudia Maria Braga de Mello, Amilcar Tanuri, Ana Tereza R Vasconcelos |
| EPI_ISL_2101740, EPI_ISL_2101741 | Unidade de apoio ao diagnóstico da COVID - UNADIG | Bioinformatics Laboratory / LNCC | Luiz G P de Almeida, Alessandra P Lamarca, Ronaldo da Silva F Jr, Liliane Cavalcante, Alexandra L Gerber, Ana Paula de C Guimaraes, Douglas Terra Machado, Cassia Alves, Diana Mariani, Cintia Policarpo, Gleidson da Silva de Oliveira, Mario Sergio Ribeiro, Silvia Carvalho, Flavio Dias da Silva, Marcio Henrique de Oliveira Garcia, Leandro Magalhaes de Souza, Cristiane Gomes da Silva, Caio Luiz Pereira Ribeiro, Andrea Cony Cavalcanti, Claudia Maria Braga de Mello, Amilcar Tanuri, Ana Tereza R Vasconcelos |
| EPI_ISL_2101742, EPI_ISL_2101743 | Laboratorio Central Noel Nutels | Bioinformatics Laboratory / LNCC | Luiz G P de Almeida, Alessandra P Lamarca, Ronaldo da Silva F Jr, Liliane Cavalcante, Alexandra L Gerber, Ana Paula de C Guimaraes, Douglas Terra Machado, Cassia Alves, Diana Mariani, Cintia Policarpo, Gleidson da Silva de Oliveira, Mario Sergio Ribeiro, Silvia Carvalho, Flavio Dias da Silva, Marcio Henrique de Oliveira Garcia, Leandro Magalhaes de Souza, Cristiane Gomes da Silva, Caio Luiz Pereira Ribeiro, Andrea Cony Cavalcanti, Claudia Maria Braga de Mello, Amilcar Tanuri, Ana Tereza R Vasconcelos |
| EPI_ISL_2101744 | Unidade de apoio ao diagnóstico da COVID - UNADIG | Bioinformatics Laboratory / LNCC | Luiz G P de Almeida, Alessandra P Lamarca, Ronaldo da Silva F Jr, Liliane Cavalcante, Alexandra L Gerber, Ana Paula de C Guimaraes, Douglas Terra Machado, Cassia Alves, Diana Mariani, Cintia Policarpo, Gleidson da Silva de Oliveira, Mario Sergio Ribeiro, Silvia Carvalho, Flavio Dias da Silva, Marcio Henrique de Oliveira Garcia, Leandro Magalhaes de Souza, Cristiane Gomes da Silva, Caio Luiz Pereira Ribeiro, Andrea Cony Cavalcanti, Claudia Maria Braga de Mello, Amilcar Tanuri, Ana Tereza R Vasconcelos |
| EPI_ISL_2101745, EPI_ISL_2101746, EPI_ISL_2101747 | Laboratorio Central Noel Nutels | Bioinformatics Laboratory / LNCC | Luiz G P de Almeida, Alessandra P Lamarca, Ronaldo da Silva F Jr, Liliane Cavalcante, Alexandra L Gerber, Ana Paula de C Guimaraes, Douglas Terra Machado, Cassia Alves, Diana Mariani, Cintia Policarpo, Gleidson da Silva de Oliveira, Mario Sergio Ribeiro, Silvia Carvalho, Flavio Dias da Silva, Marcio Henrique de Oliveira Garcia, Leandro Magalhaes de Souza, Cristiane Gomes da Silva, Caio Luiz Pereira Ribeiro, Andrea Cony Cavalcanti, Claudia Maria Braga de Mello, Amilcar Tanuri, Ana Tereza R Vasconcelos |
| EPI_ISL_2157524, EPI_ISL_2157525, EPI_ISL_2157526, EPI_ISL_2157528, EPI_ISL_2157529, EPI_ISL_2157530, EPI_ISL_2157536, EPI_ISL_2157537, EPI_ISL_2157538, EPI_ISL_2157539 | Laboratório Central de Saude Publica do Estado do Rio de Janeiro (LACEN/RJ) | Laboratory of Respiratory Viruses and Measles, Oswaldo Cruz Institute, FIOCRUZ | Paola Resende, Luciana Appolinario, Fernando Motta, Anna Carolina Paixao, Ana Carolina Mendonca, Alice Sampaio Rocha, Taina Venas, Elisa Cavalcante Pereira, Renata Serrano Lopes, Andrea Cony Cavalcanti, Marilda Siqueira on behalf of the Fiocruz COVID-19 Genomic Surveillance Network |
| EPI_ISL_2157540, EPI_ISL_2157545 | Laboratory of Respiratory Viruses and Measles, Oswaldo Cruz Institute, FIOCRUZ | Laboratory of Respiratory Viruses and Measles, Oswaldo Cruz Institute, FIOCRUZ | Paola Resende, Luciana Appolinario, Fernando Motta, Anna Carolina Paixao, Ana Carolina Mendonca, Alice Sampaio Rocha, Taina Venas, Elisa Cavalcante Pereira, Renata Serrano Lopes, Andrea Cony Cavalcanti, Marilda Siqueira on behalf of the Fiocruz COVID-19 Genomic Surveillance Network |
| EPI_ISL_2157549 | Laboratório Central de Saude Publica do Estado do Rio de Janeiro (LACEN/RJ) | Laboratory of Respiratory Viruses and Measles, Oswaldo Cruz Institute, FIOCRUZ | Paola Resende, Luciana Appolinario, Fernando Motta, Anna Carolina Paixao, Ana Carolina Mendonca, Alice Sampaio Rocha, Taina Venas, Elisa Cavalcante Pereira, Renata Serrano Lopes, Andrea Cony Cavalcanti, Marilda Siqueira on behalf of the Fiocruz COVID-19 Genomic Surveillance Network |
| EPI_ISL_2157561 | Laboratorio Central de Saude Publica do Estado do Rio de Janeiro (LACEN/RJ) | Laboratory of Respiratory Viruses and Measles, Oswaldo Cruz Institute, FIOCRUZ | Paola Resende, Luciana Appolinario, Fernando Motta, Anna Carolina Paixao, Ana Carolina Mendonca, Alice Sampaio Rocha, Taina Venas, Elisa Cavalcante Pereira, Renata Serrano Lopes, Andrea Cony Cavalcanti, Marilda Siqueira on behalf of the Fiocruz COVID-19 Genomic Surveillance Network |
| EPI_ISL_2157573, EPI_ISL_2157575 | Laboratory of Respiratory Viruses and Measles, Oswaldo Cruz Institute, FIOCRUZ | Laboratory of Respiratory Viruses and Measles, Oswaldo Cruz Institute, FIOCRUZ | Paola Resende, Luciana Appolinario, Fernando Motta, Anna Carolina Paixao, Ana Carolina Mendonca, Alice Sampaio Rocha, Taina Venas, Elisa Cavalcante Pereira, Renata Serrano Lopes, Andrea Cony Cavalcanti, Marilda Siqueira on behalf of the Fiocruz COVID-19 Genomic Surveillance Network |
| EPI_ISL_2157599 | Laboratório Central de Saude Publica do Estado do Rio de Janeiro (LACEN/RJ) | Laboratory of Respiratory Viruses and Measles, Oswaldo Cruz Institute, FIOCRUZ | Paola Resende, Luciana Appolinario, Fernando Motta, Anna Carolina Paixao, Ana Carolina Mendonca, Alice Sampaio Rocha, Taina Venas, Elisa Cavalcante Pereira, Renata Serrano Lopes, Andrea Cony Cavalcanti, Marilda Siqueira on behalf of the Fiocruz COVID-19 Genomic Surveillance Network |
| EPI_ISL_2196185, EPI_ISL_2196186, EPI_ISL_2196187 | Laboratory of Respiratory Viruses and Measles, Oswaldo Cruz Institute, FIOCRUZ | Laboratory of Respiratory Viruses and Measles, Oswaldo Cruz Institute, FIOCRUZ | Paola Resende, Luciana Appolinario, Fernando Motta, Anna Carolina Paixao, Ana Carolina Mendonca, Alice Sampaio Rocha, Taina Venas, Elisa Cavalcante Pereira, Renata Serrano Lopes, Marilda Siqueira on behalf of the Fiocruz COVID-19 Genomic Surveillance Network |
| EPI_ISL_2196189 | Laboratorio Central de Saude Publica do Estado do Rio de Janeiro (LACEN-RJ) | Laboratory of Respiratory Viruses and Measles, Oswaldo Cruz Institute, FIOCRUZ | Paola Resende, Luciana Appolinario, Fernando Motta, Anna Carolina Paixao, Ana Carolina Mendonca, Alice Sampaio Rocha, Taina Venas, Elisa Cavalcante Pereira, Renata Serrano Lopes, Andrea Cony Cavalcanti, Marilda Siqueira on behalf of the Fiocruz COVID-19 Genomic Surveillance Network |
| EPI_ISL_2196190, EPI_ISL_2196191, EPI_ISL_2196192, EPI_ISL_2196193, EPI_ISL_2196194, EPI_ISL_2196195, EPI_ISL_2196196 | Laboratory of Respiratory Viruses and Measles, Oswaldo Cruz Institute, FIOCRUZ | Laboratory of Respiratory Viruses and Measles, Oswaldo Cruz Institute, FIOCRUZ | Paola Resende, Luciana Appolinario, Fernando Motta, Anna Carolina Paixao, Ana Carolina Mendonca, Alice Sampaio Rocha, Taina Venas, Elisa Cavalcante Pereira, Renata Serrano Lopes, Marilda Siqueira on behalf of the Fiocruz COVID-19 Genomic Surveillance Network |
| EPI_ISL_2196197, EPI_ISL_2196198, EPI_ISL_2196199, EPI_ISL_2196200, EPI_ISL_2196201, EPI_ISL_2196202, EPI_ISL_2196203, EPI_ISL_2196204, EPI_ISL_2196205, EPI_ISL_2196206, EPI_ISL_2196207, EPI_ISL_2196208, EPI_ISL_2196209, EPI_ISL_2196236 | | | |
| see above | Laboratorio Central de Saude Publica do Estado do Rio de Janeiro (LACEN-RJ) | Laboratory of Respiratory Viruses and Measles, Oswaldo Cruz Institute, FIOCRUZ | Paola Resende, Luciana Appolinario, Fernando Motta, Anna Carolina Paixao, Ana Carolina Mendonca, Alice Sampaio Rocha, Taina Venas, Elisa Cavalcante Pereira, Renata Serrano Lopes, Andrea Cony Cavalcanti, Marilda Siqueira on behalf of the Fiocruz COVID-19 Genomic Surveillance Network |
| EPI_ISL_2196242, EPI_ISL_2196245 | Labortorio Central de Saude Publica do Estado do Rio de Janeiro (LACEN/RJ) | Laboratory of Respiratory Viruses and Measles, Oswaldo Cruz Institute, FIOCRUZ | Paola Resende, Luciana Appolinario, Fernando Motta, Anna Carolina Paixao, Ana Carolina Mendonca, Alice Sampaio Rocha, Taina Venas, Elisa Cavalcante Pereira, Renata Serrano Lopes, Andrea Cony Cavalcanti, Marilda Siqueira on behalf of the Fiocruz COVID-19 Genomic Surveillance Network |
| EPI_ISL_2196251 | Laboratory of Respiratory Viruses and Measles, Oswaldo Cruz Institute, FIOCRUZ | Laboratory of Respiratory Viruses and Measles, Oswaldo Cruz Institute, FIOCRUZ | Paola Resende, Luciana Appolinario, Fernando Motta, Anna Carolina Paixao, Ana Carolina Mendonca, Alice Sampaio Rocha, Taina Venas, Elisa Cavalcante Pereira, Renata Serrano Lopes, Marilda Siqueira on behalf of the Fiocruz COVID-19 Genomic Surveillance Network |
| EPI_ISL_2196313, EPI_ISL_2196314, EPI_ISL_2196315, EPI_ISL_2196316, EPI_ISL_2196317, EPI_ISL_2196318, EPI_ISL_2196319, EPI_ISL_2196320, EPI_ISL_2196321, EPI_ISL_2196322, EPI_ISL_2196323, EPI_ISL_2196324, EPI_ISL_2196325, EPI_ISL_2196326, EPI_ISL_2196327, EPI_ISL_2196328, EPI_ISL_2196329, EPI_ISL_2196330, EPI_ISL_2196331, EPI_ISL_2196332, EPI_ISL_2196333, EPI_ISL_2196334, EPI_ISL_2196356 | | | |
| see above | Labortorio Central de Saude Publica do Estado do Rio de Janeiro (LACEN/RJ) | Laboratory of Respiratory Viruses and Measles, Oswaldo Cruz Institute, FIOCRUZ | Paola Resende, Luciana Appolinario, Fernando Motta, Anna Carolina Paixao, Ana Carolina Mendonca, Alice Sampaio Rocha, Taina Venas, Elisa Cavalcante Pereira, Renata Serrano Lopes, Andrea Cony Cavalcanti, Marilda Siqueira on behalf of the Fiocruz COVID-19 Genomic Surveillance Network |
| EPI_ISL_2196361 | Laboratory of Respiratory Viruses and Measles, Oswaldo Cruz Institute, FIOCRUZ | Laboratory of Respiratory Viruses and Measles, Oswaldo Cruz Institute, FIOCRUZ | Paola Resende, Luciana Appolinario, Fernando Motta, Anna Carolina Paixao, Ana Carolina Mendonca, Alice Sampaio Rocha, Taina Venas, Elisa Cavalcante Pereira, Renata Serrano Lopes, Marilda Siqueira on behalf of the Fiocruz COVID-19 Genomic Surveillance Network |
| EPI_ISL_2249404, EPI_ISL_2249405, EPI_ISL_2249406, EPI_ISL_2249407, EPI_ISL_2249408, EPI_ISL_2249409, EPI_ISL_2249410, EPI_ISL_2249411, EPI_ISL_2249412, EPI_ISL_2249413, EPI_ISL_2249414, EPI_ISL_2249415, EPI_ISL_2249416, EPI_ISL_2249417, EPI_ISL_2249418, EPI_ISL_2249419, EPI_ISL_2249420, EPI_ISL_2249421, EPI_ISL_2249422, EPI_ISL_2249423, EPI_ISL_2249424, EPI_ISL_2249425, EPI_ISL_2249426, EPI_ISL_2249427, EPI_ISL_2249428, EPI_ISL_2249429, EPI_ISL_2249430, EPI_ISL_2249431, EPI_ISL_2249432, EPI_ISL_2249433, EPI_ISL_2249434, EPI_ISL_2249440, EPI_ISL_2249441, EPI_ISL_2249442, EPI_ISL_2249443, EPI_ISL_2249444, EPI_ISL_2249445 | | | |
| see above | Laboratório Central de Saúde Pública do Rio de Janeiro | Coordenação Geral de Laboratórios de Saúde Pública (CGLAB/DAEVS/SVS/MS) | Vagner Fonseca, et al. |
| EPI_ISL_2274097 | Laboratory of Respiratory Viruses and Measles, Oswaldo Cruz Institute, FIOCRUZ | Laboratory of Respiratory Viruses and Measles, Oswaldo Cruz Institute, FIOCRUZ | Paola Resende, Luciana Appolinario, Fernando Motta, Anna Carolina Paixao, Ana Carolina Mendonca, Alice Sampaio Rocha, Taina Venas, Elisa Cavalcante Pereira, Renata Serrano Lopes, Marilda Siqueira on behalf of the Fiocruz COVID-19 Genomic Surveillance Network |
| EPI_ISL_2274098, EPI_ISL_2274099 | Labortorio Central de Saude Publica do Estado do Rio de Janeiro (LACEN/RJ) | Laboratory of Respiratory Viruses and Measles, Oswaldo Cruz Institute, FIOCRUZ | Paola Resende, Luciana Appolinario, Fernando Motta, Anna Carolina Paixao, Ana Carolina Mendonca, Alice Sampaio Rocha, Taina Venas, Elisa Cavalcante Pereira, Renata Serrano Lopes, Andrea Cony Cavalcanti, Marilda Siqueira on behalf of the Fiocruz COVID-19 Genomic Surveillance Network |
| EPI_ISL_2274100, EPI_ISL_2274101, EPI_ISL_2274102, EPI_ISL_2274103, EPI_ISL_2274104, EPI_ISL_2274105, EPI_ISL_2274106, EPI_ISL_2274107 | Laboratory of Respiratory Viruses and Measles, Oswaldo Cruz Institute, FIOCRUZ | Laboratory of Respiratory Viruses and Measles, Oswaldo Cruz Institute, FIOCRUZ | Paola Resende, Luciana Appolinario, Fernando Motta, Anna Carolina Paixao, Ana Carolina Mendonca, Alice Sampaio Rocha, Taina Venas, Elisa Cavalcante Pereira, Renata Serrano Lopes, Marilda Siqueira on behalf of the Fiocruz COVID-19 Genomic Surveillance Network |
| EPI_ISL_2344442 | Instituto Butantan | Instituto de Medicina Tropical de Sao Paulo | Brazil-UK Centre for Arbovirus Discovery Diagnosis Genomics and Epidemiology (CADDE) Genomic Network - Instituto de Medicina Tropical |
| EPI_ISL_2385432, EPI_ISL_2385433, EPI_ISL_2385434, EPI_ISL_2385435, EPI_ISL_2385436, EPI_ISL_2385437, EPI_ISL_2385438, EPI_ISL_2385439, EPI_ISL_2385440, EPI_ISL_2385441, EPI_ISL_2385442, EPI_ISL_2385443, EPI_ISL_2385444, EPI_ISL_2385445, EPI_ISL_2385446, EPI_ISL_2385447, EPI_ISL_2385448, EPI_ISL_2385449, EPI_ISL_2385450, EPI_ISL_2385451, EPI_ISL_2385452, EPI_ISL_2385453, EPI_ISL_2385454, EPI_ISL_2385455, EPI_ISL_2385456, EPI_ISL_2385457, EPI_ISL_2385458, EPI_ISL_2385459, EPI_ISL_2385460, EPI_ISL_2385461, EPI_ISL_2385462, EPI_ISL_2385463, EPI_ISL_2385464, EPI_ISL_2385465, EPI_ISL_2385466, EPI_ISL_2385467, EPI_ISL_2385468, EPI_ISL_2385469, EPI_ISL_2385470, EPI_ISL_2385471, EPI_ISL_2385472, EPI_ISL_2385474, EPI_ISL_2385475, EPI_ISL_2385476, EPI_ISL_2385477, EPI_ISL_2385478, EPI_ISL_2385479, EPI_ISL_2385480, EPI_ISL_2385481, EPI_ISL_2385482, EPI_ISL_2385483, EPI_ISL_2385484, EPI_ISL_2385485, EPI_ISL_2385486, EPI_ISL_2385487, EPI_ISL_2385488, EPI_ISL_2385489, EPI_ISL_2385490, EPI_ISL_2385491, EPI_ISL_2385492, EPI_ISL_2385493, EPI_ISL_2385494, EPI_ISL_2385495, EPI_ISL_2385496, EPI_ISL_2385498, EPI_ISL_2385499, EPI_ISL_2385500, EPI_ISL_2385501, EPI_ISL_2385502, EPI_ISL_2385503, EPI_ISL_2385504, EPI_ISL_2385505, EPI_ISL_2385506, EPI_ISL_2385507, EPI_ISL_2385508, EPI_ISL_2385510, EPI_ISL_2385511, EPI_ISL_2385512, EPI_ISL_2385513, EPI_ISL_2385514, EPI_ISL_2385515, EPI_ISL_2385516, EPI_ISL_2385517, EPI_ISL_2385518, EPI_ISL_2385519, EPI_ISL_2385520, EPI_ISL_2385521, EPI_ISL_2385522, EPI_ISL_2385523, EPI_ISL_2385524, EPI_ISL_2385525, EPI_ISL_2385526, EPI_ISL_2385527, EPI_ISL_2385529, EPI_ISL_2385530, EPI_ISL_2385531, EPI_ISL_2385532, EPI_ISL_2385533, EPI_ISL_2385534, EPI_ISL_2385535, EPI_ISL_2385536, EPI_ISL_2385537, | | | |

| | | | |
|---|---|---|---|
| EPI_ISL_2385538, EPI_ISL_2385539, EPI_ISL_2385541, EPI_ISL_2385542, EPI_ISL_2385543, EPI_ISL_2385544, EPI_ISL_2385545, EPI_ISL_2385546, EPI_ISL_2385547, EPI_ISL_2385548, EPI_ISL_2385549, EPI_ISL_2385550, EPI_ISL_2385551, EPI_ISL_2385552, EPI_ISL_2385553, EPI_ISL_2385554, EPI_ISL_2385555, EPI_ISL_2385556, EPI_ISL_2385558, EPI_ISL_2385559, EPI_ISL_2385561, EPI_ISL_2385562, EPI_ISL_2385563, EPI_ISL_2385564, EPI_ISL_2385565, EPI_ISL_2385566, EPI_ISL_2385567, EPI_ISL_2385568, EPI_ISL_2385570, EPI_ISL_2385571, EPI_ISL_2385572, EPI_ISL_2385573, EPI_ISL_2385574, EPI_ISL_2385575, EPI_ISL_2385576, EPI_ISL_2385577, EPI_ISL_2385578, EPI_ISL_2385579, EPI_ISL_2385580, EPI_ISL_2385581, EPI_ISL_2385582, EPI_ISL_2385583, EPI_ISL_2385584, EPI_ISL_2385586, EPI_ISL_2385587, EPI_ISL_2385588, EPI_ISL_2385589, EPI_ISL_2385590, EPI_ISL_2385591, EPI_ISL_2385592, EPI_ISL_2385593, EPI_ISL_2385594, EPI_ISL_2385595, EPI_ISL_2385596, EPI_ISL_2385597, EPI_ISL_2385598, EPI_ISL_2385599, EPI_ISL_2385600, EPI_ISL_2385601, EPI_ISL_2385602, EPI_ISL_2385603, EPI_ISL_2385604, EPI_ISL_2385605, EPI_ISL_2385606, EPI_ISL_2385607, EPI_ISL_2385608, EPI_ISL_2385609, EPI_ISL_2385610, EPI_ISL_2385611, EPI_ISL_2385612, EPI_ISL_2385613, EPI_ISL_2385616, EPI_ISL_2385620, EPI_ISL_2385624, EPI_ISL_2385625, EPI_ISL_2385627, EPI_ISL_2385629, EPI_ISL_2385630, EPI_ISL_2385631, EPI_ISL_2385632, EPI_ISL_2385633, EPI_ISL_2385634, EPI_ISL_2385636, EPI_ISL_2385637, EPI_ISL_2385639, EPI_ISL_2385641, EPI_ISL_2385642, EPI_ISL_2385644, EPI_ISL_2385647, EPI_ISL_2385648, EPI_ISL_2385649, EPI_ISL_2385650, EPI_ISL_2385651, EPI_ISL_2385652, EPI_ISL_2385653, EPI_ISL_2385654, EPI_ISL_2385655, EPI_ISL_2385656, EPI_ISL_2385657, EPI_ISL_2385658, EPI_ISL_2385659, EPI_ISL_2385660, EPI_ISL_2385661, EPI_ISL_2385662, EPI_ISL_2385663, EPI_ISL_2385664, EPI_ISL_2385665, EPI_ISL_2385666, EPI_ISL_2385667, EPI_ISL_2385668 | | | |
| see above | Laboratorio Central Noel Nutels | Bioinformatics Laboratory / LNCC | Luiz G P de Almeida, Alessandra P Lamarca, Ronaldo da Silva F Jr, Liliane Cavalcante, Alexandra L Gerber, Ana Paula de C Guimaraes, Douglas Terra Machado, Cassia Alves, Diana Mariani, Cintia Policarpo, Gleidson da Silva de Oliveira, Mario Sergio Ribeiro, Silvia Carvalho, Flavio Dias da Silva, Marcio Henrique de Oliveira Garcia, Leandro Magalhaes de Souza, Cristiane Gomes da Silva, Caio Luiz Pereira Ribeiro, Andrea Cony Cavalcanti, Claudia Maria Braga de Mello, Amilcar Tanuri, Ana Tereza R Vasconcelos |
| EPI_ISL_2385669, EPI_ISL_2385670, EPI_ISL_2385671, EPI_ISL_2385674, EPI_ISL_2385675, EPI_ISL_2385676, EPI_ISL_2385677, EPI_ISL_2385678, EPI_ISL_2385679, EPI_ISL_2385680, EPI_ISL_2385681, EPI_ISL_2385682, EPI_ISL_2385683, EPI_ISL_2385684, EPI_ISL_2385685, EPI_ISL_2385686, EPI_ISL_2385689, EPI_ISL_2385690, EPI_ISL_2385691, EPI_ISL_2385692, EPI_ISL_2385693, EPI_ISL_2385694, EPI_ISL_2385695, EPI_ISL_2385696, EPI_ISL_2385697, EPI_ISL_2385698, EPI_ISL_2385699, EPI_ISL_2385700, EPI_ISL_2385701, EPI_ISL_2385702, EPI_ISL_2385704, EPI_ISL_2385705, EPI_ISL_2385707, EPI_ISL_2385708, EPI_ISL_2385709, EPI_ISL_2385711, EPI_ISL_2385712, EPI_ISL_2385713, EPI_ISL_2385714, EPI_ISL_2385717, EPI_ISL_2385718, EPI_ISL_2385719, EPI_ISL_2385720, EPI_ISL_2385723, EPI_ISL_2385724, EPI_ISL_2385725, EPI_ISL_2385726, EPI_ISL_2385727, EPI_ISL_2385728, EPI_ISL_2385729, EPI_ISL_2385730, EPI_ISL_2385731, EPI_ISL_2385732, EPI_ISL_2385733, EPI_ISL_2385734, EPI_ISL_2385735, EPI_ISL_2385736, EPI_ISL_2385737, EPI_ISL_2385739, EPI_ISL_2385740, EPI_ISL_2385741, EPI_ISL_2385742, EPI_ISL_2385743, EPI_ISL_2385745, EPI_ISL_2385748, EPI_ISL_2385749, EPI_ISL_2385750, EPI_ISL_2385751, EPI_ISL_2385752, EPI_ISL_2385753, EPI_ISL_2385754, EPI_ISL_2385755, EPI_ISL_2385756, EPI_ISL_2385757, EPI_ISL_2385758, EPI_ISL_2385759, EPI_ISL_2385760, EPI_ISL_2385761, EPI_ISL_2385764, EPI_ISL_2385765, EPI_ISL_2385766, EPI_ISL_2385768, EPI_ISL_2385769, EPI_ISL_2385770, EPI_ISL_2385771, EPI_ISL_2385772, EPI_ISL_2385773, EPI_ISL_2385774, EPI_ISL_2385775, EPI_ISL_2385776, EPI_ISL_2385777, EPI_ISL_2385778, EPI_ISL_2385779, EPI_ISL_2385780, EPI_ISL_2385781, EPI_ISL_2385782, EPI_ISL_2385784, EPI_ISL_2385785, EPI_ISL_2385786 | | | |
| see above | Unidade de apoio ao diagnóstico da COVID - UNADIG | Bioinformatics Laboratory / LNCC | Luiz G P de Almeida, Alessandra P Lamarca, Ronaldo da Silva F Jr, Liliane Cavalcante, Alexandra L Gerber, Ana Paula de C Guimaraes, Douglas Terra Machado, Cassia Alves, Diana Mariani, Cintia Policarpo, Gleidson da Silva de Oliveira, Mario Sergio Ribeiro, Silvia Carvalho, Flavio Dias da Silva, Marcio Henrique de Oliveira Garcia, Leandro Magalhaes de Souza, Cristiane Gomes da Silva, Caio Luiz Pereira Ribeiro, Andrea Cony Cavalcanti, Claudia Maria Braga de Mello, Amilcar Tanuri, Ana Tereza R Vasconcelos |
| EPI_ISL_2443578, EPI_ISL_2443579, EPI_ISL_2443580, EPI_ISL_2443581, EPI_ISL_2443582, EPI_ISL_2443583, EPI_ISL_2443584, EPI_ISL_2443585, EPI_ISL_2443586, EPI_ISL_2443587, EPI_ISL_2443588, EPI_ISL_2443589, EPI_ISL_2443590, EPI_ISL_2443591, EPI_ISL_2443592, EPI_ISL_2443593, EPI_ISL_2443594, EPI_ISL_2443595, EPI_ISL_2443596, EPI_ISL_2443597, EPI_ISL_2443598, EPI_ISL_2443599, EPI_ISL_2443600, EPI_ISL_2443601, EPI_ISL_2443602, EPI_ISL_2443603, EPI_ISL_2443604, EPI_ISL_2443605, EPI_ISL_2443606, EPI_ISL_2443607, EPI_ISL_2443608, EPI_ISL_2443609, EPI_ISL_2443610, EPI_ISL_2443611, EPI_ISL_2443612, EPI_ISL_2443613 | | | |
| see above | Laboratory of Respiratory Viruses and Measles, Oswaldo Cruz Institute, FIOCRUZ | Laboratory of Respiratory Viruses and Measles, Oswaldo Cruz Institute, FIOCRUZ | Paola Resende, Luciana Appolinario, Fernando Motta, Anna Carolina Paixao, Ana Carolina Mendonca, Alice Sampaio Rocha, Taina Venas, Elisa Cavalcante Pereira, Renata Serrano Lopes, Marilda Siqueira on behalf of the Fiocruz COVID-19 Genomic Surveillance Network |
| EPI_ISL_2443614, EPI_ISL_2443615, EPI_ISL_2443616, EPI_ISL_2443617, EPI_ISL_2443618, EPI_ISL_2443619, EPI_ISL_2443620, EPI_ISL_2443621, EPI_ISL_2443622, EPI_ISL_2443623, EPI_ISL_2443624, EPI_ISL_2443625, EPI_ISL_2443626, EPI_ISL_2443627, EPI_ISL_2443628, EPI_ISL_2443629, EPI_ISL_2443630, EPI_ISL_2443631, EPI_ISL_2443632 | | | |
| see above | Labortorio Central de Saude Publica do Estado do Rio de Janeiro (LACEN/RJ) | Laboratory of Respiratory Viruses and Measles, Oswaldo Cruz Institute, FIOCRUZ | Paola Resende, Luciana Appolinario, Fernando Motta, Anna Carolina Paixao, Ana Carolina Mendonca, Alice Sampaio Rocha, Taina Venas, Elisa Cavalcante Pereira, Renata Serrano Lopes, Andrea Cony Cavalcanti, Marilda Siqueira on behalf of the Fiocruz COVID-19 Genomic Surveillance Network |
| EPI_ISL_2443633, EPI_ISL_2443634, EPI_ISL_2443635, EPI_ISL_2443636 | Laboratory of Respiratory Viruses and Measles, Oswaldo Cruz Institute, FIOCRUZ | Laboratory of Respiratory Viruses and Measles, Oswaldo Cruz Institute, FIOCRUZ | Paola Resende, Luciana Appolinario, Fernando Motta, Anna Carolina Paixao, Ana Carolina Mendonca, Alice Sampaio Rocha, Taina Venas, Elisa Cavalcante Pereira, Renata Serrano Lopes, Marilda Siqueira on behalf of the Fiocruz COVID-19 Genomic Surveillance Network |
| EPI_ISL_2443637, EPI_ISL_2443638, EPI_ISL_2443639, EPI_ISL_2443640, EPI_ISL_2443641, EPI_ISL_2443642, EPI_ISL_2443643, EPI_ISL_2443644, EPI_ISL_2443645, EPI_ISL_2443646, EPI_ISL_2443647, EPI_ISL_2443648, EPI_ISL_2443649, EPI_ISL_2443650, EPI_ISL_2443651, EPI_ISL_2443652, EPI_ISL_2443653, EPI_ISL_2443654, EPI_ISL_2443655, EPI_ISL_2443656, EPI_ISL_2443657, EPI_ISL_2443658 | | | |
| see above | Labortorio Central de Saude Publica do Estado do Rio de Janeiro (LACEN/RJ) | Laboratory of Respiratory Viruses and Measles, Oswaldo Cruz Institute, FIOCRUZ | Paola Resende, Luciana Appolinario, Fernando Motta, Anna Carolina Paixao, Ana Carolina Mendonca, Alice Sampaio Rocha, Taina Venas, Elisa Cavalcante Pereira, Renata Serrano Lopes, Andrea Cony Cavalcanti, Marilda Siqueira on behalf of the Fiocruz COVID-19 Genomic Surveillance Network |
| EPI_ISL_2443659, EPI_ISL_2443660, EPI_ISL_2443661, EPI_ISL_2443662, EPI_ISL_2443663, EPI_ISL_2443664, EPI_ISL_2443665, EPI_ISL_2443666, EPI_ISL_2443667, EPI_ISL_2443668, EPI_ISL_2443669, EPI_ISL_2443670, EPI_ISL_2443671, EPI_ISL_2466122, EPI_ISL_2466123, EPI_ISL_2466124 | | | |
| see above | Laboratory of Respiratory Viruses and Measles, Oswaldo Cruz Institute, FIOCRUZ | Laboratory of Respiratory Viruses and Measles, Oswaldo Cruz Institute, FIOCRUZ | Paola Resende, Luciana Appolinario, Fernando Motta, Anna Carolina Paixao, Ana Carolina Mendonca, Alice Sampaio Rocha, Taina Venas, Elisa Cavalcante Pereira, Renata Serrano Lopes, Marilda Siqueira on behalf of the Fiocruz COVID-19 Genomic Surveillance Network |
| EPI_ISL_2466230 | Labortorio Central de Saude Publica do Estado do Rio de Janeiro (LACEN/RJ) | Laboratory of Respiratory Viruses and Measles, Oswaldo Cruz Institute, FIOCRUZ | Paola Resende, Luciana Appolinario, Fernando Motta, Anna Carolina Paixao, Ana Carolina Mendonca, Alice Sampaio Rocha, Taina Venas, Elisa Cavalcante Pereira, Renata Serrano Lopes, Andrea Cony Cavalcanti, Marilda Siqueira on behalf of the Fiocruz COVID-19 Genomic Surveillance Network |
| EPI_ISL_2466268 | Laboratorio Central de Saude Publica do Estado do Rio de Janeiro (LACEN-RJ) | Laboratory of Respiratory Viruses and Measles, Oswaldo Cruz Institute, FIOCRUZ | Paola Resende, Luciana Appolinario, Fernando Motta, Anna Carolina Paixao, Ana Carolina Mendonca, Alice Sampaio Rocha, Taina Venas, Elisa Cavalcante Pereira, Renata Serrano Lopes, Andrea Cony Cavalcanti, Marilda Siqueira on behalf of the Fiocruz COVID-19 Genomic Surveillance Network |
| EPI_ISL_2534862, EPI_ISL_2534863, EPI_ISL_2534864, EPI_ISL_2534865, EPI_ISL_2534866, EPI_ISL_2534867, EPI_ISL_2534868, EPI_ISL_2534869, EPI_ISL_2534870, EPI_ISL_2534871, EPI_ISL_2534872, EPI_ISL_2534873, EPI_ISL_2534874, EPI_ISL_2534875, EPI_ISL_2534876, EPI_ISL_2534877, EPI_ISL_2534878, EPI_ISL_2534879, EPI_ISL_2534880, EPI_ISL_2534881, EPI_ISL_2534882, EPI_ISL_2534883, EPI_ISL_2534884, EPI_ISL_2534885, EPI_ISL_2534886, EPI_ISL_2534887, EPI_ISL_2534888, EPI_ISL_2534889, EPI_ISL_2534890, EPI_ISL_2534891, EPI_ISL_2534892, EPI_ISL_2534893, EPI_ISL_2534894, EPI_ISL_2534895, EPI_ISL_2534896, EPI_ISL_2534897, EPI_ISL_2534898, EPI_ISL_2534899, EPI_ISL_2534900, EPI_ISL_2534901, EPI_ISL_2534902, EPI_ISL_2534903, EPI_ISL_2534904, EPI_ISL_2534905, EPI_ISL_2534906, EPI_ISL_2534907, EPI_ISL_2534908, EPI_ISL_2534909, EPI_ISL_2534910, EPI_ISL_2534911, EPI_ISL_2534912, EPI_ISL_2534913, EPI_ISL_2534914, EPI_ISL_2534915, EPI_ISL_2534916, EPI_ISL_2534917, EPI_ISL_2534918, EPI_ISL_2534919, EPI_ISL_2534920, EPI_ISL_2534921, EPI_ISL_2534922, EPI_ISL_2534924, EPI_ISL_2534925, EPI_ISL_2534926, EPI_ISL_2534927, EPI_ISL_2534928, EPI_ISL_2534929, EPI_ISL_2534930, EPI_ISL_2534931, EPI_ISL_2534933, EPI_ISL_2534934, EPI_ISL_2534935, EPI_ISL_2534936, EPI_ISL_2534937, EPI_ISL_2534938, EPI_ISL_2534939, EPI_ISL_2534940, EPI_ISL_2534941, EPI_ISL_2534942, EPI_ISL_2534943, EPI_ISL_2534944, EPI_ISL_2534946, EPI_ISL_2534947, EPI_ISL_2534948, EPI_ISL_2534949, EPI_ISL_2534950, EPI_ISL_2534951, EPI_ISL_2534952, EPI_ISL_2534955, EPI_ISL_2534956, EPI_ISL_2534957, EPI_ISL_2534958, EPI_ISL_2534959, EPI_ISL_2534960, EPI_ISL_2534961, EPI_ISL_2534962, EPI_ISL_2534963, EPI_ISL_2534964, EPI_ISL_2534965, EPI_ISL_2534966, EPI_ISL_2534967, EPI_ISL_2534968, EPI_ISL_2534969, EPI_ISL_2534970, EPI_ISL_2534971, EPI_ISL_2534972, EPI_ISL_2534973, EPI_ISL_2534974, EPI_ISL_2534975, EPI_ISL_2534976, EPI_ISL_2534977, EPI_ISL_2534978, EPI_ISL_2534979, EPI_ISL_2534980, EPI_ISL_2534981, EPI_ISL_2534982, EPI_ISL_2534983, EPI_ISL_2534984, EPI_ISL_2534985, EPI_ISL_2534986, EPI_ISL_2534987, EPI_ISL_2534988, EPI_ISL_2534989, EPI_ISL_2534990, EPI_ISL_2534991, EPI_ISL_2534992, EPI_ISL_2534993, EPI_ISL_2534994, EPI_ISL_2534996, EPI_ISL_2534997, EPI_ISL_2534999, EPI_ISL_2535000, EPI_ISL_2535001, EPI_ISL_2535002, EPI_ISL_2535003, EPI_ISL_2535004, EPI_ISL_2535005, EPI_ISL_2535006, EPI_ISL_2535007, EPI_ISL_2535009, EPI_ISL_2535010, EPI_ISL_2535011, EPI_ISL_2535012, EPI_ISL_2535013, EPI_ISL_2535014, EPI_ISL_2535015, EPI_ISL_2535016, EPI_ISL_2535018, EPI_ISL_2535019, EPI_ISL_2535021, EPI_ISL_2535022, EPI_ISL_2535023, EPI_ISL_2535024, EPI_ISL_2535025, EPI_ISL_2535026, EPI_ISL_2535027, EPI_ISL_2535028, EPI_ISL_2535029, EPI_ISL_2535030, EPI_ISL_2535031, EPI_ISL_2535032, EPI_ISL_2535033, EPI_ISL_2535034, EPI_ISL_2535035, EPI_ISL_2535036, EPI_ISL_2535037, EPI_ISL_2535038, EPI_ISL_2535039, EPI_ISL_2535040, EPI_ISL_2535041, EPI_ISL_2535043, EPI_ISL_2535044, EPI_ISL_2535045, EPI_ISL_2535046, EPI_ISL_2535047, EPI_ISL_2535048, EPI_ISL_2535049, EPI_ISL_2535050, EPI_ISL_2535051, EPI_ISL_2535052, EPI_ISL_2535053, EPI_ISL_2535054, EPI_ISL_2535055, EPI_ISL_2535056, EPI_ISL_2535057, EPI_ISL_2535058, EPI_ISL_2535059, EPI_ISL_2535060, EPI_ISL_2535061, EPI_ISL_2535062, EPI_ISL_2535063, EPI_ISL_2535064, EPI_ISL_2535065, EPI_ISL_2535066, EPI_ISL_2535067, EPI_ISL_2535068, EPI_ISL_2535069, EPI_ISL_2535070, EPI_ISL_2535071, EPI_ISL_2535072, EPI_ISL_2535073, EPI_ISL_2535074, EPI_ISL_2535075, EPI_ISL_2535076, EPI_ISL_2535077, EPI_ISL_2535078, EPI_ISL_2535080, EPI_ISL_2535081, EPI_ISL_2535082, EPI_ISL_2535083, EPI_ISL_2535084, EPI_ISL_2535085, EPI_ISL_2535086, EPI_ISL_2535087, EPI_ISL_2535088, EPI_ISL_2535089, EPI_ISL_2535090, EPI_ISL_2535091, EPI_ISL_2535092, EPI_ISL_2535093, EPI_ISL_2535094, EPI_ISL_2535095, EPI_ISL_2535096, EPI_ISL_2535097, EPI_ISL_2535098, EPI_ISL_2535099, EPI_ISL_2535100, EPI_ISL_2535101, EPI_ISL_2535102, EPI_ISL_2535103, EPI_ISL_2535104, EPI_ISL_2535105, EPI_ISL_2535106, EPI_ISL_2535107, EPI_ISL_2535108, EPI_ISL_2535109, EPI_ISL_2535110 | | | |
| see above | Laboratorio Central Noel Nutels | Bioinformatics Laboratory / LNCC | Luiz G P de Almeida, Alessandra P Lamarca, Ronaldo da Silva F Jr, Liliane Cavalcante, Alexandra L Gerber, Ana Paula de C Guimaraes, Douglas Terra Machado, Cassia Alves, Diana Mariani, Cintia Policarpo, Gleidson da Silva de Oliveira, Mario Sergio Ribeiro, Silvia Carvalho, Flavio Dias da Silva, Marcio Henrique de Oliveira Garcia, Leandro Magalhaes de Souza, Cristiane Gomes da Silva, Caio Luiz Pereira Ribeiro, Andrea Cony Cavalcanti, Claudia Maria Braga de Mello, Amilcar Tanuri, Ana Tereza R Vasconcelos |
| EPI_ISL_2535111, EPI_ISL_2535112, EPI_ISL_2535113, EPI_ISL_2535114, EPI_ISL_2535115, EPI_ISL_2535117, EPI_ISL_2535118, EPI_ISL_2535119, EPI_ISL_2535120, EPI_ISL_2535121, EPI_ISL_2535123, EPI_ISL_2535125, EPI_ISL_2535126, EPI_ISL_2535127, EPI_ISL_2535128, EPI_ISL_2535131, EPI_ISL_2535132, EPI_ISL_2535133, EPI_ISL_2535134, EPI_ISL_2535135, EPI_ISL_2535136, EPI_ISL_2535137, EPI_ISL_2535138, EPI_ISL_2535140, EPI_ISL_2535141, EPI_ISL_2535142, EPI_ISL_2535143, EPI_ISL_2535144, EPI_ISL_2535145, EPI_ISL_2535146, EPI_ISL_2535147, EPI_ISL_2535148, EPI_ISL_2535149, EPI_ISL_2535151, EPI_ISL_2535152, EPI_ISL_2535153, EPI_ISL_2535154, EPI_ISL_2535155, EPI_ISL_2535156, EPI_ISL_2535157, EPI_ISL_2535158, EPI_ISL_2535160, EPI_ISL_2535161, EPI_ISL_2535162, EPI_ISL_2535163, EPI_ISL_2535164, EPI_ISL_2535165, EPI_ISL_2535166, EPI_ISL_2535167, EPI_ISL_2535168, EPI_ISL_2535169, EPI_ISL_2535170, EPI_ISL_2535171, EPI_ISL_2535172, EPI_ISL_2535173, EPI_ISL_2535174, EPI_ISL_2535175, EPI_ISL_2535176, EPI_ISL_2535177, EPI_ISL_2535178, EPI_ISL_2535179, EPI_ISL_2535180, EPI_ISL_2535181, EPI_ISL_2535182, EPI_ISL_2535184, EPI_ISL_2535185, EPI_ISL_2535187, EPI_ISL_2535188, EPI_ISL_2535189, EPI_ISL_2535190, EPI_ISL_2535191, EPI_ISL_2535192, EPI_ISL_2535193, EPI_ISL_2535194, EPI_ISL_2535195, EPI_ISL_2535196, EPI_ISL_2535197, EPI_ISL_2535198, EPI_ISL_2535199, EPI_ISL_2535200, EPI_ISL_2535201, EPI_ISL_2535202, EPI_ISL_2535203, EPI_ISL_2535204, EPI_ISL_2535205, EPI_ISL_2535206, EPI_ISL_2535207, EPI_ISL_2535208, EPI_ISL_2535209, EPI_ISL_2535210, EPI_ISL_2535211, EPI_ISL_2535212, EPI_ISL_2535213, EPI_ISL_2535214, EPI_ISL_2535216, EPI_ISL_2535217, EPI_ISL_2535218, EPI_ISL_2535219, EPI_ISL_2535220, EPI_ISL_2535221, EPI_ISL_2535222, EPI_ISL_2535224, EPI_ISL_2535225, EPI_ISL_2535226, EPI_ISL_2535227, EPI_ISL_2535228, EPI_ISL_2535229, EPI_ISL_2535230, EPI_ISL_2535231, EPI_ISL_2535232, EPI_ISL_2535233, EPI_ISL_2535234, EPI_ISL_2535235 | | | |
| see above | Unidade de apoio ao diagnostico da COVID - UNADIG | Bioinformatics Laboratory / LNCC | Luiz G P de Almeida, Alessandra P Lamarca, Ronaldo da Silva F Jr, Liliane Cavalcante, Alexandra L Gerber, Ana Paula de C Guimaraes, Douglas Terra Machado, Cassia Alves, Diana Mariani, Cintia Policarpo, Gleidson da Silva de Oliveira, Mario Sergio Ribeiro, Silvia Carvalho, Flavio Dias da Silva, Marcio Henrique de Oliveira Garcia, Leandro Magalhaes de Souza, Cristiane Gomes da Silva, Caio Luiz Pereira Ribeiro, Andrea Cony Cavalcanti, Claudia Maria Braga de Mello, Amilcar Tanuri, Ana Tereza R Vasconcelos |
| EPI_ISL_2536269, EPI_ISL_2536270, EPI_ISL_2536271, EPI_ISL_2536272, EPI_ISL_2536273, EPI_ISL_2536274 | Laboratory of Respiratory Viruses and Measles, Oswaldo Cruz Institute, FIOCRUZ | Laboratory of Respiratory Viruses and Measles, Oswaldo Cruz Institute, FIOCRUZ | Paola Resende, Patricia Brasil, Luciana Appolinario, Fernando Motta, Anna Carolina Paixao, Ana Carolina Mendonca, Alice Sampaio Rocha, Taina Venas, Elisa Cavalcante Pereira, Renata Serrano Lopes, Marilda Siqueira on behalf of the Fiocruz COVID-19 Genomic Surveillance Network |
| EPI_ISL_2536275, EPI_ISL_2536276, EPI_ISL_2536277, EPI_ISL_2536278, EPI_ISL_2536279, EPI_ISL_2536281, EPI_ISL_2536282, EPI_ISL_2536283, EPI_ISL_2536284, EPI_ISL_2536285, EPI_ISL_2536286, EPI_ISL_2536287, EPI_ISL_2536288, EPI_ISL_2536289, EPI_ISL_2536290, EPI_ISL_2536291, EPI_ISL_2536292, EPI_ISL_2536293, EPI_ISL_2536294, EPI_ISL_2536295, EPI_ISL_2536296, EPI_ISL_2536297, EPI_ISL_2536298, EPI_ISL_2536299, EPI_ISL_2536300, EPI_ISL_2536301, EPI_ISL_2536302, EPI_ISL_2557376, EPI_ISL_2557377, EPI_ISL_2557378, EPI_ISL_2557379, EPI_ISL_2557380, EPI_ISL_2557381, EPI_ISL_2557382, | | | |

| | | | |
|---|---|---|---|
| EPI_ISL_2557383, EPI_ISL_2557384 | | | |
| see above | Labortorio Central de Saude Publica do Estado do Rio de Janeiro (LACEN/RJ) | Laboratory of Respiratory Viruses and Measles, Oswaldo Cruz Institute, FIOCRUZ | Paola Resende, Luciana Appolinario, Fernando Motta, Anna Carolina Paixao, Ana Carolina Mendonca, Alice Sampaio Rocha, Taina Venas, Elisa Cavalcante Pereira, Renata Serrano Lopes, Andrea Cony Cavalcanti, Marilda Siqueira on behalf of the Fiocruz COVID-19 Genomic Surveillance Network |
| EPI_ISL_2557385, EPI_ISL_2557386, EPI_ISL_2557387, EPI_ISL_2557388, EPI_ISL_2557389, EPI_ISL_2557390, EPI_ISL_2557391, EPI_ISL_2557392, EPI_ISL_2557393, EPI_ISL_2557394, EPI_ISL_2557395, EPI_ISL_2557396, EPI_ISL_2557397, EPI_ISL_2557398, EPI_ISL_2557399, EPI_ISL_2557400, EPI_ISL_2557401, EPI_ISL_2557402, EPI_ISL_2557403 | | | |
| see above | Laboratory of Respiratory Viruses and Measles, Oswaldo Cruz Institute, FIOCRUZ | Laboratory of Respiratory Viruses and Measles, Oswaldo Cruz Institute, FIOCRUZ | Paola Resende, Luciana Appolinario, Fernando Motta, Anna Carolina Paixao, Ana Carolina Mendonca, Alice Sampaio Rocha, Taina Venas, Elisa Cavalcante Pereira, Renata Serrano Lopes, Marilda Siqueira on behalf of the Fiocruz COVID-19 Genomic Surveillance Network |
| EPI_ISL_2557404 | Labortorio Central de Saude Publica do Estado do Rio de Janeiro (LACEN/RJ) | Laboratory of Respiratory Viruses and Measles, Oswaldo Cruz Institute, FIOCRUZ | Paola Resende, Luciana Appolinario, Fernando Motta, Anna Carolina Paixao, Ana Carolina Mendonca, Alice Sampaio Rocha, Taina Venas, Elisa Cavalcante Pereira, Renata Serrano Lopes, Andrea Cony Cavalcanti, Marilda Siqueira on behalf of the Fiocruz COVID-19 Genomic Surveillance Network |
| EPI_ISL_2557405 | Laboratory of Respiratory Viruses and Measles, Oswaldo Cruz Institute, FIOCRUZ | Laboratory of Respiratory Viruses and Measles, Oswaldo Cruz Institute, FIOCRUZ | Paola Resende, Luciana Appolinario, Fernando Motta, Anna Carolina Paixao, Ana Carolina Mendonca, Alice Sampaio Rocha, Taina Venas, Elisa Cavalcante Pereira, Renata Serrano Lopes, Marilda Siqueira on behalf of the Fiocruz COVID-19 Genomic Surveillance Network |
| EPI_ISL_2557406 | Labortorio Central de Saude Publica do Estado do Rio de Janeiro (LACEN/RJ) | Laboratory of Respiratory Viruses and Measles, Oswaldo Cruz Institute, FIOCRUZ | Paola Resende, Luciana Appolinario, Fernando Motta, Anna Carolina Paixao, Ana Carolina Mendonca, Alice Sampaio Rocha, Taina Venas, Elisa Cavalcante Pereira, Renata Serrano Lopes, Andrea Cony Cavalcanti, Marilda Siqueira on behalf of the Fiocruz COVID-19 Genomic Surveillance Network |
| EPI_ISL_2557407, EPI_ISL_2557408, EPI_ISL_2557409, EPI_ISL_2557410, EPI_ISL_2603472, EPI_ISL_2603473, EPI_ISL_2603474, EPI_ISL_2603475, EPI_ISL_2603476, EPI_ISL_2603477, EPI_ISL_2603478, EPI_ISL_2603479, EPI_ISL_2603480, EPI_ISL_2603482, EPI_ISL_2603483, EPI_ISL_2603484, EPI_ISL_2603485, EPI_ISL_2603486, EPI_ISL_2603487, EPI_ISL_2603488, EPI_ISL_2603489, EPI_ISL_2603490, EPI_ISL_2603491, EPI_ISL_2603492, EPI_ISL_2603493, EPI_ISL_2603494, EPI_ISL_2603495, EPI_ISL_2603496, EPI_ISL_2603497, EPI_ISL_2603498, EPI_ISL_2603499, EPI_ISL_2603500, EPI_ISL_2603501, EPI_ISL_2603502, EPI_ISL_2603503, EPI_ISL_2603504, EPI_ISL_2603505, EPI_ISL_2603506, EPI_ISL_2603507, EPI_ISL_2603508, EPI_ISL_2603509, EPI_ISL_2603510, EPI_ISL_2603511, EPI_ISL_2603512, EPI_ISL_2603513, EPI_ISL_2603514, EPI_ISL_2603515, EPI_ISL_2603516, EPI_ISL_2603517, EPI_ISL_2603518, EPI_ISL_2603519, EPI_ISL_2603520 | | | |
| see above | Laboratory of Respiratory Viruses and Measles, Oswaldo Cruz Institute, FIOCRUZ | Laboratory of Respiratory Viruses and Measles, Oswaldo Cruz Institute, FIOCRUZ | Paola Resende, Luciana Appolinario, Fernando Motta, Anna Carolina Paixao, Ana Carolina Mendonca, Alice Sampaio Rocha, Taina Venas, Elisa Cavalcante Pereira, Renata Serrano Lopes, Marilda Siqueira on behalf of the Fiocruz COVID-19 Genomic Surveillance Network |
| EPI_ISL_2614070, EPI_ISL_2614071, EPI_ISL_2614072, EPI_ISL_2614073, EPI_ISL_2614074, EPI_ISL_2614075, EPI_ISL_2614076, EPI_ISL_2614077, EPI_ISL_2614078, EPI_ISL_2614079, EPI_ISL_2614080, EPI_ISL_2614081, EPI_ISL_2614082, EPI_ISL_2614083, EPI_ISL_2614084, EPI_ISL_2614085, EPI_ISL_2614086, EPI_ISL_2614087, EPI_ISL_2614088, EPI_ISL_2614089, EPI_ISL_2614090, EPI_ISL_2614091 | | | |
| see above | Laboratory of Molecular Virology, Federal University of Rio de Janeito, UFRJ | Laboratory of Respiratory Viruses and Measles, Oswaldo Cruz Institute, FIOCRUZ | Paola Resende, Amilcar Tanuri, Luciana Appolinario, Fernando Motta, Anna Carolina Paixao, Ana Carolina Mendonca, Alice Sampaio Rocha, Taina Venas, Elisa Cavalcante Pereira, Renata Serrano Lopes, Marilda Siqueira on behalf of the Fiocruz COVID-19 Genomic Surveillance Network |
| EPI_ISL_2614092, EPI_ISL_2614093, EPI_ISL_2614094, EPI_ISL_2614095, EPI_ISL_2614096, EPI_ISL_2614097, EPI_ISL_2614098, EPI_ISL_2614099, EPI_ISL_2614100, EPI_ISL_2614101, EPI_ISL_2614102, EPI_ISL_2614103, EPI_ISL_2614104, EPI_ISL_2614105, EPI_ISL_2614106, EPI_ISL_2614107, EPI_ISL_2614108, EPI_ISL_2614109, EPI_ISL_2614110, EPI_ISL_2614111, EPI_ISL_2614112, EPI_ISL_2614113, EPI_ISL_2614114 | | | |
| see above | Laboratory of Respiratory Viruses and Measles, Oswaldo Cruz Institute, FIOCRUZ | Laboratory of Respiratory Viruses and Measles, Oswaldo Cruz Institute, FIOCRUZ | Paola Resende, Luciana Appolinario, Fernando Motta, Anna Carolina Paixao, Ana Carolina Mendonca, Alice Sampaio Rocha, Taina Venas, Elisa Cavalcante Pereira, Renata Serrano Lopes, Marilda Siqueira on behalf of the Fiocruz COVID-19 Genomic Surveillance Network |
| EPI_ISL_2614115 | Laboratory of Respiratory Viruses and Measles, Oswaldo Cruz Institute, FIOCRUZ | Laboratory of Respiratory Viruses and Measles, Oswaldo Cruz Institute, FIOCRUZ | Paola Resende, Amilcar Tanuri, Luciana Appolinario, Fernando Motta, Anna Carolina Paixao, Ana Carolina Mendonca, Alice Sampaio Rocha, Taina Venas, Elisa Cavalcante Pereira, Renata Serrano Lopes, Marilda Siqueira on behalf of the Fiocruz COVID-19 Genomic Surveillance Network |
| EPI_ISL_2614116, EPI_ISL_2614117, EPI_ISL_2614118, EPI_ISL_2614119, EPI_ISL_2614120, EPI_ISL_2614121, EPI_ISL_2614122, EPI_ISL_2614123, EPI_ISL_2614124, EPI_ISL_2614125, EPI_ISL_2614126, EPI_ISL_2614127, EPI_ISL_2614128, EPI_ISL_2614129, EPI_ISL_2614130, EPI_ISL_2614131, EPI_ISL_2614132, EPI_ISL_2614133, EPI_ISL_2614134, EPI_ISL_2614135, EPI_ISL_2614136, EPI_ISL_2614137, EPI_ISL_2614138, EPI_ISL_2614139, EPI_ISL_2614140, EPI_ISL_2614141, EPI_ISL_2614142, EPI_ISL_2614143, EPI_ISL_2614144, EPI_ISL_2614145, EPI_ISL_2614146, EPI_ISL_2614147, EPI_ISL_2614148, EPI_ISL_2614149, EPI_ISL_2614150, EPI_ISL_2614151, EPI_ISL_2614152, EPI_ISL_2614153, EPI_ISL_2614154, EPI_ISL_2614155, EPI_ISL_2614156, EPI_ISL_2614157, EPI_ISL_2614158, EPI_ISL_2614159, EPI_ISL_2614160, EPI_ISL_2614161, EPI_ISL_2614162, EPI_ISL_2614163, EPI_ISL_2614164, EPI_ISL_2614165, EPI_ISL_2614166, EPI_ISL_2614167, EPI_ISL_2614168, EPI_ISL_2614169, EPI_ISL_2614170, EPI_ISL_2614171, EPI_ISL_2614172, EPI_ISL_2614173, EPI_ISL_2614174, EPI_ISL_2614175, EPI_ISL_2614176, EPI_ISL_2614177, EPI_ISL_2614178, EPI_ISL_2614181, EPI_ISL_2614182, EPI_ISL_2614183, EPI_ISL_2614184, EPI_ISL_2614185, EPI_ISL_2614186, EPI_ISL_2614187, EPI_ISL_2614188, EPI_ISL_2614189, EPI_ISL_2614190, EPI_ISL_2614191, EPI_ISL_2614192, EPI_ISL_2614193, EPI_ISL_2614194, EPI_ISL_2614195, EPI_ISL_2614196, EPI_ISL_2614197, EPI_ISL_2614198, EPI_ISL_2614199, EPI_ISL_2614200, EPI_ISL_2614201, EPI_ISL_2614202, EPI_ISL_2614203, EPI_ISL_2614204, EPI_ISL_2614205, EPI_ISL_2614206, EPI_ISL_2614207, EPI_ISL_2614209, EPI_ISL_2614210, EPI_ISL_2614211, EPI_ISL_2614212, EPI_ISL_2614213, EPI_ISL_2614214, EPI_ISL_2614215, EPI_ISL_2614216, EPI_ISL_2614217, EPI_ISL_2614218, EPI_ISL_2614219, EPI_ISL_2614220, EPI_ISL_2614221, EPI_ISL_2614222, EPI_ISL_2614223, EPI_ISL_2614224, EPI_ISL_2614225, EPI_ISL_2614226, EPI_ISL_2614227, EPI_ISL_2614228, EPI_ISL_2614229, EPI_ISL_2614230, EPI_ISL_2614231, EPI_ISL_2614232, EPI_ISL_2614233, EPI_ISL_2614234, EPI_ISL_2614235, EPI_ISL_2614236, EPI_ISL_2614237, EPI_ISL_2614238, EPI_ISL_2614239, EPI_ISL_2614240, EPI_ISL_2614241, EPI_ISL_2614242, EPI_ISL_2614243, EPI_ISL_2614244, EPI_ISL_2614245, EPI_ISL_2614246, EPI_ISL_2614247, EPI_ISL_2614248, EPI_ISL_2614249, EPI_ISL_2614250, EPI_ISL_2614251, EPI_ISL_2614252, EPI_ISL_2614253, EPI_ISL_2614254, EPI_ISL_2614255, EPI_ISL_2614256, EPI_ISL_2614257, EPI_ISL_2614258, EPI_ISL_2614259, EPI_ISL_2614260, EPI_ISL_2614261, EPI_ISL_2614262, EPI_ISL_2614263, EPI_ISL_2614264, EPI_ISL_2614265, EPI_ISL_2614266, EPI_ISL_2614267, EPI_ISL_2614268, EPI_ISL_2614269, EPI_ISL_2614270, EPI_ISL_2614271, EPI_ISL_2614272, EPI_ISL_2614273, EPI_ISL_2614274, EPI_ISL_2614275, EPI_ISL_2614276, EPI_ISL_2614277, EPI_ISL_2614278, EPI_ISL_2614279, EPI_ISL_2614280, EPI_ISL_2614281, EPI_ISL_2614282, EPI_ISL_2614283, EPI_ISL_2614284, EPI_ISL_2614285, EPI_ISL_2614286, EPI_ISL_2614287, EPI_ISL_2614288, EPI_ISL_2614289, EPI_ISL_2614290, EPI_ISL_2614291, EPI_ISL_2614292, EPI_ISL_2614293, EPI_ISL_2614294, EPI_ISL_2614295, EPI_ISL_2614296, EPI_ISL_2614297, EPI_ISL_2614298, EPI_ISL_2614299, EPI_ISL_2614300, EPI_ISL_2614301, EPI_ISL_2614302, EPI_ISL_2614303, EPI_ISL_2614304, EPI_ISL_2614305, EPI_ISL_2614306, EPI_ISL_2614307, EPI_ISL_2614308, EPI_ISL_2614309, EPI_ISL_2614310, EPI_ISL_2614311, EPI_ISL_2614312, EPI_ISL_2614313, EPI_ISL_2614314, EPI_ISL_2614315, EPI_ISL_2614316, EPI_ISL_2614317, EPI_ISL_2614318, EPI_ISL_2614319, EPI_ISL_2614320, EPI_ISL_2614321, EPI_ISL_2614322, EPI_ISL_2614323, EPI_ISL_2614324, EPI_ISL_2614325, EPI_ISL_2614326, EPI_ISL_2614327, EPI_ISL_2614328, EPI_ISL_2614329, EPI_ISL_2614330, EPI_ISL_2614331, EPI_ISL_2614332, EPI_ISL_2614333, EPI_ISL_2614334, EPI_ISL_2614335, EPI_ISL_2614336, EPI_ISL_2614337, EPI_ISL_2614338, EPI_ISL_2614339, EPI_ISL_2614340, EPI_ISL_2614341, EPI_ISL_2614342, EPI_ISL_2614343, EPI_ISL_2614344, EPI_ISL_2614345, EPI_ISL_2614346, EPI_ISL_2614347, EPI_ISL_2614348, EPI_ISL_2614349, EPI_ISL_2614350 | | | |
| see above | Laboratory of Respiratory Viruses and Measles, Oswaldo Cruz Institute, FIOCRUZ | Laboratory of Respiratory Viruses and Measles, Oswaldo Cruz Institute, FIOCRUZ | Paola Resende, Luciana Appolinario, Fernando Motta, Anna Carolina Paixao, Ana Carolina Mendonca, Alice Sampaio Rocha, Taina Venas, Elisa Cavalcante Pereira, Renata Serrano Lopes, Marilda Siqueira on behalf of the Fiocruz COVID-19 Genomic Surveillance Network |
| EPI_ISL_2614351, EPI_ISL_2614352, EPI_ISL_2614353, EPI_ISL_2614354, EPI_ISL_2614355, EPI_ISL_2614356, EPI_ISL_2614357, EPI_ISL_2614358, EPI_ISL_2614359, EPI_ISL_2614360, EPI_ISL_2614361, EPI_ISL_2614362, EPI_ISL_2614363, EPI_ISL_2614364, EPI_ISL_2614365, EPI_ISL_2614366, EPI_ISL_2614367, EPI_ISL_2614368, EPI_ISL_2614369, EPI_ISL_2614370, EPI_ISL_2614371, EPI_ISL_2614372, EPI_ISL_2614373, EPI_ISL_2614374, EPI_ISL_2614375, EPI_ISL_2614376, EPI_ISL_2614377, EPI_ISL_2614378, EPI_ISL_2614379, EPI_ISL_2614380 | | | |
| see above | Labortorio Central de Saude Publica do Estado do Rio de Janeiro (LACEN/RJ) | Laboratory of Respiratory Viruses and Measles, Oswaldo Cruz Institute, FIOCRUZ | Paola Resende, Luciana Appolinario, Fernando Motta, Anna Carolina Paixao, Ana Carolina Mendonca, Alice Sampaio Rocha, Taina Venas, Elisa Cavalcante Pereira, Renata Serrano Lopes, Andrea Cony Cavalcanti, Marilda Siqueira on behalf of the Fiocruz COVID-19 Genomic Surveillance Network |
| EPI_ISL_2614381, EPI_ISL_2614382, EPI_ISL_2614383 | Instituto Estadual do Cerebro Paulo Niemeyer | Laboratory of Respiratory Viruses and Measles, Oswaldo Cruz Institute, FIOCRUZ | Paola Resende,Carlos Azevedo, Luciana Appolinario, Fernando Motta, Anna Carolina Paixao, Ana Carolina Mendonca, Alice Sampaio Rocha, Taina Venas, Elisa Cavalcante Pereira, Renata Serrano Lopes, Marilda Siqueira on behalf of the Fiocruz COVID-19 Genomic Surveillance Network |
| EPI_ISL_406798 | General Hospital of Central Theater Command of People's Liberation Army of China | BGI & Institute of Microbiology, Chinese Academy of Sciences & Shandong First Medical University & Shandong Academy of Medical Sciences & General Hospital of Central Theater Command of People's Liberation Army of China | Weijun Chen, Yuhai Bi, Weifeng Shi and Zhenhong Hu |
| EPI_ISL_414045 | Laboratório Central de Saúde Pública Noel Nutels (LACEN-RJ) | Laboratory of Respiratory Viruses and Measles, Oswaldo Cruz Institute, FIOCRUZ | Paola Resende, Alisson Fabri, Joilson Xavier, Sunando Roy, Fernando Motta, Aline Mattos, Milene Miranda, Cristiana Garcia, Braulia Caetano, Maria Ogrzewalska, Jonathan Lopes, Luciana Appolinario, Maria Nóbrega, Marilda SiqueiraFiocruz COVID-19 Genomic Surveillance Network |
| EPI_ISL_427299, EPI_ISL_427300, EPI_ISL_427301, EPI_ISL_427302, EPI_ISL_427303, EPI_ISL_427304 | Laboratory of Respiratory Viruses and Measles, Oswaldo Cruz Institute, FIOCRUZ | Laboratory of Respiratory Viruses and Measles, Oswaldo Cruz Institute, FIOCRUZ | Paola Resende, Fernando Motta, Luciana Appolinario, Sunando Roy, Aline Mattos, Milene Miranda, Cristiana Garcia, Braulia Caetano, Maria Ogrzewalska, Priscila Born, Jonathan Lopes, Marilda Siqueira on behalf of the Fiocruz COVID-19 Genomic Surveillance Network |
| EPI_ISL_456071, EPI_ISL_456072, EPI_ISL_456073, EPI_ISL_456074, EPI_ISL_456075 | Laboratory of Respiratory Viruses and Measles, Oswaldo Cruz Institute, FIOCRUZ | Laboratory of Respiratory Viruses and Measles, Oswaldo Cruz Institute, FIOCRUZ | Paola Resende, Luciana Appolinario, Fernando Motta, Aline Mattos, Milene Miranda, Cristiana Garcia, Braulia Caetano, Maria Ogrzewalska, Jonathan Lopes, Marilda Siqueira on behalf of the Fiocruz COVID-19 Genomic Surveillance Network |
| EPI_ISL_456076, EPI_ISL_456077 | Laboratório Central de Saúde Pública Noel Nutels (LACEN-RJ) | Laboratory of Respiratory Viruses and Measles, Oswaldo Cruz Institute, FIOCRUZ | Paola Resende, Luciana Appolinario, Fernando Motta, Aline Mattos, Milene Miranda, Cristiana Garcia, Braulia Caetano, Maria Ogrzewalska, Jonathan Lopes, Marilda Siqueira on behalf of the Fiocruz COVID-19 Genomic Surveillance Network |
| EPI_ISL_456079, EPI_ISL_456080, EPI_ISL_456081 | Laboratory of Respiratory Viruses and Measles, Oswaldo Cruz Institute, FIOCRUZ | Laboratory of Respiratory Viruses and Measles, Oswaldo Cruz Institute, FIOCRUZ | Paola Resende, Luciana Appolinario, Fernando Motta, Aline Mattos, Milene Miranda, Cristiana Garcia, Braulia Caetano, Maria Ogrzewalska, Jonathan Lopes, Marilda Siqueira on behalf of the Fiocruz COVID-19 Genomic Surveillance Network |
| EPI_ISL_456082, EPI_ISL_456083 | Laboratório Central de Saúde Pública Noel Nutels (LACEN-RJ) | Laboratory of Respiratory Viruses and Measles, Oswaldo Cruz Institute, FIOCRUZ | Paola Resende, Luciana Appolinario, Fernando Motta, Aline Mattos, Milene Miranda, Cristiana Garcia, Braulia Caetano, Maria Ogrzewalska, Jonathan Lopes, Marilda Siqueira on behalf of the Fiocruz COVID-19 Genomic Surveillance Network |
| EPI_ISL_456084, EPI_ISL_456085, EPI_ISL_456086, EPI_ISL_456087 | Laboratory of Respiratory Viruses and Measles, Oswaldo Cruz Institute, FIOCRUZ | Laboratory of Respiratory Viruses and Measles, Oswaldo Cruz Institute, FIOCRUZ | Paola Resende, Luciana Appolinario, Fernando Motta, Aline Mattos, Milene Miranda, Cristiana Garcia, Braulia Caetano, Maria Ogrzewalska, Jonathan Lopes, Marilda Siqueira on behalf of the Fiocruz COVID-19 Genomic Surveillance Network |
| EPI_ISL_456088 | Laboratório Central de Saúde Pública Noel Nutels (LACEN-RJ) | Laboratory of Respiratory Viruses and Measles, Oswaldo Cruz Institute, FIOCRUZ | Paola Resende, Luciana Appolinario, Fernando Motta, Aline Mattos, Milene Miranda, Cristiana Garcia, Braulia Caetano, Maria Ogrzewalska, Jonathan Lopes, Marilda Siqueira on behalf of the Fiocruz COVID-19 Genomic Surveillance Network |

| | | | |
|---|---|---|---|
| EPI_ISL_456089, EPI_ISL_456090, EPI_ISL_456091, EPI_ISL_456092, EPI_ISL_456093, EPI_ISL_456094, EPI_ISL_456095, EPI_ISL_456096, EPI_ISL_456097, EPI_ISL_456098, EPI_ISL_456099, EPI_ISL_456100, EPI_ISL_456101, EPI_ISL_456102, EPI_ISL_456103, EPI_ISL_456104, EPI_ISL_456105, EPI_ISL_456106 | | | |
| see above | Laboratory of Respiratory Viruses and Measles, Oswaldo Cruz Institute, FIOCRUZ | Laboratory of Respiratory Viruses and Measles, Oswaldo Cruz Institute, FIOCRUZ | Paola Resende, Luciana Appolinario, Fernando Motta, Aline Mattos, Milene Miranda, Cristiana Garcia, Braulia Caetano, Maria Ogrzewalska, Jonathan Lopes, Marilda Siqueira on behalf of the Fiocruz COVID-19 Genomic Surveillance Network |
| EPI_ISL_467344, EPI_ISL_467345, EPI_ISL_467346, EPI_ISL_467347, EPI_ISL_467348, EPI_ISL_467349, EPI_ISL_467350, EPI_ISL_467351, EPI_ISL_467352, EPI_ISL_467353, EPI_ISL_467354, EPI_ISL_467355, EPI_ISL_467356, EPI_ISL_467357, EPI_ISL_467358, EPI_ISL_467359, EPI_ISL_467360, EPI_ISL_467362, EPI_ISL_467363, EPI_ISL_467364, EPI_ISL_467365, EPI_ISL_467366, EPI_ISL_467367, EPI_ISL_467368, EPI_ISL_467369, EPI_ISL_467370, EPI_ISL_467371 | | | |
| see above | Laboratory of Respiratory Viruses and Measles, Oswaldo Cruz Institute, FIOCRUZ | Laboratory of Respiratory Viruses and Measles, Oswaldo Cruz Institute, FIOCRUZ | Paola Resende, Luciana Appolinario, Fernando Motta, Anna Carolina Paixão, Ana Carolina Mendonça, Aline Mattos, Milene Miranda, Cristiana Garcia, Braulia Caetano, Maria Ogrzewalska, Jonathan Lopes, Marilda Siqueira on behalf of the Fiocruz COVID-19 Genomic Surveillance Network |
| EPI_ISL_483065 | Centro de Desenvolvimento Tecnologico em Saude, Fundacao Oswaldo Cruz | Centro de Desenvolvimento Tecnologico em Saude, Fundacao Oswaldo Cruz | Souza,T.M., Fintelman-Rodrigues,N., De Paula,A.D., Tschoeke,D., Barroso,S.P., Gregorio,M.L., Oliveira,J.S., Saraiva,F.B., Ferreira,M.A., Sacramento,C.Q. |
| EPI_ISL_492032 | Instituto de Biologia do Exército | Laboratório Metabolismo Macromolecular FirminoTorres de Castro, Instituto de Biofísica Carlos Chagas Filho, Universidade Federal do Rio de Janeiro | Bianca Catarina Azevedo Cabral, Aline Rosa Vianna de Souza , Marcos Dornelas-Ribeiro, Tatiana LS Nogueira, Nádia Vaez Gonçalves da Cruz, Caleb GM Santos, Elizabeth Valentin, Marcio da Costa Cipitelli, Virginia Sara Grancieri do Amaral, Rodrigo Soares de Moura Neto, Clarissa Damaso, Rosane Silva |
| EPI_ISL_492033 | Instituto de Biologia do Exército | Laboratório Metabolismo Macromolecular FirminoTorres de Castro, Instituto de Biofísica Carlos Chagas Filho, Universidade Federal do Rio de Janeiro | Bianca Catarina Azevedo Cabral, Aline Rosa Vianna de Souza, Caleb GM Santos, Marcos Dornelas-Ribeiro, Tatiana LS Nogueira, Nádia Vaez Gonçalves da Cruz, Elizabeth Valentin, Marcio da Costa Cipitelli, Virginia Sara Grancieri do Amaral, Rodrigo Soares de Moura Neto, Clarissa Damaso, Rosane Silva |
| EPI_ISL_492034 | Instituto de Biologia do Exército | Laboratório Metabolismo Macromolecular FirminoTorres de Castro, Instituto de Biofísica Carlos Chagas Filho, Universidade Federal do Rio de Janeiro | Bianca Catarina Azevedo Cabral, Aline Rosa Vianna de Souza, Nádia Vaez Gonçalves da Cruz, Caleb GM Santos, Marcos Dornelas-Ribeiro, Tatiana LS Nogueira, Elizabeth Valentin, Marcio da Costa Cipitelli, Virginia Sara Grancieri do Amaral, Rodrigo Soares de Moura Neto, Clarissa Damaso, Rosane Silva |
| EPI_ISL_492035 | Instituto de Biologia do Exército | Laboratório Metabolismo Macromolecular FirminoTorres de Castro, Instituto de Biofísica Carlos Chagas Filho, Universidade Federal do Rio de Janeiro | Bianca Catarina Azevedo Cabral, Aline Rosa Vianna de Souza, Tatiana LS Nogueira, Nádia Vaez Gonçalves da Cruz, Caleb GM Santos, Marcos Dornelas-Ribeiro, Elizabeth Valentin, Marcio da Costa Cipitelli, Virginia Sara Grancieri do Amaral, Rodrigo Soares de Moura Neto, Clarissa Damaso, Rosane Silva |
| EPI_ISL_492036 | Instituto de Biologia do Exército | Laboratório Metabolismo Macromolecular FirminoTorres de Castro, Instituto de Biofísica Carlos Chagas Filho, Universidade Federal do Rio de Janeiro | Bianca Catarina Azevedo Cabral, Aline Rosa Vianna de Souza , Marcos Dornelas-Ribeiro, Tatiana LS Nogueira, Nádia Vaez Gonçalves da Cruz, Caleb GM Santos, Elizabeth Valentin, Marcio da Costa Cipitelli, Virginia Sara Grancieri do Amaral, Rodrigo Soares de Moura Neto, Clarissa Damaso, Rosane Silva |
| EPI_ISL_492037 | Instituto de Biologia do Exército | Laboratório Metabolismo Macromolecular FirminoTorres de Castro, Instituto de Biofísica Carlos Chagas Filho, Universidade Federal do Rio de Janeiro | Bianca Catarina Azevedo Cabral, Aline Rosa Vianna de Souza, Caleb GM Santos, Marcos Dornelas-Ribeiro, Tatiana LS Nogueira, Nádia Vaez Gonçalves da Cruz, Elizabeth Valentin, Marcio da Costa Cipitelli, Virginia Sara Grancieri do Amaral, Rodrigo Soares de Moura Neto, Clarissa Damaso, Rosane Silva |
| EPI_ISL_492038 | Instituto de Biologia do Exército | Laboratório Metabolismo Macromolecular FirminoTorres de Castro, Instituto de Biofísica Carlos Chagas Filho, Universidade Federal do Rio de Janeiro | Bianca Catarina Azevedo Cabral, Aline Rosa Vianna de Souza, Nádia Vaez Gonçalves da Cruz, Caleb GM Santos, Marcos Dornelas-Ribeiro, Tatiana LS Nogueira, Elizabeth Valentin, Marcio da Costa Cipitelli, Virginia Sara Grancieri do Amaral, Rodrigo Soares de Moura Neto, Clarissa Damaso, Rosane Silva |
| EPI_ISL_492039 | Instituto de Biologia do Exército | Laboratório Metabolismo Macromolecular FirminoTorres de Castro, Instituto de Biofísica Carlos Chagas Filho, Universidade Federal do Rio de Janeiro | Bianca Catarina Azevedo Cabral, Aline Rosa Vianna de Souza, Tatiana LS Nogueira, Nádia Vaez Gonçalves da Cruz, Caleb GM Santos, Marcos Dornelas-Ribeiro, Elizabeth Valentin, Marcio da Costa Cipitelli, Virginia Sara Grancieri do Amaral, Rodrigo Soares de Moura Neto, Clarissa Damaso, Rosane Silva |
| EPI_ISL_492040 | Instituto de Biologia do Exército | Laboratório Metabolismo Macromolecular FirminoTorres de Castro, Instituto de Biofísica Carlos Chagas Filho, Universidade Federal do Rio de Janeiro | Bianca Catarina Azevedo Cabral, Aline Rosa Vianna de Souza , Marcos Dornelas-Ribeiro, Tatiana LS Nogueira, Nádia Vaez Gonçalves da Cruz, Caleb GM Santos, Elizabeth Valentin, Marcio da Costa Cipitelli, Virginia Sara Grancieri do Amaral, Rodrigo Soares de Moura Neto, Clarissa Damaso, Rosane Silva |
| EPI_ISL_492041 | Instituto de Biologia do Exército | Laboratório Metabolismo Macromolecular FirminoTorres de Castro, Instituto de Biofísica Carlos Chagas Filho, Universidade Federal do Rio de Janeiro | Bianca Catarina Azevedo Cabral, Aline Rosa Vianna de Souza, Caleb GM Santos, Marcos Dornelas-Ribeiro, Tatiana LS Nogueira, Nádia Vaez Gonçalves da Cruz, Elizabeth Valentin, Marcio da Costa Cipitelli, Virginia Sara Grancieri do Amaral, Rodrigo Soares de Moura Neto, Clarissa Damaso, Rosane Silva |
| EPI_ISL_492042 | Instituto de Biologia do Exército | Laboratório Metabolismo Macromolecular FirminoTorres de Castro, Instituto de Biofísica Carlos Chagas Filho, Universidade Federal do Rio de Janeiro | Bianca Catarina Azevedo Cabral, Aline Rosa Vianna de Souza, Nádia Vaez Gonçalves da Cruz, Caleb GM Santos, Marcos Dornelas-Ribeiro, Tatiana LS Nogueira, Elizabeth Valentin, Marcio da Costa Cipitelli, Virginia Sara Grancieri do Amaral, Rodrigo Soares de Moura Neto, Clarissa Damaso, Rosane Silva |
| EPI_ISL_492043 | Instituto de Biologia do Exército | Laboratório Metabolismo Macromolecular FirminoTorres de Castro, Instituto de Biofísica Carlos Chagas Filho, Universidade Federal do Rio de Janeiro | Bianca Catarina Azevedo Cabral, Aline Rosa Vianna de Souza, Tatiana LS Nogueira, Nádia Vaez Gonçalves da Cruz, Caleb GM Santos, Marcos Dornelas-Ribeiro, Elizabeth Valentin, Marcio da Costa Cipitelli, Virginia Sara Grancieri do Amaral, Rodrigo Soares de Moura Neto, Clarissa Damaso, Rosane Silva |
| EPI_ISL_492044 | Instituto de Biologia do Exército | Laboratório Metabolismo Macromolecular FirminoTorres de Castro, Instituto de Biofísica Carlos Chagas Filho, Universidade Federal do Rio de Janeiro | Bianca Catarina Azevedo Cabral, Aline Rosa Vianna de Souza , Marcos Dornelas-Ribeiro, Tatiana LS Nogueira, Nádia Vaez Gonçalves da Cruz, Caleb GM Santos, Elizabeth Valentin, Marcio da Costa Cipitelli, Virginia Sara Grancieri do Amaral, Rodrigo Soares de Moura Neto, Clarissa Damaso, Rosane Silva |
| EPI_ISL_492045 | Instituto de Biologia do Exército | Laboratório Metabolismo Macromolecular FirminoTorres de Castro, Instituto de Biofísica Carlos Chagas Filho, Universidade Federal do Rio de Janeiro | Bianca Catarina Azevedo Cabral, Aline Rosa Vianna de Souza, Caleb GM Santos, Marcos Dornelas-Ribeiro, Tatiana LS Nogueira, Nádia Vaez Gonçalves da Cruz, Elizabeth Valentin, Marcio da Costa Cipitelli, Virginia Sara Grancieri do Amaral, Rodrigo Soares de Moura Neto, Clarissa Damaso, Rosane Silva |
| EPI_ISL_492048 | Instituto de Biologia do Exército | Laboratório Metabolismo Macromolecular FirminoTorres de Castro, Instituto de Biofísica Carlos Chagas Filho, Universidade Federal do Rio de Janeiro | Bianca Catarina Azevedo Cabral, Aline Rosa Vianna de Souza , Marcos Dornelas-Ribeiro, Tatiana LS Nogueira, Nádia Vaez Gonçalves da Cruz, Caleb GM Santos, Elizabeth Valentin, Marcio da Costa Cipitelli, Virginia Sara Grancieri do Amaral, Rodrigo Soares de Moura Neto, Clarissa Damaso, Rosane Silva |
| EPI_ISL_509430, EPI_ISL_509431, EPI_ISL_509432, EPI_ISL_509433, EPI_ISL_509434, EPI_ISL_509435 | Centro de Desenvolvimento Tecnologico em Saude, Fundacao Oswaldo Cruz | Centro de Desenvolvimento Tecnologico em Saude, Fundacao Oswaldo Cruz | Souza,T.M., Fintelman-Rodrigues,N., De Paula,A.D., Saraiva,F.B., Ferreira,M.A., Sacramento,C.Q., Medeiros,M.A. |
| EPI_ISL_510536 | Centro de Desenvolvimento Tecnologico em Saude, Fundacao Oswaldo Cruz | Centro de Desenvolvimento Tecnologico em Saude, Fundacao Oswaldo Cruz | Souza,T.M., Fintelman-Rodrigues,N., De Paula,A.D., Saraiva,F.B., Ferreira,M.A., Sacramento,C.Q. and Medeiros,M.A. |
| EPI_ISL_510541 | Centro de Desenvolvimento Tecnologico em Saude, Fundacao Oswaldo Cruz | Centro de Desenvolvimento Tecnologico em Saude, Fundacao Oswaldo Cruz | Souza,T.M., Fintelman-Rodrigues,N., De Paula,A.D., Saraiva,F.B., Ferreira,M.A., Sacramento,C.Q. and Medeiros,M.A. |
| EPI_ISL_513513, EPI_ISL_513514, EPI_ISL_513515, EPI_ISL_513516, EPI_ISL_513518, EPI_ISL_513519, EPI_ISL_513520, EPI_ISL_513521, EPI_ISL_513522, EPI_ISL_513524, EPI_ISL_513525, EPI_ISL_513526, EPI_ISL_513527, EPI_ISL_513529, EPI_ISL_513530, EPI_ISL_513531, EPI_ISL_513532, EPI_ISL_513533, EPI_ISL_513534, EPI_ISL_513535, EPI_ISL_513536, EPI_ISL_513537, EPI_ISL_513538, EPI_ISL_513539, EPI_ISL_513540, EPI_ISL_513541, EPI_ISL_513542, EPI_ISL_513543, EPI_ISL_513544, EPI_ISL_513545, EPI_ISL_513546, EPI_ISL_513547, EPI_ISL_513548, EPI_ISL_513549, EPI_ISL_513550, EPI_ISL_513551, EPI_ISL_513553, EPI_ISL_513556, EPI_ISL_513557, EPI_ISL_513558, EPI_ISL_513559, EPI_ISL_513560, EPI_ISL_513561, EPI_ISL_513562, EPI_ISL_513563, EPI_ISL_513564, EPI_ISL_513567, EPI_ISL_513568, EPI_ISL_513571, EPI_ISL_513572, EPI_ISL_513573, EPI_ISL_513574, EPI_ISL_513575, EPI_ISL_513576, EPI_ISL_513577, EPI_ISL_513578, EPI_ISL_513579, EPI_ISL_513580, EPI_ISL_513581, EPI_ISL_513582, EPI_ISL_513583 | | | |
| see above | Programa de Oncovirologia, Instituto Nacional de Câncer | Programa de Oncovirologia, Instituto Nacional de Câncer | Juliana D. Siqueira, Livia R. Goes, Brunna M. Alves, Claudia Cicala,James Arthos, João P.B. Viola, Andreia C. de Melo, Marcelo A. Soares |
| EPI_ISL_528539 | LVM/UFRJ | LNCC | Gustavo M. Romário M. de Souza; Bruno B. Bezerra; Lucio A. Caldas; Fabio Limonte; Elena Cobos; Sharton V. A. Coelho; Luiz Almeida; Luiza Higga; Isadora A. Correa; Diana Marianni; Luciana B. Arruda; Marcelo Bozza; Orlando Ferreira; Wanderley de Souza; Ana Teresa R. Vasconcelos; Terezinha M. Castineiras; Amilcar Tanuri; Luciana J. Costa. |
| EPI_ISL_528637, EPI_ISL_528638 | LVM/UFRJ | Bioinformatics Laboratory / LNCC | Gustavo D. P. Silva; M. Romário M. de Souza; Bruno B. Bezerra; Lucio A. Caldas; Fabio Limonte; Elena Cobos; Sharton V. A. Coelho; Luiz Almeida; Luiza Higga; Isadora A. Correa; Diana Marianni; Luciana B. Arruda; Marcelo Bozza; Orlando Ferreira; Wanderley de Souza; Ana Teresa R. Vasconcelos; Terezinha M. Castineiras; Amilcar Tanuri; Luciana J. Costa |
| EPI_ISL_529139 | Centro de Desenvolvimento Tecnologico em Saude, Fundacao Oswaldo Cruz | Centro de Desenvolvimento Tecnologico em Saude, Fundacao Oswaldo Cruz | Souza,T.M., Fintelman-Rodrigues,N., De Paula,A.D., Saraiva,F.B., Ferreira,M.A., Sacramento,C.Q., Medeiros,M.A. |

| | | | |
|---|---|---|---|
| EPI_ISL_529140 | Centro de Desenvolvimento Tecnologico em Saude, Fundacao Oswaldo Cruz | Centro de Desenvolvimento Tecnologico em Saude, Fundacao Oswaldo Cruz | Souza,T.M., Fintelman-Rodrigues,N., De Paula,A.D., Saraiva,F.B., Ferreira,M.A., Sacramento,C.Q., Medeiros,M.A. |
| EPI_ISL_541347, EPI_ISL_541348, EPI_ISL_541349, EPI_ISL_541350, EPI_ISL_541351 | Laboratory of Respiratory Viruses and Measles, Oswaldo Cruz Institute, FIOCRUZ | Laboratory of Respiratory Viruses and Measles, Oswaldo Cruz Institute, FIOCRUZ | Paola Resende, Luciana Appolinario, Fernando Motta, Anna Carolina Paixão, Ana Carolina Mendonça, Jonathan Lopes, Marilda Siqueira on behalf of the Fiocruz COVID-19 Genomic Surveillance Network |
| EPI_ISL_541352 | Laboratory of Respiratory Viruses and Measles, Oswaldo Cruz Institute, FIOCRUZ | Laboratory of Respiratory Viruses and Measles, Oswaldo Cruz Institute, FIOCRUZ | Paola Resende, Roxana Loayza, Cinthia Avila, Luciana Appolinario, Fernando Motta, Anna Carolina Paixao, Ana Carolina Mendonca, Marilda Siqueira on behalf of the Fiocruz COVID-19 Genomic Surveillance Network |
| EPI_ISL_541353, EPI_ISL_541354, EPI_ISL_541355 | Laboratory of Respiratory Viruses and Measles, Oswaldo Cruz Institute, FIOCRUZ | Laboratory of Respiratory Viruses and Measles, Oswaldo Cruz Institute, FIOCRUZ | Paola Resende, Luciana Appolinario, Fernando Motta, Anna Carolina Paixão, Ana Carolina Mendonça, Jonathan Lopes, Marilda Siqueira on behalf of the Fiocruz COVID-19 Genomic Surveillance Network |
| EPI_ISL_541356 | Laboratory of Respiratory Viruses and Measles, Oswaldo Cruz Institute, FIOCRUZ | Laboratory of Respiratory Viruses and Measles, Oswaldo Cruz Institute, FIOCRUZ | Paola Resende, Roxana Loayza, Cinthia Avila, Luciana Appolinario, Fernando Motta, Anna Carolina Paixao, Ana Carolina Mendonca, Marilda Siqueira on behalf of the Fiocruz COVID-19 Genomic Surveillance Network |
| EPI_ISL_541357, EPI_ISL_541358 | Laboratory of Respiratory Viruses and Measles, Oswaldo Cruz Institute, FIOCRUZ | Laboratory of Respiratory Viruses and Measles, Oswaldo Cruz Institute, FIOCRUZ | Paola Resende, Luciana Appolinario, Fernando Motta, Anna Carolina Paixão, Ana Carolina Mendonça, Jonathan Lopes, Marilda Siqueira on behalf of the Fiocruz COVID-19 Genomic Surveillance Network |
| EPI_ISL_541359, EPI_ISL_541360, EPI_ISL_541361 | Laboratory of Respiratory Viruses and Measles, Oswaldo Cruz Institute, FIOCRUZ | Laboratory of Respiratory Viruses and Measles, Oswaldo Cruz Institute, FIOCRUZ | Paola Resende, Roxana Loayza, Cinthia Avila, Luciana Appolinario, Fernando Motta, Anna Carolina Paixao, Ana Carolina Mendonca, Marilda Siqueira on behalf of the Fiocruz COVID-19 Genomic Surveillance Network |
| EPI_ISL_541362, EPI_ISL_541363 | Laboratory of Respiratory Viruses and Measles, Oswaldo Cruz Institute, FIOCRUZ | Laboratory of Respiratory Viruses and Measles, Oswaldo Cruz Institute, FIOCRUZ | Paola Resende, Luciana Appolinario, Fernando Motta, Anna Carolina Paixão, Ana Carolina Mendonça, Jonathan Lopes, Marilda Siqueira on behalf of the Fiocruz COVID-19 Genomic Surveillance Network |
| EPI_ISL_541364, EPI_ISL_541365, EPI_ISL_541366 | Laboratory of Respiratory Viruses and Measles, Oswaldo Cruz Institute, FIOCRUZ | Laboratory of Respiratory Viruses and Measles, Oswaldo Cruz Institute, FIOCRUZ | Paola Resende, Roxana Loayza, Cinthia Avila, Luciana Appolinario, Fernando Motta, Anna Carolina Paixao, Ana Carolina Mendonca, Marilda Siqueira on behalf of the Fiocruz COVID-19 Genomic Surveillance Network |
| EPI_ISL_541367 | Laboratory of Respiratory Viruses and Measles, Oswaldo Cruz Institute, FIOCRUZ | Laboratory of Respiratory Viruses and Measles, Oswaldo Cruz Institute, FIOCRUZ | Paola Resende, Luciana Appolinario, Fernando Motta, Anna Carolina Paixão, Ana Carolina Mendonça, Jonathan Lopes, Marilda Siqueira on behalf of the Fiocruz COVID-19 Genomic Surveillance Network |
| EPI_ISL_541368, EPI_ISL_541369 | Laboratory of Respiratory Viruses and Measles, Oswaldo Cruz Institute, FIOCRUZ | Laboratory of Respiratory Viruses and Measles, Oswaldo Cruz Institute, FIOCRUZ | Paola Resende, Roxana Loayza, Cinthia Avila, Luciana Appolinario, Fernando Motta, Anna Carolina Paixao, Ana Carolina Mendonca, Marilda Siqueira on behalf of the Fiocruz COVID-19 Genomic Surveillance Network |
| EPI_ISL_623108, EPI_ISL_623110, EPI_ISL_623112, EPI_ISL_623114, EPI_ISL_623116, EPI_ISL_623118, EPI_ISL_623119, EPI_ISL_623120, EPI_ISL_623124, EPI_ISL_623126, EPI_ISL_623129, EPI_ISL_623130, EPI_ISL_623132, EPI_ISL_623134, EPI_ISL_623136, EPI_ISL_623138, EPI_ISL_623140, EPI_ISL_623142, EPI_ISL_623143, EPI_ISL_623144, EPI_ISL_623145, EPI_ISL_623146, EPI_ISL_623147, EPI_ISL_623148, EPI_ISL_623149, EPI_ISL_623152, EPI_ISL_623154, EPI_ISL_623157, EPI_ISL_623158, EPI_ISL_623160, EPI_ISL_623161, EPI_ISL_623162, EPI_ISL_623164, EPI_ISL_623165, EPI_ISL_623166, EPI_ISL_623168 | | | |
| see above | Laboratorio de Virologia Molecular / UFRJ | Bioinformatics Laboratory / LNCC | Carolina M Voloch, Ronaldo S Francisco Jr, Luiz G P de Almeida, Otavio J. Brustolini, Cynthia C Cardoso, Alexandra L Gerber, Ana Paula de C Guimarães, Diana Mariani, Covid19-UFRJ Workgroup, Luís Cristóvão Pôrto, Renato S Aguiar, Terezinha M P P Castiñeiras, Orlando C. Ferreira, Amilcar Tanuri, Ana Tereza R de Vasconcelos |
| EPI_ISL_636737, EPI_ISL_636834, EPI_ISL_636835, EPI_ISL_636837 | Laboratório de Imunofarmacologia - Instituto Oswaldo Cruz | Laboratório de Imunofarmacologia - Instituto Oswaldo Cruz | Souza,T.M., Fintelman-Rodrigues,N., De Paula,A.D., Saraiva,F.B., Ferreira,M.A. and Sacramento,C.Q. |
| EPI_ISL_717806, EPI_ISL_717807, EPI_ISL_717808 | Laboratorio de Virologia Molecular / UFRJ | Bioinformatics Laboratory / LNCC | Carolina M Voloch, Ronaldo da Silva F Jr, Luiz G P de Almeida, Cynthia C Cardoso, Otavio Bustrolini, Alexandra L Gerber, Ana Paula de C Guimarães, Diana Mariani, Andréa Cony Cavalcanti, Claudia dos Santos Rodrigues, Terezinha M P P Castiñeira, Amilcar Tanuri, Ana Tereza R de Vasconcelos |
| EPI_ISL_717809 | LACEN Dr. Francisco Rimolo Neto | Bioinformatics Laboratory / LNCC | Carolina M Voloch, Ronaldo da Silva F Jr, Luiz G P de Almeida, Cynthia C Cardoso, Otavio Bustrolini, Alexandra L Gerber, Ana Paula de C Guimarães, Diana Mariani, Andréa Cony Cavalcanti, Claudia dos Santos Rodrigues, Terezinha M P P Castiñeira, Amilcar Tanuri, Ana Tereza R de Vasconcelos |
| EPI_ISL_717810, EPI_ISL_717811, EPI_ISL_717812, EPI_ISL_717813, EPI_ISL_717814, EPI_ISL_717815, EPI_ISL_717816, EPI_ISL_717817, EPI_ISL_717818, EPI_ISL_717819, EPI_ISL_717820, EPI_ISL_717821, EPI_ISL_717822, EPI_ISL_717823, EPI_ISL_717824, EPI_ISL_717825, EPI_ISL_717826, EPI_ISL_717827, EPI_ISL_717828, EPI_ISL_717829, EPI_ISL_717830, EPI_ISL_717831 | | | |
| see above | Laboratorio de Virologia Molecular / UFRJ | Bioinformatics Laboratory / LNCC | Carolina M Voloch, Ronaldo da Silva F Jr, Luiz G P de Almeida, Cynthia C Cardoso, Otavio Bustrolini, Alexandra L Gerber, Ana Paula de C Guimarães, Diana Mariani, Andréa Cony Cavalcanti, Claudia dos Santos Rodrigues, Terezinha M P P Castiñeira, Amilcar Tanuri, Ana Tereza R de Vasconcelos |
| EPI_ISL_717832, EPI_ISL_717833, EPI_ISL_717834, EPI_ISL_717835, EPI_ISL_717836 | LACEN Dr. Francisco Rimolo Neto | Bioinformatics Laboratory / LNCC | Carolina M Voloch, Ronaldo da Silva F Jr, Luiz G P de Almeida, Cynthia C Cardoso, Otavio Bustrolini, Alexandra L Gerber, Ana Paula de C Guimarães, Diana Mariani, Andréa Cony Cavalcanti, Claudia dos Santos Rodrigues, Terezinha M P P Castiñeira, Amilcar Tanuri, Ana Tereza R de Vasconcelos |
| EPI_ISL_717837, EPI_ISL_717838, EPI_ISL_717839, EPI_ISL_717840 | Laboratorio de Virologia Molecular / UFRJ | Bioinformatics Laboratory / LNCC | Carolina M Voloch, Ronaldo da Silva F Jr, Luiz G P de Almeida, Cynthia C Cardoso, Otavio Bustrolini, Alexandra L Gerber, Ana Paula de C Guimarães, Diana Mariani, Andréa Cony Cavalcanti, Claudia dos Santos Rodrigues, Terezinha M P P Castiñeira, Amilcar Tanuri, Ana Tereza R de Vasconcelos |
| EPI_ISL_717841 | LACEN Dr. Francisco Rimolo Neto | Bioinformatics Laboratory / LNCC | Carolina M Voloch, Ronaldo da Silva F Jr, Luiz G P de Almeida, Cynthia C Cardoso, Otavio Bustrolini, Alexandra L Gerber, Ana Paula de C Guimarães, Diana Mariani, Andréa Cony Cavalcanti, Claudia dos Santos Rodrigues, Terezinha M P P Castiñeira, Amilcar Tanuri, Ana Tereza R de Vasconcelos |
| EPI_ISL_717842, EPI_ISL_717843, EPI_ISL_717844, EPI_ISL_717845, EPI_ISL_717846, EPI_ISL_717847, EPI_ISL_717848, EPI_ISL_717849, EPI_ISL_717850, EPI_ISL_717851, EPI_ISL_717852, EPI_ISL_717853, EPI_ISL_717854, EPI_ISL_717855, EPI_ISL_717856, EPI_ISL_717857, EPI_ISL_717858, EPI_ISL_717859, EPI_ISL_717860, EPI_ISL_717861, EPI_ISL_717862, EPI_ISL_717863, EPI_ISL_717864, EPI_ISL_717865, EPI_ISL_717866, EPI_ISL_717867, EPI_ISL_717868, EPI_ISL_717869, EPI_ISL_717870, EPI_ISL_717871, EPI_ISL_717872, EPI_ISL_717873, EPI_ISL_717874, EPI_ISL_717875, EPI_ISL_717876, EPI_ISL_717877, EPI_ISL_717878, EPI_ISL_717879, EPI_ISL_717880, EPI_ISL_717881, EPI_ISL_717882, EPI_ISL_717883, EPI_ISL_717884, EPI_ISL_717885, EPI_ISL_717886, EPI_ISL_717887, EPI_ISL_717888, EPI_ISL_717889, EPI_ISL_717890, EPI_ISL_717891, EPI_ISL_717892, EPI_ISL_717893, EPI_ISL_717894, EPI_ISL_717895, EPI_ISL_717896, EPI_ISL_717897, EPI_ISL_717898 | | | |
| see above | Laboratorio de Virologia Molecular / UFRJ | Bioinformatics Laboratory / LNCC | Carolina M Voloch, Ronaldo da Silva F Jr, Luiz G P de Almeida, Cynthia C Cardoso, Otavio Bustrolini, Alexandra L Gerber, Ana Paula de C Guimarães, Diana Mariani, Andréa Cony Cavalcanti, Claudia dos Santos Rodrigues, Terezinha M P P Castiñeira, Amilcar Tanuri, Ana Tereza R de Vasconcelos |
| EPI_ISL_717899, EPI_ISL_717900, EPI_ISL_717901, EPI_ISL_717902, EPI_ISL_717903, EPI_ISL_717904, EPI_ISL_717905, EPI_ISL_717906, EPI_ISL_717907, EPI_ISL_717908, EPI_ISL_717909 | | | |
| see above | LACEN RJ - Noel Nutels | Bioinformatics Laboratory / LNCC | Carolina M Voloch, Ronaldo da Silva F Jr, Luiz G P de Almeida, Cynthia C Cardoso, Otavio Bustrolini, Alexandra L Gerber, Ana Paula de C Guimarães, Diana Mariani, Andréa Cony Cavalcanti, Claudia dos Santos Rodrigues, Terezinha M P P Castiñeira, Amilcar Tanuri, Ana Tereza R de Vasconcelos |
| EPI_ISL_717910, EPI_ISL_717911, EPI_ISL_717912, EPI_ISL_717913, EPI_ISL_717914, EPI_ISL_717915, EPI_ISL_717916, EPI_ISL_717917, EPI_ISL_717918, EPI_ISL_717919 | LACEN Dr. Francisco Rimolo Neto | Bioinformatics Laboratory / LNCC | Carolina M Voloch, Ronaldo da Silva F Jr, Luiz G P de Almeida, Cynthia C Cardoso, Otavio Bustrolini, Alexandra L Gerber, Ana Paula de C Guimarães, Diana Mariani, Andréa Cony Cavalcanti, Claudia dos Santos Rodrigues, Terezinha M P P Castiñeira, Amilcar Tanuri, Ana Tereza R de Vasconcelos |
| EPI_ISL_717920, EPI_ISL_717921, EPI_ISL_717922, EPI_ISL_717923, EPI_ISL_717924, EPI_ISL_717925, EPI_ISL_717926, EPI_ISL_717927, EPI_ISL_717928, EPI_ISL_717929, EPI_ISL_717930, EPI_ISL_717931, EPI_ISL_717932, EPI_ISL_717933, EPI_ISL_717934, EPI_ISL_717935, EPI_ISL_717936, EPI_ISL_717937, EPI_ISL_717938, EPI_ISL_717939, EPI_ISL_717940, EPI_ISL_717941, EPI_ISL_717942, EPI_ISL_717943, EPI_ISL_717944, EPI_ISL_717945, EPI_ISL_717946, EPI_ISL_717947, EPI_ISL_717948, EPI_ISL_717949, EPI_ISL_717950, EPI_ISL_717951, EPI_ISL_717952, EPI_ISL_717953, EPI_ISL_717954, EPI_ISL_717955, EPI_ISL_717956, EPI_ISL_717957 | | | |
| see above | Laboratorio de Virologia Molecular / UFRJ | Bioinformatics Laboratory / LNCC | Carolina M Voloch, Ronaldo da Silva F Jr, Luiz G P de Almeida, Cynthia C Cardoso, Otavio Bustrolini, Alexandra L Gerber, Ana Paula de C Guimarães, Diana Mariani, Andréa Cony Cavalcanti, Claudia dos Santos Rodrigues, Terezinha M P P Castiñeira, Amilcar Tanuri, Ana Tereza R de Vasconcelos |
| EPI_ISL_717958 | LACEN Dr. Francisco Rimolo Neto | Bioinformatics Laboratory / LNCC | Carolina M Voloch, Ronaldo da Silva F Jr, Luiz G P de Almeida, Cynthia C Cardoso, Otavio Bustrolini, Alexandra L Gerber, Ana Paula de C Guimarães, Diana Mariani, Andréa Cony Cavalcanti, Claudia dos Santos Rodrigues, Terezinha M P P Castiñeira, Amilcar Tanuri, Ana Tereza R de Vasconcelos |
| EPI_ISL_717959, EPI_ISL_717960, EPI_ISL_717961 | Laboratorio de Virologia Molecular / UFRJ | Bioinformatics Laboratory / LNCC | Carolina M Voloch, Ronaldo da Silva F Jr, Luiz G P de Almeida, Cynthia C Cardoso, Otavio Bustrolini, Alexandra L Gerber, Ana Paula de C Guimarães, Diana Mariani, Andréa Cony Cavalcanti, Claudia dos Santos Rodrigues, Terezinha M P P Castiñeira, Amilcar Tanuri, Ana Tereza R de Vasconcelos |
| EPI_ISL_717962 | LACEN RJ - Noel Nutels | Bioinformatics Laboratory / LNCC | Carolina M Voloch, Ronaldo da Silva F Jr, Luiz G P de Almeida, Cynthia C Cardoso, Otavio Bustrolini, Alexandra L Gerber, Ana Paula de C Guimarães, Diana Mariani, Andréa Cony Cavalcanti, Claudia dos Santos Rodrigues, Terezinha M P P Castiñeira, Amilcar Tanuri, Ana Tereza R de Vasconcelos |
| EPI_ISL_717963, EPI_ISL_717964 | LACEN Dr. Francisco Rimolo Neto | Bioinformatics Laboratory / LNCC | Carolina M Voloch, Ronaldo da Silva F Jr, Luiz G P de Almeida, Cynthia C Cardoso, Otavio Bustrolini, Alexandra L Gerber, Ana Paula de C Guimarães, Diana Mariani, Andréa Cony Cavalcanti, Claudia dos Santos Rodrigues, Terezinha M P P Castiñeira, Amilcar Tanuri, Ana Tereza R de Vasconcelos |
